# Detection of B.1.351 and B.1.1.7 SARS-CoV-2 variants in Monterrey metropolitan area in Northeast Mexico

**DOI:** 10.1101/2021.06.21.21258579

**Authors:** Kame A. Galán-Huerta, Eduardo Garza-de-la-Peña, Gabriela V. Elizondo-Valdez, María F. Herrera-Saldivar, Ana M. Rivas-Estilla, Sonia A. Lozano-Sepúlveda, Natalia Martínez-Acuña, Daniel Arellanos-Soto, Javier Ramos-Jiménez

**Affiliations:** Centro de Investigación e Innovación en Virología Médica, Departamento de Bioquímica y Medicina Molecular, Facultad de Medicina, Universidad Autónoma de Nuevo León; Laboratorio de Patología Clínica y Genética Molecular; Servicio de Infectología, Hospital Universitario “Dr. José Eleuterio González”, Universidad Autónoma de Nuevo León

**Keywords:** SARS-CoV-2, Variant of concern, Phylogeny, Mexico

## Abstract

SARS-CoV-2 variants of concern (VOC) have spread throughout the world. In Mexico, VOC B.1.351 has been detected on one occasion and B.1.1.7 several times. We detected lineages B.1.351 and B.1.1.7 in patients who traveled to USA and B.1.1.7 in a patient with no travel history.

**Article summary line:** Introduction of B.1.351 and B.1.1.7 S ARS-CoV-2 variants in Monterrey, Mexico.

---

Since late 2020, several variants of SARS-CoV-2 have emerged. These variants, named variants of concern (VOC), are monitored because they are associated with an increase in transmissibility, increase in severity, and vaccine effectiveness reduction (*1-3*). VOC Alpha (Lineage B.1.1.7) was first detected in Mexico in December 2020, and Nuevo Leon in January 2021. VOC Beta (Lineage B.1.351) was detected in Mexico in April 2021 and not previously detected in Nuevo Leon.

We randomly selected, via randomizer.org, residual RNA samples derived from nasopharyngeal swabs from patients living in Nuevo Leon, Mexico. Samples were positive for SARS-CoV-2 and had Ct values <30 in TaqPath COVID□19 CE□IVD RT□PCR Kit (Thermo Fisher Scientific, Waltham, MA). We used samples after a two-week retention period. To maintain anonymity, samples were deidentified and only age and gender were obtained. We sequenced the whole genome with Artic Network’s amplicon sequencing protocol (*4*) in a MinION (Oxford Nanopore Technologies, Oxford, UK).

Global reference sequences were selected from Nextstrain’s global dataset (n=3644) and all the B.1.1.7 and B.1.351 genomes from Mexico until April 29^th^, 2021 (n=77) and retrieved them from the GISAID database. We did a BLAST search for each of the recently obtained sequences and retrieved the five most similar sequences.

Afterwards, we used Nextstrain (*5*) to analyze the newly obtained sequences and to get the global perspective of our data. Posteriorly, we made two alignments. One with all the sequences, from lineage B.1.1.7, that were included in the clades where the sequenced samples were grouped. The other alignment included all the sequences belonging to the lineage B.1.351. We constructed a maximum-likelihood tree in IQ-TREE v2.0.3 (*6*) for each alignment, searched for temporal data in TempEst v1.5.3 and eliminated outlier sequences shown in the root-to-tip plot.

Next, we inferred maximum clade credibility trees for each alignment with BEAST v1.10.4 (*7*). We used a strict molecular clock, HKY+G substitution model, and a coalescent exponential growth tree prior. We used a chain length of 100 million and sampled every 10,000 steps. Chain convergence was evaluated using Tracer v.1.7.1 and trees were summarized using TreeAnnotator discarding 10% as burn-in. Sequences generated in this report were deposited in GISAID with the following numbers: EPI_ISL_1789696 - EPI_ISL_1789699.

The Ethics Committee from Facultad de Medicina y Hospital Universitario Universidad Autónoma de Nuevo León reviewed and approved this work with the following registration number: BI20-0004.

We detected lineage B.1.351 in one patient and lineage B.1.1.7 in three patients. The four patients were males and three of them had traveled to USA in the previous two weeks. Lineage B.1.351 was detected in a middle-aged man with recent travel to Pennsylvania, USA. He had no contact with similar cases and had onset of symptoms the second week of April. Lineage B.1.1.7 was detected in two teenagers and one middle-aged man. The two teenagers had recent travel history to Colorado, USA, contact with similar cases and had onset of symptoms the first week of April. The middle-aged man had no recent travel history, no contact with similar cases and had onset of symptoms the second week of April. These four patients were treated ambulatory.

Initial phylogenetic analysis places the sample NLE-UANL-074 in lineage B.1.351 and samples NLE-UANL-067, 068 and 075 in lineage B.1.1.7 (Figure panel A). Until May 2021, there were only three B.1.351 viruses detected in Mexico: two from Campeche, and one from Baja California Norte. Sample NLE-UANL-074 groups in a well-supported clade with viruses isolated in Pennsylvania state (Figure panel B and D). This clade does not group with the previously reported samples from Mexico, confirming an introduction.

**Figure.**
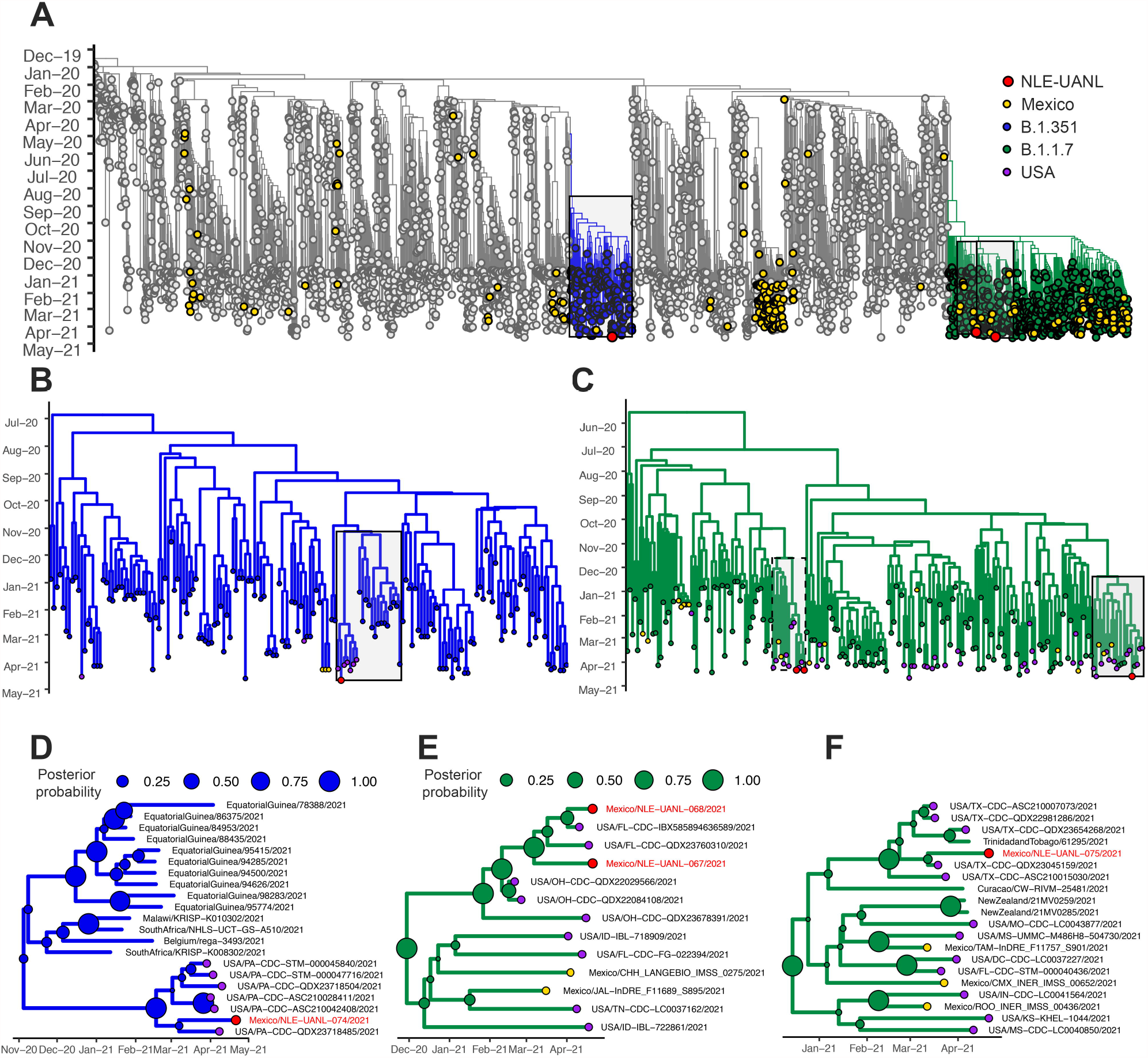
Phylogenetic relationship of SARS-CoV-2, 2019-2021. A) Global context tree highlighting lineages B.1.351 (blue) and B.1.1.7 (green). Black boxes show sequences that were analyzed. B) Phylogenetic tree of B.1.351 lineage showing the relationship of samples from this report with samples from Mexico and USA. The black box shows posterior magnification. C) Phylogenetic tree of B.1.1.7 lineage showing the relationship of samples from this report with samples from Mexico and USA. Black boxes show posterior magnification. D) Magnification of B.1.351 tree showing the clade in which the sequenced virus groups. E) Magnification of the discontinuous box in the B.1.1.7 tree showing the relationship of the sequenced viruses. F) Magnification of the continuous box in the B.1.1.7 tree showing the relationship of the sequenced virus. Posterior probabilities are shown in the nodes of the magnifications. Viruses reported in this research letter are in red. Phylogenetic trees were visualized in ggtree (*8*). Sequences used in this analysis were downloaded from GISAID. The authors of sequences used are listed in the Appendix Table.

B.1.1.7 lineage was previously introduced to Mexico and explains the samples dispersed though the phylogenetic tree (Figure panel C). Further analysis revealed that samples NLE-UANL-067 and 068 grouped with samples isolated in Florida in a well-supported clade (Figure panel E). These viruses differ from previously reported in Mexico, suggesting an introduction from USA. Sample NLE-UANL-075, isolated from the patient without travel history, groups with viruses isolated in Texas in a well-supported clade (Figure panel F). This finding could be explained if the patient had contact with people who traveled to Texas and were pre-symptomatic.

Even though samples NLE-UANL-067, 068 and 075 belong to lineage B.1.1.7 and were isolated in Nuevo Leon, they are not related because they group in different well-supported clades. Even though lineage B.1.1.7 was previously detected in Nuevo Leon, there was no widespread transmission throughout the state.

This is the first report of B.1.351 and the third detection of B.1.1.7 in Nuevo Leon state. We need posterior surveillance to evaluate community transmission and association of these lineages with a new wave of COVID-19 cases.

## Data Availability

Sequences generated in this report were deposited in GISAID with the following numbers: EPI_ISL_1789696 - EPI_ISL_1789699.

## Acknowledgments

This work was funded by Consejo Nacional de Ciencia y Tecnología (CONACyT) with the following grant number 312328, awarded to Kame A. Galán-Huerta. The authors gratefully acknowledge the authors and submitting laboratories that generated genome data and shared it via GISAID. A full list acknowledging the authors submitting data used in this report can be found in Appendix Table.

## Author bio

Dr. Galán-Huerta is an assistant professor and founder member of the Center for Investigation and Innovation in Medical Virology, in the Faculty of Medicine of the Autonomous University of Nuevo Leon. His primary research interests are emerging and reemerging viral diseases and epidemiology.

