## Appendix table for "Detection of B.1.351 and B.1.1.7 SARS-CoV-2 variants in Monterrey metropolitan area in Northeast Mexico"

We gratefully acknowledge the following Authors from the Originating laboratories responsible for obtaining the specimens, as well as the Submitting laboratories where the genome data were generated and shared via GISAID, on which this research is based.

All Submitters of data may be contacted directly via www.gisaid.org

Authors are sorted alphabetically.

| Accession ID | Originating Laboratory | Submitting Laboratory | Authors |
|---|---|---|---|
| EPI_ISL_1001004 | Bundeswehr Institute of Microbiology | Bundeswehr Institute of Microbiology | Markus Antwerpen, Alexandra Rehn, Mathias Walter, Malena Bestehorn-Willmann, Sabine Zange, Enrico Georgi, Roman Wölfel |
| EPI_ISL_1001167 | National Virus Reference Laboratory | National Virus Reference Laboratory | Michael Carr, Gabriel Gonzalez, Zoe Yandle, Jonathan Dean, Cillian F De Gascun |
| EPI_ISL_1002335 | Viollier AG | Department of Biosystems Science and Engineering, ETH Zürich | Chaoran Chen, Sarah Nadeau, Catharine Aquino, Ivan Topolsky, Philipp Jablonski, Lara Fuhrmann, David Dreifuss, Katharina Jahn, Andreia Cabral de Gouvea, Maria Domenica Moccia, Simon Grüter, Timothy Sykes, Lennart Opitz, Griffin White, Laura Neff, Doris Popovic, Andrea Patrignani, Jay Tracy, Ralph Schlapbach, Christiane Beckmann, Maurice Redondo, Olivier Kobel, Christoph Noppen, Sophie Seidel, Noemie Santamaria de Souza, Niko Beerenwinkel, Tanja Stadler |
| EPI_ISL_1004884 | Viollier AG | Department of Biosystems Science and Engineering, ETH Zürich | Chaoran Chen, Sarah Nadeau, Ivan Topolsky, Emmanouil Dermitzakis, Keith Harshman, Ioannis Xenarios, Henri Pegeot, Lorenzo Cerutti, Deborah Penet, Philipp Jablonski, Lara Fuhrmann, David Dreifuss, Katharina Jahn, Christiane Beckmann, Maurice Redondo, Olivier Kobel, Christoph Noppen, Sophie Seidel, Noemie Santamaria de Souza, Niko Beerenwinkel, Tanja Stadler |
| EPI_ISL_1005309 | Vilnius university hospital Santaros Klinikos, Center of Laboratory Medicine | Vilnius university hospital Santaros Klinikos, Center of Laboratory Medicine | Gytis Dudas, Ingrida Olendraite, Justinas Slikas, Daniel Naumovas, Rimvydas Norvilas, Dovile Ezerskyte, Laimonas Griskevicius |
| EPI_ISL_1005697 | RSUD Dr. Tjitrowardojo | Genetics Working Group (Pokja Genetik) Faculty of Medicine, Public Health and Nursing Universitas Gadjah Mada (FK-KMK UGM); Disease Investigation Center Wates Ministry of Agriculture Indonesia; Department of Microbiology FK-KMK UGM; Laboratorium Diagnostik Yayasan Tahija World Mosquito Program (WMP) Yogyakarta Center for Tropical Medicine FK-KMK UGM; Integrated Research Center FK-KMK UGM; Department of Computer Science and Electronics FMIPA UGM; Balai Besar Teknik Kesehatan Lingkungan dan Pengendalian Penyakit (BBTKLPP) Yogyakarta | Gunadi, Hendra Wibawa, Marcellus, Mohamad S. Hakim, Edwin W. Daniwijaya, Ludhang P. Rizki, Endah Supriyati, Eggi Arguni, Titik Nuryastuti, Tri Wibawa, Dwi AA Nugrahaningsih, Afiahayati, Siswanto, Kristy Iskandar, Nungki Anggorowati, Irene, Indaryati, Havid Setyawan, Arry Setyowati, Chusni Mubarakh, Try Nirmala Sari, Kemala Athollah, Dyah Ayu Puspitarani |
| EPI_ISL_1007652 | Division of Emerging Infectious Diseases, Bureau of Infectious Diseases Diagnosis Control, Korea Disease Control and Prevention Agency | Division of Emerging Infectious Diseases, Bureau of Infectious Diseases Diagnosis Control, Korea Disease Control and Prevention Agency | Ae Kyung Park, Il-Hwan Kim, Heui Man Kim, Jeong-Min Kim, Namjoo Lee, Chae Young Lee, Sang Hee Woo, Eun-Jin Kim |
| EPI_ISL_1008045, EPI_ISL_1008107, EPI_ISL_1008144, EPI_ISL_1008167 | Austrian Agency for Health and Food Safety (AGES) | Bergthaler laboratory, CeMM Research Center for Molecular Medicine of the Austrian Academy of Sciences | Lukas Endler, Anna Schedl, Thomas Penz, Benedikt Agerer, Maelle Le Moing, Michael Schuster, Bekir Erguner, Jan Laine, Martin Senekowitsch, Christoph Bock, Andreas Bergthaler |
| EPI_ISL_1008270 | Institute for Water Quality and Resource Management, Technical University Vienna | Bergthaler laboratory, CeMM Research Center for Molecular Medicine of the Austrian Academy of Sciences | Lukas Endler, Anna Schedl, Thomas Penz, Benedikt Agerer, Maelle Le Moing, Michael Schuster, Bekir Erguner, Jan Laine, Martin Senekowitsch, Christoph Bock, Andreas Bergthaler |
| EPI_ISL_1008285, EPI_ISL_1008286 | Department of Microbiology, University Innsbruck | Bergthaler laboratory, CeMM Research Center for Molecular Medicine of the Austrian Academy of Sciences | Lukas Endler, Anna Schedl, Thomas Penz, Benedikt Agerer, Maelle Le Moing, Michael Schuster, Bekir Erguner, Jan Laine, Martin Senekowitsch, Christoph Bock, Andreas Bergthaler |
| EPI_ISL_1008367 | Department of Laboratory Medicine, Division of Clinical Virology, University of Medicine, Vienna | Bergthaler laboratory, CeMM Research Center for Molecular Medicine of the Austrian Academy of Sciences | Lukas Endler, Anna Schedl, Thomas Penz, Benedikt Agerer, Maelle Le Moing, Michael Schuster, Bekir Erguner, Jan Laine, Martin Senekowitsch, Christoph Bock, Andreas Bergthaler |
| EPI_ISL_1008713 | Lister Laboratorio de Referencia S.A. de C.V. | Instituto de diagnóstico y Referencia Epidemiologicos (INDRE) Departamento de Virología | Claudia Wong-Arambula, Abril Rodriguez-Maldonado, Fabiola Garces-Ayala, Natividad Cruz-Ortiz, Tatiana Nunez-Garcia, Gisela Barrera-Badillo, Lucia Hernandez-Rivas, Irma Lopez-Martinez, Ernesto Ramirez-Gonzalez. |
| EPI_ISL_1010705 | Area of Virology, Serology and Virology Division (SAViD), New South Wales Health Pathology Randwick | Microbiology RPAH | Foster, C.; Au, J.; Ruiz Silva, M.; Deveson, I.; Bull, R.; Van Hal, S.; Rawlinson, W. |
| EPI_ISL_1010713, EPI_ISL_1010714 | New South Wales Health Pathology Royal Prince Alfred Hospital | Microbiology RPAH | Foster, C.; Au, J.; Ruiz Silva, M.; Deveson, I.; Bull, R.; Van Hal, S.; Rawlinson, W. |
| EPI_ISL_1010728 | Department of Laboratory Medicine, National Taiwan University Hospital | Microbial Genomics Core Lab, National Taiwan University Centers of Genomic and Precision Medicine | Shiou-Hwei Yeh, You-Yu Lin, Ya-Yun Lai, Chiao-Ling Li, Shan-Chwen Chang, Pei-Jer Chen, Sui-Yuan Chang |
| EPI_ISL_1011756 | General Hospital - Veles | Research Center for Genetic Engineering and Biotechnology "Georgi D. Efremov" , Macedonian Academy of Sciences and Arts | Aleksandar J. Dimovski, Dijana Plasheska-Karanfilska, Predrag Noveski, Gjorgji Bozinovski, Milena Jakimovska |
| EPI_ISL_1013216, EPI_ISL_1013432 | Institut Pasteur de Guadeloupe | National Reference Center for Viruses of Respiratory Infections, Institut Pasteur, Paris | Marion Barbet, Sylvie Behillil, Méline Bizard, Angela Brisebarre, Camille Capel, Etienne Simon-Lorière, Vincent Enouf, Maud Vanpeene, Sylvie van der Werf,Talarmin Antoine |
| EPI_ISL_1013603, EPI_ISL_1013605 | Department for Molecular Diagnostics, Centre for Medical Microbiology, Institute of Public Health, Montenegro | Charité Universitätsmedizin Berlin, Institut für Virologie | Victor M Corman, Barbara Mühlemann, Jörn Beheim-Schwarzbach, Julia Tesch, Tobias Bleicker, Danijela Vujoševi, Marija Govedarica, Talitha Veith, Julia Schneider, Terry Jones, Christian Drosten |
| EPI_ISL_1013864, EPI_ISL_1014351, EPI_ISL_1014352, EPI_ISL_1014549, EPI_ISL_1014555, EPI_ISL_1014558, EPI_ISL_1014559, EPI_ISL_1014565, EPI_ISL_1014568, EPI_ISL_1014574, EPI_ISL_1014576, EPI_ISL_1014580, EPI_ISL_1014587, EPI_ISL_1014590, EPI_ISL_1014592, EPI_ISL_1014594, EPI_ISL_1014595, EPI_ISL_1014603, EPI_ISL_1014631, EPI_ISL_1014641, EPI_ISL_1014649, EPI_ISL_1014652, EPI_ISL_1014654 | | | |
| see above | Dutch COVID-19 response team | National Institute for Public Health and the Environment (RIVM) | Adam Meijer, Harry Vennema, Dirk Eggink, Jeroen Cremer, Sharon van den Brink, Bas van der Veer, AnneMarie van den Brandt, Florian Zwagemaker, Dennis Schmitz, Chantal Reusken, on behalf of the national COVID-19 response team |
| EPI_ISL_1014676 | National Influenza Center, Virology Department | National Influenza Center | V Salimi,NZ Shafiei Jandaghi, J Yavarian, A Nejati, K Sadeghi, N Ghavvami,F Ajaminejad and T Mokhtari Azad |
| EPI_ISL_1014684 | National Influenza Center, Virology Department | National Influenza Center | A Nejati, J Yavarian,K Sadeghi, NZ Shafiei Jandaghi, V Salimi, N Ghavvami,F Ajaminejad and T Mokhtari Azad |
| EPI_ISL_1014685 | National Influenza Center, Virology Department | National Influenza Center | K Sadeghi, A Nejati, J Yavarian, NZ Shafiei Jandaghi, V Salimi, F Ajaminejad,N Ghavvami and T Mokhtari Azad |
| EPI_ISL_1014816 | University of Michigan Clinical Microbiology Laboratory | Lauring Lab, University of Michigan, Department of Microbiology and Immunology | Valesano |
| EPI_ISL_1016681 | Helix/Illumina | Respiratory Viruses Branch, Division of Viral Diseases, Centers for Disease Control and Prevention | Peter W. Cook, Dakota Howard, Dhwani Batra, Ben L. Rambo-Martin, Eileen de Feo, Jan Antico, Christine Tran, Matthew Tolentino, Shannon Wickline, Kim Gietzen, Brad Sickler, Jingtao Liu, Eric Allen, Phil Febbo, Summer Galloway, Nicole L. Washington, Simon White, Geraint Levan, Kelly Schiabor Barrett, Elizabeth Cirulli, Alexandre Bolze, Ary Ascencio, Charlotte Rivera-Garcia, Ryan Cho, Jason Nguyen, Sherry Wang, Jimmy Ramirez, Tyler Cassens, Efren Sandoval, Magnus Isaksson, William Lee, David Becker, Marc Laurent, James Lu, Clinton R. Paden, Suxiang Tong, Duncan MacCannell |
| EPI_ISL_1016842 | University of Sarajevo, Veterinary Faculty, Laboratory for Molecular Diagnostic and Research Laboratory | University of Sarajevo, Veterinary Faculty, Laboratory for Molecular Diagnostic and Research Laboratory | Goletic T., Goletic S., Softic A., Alic-Seho A., Nicevic M., Jazic A., Hodzic A., Terzic I., Sabic E. |

| | | | |
|---|---|---|---|
| EPI_ISL_1016846, EPI_ISL_1016851, EPI_ISL_1016853, EPI_ISL_1016854, EPI_ISL_1016867, EPI_ISL_1016873 | LabPLUS | Institute of Environmental Science and Research (ESR) | Xiaoyun Ren, Matt Storey, Nikki Freed, Muhammad Faisal, Jing Wang, Hermes Perez, Anja Werno, Antje van der Linden, Arlo Upton, Chris Mansell, David Hammer, Dragana Drinkovic, Gary McAuliffe, Hana Sofia Andersson, James Ussher, Jill Sherwood, Josh Freeman, Julia Howard, Juliet Elvy, Mary DeAlmeida, Matt Blakiston, Matthew Rogers, Max Bloomfield, Michael Addidle, Michelle Balm, Sally Roberts, Sarah Jefferies, Sharmini Muttaiyah, Susan Morpeth, Susan Taylor, Timothy Blackmore, Vani Sathyendran, Veronica Playle, Virginia Hope, Erasmus Smit, Lauren Jelly, Olin Silander, Joep de Ligt |
| EPI_ISL_1016878 | Wellington SCL (WN) | Institute of Environmental Science and Research (ESR) | Xiaoyun Ren, Matt Storey, Nikki Freed, Muhammad Faisal, Jing Wang, Hermes Perez, Anja Werno, Antje van der Linden, Arlo Upton, Chris Mansell, David Hammer, Dragana Drinkovic, Gary McAuliffe, Hana Sofia Andersson, James Ussher, Jill Sherwood, Josh Freeman, Julia Howard, Juliet Elvy, Mary DeAlmeida, Matt Blakiston, Matthew Rogers, Max Bloomfield, Michael Addidle, Michelle Balm, Sally Roberts, Sarah Jefferies, Sharmini Muttaiyah, Susan Morpeth, Susan Taylor, Timothy Blackmore, Vani Sathyendran, Veronica Playle, Virginia Hope, Erasmus Smit, Lauren Jelly, Olin Silander, Joep de Ligt |
| EPI_ISL_1016879 | Middlemore Hospital | Institute of Environmental Science and Research (ESR) | Xiaoyun Ren, Matt Storey, Nikki Freed, Muhammad Faisal, Jing Wang, Hermes Perez, Anja Werno, Antje van der Linden, Arlo Upton, Chris Mansell, David Hammer, Dragana Drinkovic, Gary McAuliffe, Hana Sofia Andersson, James Ussher, Jill Sherwood, Josh Freeman, Julia Howard, Juliet Elvy, Mary DeAlmeida, Matt Blakiston, Matthew Rogers, Max Bloomfield, Michael Addidle, Michelle Balm, Sally Roberts, Sarah Jefferies, Sharmini Muttaiyah, Susan Morpeth, Susan Taylor, Timothy Blackmore, Vani Sathyendran, Veronica Playle, Virginia Hope, Erasmus Smit, Lauren Jelly, Olin Silander, Joep de Ligt |
| EPI_ISL_1016926 | Arizona State Public Health Laboratory | Arizona State Public Health Laboratory | Trung Huynh, Jessica Escobar, Katherine Fullerton, Nobuko Fukushima, Stacy White, Linda Getsinger, Victor Waddell |
| EPI_ISL_1017084 | Providence St. Joseph Health Orange - Chapman | Providence St. Joseph Health Molecular Genomics Laboratory | Alexa K Dowdell, Brian D Piening, Fred L Robinson, Carlo B Bifulco, Mary Campbell |
| EPI_ISL_1017522 | Murphy Medical Associates | Grubaugh Lab - Yale School of Public Health | Mary Petrone, Joseph Fauver, Caleb Neal, Steven Murphy, Chantal Vogels, Mallery Breban, Annie Watkins, Tara Alpert, Nathan Grubaugh |
| EPI_ISL_1017678 | PathWest Laboratory Medicine WA | PathWest Laboratory Medicine WA Microbial Surveillance Unit | PathWest Laboratory Medicine WA Microbial Surveillance Unit |
| EPI_ISL_1017686 | Institute of Virology, Vaccines and Sera "Torlak" | Institute of microbiology and Immunology, Faculty of Medicine, University of Belgrade | Knezevic,A., Jankovic,M., Vidanovic,D., Milicevic,O., Tesovic,B., Sekler,M., Jovanovic,T. |
| EPI_ISL_1018093, EPI_ISL_1018094, EPI_ISL_1018095, EPI_ISL_1018096, EPI_ISL_1018097, EPI_ISL_1018098 | Immunology, Noguchi Memorial Institute for Medical Research | Immunology, Noguchi Memorial Institute for Medical Research | Adu,B., Egyir,B., Kumordjie,S., Agbodzi,B., Yeboah,C., Mohktar,Q., Oteng,F., Owusu-Nyantakyi,C., Asare,K.M., Appiah-Kubi,J., Adusei-Poku,M.A., Odoom,J.K., Ampofo,W.K., Bonney,J.K. |
| EPI_ISL_1018273, EPI_ISL_1018291, EPI_ISL_1019800, EPI_ISL_1020239, EPI_ISL_1020241 | Department of Health Technology and Informatics, The Hong Kong Polytechnic University | Department of Health Technology and Informatics, The Hong Kong Polytechnic University | Gilman Kit-Hang Siu, Lam-Kwong Lee, Kenneth Siu-Sing Leung, Jake Siu-Lun Leung, Timothy Ting-Leung Ng, Chloe Toi-Mei Chan, Kingsley King-Gee Tam, Hiu-Yin Lao, Denise Sze-Hang Wong, Alan Ka-Lun Wu, Miranda Chong-Yee Yau, Yvette Wai-Man Lai, Kitty Sau-Chun Fung, Sandy Ka-Yee Chau, Barry Kin-Chung Wong, Wing-Kin To, Kristine Luk, Alex Yat-Man Ho, Tak-Lun Que, Kam-Tong Yip, Wing Cheong Yam, David Ho-Keung Shum, Shea Ping Yip |
| EPI_ISL_1020315, EPI_ISL_1020316 | Department of Laboratory Medicine, National Taiwan University Hospital | Microbial Genomics Core Lab, National Taiwan University Centers of Genomic and Precision Medicine | Shiou-Hwei Yeh, You-Yu Lin, Ya-Yun Lai, Chiao-Ling Li, Shan-Chwen Chang, Pei-Jer Chen, Sui-Yuan Chang |
| EPI_ISL_1021320, EPI_ISL_1021798, EPI_ISL_1022178 | Laboratory Corporation of America | Respiratory Viruses Branch, Division of Viral Diseases, Centers for Disease Control and Prevention | Peter W. Cook, Dakota Howard, Dhwani Batra, Ben L. Rambo-Martin, Clinton R. Paden, Suxiang Tong, Duncan MacCannell |
| EPI_ISL_1022519 | Department of Virus and Microbiological Special Diagnostics, Statens Serum Institut, Copenhagen, Denmark | Aalborg University | Danish Covid-19 Genome Consortium |
| EPI_ISL_1023463 | Instituto Nacional de Saude (INSA) | Instituto Nacional de Saude (INSA) | Borges et al |
| EPI_ISL_1024488, EPI_ISL_1025624 | Department of Virus and Microbiological Special Diagnostics, Statens Serum Institut, Copenhagen, Denmark | Aalborg University | Danish Covid-19 Genome Consortium |
| EPI_ISL_1027650, EPI_ISL_1027657 | Department of Microbiology, National Institute for Public Health of Kosova | Charité Universitätsmedizin Berlin, Institut für Virologie | Victor M Corman, Julia Schneider, Donjeta Hajdari, Zana Deva, Xhevat Jakupi, Barbara Mühlemann, Jörn Beheim-Schwarzbach, Talitha Veith, Terry Jones, Christian Drosten |
| EPI_ISL_1027694, EPI_ISL_1029064, EPI_ISL_1029254, EPI_ISL_1029398, EPI_ISL_1029543, EPI_ISL_1029549, EPI_ISL_1029884, EPI_ISL_1029912, EPI_ISL_1029927 | Laboratory Corporation of America | Respiratory Viruses Branch, Division of Viral Diseases, Centers for Disease Control and Prevention | Peter W. Cook, Dakota Howard, Dhwani Batra, Ben L. Rambo-Martin, Clinton R. Paden, Suxiang Tong, Duncan MacCannell |
| EPI_ISL_1029956 | SA Pathology | SA Pathology | Lex Leong, Julien Soubrier, Chuan Kok Lim, Song Gao, Mark Turra, Karin Kassahn, Ivan Bastian, Geoff Higgins |
| EPI_ISL_1030513, EPI_ISL_1032090 | Laboratory Corporation of America | Respiratory Viruses Branch, Division of Viral Diseases, Centers for Disease Control and Prevention | Peter W. Cook, Dakota Howard, Dhwani Batra, Ben L. Rambo-Martin, Clinton R. Paden, Suxiang Tong, Duncan MacCannell |
| EPI_ISL_1032883, EPI_ISL_1032912 | Illinois Department of Public Health | Gagnon Lab, Southern Illinois University | Keith Gagnon |
| EPI_ISL_1033145, EPI_ISL_1033147, EPI_ISL_1033150 | Microbiological Diagnostic Unit - Public Health Laboratory (MDU-PHL) | MDU-PHL | Seemann T., Sait, M.L., Sherry, N.L. |
| EPI_ISL_1033154 | Victorian Infectious Diseases Reference Laboratory (VIDRL) | VIDRL and MDU-PHL | Caly L., Seemann T., Sait, M.L., Druce J., Sherry, N.L. |
| EPI_ISL_1033156 | Microbiological Diagnostic Unit - Public Health Laboratory (MDU-PHL) | MDU-PHL | Seemann T., Sait, M.L., Sherry, N.L. |
| EPI_ISL_1034592, EPI_ISL_1034659, EPI_ISL_1034698, EPI_ISL_1034720 | Department of Microbiology, The University of Hong Kong | Department of Microbiology, The University of Hong Kong | Kelvin K.W. To, Kwok-Yung Yuen |
| EPI_ISL_1034756, EPI_ISL_1034757, EPI_ISL_1034758 | Bundeswehr Institute of Microbiology | Bundeswehr Institute of Microbiology | Markus Antwerpen, Alexandra Rehn, Mathias Walter, Malena Bestehorn-Willmann, Mike Pillukat, Sabine Zange, Enrico Georgi, Roman Wölfel |
| EPI_ISL_1035764, EPI_ISL_1035765, EPI_ISL_1035767, EPI_ISL_1035768, EPI_ISL_1035769, EPI_ISL_1035808 | Dutch COVID-19 response team | National Institute for Public Health and the Environment (RIVM) | Adam Meijer, Harry Vennema, Dirk Eggink, Jeroen Cremer, Sharon van den Brink, Bas van der Veer, AnneMarie van den Brandt, Florian Zwagemaker, Dennis Schmitz, Chantal Reusken, on behalf of the national COVID-19 response team |
| EPI_ISL_1036102 | Labormedizinisches Zentrum Dr Risch | University Hospital Basel, Clinical Bacteriology | Tim Roloff, Madlen Stange, Helena MB Seth-Smith, Alfredo Mari, Karoline Leuzinger, Julia Bielicki, Nadia Wohlwend,Martin Risch, Lorenz Risch, Manuel Battegay, Hans Hirsch, Adrian Egli |
| EPI_ISL_1036185 | SIESP DIP PREV CHIETI | Istituto Zooprofilattico Sperimentale dell'Abruzzo e Molise "G. Caporale" | Lorusso A, Marcacci M, Di Domenico M, Ancora M, Curini V, Mangone I, Rinaldi A, Scialabba S, Di Pasquale A, Cammà C, Puglia I, Calistri P, Savini G |
| EPI_ISL_1036751 | National Institute of Laboratory Medicine and Referral Center | Genomic Research Lab, BCSIR | Shahina Akter, Mohammad Samir Uzzaman, Eshrar Osman, Md. Ahashan Habib, Tanjina Akhtar Banu, Abu Sayeed Mohammad Mahmud, Md. Murshed Hasan Sarkar, Barna Goswami, Iffat Jahan, Md. Saddam Hossain, Tasnim Nafisa, Md. Maruf Ahmed Molla, Mahmuda Yeasmin, Asish Kumar Ghosh, Arifa Akram, A. K. M.Shamsuzzaman, Md. Salim Khan |
| EPI_ISL_1036752 | National Institute of Laboratory Medicine and Referral Center | Genomic Research Lab, BCSIR, Dr. Qudrat-E-Khuda Road, Dhaka 1205, Bangladesh | Tanjina Akhtar Banu, Mohammad Samir Uzzaman, Eshrar Osman, Md. Ahashan Habib, Shahina Akter, Abu Sayeed Mohammad Mahmud, Md. Murshed Hasan Sarkar, Barna Goswami, Iffat Jahan, Md. Saddam Hossain, Tasnim Nafisa, Md. Maruf Ahmed Molla, Mahmuda Yeasmin, Asish Kumar Ghosh, Arifa Akram, A. K. M.Shamsuzzaman, Md. Salim Khan, |
| EPI_ISL_1038710 | Laboratory Corporation of America | Respiratory Viruses Branch, Division of Viral Diseases, Centers for Disease Control and Prevention | Peter W. Cook, Dakota Howard, Dhwani Batra, Ben L. Rambo-Martin, Clinton R. Paden, Suxiang Tong, Duncan MacCannell |
| EPI_ISL_1039130 | HELIX LLC | WHO National Influenza Centre Russian Federation | Andrey Komissarov, Artem Fadeev, Anna Ivanova, Kseniya Komissarova, Dmitry Bazhenov, Tamila Musaeva, Maria Timofeeva, Veronika Eder, Maria Pisareva, Daria Danilenko, Ksenia Safina, Elena Nabieva, Georgii Bazykin, Dmitry Lioznov |
| EPI_ISL_1039156 | WHO National Influenza Centre Russian Federation | WHO National Influenza Centre Russian Federation | Andrey Komissarov, Artem Fadeev, Anna Ivanova, Kseniya Komissarova, Dmitry Bazhenov, Tamila Musaeva, Maria Timofeeva, Veronika Eder, Maria |

| | | | |
|---|---|---|---|
| | | | Pisareva, Daria Danilenko, Ksenia Safina, Elena Nabieva, Georgii Bazykin, Dmitry Lioznov |
| EPI_ISL_1040028 | Original detection - Virology Unit, Institut Pasteur du Cambodge; Sequencing - US National Institute of Allergy and Infectious Diseases Cambodia | Virology Unit, Institut Pasteur du Cambodge | Jennifer Bohl, Sophana Chea, Sreyngim Lay, Ly Sovann, Kraing Sidonn, Yi Sengdoeurn, Chin Savuth, Chau Darapheak, Veasna Duong, Jessica Manning, Erik A Karlsson |
| EPI_ISL_1040066 | UCLA Clinical Micro Lab | Los Angeles County PHL | P. Hemarajata et al. |
| EPI_ISL_1040659, EPI_ISL_1040818 | Groote Schuur Hospital wc GSH | NHLS/UCT | Arash Iranzadeh, Deelan Doolabh, Lynn Tyers, Bruna Galvao, Innocent Mudau, Marvin Hsiao, Kruger Marais, Diana Hardie, Stephen Korsman, Carolyn Williamson |
| EPI_ISL_1040820 | CoVid WC Cape Town Metro | NHLS/UCT | Arash Iranzadeh, Deelan Doolabh, Lynn Tyers, Bruna Galvao, Innocent Mudau, Marvin Hsiao, Kruger Marais, Diana Hardie, Stephen Korsman, Carolyn Williamson |
| EPI_ISL_1041197, EPI_ISL_1041200, EPI_ISL_1041203 | Hungarian Defence Forces Military Medical Centre | National Laboratory of Virology, Szentágothai Research Centre | Endre Gábor Tóth, Balázs Somogyi, Ágnes Balázs-Nagy, Csaba Pereszlényi,Ferenc Jakab, Gábor Kemenesi |
| EPI_ISL_1046791 | Bundeswehr Institute of Microbiology | Bundeswehr Institute of Microbiology | Markus Antwerpen, Alexandra Rehn, Mathias Walter, Malena Bestehorn-Willmann, Mike Pillukat, Sabine Zange, Enrico Georgi, Roman Wölfel |
| EPI_ISL_1048814, EPI_ISL_1049198 | KU Leuven, Rega Institute, Clinical and Epidemiological Virology | KU Leuven, Rega Institute, Clinical and Epidemiological Virology | Tony Wawina-Bokalanga, Bert Vanmechelen, Joan Marti-Carerras, Piet Maes |
| EPI_ISL_1054965, EPI_ISL_1054981, EPI_ISL_1054982, EPI_ISL_1054994, EPI_ISL_1054995 | Instituto de Diagnostico y Referencia Epidemiologicos INDRE_RNLSP | Instituto de Diagnostico y Referencia Epidemiologicos (INDRE) | Claudia Wong-Arambula, Abril Rodriguez-Maldonado, Fabiola Garces-Ayala, Adnan Araiza-Rodriguez, David Fragoso-Fonseca, Sergio Rangel-Guerrero, Mayra Jimenez-Morales, Nancy Munoz-Hernandez, Natividad Cruz-Ortiz, Tatiana Nunez-Garcia, Gisela Barrera-Badillo, Lucia Hernandez-Rivas, Irma Lopez-Martinez, Ernesto Ramirez-Gonzalez. |
| EPI_ISL_1055447, EPI_ISL_1055448, EPI_ISL_1055451 | Dr. Leonard A. Miller Centre for Health Services | National Microbiology Laboratory (NML) | Anna Majer, Shari Tyson, Grace Seo, Philip Mabon, Elsie Grudeski, Rhiannon Huzarewich, Russell Mandes, Anneliese Landgraff, Jennifer Tanner, Natalie Knox, Morag Graham, Gary Van Domselaar, Robert Needle, Yang Yu, Adel Malek, Laura Gilbert, George Zahariadis, Nathalie Bastien, Yan Li, Timothy Booth, Darian Hole, Madison Chapel, Kirsten Biggar, Kerri Smith, CanCOGeN's metadata curation team, Public Health Agency of Canada CanCOGeN team |
| EPI_ISL_1055464, EPI_ISL_1055683 | QEII Health Sciences Centre | National Microbiology Laboratory (NML) | Anna Majer, Shari Tyson, Grace Seo, Philip Mabon, Elsie Grudeski, Rhiannon Huzarewich, Russell Mandes, Anneliese Landgraff, Jennifer Tanner, Natalie Knox, Morag Graham, Gary Van Domselaar, Todd Hatchette, Jason LeBlanc, Janice Pettipas, Dan Gaston, Nathalie Bastien, Yan Li, Timothy Booth, Darian Hole, Madison Chapel, Kirsten Biggar, CanCOGeN's metadata curation team, Public Health Agency of Canada CanCOGeN team |
| EPI_ISL_1056301 | National Laboratory for Health, Environment and Food | CISLD (Clinical Institute of Special Laboratory Diagnostics), University Children's Hospital, University Medical Center Ljubljana | Jernej Kova, Barbara Jenko Bizjan, Tine Tesovnik, Robert Šket, Katarina Kozmos, Ana Grom, Maruša Debeljak, Marko Pokorn, Tadej Battelino |
| EPI_ISL_1056974, EPI_ISL_1057098 | Institute of Biocides and Medical Ecology, Belgarde, Serbia | Virology Department Institute of Microbiology and Immunology Faculty of Medicine University of Belgrade | Banko Ana, Miljanovic Danijela, Milicevic Ognjen, Loncar Ana, Abazovic Dzihan, Despot Dragana |
| EPI_ISL_1059913, EPI_ISL_1059914 | Dutch COVID-19 response team | National Institute for Public Health and the Environment (RIVM) | Adam Meijer, Harry Vennema, Dirk Eggink, Jeroen Cremer, Sharon van den Brink, Bas van der Veer, AnneMarie van den Brandt, Florian Zwagemaker, Dennis Schmitz, Chantal Reusken, on behalf of the national COVID-19 response team |
| EPI_ISL_1060728, EPI_ISL_1060752, EPI_ISL_1060754 | Instituto de Diagnostico y Referencia Epidemiologicos (INDRE)_RNLSP | Instituto de Diagnostico y Referencia Epidemiologicos (INDRE) | Claudia Wong-Arambula, Abril Rodriguez-Maldonado, Fabiola Garces-Ayala, Adnan Araiza-Rodriguez, David Fragoso-Fonseca, Sergio Rangel-Guerrero, Mayra Jimenez-Morales, Nancy Munoz-Hernandez, Natividad Cruz-Ortiz, Tatiana Nunez-Garcia, Gisela Barrera-Badillo, Lucia Hernandez-Rivas, Irma Lopez-Martinez, Ernesto Ramirez-Gonzalez. |
| EPI_ISL_1061309 | Public Health Virology-Forensic and Scientific Services | Public Health Virology-Forensic and Scientific Services | Son Nguyen |
| EPI_ISL_1063607, EPI_ISL_1063726, EPI_ISL_1063729 | Division of Emerging Infectious Diseases, Bureau of Infectious Diseases Diagnosis Control, Korea Disease Control and Prevention Agency | Division of Emerging Infectious Diseases, Bureau of Infectious Diseases Diagnosis Control, Korea Disease Control and Prevention Agency | Ae Kyung Park, Il-Hwan Kim, Heui Man Kim, Jeong-Min Kim, Namjoo Lee, Chaeyoung Lee, Sang Hee Woo, Eun-Jin Kim |
| EPI_ISL_1065954 | Department of Virus and Microbiological Special Diagnostics, Statens Serum Institut, Copenhagen, Denmark | Aalborg University | Danish Covid-19 Genome Consortium |
| EPI_ISL_1067591 | AREA DE SALUD GOICOECHEA 2 - CLINICA DR. JIMENEZ NUÑEZ | Inciensa, Instituto Costarricense de Investigación y Enseñanza en Nutrición y Salud | Francisco Duarte, Hebleen Porras, Claudio Soto-Garita, Estela Cordero, Adriana Godínez, Melany Calderón & Mariel López |
| EPI_ISL_1067595 | TAMIZAJE COMUNITARIO - PASO CANOAS | Inciensa, Instituto Costarricense de Investigación y Enseñanza en Nutrición y Salud | Francisco Duarte, Hebleen Porras, Claudio Soto-Garita, Estela Cordero, Adriana Godínez, Melany Calderón & Mariel López |
| EPI_ISL_1067599 | AREA DE SALUD TURRIALBA-JIMENEZ | Inciensa, Instituto Costarricense de Investigación y Enseñanza en Nutrición y Salud | Francisco Duarte, Hebleen Porras, Claudio Soto-Garita, Estela Cordero, Adriana Godínez, Melany Calderón & Mónica Charpentier-Artavia |
| EPI_ISL_1067604 | HOSPITAL SAN JUAN DE DIOS | Inciensa, Instituto Costarricense de Investigación y Enseñanza en Nutrición y Salud | Francisco Duarte, Hebleen Porras, Claudio Soto-Garita, Estela Cordero, Adriana Godínez, Melany Calderón & Daniel Cascante-Serrano |
| EPI_ISL_1067608 | unknown | Inciensa, Instituto Costarricense de Investigación y Enseñanza en Nutrición y Salud | Francisco Duarte, Hebleen Porras, Claudio Soto-Garita, Estela Cordero, Adriana Godínez, Melany Calderón, Caterina Guzmán, Nazareth Ruiz & Adriana Bermúdez-Espinoza |
| EPI_ISL_1067614 | HOSPITAL MEXICO | Inciensa, Instituto Costarricense de Investigación y Enseñanza en Nutrición y Salud | Francisco Duarte, Hebleen Porras, Claudio Soto-Garita, Estela Cordero, Adriana Godínez, Melany Calderón, Caterina Guzmán, Nazareth Ruiz & Teresita Somogyi |
| EPI_ISL_1067617 | AREA DE SALUD GOICOECHEA 2 - CLINICA DR. JIMENEZ NUÑEZ | Inciensa, Instituto Costarricense de Investigación y Enseñanza en Nutrición y Salud | Francisco Duarte, Hebleen Porras, Claudio Soto-Garita, Estela Cordero, Adriana Godínez, Melany Calderón & Mariel López |
| EPI_ISL_1067767 | Broad Institute Clinical Research Sequencing Platform | Infectious Disease Program, Broad Institute of Harvard and MIT | Lemieux,J.E., Siddle,K.J., Adams,G., Gladden-Young,A., Lagerborg,K., Rudy,M., DeRuff,K., Carter,A., Normandin,E., Bauer,M., Reilly,S., Tomkins-Tinch,C., Loreth,C., Chaluvadi,S., Birren,B.W., Gallagher,G., Smole,S., Park,D.J., MacInnis,B.L., and Sabeti,P.C. |
| EPI_ISL_1068157, EPI_ISL_1068160, EPI_ISL_1068170, EPI_ISL_1068198 | Laboratorio de Ecologia de Doencas Transmissiveis na Amazonia, Instituto Leonidas e Maria Deane - Fiocruz Amazonia | Laboratorio de Ecologia de Doencas Transmissiveis na Amazonia, Instituto Leonidas e Maria Deane - Fiocruz Amazonia | Valdinete Nascimento, Victor Souza, André Corado, Fernanda Nascimento, George Silva, Ágatha Costa, Debora Duarte, Karina Pessoa, Matilde Mejía, Luciana Gonçalves, Maria Júlia Brandão, Michele Jesus, Felipe Naveca on behalf of the Fiocruz COVID-19 Genomic Surveillance Network |
| EPI_ISL_1069181 | Pagbilao Rural Health Unit | Research Institute for Tropical Medicine | Hannah Leah Morito, Othoniel Jan Onza, John Leonard Chan, Ma Angelica Tujan, Francisco Gerardo Polotan, Inez Andrea Medado, Kirstyn Brunker, Edelwisa Mercado, Daria Manalo, Catalino Demetria, Joseph Hughes |
| EPI_ISL_1069185 | H.M. Corachea General Hospital | Research Institute for Tropical Medicine | Hannah Leah Morito, Othoniel Jan Onza, John Leonard Chan, Ma Angelica Tujan, Francisco Gerardo Polotan, Inez Andrea Medado, Kirstyn Brunker, Edelwisa Mercado, Daria Manalo, Catalino Demetria, Joseph Hughes |
| EPI_ISL_1069391, EPI_ISL_1069392, EPI_ISL_1069393 | PathWest Laboratory Medicine WA | PathWest Laboratory Medicine WA Microbial Surveillance Unit | PathWest Laboratory Medicine WA Microbial Surveillance Unit |
| EPI_ISL_1072961 | National Institute of Laboratory Medicine and Referral Center | Genomic Research Lab, BCSIR | Md. Maruf Ahmed Molla, Mohammad Samir Uzzaman, Eshrar Osman, Md. Ahashan Habib, Shahina Akter,Tanjina Akhtar Banu, Abu Sayeed Mohammad Mahmud, Md. Murshed Hasan Sarkar, Barna Goswami, Iffat Jahan, Md. Saddam Hossain, Tasnim Nafisa, Mahmuda Yeasmin, Asish Kumar Ghosh, Arifa Akram, A. K. M.Shamsuzzaman, Md. Salim Khan |
| EPI_ISL_1073648, EPI_ISL_1073747 | Ministry of Health Turkey | Ministry of Health Turkey | Fatma Bayrakdar, Yasemin Cogun, Süleyman Yalcin, Aye Baak Alta, Gülay Korukluolu |
| EPI_ISL_1073960, EPI_ISL_1073966 | Ramathibodi Hospital | COVID-19 Network Investigations (CONI) Alliance | Elizabeth Batty, Wasun Chantratita, Thanat Chookajorn, Stefan Fernandez, Angkana Huang, Anthony R. Jones, Khajohn Joonlasak, Chonticha Klungtong, Theerarat Kochakarn, Namfon Kotanan, Krittikorn Kumpornsin, Duangkamon Loesbanluechai, Wudtichai Manasatienkij, Bhakbhoom Panthan, Ekawat Pasomsub, Kingkan Rakmanee, Insee Sensorn, Janjira Thaipadungpanit, Arporn Wangwiwatsin, Treewat Watthanachockchai |

| | | | |
|---|---|---|---|
| EPI_ISL_1074004, EPI_ISL_1074006, EPI_ISL_1074008 | Office of Diseases Prevention and Control Region 4 Saraburi | COVID-19 Network Investigations (CONI) Alliance | Elizabeth Batty, Wasun Chantratita, Thanat Chookajorn, Stefan Fernandez, Angkana Huang, Anthony R. Jones, Khajohn Joonlasak, Chonticha Klungtong, Theerarat Kochakarn, Namfon Kotanan, Krittikorn Kumpornsin, Duangkamon Loesbanluechai, Wudtichai Manasatienkij, Bhakbhoom Panthan, Ekawat Pasomsub, Kingkan Rakmanee, Insee Sensorn, Janjira Thaipadungpanit, Arporn Wangwiwatsin, Treewat Watthanachockchai, Suttiruk Changchawai, Sataporn Hatsadichart, Jutikul Kaewmalakul, Siriporn Lakesukthom, Wonvimol Lemprasert, Praima Moonmuang, Payon Pengyo, Pakjira Rimdusit, Nathamon Runnachot, Sirinapa Singthong, Siriwan Yaemnimnual |
| EPI_ISL_1077961 | Houston Methodist Hospital | Houston Methodist Hospital | S. Wesley Long, Randall J. Olsen, Paul A. Christensen, Sishir Subedi, Robert Olson, James J. Davis, Matthew Ojeda Saavedra, Prasanti Yerramilli, Layne Pruitt, Kristina Reppond, Madison N. Shyer, Jessica Cambric, Ilya J. Finkelstein, Jimmy Gollihar, and James M. Musser |
| EPI_ISL_1078996 | IAL Regional de Bauru | Instituto Adolfo Lutz, Interdiciplinary Procedures Center, Strategic Laboratory | Claudio Tavares Sacchi, Claudia Regina Gonçalves, Erica Valessa Ramos Gomes, Karoline Rodrigues Campos |
| EPI_ISL_1080343 | NJ Rapid Response Teams | NJ_PHEL | Lindsey Bodnar, Shiv Verma, Dana Woell, Byeong Jeong |
| EPI_ISL_1080442 | Instituto Nacional de Medicina Genomica | Instituto Nacional de Medicina Genomica | Hidalgo-Miranda A, Mendoza-Vargas A, Reyes-Grajeda JP, Cedro-Tanda A, Alcaraz N, Gonzalez-Barrera D, Rangel-DeLeon D, Ramirez-Vega O, Sifuentes-Rojas C, Cisneros-Villanueva M, Herrera-Montalvo LA |
| EPI_ISL_1081452 | Genomica Lab Molecular, M©xico | Andersen lab at Scripps Research | SEARCH Alliance San Diego with Jonathan Gonzalez Garcia, Jose Roman Chavez Mendez, Jose Horacio Reyna Verdugo, Martin Gonzalez Ibarra, Luis Alberto Rangel Gonzalez |
| EPI_ISL_1081793, EPI_ISL_1081794 | Philippine Red Cross | Philippine Genome Center | Francis A. Tablizo, Cynthia P. Saloma, Marc Jerrone R. Castro, Kenneth M. Kim, Maria Sofia L. Yangzon, Carlo M. Lapid, Benedict A. Maralit, Marc Edsel C. Ayes, Jan Michael C. Yap, Jo-Hannah S. Llames, Sheila Mae M. Araiza, Kris P. Punayan, Irish Coleen A. Asin, Candice Francheska B. Tambaoan, Asia Louisa U. Chong, Karol Sophia Agape R. Padilla, Rianna Patricia S. Cruz, El King D. Morado, Joshua Gregor A. Dizon, Eva Maria Cutiongco-de la Paz, Alethea R. de Guzman, Razel Nikka M. Hao, Arianne A. Zamora, Devon Ray Pacial, Juan Antonio R. Magalang, Marissa Alejandria, Celia Carlos, Anna Ong-Lim, Edsel Maurice Salvaña, John Q. Wong, Jaime C. Montoya, and Maria Rosario Singh-Vergeire |
| EPI_ISL_1081941, EPI_ISL_1081947 | National Public Health Laboratory, National Centre for Infectious Diseases | National Public Health Laboratory, National Centre for Infectious Diseases | Tze Minn Mak, Zhenyang Zhou, Lin Cui, Raymond Tzer Pin Lin |
| EPI_ISL_1081961 | "Stefan S. Nicolau"Institute of Virology | "Stefan S. Nicolau"Institute of Virology | Anca Botezatu, Iulia Virginia Iancu, Coralia Bleotu, Mihaela Economescu, Laura Necula, Denisa Dragu, Lilia Matei, Ioana Pitica, Marinela Bostan, Mirela Mihaila, Laura Grecu, Camelia Sultana, Simona Ruta, Ana Iulia Neagu, Adriana Plesa, Saviana Nedeianu, Alina Nastasie, Cristina Mambet,Gabriela Anton, Carmen Cristina Diaconu |
| EPI_ISL_1082071, EPI_ISL_1082169 | Department of Virology and Immunology, University of Helsinki and Helsinki University Hospital, Huslab Finland | Department of Virology, Faculty of Medicine, University of Helsinki, Helsinki, Finland | Teemu Smura, Ravi Kant, Phuoc Truong, Hussein Alburkat, Hannimari Kallio-Kokko, Jenni Virtanen, Maija Suvanto, Essi Korhonen, Sari Hannula, Harri Kangas, Hanna Liimatainen, Satu Kurkela, Hanna Jarva, Maija Lappalainen, Pekka Ellonen, Olli Vapalahti |
| EPI_ISL_1082268 | LabPLUS | Institute of Environmental Science and Research (ESR) | Rachel Boyle, SallyAnn Harbison, Olivia Stroeven, Xiaoyun Ren, Matt Storey, Nikki Freed, Muhammad Faisal, Jing Wang, Hermes Perez, Anja Werno, Antje van der Linden, Arlo Upton, Chris Mansell, David Hammer, Dragana Drinkovic, Gary McAuliffe, Hana Sofia Andersson, James Ussher, Jill Sherwood, Josh Freeman, Julia Howard, Juliet Elvy, Mary DeAlmeida, Matt Blakiston, Matthew Rogers, Max Bloomfield, Michael Addidle, Michelle Balm, Sally Roberts, Sarah Jefferies, Sharmini Muttaiyah, Susan Morpeth, Susan Taylor, Timothy Blackmore, Vani Sathyendran, Veronica Playle, Virginia Hope, Erasmus Smit, Lauren Jelly, Olin Silander, Joep de Ligt |
| EPI_ISL_1082425 | Ministry of Health Turkey | Ministry of Health Turkey | Fatma Bayrakdar, Yasemin Cogun, Süleyman Yalcin, Aye Baak Alta, Gülay Korukluolu |
| EPI_ISL_1085553 | Hôpital Henri Mondor | Department of Virology, Henri Mondor University Hospital, Assistance Publique Hôpitaux de Paris, Université Paris-Est Créteil, INSERM U955 | Christophe Rodriguez, Slim Fourati, Vanessa Demontant, Guillaume Gricourt, Melissa N'Debi, Alexandre Soulier, Elisabeth Trawinski, Jean-Michel Pawlotsky |
| EPI_ISL_1085783 | CHI VILLENEUVE ST GEORGES | Department of Virology, Henri Mondor University Hospital, Assistance Publique Hôpitaux de Paris, Université Paris-Est Créteil, INSERM U955 | Christophe Rodriguez, Slim Fourati, Vanessa Demontant, Guillaume Gricourt, Melissa N'Debi, Alexandre Soulier, Elisabeth Trawinski, Jean-Michel Pawlotsky |
| EPI_ISL_1085935 | Besançon Laboratoire CBM25 Terre Rouge | Department of Virology, Henri Mondor University Hospital, Assistance Publique Hôpitaux de Paris, Université Paris-Est Créteil, INSERM U955 | Christophe Rodriguez, Slim Fourati, Vanessa Demontant, Guillaume Gricourt, Melissa N'Debi, Alexandre Soulier, Elisabeth Trawinski, Jean-Michel Pawlotsky |
| EPI_ISL_1085967 | Netcare/AMPATH | KRISP, KZN Research Innovation and Sequencing Platform | Giandhari J, Pillay S, Maslo C, Sitharam L, Lessells R, Mdlalose K, York D, Khan S, Emmanuel SJ, Tegally H, Wilkinson E, de Oliveira T |
| EPI_ISL_1086051 | IAL Regional de Bauru | Instituto Adolfo Lutz, Interdiciplinary Procedures Center, Strategic Laboratory | Claudio Tavares Sacchi, Claudia Regina Gonçalves, Erica Valessa Ramos Gomes, Karoline Rodrigues Campos, Caio Vinicius Dias Lopes |
| EPI_ISL_1086485, EPI_ISL_1086603, EPI_ISL_1087238, EPI_ISL_1087681, EPI_ISL_1088345 | Quest Diagnostics Incorporated | Respiratory Viruses Branch, Division of Viral Diseases, Centers for Disease Control and Prevention | Peter W. Cook, Dakota Howard, Dhwani Batra, Ben L. Rambo-Martin, S. H. Rosenthal, A. Gerasimova, R. M. Kagan, B. Anderson, M. Hua, Y. Liu, L.E. Bernstein, K.E. Livingston, A. Perez, I. A. Shlyakhter, R. V. Rolando, R. Owen, P. Tanpaiboon, F. Lacbawan, Clinton R. Paden, Suxiang Tong, Duncan MacCannell |
| EPI_ISL_1089223, EPI_ISL_1089294 | Helix / Illumina | Respiratory Viruses Branch, Division of Viral Diseases, Centers for Disease Control and Prevention | Peter W. Cook, Dakota Howard, Dhwani Batra, Ben L. Rambo-Martin, Eileen de Feo, Jan Antico, Christine Tran, Matthew Tolentino, Shannon Wickline, Kim Gietzen, Brad Sickler, Jingtao Liu, Eric Allen, Phil Febbo, Summer Galloway, Nicole L. Washington, Simon White, Geraint Levan, Kelly Schiabor Barrett, Elizabeth Cirulli, Alexandre Bolze, Ary Ascencio, Charlotte Rivera-Garcia, Ryan Cho, Jason Nguyen, Sherry Wang, Jimmy Ramirez, Tyler Cassens, Efren Sandoval, Magnus Isaksson, William Lee, David Becker, Marc Laurent, James Lu, Clinton R. Paden, Suxiang Tong, Duncan MacCannell |
| EPI_ISL_1089774, EPI_ISL_1089817 | Dutch COVID-19 response team | National Institute for Public Health and the Environment (RIVM) | Adam Meijer, Harry Vennema, Dirk Eggink, Jeroen Cremer, Sharon van den Brink, Bas van der Veer, AnneMarie van den Brandt, Florian Zwagemaker, Dennis Schmitz, Chantal Reusken, on behalf of the national COVID-19 response team |
| EPI_ISL_1090639 | Quest Diagnostics Incorporated | Respiratory Viruses Branch, Division of Viral Diseases, Centers for Disease Control and Prevention | Peter W. Cook, Dakota Howard, Dhwani Batra, Ben L. Rambo-Martin, S. H. Rosenthal, A. Gerasimova, R. M. Kagan, B. Anderson, M. Hua, Y. Liu, L.E. Bernstein, K.E. Livingston, A. Perez, I. A. Shlyakhter, R. V. Rolando, R. Owen, P. Tanpaiboon, F. Lacbawan, Clinton R. Paden, Suxiang Tong, Duncan MacCannell |
| EPI_ISL_1091259 | Laboratorio PGM | Laboratorio de Infectología Molecular, Departamento de Bioquímica y Medicina Molecular,Facultad de Medicina - Universidad Autónoma de Nuevo León | Kame A. Galán-Huerta, María F. Herrera-Saldivar, Natalia Martínez-Acuña, Sonia A. Lozano-Sepúlveda, Daniel Arellanos-Soto, Ana M. Rivas-Estilla, Javier Ramos-Jimenez, Gabriela Elizondo, Eduardo Garza-de-la-Peña |
| EPI_ISL_1091286 | Azienda Ospedaliera San Camillo Forlanini | INMI Lazzaro Spallanzani IRCCS | CEM Gruber, B Bartolini, E Giombini, F Messina, O Butera, M Rueca, G Parisi, ML Guarino, A Di Caro, MR Capobianchi |
| EPI_ISL_1091345 | KU Leuven, Rega Institute, Clinical and Epidemiological Virology | KU Leuven, Rega Institute, Clinical and Epidemiological Virology | Tony Wawina-Bokalanga, Bert Vanmechelen, Joan Marti-Carerras, Piet Maes |
| EPI_ISL_1091678 | US Air Force School of Aerospace Medicine | US Air Force School of Aerospace Medicine | Anthony Fries, Jennifer Meyer, William Gruner, William Buggele, Amanda Javorina, Sarah Purves, Clarise Starr, Elizabeth Macias |
| EPI_ISL_1091781 | ICMT | Universidad Nacional de Colombia - Laboratorio Genómico One Health | Andres F. Cardona-Rios, Daniel O. Maldonado-Perez, Laura Silvana Perez, Karl A Ciuoderis, Idabely Betancur Ortiz, Sandra Ines Cano, Diego A. Álvarez-Díaz, Carlos Franco-Muñoz, Marcela Mercado-Reyes, Jorge E. Osorio, Juan P. Hernandez-Ortiz |
| EPI_ISL_1091785 | LDSP | Universidad Nacional de Colombia - Laboratorio Genómico One Health | Andres F. Cardona-Rios, Daniel O. Maldonado-Perez, Laura Silvana Perez, Karl A Ciuoderis, Maria Angélica Maya, Idabely Betancur Ortiz, Sandra Ines Cano, Diego A. Álvarez-Díaz, Carlos Franco-Muñoz, Marcela Mercado-Reyes, Jorge E. Osorio, Juan P. Hernandez-Ortiz |
| EPI_ISL_1091824 | Murphy Medical Associates | Grubaugh Lab - Yale School of Public Health | Mary Petrone, Joseph Fauver, Caleb Neal, Steven Murphy, Chantal Vogels, Mallery Breban, Annie Watkins, Tara Alpert, Nathan Grubaugh |
| EPI_ISL_1092006 | Laboratorio IMAT | Instituto Nacional de Salud- Dirección de Investigación en Salud Pública | Katherine Laiton-Donato, Diego A. Álvarez-Díaz, Carlos Franco-Muñoz, Hector Alejandro Ruiz-Moreno, Maria T. Herrera-Sepúlveda, Diego Andrés Prada, Jhonnatan Reales-González, Sheryll Corchuelo, Julian Naizaque, Gerardo Santamaría, Magdalena Wiesner, Martha Lucia Ospina Martinez, Marcela Mercado-Reyes |
| EPI_ISL_1093361 | KU Leuven, Rega Institute, Clinical and Epidemiological Virology | KU Leuven, Rega Institute, Clinical and Epidemiological Virology | Tony Wawina-Bokalanga, Bert Vanmechelen, Joan Marti-Carerras, Piet Maes |

| | | | |
|---|---|---|---|
| EPI_ISL_1093447 | Nigerian Centre for Disease Control (NCDC) | African Centre of Excellence for Genomics of Infectious Diseases (ACEGID), Redeemer's University | Olawoye, I.B., et al |
| EPI_ISL_1094250 | IL Dept. of Public Health Springfield Laboratory | Respiratory Viruses Branch, Division of Viral Diseases, Centers for Disease Control and Prevention | Krista Queen, Yan Li, Ying Tao, Jing Zhang, Anna Uehara, Anna Montmayeur, Clinton R. Paden, Peter W. Cook, Rachel Marine, Mili Sheth, Jasmine Padilla, Sarah Nobles, Mark Burroughs, Lori Rowe, Haibin Wang, Ben L. Rambo-Martin, Dhwani Batra, Justin Lee, Suxiang Tong |
| EPI_ISL_1094677 | ME Health and Environmental Testing Laboratory | Respiratory Viruses Branch, Division of Viral Diseases, Centers for Disease Control and Prevention | Krista Queen, Yan Li, Ying Tao, Jing Zhang, Anna Uehara, Anna Montmayeur, Clinton R. Paden, Peter W. Cook, Rachel Marine, Mili Sheth, Jasmine Padilla, Sarah Nobles, Mark Burroughs, Lori Rowe, Haibin Wang, Ben L. Rambo-Martin, Dhwani Batra, Justin Lee, Suxiang Tong |
| EPI_ISL_1094719 | AK State Public Health Lab, State Health Department | Respiratory Viruses Branch, Division of Viral Diseases, Centers for Disease Control and Prevention | Krista Queen, Yan Li, Ying Tao, Jing Zhang, Anna Uehara, Anna Montmayeur, Clinton R. Paden, Peter W. Cook, Rachel Marine, Mili Sheth, Jasmine Padilla, Sarah Nobles, Mark Burroughs, Lori Rowe, Haibin Wang, Ben L. Rambo-Martin, Dhwani Batra, Justin Lee, Suxiang Tong |
| EPI_ISL_1094738 | NE Public Health Laboratory | Respiratory Viruses Branch, Division of Viral Diseases, Centers for Disease Control and Prevention | Krista Queen, Yan Li, Ying Tao, Jing Zhang, Anna Uehara, Anna Montmayeur, Clinton R. Paden, Peter W. Cook, Rachel Marine, Mili Sheth, Jasmine Padilla, Sarah Nobles, Mark Burroughs, Lori Rowe, Haibin Wang, Ben L. Rambo-Martin, Dhwani Batra, Justin Lee, Suxiang Tong |
| EPI_ISL_1094896 | AL Dept. of Public Health Bureau of Clinical Laboratories | Respiratory Viruses Branch, Division of Viral Diseases, Centers for Disease Control and Prevention | Krista Queen, Yan Li, Ying Tao, Jing Zhang, Anna Uehara, Anna Montmayeur, Clinton R. Paden, Peter W. Cook, Rachel Marine, Mili Sheth, Jasmine Padilla, Sarah Nobles, Mark Burroughs, Lori Rowe, Haibin Wang, Ben L. Rambo-Martin, Dhwani Batra, Justin Lee, Suxiang Tong |
| EPI_ISL_1094934 | MT Public Health Laboratory | Respiratory Viruses Branch, Division of Viral Diseases, Centers for Disease Control and Prevention | Krista Queen, Yan Li, Ying Tao, Jing Zhang, Anna Uehara, Anna Montmayeur, Clinton R. Paden, Peter W. Cook, Rachel Marine, Mili Sheth, Jasmine Padilla, Sarah Nobles, Mark Burroughs, Lori Rowe, Haibin Wang, Ben L. Rambo-Martin, Dhwani Batra, Justin Lee, Suxiang Tong |
| EPI_ISL_1096139, EPI_ISL_1096140, EPI_ISL_1096348 | Cambodian National Public Health Laboratory, National Institute of Public Health | Virology Unit, Institut Pasteur du Cambodge | Sokhoun Yann, Ly Sovann, Kraing Sidonn, Yi Sengdoeurn, Chin Savuth, Chau Darapheak, Veasna Duong, Erik A Karlsson |
| EPI_ISL_1096746 | ASL Napoli 1 Centro | AMES Centro Polidiagnostico Strumentale S.r.l. | "Giovanni Savarese, Raffaella Ruggiero, Eloisa Evangelista, Antonella Di Carlo, Luisa Circelli, Luigi D'Amore, Roberto Sirica, Antonio Fico" |
| EPI_ISL_1097629 | Colorado Department of Public Health and Environment | Colorado Department of Puplic Health and Environment | Laura Bankers, Molly C. Hetherington-Rauth, Diana Ir, Shannon Ely, Shannon R. Matzinger, Sarah Elizabeth Totten, Emily A. Travanty |
| EPI_ISL_1097649 | Eurofins Diatherix | Hudsonalpha Genome Sequencing Center | Jane Grimwood, Melissa Williams, Lori H. Handley, Joshua Stough, Leslie Malone, Stefan Brzezinski, Ada Stewart, Teresa Jones, Jenell Webber, John Lovell, Jennifer Cart, and Jeremy Schmutz |
| EPI_ISL_1098644, EPI_ISL_1098645 | Public Health Virology-Forensic and Scientific Services | Public Health Virology-Forensic and Scientific Services | Son Nguyen |
| EPI_ISL_1098798 | Area of Virology, Serology and Virology Division (SAViD), New South Wales Health Pathology Randwick | Virology Research Laboratory; Area of Virology, Serology and Virology Division (SAViD), New South Wales Health Pathology Randwick | Foster, C.; Au, J.; Ruiz Silva, M.; Deveson, I.; Bull, R.; Van Hal, S.; Rawlinson, W. |
| EPI_ISL_1103576 | Laboratoire central de Virologie | Laboratoire de Biotechnologie | Myriam Seffar, Hakima Kabbaj, Ghizlane EL Amin, Amal Zouaki, Mouna Ouadghiri, Tarik Aanniz, Abdelomunim Essabbar, Saaïd Amzazi, Lahcen Belyamani and Azeddine Ibrahimi |
| EPI_ISL_1107189 | Laboratoire central de Virologie | Laboratoire de Biotechnologie | Hakima Kabbaj, Myriam Seffar, Ghizlane EL Amin, Amal Zouaki, Abdelomunim Essabbar, Tarik Aanniz, Mouna Ouadghiri, Saaïd Amzazi, Lahcen Belyamani and Azeddine Ibrahimi |
| EPI_ISL_1109485 | Medical Ain Shams Research Institute (MASRI), Ain Shams University | Medical Ain Shams Research Institute (MASRI), Ain Shams University | Hesham Elghazaly, Sara Hassan Agwa, Hala Hafez , Fatma Ebied, Manal Hamdy Elsaid, Mohamed Elhadidi, Aya Mohamed , Shimaa Moustafa, Reham Kassab, Iman Foda , Samia Abdou Girgis, Hoda Ezz Elarab, Hagar Elshora, Osama Mansour. |
| EPI_ISL_1109548 | Croatian Institute of Public Health | Croatian Institute of Public Health | Irena Tabain, Ivana Ferenak |
| EPI_ISL_1109622 | Laboratoire central de Virologie | Laboratoire de Biotechnologie | Hakima Kabbaj, Myriam Seffar, Ghizlane EL Amin, Amal Zouaki, Abdelomunim Essabbar, Tarik Aanniz, Mouna Ouadghiri, Saaïd Amzazi, Lahcen Belyamani and Azeddine Ibrahim |
| EPI_ISL_1109674 | Servicio de Microbiología. Hospital Clínico Universitario de Valencia | SeqCOVID-SPAIN consortium/IBV(CSIC) | David Navarro Ortega, Eliseo Albert Vicent, Ignacio Torres and SeqCOVID-SPAIN consortium |
| EPI_ISL_1111077, EPI_ISL_1111144, EPI_ISL_1111146, EPI_ISL_1111148, EPI_ISL_1111154, EPI_ISL_1111159, EPI_ISL_1111160, EPI_ISL_1111284, EPI_ISL_1111286, EPI_ISL_1111307, EPI_ISL_1111313, EPI_ISL_1111314, EPI_ISL_1111335, EPI_ISL_1111349, EPI_ISL_1111350, EPI_ISL_1111446, EPI_ISL_1111467, EPI_ISL_1111475, EPI_ISL_1111476, EPI_ISL_1111483, EPI_ISL_1111485, EPI_ISL_1111489, EPI_ISL_1111490, EPI_ISL_1111491, EPI_ISL_1111503 | | | |
| see above | Laboratorio de Referencia Nacional de Virus Respiratorio. Instituto Nacional de Salud Perú | Laboratorio de Referencia Nacional de Enteropatógenos. Instituto Nacional de Salud del Perú | Ronnie Gavilan Chavez, Junior Caro Castro, Willi Quino Sifuentes, Veronica Hurtado Vela, Iris Silva Molina, Fiorella Orellana Peralta |
| EPI_ISL_1111986 | National Laboratory for Health, Environment and Food, OMM, Koper | CISLD (Clinical Institute of Special Laboratory Diagnostics), University Children's Hospital, University Medical Center Ljubljana | Jernej Kova, Barbara Jenko Bizjan, Tine Tesovnik, Robert Šket, Katarina Kozmos, Ana Grom, Maruša Debeljak, Marko Pokorn, Tadej Battelino |
| EPI_ISL_1112304, EPI_ISL_1112309, EPI_ISL_1112326, EPI_ISL_1112753 | Public Health Center of Ukraine | Charite Universitatsmedizin Berlin, Institute of Virology | Victor M Corman, Barbara Mühlemann, Jörn Beheim-Schwarzbach, Talitha Veith, Julia Schneider, Terry Jones, Roman Rodyna, Ihor Kuzin, Liudmyla Chernenko, Iryna Demchyshyna, Christian Drosten |
| EPI_ISL_1114719 | Institute for Medical Research, Infectious Disease Research Centre, National Institutes of Health, Ministry of Health Malaysia | Institute for Medical Research, Infectious Disease Research Centre, National Institutes of Health, Ministry of Health Malaysia | Suppiah J, Kamel K, Azizan MA, Thayan R |
| EPI_ISL_1116466 | Laboratoire central de Virologie | Laboratoire de Biotechnologie | Ghizlane EL Amin, Hakima Kabbaj, Myriam Seffar, Amal Zouaki, Abdelomunim Essabbar, Tarik Aanniz, Mouna Ouadghiri, Saaïd Amzazi, Lahcen Belyamani and Azeddine Ibrahimi |
| EPI_ISL_1116468 | Laboratoire de Microbiologie- CHU Habib Bourguiba - Sfax | Laboratoire des Procédés de Criblage Moléculaire et Cellulaire-Centre de Biotechnologie de Sfax | Souissi,A., Abid,N., Ben Ayed,I., Gargouri,S., Abdelmoulah,F.,Elargoubi,A., Smeti,I., Bensaid,M., Stambouli,N., Kharat,N., Ajili,F., Fki-berrajah,L., Mhalla,S., Chtourou,A., Gaaloul,I., Nabli,A., Turki,M., Aouni,M., Hammami,A., Mastouri,M., Karray Hakim,H., Kamoun,S., Rebai,A. and Masmoudi,S. |
| EPI_ISL_1116470 | Laboratoire central de Virologie | Laboratoire de Biotechnologie | Hakima Kabbaj, Myriam Seffar, Ghizlane EL Amin, Amal Zouaki, Abdelomunim Essabbar, Tarik Aanniz, Mouna Ouadghiri, Saaïd Amzazi, Lahcen Belyamani and Azeddine Ibrahimi |
| EPI_ISL_1117124 | Instituto Nacional de Saude (INSA) | Instituto Nacional de Saude (INSA) | Borges et al |
| EPI_ISL_1117312 | Instituto Nacional de Saude (INSA) and Instituto Gulbenkian de Ciencia (IGC) | Instituto Nacional de Saude (INSA) and Instituto Gulbenkian de Ciencia (IGC) | Borges et al |
| EPI_ISL_1117396 | Nucleo de Pesquisa em Inovacao Terapeutica - UFPE | LABBE, Federal University of Pernambuco | Wilson Jose da Silva Junior, Marcos da Silveira Regueira Neto, Heidi Lacerda Alves da Cruz, Bruno Sampaio, Reginaldo Goncalves de Lima Neto, Maira Galdino da Rocha Pitta, Michelly Cristiny Pereira, Marco Katzenberger, Valdir de Queiroz Balbino |
| EPI_ISL_1117828 | Austrian Agency for Health and Food Safety (AGES) | Bergthaler laboratory, CeMM Research Center for Molecular Medicine of the Austrian Academy of Sciences | Lukas Endler, Anna Schedl, Thomas Penz, Benedikt Agerer, Maelle Le Moing, Michael Schuster, Bekir Erguner, Jan Laine, Martin Senekowitsch, Christoph Bock, Andreas Bergthaler |
| EPI_ISL_1120732 | Laboratoire central de Virologie | Laboratoire de Biotechnologie | Hakima Kabbaj, Myriam Seffar, Ghizlane EL Amin, Amal Zouaki, Abdelomunim Essabbar, Tarik Aanniz, Mouna Ouadghiri, Saaïd Amzazi, Lahcen Belyamani and Azeddine Ibrahimi |
| EPI_ISL_1120949 | Hospital Universitario 12 de Octubre | SeqCOVID-SPAIN consortium/IBV(CSIC) | Esther Viedma, Raúl Recio, Irene Múñoz-Gallego, Jennifer Villa, Rafael Delgado, Mª Dolores Folgueira and SeqCOVID-SPAIN consortiumconsortium |
| EPI_ISL_1121039, EPI_ISL_1121040 | Public Health Virology-Forensic and Scientific Services | Public Health Virology-Forensic and Scientific Services | Son Nguyen |
| EPI_ISL_1121651 | Broad Institute Clinical Research Sequencing Platform | Infectious Disease Program, Broad Institute of Harvard and MIT | Lemieux,J.E., Siddle,K.J., Adams,G., Gladden-Young,A., Lagerborg,K., Rudy,M., DeRuff,K., Carter,A., Normandin,E., Bauer,M., Reilly,S., Tomkins-Tinch,C., Loreth,C., Chaluvadi,S., Birren,B.W., Gallagher,G., Smole,S., Park,D.J., MacInnis,B.L., and Sabeti,P.C. |
| EPI_ISL_1121855 | Rhode Island Department of Health | Infectious Disease Program, Broad Institute of Harvard and MIT | Lemieux,J.E., Siddle,K.J., Huard,R., King,E., Azevedo,K., Miller,A., Adams,G., Gladden-Young,A., Lagerborg,K., Rudy,M., DeRuff,K., Carter,A., Normandin,E., Bauer,M., Reilly,S., Tomkins-Tinch,C., Loreth,C., Chaluvadi,S., Birren,B.W., Gallagher,G., Smole,S., Park,D.J., MacInnis,B.L., and Sabeti,P.C. |

| | | | |
|---|---|---|---|
| EPI_ISL_1121978, EPI_ISL_1121981, EPI_ISL_1121987, EPI_ISL_1121988, EPI_ISL_1121989 | New South Wales Health Pathology Royal Prince Alfred Hospital | Microbiology RPAH | Foster, C.; Au, J.; Ruiz Silva, M.; Deveson, I.; Bull, R.; Van Hal, S.; Rawlinson, W. |
| EPI_ISL_1122013 | Kentucky State Public Health Lab | Kentucky State Public Health Lab | Stephanie Lunn, Karim George, Joshua Tobias, William Grooms, Vaneet Arora, Matthew Johnson, Rachel Zinner, Rhonda Lucas |
| EPI_ISL_1122017 | Emergency department of COVID-19, Beijing Ditan Hospital, Capital Medical University, Beijing, China. | National Institute for Viral Disease Control and Prevention | Yang Song2*, Ziruo Ge1*, Shuping Cui1, 3*, Di Tian1, Gang Wan1, Shuangli Zhu2, Xianbo Wang1, Yu Wang1, Xiang Zhao2, Pan Xiang1, Yanli Xu1, Tingyu Zhang1, Long Liu1, Gang Liu1, Yanhai Wang2, Jianbo Tan1, Wei Zhang1#, Wenbo Xu2# and Zhihai Chen1# |
| EPI_ISL_1122419, EPI_ISL_1122420 | National Public Health Laboratory, National Centre for Infectious Diseases | National Virology Reference Laboratory | Tze Minn Mak, Zhenyang Zhou, Zainun Zaini, Surita Taib, Lin Cui, Raymond Tzer Pin Lin |
| EPI_ISL_1122445 | Cebu TB Reference Laboratory | Philippine Genome Center | Francis A. Tablizo, Kenneth M. Kim, Carlo M. Lapid, Marc Jerrone R. Castro, Maria Sofia L. Yangzon, Benedict A. Maralit, Marc Edsel C. Ayes, Eva Maria Cutiongco-de la Paz, Alethea R. de Guzman, Jan Michael C. Yap, Jo-Hannah S. Llames, Sheila Mae M. Araiza, Kris P. Punayan, Irish Coleen A. Asin, Candice Francheska B. Tambaoan, Asia Louisa U. Chong, Karol Sophia Agape R. Padilla, Rianna Patricia S. Cruz, El King D. Morado, Joshua Gregor A. Dizon, Razel Nikka M. Hao, Arianne A. Zamora, Devon Ray Pacial, Juan Antonio R. Magalang, Marissa Alejandria, Celia Carlos, Anna Ong-Lim, Edsel Maurice Salvaña, John Q. Wong, Jaime C. Montoya, Maria Rosario Singh-Vergeire and Cynthia P. Saloma |
| EPI_ISL_1123937, EPI_ISL_1124275, EPI_ISL_1131764 | Pathogen Genomics Center, National Institute of Infectious Diseases | Pathogen Genomics Center, National Institute of Infectious Diseases | Tsuyoshi Sekizuka, Kentaro Itokawa, Rina Tanaka, Masanori Hashino, Makoto Kuroda |
| EPI_ISL_1132078 | SARS-CoV-2 testing team, National Institute of Infectious Diseases | Pathogen Genomics Center, National Institute of Infectious Diseases | Tsuyoshi Sekizuka, Kentaro Itokawa, Rina Tanaka, Masanori Hashino, Ai Kawana-Tachikawa, Shoji Miki, Shigeru Kusagawa, Makoto Kuroda |
| EPI_ISL_1132829 | Instituto Nacional de Saude (INS), Mozambique | KRISP, KZN Research Innovation and Sequencing Platform | Nalia Ismael, Nadia Sitoe, Paulo Arnaldo, Nedio Mabunda, Giandhari J, Pillay S, Emmanuel S, Tegally H, Wilkinson E, de Oliveira T |
| EPI_ISL_1133274 | Laboratório Hermes Pardini | Laboratório de Biologia Integrativa, Instituto de Ciências Biológicas, Universidade Federal de Minas Gerais | Filipe Romero Rebello Moreira, Diego Menezes Bonfim, Danielle Alves Gomes Zauli, Joice do Prado Silva, Aline Brito de Lima, Frederico Scott Varella Malta, Alessandro Clayton de Souza Ferreira, Victor Cavalcanti Pardini, Daniel Costa Queiroz, Rafael Marques de Souza, Victor Emmanuel Viana Geddes, Walyson Coelho Costa, Wagner Carlos Santos Magalhaes, Rennan Garcias Moreira, Carolina Moreira Voloch, Renan Pedra de Souza, Renato Santana Aguiar. |
| EPI_ISL_1137503, EPI_ISL_1137504, EPI_ISL_1137505 | Public Health Virology-Forensic and Scientific Services | Public Health Virology-Forensic and Scientific Services | Son Nguyen |
| EPI_ISL_1138421, EPI_ISL_1138422 | Laboratorio de Referencia Nacional de Virus Respiratorio. Instituto Nacional de Salud Perú | Laboratorio de Referencia Nacional de Biotecnología y Biología Molecular. Instituto Nacional de Salud Perú | Carlos Padilla Rojas, Karolyn Vega Chozo, Luis Barcena, Priscila Lope Pari, Omar Caceres Rey, Marco Galarza Perez, Maribel Huaringa Nuñez, Johanna Balbuena Torrez, Henri Bailon Calderon, Nancy Rojas Serrano |
| EPI_ISL_1138512, EPI_ISL_1138513, EPI_ISL_1138515 | SA Pärnu Hospital Laboratory | 1. Laboratory of Communicable Diseases (Estonia); 2. Eurofins Genomics Europe Sequencing GmbH | Liidia Dotsenko |
| EPI_ISL_1138757 | General Hospital - Strumica | Research Center for Genetic Engineering and Biotechnology "Georgi D. Efremov" , Macedonian Academy of Sciences and Arts | Aleksandar J. Dimovski, Dijana Plasheska-Karanfilska, Predrag Noveski, Gjorgji Bozinovski, Milena Jakimovska |
| EPI_ISL_1138758 | General Hospital - Struga | Research Center for Genetic Engineering and Biotechnology "Georgi D. Efremov" , Macedonian Academy of Sciences and Arts | Aleksandar J. Dimovski, Dijana Plasheska-Karanfilska, Predrag Noveski, Gjorgji Bozinovski, Milena Jakimovska |
| EPI_ISL_1138834, EPI_ISL_1138870 | Office of Diseases Prevention and Control Region 4 Saraburi | COVID-19 Network Investigations (CONI) Alliance | Elizabeth Batty, Wasun Chantratita, Thanat Chookajorn, Stefan Fernandez, Angkana Huang, Anthony R. Jones, Khajohn Joonlasak, Chonticha Klungtong, Theerarat Kochakarn, Namfon Kotanan, Krittikorn Kumpornsin, Duangkamon Loesbanluechai, Wudtichai Manasatienkij, Bhakbhoom Panthan, Ekawat Pasomsub, Kingkan Rakmanee, Insee Sensorn, Janjira Thaipadungpanit, Arporn Wangwiwatsin, Treewat Watthanachockchai, Suttiruk Changchawai, Sataporn Hatsadichart, Jutikul Kaewmalakul, Siriporn Lakesukthom, Wonvimol Lemprasert, Praima Moonmuang, Payon Pengyo, Pakjira Rimdusit, Nathamon Runnachot, Sirinapa Singthong, Siriwan Yaemnimnual |
| EPI_ISL_1138954 | Division of Emerging Infectious Diseases, Bureau of Infectious Diseases Diagnosis Control, Korea Disease Control and Prevention Agency | Division of Emerging Infectious Diseases, Bureau of Infectious Diseases Diagnosis Control, Korea Disease Control and Prevention Agency | Ae Kyung Park, Il-Hwan Kim, Heui Man Kim, Jeong-Min Kim, Namjoo Lee, Chae Young Lee, Sang Hee Woo, Eun-Jin Kim |
| EPI_ISL_1138983 | Laboratory for HIV and opportunistic infections diagnosis The Republican Research and Practical Center for Epidemiology and Microbiology (RRPCEM) | Laboratory for HIV and opportunistic infections diagnosis The Republican Research and Practical Center for Epidemiology and Microbiology (RRPCEM) | Elena Gasich, Kirill Bulda, Artur Akhremchuk, Leonid Valentovich, Anatoly Krasko, Vladimir Gorbunov |
| EPI_ISL_1139056 | LACEN do Mato Grosso do Sul | Instituto Adolfo Lutz, Interdiciplinary Procedures Center, Strategic Laboratory | Claudio Tavares Sacchi, Claudia Regina Gonçalves, Erica Valessa Ramos Gomes, Karoline Rodrigues Campos, Caio Vinicius Dias Lopes |
| EPI_ISL_1139157 | National Center of Disease Control and Prevention of the Republic of Armenia | Institute of Molecular Biology NAS RA, Republic of Armenia, Department of Bioengineering, BioinformaticsInstitute and Molecular Biology IBMPh RAU, Republic of Armenia | Arsen Arakelyan, Diana Avetyan, Siras Hakobyan, Gisane Khachatyan, Maria Nikoghosyan, Tamara Sirunyan, Nelli Muradyan, Andranik Chavushyan, Hovsep Ghazaryan, Roksana Zakharyan, Shushan Sargrsyan, Gayane Melik-Pashayan |
| EPI_ISL_1140120 | General Hospital - Shtip | Research Center for Genetic Engineering and Biotechnology "Georgi D. Efremov" , Macedonian Academy of Sciences and Arts | Aleksandar J. Dimovski, Dijana Plasheska-Karanfilska, Predrag Noveski, Gjorgji Bozinovski, Milena Jakimovska |
| EPI_ISL_1141116 | amedes MVZ Halle/Leipzig | Robert Koch Institute | unknown |
| EPI_ISL_1141939 | SYNLAB Jena Oncoscreen | Robert Koch Institute | unknown |
| EPI_ISL_1142461 | Bioscientia Labor Wermsdorf | Robert Koch Institute | unknown |
| EPI_ISL_1143740, EPI_ISL_1143963 | Bayerisches Landesamt für Gesundheit und Lebensmittelsicherheit (LGL) | Robert Koch Institute | unknown |
| EPI_ISL_1151450 | Praxisgemeinschaft für Laboratoriumsmedizin Labor Blumenstraße; Praxis Dr. E. Wietschel & Dr. F. Wietschel | Robert Koch Institute | unknown |
| EPI_ISL_1157744 | SYNLAB MVZ Weiden | Robert Koch Institute | unknown |
| EPI_ISL_1159294 | Quest Diagnostics Incorporated | Centers for Disease Control and Prevention Division of Viral Diseases, Pathogen Discovery | Peter W. Cook, Dakota Howard, Dhwani Batra, Ben L. Rambo-Martin, S. H. Rosenthal, A. Gerasimova, R. M. Kagan, B. Anderson, M. Hua, Y. Liu, L.E. Bernstein, K.E. Livingston, A. Perez, I. A. Shlyakhter, R. V. Rolando, R. Owen, P. Tanpaiboon, F. Lacbawan, Clinton R. Paden, Suxiang Tong, Duncan MacCannell |
| EPI_ISL_1159698, EPI_ISL_1159700 | Laboratoire central de Virologie | Laboratoire de Biotechnologie | Tarik Aanniz, Mouna Ouadghiri, Abdelomunim Essabbar, Ghizlane EL Amin, Amal Zouaki, Myriam Seffar, Hakima Kabbaj, Saaïd Amzazi, Lahcen Belyamani and Azeddine Ibrahimi |
| EPI_ISL_1161478, EPI_ISL_1161704 | Laboratory Corporation of America | Respiratory Viruses Branch, Division of Viral Diseases, Centers for Disease Control and Prevention | Peter W. Cook, Dakota Howard, Dhwani Batra, Ben L. Rambo-Martin, Minoo Agarwal, Eyad Almasri Debbie Boles, Ayla Burns, Nuthawin Charoensri, Oren Cohen, Susan Countryman, Mary Ann Cristobal, Bobbi Croy, Suzanne Dale, Hrushikesh Deshmukh, Amanda Douglas, Vincent Drouillon, Marcia Eisenberg, Howard Engler, Rama Ghatti, Prashant Gupta, Susan Hicks, Jake Humphrey, Lax Iyer, Manoj Jain, Mohan Kolli, Brian Krueger, Tim Kuphal, Stanley Letovsky, Michael Levandoski, Craig Lukasik, Jonathan Meltzer, Brian Norvell, Mindy Nye, Scott Parker, Christos Petropoulos, John Pruitt, Steven Ragan, Scott Ryan, Mike Sapeta, Jana Schroth, Suresh Babu Selvaraju, Goran Stevovic, Amanda Suchanek, Andrea Throop, Lyndon Tilson, Thomas Urban, Joe Voshell, Kimberly Wagner, Jonathan Williams, Mary Williamson, Qian Zeng, Tricia Zwiefelhofer, Clinton R. Paden, Suxiang Tong, Duncan MacCannell |

| | | | |
|---|---|---|---|
| EPI_ISL_1164326 | Centro Hospitalar de Entre o Douro e Vouga (CHEDV) | Institute of Biomedicine (IBiMED), Universidade de Aveiro | Gabriela Moura, Sofia Marques, Patricia Arinto, Miguel Pinheiro and Manuel Santos |
| EPI_ISL_1164606 | Salzkammergutklinikum Vöcklabruck, Institut für Pathologie | Salzkammergutklinikum Vöcklabruck, Institut für Pathologie | Regina Stitz |
| EPI_ISL_1164627, EPI_ISL_1164632, EPI_ISL_1164634, EPI_ISL_1164662, EPI_ISL_1164688, EPI_ISL_1164699, EPI_ISL_1164703, EPI_ISL_1164706, EPI_ISL_1164709, EPI_ISL_1164717, EPI_ISL_1164718, EPI_ISL_1164721, EPI_ISL_1164724, EPI_ISL_1164728, EPI_ISL_1164732, EPI_ISL_1164736, EPI_ISL_1164738, EPI_ISL_1164739, EPI_ISL_1164745 | | | |
| see above | Department of Molecular Virology, Cyprus Institute of Neurology and Genetics | Department of Molecular Virology, Cyprus Institute of Neurology and Genetics | Jan Richter, Pavlos Fanis, Christina Tryfonos, Dana Koptides, George Krashias, Stavros Bashiardes, Andreas Hadjisavvas, Maria Loizidou, Anastasis Oulas, Denise Alexandrou, Olga Kalakouta, Mihalis Panayiotidis, George Spyrou, Christina Christodoulou |
| EPI_ISL_1164791, EPI_ISL_1164810 | Idaho Bureau of Laboratories | IBL | "R. Beukelman, Matthew C. Burns, Aimee Ceniseros, Robert L. Voermans, Christopher Ball" |
| EPI_ISL_1165024 | Division of Emerging Infectious Diseases, Bureau of Infectious Diseases Diagnosis Control, Korea Disease Control and Prevention Agency | Division of Emerging Infectious Diseases, Bureau of Infectious Diseases Diagnosis Control, Korea Disease Control and Prevention Agency | Ae Kyung Park, Il-Hwan Kim, Heui Man Kim, Jeong-Min Kim, Namjoo Lee, Chae Young Lee, Sang Hee Woo, Eun-Jin Kim |
| EPI_ISL_1165379, EPI_ISL_1165512, EPI_ISL_1165593 | Dutch COVID-19 response team | National Institute for Public Health and the Environment (RIVM) | Adam Meijer, Harry Vennema, Dirk Eggink, Jeroen Cremer, Sharon van den Brink, Bas van der Veer, AnneMarie van den Brandt, Florian Zwagemaker, Dennis Schmitz, Chantal Reusken, on behalf of the national COVID-19 response team |
| EPI_ISL_1166981 | Hopital | National Reference Center for Viruses of Respiratory Infections, Institut Pasteur, Paris | Marion Barbet, Sylvie Behillil, Méline Bizard, Angela Brisebarre, Camille Capel, Etienne Simon-Lorière, Vincent Enouf, Maud Vanpeene, Sylvie van der Werf,Combe Patrice |
| EPI_ISL_1167030, EPI_ISL_1167032, EPI_ISL_1167034, EPI_ISL_1167057, EPI_ISL_1167067, EPI_ISL_1167074, EPI_ISL_1167083, EPI_ISL_1167084 | "Dr. Andrija Stampar" Teaching Institute of Public Health, Department of Clinical Microbiology | Istituto di Genomica Applicata | Jasmina Vranes, Slobodanka Radovic, Fedrica Cattonaro, Irena Jurman, Vera Vendramin, Gabriele Magris, Eleonora Paparelli, Davide Scaglione, Michele Morgante |
| EPI_ISL_1167127, EPI_ISL_1167128, EPI_ISL_1167134, EPI_ISL_1167139, EPI_ISL_1167141, EPI_ISL_1167146, EPI_ISL_1167157, EPI_ISL_1167163, EPI_ISL_1167181 | Iressef Genomics lab | L'institut de Recherche en Santé, de Surveillance Épidémiologique et de Formation (IRESSEF) | Souleymane MBOUP, Abdou PADANE, Abdoulie KANTEH ,Abdul Karim SESAY , Khadim GUEYE, Papa Alassane DIAW, Birahim Piere NDIAYE, Barada CISSE, Aminata MBOUP, Moustapha MBOW, Ndeye Coumba Toure KANE, Nafisatou LEYE, Gora LO, Ambroise AHOUIDI , Astou Gaye GAYE, Aminata DIA, Yacine DIA |
| EPI_ISL_1167549 | Kansas Health and Environmental Lab | Kansas Health and Environmental Lab | Mike Grose, Jonathan Barnell, Ben Olsen, and Phil Adam |
| EPI_ISL_1167672, EPI_ISL_1167674, EPI_ISL_1167687, EPI_ISL_1167753, EPI_ISL_1167754, EPI_ISL_1167806, EPI_ISL_1167810, EPI_ISL_1167812, EPI_ISL_1167825, EPI_ISL_1167829, EPI_ISL_1167841, EPI_ISL_1167852, EPI_ISL_1167865, EPI_ISL_1167878, EPI_ISL_1167881, EPI_ISL_1167883, EPI_ISL_1167899, EPI_ISL_1167902, EPI_ISL_1167905, EPI_ISL_1167906, EPI_ISL_1167911, EPI_ISL_1167924, EPI_ISL_1167930 | | | |
| see above | Genetica Molecular and Subdepartamento de Virologia ISP Chile | Instituto de Salud Publica de Chile | Javier Tognarelli, Karen Orostica, Barbara Parra, Loredana Arata, Jaime Lagos, Gisselle Barra, Patricia Bustos, Rodrigo Fasce, Andres Castillo, Jorge Fernandez |
| EPI_ISL_1168193, EPI_ISL_1168194 | Hopital | National Reference Center for Viruses of Respiratory Infections, Institut Pasteur, Paris | Marion Barbet, Sylvie Behillil, Méline Bizard, Angela Brisebarre, Camille Capel, Etienne Simon-Lorière, Vincent Enouf, Maud Vanpeene, Sylvie van der Werf,Hermann CéCile |
| EPI_ISL_1168457, EPI_ISL_1168521, EPI_ISL_1168522, EPI_ISL_1168524, EPI_ISL_1168527, EPI_ISL_1168530, EPI_ISL_1168541, EPI_ISL_1168575, EPI_ISL_1168608, EPI_ISL_1168609, EPI_ISL_1168614, EPI_ISL_1168615, EPI_ISL_1168616, EPI_ISL_1168618, EPI_ISL_1168626, EPI_ISL_1168627 | | | |
| see above | Instituto de Diagnostico y Referencia Epidemiologicos INDRE_RNLSP | Instituto de Diagnostico y Referencia Epidemiologicos (INDRE) | Claudia Wong-Arambula, Abril Rodriguez-Maldonado, Vanessa Rivero-Arredondo, Ariadna Medina-Benitez, Joaquin Quiroz-Mercado,David Fragoso-Fonseca, Sergio Rangel-Guerrero, Natividad Cruz-Ortiz, Tatiana Nunez-Garcia, Gisela Barrera-Badillo, Lucia Hernandez-Rivas, Irma Lopez-Martinez, Ernesto Ramirez-Gonzalez. |
| EPI_ISL_1169046 | National Institute of Health Research and Development | National Institute of Health Research and Development | Hana Apsari Pawestri, Subangkit, Kartika Dewi Puspa, Arie Ardiansyah Nugraha, Hartanti Dian Ikawati, Krisna Nur Andriana Pangesti, Yuni Rukminiati, Ririn Ramadhany, Agustiningsih, Kindi Adam, Holy Arif Wibowo, Triyani Soekarso, Ni Ketut Susilarini, Nurika Hariastuti, Ully Alfi Nikmah, Reni Herman, Nike Susanti, Herna, Tati Febriyanti, Natalie Laurencia Kipuw, Fauzul Muna, Irene Lorinda Indalao, Nelly Puspandari, Vivi Setiawaty. |
| EPI_ISL_1169058 | National Institute of Health Research and Development | National Institute of Health Research and Development | Arie Ardiansyah Nugraha, Kartika Dewi Puspa, Hana Apsari Pawestri, Subangkit, , Hartanti Dian Ikawati, Krisna Nur Andriana Pangesti, Yuni Rukminiati, Ririn Ramadhany, Agustiningsih, Kindi Adam, Holy Arif Wibowo, Triyani Soekarso, Ni Ketut Susilarini, Nurika Hariastuti, Ully Alfi Nikmah, Reni Herman, Nike Susanti, Herna, Tati Febriyanti, Natalie Laurencia Kipuw, Fauzul Muna, Irene Lorinda Indalao, Nelly Puspandari, Vivi Setiawaty. |
| EPI_ISL_1169497, EPI_ISL_1169524, EPI_ISL_1169543, EPI_ISL_1169549 | Commonwealth Healthcare Center | Genomics and Discovery, Respiratory Viruses Branch, Division of Viral Diseases, Centers for Disease Control and Prevention | Krista Queen, Yan Li, Ying Tao, Jing Zhang, Anna Uehara, Anna Montmayeur, Clinton R. Paden, Peter W. Cook, Rachel Marine, Mili Sheth, Jasmine Padilla, Sarah Nobles, Mark Burroughs, Lori Rowe, Haibin Wang, Ben L. Rambo-Martin, Dhwani Batra, Justin Lee, Suxiang Tong |
| EPI_ISL_1170946, EPI_ISL_1170947 | ACT Pathology | Schwessinger Lab | Ashley Jones, Benjamin Schwessinger, Robert Lanfear, Megan McDonald, Ming-Dao Chia, Kevin Murray, Robyn N Hall, Craig Kennedy, Karina Kennedy |
| EPI_ISL_1170953 | PHV-FSS | PHV-FSS | Son Nguyen |
| EPI_ISL_1171204, EPI_ISL_1171231 | OHSU Lab Services Molecular Microbiology Lab | Ginkgo Bioworks Clinical Laboratory | Brendan L. O'Connell, Sally Grindstaff, Kayla Carter, Ruth V. Nichols, Malaika Mckenzie-Bennett, James McGann, Jim Griffin, Keith Robison, Alex Plocik, Becky Schilling, Martha Pierson, Rebecca Littlefield, Michelle Spencer, Amanda Riccardi, Birgitte Simen, Alec J. Hirsch, Donna Hansel, Guang Fan, Xuan Qin, Daniel N. Streblow, William B. Messer, Andrew C. Adey, Benjamin N. Bimber, Brian J. O'Roak |
| EPI_ISL_1171642 | IAL Regional de Marilia | Instituto Adolfo Lutz, Interdiciplinary Procedures Center, Strategic Laboratory | Claudio Tavares Sacchi, Claudia Regina Gonçalves, Erica Valessa Ramos Gomes, Karoline Rodrigues Campos, Caio Vinicius Dias Lopes |
| EPI_ISL_1172030, EPI_ISL_1172032 | LabPLUS | Institute of Environmental Science and Research (ESR) | Rachel Boyle, SallyAnn Harbison, Olivia Stroeven, Xiaoyun Ren, Matt Storey, Nikki Freed, Muhammad Faisal, Jing Wang, Hermes Perez, Anja Werno, Antje van der Linden, Arlo Upton, Chris Mansell, David Hammer, Dragana Drinkovic, Gary McAuliffe, Hana Sofia Andersson, James Ussher, Jill Sherwood, Josh Freeman, Julia Howard, Juliet Elvy, Mary DeAlmeida, Matt Blakiston, Matthew Rogers, Max Bloomfield, Michael Addidle, Michelle Balm, Sally Roberts, Sarah Jefferies, Sharmini Muttaiyah, Susan Morpeth, Susan Taylor, Timothy Blackmore, Vani Sathyendran, Veronica Playle, Virginia Hope, Erasmus Smit, Lauren Jelly, Olin Silander, Joep de Ligt |
| EPI_ISL_1180152 | Laboratory of virology and molecular diagnostics | Laboratory of virology and molecular diagnostics | Kuzmanovska Maja, Boshevska Golubinka, Elizabeta Janchevska |
| EPI_ISL_1180454 | National Virus Reference Laboratory | National Virus Reference Laboratory | Zoe Yandle, Charlene Bennett, Gabriel Gonzalez, Michael Carr, Jonathan Dean, Cillian F De Gascun |
| EPI_ISL_1180807 | Austrian Agency for Health and Food Safety (AGES) | Bergthaler laboratory, CeMM Research Center for Molecular Medicine of the Austrian Academy of Sciences | Lukas Endler, Anna Schedl, Fabian Amman, Thomas Penz, Benedikt Agerer, Maelle Le Moing, Michael Schuster, Bekir Erguner, Jan Laine, Martin Senekowitsch, Christoph Bock, Andreas Bergthaler |
| EPI_ISL_1180844 | Department of Laboratory Medicine, Division of Clinical Virology, University of Medicine, Vienna | Bergthaler laboratory, CeMM Research Center for Molecular Medicine of the Austrian Academy of Sciences | Lukas Endler, Anna Schedl, Fabian Amman, Thomas Penz, Benedikt Agerer, Maelle Le Moing, Michael Schuster, Bekir Erguner, Jan Laine, Martin Senekowitsch, Christoph Bock, Andreas Bergthaler |
| EPI_ISL_1181116 | genXone SA, Molecular Diagnostics Laboratory / NZOZ | genXone SA, Research & Development Laboratory | Maciej Sykulski, Grzegorz Nowicki, Jakub Grabowski, Natalia Drweska-Matelska, Anna Brylak-Blaszkow, Aleksandra Gidlewicz, Karol Szeszko, Lukasz Krych, Michal Kaszuba |
| EPI_ISL_1181360 | Laboratory of Respiratory Viruses and Measles, Oswaldo Cruz Institute, FIOCRUZ | Laboratory of Respiratory Viruses and Measles, Oswaldo Cruz Institute, FIOCRUZ | Paola Resende, Luciana Appolinario, Fernando Motta, Anna Carolina Paixao, Ana Carolina Mendonca, Alice Sampaio Rocha, Renata Serrano Lopes, Marilda Siqueira on behalf of the Fiocruz COVID-19 Genomic Surveillance Network |
| EPI_ISL_1181409 | Laboratorio Central de Saude Publica do Estado de Sergipe (LACEN-SE) | Laboratory of Respiratory Viruses and Measles, Oswaldo Cruz Institute, FIOCRUZ | Paola Resende, Luciana Appolinario, Fernando Motta, Anna Carolina Paixao, Ana Carolina Mendonca, Alice Sampaio Rocha, Renata Serrano Lopes, Cliomar Alves dos Santos, Marilda Siqueira on behalf of the Fiocruz COVID-19 Genomic Surveillance Network |
| EPI_ISL_1181415 | Laboratory of Respiratory Viruses and Measles, Oswaldo Cruz Institute, FIOCRUZ | Laboratory of Respiratory Viruses and Measles, Oswaldo Cruz Institute, FIOCRUZ | Paola Resende, Luciana Appolinario, Fernando Motta, Anna Carolina Paixao, Ana Carolina Mendonca, Alice Sampaio Rocha, Renata Serrano Lopes, Marilda Siqueira on behalf of the Fiocruz COVID-19 Genomic Surveillance Network |
| EPI_ISL_1181559, EPI_ISL_1181564 | Laboratorio Central de Saude Publica do Estado do Parana (LACEN-PR) | Laboratory of Respiratory Viruses and Measles, Oswaldo Cruz Institute, FIOCRUZ | Paola Resende, Luciana Appolinario, Fernando Motta, Anna Carolina Paixao, Ana Carolina Mendonca, Alice Sampaio Rocha, Renata Serrano Lopes, Maria do Carmo Debur, Irina Nastassja Riediger, Marilda Siqueira on behalf of the Fiocruz COVID-19 Genomic Surveillance Network |

| | | | |
|---|---|---|---|
| EPI_ISL_1182581 | Laboratório Central do Estado do Paraná | Coordenação Geral de Laboratórios de Saúde Pública (CGLAB/DAEVS/SVS/MS) | Vagner Fonseca, et al. |
| EPI_ISL_1182727 | Alaska State Virology Laboratory | Alaska State Virology Laboratory | Stephanie DeRonde, Lisa Smith, Ph.D., Jack Chen, Ph.D. |
| EPI_ISL_1184175 | Indiana Animal Disease Diagnostic Laboratory | Carpi Laboratory - Purdue University | Jack Dorman, Ilinca I Ciubotariu, Lev Gorenstein, Abebe A Fola, G Kenitra Hendrix, Rebecca P Wilkes, Giovanna Carpi |
| EPI_ISL_1184505, EPI_ISL_1184506 | Area of Virology, Serology and Virology Division (SAViD), New South Wales Health Pathology Randwick | Virology Research Laboratory; Area of Virology, Serology and Virology Division (SAViD), New South Wales Health Pathology Randwick | Foster, C.; Au, J.; Ruiz Silva, M.; Deveson, I.; Bull, R.; Van Hal, S.; Rawlinson, W. |
| EPI_ISL_1184507 | PathWest Laboratory Medicine WA | PathWest Laboratory Medicine WA Microbial Surveillance Unit | PathWest Laboratory Medicine WA Microbial Surveillance Unit |
| EPI_ISL_1186028 | Institute for Medical Research, Infectious Disease Research Centre, National Institutes of Health, Ministry of Health Malaysia | Institute for Medical Research, Infectious Disease Research Centre, National Institutes of Health, Ministry of Health Malaysia | Suppiah J, Kamel K, Mohd Zawawi Z, Ramly N, Robert F, Thayan R |
| EPI_ISL_1190751, EPI_ISL_1190754, EPI_ISL_1190765 | CREMER(Centre de Rechercherches sur les Maladies Emergentes et Ré-émergentes) | TransVIHMI(Recherches Translationnelles sur le VIH et les Maladies Infectieuses) | Celestin Godwe, Christelle Butel, Martin Maidadi Foudi, Laetitia Serrano, Dowbiss Meta Djomsi, Nicole Vidal, Esemu Livo, Rodrigue Kamga, Marie Amougou , Eitel Mpoudi Ngole,Ahidjo Ayouba, Marcel Tongo, Martine Peeters, Eric Delaporte |
| EPI_ISL_1191600, EPI_ISL_1191603, EPI_ISL_1191604, EPI_ISL_1191605 | Virology Department, Victoria Hospital, Plaine-Wilhems, Mauritius | National Institute for Communicable Diseases of the National Health Laboratory Service | Ramuth M, Manraj SS, Sonoo J, Baboo SB, Amoako DG, Scheepers C, Mohale T, Ntuli N, Mahlangu B, Ismail A, Bhiman JN |
| EPI_ISL_1191740 | Laboratory of virology and molecular diagnostics | Laboratory of virology and molecular diagnostics | Kuzmanovska Maja, Boshevska Golubinka, Janchevska Elizabeta |
| EPI_ISL_1191790, EPI_ISL_1191792, EPI_ISL_1191809, EPI_ISL_1191818, EPI_ISL_1191834, EPI_ISL_1191845, EPI_ISL_1191855, EPI_ISL_1191871, EPI_ISL_1191873, EPI_ISL_1191875, EPI_ISL_1191876, EPI_ISL_1191887, EPI_ISL_1191896, EPI_ISL_1191915, EPI_ISL_1191922, EPI_ISL_1191927, EPI_ISL_1191940, EPI_ISL_1191970, EPI_ISL_1191979, EPI_ISL_1191994, EPI_ISL_1192012, EPI_ISL_1192022, EPI_ISL_1192026, EPI_ISL_1192028, EPI_ISL_1192032, EPI_ISL_1192047 | | | |
| see above | National Microbiology Reference Laboratory | Quadram Institute Bioscience | Tapfumanei Mashe, Faustinos T Takawira, Hlanai Gumbo, Kenneth K Maeka, Agnes Juru, Charles Nyagupe, Sekesai Zinyowera, Muchaneta Mugabe, Thanh Le Viet, Justin O'Grady, Gemma Kay, David Baker, Gaetan Thilliez, Ana-Victoria Gutierrez, Robert Kingsley, Leonardo de Oliveira Martins, Andrew Tarupiwa, Andrew J. Page, Raiva Simbi |
| EPI_ISL_1192613, EPI_ISL_1192885, EPI_ISL_1193023 | Swedish national genomic surveillance program of SARS-CoV-2 | The Public Health Agency of Sweden | Swedish national genomic surveillance program of SARS-CoV-2 |
| EPI_ISL_1193292 | Laboratory Corporation of America | Centers for Disease Control and Prevention Division of Viral Diseases, Pathogen Discovery | Peter W. Cook, Dakota Howard, Dhwani Batra, Ben L. Rambo-Martin, Minoo Agarwal, Eyad Almasri Debbie Boles, Ayla Burns, Nuthawin Charoensri, Oren Cohen, Susan Countryman, Mary Ann Cristobal, Bobbi Croy, Suzanne Dale, Hrushikesh Deshmukh, Amanda Douglas, Vincent Drouillon, Marcia Eisenberg, Howard Engler, Rama Ghatti, Prashant Gupta, Susan Hicks, Jake Humphrey, Lax Iyer, Manoj Jain, Mohan Kolli, Brian Krueger, Tim Kuphal, Stanley Letovsky, Michael Levandoski, Craig Lukasik, Jonathan Meltzer, Brian Norvell, Mindy Nye, Scott Parker, Christos Petropoulos, John Pruitt, Steven Ragan, Scott Ryan, Mike Sapeta, Jana Schroth, Suresh Babu Selvaraju, Goran Stevovic, Amanda Suchanek, Andrea Throop, Lyndon Tilson, Thomas Urban, Joe Voshell, Kimberly Wagner, Jonathan Williams, Mary Williamson, Qian Zeng, Tricia Zwiefelhofer, Clinton R. Paden, Suxiang Tong, Duncan MacCannell |
| EPI_ISL_1193544, EPI_ISL_1193914 | Quest Diagnostics Incorporated | Centers for Disease Control and Prevention Division of Viral Diseases, Pathogen Discovery | Peter W. Cook, Dakota Howard, Dhwani Batra, Ben L. Rambo-Martin, S. H. Rosenthal, A. Gerasimova, R. M. Kagan, B. Anderson, M. Hua, Y. Liu, L.E. Bernstein, K.E. Livingston, A. Perez, I. A. Shlyakhter, R. V. Rolando, R. Owen, P. Tanpaiboon, F. Lacbawan, Clinton R. Paden, Suxiang Tong, Duncan MacCannell |
| EPI_ISL_1195198, EPI_ISL_1195202, EPI_ISL_1195207 | National Public Health Center, COVID Laboratory | National Public Health Center, National Biosafety Laboratory | Bernadett Pályi, Zoltán Kis, Nóra Magyar, Judit Henczkó, Dániel Déri, Norbert Solymosi |
| EPI_ISL_1195498 | DC Public Health Lab/ Dept. of Forensic Sciences | DC Public Health Lab/ Dept. of Forensic Sciences | Janis Doss, Scott Nguyen, Elizabeth Zelaya, Sarah Scott, Connie Maza, Monica Mann, Brittany Hamilton, David Payne, Jocelyn Hauser |
| EPI_ISL_1195726 | Public Health Institute of Šibenik-Knin County | Croatian Institute of Public Health | Irena Tabain, Ivana Ferenak |
| EPI_ISL_1195975 | Alaska State Virology Laboratory | Alaska State Virology Laboratory | Stephanie DeRonde, Elva House, Lisa Smith, Ph.D., Jack Chen, Ph.D. |
| EPI_ISL_1196005 | RSP Clinic | National Institute for Communicable Diseases of the National Health Laboratory Service | Maphalala GP, Amoako DG, Scheepers C, Mohale T, Ntuli N, Mahlangu B, Ismail A, Bhiman JN |
| EPI_ISL_1196006 | TLC Clinic | National Institute for Communicable Diseases of the National Health Laboratory Service | Maphalala GP, Amoako DG, Scheepers C, Mohale T, Ntuli N, Mahlangu B, Ismail A, Bhiman JN |
| EPI_ISL_1196008 | Nhlangano Health Centre | National Institute for Communicable Diseases of the National Health Laboratory Service | Maphalala GP, Amoako DG, Scheepers C, Mohale T, Ntuli N, Mahlangu B, Ismail A, Bhiman JN |
| EPI_ISL_1196011, EPI_ISL_1196012 | Homecare | National Institute for Communicable Diseases of the National Health Laboratory Service | Maphalala GP, Amoako DG, Scheepers C, Mohale T, Ntuli N, Mahlangu B, Ismail A, Bhiman JN |
| EPI_ISL_1196145 | Ministry of Health Turkey | Ministry of Health Turkey | Fatma Bayrakdar, Yasemin Cosgun, Suleyman Yalcin, Gulay Korukluoglu |
| EPI_ISL_1196421 | AREA DE SALUD CORREDORES | Inciensa, Instituto Costarricense de Investigación y Enseñanza en Nutrición y Salud | Francisco Duarte, Hebleen Porras, Claudio Soto-Garita, Estela Cordero, Adriana Godínez, Melany Calderón & Mariel López |
| EPI_ISL_1196426, EPI_ISL_1196427 | LABORATORIO CLINICO CENAHCE | Inciensa, Instituto Costarricense de Investigación y Enseñanza en Nutrición y Salud | Francisco Duarte, Hebleen Porras, Claudio Soto-Garita, Estela Cordero, Adriana Godínez, Melany Calderón & Andrés Feoli-Grant |
| EPI_ISL_1196430 | HOSPITAL DR. MAX PERALTA JIMENEZ | Inciensa, Instituto Costarricense de Investigación y Enseñanza en Nutrición y Salud | Francisco Duarte, Hebleen Porras, Claudio Soto-Garita, Estela Cordero, Adriana Godínez, Melany Calderón & Mónica Charpentier-Artavia |
| EPI_ISL_1196431 | HOSPITAL MEXICO | Inciensa, Instituto Costarricense de Investigación y Enseñanza en Nutrición y Salud | Francisco Duarte, Hebleen Porras, Claudio Soto-Garita, Estela Cordero, Adriana Godínez, Melany Calderón & Teresita Somogyi |
| EPI_ISL_1196433 | HOSPITAL DR. ENRIQUE BALTODANO BRICEÑO | Inciensa, Instituto Costarricense de Investigación y Enseñanza en Nutrición y Salud | Francisco Duarte, Hebleen Porras, Claudio Soto-Garita, Estela Cordero, Adriana Godínez, Melany Calderón & Adriana Bermúdez-Espinoza |
| EPI_ISL_1196866 | Institute for Medical Research, Infectious Disease Research Centre, National Institutes of Health, Ministry of Health Malaysia | Institute for Medical Research, Infectious Disease Research Centre, National Institutes of Health, Ministry of Health Malaysia | Suppiah J, Kamel K, Mohd Zawawi Z, Ramly N, Robert F, Thayan R |
| EPI_ISL_1196893 | MT Public Health Laboratory | Pathogen Discovery, Respiratory Viruses Branch, Division of Viral Diseases, Centers for Disease Control and Prevention | Ying Tao, Jing Zhang, Yan Li, Brian Lynch, Anna Kelleher, Krista Queen, Anna Uehara, Peter Cook, Han Jia Justin Ng, Clinton R. Paden, Haibin Wang, Suxiang Tong |
| EPI_ISL_1197037 | Laboratoire de virologie clinique - Institut Pasteur de Tunis | 1-Laboratory of Microbiology, National Reference Lab, Charles Nicolle Hospital; 2-University of Tunis ElManar, Faculty of Medicine of Tunis, LR99ES09, Tunis, Tunisia | Sameh Trabelsi, Nissaf Ben Alaya, Mouna Ben Sassi, Sana Ferjani, Mouna Safer, Salma Abid, Fares Wasfi, Mariem Gdoura, Sondes Haddad, Anissa Chouikha, Roua Ben Othman, Sara Chammam, Ines Mdini, Manel Ben Sassi, Imen Kacem, Maher Kharrat, Alia BenKahla, Jalila Ben Khelil, Riadh Daghfous, Riadh Gouider, Henda Triki, Ilhem Boutiba-Ben Boubaker. |
| EPI_ISL_1197064 | Nigeria Centre For Disease Control | National reference Laboratory, NCDC, Gaduwa, Abuja | Dr Ndodo Nnaemeka, Olusola Anuoluwapo Akanbi, Chimaobi Chukwu, Dr Omoare Adesuyi, Catherine Okoi, Anthony Ahumibe, Naidoo Dhamari, Nwando Mba, Dr Chikwe Ihekweazu |
| EPI_ISL_1197084, EPI_ISL_1197090, EPI_ISL_1197102, EPI_ISL_1197106, EPI_ISL_1197109 | Department of Microbiology, The University of Hong Kong | Department of Microbiology, The University of Hong Kong | Kelvin K.W. To, Kwok-Yung Yuen |
| EPI_ISL_1200660 | BIOMNIS LYON | CNR Virus des Infections Respiratoires - France SUD | Antonin Bal, Gregory Destras, Gwendolyne Burfin, Hadrien Regue, Quentin Semanas, Martine Valette, Bruno Lina, Laurence Josset |

| | | | |
|---|---|---|---|
| EPI_ISL_1201431 | Laboratory of Genetics and Personalized Medicine, Zan Mitrev Clinic | Laboratory of Genetics and Personalized Medicine, Zan Mitrev Clinic | Kungulovski G. et al. |
| EPI_ISL_1201507 | Hopital | National Reference Center for Viruses of Respiratory Infections, Institut Pasteur, Paris | Marion Barbet, Sylvie Behillil, Méline Bizard, Angela Brisebarre, Camille Capel, Etienne Simon-Lorière, Vincent Enouf, Maud Vanpeene, Sylvie van der Werf,Durivault JéRôMe |
| EPI_ISL_1201736 | WSSE Olsztyn | 1. National Institute of Public Health - National Institute of Hygiene; 2. Eurofins Genomics Europe Sequencing GmbH | Wokowicz Tomasz, Zacharczuk Katarzyna, Sadkowska-Todys Magorzata, Gierczyki Rafa, Eurofins Genomics Europe Sequencing Team, ECDC COVID-19 WGS support team |
| EPI_ISL_1201895 | Diagnosticos da America - DASA | Instituto Adolfo Lutz, Interdiciplinary Procedures Center, Strategic Laboratory | Claudio Tavares Sacchi, Claudia Regina Gonçalves, Erica Valessa Ramos Gomes, Karoline Rodrigues Campos, Caio Vinicius Dias Lopes |
| EPI_ISL_1208399 | Laboratoire de virologie clinique - Institut Pasteur de Tunis | 1-Laboratory of Microbiology, National Reference Lab, Charles Nicolle Hospital; 2-University of Tunis ElManar, Faculty of Medicine of Tunis, LR99ES09, Tunis, Tunisia | Sameh Trabelsi, Nissaf Ben Alaya, Mouna Ben Sassi, Sana Ferjani, Mouna Safer, Salma Abid, Fares Wasfi, Mariem Gdoura, Sondes Haddad, Anissa Chouikha, Roua Ben Othman, Sara Chammam, Ines Mdini, Manel Ben Sassi, Imen Kacem, Maher Kharrat, Alia BenKahla, Jalila Ben Khelil, Riadh Daghfous, Riadh Gouider, Henda Triki, Ilhem Boutiba-Ben Boubaker. |
| EPI_ISL_1208400, EPI_ISL_1208401, EPI_ISL_1208402 | ACT Pathology | Schwessinger Lab | Ashley Jones, Benjamin Schwessinger, Robert Lanfear, Megan McDonald, Ming-Dao Chia, Kevin Murray, Robyn N Hall, Craig Kennedy, Karina Kennedy |
| EPI_ISL_1209356 | Institute for Water Quality and Resource Management, Technical University Vienna | Bergthaler laboratory, CeMM Research Center for Molecular Medicine of the Austrian Academy of Sciences | Lukas Endler, Anna Schedl, Fabian Amman, Thomas Penz, Benedikt Agerer, Maelle Le Moing, Michael Schuster, Bekir Erguner, Jan Laine, Martin Senekowitsch, Christoph Bock, Andreas Bergthaler |
| EPI_ISL_1209407 | Laboratory for HIV and opportunistic infections diagnosis The Republican Research and Practical Center for Epidemiology and Microbiology (RRPCEM) | Laboratory for HIV and opportunistic infections diagnosis The Republican Research and Practical Center for Epidemiology and Microbiology (RRPCEM) | Elena Gasich, Kirill Bulda, Artur Akhremchuk, Leonid Valentovich, Anatoly Krasko, Vladimir Gorbunov |
| EPI_ISL_1209423 | Division of Emerging Infectious Diseases, Bureau of Infectious Diseases Diagnosis Control, Korea Disease Control and Prevention Agency | Division of Emerging Infectious Diseases, Bureau of Infectious Diseases Diagnosis Control, Korea Disease Control and Prevention Agency | Ae Kyung Park, Il-Hwan Kim, Heui Man Kim, Jeong-Min Kim, Chae Young Lee, Jin Sun No, Namjoo Lee, Sang Hee Woo, Eun-Jin Kim |
| EPI_ISL_1210050, EPI_ISL_1210291 | Israel Central Virology laboratory | Israel National Consortium for SARS-CoV-2 sequencing | Neta Zuckerman, Efrat Dahan Bucris, Michal Mandelboim, Dana Bar-Ilan, Oran Erster, Tzvia Mann, Omer Murik, David A. Zeevi, Assaf Rokney, Joseph Jaffe, Eva Nachum, Maya Davidovich Cohen, Ephraim Fass, Gal Zizelski Valenci, Mor Rubinstein, Efrat Rorman, Israel Nissan, Efrat Glick-Saar, Omri Nayshool, Gideon Rechavi, Ella Mendelson, Orna Mor |
| EPI_ISL_1213201, EPI_ISL_1213207, EPI_ISL_1213369 | LBM/UFPB | Bioinformatics Laboratory / LNCC | Alessandra P Lamarca, Luiz G P de Almeida, Ronaldo da Silva Francisco Jr, Lucymara Fassarella Agnez Lima, Kátia Castanho Scortecci, Vinícius Pietta Perez, Otavio J. Brustolini, Eduardo Sérgio Soares Sousa, Danielle Angst Secco, Angela Maria Guimarães Santos, George Rego Albuquerque, Ana Paula Melo Mariano, Bianca Mendes Maciel, Alexandra L Gerber, Ana Paula de C Guimarães, Paulo Ricardo Nascimento, Francisco Paulo Freire Neto, Sandra Rocha Gadelha, Luís Cristóvão Porto, Eloiza Helena Campana, Selma Maria Bezerra Jeronimo, Ana Tereza R Vasconcelos |
| EPI_ISL_1213425 | Laboratório HLA/UERJ | Bioinformatics Laboratory / LNCC | Alessandra P Lamarca, Luiz G P de Almeida, Ronaldo da Silva Francisco Jr, Lucymara Fassarella Agnez Lima, Kátia Castanho Scortecci, Vinícius Pietta Perez, Otavio J. Brustolini, Eduardo Sérgio Soares Sousa, Danielle Angst Secco, Angela Maria Guimarães Santos, George Rego Albuquerque, Ana Paula Melo Mariano, Bianca Mendes Maciel, Alexandra L Gerber, Ana Paula de C Guimarães, Paulo Ricardo Nascimento, Francisco Paulo Freire Neto, Sandra Rocha Gadelha, Luís Cristóvão Porto, Eloiza Helena Campana, Selma Maria Bezerra Jeronimo, Ana Tereza R Vasconcelos |
| EPI_ISL_1213504, EPI_ISL_1213509, EPI_ISL_1213524 | Bohol Containerized PCR Laboratory | Philippine Genome Center | Francis A. Tablizo, Kenneth M. Kim, Carlo M. Lapid, Marc Jerrone R. Castro, Maria Sofia L. Yangzon, Benedict A. Maralit, Marc Edsel C. Ayes, Eva Maria Cutiongco-de la Paz, Alethea R. de Guzman, Jan Michael C. Yap, Jo-Hannah S. Llames, Sheila Mae M. Araiza, Kris P. Punayan, Irish Coleen A. Asin, Candice Francheska B. Tambaoan, Asia Louisa U. Chong, Karol Sophia Agape R. Padilla, Rianna Patricia S. Cruz, El King D. Morado, Joshua Gregor A. Dizon, Razel Nikka M. Hao, Arianne A. Zamora, Devon Ray Pacial, Juan Antonio R. Magalang, Marissa Alejandria, Celia Carlos, Anna Ong-Lim, Edsel Maurice Salvaña, John Q. Wong, Jaime C. Montoya, Maria Rosario Singh-Vergeire and Cynthia P. Saloma |
| EPI_ISL_1213547 | Cebu TB Reference Laboratory | Philippine Genome Center | Francis A. Tablizo, Kenneth M. Kim, Carlo M. Lapid, Marc Jerrone R. Castro, Maria Sofia L. Yangzon, Benedict A. Maralit, Marc Edsel C. Ayes, Eva Maria Cutiongco-de la Paz, Alethea R. de Guzman, Jan Michael C. Yap, Jo-Hannah S. Llames, Sheila Mae M. Araiza, Kris P. Punayan, Irish Coleen A. Asin, Candice Francheska B. Tambaoan, Asia Louisa U. Chong, Karol Sophia Agape R. Padilla, Rianna Patricia S. Cruz, El King D. Morado, Joshua Gregor A. Dizon, Razel Nikka M. Hao, Arianne A. Zamora, Devon Ray Pacial, Juan Antonio R. Magalang, Marissa Alejandria, Celia Carlos, Anna Ong-Lim, Edsel Maurice Salvaña, John Q. Wong, Jaime C. Montoya, Maria Rosario Singh-Vergeire and Cynthia P. Saloma |
| EPI_ISL_1213571 | Negros Oriental Provincial Hospital | Philippine Genome Center | Francis A. Tablizo, Kenneth M. Kim, Carlo M. Lapid, Marc Jerrone R. Castro, Maria Sofia L. Yangzon, Benedict A. Maralit, Marc Edsel C. Ayes, Eva Maria Cutiongco-de la Paz, Alethea R. de Guzman, Jan Michael C. Yap, Jo-Hannah S. Llames, Sheila Mae M. Araiza, Kris P. Punayan, Irish Coleen A. Asin, Candice Francheska B. Tambaoan, Asia Louisa U. Chong, Karol Sophia Agape R. Padilla, Rianna Patricia S. Cruz, El King D. Morado, Joshua Gregor A. Dizon, Razel Nikka M. Hao, Arianne A. Zamora, Devon Ray Pacial, Juan Antonio R. Magalang, Marissa Alejandria, Celia Carlos, Anna Ong-Lim, Edsel Maurice Salvaña, John Q. Wong, Jaime C. Montoya, Maria Rosario Singh-Vergeire and Cynthia P. Saloma |
| EPI_ISL_1214444 | SARS-CoV-2 Sequencing Castilla y Leon-Spain Consortium | SARS-CoV-2 Sequencing Castilla y Leon-Spain Consortium | Antonio Orduña-Domingo, Marta Hernandez, David Abad, Marta Dominguez-Gil, Silvia Rojo, Laura Sánchez de Prada, Sonsoles Garcinuño Pérez, Carmen Aldea-Mansilla, Mª Fe Brezmes-Valdivieso, Gregoria Megías Lobón, María Antonia García Castro, Carmen Gimeno Crespo, Noelia Arenal Andrés, Carlos Fuster Foz, M. Isabel Fernandez-Natal, Jose María Eiros Bouza |
| EPI_ISL_1216080 | Bioscientia Labor Wermsdorf | Robert Koch Institute | unknown |
| EPI_ISL_1219432, EPI_ISL_1219454 | BIOR | Latvian Biomedical Research and Study Centre | Janis Pjalkovskis, Nikita Zrelovs, Monta Ustinova, Ivars Silamikelis, Liga Birzniece, Kaspars Megnis, Una Krumina, Guntars Zarins, Vita Rovite, Lauma Freimane, Laila Silamikele, Laura Ansone, Davids Fridmanis, Irena Meistere, Daina Pule, Jurijs Perevoscikovs, Uga Dumpis, Janis Klovins |
| EPI_ISL_1219803 | South Dakota Public Health Laboratory | South Dakota Public Health Laboratory | Jacob Garfin and Chris Carlson |
| EPI_ISL_1219949 | Labo Analyses Med | National Reference Center for Viruses of Respiratory Infections, Institut Pasteur, Paris | Marion Barbet, Sylvie Behillil, Méline Bizard, Angela Brisebarre, Camille Capel, Etienne Simon-Lorière, Vincent Enouf, Maud Vanpeene, Sylvie van der Werf,Durivault JéRôMe |
| EPI_ISL_1219950 | Labo Analyses Med | National Reference Center for Viruses of Respiratory Infections, Institut Pasteur, Paris | Marion Barbet, Sylvie Behillil, Méline Bizard, Angela Brisebarre, Camille Capel, Etienne Simon-Lorière, Vincent Enouf, Maud Vanpeene, Sylvie van der Werf,Ebel Anne |
| EPI_ISL_1219960 | Labo Analyses Med | National Reference Center for Viruses of Respiratory Infections, Institut Pasteur, Paris | Marion Barbet, Sylvie Behillil, Méline Bizard, Angela Brisebarre, Camille Capel, Etienne Simon-Lorière, Vincent Enouf, Maud Vanpeene, Sylvie van der Werf,Oliel Jonathan |
| EPI_ISL_1219962, EPI_ISL_1219963 | Labo Analyses Med | National Reference Center for Viruses of Respiratory Infections, Institut Pasteur, Paris | Marion Barbet, Sylvie Behillil, Méline Bizard, Angela Brisebarre, Camille Capel, Etienne Simon-Lorière, Vincent Enouf, Maud Vanpeene, Sylvie van der Werf,Durivault JéRôMe |
| EPI_ISL_1220047 | Hospital Universitario de San Rafael de Tunja | Instituto Nacional de Salud- Dirección de Investigación en Salud Pública | Katherine Laiton-Donato, Carlos Franco-Muñoz, Diego A. Álvarez-Díaz, Hector Alejandro Ruiz-Moreno, Jhonnatan Reales-González, Diego Andrés Prada, Sheryll Corchuelo, Maria T. Herrera-Sepúlveda, Julian Naizaque, Gerardo Santamaría, Magdalena Wiesner, Martha Lucia Ospina Martínez, Marcela Mercado-Reyes. |
| EPI_ISL_1220368, EPI_ISL_1220638 | Quest Diagnostics Incorporated | Centers for Disease Control and Prevention Division of Viral Diseases, Pathogen Discovery | Peter W. Cook, Dakota Howard, Dhwani Batra, Ben L. Rambo-Martin, S. H. Rosenthal, A. Gerasimova, R. M. Kagan, B. Anderson, M. Hua, Y. Liu, L.E. Bernstein, K.E. Livingston, A. Perez, I. A. Shlyakhter, R. V. Rolando, R. Owen, P. Tanpaiboon, F. Lacbawan, Clinton R. Paden, Suxiang Tong, Duncan MacCannell |
| EPI_ISL_1222766 | Central Scientific Research Department, Gomel State Medical University, | Laboratory of Genomics and Bioinformatics of the Forest Research Institute of the NAS of Belarus | Stoma,I.O., Baranov,O.Y., Voropaev,E.V., Osipkina,O.V.,Zyatskov,A.A., Shaforost,A.S., Padutov,V.E., Panteleev,S.V., Kiryanov,P.S. and Mozharovskaya,L.V. |
| EPI_ISL_1225330, EPI_ISL_1225393, EPI_ISL_1225453, EPI_ISL_1225466, EPI_ISL_1225483, EPI_ISL_1225522, EPI_ISL_1225531 | Gorgas Memorial Laboratory of Health Studies | Gorgas Memorial Laboratory of Health Studies | Yamilka Diaz, Anyuri Ortiz, Adriana Weeden, Daniel Castillo, Claudia Gonzalez, Brechla Moreno, Mabel Martinez-Montero, Marlene Castillo, Gretel Vasquez, Lisseth Saenz, Danilo Franco, Yaneth Pitti, Oris Chavarria, Jessica Gondola, Ambar Moreno, Layda Abrego, Davis Beltran, Ilka Guerra, Jim Chang, Zumara Chaverra, Isela Guerrero, Alejandra Valoy, Melissa Gaitan, Dimelza Arauz, Maria Chen-German, Elimelec Valdespino, Rita Rodriguez, Rita Corrales, Juan Miguel Pascale, Alexander Martinez, Sandra Lopez-Verges |

| | | | |
|---|---|---|---|
| EPI_ISL_1225875 | "AK State Public Health Lab, State Health Department" | Genomics and Discovery, Respiratory Viruses Branch, Division of Viral Diseases, Centers for Disease Control and Prevention | Krista Queen, Yan Li, Ying Tao, Jing Zhang, Anna Uehara, Anna Montmayeur, Clinton R. Paden, Kristen Knipe, Matthew Schmerer, Shoshona Le, Katie Dillon, Peter W. Cook, Rachel Marine, Mili Sheth, Jasmine Padilla, Sarah Nobles, Mark Burroughs, Lori Rowe, Haibin Wang, Ben L. Rambo-Martin, Kristine Lacek, Sam Shepard, Dhwani Batra, Suxiang Tong, Justin Lee |
| EPI_ISL_1227690 | NYC Pandemic Response Lab | Wadsworth Center, New York State Department of Health | Kirsten St. George, Daryl M. Lamson, Alexis Russel, Matthew Shudt, Melissa A Leisner, Jonathan Plitnick, Navjot Singh, John Kelly, Erasmus Schneider, Erica Lasek-Nesselquist |
| EPI_ISL_1228612 | Division of Emerging Infectious Diseases, Bureau of Infectious Diseases Diagnosis Control, Korea Disease Control and Prevention Agency | Division of Emerging Infectious Diseases, Bureau of Infectious Diseases Diagnosis Control, Korea Disease Control and Prevention Agency | Ae Kyung Park, Il-Hwan Kim, Heui Man Kim, Jeong-Min Kim, Jeong-Ah Kimg, Chae Young Lee, Jin Sun No, Eun-Jin Kim |
| EPI_ISL_1229164 | National Public Health Laboratory, National Centre for Infectious Diseases | National Public Health Laboratory, National Centre for Infectious Diseases | Tze Minn Mak, Zhenyang Zhou, Grace Jie Yin Ngan, Royce Ang, Lin Cui, Raymond Tzer Pin Lin |
| EPI_ISL_1229410, EPI_ISL_1229411 | Hematology Laboratory, Section of Molecular Diagnostics, University Clinical Centre, Medical University of Gdansk | Laboratory of Recombinant Vaccines | Lukasz Rabalski, Maciej Kosinski, Maciej Grzybek, Adam Sodol, Aneta Szulc, Krzysztof Lewandowski, Ewa Milosz, Marlena Robakowska, Boguslaw Szewczyk, Krystyna Bienkowska-Szewczyk |
| EPI_ISL_1229416, EPI_ISL_1229471 | Ministry of Health Turkey | Ministry of Health Turkey | Fatma Bayrakdar, Yasemin Cosgun, Suleyman Yalcin, Gulay Korukluoglu |
| EPI_ISL_1231593, EPI_ISL_1231750 | National Center for Infectious and Parasitic Diseases (NCIPD) | National Center for Infectious and Parasitic Diseases (NCIPD) | Alexiev et al |
| EPI_ISL_1232245, EPI_ISL_1232280, EPI_ISL_1232313, EPI_ISL_1232314, EPI_ISL_1232318, EPI_ISL_1232327, EPI_ISL_1232537, EPI_ISL_1232576, EPI_ISL_1232583, EPI_ISL_1232625 | Dutch COVID-19 response team | National Institute for Public Health and the Environment (RIVM) | Adam Meijer, Harry Vennema, Dirk Eggink, Jeroen Cremer, Sharon van den Brink, Bas van der Veer, AnneMarie van den Brandt, Florian Zwagemaker, Dennis Schmitz, Chantal Reusken, on behalf of the national COVID-19 response team |
| EPI_ISL_1233032 | North Dakota Department of Health, Public Health Laboratory | North Dakota Department of Health, Public Health Laboratory | Lisa Wingerter |
| EPI_ISL_1233064, EPI_ISL_1233094, EPI_ISL_1233096, EPI_ISL_1233107, EPI_ISL_1233133 | Centre for Dengue Research and AICBU, Department of Immunology and Molecular Medicine | Centre for Dengue Research and AICBU, Department of Immunology and Molecular Medicine | Chandima Jeewandara, Deshni Jayathilaka, Dinuka Ariyaratne, Tibutius Thanesh Pramanayagam, Diyanath Ranasinghe, Laksiri Gomes, Gathsaurie Neelika Malavige |
| EPI_ISL_1233500 | National Center for Infectious and Parasitic Diseases (NCIPD) | National Center for Infectious and Parasitic Diseases (NCIPD) | Alexiev et al |
| EPI_ISL_1233663, EPI_ISL_1233667 | Labormedizinisches Zentrum Dr Risch | University Hospital Basel, Clinical Bacteriology | Tim Roloff, Madlen Stange, Helena MB Seth-Smith, Alfredo Mari, Karoline Leuzinger, Julia Bielicki, Nadia Wohlwend,Martin Risch, Lorenz Risch, Manuel Battegay, Hans Hirsch, Adrian Egli |
| EPI_ISL_1233829 | Institute of Molecular and Translational Medicine / Laboratory of Experimental Medicine, Faculty of Medicine and Dentistry, Palacky University | Institute of Molecular and Translational Medicine / Laboratory of Experimental Medicine | Rastislav Slavkovský, Hana Jaworek, Vladimíra Koudeláková, Marián Hajdúch |
| EPI_ISL_1233878 | Department of Laboratory Medicine, Clinical Center, National Institutes of Health | Laboratory of Parasitic Diseases, Systems Genomics Section, National Institute of Allergy and Infectious Diseases, National Institutes of Health | Allison Roder, Stephanie Banakis, Matthew Chung, Jung-ho Youn, Rachel Mercado, Wei Wang, Tara Palmore, Michael Bell, Heike Bailin, Jessica McCormick-Ell, Sanchita Das, Jennifer Kwan, Elodie Ghedin |
| EPI_ISL_1234530 | MRCG at LSHTM Genomics lab | MRCG at LSHTM Genomics lab | Abdul Karim sesay, Abdoulie Kanteh, Jarra Manneh, Mariama Kujabi, Bakary Sanyang |
| EPI_ISL_1235859, EPI_ISL_1236768, EPI_ISL_1237932 | Houston Methodist Hospital | Houston Methodist Hospital | S. Wesley Long, Randall J. Olsen, Paul A. Christensen, Sishir Subedi, Robert Olson, James J. Davis, Matthew Ojeda Saavedra, Prasanti Yerramilli, Layne Pruitt, Kristina Reppond, Madison N. Shyer, Jessica Cambric, Ilya J. Finkelstein, Jimmy Gollihar, and James M. Musser |
| EPI_ISL_1238775, EPI_ISL_1238782 | North Dakota Department of Health, Public Health Laboratory | North Dakota Department of Health, Public Health Laboratory | Lisa Wingerter |
| EPI_ISL_1239115, EPI_ISL_1239133 | Laboratório Central de Saúde Pública do Espírito Santo | Coordenação Geral de Laboratórios de Saúde Pública (CGLAB) | Vagner Fonseca et al, |
| EPI_ISL_1239262 | Kimberley Hospital, National Health Laboratory Services, Northern Cape, South Africa | National Institute for Communicable Diseases of the National Health Laboratory Service | Amoako DG, Scheepers C, Mohale T, Ntuli N, Mahlangu B, Ismail A, Bhiman JN |
| EPI_ISL_1240071 | Clinical Center, University of Sarajevo; Unit for Clinical Microbiology | Clinical Center, University of Sarajevo; Unit for Clinical Microbiology | Irma Salimovi-Beši, Amela Dedei-Ljubovi, Edina Zahirovi, Suzana Arapi, Sebija Izetbegovi, Sandra Vegar-Zubovi |
| EPI_ISL_1240296 | National Laboratory for Health, Environment and Food, OMM, Celje | CISLD (Clinical Institute of Special Laboratory Diagnostics), University Children's Hospital, University Medical Center Ljubljana | Jernej Kova, Barbara Jenko Bizjan, Tine Tesovnik, Robert Šket, Katarina Kozmos, Ana Grom, Maruša Debeljak, Marko Pokorn, Tadej Battelino |
| EPI_ISL_1240648 | Israel Central Virology Laboratory | Israel National Consortium for SARS-CoV-2 sequencing | Neta Zuckerman, Efrat Dahan Bucris, Michal Mandelboim, Dana Bar-Ilan, Oran Erster, Tzvia Mann, Omer Murik, David A. Zeevi, Assaf Rokney, Joseph Jaffe, Eva Nachum, Maya Davidovich Cohen, Ephraim Fass, Gal Zizelski Valenci, Mor Rubinstein, Efrat Rorman, Israel Nissan, Efrat Glick-Saar, Omri Nayshool, Gideon Rechavi, Ella Mendelson, Orna Mor |
| EPI_ISL_1240948, EPI_ISL_1240964 | Department of Virology and Immunology, University of Helsinki and Helsinki University Hospital, Huslab Finland | Department of Virology, Faculty of Medicine, University of Helsinki, Helsinki, Finland | Teemu Smura, Ravi Kant, Phuoc Truong, Hussein Alburkat, Mert Erdin, Fathiah Zakham, Hannimari Kallio-Kokko, Jenni Virtanen, Maija Suvanto, Essi Korhonen, Sari Hannula, Harri Kangas, Hanna Liimatainen, Satu Kurkela, Hanna Jarva, Maija Lappalainen, Pekka Ellonen, Olli Vapalahti |
| EPI_ISL_1240991 | Area of Virology, Serology and Virology Division (SAViD), New South Wales Health Pathology Randwick | Area of Virology, Serology and Virology Division (SAViD), New South Wales Health Pathology Randwick | Foster, C.; Au, J.; Ruiz Silva, M.; Deveson, I.; Bull, R.; Van Hal, S.; Rawlinson, W. |
| EPI_ISL_1241735 | SYNLAB | GIGA Medical Genomics | Keith Durkin, Maria Artesi, Sébastien Bontems, Raphaël Boreux, Bouchra Boujemla, Nathalie Renotte, Cécile Meex, Pierrette Melin, Marie-Pierre Hayette, Vincent Bours |
| EPI_ISL_1241840 | PHV-FSS | PHV-FSS | Son Nguyen |
| EPI_ISL_1242014, EPI_ISL_1242016, EPI_ISL_1242017 | Nigeria Centre for Disease Control (NCDC) | African Centre of Excellence for Genomics of Infectious Diseases (ACEGID), Redeemer's University | Oluniyi P.E. et al |
| EPI_ISL_1249993 | Victorian Infectious Diseases Reference Laboratory (VIDRL) | VIDRL and MDU-PHL | Caly L., Seemann T., Sait, M.L., Druce J., Sherry, N.L. |
| EPI_ISL_1249999, EPI_ISL_1250002, EPI_ISL_1250005 | Microbiological Diagnostic Unit - Public Health Laboratory (MDU-PHL) | MDU-PHL | Seemann T., Sait, M.L., Sherry, N.L. |
| EPI_ISL_1250695 | Wellington SCL (WN) | Institute of Environmental Science and Research (ESR) | Rachel Boyle, SallyAnn Harbison, Olivia Stroeven, Xiaoyun Ren, Matt Storey, Nikki Freed, Muhammad Faisal, Jing Wang, Hermes Perez, Anja Werno, Antje van der Linden, Arlo Upton, Chris Mansell, David Hammer, Dragana Drinkovic, Gary McAuliffe, Hana Sofia Andersson, James Ussher, Jill Sherwood, Josh Freeman, Julia Howard, Juliet Elvy, Mary DeAlmeida, Matt Blakiston, Matthew Rogers, Max Bloomfield, Michael Addidle, Michelle Balm, Sally Roberts, Sarah Jefferies, Sharmini Muttaiyah, Susan Morpeth, Susan Taylor, Timothy Blackmore, Vani Sathyendran, Veronica Playle, Virginia Hope, Erasmus Smit, Lauren Jelly, Olin Silander, Joep de Ligt |
| EPI_ISL_1250706 | Middlemore Hospital | Institute of Environmental Science and Research (ESR) | Rachel Boyle, SallyAnn Harbison, Olivia Stroeven, Xiaoyun Ren, Matt Storey, Nikki Freed, Muhammad Faisal, Jing Wang, Hermes Perez, Anja Werno, Antje van der Linden, Arlo Upton, Chris Mansell, David Hammer, Dragana Drinkovic, Gary McAuliffe, Hana Sofia Andersson, James Ussher, Jill Sherwood, Josh Freeman, Julia Howard, Juliet Elvy, Mary DeAlmeida, Matt Blakiston, Matthew Rogers, Max Bloomfield, Michael Addidle, Michelle Balm, Sally Roberts, Sarah Jefferies, Sharmini Muttaiyah, Susan Morpeth, Susan Taylor, Timothy Blackmore, Vani Sathyendran, Veronica Playle, Virginia Hope, Erasmus Smit, Lauren Jelly, Olin Silander, Joep de Ligt |
| EPI_ISL_1250842 | Clin & Gen Lab | Molecular Genetics Laboratory, Instituto de Investigaciones Químicas, Universidad Mayor de San Andrés | Oscar M. Rollano-Peñaloza, Aneth Vasquez Michel |

| | | | |
|---|---|---|---|
| EPI_ISL_1251435 | Nordland Hospital - Bodo, Laboratory Department, Molecular Biology Unit | Norwegian Institute of Public Health, Department of Virology | Kathrine Stene-Johansen, Kamilla Heddeland Instefjord, Hilde Elshaug, Garcia Llorente Ignacio, Jon Bråte, Engebretsen Serina Beate,Pedersen Benedikte Nevjen, Debech Nadia, Atiya R Ali,Marie Paulsen Madsen, Rasmus Riis Kopperud, Hilde Vollan, Karoline Bragstad, Olav Hungnes |
| EPI_ISL_1252451 | National Public Health Laboratory, National Centre for Infectious Diseases | National Public Health Laboratory, National Centre for Infectious Diseases | Tze Minn Mak, Zhenyang Zhou, Grace Jie Yin Ngan, Royce Ang, Lin Cui, Raymond Tzer Pin Lin |
| EPI_ISL_1253694 | South Dakota Public Health Laboratory | South Dakota Public Health Laboratory | Jacob Garfin and Chris Carlson |
| EPI_ISL_1255105 | West African Centre for Cell Biology of Infectious Pathogens (WACCBIP), University of Ghana, Volta Road, Legon-Accra, Ghana | West African Centre for Cell Biology of Infectious Pathogens (WACCBIP), University of Ghana, Volta Road, Legon-Accra, Ghana | Collins M. Morang'a, Joyce M. Ngoi, Evelyn B. Quansah, Samirah Saiid, Dominic S.Y. Amuzu, Vincent Appiah, Philip M. Soglo, Vanessa Magnussen, Aisha Mohammed, Kesego Tapela, Nelson Kibinge, Abdoulaye B Diallo, Frederick Kumi-Ansah, Theophilus Odoom, Oliver D Boakye5, Emmanuella Amoako4, Abdul-Karim Abass, , Samuel Kaba Akoriyea, Frederick Tei-Maya, Lucas N. Amenga-Etego, Dam Kenneth Mibut, Yaw Bediako, Benjamin Demah Nuertey, Gordon A Awandare, Peter K Quashie, Gordon A Awandare, Yaw Bediako |
| EPI_ISL_1255109, EPI_ISL_1255179, EPI_ISL_1255215, EPI_ISL_1255229, EPI_ISL_1255250, EPI_ISL_1255263, EPI_ISL_1255265, EPI_ISL_1255267, EPI_ISL_1255270, EPI_ISL_1255273, EPI_ISL_1255274, EPI_ISL_1255277 | | | |
| see above | West African Centre for Cell Biology of Infectious Pathogens (WACCBIP), University of Ghana, Accra, Ghana | West African Centre for Cell Biology of Infectious Pathogens (WACCBIP), University of Ghana, Volta Road, Legon-Accra, Ghana | Collins M. Morang'a, Joyce M. Ngoi, Evelyn B. Quansah, Samirah Saiid, Dominic S.Y. Amuzu, Vincent Appiah, Philip M. Soglo, Vanessa Magnussen, Aisha Mohammed, Kesego Tapela, Nelson Kibinge, Abdoulaye B Diallo, Frederick Kumi-Ansah, Theophilus Odoom, Oliver D Boakye5, Emmanuella Amoako4, Abdul-Karim Abass, , Samuel Kaba Akoriyea, Frederick Tei-Maya, Lucas N. Amenga-Etego, Dam Kenneth Mibut, Yaw Bediako, Benjamin Demah Nuertey, Gordon A Awandare, Peter K Quashie, Gordon A Awandare, Yaw Bediako |
| EPI_ISL_1257828 | RSUP Dr. Sardjito | National Institute of Health Research and Development | Vivi Setiawaty, Hana Apsari Pawestri, Subangkit, Kartika Dewi Puspa, Arie Ardiansyah Nugraha, Hartanti Dian Ikawati, Nelly Puspandari, Krisna Pangesti |
| EPI_ISL_1258314, EPI_ISL_1258316, EPI_ISL_1258370, EPI_ISL_1258375 | MB-Cadham Provincial laboratory | National Microbiology Laboratory (NML) | Anna Majer, Shari Tyson, Grace Seo, Philip Mabon, Elsie Grudeski, Rhiannon Huzarewich, Russell Mandes, Anneliese Landgraff, Jennifer Tanner, Natalie Knox, Morag Graham, Gary Van Domselaar, Paul Van Caeseele, Jared Bullard, David Alexander, Kerry Dust, Nathalie Bastien, Yan Li, Timothy Booth, Darian Hole, Madison Chapel, Kirsten Biggar, CanCOGeN's metadata curation team, Public Health Agency of Canada CanCOGeN team |
| EPI_ISL_1259042, EPI_ISL_1259072 | Synlab Eesti OÜ | 1. Laboratory of Communicable Diseases (Estonia); 2. Eurofins Genomics Europe Sequencing GmbH | Liidia Dotsenko et al. |
| EPI_ISL_1259296 | Hopital | National Reference Center for Viruses of Respiratory Infections, Institut Pasteur, Paris | Marion Barbet, Sylvie Behillil, Méline Bizard, Angela Brisebarre, Camille Capel, Etienne Simon-Lorière, Vincent Enouf, Maud Vanpeene, Sylvie van der Werf,Hermann CéCile |
| EPI_ISL_1260912 | ULSS 6 Euganea | Istituto Zooprofilattico Sperimentale delle Venezie | Adelaide Milani, Alessia Schivo, Annalisa Salviato, Erika Giorgia Quaranta, Luca Tassoni, Ambra Pastori, Edoardo Giussani, Alice Fusaro, Isabella Monne, Calogero Terregino, Antonia Ricci |
| EPI_ISL_1261268, EPI_ISL_1261339 | National Institute of Public Health | National Reference Laboratory for Influenza and Respiratory Viruses CZE | Helena Jirincova, Jaromira Vecerova, Timotej Suri, Dusan Trnka, Alexander Nagy |
| EPI_ISL_1261697 | LACEN - Laboratório Central de Saúde Pública do Ceará | Evandro Chagas Institute | Santos, M.C.; Silva, A.M.; Junior, W.D.C.; Barbagelata, L.S.; Ferreira, J.A.; Sousa, E.M.A.; da Silva, P.S.; Pinheiro, K.C.; L.C.; Sousa Junior, E.C. |
| EPI_ISL_1262175 | Helix/Illumina | Centers for Disease Control and Prevention Division of Viral Diseases, Pathogen Discovery | Peter W. Cook, Dakota Howard, Dhwani Batra, Ben L. Rambo-Martin, Eileen de Feo, Jan Antico, Christine Tran, Matthew Tolentino, Shannon Wickline, Kim Gietzen, Brad Sickler, Jingtao Liu, Eric Allen, Phil Febbo, Summer Galloway, Nicole L. Washington, Simon White, Geraint Levan, Kelly Schiabor Barrett, Elizabeth Cirulli, Alexandre Bolze, Ary Ascencio, Charlotte Rivera-Garcia, Ryan Cho, Jason Nguyen, Sherry Wang, Jimmy Ramirez, Tyler Cassens, Efren Sandoval, Magnus Isaksson, William Lee, David Becker, Marc Laurent, James Lu, Clinton R. Paden, Suxiang Tong, Duncan MacCannell |
| EPI_ISL_1262467 | GA Department of Public Health | GA Department of Public Health | Stacy Reeves, Jonathan Edwards, Cynthia Dixey, Tonia Parrott, Aliyah Fields, Taylor Smith |
| EPI_ISL_1262697 | Instituto Nacional de Medicina Genomica | Instituto Nacional de Medicina Genomica | Hidalgo-Miranda A, Mendoza-Vargas A, Reyes-Grajeda JP, Cedro-Tanda A, Alcaraz N, Gonzalez-Barrera D, Rangel-DeLeon D, Ramirez-Vega O, Herrera-Montalvo LA |
| EPI_ISL_1265371 | "Stefan S. Nicolau"Institute of Virology | "Stefan S. Nicolau"Institute of Virology | Anca Botezatu, Iulia Virginia Iancu, Coralia Bleotu, Mihaela Economescu, Laura Necula, Denisa Dragu, Lilia Matei, Ioana Pitica, Marinela Bostan, Mirela Mihaila, Laura Grecu, Camelia Sultana, Simona Ruta, Ana Iulia Neagu, Adriana Plesa, Saviana Nedeianu, Alina Nastasie, Cristina Mambet,Gabriela Anton, Carmen Cristina Diaconu |
| EPI_ISL_1265420 | 1. Wojewódzka Stacja Sanitarno - Epidemiologiczna w Katowicach (WSSE Katowice); 2. Wojewódzka Stacja Sanitarno - Epidemiologiczna w Kielcach (WSSE Kielce); 3. Wojewódzka Stacja Sanitarno - Epidemiologiczna w Szczecinie (WSSE Szczecin); 4. Wojewódzka Stacja Sanitarno - Epidemiologiczna w Lublinie (WSSE Lublin); 5. Wojewódzka Stacja Sanitarno - Epidemiologiczna w Gorzowie Wielkopolskim (WSSE Gorzów Wielkopolski); 6. Wojewódzka Stacja Sanitarno-Epidemiologiczna w Gdasku (WSSE Gdask); 7. Wojewódzka Stacja Sanitarno-Epidemiologiczna w Poznaniu (WSSE Pozna); 8. Wojewódzka Stacja Sanitarno-Epidemiologiczna w Opolu (WSSE Opole); 9. Wojewódzka Stacja Sanitarno-Epidemiologiczna we Wrocawiu (WSSE Wrocaw); 10. Wojewódzka Stacja Sanitarno-Epidemiologiczna w Krakowie (WSSE Kraków); 11. Wojewódzka Stacja Sanitarno-Epidemiologiczna w Olsztynie (WSSE Olsztyn); 12. Wojewódzka Stacja Sanitarno-Epidemiologiczna w Bydgoszczy (WSSE Bydgoszcz); 13. Wojewódzka Stacja Sanitarno-Epidemiologiczna w Rzeszowie (WSSE Rzeszów); 14. Wojewódzka Stacja Sanitarno-Epidemiologiczna w Biaymstoku (WSSE Biaystok); | 1. ViroGenetics - BSL3 Laboratory of Virology, Maopolska Centre of Biotechnology, Jagiellonian University; 2. Human Genome Variation Research Group, Malopolska Centre of Biotechnology, Jagiellonian University; | Klajmon,A., Mazur-Panasiuk,N., Szulc,P., Kowalski,M., Gromowski,T., Marszalek,K., Labaj,P.P., Branicki,W., Pyrc,K. |
| EPI_ISL_1265916 | Quest Diagnostics Incorporated | Centers for Disease Control and Prevention Division of Viral Diseases, Pathogen Discovery | Peter W. Cook, Dakota Howard, Dhwani Batra, Ben L. Rambo-Martin, S. H. Rosenthal, A. Gerasimova, R. M. Kagan, B. Anderson, M. Hua, Y. Liu, L.E. Bernstein, K.E. Livingston, A. Perez, I. A. Shlyakhter, R. V. Rolando, R. Owen, P. Tanpaiboon, F. Lacbawan, Clinton R. Paden, Suxiang Tong, Duncan MacCannell |
| EPI_ISL_1266540 | National Laboratory for Health, Environment and Food, OMM, Celje | CISLD (Clinical Institute of Special Laboratory Diagnostics), University Children's Hospital, University Medical Center Ljubljana | Jernej Kova, Barbara Jenko Bizjan, Tine Tesovnik, Robert Šket, Katarina Kozmos, Ana Grom, Maruša Debeljak, Marko Pokorn, Tadej Battelino |
| EPI_ISL_1267443 | Quest Diagnostics Incorporated | Centers for Disease Control and Prevention Division of Viral Diseases, Pathogen Discovery | Peter W. Cook, Dakota Howard, Dhwani Batra, Ben L. Rambo-Martin, S. H. Rosenthal, A. Gerasimova, R. M. Kagan, B. Anderson, M. Hua, Y. Liu, L.E. Bernstein, K.E. Livingston, A. Perez, I. A. Shlyakhter, R. V. Rolando, R. Owen, P. Tanpaiboon, F. Lacbawan, Clinton R. Paden, Suxiang Tong, Duncan MacCannell |
| EPI_ISL_1267524, EPI_ISL_1270037 | Helix/Illumina | Centers for Disease Control and Prevention Division of Viral Diseases, Pathogen Discovery | Peter W. Cook, Dakota Howard, Dhwani Batra, Ben L. Rambo-Martin, Eileen de Feo, Jan Antico, Christine Tran, Matthew Tolentino, Shannon Wickline, Kim Gietzen, Brad Sickler, Jingtao Liu, Eric Allen, Phil Febbo, Summer Galloway, Nicole L. Washington, Simon White, Geraint Levan, Kelly Schiabor Barrett, Elizabeth Cirulli, Alexandre Bolze, Ary Ascencio, Charlotte Rivera-Garcia, Ryan Cho, Jason Nguyen, Sherry Wang, Jimmy Ramirez, Tyler Cassens, Efren Sandoval, Magnus Isaksson, William Lee, David Becker, Marc Laurent, James Lu, Clinton R. Paden, Suxiang Tong, Duncan MacCannell |
| EPI_ISL_1272258 | NB-Hôpital Georges L. Dumont | National Microbiology Laboratory (NML) | Anna Majer, Shari Tyson, Grace Seo, Philip Mabon, Elsie Grudeski, Rhiannon Huzarewich, Russell Mandes, Anneliese Landgraff, Jennifer Tanner, Natalie Knox, Morag Graham, Gary Van Domselaar, Richard Garceau, Guillaume Desnoyers, Nathalie Bastien, Yan Li, Timothy Booth, Darian Hole, Madison Chapel, Kirsten Biggar, CanCOGeN's metadata curation team, Public Health Agency of Canada CanCOGeN team |
| EPI_ISL_1272357 | NS-QEII Health Sciences Centre | National Microbiology Laboratory (NML) | Anna Majer, Shari Tyson, Grace Seo, Philip Mabon, Elsie Grudeski, Rhiannon Huzarewich, Russell Mandes, Anneliese Landgraff, Jennifer Tanner, Natalie |

| | | | |
|---|---|---|---|
| | | | Knox, Morag Graham, Gary Van Domselaar, Todd Hatchette, Jason LeBlanc, Janice Pettipas, Dan Gaston, Nathalie Bastien, Yan Li, Timothy Booth, Darian Hole, Madison Chapel, Kirsten Biggar, CanCOGeN's metadata curation team, Public Health Agency of Canada CanCOGeN team |
| EPI_ISL_1272627, EPI_ISL_1272633 | MS Public Health Laboratory | Centers for Disease Control and Prevention Division of Viral Diseases, Pathogen Discovery | Krista Queen, Yan Li, Ying Tao, Jing Zhang, Anna Uehara, Anna Montmayeur, Clinton R. Paden, Kristen Knipe, Matthew Schmerer, Shoshona Le, Katie Dillon, Peter W. Cook, Rachel Marine, Mili Sheth, Jasmine Padilla, Sarah Nobles, Mark Burroughs, Lori Rowe, Haibin Wang, Ben L. Rambo-Martin, Kristine Lacek, Sam Shepard, Dhwani Batra, Suxiang Tong, Justin Lee |
| EPI_ISL_1272918 | PR Public Health Lab | Centers for Disease Control and Prevention Division of Viral Diseases, Pathogen Discovery | Krista Queen, Yan Li, Ying Tao, Jing Zhang, Anna Uehara, Anna Montmayeur, Clinton R. Paden, Kristen Knipe, Matthew Schmerer, Shoshona Le, Katie Dillon, Peter W. Cook, Rachel Marine, Mili Sheth, Jasmine Padilla, Sarah Nobles, Mark Burroughs, Lori Rowe, Haibin Wang, Ben L. Rambo-Martin, Kristine Lacek, Sam Shepard, Dhwani Batra, Suxiang Tong, Justin Lee |
| EPI_ISL_1273054, EPI_ISL_1273056, EPI_ISL_1273058, EPI_ISL_1273063, EPI_ISL_1273071, EPI_ISL_1273073 | Guam Public Health Laboratory | Centers for Disease Control and Prevention Division of Viral Diseases, Pathogen Discovery | Krista Queen, Yan Li, Ying Tao, Jing Zhang, Anna Uehara, Anna Montmayeur, Clinton R. Paden, Kristen Knipe, Matthew Schmerer, Shoshona Le, Katie Dillon, Peter W. Cook, Rachel Marine, Mili Sheth, Jasmine Padilla, Sarah Nobles, Mark Burroughs, Lori Rowe, Haibin Wang, Ben L. Rambo-Martin, Kristine Lacek, Sam Shepard, Dhwani Batra, Suxiang Tong, Justin Lee |
| EPI_ISL_1273102 | Faculty of Medicine, Al-Quds University | Faculty of Medicine, Al-Quds University | Ereqat,S., Al-Jawabreh,A., Dumaidi,K., Nasereddin,A. |
| EPI_ISL_1273214 | Oxford University Clinical Research Unit (OUCRU) | Oxford University Clinical Research Unit (OUCRU) | Nguyen Van Vinh Chau, Nguyen Thi Thu Hong, Nghiem My Ngoc, Nguyen To Anh, Huynh Trung Trieu, Le Nguyen Truc Nhu, Lam Minh Yen, Ngo Ngoc Quang Minh, Nguyen Thanh Phong, Nguyen Thanh Truong, le Thi Thu Huong, Tran Nguyen Hoang Tu, Le Manh Hung, Tran Tan Thanh, Nguyen Thanh Dung, Nguyen Tri Dung, Guy Thwaites, Le Van Tan |
| EPI_ISL_1273392, EPI_ISL_1273393, EPI_ISL_1273395, EPI_ISL_1273396, EPI_ISL_1273401, EPI_ISL_1273406, EPI_ISL_1273408 | National Reference Laboratory - Ministry of Health Maseru Lesotho | National Institute for Communicable Diseases of the National Health Laboratory Service | Gorova V, Mathabo M, Mooko M, Banda R, Amoako DG, Scheepers C, Mohale T, Ntuli N, Mahlangu B, Ismail A, Bhiman JN |
| EPI_ISL_1273720 | University Hospital Basel, Clinical Virology | University Hospital Basel, Clinical Bacteriology | Tim Roloff, Madlen Stange, Helena MB Seth-Smith, Alfredo Mari, Karoline Leuzinger, Julia Bielicki, Manuel Battegay, Hans Hirsch, Adrian Egli |
| EPI_ISL_1278275, EPI_ISL_1278276 | Clin & Gen Lab | Molecular Genetics Laboratory, Instituto de Investigaciones Químicas, Universidad Mayor de San Andrés | Oscar M. Rollano-Peñaloza, Aneth Vasquez Michel |
| EPI_ISL_1278277 | Laboratorio de Biologia Molecular, Hospital San Pedro Claver | Molecular Genetics Laboratory, Instituto de Investigaciones Químicas, Universidad Mayor de San Andrés | Oscar M. Rollano-Peñaloza, Carmen Delgado Barrera, Aneth Vasquez Michel |
| EPI_ISL_1278281, EPI_ISL_1278282, EPI_ISL_1278283, EPI_ISL_1278284 | Laboratorio de Biologia Molecular, SEDES-Potosi | Molecular Genetics Laboratory, Instituto de Investigaciones Químicas, Universidad Mayor de San Andrés | Oscar M. Rollano-Peñaloza, Aneth Vasquez Michel |
| EPI_ISL_1279265 | Unidad de Investigación Biomédica de Zacatecas (UIBZ) | Instituto Nacional de Enfermedades Respiratorias (INER): Centro de Investigación en Enfermedades Infecciosas (CIENI) | Consorcio Mexicano de Vigilancia Genómica (CoViGen-Mex). Authors (in alphabetical order): Julio Elias Alvarado-Yaah, Carlos F. Arias, Santiago Ávila-Ríos, Víctor Hugo Borja-Aburto, Celia Boukadida, Juan Bautista Chale-Dzul , José Antonio Enciso-Moreno, Gloria Elena Espinoza-Ayala, Fernando Fontove-Herrera, Concepción Grajales-Muñiz, Ricardo Grande, Alfredo Herrera-Estrella, Carla Ivón Herrera-Najera, Pavel Isa, Brenda Irasema Maldonado-Meza, Bernardo Martínez-Miguel, Margarita Matías-Florentino, María Guadalupe de Jesús Mireles-Rivera, Gloria María Molina-Salinas, Hector Montoya-Fuentes, José Esteban Muñoz-Medina, José de Jesús Nuñez-Contreras, Alicia Ocaña-Mondragón, Luis Alberto Ochoa-Carrera, Hector Esteban Paz-Juárez, Francisco Pulido, Helen Haydee Fernanda Ramírez-Plascencia, Angel Gustavo Salas-Lais, Jorge Ivan Salinal-Nevarez, Alejandro Sanchez-Flores, Clara Esperanza Santacruz-Tinoco, María Guadalupe Santiago-Mauricio , Nelly Sélem-Mojica, Blanca Taboada , Gloria Vazquez |
| EPI_ISL_1279266 | Centro de Investigación Biomédica del Noreste (CIBIN) | Instituto Nacional de Enfermedades Respiratorias (INER): Centro de Investigación en Enfermedades Infecciosas (CIENI) | Consorcio Mexicano de Vigilancia Genómica (CoViGen-Mex). Authors (in alphabetical order): Julio Elias Alvarado-Yaah, Carlos F. Arias, Santiago Ávila-Ríos, Víctor Hugo Borja-Aburto, Celia Boukadida, Juan Bautista Chale-Dzul , José Antonio Enciso-Moreno, Gloria Elena Espinoza-Ayala, Fernando Fontove-Herrera, Concepción Grajales-Muñiz, Ricardo Grande, Alfredo Herrera-Estrella, Carla Ivón Herrera-Najera, Pavel Isa, Brenda Irasema Maldonado-Meza, Bernardo Martínez-Miguel, Margarita Matías-Florentino, María Guadalupe de Jesús Mireles-Rivera, Gloria María Molina-Salinas, Hector Montoya-Fuentes, José Esteban Muñoz-Medina, José de Jesús Nuñez-Contreras, Alicia Ocaña-Mondragón, Luis Alberto Ochoa-Carrera, Hector Esteban Paz-Juárez, Francisco Pulido, Helen Haydee Fernanda Ramírez-Plascencia, Angel Gustavo Salas-Lais, Jorge Ivan Salinal-Nevarez, Alejandro Sanchez-Flores, Clara Esperanza Santacruz-Tinoco, María Guadalupe Santiago-Mauricio , Nelly Sélem-Mojica, Blanca Taboada , Gloria Vazquez |
| EPI_ISL_1279281 | Unidad de Investigación Biomédica de Zacatecas (UIBZ) | Instituto Nacional de Enfermedades Respiratorias (INER): Centro de Investigación en Enfermedades Infecciosas (CIENI) | Consorcio Mexicano de Vigilancia Genómica (CoViGen-Mex). Authors (in alphabetical order): Julio Elias Alvarado-Yaah, Carlos F. Arias, Santiago Ávila-Ríos, Víctor Hugo Borja-Aburto, Celia Boukadida, Juan Bautista Chale-Dzul , José Antonio Enciso-Moreno, Gloria Elena Espinoza-Ayala, Fernando Fontove-Herrera, Concepción Grajales-Muñiz, Ricardo Grande, Alfredo Herrera-Estrella, Carla Ivón Herrera-Najera, Pavel Isa, Brenda Irasema Maldonado-Meza, Bernardo Martínez-Miguel, Margarita Matías-Florentino, María Guadalupe de Jesús Mireles-Rivera, Gloria María Molina-Salinas, Hector Montoya-Fuentes, José Esteban Muñoz-Medina, José de Jesús Nuñez-Contreras, Alicia Ocaña-Mondragón, Luis Alberto Ochoa-Carrera, Hector Esteban Paz-Juárez, Francisco Pulido, Helen Haydee Fernanda Ramírez-Plascencia, Angel Gustavo Salas-Lais, Jorge Ivan Salinal-Nevarez, Alejandro Sanchez-Flores, Clara Esperanza Santacruz-Tinoco, María Guadalupe Santiago-Mauricio , Nelly Sélem-Mojica, Blanca Taboada , Gloria Vazquez |
| EPI_ISL_1279391 | Laboratorio Central de Epidemiología (LCE) | Instituto Nacional de Enfermedades Respiratorias (INER): Centro de Investigación en Enfermedades Infecciosas (CIENI) | Consorcio Mexicano de Vigilancia Genómica (CoViGen-Mex). Authors (in alphabetical order): Julio Elias Alvarado-Yaah, Carlos F. Arias, Santiago Ávila-Ríos, Víctor Hugo Borja-Aburto, Celia Boukadida, Juan Bautista Chale-Dzul , José Antonio Enciso-Moreno, Gloria Elena Espinoza-Ayala, Fernando Fontove-Herrera, Concepción Grajales-Muñiz, Ricardo Grande, Alfredo Herrera-Estrella, Carla Ivón Herrera-Najera, Pavel Isa, Brenda Irasema Maldonado-Meza, Bernardo Martínez-Miguel, Margarita Matías-Florentino, María Guadalupe de Jesús Mireles-Rivera, Gloria María Molina-Salinas, Hector Montoya-Fuentes, José Esteban Muñoz-Medina, José de Jesús Nuñez-Contreras, Alicia Ocaña-Mondragón, Luis Alberto Ochoa-Carrera, Hector Esteban Paz-Juárez, Francisco Pulido, Helen Haydee Fernanda Ramírez-Plascencia, Angel Gustavo Salas-Lais, Jorge Ivan Salinal-Nevarez, Alejandro Sanchez-Flores, Clara Esperanza Santacruz-Tinoco, María Guadalupe Santiago-Mauricio , Nelly Sélem-Mojica, Blanca Taboada , Gloria Vazquez |
| EPI_ISL_1279479 | Centro de Investigación Biomédica del Noreste (CIBIN) | Instituto Nacional de Enfermedades Respiratorias (INER): Centro de Investigación en Enfermedades Infecciosas (CIENI) | Consorcio Mexicano de Vigilancia Genómica (CoViGen-Mex). Authors (in alphabetical order): Julio Elias Alvarado-Yaah, Carlos F. Arias, Santiago Ávila-Ríos, Víctor Hugo Borja-Aburto, Celia Boukadida, Juan Bautista Chale-Dzul , José Antonio Enciso-Moreno, Gloria Elena Espinoza-Ayala, Fernando Fontove-Herrera, Concepción Grajales-Muñiz, Ricardo Grande, Alfredo Herrera-Estrella, Carla Ivón Herrera-Najera, Pavel Isa, Brenda Irasema Maldonado-Meza, Bernardo Martínez-Miguel, Margarita Matías-Florentino, María Guadalupe de Jesús Mireles-Rivera, Gloria María Molina-Salinas, Hector Montoya-Fuentes, José Esteban Muñoz-Medina, José de Jesús Nuñez-Contreras, Alicia Ocaña-Mondragón, Luis Alberto Ochoa-Carrera, Hector Esteban Paz-Juárez, Francisco Pulido, Helen Haydee Fernanda Ramírez-Plascencia, Angel Gustavo Salas-Lais, Jorge Ivan Salinal-Nevarez, Alejandro Sanchez-Flores, Clara Esperanza Santacruz-Tinoco, María Guadalupe Santiago-Mauricio , Nelly Sélem-Mojica, Blanca Taboada , Gloria Vazquez |
| EPI_ISL_1279540, EPI_ISL_1279556, EPI_ISL_1279578 | Laboratorio Central de Epidemiología (LCE) | Instituto Nacional de Enfermedades Respiratorias (INER): Centro de Investigación en Enfermedades Infecciosas (CIENI) | Consorcio Mexicano de Vigilancia Genómica (CoViGen-Mex). Authors (in alphabetical order): Julio Elias Alvarado-Yaah, Carlos F. Arias, Santiago Ávila-Ríos, Víctor Hugo Borja-Aburto, Celia Boukadida, Juan Bautista Chale-Dzul , José Antonio Enciso-Moreno, Gloria Elena Espinoza-Ayala, Fernando Fontove-Herrera, Concepción Grajales-Muñiz, Ricardo Grande, Alfredo Herrera-Estrella, Carla Ivón Herrera-Najera, Pavel Isa, Brenda Irasema Maldonado-Meza, Bernardo Martínez-Miguel, Margarita Matías-Florentino, María Guadalupe de Jesús Mireles-Rivera, Gloria María Molina-Salinas, Hector Montoya-Fuentes, José Esteban Muñoz-Medina, José de Jesús Nuñez-Contreras, Alicia Ocaña-Mondragón, Luis Alberto Ochoa-Carrera, Hector Esteban Paz-Juárez, Francisco Pulido, Helen Haydee Fernanda Ramírez-Plascencia, Angel Gustavo Salas-Lais, Jorge Ivan Salinal-Nevarez, Alejandro Sanchez-Flores, Clara Esperanza Santacruz-Tinoco, María Guadalupe Santiago-Mauricio , Nelly Sélem-Mojica, Blanca Taboada , Gloria Vazquez |
| EPI_ISL_1279604 | Unidad de Investigación Médica de Yucatán (UIMY) | Instituto Nacional de Enfermedades Respiratorias (INER): Centro de Investigación en Enfermedades Infecciosas (CIENI) | Consorcio Mexicano de Vigilancia Genómica (CoViGen-Mex). Authors (in alphabetical order): Julio Elias Alvarado-Yaah, Carlos F. Arias, Santiago Ávila-Ríos, Víctor Hugo Borja-Aburto, Celia Boukadida, Juan Bautista Chale-Dzul , José Antonio Enciso-Moreno, Gloria Elena Espinoza-Ayala, Fernando Fontove-Herrera, Concepción Grajales-Muñiz, Ricardo Grande, Alfredo Herrera-Estrella, Carla Ivón Herrera-Najera, Pavel Isa, Brenda Irasema Maldonado-Meza, Bernardo Martínez-Miguel, Margarita Matías-Florentino, María Guadalupe de Jesús Mireles-Rivera, Gloria María Molina-Salinas, Hector Montoya-Fuentes, José Esteban Muñoz-Medina, José de Jesús Nuñez-Contreras, Alicia Ocaña-Mondragón, Luis Alberto Ochoa-Carrera, Hector Esteban Paz-Juárez, Francisco Pulido, Helen Haydee Fernanda Ramírez-Plascencia, Angel Gustavo Salas-Lais, Jorge Ivan Salinal-Nevarez, Alejandro Sanchez-Flores, Clara Esperanza Santacruz-Tinoco, María Guadalupe Santiago-Mauricio , Nelly Sélem-Mojica, Blanca Taboada , Gloria Vazquez |
| EPI_ISL_1279959 | National Institute of Infectious Diseases-Prof. Dr. Matei Bals Molecular Diagnostics Laboratory | National Institute of Infectious Diseases-Prof. Dr. Matei Bals Molecular Diagnostics Laboratory | Leontina Banica, Marius Surleac, Corina Casangiu, Petre Milu, Andreea Tudor, Simona Paraschiv, Dan Otelea |
| EPI_ISL_1280104 | Department of Genetics, Medirex | Laboratory of Genomics and Bioinformatics, Comenius | Tatiana Sedláková, Miroslav Böhmer, Renáta Lukaková, Gabriel Minárik, Anna Giová, Werner Krampl, Diana Rusáková, Jaroslav Budiš, Tomáš Szemes |

| | | | |
|---|---|---|---|
| | | University Science Park | |
| EPI_ISL_1287349 | Botswana Harvard HIV Reference Laboratory | Botswana Harvard HIV Reference Laboratory | Sikhulile Moyo, Wonderful T. Choga, Dorcas Maruapula,Thongbotho Mphoyakgosi, Boitumelo Zuze, Botshelo Radibe, Legodile Kooepile, David Lawrence, Roger Shapiro, Shahin Lockman, Mosepele Mosepele, Joseph Makhema, Simani Gaseitsiwe |
| EPI_ISL_1287761, EPI_ISL_1287762, EPI_ISL_1287763, EPI_ISL_1287764, EPI_ISL_1287765, EPI_ISL_1287767 | Unidad de Investigación Biomédica de Zacatecas (UIBZ) | Instituto Nacional de Enfermedades Respiratorias (INER): Centro de Investigación en Enfermedades Infecciosas (CIENI) | Consorcio Mexicano de Vigilancia Genómica (CoViGen-Mex). Authors (in alphabetical order): Julio Elias Alvarado-Yaah, Carlos F. Arias, Santiago Ávila-Ríos, Víctor Hugo Borja-Aburto, Celia Boukadida, Juan Bautista Chale-Dzul , José Antonio Enciso-Moreno, Gloria Elena Espinoza-Ayala, Fernando Fontove-Herrera, Concepción Grajales-Muñiz, Ricardo Grande, Alfredo Herrera-Estrella, Carla Ivón Herrera-Najera, Pavel Isa, Brenda Irasema Maldonado-Meza, Bernardo Martínez-Miguel, Margarita Matías-Florentino, María Guadalupe de Jesús Mireles-Rivera, Gloria María Molina-Salinas, Hector Montoya-Fuentes, José Esteban Muñoz-Medina, José de Jesús Nuñez-Contreras, Alicia Ocaña-Mondragón, Luis Alberto Ochoa-Carrera, Hector Esteban Paz-Juárez, Francisco Pulido, Helen Haydee Fernanda Ramírez-Plascencia, Angel Gustavo Salas-Lais, Jorge Ivan Salinal-Nevarez, Alejandro Sanchez-Flores, Clara Esperanza Santacruz-Tinoco, María Guadalupe Santiago-Mauricio , Nelly Sélem-Mojica, Blanca Taboada , Gloria Vazquez |
| EPI_ISL_1288129, EPI_ISL_1288139, EPI_ISL_1288149 | Dutch COVID-19 response team | National Institute for Public Health and the Environment (RIVM) | Adam Meijer, Harry Vennema, Dirk Eggink, Jeroen Cremer, Sharon van den Brink, Bas van der Veer, AnneMarie van den Brandt, Florian Zwagemaker, Dennis Schmitz, Chantal Reusken, on behalf of the national COVID-19 response team |
| EPI_ISL_1288221, EPI_ISL_1288250, EPI_ISL_1288253, EPI_ISL_1288314, EPI_ISL_1288326, EPI_ISL_1288330, EPI_ISL_1288388, EPI_ISL_1288402, EPI_ISL_1288453, EPI_ISL_1288495 | Laboratorio Central de Epidemiología (LCE) | Instituto de Biotecnología de la UNAM | Consorcio Mexicano de Vigilancia Genómica (CoViGen-Mex). Authors (in alphabetical order): Julio Elias Alvarado-Yaah, Carlos F. Arias, Santiago Ávila-Ríos, Víctor Hugo Borja-Aburto, Celia Boukadida, Juan Bautista Chale-Dzul , José Antonio Enciso-Moreno, Gloria Elena Espinoza-Ayala, Fernando Fontove-Herrera, Concepción Grajales-Muñiz, Ricardo Grande, Alfredo Herrera-Estrella, Carla Ivón Herrera-Najera, Pavel Isa, Brenda Irasema Maldonado-Meza, Bernardo Martínez-Miguel, Margarita Matías-Florentino, María Guadalupe de Jesús Mireles-Rivera, Gloria María Molina-Salinas, Hector Montoya-Fuentes, José Esteban Muñoz-Medina, José de Jesús Nuñez-Contreras, Alicia Ocaña-Mondragón, Luis Alberto Ochoa-Carrera, Hector Esteban Paz-Juárez, Francisco Pulido, Helen Haydee Fernanda Ramírez-Plascencia, Angel Gustavo Salas-Lais, Jorge Ivan Salinal-Nevarez, Alejandro Sanchez-Flores, Clara Esperanza Santacruz-Tinoco, María Guadalupe Santiago-Mauricio , Nelly Sélem-Mojica, Blanca Taboada , Gloria Vazquez |
| EPI_ISL_1289388, EPI_ISL_1289401 | Dutch COVID-19 response team | National Institute for Public Health and the Environment (RIVM) | Adam Meijer, Harry Vennema, Dirk Eggink, Jeroen Cremer, Sharon van den Brink, Bas van der Veer, AnneMarie van den Brandt, Florian Zwagemaker, Dennis Schmitz, Chantal Reusken, on behalf of the national COVID-19 response team |
| EPI_ISL_1289562 | Michigan Department of Health and Human Services, Bureau of Laboratories | Michigan Department of Health and Human Services, Bureau of Laboratories | Blankenship HM, Riner D, Soehnlen MK |
| EPI_ISL_1289876, EPI_ISL_1289885, EPI_ISL_1289915, EPI_ISL_1289920, EPI_ISL_1289921, EPI_ISL_1289924 | Department of Public Health Microbiology Ljubljana, National Laboratory for Health, Environment and Food | Department of Public Health Microbiology Ljubljana, National Laboratory for Health, Environment and Food | José Gonçalves, Tom Koritnik, Martin Bosilj, Katarina Prosenc, Metka Paragi |
| EPI_ISL_1290557 | Swedish national genomic surveillance program of SARS-CoV-2 | The Public Health Agency of Sweden | Swedish national genomic surveillance program of SARS-CoV-2 |
| EPI_ISL_1291044, EPI_ISL_1291381, EPI_ISL_1291581 | Quest Diagnostics Incorporated | Centers for Disease Control and Prevention Division of Viral Diseases, Pathogen Discovery | Peter W. Cook, Dakota Howard, Dhwani Batra, Ben L. Rambo-Martin, S. H. Rosenthal, A. Gerasimova, R. M. Kagan, B. Anderson, M. Hua, Y. Liu, L.E. Bernstein, K.E. Livingston, A. Perez, I. A. Shlyakhter, R. V. Rolando, R. Owen, P. Tanpaiboon, F. Lacbawan, Clinton R. Paden, Suxiang Tong, Duncan MacCannell |
| EPI_ISL_1292548 | State Laboratories Division, Hawaii State Department of Health | State Laboratories Division, Hawaii State Department of Health | Pamela O'Brien, Drew Kuwazaki, Ayana Garnet, Razvan Sultana, Edward Desmond |
| EPI_ISL_1293036 | MD Public Health Laboratory | MD Public Health Laboratory | Maryland Department of Health Laboratories Administration |
| EPI_ISL_1293114 | Tempus | Grubaugh Lab - Yale School of Public Health | Joseph Fauver, Mallery Breban, Isabell Ott, Tara Alpert, Mary Petrone, Anderson Brito, Chantal Vogels, Annie Watkins, Chaney Kalinich, Jessica Rothman, Matthew J. MacKay, Gaurav Khullar, Jessica Metti, Joel T. Dudley, Megan Nash, Nike Beaubier, Christopher E. Mason, Nathan Grubaugh |
| EPI_ISL_1293350 | Oxford University Clinical Research Unit (OUCRU) | Oxford University Clinical Research Unit (OUCRU) | Nguyen Van Vinh Chau, Nguyen Thi Thu Hong, Nghiem My Ngoc, Nguyen To Anh, Huynh Trung Trieu, Le Nguyen Truc Nhu, Lam Minh Yen, Ngo Ngoc Quang Minh, Nguyen Thanh Phong, Nguyen Thanh Truong, Le Thi Thu Huong, Tran Nguyen Hoang Tu, Le Manh Hung, Tran Tan Thanh, Nguyen Thanh Dung, Nguyen Tri Dung, Guy Thwaites, le Van Tan |
| EPI_ISL_1295843 | Sharp HealthCare Laboratory | Andersen lab at Scripps Research | SEARCH Alliance San Diego with Aaron Harding, Jacquelyn Berumen, Cathy Woerle, Liam McGinnis, Art Mendoza, Omid Bakhtar |
| EPI_ISL_1295933, EPI_ISL_1295934 | PathWest Laboratory Medicine WA | PathWest Laboratory Medicine WA Microbial Surveillance Unit | PathWest Laboratory Medicine WA Microbial Surveillance Unit |
| EPI_ISL_1296992 | Sonora Quest Laboratories | TGen North | "Jolene Bowers, Heather Centner, Chris French, Hayley Yaglom, Ashlyn Pfeiffer, Darrin Lemmer, Dave Engelthaler, The Arizona COVID Genomics Union (ACGU)" |
| EPI_ISL_1297497 | Quest Diagnostics Incorporated | Centers for Disease Control and Prevention Division of Viral Diseases, Pathogen Discovery | Peter W. Cook, Dakota Howard, Dhwani Batra, Ben L. Rambo-Martin, S. H. Rosenthal, A. Gerasimova, R. M. Kagan, B. Anderson, M. Hua, Y. Liu, L.E. Bernstein, K.E. Livingston, A. Perez, I. A. Shlyakhter, R. V. Rolando, R. Owen, P. Tanpaiboon, F. Lacbawan, Clinton R. Paden, Suxiang Tong, Duncan MacCannell |
| EPI_ISL_1298458 | Instituto Nacional de Medicina Genomica | Instituto Nacional de Medicina Genomica | Hidalgo-Miranda A, Mendoza-Vargas A, Reyes-Grajeda JP, Cedro-Tanda A, Alcaraz N, Gonzalez-Barrera D, Rangel-DeLeon D, Ramirez-Vega O, Herrera-Montalvo LA |
| EPI_ISL_1298474 | Burshtyn CCH | The Institute of Molecular Biology and Genetics of NASU | M.Tukalo et al. |
| EPI_ISL_1298476, EPI_ISL_1298477 | Ivano-Frankivsk CCH#1 | The Institute of Molecular Biology and Genetics of NASU | M.Tukalo et al. |
| EPI_ISL_1298480 | Nadvirna CRH | The Institute of Molecular Biology and Genetics of NASU | M.Tukalo et al. |
| EPI_ISL_1298482 | Tysmenytsia CH | The Institute of Molecular Biology and Genetics of NASU | M.Tukalo et al. |
| EPI_ISL_1299860, EPI_ISL_1299865, EPI_ISL_1299866, EPI_ISL_1299868, EPI_ISL_1299871 | Unit of lab surveillance of viral emerging diseases, National Lab of Influenza | Respiratory Virus Unit, National Infection Service, Public Health England | PHE Covid Sequencing Team, Iris Hasibra, Prof Silvia Bino, Prof Albana Fico |
| EPI_ISL_1299913, EPI_ISL_1299916, EPI_ISL_1299917, EPI_ISL_1299918, EPI_ISL_1299920 | Labormedizinisches Zentrum Dr Risch | University Hospital Basel, Clinical Bacteriology | Tim Roloff, Madlen Stange, Helena MB Seth-Smith, Alfredo Mari, Karoline Leuzinger, Julia Bielicki, Nadia Wohlwend,Martin Risch, Lorenz Risch, Manuel Battegay, Hans Hirsch, Adrian Egli |
| EPI_ISL_1300452, EPI_ISL_1300474, EPI_ISL_1300481, EPI_ISL_1300486, EPI_ISL_1300501, EPI_ISL_1300508 | Genetica Molecular and Subdepartamento de Virologia ISP Chile | Instituto de Salud Publica de Chile | Javier Tognarelli, Karen Orostica, Barbara Parra, Loredana Arata, Jaime Lagos, Gisselle Barra, Patricia Bustos, Rodrigo Fasce, Andres Castillo, Jorge Fernandez |
| EPI_ISL_1300526, EPI_ISL_1300529, EPI_ISL_1300530 | Public Health Virology-Forensic and Scientific Services (PHV-FSS) | Public Health Virology-Forensic and Scientific Services (PHV-FSS) | Son Nguyen |
| EPI_ISL_1300531 | PHV-FSS | PHV-FSS | Son Nguyen |
| EPI_ISL_1300532, EPI_ISL_1300533 | Public Health Virology-Forensic and Scientific Services (PHV-FSS) | Public Health Virology-Forensic and Scientific Services (PHV-FSS) | Son Nguyen |
| EPI_ISL_1300647, EPI_ISL_1300649, EPI_ISL_1300650 | Clinical Center, University of Sarajevo; Unit for Clinical Microbiology | Clinical Center, University of Sarajevo; Unit for Clinical Microbiology | Irma Salimovi-Beši, Amela Dedei-Ljubovi, Edina Zahirovi, Suzana Arapi, Sebija Izetbegovi, Sandra Vegar-Zubovi, Maja Kuzmanovska, Golubinka Boshevska |
| EPI_ISL_1301483, EPI_ISL_1301549, EPI_ISL_1301561, EPI_ISL_1301567, EPI_ISL_1301569 | Instituto de Diagnostico y Referencia Epidemiologicos InDRE_RNLSP | Instituto de Biotecnología de la UNAM | Authors from IBT, IMSS, InDRE and INER (in alphabetical order): Carlos F. Arias, Santiago Ávila-Ríos, Gisela Barrera-Badillo, Eduardo Becerril-Vargas, Celia Boukadida, Natividad Cruz-Ortiz, Larissa Fernandes-Matano, Ricardo Grande, Lucia Hernandez-Rivas, Alejandra Hernández-Terán, Pavel Isa, Irma Lopez-Martinez, José Arturo Martínez-Orozco, Margarita Matías-Florentino, Fidencio Mejía-Nepomuceno, Edgar Mendieta-Condado, Mario |

| | | | |
|---|---|---|---|
| | | | Mújica-Sánchez, José Esteban Muñoz-Medina, Tatiana Nunez-Garcia, Luis Alberto Ochoa-Carrera, Hector Esteban Paz-Juárez, Francisco Pulido, José Ernesto Ramírez-González, Alma Rincón-Rubio, Teresita Rojas-Mendoza, Jorge Salas-Hernández, Alejandro Sanchez-Flores, Clara Esperanza Santacruz-Tinoco, Andrea Santos Coy-Arechavaleta, Blanca Taboada, Gloria Vazquez, Joel Armando Vázquez-Pérez, Jerome Jean Verleyen |
| EPI_ISL_1301622, EPI_ISL_1301646 | Laboratorio Central de Epidemiología IMSS | Instituto de Biotecnología de la UNAM | Authors from IBT, IMSS, InDRE and INER (in alphabetical order): Carlos F. Arias, Santiago Ávila-Ríos, Gisela Barrera-Badillo, Eduardo Becerril-Vargas, Celia Boukadida, Natividad Cruz-Ortiz, Larissa Fernandes-Matano, Ricardo Grande, Lucia Hernandez-Rivas, Alejandra Hernández-Terán, Pavel Isa, Irma Lopez-Martinez, José Arturo Martínez-Orozco, Margarita Matías-Florentino, Fidencio Mejía-Nepomuceno, Edgar Mendieta-Condado, Mario Mújica-Sánchez, José Esteban Muñoz-Medina, Tatiana Nunez-Garcia, Luis Alberto Ochoa-Carrera, Hector Esteban Paz-Juárez, Francisco Pulido, José Ernesto Ramírez-González, Alma Rincón-Rubio, Teresita Rojas-Mendoza, Jorge Salas-Hernández, Alejandro Sanchez-Flores, Clara Esperanza Santacruz-Tinoco, Andrea Santos Coy-Arechavaleta, Blanca Taboada, Gloria Vazquez, Joel Armando Vázquez-Pérez, Jerome Jean Verleyen |
| EPI_ISL_1301688 | Instituto de Diagnostico y Referencia Epidemiologicos InDRE_RNLSP | Instituto de Biotecnología de la UNAM | Authors from IBT, IMSS, InDRE and INER (in alphabetical order): Carlos F. Arias, Santiago Ávila-Ríos, Gisela Barrera-Badillo, Eduardo Becerril-Vargas, Celia Boukadida, Natividad Cruz-Ortiz, Larissa Fernandes-Matano, Ricardo Grande, Lucia Hernandez-Rivas, Alejandra Hernández-Terán, Pavel Isa, Irma Lopez-Martinez, José Arturo Martínez-Orozco, Margarita Matías-Florentino, Fidencio Mejía-Nepomuceno, Edgar Mendieta-Condado, Mario Mújica-Sánchez, José Esteban Muñoz-Medina, Tatiana Nunez-Garcia, Luis Alberto Ochoa-Carrera, Hector Esteban Paz-Juárez, Francisco Pulido, José Ernesto Ramírez-González, Alma Rincón-Rubio, Teresita Rojas-Mendoza, Jorge Salas-Hernández, Alejandro Sanchez-Flores, Clara Esperanza Santacruz-Tinoco, Andrea Santos Coy-Arechavaleta, Blanca Taboada, Gloria Vazquez, Joel Armando Vázquez-Pérez, Jerome Jean Verleyen |
| EPI_ISL_1301734, EPI_ISL_1301737, EPI_ISL_1301741, EPI_ISL_1301742, EPI_ISL_1301748, EPI_ISL_1301751, EPI_ISL_1301759, EPI_ISL_1301760, EPI_ISL_1301761, EPI_ISL_1301762, EPI_ISL_1301773, EPI_ISL_1301776, EPI_ISL_1301787, EPI_ISL_1301793 | | | |
| see above | Nucleic Acid Testing, National Reference Laboratory | GIGA Medical Genomics | Yvan Butera, Keith Durkin, Maria Artesi, Bouchra Boujemla, Robert Rutayisire, Patrick Tuyisenge, Esperence Umumararungu, Sébastien Bontems, Marie-Pierre Hayette, Nathalie Renotte, Swaibu Gatare, Jacob Souopgui, Sabin Nsanzimana, Vincent Bours, Léon Mutesa |
| EPI_ISL_1302094, EPI_ISL_1302144, EPI_ISL_1302145, EPI_ISL_1302146, EPI_ISL_1302147, EPI_ISL_1302148, EPI_ISL_1302150, EPI_ISL_1302151 | National Center of Infectious and Parasitic Diseases | National Center of Infectious and Parasitic Diseases | Alexiev et al |
| EPI_ISL_1302236, EPI_ISL_1302237, EPI_ISL_1302264 | Laboratorio Central de Epidemiología IMSS | Instituto de Biotecnología de la UNAM | Authors from IBT, IMSS, InDRE and INER (in alphabetical order): Carlos F. Arias, Santiago Ávila-Ríos, Gisela Barrera-Badillo, Eduardo Becerril-Vargas, Celia Boukadida, Natividad Cruz-Ortiz, Larissa Fernandes-Matano, Ricardo Grande, Lucia Hernandez-Rivas, Alejandra Hernández-Terán, Pavel Isa, Irma Lopez-Martinez, José Arturo Martínez-Orozco, Margarita Matías-Florentino, Fidencio Mejía-Nepomuceno, Edgar Mendieta-Condado, Mario Mújica-Sánchez, José Esteban Muñoz-Medina, Tatiana Nunez-Garcia, Luis Alberto Ochoa-Carrera, Hector Esteban Paz-Juárez, Francisco Pulido, José Ernesto Ramírez-González, Alma Rincón-Rubio, Teresita Rojas-Mendoza, Jorge Salas-Hernández, Alejandro Sanchez-Flores, Clara Esperanza Santacruz-Tinoco, Andrea Santos Coy-Arechavaleta, Blanca Taboada, Gloria Vazquez, Joel Armando Vázquez-Pérez, Jerome Jean Verleyen |
| EPI_ISL_1302335 | Instituto de Diagnostico y Referencia Epidemiologicos InDRE_RNLSP | Instituto de Biotecnología de la UNAM | Authors from IBT, IMSS, InDRE and INER (in alphabetical order): Carlos F. Arias, Santiago Ávila-Ríos, Gisela Barrera-Badillo, Eduardo Becerril-Vargas, Celia Boukadida, Natividad Cruz-Ortiz, Larissa Fernandes-Matano, Ricardo Grande, Lucia Hernandez-Rivas, Alejandra Hernández-Terán, Pavel Isa, Irma Lopez-Martinez, José Arturo Martínez-Orozco, Margarita Matías-Florentino, Fidencio Mejía-Nepomuceno, Edgar Mendieta-Condado, Mario Mújica-Sánchez, José Esteban Muñoz-Medina, Tatiana Nunez-Garcia, Luis Alberto Ochoa-Carrera, Hector Esteban Paz-Juárez, Francisco Pulido, José Ernesto Ramírez-González, Alma Rincón-Rubio, Teresita Rojas-Mendoza, Jorge Salas-Hernández, Alejandro Sanchez-Flores, Clara Esperanza Santacruz-Tinoco, Andrea Santos Coy-Arechavaleta, Blanca Taboada, Gloria Vazquez, Joel Armando Vázquez-Pérez, Jerome Jean Verleyen |
| EPI_ISL_1302451 | Medicina practica laboratorija | Lithuanian University of Health Sciences Hospital, Department of Genetics and Molecular Medicine | Rasa Ugenskiene, Darius Cereskevicius, Inga Nasvytiene, Zivile Zemeckiene, Mantas Sarauskas, Marius Sukys, Astra Vitkauskiene, Renaldas Jurkevicius |
| EPI_ISL_1302728, EPI_ISL_1302770 | Ministry of Health Turkey | Ministry of Health Turkey | Fatma Bayrakdar, Yasemin Cosgun, Suleyman Yalcin, Gulay Korukluoglu |
| EPI_ISL_1302900, EPI_ISL_1302928 | ADMED Microbiologie | Genomics and Transcriptomics, Philip Morris International | Reto Lienhard, Marie-Lise Tritten, Emmanuel Guedj, Nicolas Sierro, Rémi Dulize, David Bornand, Mehdi Auberson, Maxime Berthouzoz, Nikolai Ivanov, Manuel Peitsch |
| EPI_ISL_1303367 | Instituto Nacional de Salud- Dirección de Redes de Laboratorios de Salud Pública | Instituto Nacional de Salud- Dirección de Investigación en Salud Pública | Katherine Laiton-Donato, Diego A. Álvarez-Díaz, Carlos Franco-Muñoz, Hector Alejandro Ruiz-Moreno, Maria T. Herrera-Sepúlveda, Diego Andrés Prada, Jhonnatan Reales-González, Sheryll Corchuelo, Julian Naizaque, Gerardo Santamaría, Sergio Gomez, Lisseth Pardo, Juan Camilo Martinez, Marta Lopez Blanco, Ángela Alarcon Cruz, Diana Malo, Carmen Osorio, Magdalena Wiesner, Martha Lucia Ospina Martinez, Marcela Mercado-Reyes |
| EPI_ISL_1303559 | Houston Methodist Hospital | Houston Methodist Hospital | S. Wesley Long, Randall J. Olsen, Paul A. Christensen, Sishir Subedi, Robert Olson, James J. Davis, Matthew Ojeda Saavedra, Prasanti Yerramilli, Layne Pruitt, Kristina Reppond, Madison N. Shyer, Jessica Cambric, Ilya J. Finkelstein, Jimmy Gollihar, and James M. Musser |
| EPI_ISL_1306124 | Public Health Virology-Forensic and Scientific Services (PHV-FSS) | Public Health Virology-Forensic and Scientific Services (PHV-FSS) | Son Nguyen |
| EPI_ISL_1307659, EPI_ISL_1307665, EPI_ISL_1307671, EPI_ISL_1307676 | Central Public Health Laboratory | National Public Health Laboratory, National Centre for Infectious Diseases | Tze Minn Mak, Zhenyang Zhou, Esorom Daoni, Theresa Palou, Lin Cui, Raymond Tzer Pin Lin |
| EPI_ISL_1311324 | Dutch COVID-19 response team | Erasmus Medical Center | Bas Oude Munnink, Reina Sikkema, David Nieuwenhuijse, Irina Chestakova, Anne van der Linden, Marjan Boter, Emmanuelle Munger, Corine GeurtsvanKessel, Annemiek van der Eijk, Richard Molenkamp, Marion Koopmans, on behalf of the Dutch national COVID-19 response team. |
| EPI_ISL_1312009 | Public Health Institute of Zagreb County | Croatian Institute of Public Health | Irena Tabain, Ivana Ferenak |
| EPI_ISL_1312232 | KU Leuven, Rega Institute, Clinical and Epidemiological Virology | KU Leuven, Rega Institute, Clinical and Epidemiological Virology | Tony Wawina-Bokalanga, Bert Vanmechelen, Joan Marti-Carerras, Piet Maes |
| EPI_ISL_1312511, EPI_ISL_1312562 | Centrala laboratorija | Latvian Biomedical Research and Study Centre | Janis Pjalkovskis, Nikita Zrelovs, Monta Ustinova, Ivars Silamikelis, Liga Birzniece, Kaspars Megnis, Una Krumina, Guntars Zarins, Vita Rovite, Lauma Freimane, Laila Silamikele, Laura Ansone, Davids Fridmanis, Marta Priedite, Jana Osite, Jurijs Perevoscikovs, Uga Dumpis, Janis Klovins |
| EPI_ISL_1312683 | E. Gulbja laboratorija | Latvian Biomedical Research and Study Centre | Janis Pjalkovskis, Nikita Zrelovs, Monta Ustinova, Ivars Silamikelis, Liga Birzniece, Kaspars Megnis, Una Krumina, Guntars Zarins, Vita Rovite, Lauma Freimane, Laila Silamikele, Laura Ansone, Davids Fridmanis, Mikus Gavars, Dmitrijs Perminovs, Jurijs Perevoscikovs, Uga Dumpis, Janis Klovins |
| EPI_ISL_1312777 | Molecular & Genomic Pathology Laboratory, Thomas Jefferson University Hospital | Molecular & Genomic Pathology Laboratory, Thomas Jefferson University Hospital | Run Jin, Nitika Badjatia, Zi-xuan Wang |
| EPI_ISL_1314853 | Quest Diagnostics Incorporated | Centers for Disease Control and Prevention Division of Viral Diseases, Pathogen Discovery | Peter W. Cook, Dakota Howard, Dhwani Batra, Ben L. Rambo-Martin, S. H. Rosenthal, A. Gerasimova, R. M. Kagan, B. Anderson, M. Hua, Y. Liu, L.E. Bernstein, K.E. Livingston, A. Perez, I. A. Shlyakhter, R. V. Rolando, R. Owen, P. Tanpaiboon, F. Lacbawan, Clinton R. Paden, Suxiang Tong, Duncan MacCannell |
| EPI_ISL_1315066 | New South Wales Health Pathology Royal Prince Alfred Hospital | Microbiology RPAH | Foster, C.; Au, J.; Ruiz Silva, M.; Deveson, I.; Bull, R.; Van Hal, S.; Rawlinson, W. |
| EPI_ISL_1315312, EPI_ISL_1315326 | LabPLUS | Institute of Environmental Science and Research (ESR) | Rachel Boyle, SallyAnn Harbison, Olivia Stroeven, Xiaoyun Ren, Matt Storey, Nikki Freed, Muhammad Faisal, Jing Wang, Hermes Perez, Anja Werno, Antje van der Linden, Arlo Upton, Chris Mansell, David Hammer, Dragana Drinkovic, Gary McAuliffe, Hana Sofia Andersson, James Ussher, Jill Sherwood, Josh Freeman, Julia Howard, Juliet Elvy, Mary DeAlmeida, Matt Blakiston, Matthew Rogers, Max Bloomfield, Michael Addidle, Michelle Balm, Sally Roberts, Sarah Jefferies, Sharmini Muttaiyah, Susan Morpeth, Susan Taylor, Timothy Blackmore, Vani Sathyendran, Veronica Playle, Virginia Hope, Erasmus Smit, Lauren Jelly, Olin Silander, Joep de Ligt |
| EPI_ISL_1315347, EPI_ISL_1315357, EPI_ISL_1315395, EPI_ISL_1315398 | Division of Emerging Infectious Diseases, Bureau of Infectious Diseases Diagnosis Control, Korea Disease Control and Prevention Agency | Division of Emerging Infectious Diseases, Bureau of Infectious Diseases Diagnosis Control, Korea Disease Control and Prevention Agency | Ae Kyung Park, Il-Hwan Kim, Heui Man Kim, Jeong-Min Kim, Jeong-Ah Kim, Chae Young Lee, Jin Sun No, Eun-Jin Kim |
| EPI_ISL_1317722 | Norwegian Institute of Public Health, Department of Virology | Norwegian Institute of Public Health, Department of Virology | Kathrine Stene-Johansen, Kamilla Heddeland Instefjord, Hilde Elshaug, Garcia Llorente Ignacio, Jon Bråte, Engebretsen Serina Beate,Pedersen Benedikte Nevjen, Debech Nadia, Atiya R Ali,Marie Paulsen Madsen, Rasmus Riis Kopperud, Hilde Vollan, Karoline Bragstad, Olav Hungnes |

| | | | |
|---|---|---|---|
| EPI_ISL_1318431, EPI_ISL_1318434, EPI_ISL_1318435, EPI_ISL_1318444 | ND Dept. of Health Laboratory Services-Microbiology | Centers for Disease Control and Prevention Division of Viral Diseases, Pathogen Discovery | Krista Queen, Yan Li, Ying Tao, Jing Zhang, Anna Uehara, Anna Montmayeur, Clinton R. Paden, Kristen Knipe, Matthew Schmerer, Shoshona Le, Katie Dillon, Peter W. Cook, Rachel Marine, Mili Sheth, Jasmine Padilla, Sarah Nobles, Mark Burroughs, Lori Rowe, Haibin Wang, Ben L. Rambo-Martin, Kristine Lacek, Sam Shepard, Dhwani Batra, Suxiang Tong, Justin Lee |
| EPI_ISL_1319069 | Laboratory Corporation of America | Centers for Disease Control and Prevention Division of Viral Diseases, Pathogen Discovery | Peter W. Cook, Dakota Howard, Dhwani Batra, Ben L. Rambo-Martin, Minoo Agarwal, Eyad Almasri, Debbie Boles, Ayla Burns, Nuthawin Charoensri, Oren Cohen, Susan Countryman, Mary Ann Cristobal, Bobbi Croy, Suzanne Dale, Hrushikesh Deshmukh, Amanda Douglas, Vincent Drouillon, Marcia Eisenberg, Howard Engler, Rama Ghatti, Prashant Gupta, Susan Hicks, Jake Humphrey, Lax Iyer, Manoj Jain, Mohan Kolli, Brian Krueger, Tim Kuphal, Stanley Letovsky, Michael Levandoski, Craig Lukasik, Jonathan Meltzer, Brian Norvell, Mindy Nye, Scott Parker, Christos Petropoulos, John Pruitt, Steven Ragan, Scott Ryan, Mike Sapeta, Jana Schroth, Suresh Babu Selvaraju, Goran Stevovic, Amanda Suchanek, Andrea Throop, Lyndon Tilson, Thomas Urban, Joe Voshell, Kimberly Wagner, Jonathan Williams, Mary Williamson, Qian Zeng, Tricia Zwiefelhofer, Clinton R. Paden, Suxiang Tong, Duncan MacCannell |
| EPI_ISL_1319399 | Synlab Eesti OÜ | 1. Laboratory of Communicable Diseases (Estonia); 2. Eurofins Genomics Europe Sequencing GmbH | Liidia Dotsenko et al. |
| EPI_ISL_1320852 | Laboratory Corporation of America | Centers for Disease Control and Prevention Division of Viral Diseases, Pathogen Discovery | Peter W. Cook, Dakota Howard, Dhwani Batra, Ben L. Rambo-Martin, Minoo Agarwal, Eyad Almasri, Debbie Boles, Ayla Burns, Nuthawin Charoensri, Oren Cohen, Susan Countryman, Mary Ann Cristobal, Bobbi Croy, Suzanne Dale, Hrushikesh Deshmukh, Amanda Douglas, Vincent Drouillon, Marcia Eisenberg, Howard Engler, Rama Ghatti, Prashant Gupta, Susan Hicks, Jake Humphrey, Lax Iyer, Manoj Jain, Mohan Kolli, Brian Krueger, Tim Kuphal, Stanley Letovsky, Michael Levandoski, Craig Lukasik, Jonathan Meltzer, Brian Norvell, Mindy Nye, Scott Parker, Christos Petropoulos, John Pruitt, Steven Ragan, Scott Ryan, Mike Sapeta, Jana Schroth, Suresh Babu Selvaraju, Goran Stevovic, Amanda Suchanek, Andrea Throop, Lyndon Tilson, Thomas Urban, Joe Voshell, Kimberly Wagner, Jonathan Williams, Mary Williamson, Qian Zeng, Tricia Zwiefelhofer, Clinton R. Paden, Suxiang Tong, Duncan MacCannell |
| EPI_ISL_1321413 | National Institute of Public Health | National Reference Laboratory for Influenza and Respiratory Viruses CZE | Helena Jirincova, Jaromira Vecerova, Timotej Suri, Dusan Trnka, Alexander Nagy |
| EPI_ISL_1321479, EPI_ISL_1321518, EPI_ISL_1321537, EPI_ISL_1321563, EPI_ISL_1321572, EPI_ISL_1321573, EPI_ISL_1321587 | Genetica Molecular and Subdepartamento de Virologia ISP Chile | Instituto de Salud Publica de Chile | Javier Tognarelli, Karen Orostica, Barbara Parra, Loredana Arata, Jaime Lagos, Gisselle Barra, Patricia Bustos, Rodrigo Fasce, Andres Castillo, Jorge Fernandez |
| EPI_ISL_1322322, EPI_ISL_1322323, EPI_ISL_1322324, EPI_ISL_1322325, EPI_ISL_1322327, EPI_ISL_1322328 | Central Public Health Laboratory | National Public Health Laboratory, National Centre for Infectious Diseases | Tze Minn Mak, Zhenyang Zhou, Esorom Daoni, Theresa Palou, Lin Cui, Raymond Tzer Pin Lin |
| EPI_ISL_1322732 | KU Leuven, Rega Institute, Clinical and Epidemiological Virology | KU Leuven, Rega Institute, Clinical and Epidemiological Virology | Tony Wawina-Bokalanga, Bert Vanmechelen, Joan Marti-Carerras, Piet Maes |
| EPI_ISL_1323453, EPI_ISL_1323497 | Illinois Department of Public Health | Gagnon Lab, Southern Illinois University | Keith Gagnon |
| EPI_ISL_1324730 | UAB InMedica | Vilnius University Hospital Santaros Klinikos, Center of Laboratory Medicine | Gytis Dudas, Rimvydas Norvilas, Dovile Ezerskyte, Daniel Naumovas, Ingrida Olendraite, Ligita Raugaite, Monika Katenaite, Mindaugas Stoskus, Laimonas Griskevicius |
| EPI_ISL_1324758 | Laboratorio de Diagnostico de Enfermedades Emergentes y Reemergentes | Instituto de Diagnostico y Referencia Epidemiologicos (INDRE) | Claudia Wong-Arambula, Abril Rodriguez-Maldonado, Vanessa Rivero-Arredondo, Ariadna Medina-Benitez, Joaquin Quiroz-Mercado, Sergio Rangel-Guerrero,Natali Vega-Magana, Natividad Cruz-Ortiz, Tatiana Nunez-Garcia, Gisela Barrera-Badillo, Lucia Hernandez-Rivas, Irma Lopez-Martinez, Ernesto Ramirez-Gonzalez. |
| EPI_ISL_1327546 | Anteja laboratorija (UAB Diagnostikos laboratorija) | Vilnius University Hospital Santaros Klinikos, Center of Laboratory Medicine | Ingrida Olendraite, Gytis Dudas, Rimvydas Norvilas, Dovile Ezerskyte, Daniel Naumovas, Ligita Raugaite, Monika Katenaite, Mindaugas Stoskus, Laimonas Griskevicius |
| EPI_ISL_1334380 | Laboratorios Lister | Instituto de Diagnostico y Referencia Epidemiologicos (INDRE) | Claudia Wong-Arambula, Abril Rodriguez-Maldonado, Vanessa Rivero-Arredondo, Ariadna Medina-Benitez, Joaquin Quiroz-Mercado, Sergio Rangel-Guerrero, Natividad Cruz-Ortiz, Tatiana Nunez-Garcia, Gisela Barrera-Badillo, Lucia Hernandez-Rivas, Irma Lopez-Martinez, Ernesto Ramirez-Gonzalez. |
| EPI_ISL_1334381 | LESP Guerrero | Instituto de Diagnostico y Referencia Epidemiologicos (INDRE) | Claudia Wong-Arambula, Abril Rodriguez-Maldonado, Vanessa Rivero-Arredondo, Ariadna Medina-Benitez, Joaquin Quiroz-Mercado, Sergio Rangel-Guerrero, Natividad Cruz-Ortiz, Tatiana Nunez-Garcia, Gisela Barrera-Badillo, Lucia Hernandez-Rivas, Irma Lopez-Martinez, Ernesto Ramirez-Gonzalez. |
| EPI_ISL_1334382, EPI_ISL_1334383, EPI_ISL_1334384, EPI_ISL_1334385 | Laboratorios Lister | Instituto de Diagnostico y Referencia Epidemiologicos (INDRE) | Claudia Wong-Arambula, Abril Rodriguez-Maldonado, Vanessa Rivero-Arredondo, Ariadna Medina-Benitez, Joaquin Quiroz-Mercado, Sergio Rangel-Guerrero, Natividad Cruz-Ortiz, Tatiana Nunez-Garcia, Gisela Barrera-Badillo, Lucia Hernandez-Rivas, Irma Lopez-Martinez, Ernesto Ramirez-Gonzalez. |
| EPI_ISL_1334386 | LESP Guanajuato | Instituto de Diagnostico y Referencia Epidemiologicos (INDRE) | Claudia Wong-Arambula, Abril Rodriguez-Maldonado, Vanessa Rivero-Arredondo, Ariadna Medina-Benitez, Joaquin Quiroz-Mercado, Sergio Rangel-Guerrero, Natividad Cruz-Ortiz, Tatiana Nunez-Garcia, Gisela Barrera-Badillo, Lucia Hernandez-Rivas, Irma Lopez-Martinez, Ernesto Ramirez-Gonzalez. |
| EPI_ISL_1334395 | Wyoming Public Health Laboratory | Wyoming Public Health Laboratory | Jim Mildenberger, Wanda Manley, Noah Hull, Taylor Fearing, Lynette Gumbleton, Channing Weber, Ashley Norberg, Chayse Rowley, Marley Goetz, Brian Dominguez, Elliot Thomasson, Cari Sloma, and Rob Christensen |
| EPI_ISL_1334517 | Laboratory of virology, National center of expertise | RSE "National Center for Biotechnology" and RSE "National Center of Expertise" | Shevtsov Alexandr, Amirgazin Asylulan, Kamalova Dinara, Abdaliyev Askar, Tungushbayev Talgat, Sharipova Saule, Balykbaev Kanat, Ramankulov Yerlan |
| EPI_ISL_1334553 | Clinical Molecular Microbiology Laboratory, UNC Hospital | Dirk Dittmer | Justin T. Landis, Razia Moorad, Brent A. Eason, Melissa B. Miller, Linda Pluta, Dirk Dittmer, Angelica Juarez, Cameroon Grant, Evelyn Hoffman, Patricio Cano, Jason Wong, Carolina Caro-Vegas, Ryan McNamara, Danielle Chappell, Cecilia Thompson, Shawn Hawken, Jason Smedberg, Andrea McDaniel-Harper, Billy Fisher, Ashwini Roy-Chaudhury, Blossom Damania. |
| EPI_ISL_1334569, EPI_ISL_1334574 | Laboratory of virology, National center of expertise | RSE "National Center for Biotechnology" and RSE "National Center of Expertise" | Shevtsov Alexandr, Amirgazin Asylulan, Kamalova Dinara, Abdaliyev Askar, Tungushbayev Talgat, Sharipova Saule, Balykbaev Kanat, Ramankulov Yerlan |
| EPI_ISL_1335757 | LESP Zacatecas | Instituto de diagnóstico y Referencia Epidemiologicos (INDRE) Departamento de Virología | Claudia Wong-Arambula, Abril Rodriguez-Maldonado, Vanessa Rivero-Arredondo, Ariadna Medina-Benitez, Joaquin Quiroz-Mercado, Sergio Rangel-Guerrero, Natividad Cruz-Ortiz, Tatiana Nunez-Garcia, Gisela Barrera-Badillo, Lucia Hernandez-Rivas, Irma Lopez-Martinez, Ernesto Ramirez-Gonzalez. |
| EPI_ISL_1335895 | Martha Friska Multatuli Hospital Medan | Faculty of Medicine, Universitas Sumatera Utara; Institute of Tropical Disease, Universitas Airlangga | Krisnoadi Rahardjo, Aldise M Nastri, Jezzy R Dewantari, Rima R Prasetya, Yasuko Mori, Soetjipto, Kazufumi Shimizu, Maria I Lusida, Meliani, R Andika D Cahyadi, Mirzan Hasibuan, Irbah R Nainggolan, Ramadhan Bestari, Franciscus Ginting, Muhammad Ichwan, R Lia Kusumawati, Inke N D Lubis |
| EPI_ISL_1336649 | HOPITAL PRINCESSE GRACE | CNR Virus des Infections Respiratoires - France SUD | Antonin Bal, Gregory Destras, Gwendolyne Burfin, Hadrien Regue, Quentin Semanas, Martine Valette, Bruno Lina, Laurence Josset |
| EPI_ISL_1337373 | LESP Baja California Sur | Instituto de Diagnostico y Referencia Epidemiologicos (INDRE) | Claudia Wong-Arambula, Abril Rodriguez-Maldonado, Vanessa Rivero-Arredondo, Ariadna Medina-Benitez, Joaquin Quiroz-Mercado, Sergio Rangel-Guerrero, Natividad Cruz-Ortiz, Tatiana Nunez-Garcia, Gisela Barrera-Badillo, Lucia Hernandez-Rivas, Irma Lopez-Martinez, Ernesto Ramirez-Gonzalez. |
| EPI_ISL_1337899 | Dutch COVID-19 response team | Medical Microbiology, Maastricht University Medical Centre | Jozef Dingemans*, Brian van der Veer*, Erik Beuken, Carmen Reumkens, Lieke van Alphen, Christian Hoebe, Paul Savelkoul |
| EPI_ISL_1337989 | DC Public Health Lab/ Dept. of Forensic Sciences | DC Public Health Lab/ Dept. of Forensic Sciences | Janis Doss, Scott Nguyen, Elizabeth Zelaya, Sarah Scott, Connie Maza, Monica Mann, Brittany Hamilton, David Payne, Jocelyn Hauser |
| EPI_ISL_1339817 | Laboratory Corporation of America | Centers for Disease Control and Prevention Division of Viral Diseases, Pathogen Discovery | Peter W. Cook, Dakota Howard, Dhwani Batra, Ben L. Rambo-Martin, Minoo Agarwal, Eyad Almasri, Debbie Boles, Ayla Burns, Nuthawin Charoensri, Oren Cohen, Susan Countryman, Mary Ann Cristobal, Bobbi Croy, Suzanne Dale, Hrushikesh Deshmukh, Amanda Douglas, Vincent Drouillon, Marcia Eisenberg, Howard Engler, Rama Ghatti, Prashant Gupta, Susan Hicks, Jake Humphrey, Lax Iyer, Manoj Jain, Mohan Kolli, Brian Krueger, Tim Kuphal, Stanley Letovsky, Michael Levandoski, Craig Lukasik, Jonathan Meltzer, Brian Norvell, Mindy Nye, Scott Parker, Christos Petropoulos, John Pruitt, Steven |

| | | | |
|---|---|---|---|
| | | | Ragan, Scott Ryan, Mike Sapeta, Jana Schroth, Suresh Babu Selvaraju, Goran Stevovic, Amanda Suchanek, Andrea Throop, Lyndon Tilson, Thomas Urban, Joe Voshell, Kimberly Wagner, Jonathan Williams, Mary Williamson, Qian Zeng, Tricia Zwiefelhofer, Clinton R. Paden, Suxiang Tong, Duncan MacCannell |
| EPI_ISL_1339991 | DPHL | Delaware Public Health Lab | Rebecca Savage |
| EPI_ISL_1340630 | LESP Guanajuato | Instituto de Diagnostico y Referencia Epidemiologicos (INDRE) | Claudia Wong-Arambula, Abril Rodriguez-Maldonado, Vanessa Rivero-Arredondo, Ariadna Medina-Benitez, Joaquin Quiroz-Mercado, Sergio Rangel-Guerrero, Natividad Cruz-Ortiz, Tatiana Nunez-Garcia, Gisela Barrera-Badillo, Lucia Hernandez-Rivas, Irma Lopez-Martinez, Ernesto Ramirez-Gonzalez. |
| EPI_ISL_1340752, EPI_ISL_1340753, EPI_ISL_1340755, EPI_ISL_1340759, EPI_ISL_1340761, EPI_ISL_1340762, EPI_ISL_1340763, EPI_ISL_1340764 | Departamento de Virologia, Laboratorio Central de Salud Pública, Avenida Venezuela y Teniente Escurra, Asunción, Paraguay | Laboratory of Respiratory Viruses and Measles, Oswaldo Cruz Institute, FIOCRUZ | Paola Resende, Cynthia Vazquez, Luciana Appolinario, Fernando Motta, Anna Carolina Paixao, Ana Carolina Mendonca, Alice Sampaio Rocha, Renata Serrano Lopes, Marilda Siqueira on behalf of the Fiocruz COVID-19 Genomic Surveillance Network |
| EPI_ISL_1340867 | West Bandung Public Health | West Java Health Laboratory; School of Life Sciences and Technology, Institut Teknologi Bandung | Azzania Fibriani, Ema Rahmawati, Ryan Bayusantika Ristandi, Rifky Waluyajati Rachman, Cut Nur Cinthia Alamanda, Isak Solihin, Rini Robiani, Miftahul Faridl, Karimatu Khoirunnisa |
| EPI_ISL_1341378, EPI_ISL_1341501, EPI_ISL_1341505 | Laboratory of Virology, National center of expertise | RSE "National Center of Expertise" and RSE "National center for Biotechnology" | Abdaliyev Askar, Tungushbayev Talgat, Sharipova Saule, Shevtsov Alexandr, Amirgazin Asylulan, Kamalova Dinara, Ramankulov Erlan, Balykbaev Kanat |
| EPI_ISL_1341507 | New South Wales Health Pathology Royal Prince Alfred Hospital | Microbiology RPAH | Foster, C.; Au, J.; Ruiz Silva, M.; Deveson, I.; Bull, R.; Van Hal, S.; Rawlinson, W. |
| EPI_ISL_1341641, EPI_ISL_1341642 | Laboratory of Virology, National center of expertise | RSE "National Center of Expertise" and RSE "National center for Biotechnology" | Abdaliyev Askar, Tungushbayev Talgat, Sharipova Saule, Shevtsov Alexandr, Amirgazin Asylulan, Kamalova Dinara, Ramankulov Erlan, Balykbaev Kanat |
| EPI_ISL_1341651 | UW Virology Lab | UW Virology Lab | Pavitra Roychoudhury, Hong Xie, Lasata Shrestha, Shah Mohamed Bakhash, Michelle Lin, Noah R. Baker, Sean Ellis, Saraswathi Sathees, Meei-Li Huang, Keith R Jerome, Alexander Greninger |
| EPI_ISL_1347023 | Microbiology Department, Laboratori Clínic Metropolitana Nord. Hospital Universitari Germans Trias i Pujol. | Can Ruti SARS-CoV-2 Sequencing Hub (HUGTiP/IrsiCaixa/IGTP) | Marc Noguera-Julian, Pilar Armengol, Ignacio Blanco, Antoni E Bordoy, Francesc Catala-Moll, Pere-Joan Cardona, Maria Casadellà, Cristina Casañ, Gemma Clara, Bonaventura Clotet, Cristina Esteban, Montserrat Giménez, Mercedes Guerrero, Anna Not, Roger Paredes, Mariona Parera, Verónica Saludes, Alba Sánchez, and Elisa Martró on behalf of the Can Ruti SARS-CoV-2 Sequencing Hub. |
| EPI_ISL_1347881, EPI_ISL_1347885, EPI_ISL_1347894, EPI_ISL_1347899, EPI_ISL_1347907, EPI_ISL_1347921, EPI_ISL_1347931 | Instituto Nacional de Investigação em Saúde | KRISP, KZN Research Innovation and Sequencing Platform | Morais J, Neto Z, Afonso P, Miranda J, David K, Inglês L, Pereira A, Paulo A Carralero RR Paixão JP, Freitas RH, Mufinda M, Lutucuta S, Giandhari J, Pillay S, Naidoo Y, Emmanuel SJ, Tegally H, Wilkinson E, de Oliveira T |
| EPI_ISL_1351435, EPI_ISL_1351443 | Centro de InvestigaciÃ³n BiomÃ©dica del Noreste (CIBIN) | Unidad de Genomica Avanzada | Consorcio Mexicano de Vigilancia Genomica (CoViGen-Mex). Authors (in alphabetical order): Julio Elias Alvarado-Yaah, Carlos F. Arias, Santiago Avila-Rios, Victor Hugo Borja-Aburto, Celia Boukadida, Juan Bautista Chale-Dzul , Jose Antonio Enciso-Moreno, Gloria Elena Espinoza-Ayala, Fernando Fontove-Herrera, Concepcion Grajales-Muniz, Ricardo Grande, Alfredo Herrera-Estrella, Carla Ivon Herrera-Najera, Pavel Isa, Brenda Irasema Maldonado-Meza, Bernardo Martinez-Miguel, Margarita Matias-Florentino, Maria Guadalupe de Jesus Mireles-Rivera, Gloria Maria Molina-Salinas, Hector Montoya-Fuentes, Jose Esteban Munoz-Medina, Jose de Jesus Nunez-Contreras, Alicia Ocana-Mondragon, Luis Alberto Ochoa-Carrera, Hector Esteban Paz-Juarez, Francisco Pulido, Helen Haydee Fernanda Ramirez-Plascencia, Angel Gustavo Salas-Lais, Jorge Ivan Salinal-Nevarez, Alejandro Sanchez-Flores, Clara Esperanza Santacruz-Tinoco, Maria Guadalupe Santiago-Mauricio, Nelly Selem-Mojica, Blanca Taboada, Gloria Vazquez |
| EPI_ISL_1351474, EPI_ISL_1351498 | Unidad de InvestigaciÃ³n MÃ©dica de YucatÃ¡n (UIMY) | Unidad de Genomica Avanzada | Consorcio Mexicano de Vigilancia Genomica (CoViGen-Mex). Authors (in alphabetical order): Julio Elias Alvarado-Yaah, Carlos F. Arias, Santiago Avila-Rios, Victor Hugo Borja-Aburto, Celia Boukadida, Juan Bautista Chale-Dzul , Jose Antonio Enciso-Moreno, Gloria Elena Espinoza-Ayala, Fernando Fontove-Herrera, Concepcion Grajales-Muniz, Ricardo Grande, Alfredo Herrera-Estrella, Carla Ivon Herrera-Najera, Pavel Isa, Brenda Irasema Maldonado-Meza, Bernardo Martinez-Miguel, Margarita Matias-Florentino, Maria Guadalupe de Jesus Mireles-Rivera, Gloria Maria Molina-Salinas, Hector Montoya-Fuentes, Jose Esteban Munoz-Medina, Jose de Jesus Nunez-Contreras, Alicia Ocana-Mondragon, Luis Alberto Ochoa-Carrera, Hector Esteban Paz-Juarez, Francisco Pulido, Helen Haydee Fernanda Ramirez-Plascencia, Angel Gustavo Salas-Lais, Jorge Ivan Salinal-Nevarez, Alejandro Sanchez-Flores, Clara Esperanza Santacruz-Tinoco, Maria Guadalupe Santiago-Mauricio, Nelly Selem-Mojica, Blanca Taboada, Gloria Vazquez |
| EPI_ISL_1351540 | Centro de InvestigaciÃ³n BiomÃ©dica de Occidente (CIBO) | Unidad de Genomica Avanzada | Consorcio Mexicano de Vigilancia Genomica (CoViGen-Mex). Authors (in alphabetical order): Julio Elias Alvarado-Yaah, Carlos F. Arias, Santiago Avila-Rios, Victor Hugo Borja-Aburto, Celia Boukadida, Juan Bautista Chale-Dzul , Jose Antonio Enciso-Moreno, Gloria Elena Espinoza-Ayala, Fernando Fontove-Herrera, Concepcion Grajales-Muniz, Ricardo Grande, Alfredo Herrera-Estrella, Carla Ivon Herrera-Najera, Pavel Isa, Brenda Irasema Maldonado-Meza, Bernardo Martinez-Miguel, Margarita Matias-Florentino, Maria Guadalupe de Jesus Mireles-Rivera, Gloria Maria Molina-Salinas, Hector Montoya-Fuentes, Jose Esteban Munoz-Medina, Jose de Jesus Nunez-Contreras, Alicia Ocana-Mondragon, Luis Alberto Ochoa-Carrera, Hector Esteban Paz-Juarez, Francisco Pulido, Helen Haydee Fernanda Ramirez-Plascencia, Angel Gustavo Salas-Lais, Jorge Ivan Salinal-Nevarez, Alejandro Sanchez-Flores, Clara Esperanza Santacruz-Tinoco, Maria Guadalupe Santiago-Mauricio, Nelly Selem-Mojica, Blanca Taboada, Gloria Vazquez |
| EPI_ISL_1351716 | Laboratorio Central de EpidemiologÃ-a (LCE) | Unidad de Genomica Avanzada | Consorcio Mexicano de Vigilancia Genomica (CoViGen-Mex). Authors (in alphabetical order): Julio Elias Alvarado-Yaah, Carlos F. Arias, Santiago Avila-Rios, Victor Hugo Borja-Aburto, Celia Boukadida, Juan Bautista Chale-Dzul , Jose Antonio Enciso-Moreno, Gloria Elena Espinoza-Ayala, Fernando Fontove-Herrera, Concepcion Grajales-Muniz, Ricardo Grande, Alfredo Herrera-Estrella, Carla Ivon Herrera-Najera, Pavel Isa, Brenda Irasema Maldonado-Meza, Bernardo Martinez-Miguel, Margarita Matias-Florentino, Maria Guadalupe de Jesus Mireles-Rivera, Gloria Maria Molina-Salinas, Hector Montoya-Fuentes, Jose Esteban Munoz-Medina, Jose de Jesus Nunez-Contreras, Alicia Ocana-Mondragon, Luis Alberto Ochoa-Carrera, Hector Esteban Paz-Juarez, Francisco Pulido, Helen Haydee Fernanda Ramirez-Plascencia, Angel Gustavo Salas-Lais, Jorge Ivan Salinal-Nevarez, Alejandro Sanchez-Flores, Clara Esperanza Santacruz-Tinoco, Maria Guadalupe Santiago-Mauricio, Nelly Selem-Mojica, Blanca Taboada, Gloria Vazquez |
| EPI_ISL_1352331 | MDI Limbach Berlin GmbH | Robert Koch Institute | unknown |
| EPI_ISL_1357801 | Israel Central Virology laboratory | Israel National Consortium for SARS-CoV-2 sequencing | Neta Zuckerman, Efrat Dahan Bucris, Michal Mandelboim, Dana Bar-Ilan, Oran Erster, Tzvia Mann, Omer Murik, David A. Zeevi, Assaf Rokney, Joseph Jaffe, Eva Nachum, Maya Davidovich Cohen, Ephraim Fass, Gal Zizelski Valenci, Mor Rubinstein, Efrat Rorman, Israel Nissan, Efrat Glick-Saar, Omri Nayshool, Gideon Rechavi, Ella Mendelson, Orna Mor |
| EPI_ISL_1358330 | Ministry of Health Turkey | Ministry of Health Turkey | Fatma Bayrakdar, Yasemin Cogun, Süleyman Yalcin, Gülay Korukluolu |
| EPI_ISL_1359067 | LESP Yucatan | Instituto de Diagnostico y Referencia Epidemiologicos (INDRE) | Claudia Wong-Arambula, Abril Rodriguez-Maldonado, Vanessa Rivero-Arredondo, Ariadna Medina-Benitez, Joaquin Quiroz-Mercado, Sergio Rangel-Guerrero, Natividad Cruz-Ortiz, Tatiana Nunez-Garcia, Gisela Barrera-Badillo, Lucia Hernandez-Rivas, Irma Lopez-Martinez, Ernesto Ramirez-Gonzalez. |
| EPI_ISL_1360051 | Klinisch Laboratorium ZNA | Klinisch Laboratorium ZNA | Verstrepen et al. |
| EPI_ISL_1361989 | Swedish national genomic surveillance program of SARS-CoV-2 | The Public Health Agency of Sweden | Swedish national genomic surveillance program of SARS-CoV-2 |
| EPI_ISL_1363281 | Sonora Quest Laboratories | TGen North | Jolene Bowers, Heather Centner, Chris French, Hayley Yaglom, Ashlyn Pfeiffer, Darrin Lemmer, Dave Engelthaler, The Arizona COVID Genomics Union (ACGU) |
| EPI_ISL_1363787 | Clin & Gen Lab | Microbiologia Molecular, Instituto SELADIS, Universidad Mayor de San Andrés | Oscar M. Rollano-Peñaloza, Aneth Vasquez Michel |
| EPI_ISL_1364540 | Public Health Virology-Forensic and Scientific Services (PHV-FSS) | Public Health Virology-Forensic and Scientific Services (PHV-FSS) | Son Nguyen |
| EPI_ISL_1364615, EPI_ISL_1364618, EPI_ISL_1364619, EPI_ISL_1364863, | Laboratory of virology, National center of expertise | RSE "National Center for Biotechnology" and RSE "National Center of Expertise" | Shevtsov Alexandr, Amirgazin Asylulan, Kamalova Dinara, Abdaliyev Askar, Tungushbayev Talgat, Sharipova Saule, Balykbaev Kanat, Ramankulov Yerlan |

| | | | |
|---|---|---|---|
| EPI_ISL_1364864, EPI_ISL_1364879 | | | |
| EPI_ISL_1365028, EPI_ISL_1365033 | Molecular diagnostic unit for viral haemorrhagic fevers and emerging viruses, Bouaké CHU Laboratory | Molecular diagnostic unit for viral haemorrhagic fevers and emerging viruses, Bouaké CHU Laboratory | Chantal Akoua-Koffi, Diané Bamourou, Etilé Anoh, Oby Wayoro, Safiatou Karidioula, Adjaratou Traoré, Soundélé Maïté, Monemo Pacome, Coulibaly Mbegnan, Bamba Fatoumata Touré, Kra Ouffoué, Grit Schubert, Essia Belarbi, Fabian Leendertz |
| EPI_ISL_1365640 | LESP Nuevo Leon | Instituto de Diagnostico y Referencia Epidemiologicos (INDRE) | Claudia Wong-Arambula, Abril Rodriguez-Maldonado, Vanessa Rivero-Arredondo, Ariadna Medina-Benitez, Joaquin Quiroz-Mercado, Sergio Rangel-Guerrero, Natividad Cruz-Ortiz, Tatiana Nunez-Garcia, Gisela Barrera-Badillo, Lucia Hernandez-Rivas, Irma Lopez-Martinez, Ernesto Ramirez-Gonzalez. |
| EPI_ISL_1365649 | LESP Tamaulipas | Instituto de Diagnostico y Referencia Epidemiologicos (INDRE) | Claudia Wong-Arambula, Abril Rodriguez-Maldonado, Vanessa Rivero-Arredondo, Ariadna Medina-Benitez, Joaquin Quiroz-Mercado, Sergio Rangel-Guerrero, Natividad Cruz-Ortiz, Tatiana Nunez-Garcia, Gisela Barrera-Badillo, Lucia Hernandez-Rivas, Irma Lopez-Martinez, Ernesto Ramirez-Gonzalez. |
| EPI_ISL_1365734, EPI_ISL_1365736, EPI_ISL_1365743 | Laboratory of Virology, National center of expertise | RSE "National Center of Expertise" and RSE "National center for Biotechnology" | Abdaliyev Askar, Tungushbayev Talgat, Sharipova Saule, Shevtsov Alexandr, Amirgazin Asylulan, Kamalova Dinara, Ramankulov Erlan, Balykbaev Kanat |
| EPI_ISL_1365827 | Laboratoire de santé publique du Québec | Laboratoire de santé publique du Québec | Sandrine Moreira, Ioannis Ragoussis, Guillaume Bourque, Jesse Shapiro, Mark Lathrop and Michel Roger on behalf of the CoVSeQ research group (http://covseq.ca/researchgroup) |
| EPI_ISL_1366083 | Dr. Soetomo Teaching Hospital | Research Center for Vaccine Technology and Development, Institute of Tropical Disease, Airlangga University | Fedik Abdul Rantam, Cita Rosita Sigit Prakoeswa, Damayanti Tinduh, Jusak Nugraha, Helen Susilowati, Andi Yasmin Wijaya, Ni Nyoman Tri Puspaningsih, Dwiyanti Puspitasari, Dominicus Husada, Neneng Dewi Kurniati, Aryati |
| EPI_ISL_1366334 | Genomica Lab Molecular, Mexico | Andersen lab at Scripps Research | SEARCH Alliance San Diego with Jonathan Gonzalez Garcia, Jose Roman Chavez Mendez, Jose Horacio Reyna Verdugo, Martin Gonzalez Ibarra, Luis Alberto Rangel Gonzalez |
| EPI_ISL_1366620 | Gundersen Clinical Microbiology Laboratory | Kabara Cancer Research Institute | Craig S. Richmond, Paraic A. Kenny |
| EPI_ISL_1366665 | LESP Nuevo Leon | Instituto de Diagnostico y Referencia Epidemiologicos (INDRE) | Claudia Wong-Arambula, Abril Rodriguez-Maldonado, Vanessa Rivero-Arredondo, Ariadna Medina-Benitez, Joaquin Quiroz-Mercado, Sergio Rangel-Guerrero, Natividad Cruz-Ortiz, Tatiana Nunez-Garcia, Gisela Barrera-Badillo, Lucia Hernandez-Rivas, Irma Lopez-Martinez, Ernesto Ramirez-Gonzalez. |
| EPI_ISL_1366678 | LESP Morelos | Instituto de Diagnostico y Referencia Epidemiologicos (INDRE) | Claudia Wong-Arambula, Abril Rodriguez-Maldonado, Vanessa Rivero-Arredondo, Ariadna Medina-Benitez, Joaquin Quiroz-Mercado, Sergio Rangel-Guerrero, Natividad Cruz-Ortiz, Tatiana Nunez-Garcia, Gisela Barrera-Badillo, Lucia Hernandez-Rivas, Irma Lopez-Martinez, Ernesto Ramirez-Gonzalez. |
| EPI_ISL_1366745 | OUCRU | OUCRU | Nguyen Van Vinh Chau, Nguyen Thi Thu Hong, Nghiem My Ngoc, Nguyen To Anh, Huynh Trung Trieu, Le Nguyen Truc Nhu, Lam Minh Yen, Ngo Ngoc Quang Minh, Nguyen Thanh Phong, Nguyen Thanh Truong, Le Thi Thu Huong, Tran Nguyen Hoang Tu, Le Manh Hung, Tran Tan Thanh, Nguyen Thanh Dung, Nguyen Tri Dung, Guy Thwaites, Le Van Tan |
| EPI_ISL_1366788 | National Health Laboratory Service, South Africa | KRISP, KZN Research Innovation and Sequencing Platform | Laguda-Akingba O, Giandhari J, Pillay S, Lessells R, Mdlalose K, York D, Khan S, Emmanuel SJ, Tegally H, Wilkinson E, de Oliveira T |
| EPI_ISL_1367678, EPI_ISL_1367682, EPI_ISL_1367697 | Molecular diagnostic unit for viral haemorrhagic fevers and emerging viruses, Bouaké CHU Laboratory | Molecular diagnostic unit for viral haemorrhagic fevers and emerging viruses, Bouaké CHU Laboratory | Chantal Akoua-Koffi, Diané Bamourou, Etilé Anoh, Oby Wayoro, Safiatou Karidioula, Adjaratou Traoré, Soundélé Maïté, Monemo Pacome, Coulibaly Mbegnan, Bamba Fatoumata Touré, Kra Ouffoué, Grit Schubert, Essia Belarbi, Fabian Leendertz |
| EPI_ISL_1370199, EPI_ISL_1370230, EPI_ISL_1370674, EPI_ISL_1371022, EPI_ISL_1371221 | Dutch COVID-19 response team | National Institute for Public Health and the Environment (RIVM) | Adam Meijer, Harry Vennema, Dirk Eggink, Jeroen Cremer, Sharon van den Brink, Bas van der Veer, AnneMarie van den Brandt, Florian Zwagemaker, Dennis Schmitz, Chantal Reusken, on behalf of the national COVID-19 response team |
| EPI_ISL_1371898, EPI_ISL_1371899, EPI_ISL_1371900, EPI_ISL_1371901, EPI_ISL_1371902, EPI_ISL_1371904 | C H DE LA POLYNESIE FRANCAISE | CNR Virus des Infections Respiratoires - France SUD | Antonin Bal, Gregory Destras, Gwendolyne Burfin, Hadrien Regue, Quentin Semanas, Martine Valette, Bruno Lina, Laurence Josset |
| EPI_ISL_1371933 | National Health Laboratory Service, South Africa | KRISP, KZn Research Innovation and Sequencing Platform | Giandhari J, Pillay S, Maslo C, Sitharam L, Lessells R, Mdlalose K, York D, Khan S, Emmanuel SJ, Tegally H, Wilkinson E, de Oliveira T |
| EPI_ISL_1372067 | Microbiology Department , Late Shri Lakhiram Agrawal Memory Hospital & College, Raigarh, Chhattisgarh | Institute of Life Sciences - INSACOG | Sunil K. Raghav, Safal Walia, Arup Ghosh, Atimukta Jha, Amol M. Kanampalliwar, Shifu Aggarwal, Rupesh Dash, Rajeeb Swain, Punit Prasad, INSACOG Consortium, Ajay Parida |
| EPI_ISL_1372369 | HELIX LLC | WHO National Influenza Centre Russian Federation | Andrey Komissarov, Artem Fadeev, Anna Ivanova, Kseniya Komissarova, Dmitry Bazhenov, Tamila Musaeva, Maria Timofeeva, Veronika Eder, Maria Pisareva, Daria Danilenko, Ksenia Safina, Elena Nabieva, Georgii Bazykin, Dmitry Lioznov |
| EPI_ISL_1372714 | Maine Health and Environmental Testing Laboratory | Tewhey Lab, The Jackson Laboratory | Matluk,N., Dewey,H., Iosue,F., Barter,M., Lynch,R., Munger,H. and Tewhey,R. |
| EPI_ISL_1373467 | Clinical Molecular Microbiology Laboratory, UNC Hospitals | Jeremy Wang | Jeremy Wang, Alexander Rubinsteyn, Colleen Rice, Jason Smedberg, Shawn Hawken, Melissa Miller, Corbin Jones, Robert Hagan |
| EPI_ISL_1373557 | Vault Health | Minnesota Department of Health, Public Health Laboratory | Alexandra Lorentz, Jacob Garfin, Matt Plumb, and Xiong Wang |
| EPI_ISL_1373733 | Washington University School of Medicine | Genomics and Discovery, Respiratory Viruses Branch, Division of Viral Diseases, Centers for Disease Control and Prevention | Ying Tao, Jing Zhang, Yan Li, Brian Lynch, Anna Kelleher, Krista Queen, Anna Uehara, Peter Cook, Han Jia Justin Ng, Clinton R. Paden, Haibin Wang, Suxiang Tong |
| EPI_ISL_1373958 | Vanderbilt University Medical Center | Pathogen Discovery, Respiratory Viruses Branch, Division of Viral Diseases, Centers for Disease Control and Prevention | Jing Zhang, Yan Li, Ying Tao, Brian Lynch, Anna Kelleher, Krista Queen, Anna Uehara, Peter Cook, Han Jia Justin Ng, Clinton R. Paden, Haibin Wang, Suxiang Tong |
| EPI_ISL_1379078 | Laboratoire de santé publique du Québec | Laboratoire de santé publique du Québec | Sandrine Moreira, Ioannis Ragoussis, Guillaume Bourque, Jesse Shapiro, Mark Lathrop and Michel Roger on behalf of the CoVSeQ research group (http://covseq.ca/researchgroup) |
| EPI_ISL_1379327 | Hackensack Meridian Health | New York Genome Center | Michael Zody, Andre Corvelo, Dayna M. Oschwald, Samantha Fennessey, Tom Maniatis, Liang Chen, Jose Mediavilla, Marcus Cunningham, Kaelea Composto, Kar Chow, David Perlin, Barry Kreiswirth |
| EPI_ISL_1379392 | HOSPITAL CIUDAD NEILY | Inciensa, Instituto Costarricense de Investigación y Enseñanza en Nutrición y Salud | Francisco Duarte, Hebleen Porras, Claudio Soto-Garita, Estela Cordero, Adriana Godínez & Melany Calderón |
| EPI_ISL_1379418 | HOSPITAL DR. TONY FACIO CASTRO | Inciensa, Instituto Costarricense de Investigación y Enseñanza en Nutrición y Salud | Francisco Duarte, Hebleen Porras, Claudio Soto-Garita, Estela Cordero, Adriana Godínez, Melany Calderón & Tashana Anglin-Williams |
| EPI_ISL_1379426 | HOSPITAL DR. MAX PERALTA JIMENEZ | Inciensa, Instituto Costarricense de Investigación y Enseñanza en Nutrición y Salud | Francisco Duarte, Hebleen Porras, Claudio Soto-Garita, Estela Cordero, Adriana Godínez, Melany Calderón & Mónica Charpentier-Artavia |
| EPI_ISL_1380069, EPI_ISL_1380120, EPI_ISL_1380124, EPI_ISL_1380188, EPI_ISL_1380370, EPI_ISL_1380464 | CSIR-Centre for Cellular and Molecular Biology | CSIR-Centre for Cellular and Molecular Biology-INSACOG | Payel Mukherjee,Pratheusa Maccha,Namami Gaur,Lamuk Zaveri,Tulasi Nagabandi,Purushotham Vodnala,Blessy B John,Viswagithe S L,B Himasri,Sofia Banu,Priya Singh,Archana Bharadwaj Siva,Karthik Bharadwaj Tallapaka,Rakesh K Mishra,Divya Tej Sowpati |
| EPI_ISL_1380515, EPI_ISL_1380608, EPI_ISL_1381041 | KU Leuven, Rega Institute, Clinical and Epidemiological Virology | KU Leuven, Rega Institute, Clinical and Epidemiological Virology | Tony Wawina-Bokalanga, Bert Vanmechelen, Joan Marti-Carerras, Piet Maes |
| EPI_ISL_1381301, EPI_ISL_1381302, EPI_ISL_1381305 | Omics Sciences Laboratory | Omics Sciences Laboratory | Derly Andrade Molina, Rubén Armas González, Gabriel Morey León, Darlyn Amaya, Katheryn Sacheri Viteri, Emily Sulay Saltos Montalvo, Paula Juliana Gavilanes Jarrín, Juan Carlos Fernández Cadena |
| EPI_ISL_1381318, EPI_ISL_1381340, EPI_ISL_1381364 | Laboratory for Respiratory Viruses, Cantacuzino National Military-Medical Institute for Research and Development | Cantacuzino Institute Virology | Luiza Ustea, Nicoleta Paraschiv, Catalina Pascu, Sorin Dinu, Mihaela Lazar |
| EPI_ISL_1381799 | Institute of Virology, Biomedical Research Center of the Slovak Academy of Sciences, Bratislava | Faculty of Natural Sciences, Comenius University, Bratislava | Brona Brejova, Viktoria Cabanova, Kristina Borsova, Viktoria Hodorova, Sabina Fumacova Havlikova, Juraj Kopacek, Martina Lickova, Lubomira Lukacikova, Martina Nebohacova, Monika Slavikova, Tomas Vinar, Jozef Nosek, Boris Klempa |
| EPI_ISL_1381910 | Virginia Division of Consolidated Laboratory Services | Virginia Division of Consolidated Laboratory Services | Virginia DCLS |

| | | | |
|---|---|---|---|
| EPI_ISL_1383243 | South Eastern Area Laboratory Services (SEALS) | NSW Health Pathology - Institute of Clinical Pathology and Medical Research; Westmead Hospital; University of Sydney | CIDM-PH et al. |
| EPI_ISL_1383272, EPI_ISL_1383779 | Laboratoire national de sante, Microbiology, Virology | Laboratoire national de sante, Microbiology, Microbial Genomics Platform | Anke Wienecke-Baldacchino, Catherine Ragimbeau,Jessica Tapp, Fatu Djabi, Lise Pignon, Raoul Salmon, Trung Nguyen Nguyen, Tamir Abdelrahman |
| EPI_ISL_1384461 | Laboratoires d'analyses medicales - Ketterthill | Laboratoire national de sante, Microbiology, Microbial Genomics Platform | Anke Wienecke-Baldacchino, Catherine Ragimbeau,Jessica Tapp, Fatu Djabi, Lise Pignon, Raoul Salmon, Serge Vedy, Caroline Scheiber, Tamir Abdelrahman |
| EPI_ISL_1384594 | Laboratoires Reunis | Laboratoire national de sante, Microbiology, Microbial Genomics Platform | Anke Wienecke-Baldacchino, Catherine Ragimbeau,Jessica Tapp, Fatu Djabi, Lise Pignon, Raoul Salmon, Bernard Weber, Tamir Abdelrahman |
| EPI_ISL_1384811, EPI_ISL_1384812 | PathWest Laboratory Medicine WA | PathWest Laboratory Medicine WA Microbial Surveillance Unit | PathWest Laboratory Medicine WA Microbial Surveillance Unit |
| EPI_ISL_1385784, EPI_ISL_1385786, EPI_ISL_1385789, EPI_ISL_1385791, EPI_ISL_1385794 | Department of Virology | Department of Virology | Massab Umair, Aamer Ikram, Muhammad Salman, Nazish Badar, Sana Tamim, Zaira Rehman, Abdul Ahad, Shannon Whitmer, Melissa Mobley, Austin Leach, Ketan Patel, Joel Montgomery, John Klena |
| EPI_ISL_1385804 | Alfa Diagnostica, Republic of Moldova | ONCOGENE LLC | ONCOGENE LLC |
| EPI_ISL_1385811 | Alfa Diagnostica LLC | ONCOGENE LLC | ONCOGENE LLC |
| EPI_ISL_1386044 | Microbiology Department, Laboratori Clínic Metropolitana Nord. Hospital Universitari Germans Trias i Pujol. | Can Ruti SARS-CoV-2 Sequencing Hub (HUGTiP/IrsiCaixa/IGTP) | Marc Noguera-Julian, Pilar Armengol, Ignacio Blanco, Antoni E Bordoy, Francesc Catala-Moll, Pere-Joan Cardona, Maria Casadellà, Cristina Casañ, Gemma Clara, Bonaventura Clotet, Cristina Esteban, Montserrat Giménez, Mercedes Guerrero, Anna Not, Roger Paredes, Mariona Parera, Verónica Saludes, Alba Sánchez, and Elisa Martró on behalf of the Can Ruti SARS-CoV-2 Sequencing Hub. |
| EPI_ISL_1390900 | Laboratoire de santé publique du Québec | Laboratoire de santé publique du Québec | Sandrine Moreira, Ioannis Ragoussis, Guillaume Bourque, Jesse Shapiro, Mark Lathrop and Michel Roger on behalf of the CoVSeQ research group |
| EPI_ISL_1391703 | Helix/Illumina | Centers for Disease Control and Prevention Division of Viral Diseases, Pathogen Discovery | Peter W. Cook, Dakota Howard, Dhwani Batra, Ben L. Rambo-Martin, Eileen de Feo, Jan Antico, Christine Tran, Matthew Tolentino, Shannon Wickline, Kim Gietzen, Brad Sickler, Jingtao Liu, Eric Allen, Phil Febbo, Summer Galloway, Nicole L. Washington, Simon White, Geraint Levan, Kelly Schiabor Barrett, Elizabeth Cirulli, Alexandre Bolze, Ary Ascencio, Charlotte Rivera-Garcia, Ryan Cho, Jason Nguyen, Sherry Wang, Jimmy Ramirez, Tyler Cassens, Efren Sandoval, Magnus Isaksson, William Lee, David Becker, Marc Laurent, James Lu, Clinton R. Paden, Suxiang Tong, Duncan MacCannell |
| EPI_ISL_1392390 | Quest Diagnostics Incorporated | Centers for Disease Control and Prevention Division of Viral Diseases, Pathogen Discovery | Peter W. Cook, Dakota Howard, Dhwani Batra, Ben L. Rambo-Martin, S. H. Rosenthal, A. Gerasimova, R. M. Kagan, B. Anderson, M. Hua, Y. Liu, L.E. Bernstein, K.E. Livingston, A. Perez, I. A. Shlyakhter, R. V. Rolando, R. Owen, P. Tanpaiboon, F. Lacbawan, Clinton R. Paden, Suxiang Tong, Duncan MacCannell |
| EPI_ISL_1393726 | WSSE Rzeszów LDM | 1. National Institute of Public Health - National Institute of Hygiene; 2. Eurofins Genomics Europe Sequencing GmbH | Wokowicz Tomasz, Zacharczuk Katarzyna, Sadkowska-Todys Magorzata, Gierczyki Rafa, Eurofins Genomics Europe Sequencing Team, ECDC COVID-19 WGS support team |
| EPI_ISL_1393832 | Laboratorium Medyczne LAB-MED | 1. National Institute of Public Health - National Institute of Hygiene; 2. Eurofins Genomics Europe Sequencing GmbH | Wokowicz Tomasz, Zacharczuk Katarzyna, Sadkowska-Todys Magorzata, Gierczyki Rafa, Eurofins Genomics Europe Sequencing Team, ECDC COVID-19 WGS support team |
| EPI_ISL_1395780 | Laboratorio de Virología del Hospital de Niños Dr. Ricardo Gutierrez | Área de Secuenciación del Laboratorio de Virología del Hospital de Niños Dr. Ricardo Gutierrez on behalf of 'Proyecto Argentino Interinstitucional de genomica de SARS-CoV-2' (PAIS Consortium) | Alexay, S; Thomas, G; Medina, C; Labarta, N; Streitenberger, C; Villegas, E; Barreda Frank, M; Grandis, E; Acevedo, ME; Alvarez Lopez, C; Jacques, O; Mistchenko, A; Nabaes Jodar, M; Goya, S; Lusso, S; Acuña, D; Natale, MI; Valinotto, LE; Viegas, M. |
| EPI_ISL_1395782, EPI_ISL_1395783, EPI_ISL_1395784, EPI_ISL_1395785 | Instituto de Investigaciones Biomédicas en Retrovirus y SIDA (INBIRS) | Área de Secuenciación del Laboratorio de Virología del Hospital de Niños Dr. Ricardo Gutierrez on behalf of 'Proyecto Argentino Interinstitucional de genomica de SARS-CoV-2' (PAIS Consortium) | Vanesa Seery; Federico Remes Lenicov; Horacio Salomón; Nabaes Jodar, M; Goya, S; Lusso, S; Acuña, D; Alexay, S; Natale, MI; Valinotto, LE; Viegas, M. |
| EPI_ISL_1395792, EPI_ISL_1395796 | Laboratorio de Virología del Hospital de Niños Dr. Ricardo Gutierrez | Área de Secuenciación del Laboratorio de Virología del Hospital de Niños Dr. Ricardo Gutierrez on behalf of 'Proyecto Argentino Interinstitucional de genomica de SARS-CoV-2' (PAIS Consortium) | Alexay, S; Thomas, G; Medina, C; Labarta, N; Streitenberger, C; Villegas, E; Barreda Frank, M; Grandis, E; Acevedo, ME; Alvarez Lopez, C; Jacques, O; Mistchenko, A; Nabaes Jodar, M; Goya, S; Lusso, S; Acuña, D; Natale, MI; Valinotto, LE; Viegas, M. |
| EPI_ISL_1395798 | Laboratorio del Hospital Interzonal General de Agudos "Evita" | Área de Secuenciación del Laboratorio de Virología del Hospital de Niños Dr. Ricardo Gutierrez on behalf of 'Proyecto Argentino Interinstitucional de genomica de SARS-CoV-2' (PAIS Consortium) | Isabel Desimone; Erica Luczak; Omar Grossi; Lorena Serrano; Rubén Pelagamos; Alejandra Musto; Nabaes Jodar, M; Goya, S; Lusso, S; Acuña, D; Alexay, S; Natale, MI; Valinotto, LE; Viegas, M. |
| EPI_ISL_1395802 | Instituto de Investigaciones Biomédicas en Retrovirus y SIDA (INBIRS) | Área de Secuenciación del Laboratorio de Virología del Hospital de Niños Dr. Ricardo Gutierrez on behalf of 'Proyecto Argentino Interinstitucional de genomica de SARS-CoV-2' (PAIS Consortium) | Vanesa Seery; Federico Remes Lenicov; Horacio Salomón; Nabaes Jodar, M; Goya, S; Lusso, S; Acuña, D; Alexay, S; Natale, MI; Valinotto, LE; Viegas, M. |
| EPI_ISL_1395807 | Laboratorio del Hospital Teresa Germani | Área de Secuenciación del Laboratorio de Virología del Hospital de Niños Dr. Ricardo Gutierrez on behalf of 'Proyecto Argentino Interinstitucional de genomica de SARS-CoV-2' (PAIS Consortium) | Anabel Luzio; Gisela Sauco; Mauricio Spacaventto; Nabaes Jodar, M; Goya, S; Lusso, S; Acuña, D; Alexay, S; Natale, MI; Valinotto, LE; Viegas, M. |
| EPI_ISL_1395825, EPI_ISL_1395832 | Inmunología del Hospital Perrando e Instituto de Medicina Regional de la UNNE | Grupo de Genómica y Bioinformática del Instituto de Investigación de la Cadena Láctea CONICET-INTA on behalf of 'Proyecto Argentino Interinstitucional de genomica de SARS-CoV-2' (PAIS Consortium) | María Delia Foussal, Gerardo Deluca, Natalia Andrea Ayala, María Verónica Gómez, Gustavo Giusiano, Horacio Lucero, Marcelo Marin, Antonieta Cayré, Laura Lescano, Eberhardt, MF, Irazoqui, Amadio, AF |
| EPI_ISL_1395936, EPI_ISL_1395939, EPI_ISL_1395942 | Laboratorio Central de Salud Pública | Grupo de Genómica y Bioinformática del Instituto de Investigación de la Cadena Láctea CONICET-INTA on behalf of 'Proyecto Argentino Interinstitucional de genomica de SARS-CoV-2' (PAIS Consortium) | Natalia Andrea Ayala y María Verónica Gómez, Erica Struss, Esteban Paredes, Antonieta Cayré, Laura Lescano, Eberhardt, MF, Irazoqui, Amadio, AF |
| EPI_ISL_1395990, EPI_ISL_1396002, EPI_ISL_1396029, EPI_ISL_1396040, EPI_ISL_1396046 | Laboratorio Central, Ministerio de Salud Cordoba | Instituto de Patologia Vegetal (CIAP-INTA) on behalf of 'Proyecto Argentino Interinstitucional de genomica de SARS-CoV-2' (PAIS Consortium) | Fernández, FD; Marquez, N.; Debat, HJ.; Re, V.; Pisano, M.B.; Castro, G.; Barbas, G. |
| EPI_ISL_1396114 | Laboratorio de Virus Respiratorios y Neurovirosis. Hospital Señor del Milagro | Instituto de Patologia Vegetal (CIAP-INTA) on behalf of 'Proyecto Argentino Interinstitucional de genomica de SARS-CoV-2' (PAIS Consortium) | Fernández, FD, Marquez, N., Debat, HJ., Irazoqui, M., Amadio, A. Dra. Raskovsky Viviana, Dr. Lavaque Esteban, Dra. Veronica Lesser. Tecnica: Pamela Cajal, Fernanda Aguero. |
| EPI_ISL_1396130, EPI_ISL_1396142 | Laboratorio de la Dirección de Epidemiología | Área de Secuenciación del Laboratorio de Virología del Hospital de Niños Dr. Ricardo Gutierrez on behalf of 'Proyecto Argentino Interinstitucional de genomica de SARS-CoV-2' (PAIS Consortium) | Rechimont, C; Usero, JC; Villasana, M; Bertone, AG; Nabaes Jodar, M; Goya, S; Lusso, S; Acuña, D; Alexay, S; Natale, MI; Valinotto, LE; Viegas, M. |
| EPI_ISL_1396203, EPI_ISL_1396205, EPI_ISL_1396218, EPI_ISL_1396221, EPI_ISL_1396222, EPI_ISL_1396223 | Laboratorio de Virología del Hospital de Niños Dr. Ricardo Gutierrez | Área de Secuenciación del Laboratorio de Virología del Hospital de Niños Dr. Ricardo Gutierrez on behalf of 'Proyecto Argentino Interinstitucional de genomica de SARS-CoV-2' (PAIS Consortium) | Alexay, S; Thomas, G; Medina, C; Labarta, N; Streitenberger, C; Villegas, E; Barreda Frank, M; Grandis, E; Acevedo, ME; Alvarez Lopez, C; Jacques, O; Mistchenko, A; Nabaes Jodar, M; Goya, S; Lusso, S; Acuña, D; Natale, MI; Valinotto, LE; Viegas, M. |

| | | | |
|---|---|---|---|
| EPI_ISL_1396250, EPI_ISL_1396266 | Laboratorio de Virología del Hospital de Niños Dr. Ricardo Gutierrez | Biocódices SA. on behalf of 'Proyecto Argentino Interinstitucional de genomica de SARS-CoV-2' (PAIS Consortium) | Alexay, S; Thomas, G; Medina, C; Labarta, N; Streitenberger, C; Villegas, E; Barreda Frank, M; Grandis, E; Acevedo, ME; Alvarez Lopez, C; Jacques, O; Mistchenko, A; J, Zubrzycki J; Berros, JM; Dopazo, H. |
| EPI_ISL_1396359 | Laboratorio del Hospital Regional Ushuaia Gdor. Ernesto Campos | Nodo de Secuenciación Tierra del Fuego - Hospital Regional Ushuaia - Centro Austral De Investigaciones Científicas - Universidad Nacional De Tierra Del Fuego on behalf of 'Proyecto Argentino Interinstitucional de genomica de SARS-CoV-2' (PAIS Consortium) | Carina Andrea De Roccis, Gabriel Alejandro Castro, Silvana Beatriz Cáceres, Carolina Beatriz Yulan, Manuel Fabian Boutureira, Alejandro Ezequiel Rojas, Fernando Gallego, Santiago Guillermo Ceballos, Cristina Fernanda Nardi, Ivan Dario Gramundi |
| EPI_ISL_1396389, EPI_ISL_1396390 | Laboratorio Salud Publica Mendoza | Nodo de Secuenciación Tierra del Fuego - Hospital Regional Ushuaia - Centro Austral De Investigaciones Científicas - Universidad Nacional De Tierra Del Fuego on behalf of 'Proyecto Argentino Interinstitucional de genomica de SARS-CoV-2' (PAIS Consortium) | Silvia Zerrer, Belén Ortiz, Silvina Denita, Luciana Martinez, Cristian Garay, Fernando Giuliani, Andrea Falaschi, Aguirre Carolina, Alberto Carena, Bosio Lia, Patricia Robledo, Maria Belen Peralta Roca, Carlos Espul, Alejandro Ezequiel Rojas, Fernando Gallego, Santiago Guillermo Ceballos, Cristina Fernanda Nardi, Ivan Dario Gramundi |
| EPI_ISL_1396460 | Centro de Tecnología en Salud Pública de la Universidad Nacional de Rosario | Laboratorio Mixto de Biotecnología Acuática (LMBA) on behalf of 'Proyecto Argentino Interinstitucional de genomica de SARS-CoV-2' (PAIS Consortium) | Joaquín Ezpeleta, Ignacio García Labari, Victoria Posner, Vanina Villanova, Pablo Casal, Pilar Bulacio, Sofía Lavista Llanos, Federico Remes Lenicov, Ana Paletta, Leandro Ciappina, Flavio Spetale, Agustina Cerri, Silvana Spinelli, Elisa Bolatti, Diego Chouhy, María Re, Gastón Viarengo, Ana Cavatorta, Julian Acosta, Javier Murillo, Laura Angelone, Adriana Giri, Silvia Arranz, Elizabeth Tapia (argenTAG) |
| EPI_ISL_1397960 | Department of Virology | Department of Virology | Massab Umair, Aamer Ikram, Muhammad Salman, Nazish Badar, Sana Tamim, Zaira Rehman, Abdul Ahad, Shannon Whitmer, Melissa Mobley, Austin Leach, Ketan Patel, Joel Montgomery, John Klena |
| EPI_ISL_1398511 | National Food and Veterinary Risk Assessment Institute | Vilnius University Hospital Santaros Klinikos, Center of Laboratory Medicine | Gytis Dudas, Ingrida Olendraite, Rimvydas Norvilas, Dovile Ezerskyte, Daniel Naumovas, Ligita Raugaite, Monika Katenaite, Mindaugas Stoskus, Laimonas Griskevicius |
| EPI_ISL_1398930 | Subang Public Health | West Java Health Laboratory; School of Life Sciences and Technology, Institut Teknologi Bandung | Azzania Fibriani, Ema Rahmawati, Ryan Bayusantika Ristandi, Rifky Waluyajati Rachman, Cut Nur Cinthia Alamanda, Isak Solihin, Rini Robiani, Miftahul Faridl, Karimatu Khoirunnisa, Kamila Tania |
| EPI_ISL_1398936 | Purwakarta Public Health | West Java Health Laboratory | Azzania Fibriani, Ema Rahmawati, Ryan Bayusantika Ristandi, Rifky Waluyajati Rachman, Cut Nur Cinthia Alamanda, Isak Solihin, Rini Robiani, Miftahul Faridl, Karimatu Khoirunnisa, Kamila Tania |
| EPI_ISL_1399259 | LESP Jalisco | Instituto de Diagnostico y Referencia Epidemiologicos (INDRE) | Claudia Wong-Arambula, Abril Rodriguez-Maldonado, Vanessa Rivero-Arredondo, Ariadna Medina-Benitez, Joaquin Quiroz-Mercado, Sergio Rangel-Guerrero, Natividad Cruz-Ortiz, Tatiana Nunez-Garcia, Gisela Barrera-Badillo, Lucia Hernandez-Rivas, Irma Lopez-Martinez, Ernesto Ramirez-Gonzalez. |
| EPI_ISL_1399274 | LESP Nayarit | Instituto de Diagnostico y Referencia Epidemiologicos (INDRE) | Claudia Wong-Arambula, Abril Rodriguez-Maldonado, Vanessa Rivero-Arredondo, Ariadna Medina-Benitez, Joaquin Quiroz-Mercado, Sergio Rangel-Guerrero, Natividad Cruz-Ortiz, Tatiana Nunez-Garcia, Gisela Barrera-Badillo, Lucia Hernandez-Rivas, Irma Lopez-Martinez, Ernesto Ramirez-Gonzalez. |
| EPI_ISL_1399750 | Instituto Nacional de Saude (INSA) and BioSystems & Integrative Sciences Institute (BioISI) Genomics Unit, FCUL | Instituto Nacional de Saude (INSA) and BioSystems & Integrative Sciences Institute (BioISI) Genomics Unit, FCUL | Borges et al |
| EPI_ISL_1399905, EPI_ISL_1400081 | Instituto Nacional de Saude (INSA) and Instituto Gulbenkian de Ciencia (IGC) | Instituto Nacional de Saude (INSA) and Instituto Gulbenkian de Ciencia (IGC) | Borges et al |
| EPI_ISL_1400301 | LESP Morelos | Instituto de Diagnostico y Referencia Epidemiologicos (INDRE) | Claudia Wong-Arambula, Abril Rodriguez-Maldonado, Vanessa Rivero-Arredondo, Ariadna Medina-Benitez, Joaquin Quiroz-Mercado, Sergio Rangel-Guerrero, Natividad Cruz-Ortiz, Tatiana Nunez-Garcia, Gisela Barrera-Badillo, Lucia Hernandez-Rivas, Irma Lopez-Martinez, Ernesto Ramirez-Gonzalez. |
| EPI_ISL_1400321 | LESP Guanajuato | Instituto de Diagnostico y Referencia Epidemiologicos (INDRE) | Claudia Wong-Arambula, Abril Rodriguez-Maldonado, Vanessa Rivero-Arredondo, Ariadna Medina-Benitez, Joaquin Quiroz-Mercado, Sergio Rangel-Guerrero, Natividad Cruz-Ortiz, Tatiana Nunez-Garcia, Gisela Barrera-Badillo, Lucia Hernandez-Rivas, Irma Lopez-Martinez, Ernesto Ramirez-Gonzalez. |
| EPI_ISL_1400565 | HELIX LLC | WHO National Influenza Centre Russian Federation | Andrey Komissarov, Artem Fadeev, Anna Ivanova, Kseniya Komissarova, Dmitry Bazhenov, Mikhail Bakaev, Daria Danilenko, Ksenia Safina, Elena Nabieva, Georgii Bazykin, Dmitry Lioznov |
| EPI_ISL_1401568 | Public Health Authority of the Slovak Republic | Laboratory of Genomics and Bioinformatics, Comenius University Science Park | Tatiana Sedláková, Diana Rusáková, Miroslav Böhmer, Anna Giová, Jaroslav Budiš, Tomáš Szemes |
| EPI_ISL_1402279 | Department of Public Health Microbiology Ljubljana, National Laboratory for Health, Environment and Food | Department of Public Health Microbiology Ljubljana, National Laboratory for Health, Environment and Food | Tom Koritnik, José Gonçalves, Martin Bosilj, Katarina Prosenc, Metka Paragi |
| EPI_ISL_1404588, EPI_ISL_1404614 | Biolab Diagnostic Laboratories | Biolab Diagnostic Laboratories | Issa Abu-Dayyeh, Ahmad Tibi, Lama Hussein, Shayma Ali, Badia Saddedin, Eiad Atwa, Amid Abdelnour |
| EPI_ISL_1404758 | Laboratoire de santé publique du Québec | Laboratoire de santé publique du Québec | Sandrine Moreira, Ioannis Ragoussis, Guillaume Bourque, Jesse Shapiro, Mark Lathrop and Michel Roger on behalf of the CoVSeQ research group (http://covseq.ca/researchgroup) |
| EPI_ISL_1404914 | PHV-FSS | PHV-FSS | Son Nguyen |
| EPI_ISL_1406078 | Institute for Medical Research, Infectious Disease Research Centre, National Institutes of Health, Ministry of Health Malaysia | Institute for Medical Research, Infectious Disease Research Centre, National Institutes of Health, Ministry of Health Malaysia | Suppiah J, Kamel K, Mohd Zawawi Z, Ramly N, Robert F, Thayan R |
| EPI_ISL_1406143, EPI_ISL_1406151 | Biolab Diagnostic Laboratories | Biolab Diagnostic Laboratories | Issa Abu-Dayyeh, Ahmad Tibi, Lama Hussein, Shayma Ali, Badia Saddedin, Eiad Atwa, Amid Abdelnour |
| EPI_ISL_1406192, EPI_ISL_1406194 | Institute for Medical Research, Infectious Disease Research Centre, National Institutes of Health, Ministry of Health Malaysia | Institute for Medical Research, Infectious Disease Research Centre, National Institutes of Health, Ministry of Health Malaysia | Suppiah J, Kamel K, Mohd Zawawi Z, Ramly N, Robert F, Thayan R |
| EPI_ISL_1406195 | Biolab Diagnostic Laboratories | Biolab Diagnostic Laboratories | Issa Abu-Dayyeh, Ahmad Tibi, Lama Hussein, Shayma Ali, Badia Saddedin, Eiad Atwa, Amid Abdelnour |
| EPI_ISL_1406242 | Institute for Medical Research, Infectious Disease Research Centre, National Institutes of Health, Ministry of Health Malaysia | Institute for Medical Research, Infectious Disease Research Centre, National Institutes of Health, Ministry of Health Malaysia | Suppiah J, Kamel K, Mohd Zawawi Z, Ramly N, Robert F, Thayan R |
| EPI_ISL_1406394 | Department of Virology | Department of Virology | Massab Umair, Aamer Ikram, Muhammad Salman, Nazish Badar, Sana Tamim, Zaira Rehman, Abdul Ahad, Shannon Whitmer, Melissa Mobley, Austin Leach, Ketan Patel, Joel Montgomery, John Klena |
| EPI_ISL_1406433 | Area of Virology, Serology and Virology Division (SAViD), New South Wales Health Pathology Randwick | Virology Research Laboratory; Area of Virology, Serology and Virology Division (SAViD), New South Wales Health Pathology Randwick | Foster, C.; Au, J.; Ruiz Silva, M.; Wong, M.; Deveson, I.; Bull, R.; Van Hal, S.; Rawlinson, W. |
| EPI_ISL_1407109, EPI_ISL_1407115, EPI_ISL_1407120, EPI_ISL_1407128, EPI_ISL_1407144, EPI_ISL_1407161, EPI_ISL_1407174, EPI_ISL_1407196, EPI_ISL_1407223, EPI_ISL_1407225, EPI_ISL_1407238 | | | |
| see above | National HIV Reference Laboratory, Ministry of Health, Public Health Institute of Malawi | KRISP, KZN Research Innovation and Sequencing Platform | Mvula B, Chilima B, Chiwaula M, Mwangomba W, Panja L, Kasambara W, Auld A, Kim L, Kampira E, Kaba M, Wadonda N, Maida A, Giandhari J, Pillay S, Naidoo Y, Lessells R, Emmanuel SJ, Tegally H, Wilkinson E, de Oliveira T |
| EPI_ISL_1415109 | National Centre For Cell Science | National Centre For Cell Science - INSACOG | Dhiraj Paul, Mitali Inamdar, Sonal Manik Chavan, Mohak P Gujare, Shivang P. Bhanushali, Manoj Kumar Bhat, Ajay Pillai, INSACOG Consortium team, Yogesh Shouche. |
| EPI_ISL_1415293 | Institut National d'hygiène | "Unité Mixte Internationale TransVIHMI (UMI 233 IRD - U1175 | Mounerou SALOU, Christelle BUTEL, Wembo A. HALATOKO,Issaka Maman, Abla A. KONOU, Amivi EHLAN, Adodo SADJI, Kokou TEGUENI,Sidonie |

| | | | |
|---|---|---|---|
| | | INSERM - Université de Montpellier) IRD (Institut de recherche pour le développement)" | A.M.KAGNISSODE, Akoélé SILIADIN, Alassane OURO-MEDELI, Messanh DOUFFAN,Déléma MABA,Sika DOSSIM, Améyo DORKENOO, Mireille PRINCE-DAVID,Anoumou DAGNRA,Laetitia SERRANO,Ahidjo AYOUBA,Eric DELAPORTE, Martine PEETERS |
| EPI_ISL_1415392, EPI_ISL_1415398, EPI_ISL_1415406, EPI_ISL_1415417 | Reference Laboratory of the Ministry of Health | Laboratory of Respiratory Viruses and Measles, Oswaldo Cruz Institute, FIOCRUZ | Paola Resende, Indira Martins, Jessica Edwards, Luciana Appolinario, Fernando Motta, Anna Carolina Paixao, Ana Carolina Mendonca, Alice Sampaio Rocha, Renata Serrano Lopes, Marilda Siqueira on behalf of the Fiocruz COVID-19 Genomic Surveillance Network |
| EPI_ISL_1415425, EPI_ISL_1415428, EPI_ISL_1416165, EPI_ISL_1416192, EPI_ISL_1416222 | Institut National d'hygiène | "Unité Mixte Internationale TransVIHMI (UMI 233 IRD - U1175 INSERM - Université de Montpellier) IRD (Institut de recherche pour le développement)" | Mounerou SALOU, Christelle BUTEL, Wembo A. HALATOKO,Issaka Maman, Abla A. KONOU, Amivi EHLAN, Adodo SADJI, Kokou TEGUENI,Sidonie A.M.KAGNISSODE, Akoélé SILIADIN, Alassane OURO-MEDELI, Messanh DOUFFAN,Déléma MABA,Sika DOSSIM, Améyo DORKENOO, Mireille PRINCE-DAVID,Anoumou DAGNRA,Laetitia SERRANO,Ahidjo AYOUBA,Eric DELAPORTE, Martine PEETERS |
| EPI_ISL_1416277 | Oregon State University TRACE Project | OSU Center for Genome Research and Biocomputing | Oregon State University TRACE Project |
| EPI_ISL_1416322 | PathWest Laboratory Medicine WA | PathWest Laboratory Medicine WA Microbial Surveillance Unit | PathWest Laboratory Medicine WA Microbial Surveillance Unit |
| EPI_ISL_1416386, EPI_ISL_1416397, EPI_ISL_1416489, EPI_ISL_1416666 | Laboratorio Central de Epidemiología (LCE) | Instituto de Biotecnología de la UNAM | Consorcio Mexicano de Vigilancia Genómica (CoViGen-Mex). Authors (in alphabetical order): Julio Elias Alvarado-Yaah, Carlos F. Arias, Santiago Ávila-Ríos, Víctor Hugo Borja-Aburto, Celia Boukadida, Juan Bautista Chale-Dzul , José Antonio Enciso-Moreno, Gloria Elena Espinoza-Ayala, Fernando Fontove-Herrera, Concepción Grajales-Muñiz, Ricardo Grande, Alfredo Herrera-Estrella, Carla Ivón Herrera-Najera, Pavel Isa, Brenda Irasema Maldonado-Meza, Bernardo Martínez-Miguel, Margarita Matías-Florentino, María Guadalupe de Jesús Mireles-Rivera, Gloria María Molina-Salinas, Hector Montoya-Fuentes, José Esteban Muñoz-Medina, José de Jesús Nuñez-Contreras, Alicia Ocaña-Mondragón, Luis Alberto Ochoa-Carrera, Hector Esteban Paz-Juárez, Francisco Pulido, Helen Haydee Fernanda Ramírez-Plascencia, Angel Gustavo Salas-Lais, Jorge Ivan Salinal-Nevarez, Alejandro Sanchez-Flores, Clara Esperanza Santacruz-Tinoco, María Guadalupe Santiago-Mauricio , Nelly Sélem-Mojica, Blanca Taboada , Gloria Vazquez |
| EPI_ISL_1416982 | Outre mer | National Reference Center for Viruses of Respiratory Infections, Institut Pasteur, Paris | Marion Barbet, Sylvie Behillil, Méline Bizard,Frédéric Lemoine,Corinne Maufrais,Christophe Malabat, Angela Brisebarre, Camille Capel, Louise Lefrançois, Etienne Simon-Lorière, Vincent Enouf, Maud Vanpeene, Sylvie van der Werf,GréGoire Potiron |
| EPI_ISL_1417220, EPI_ISL_1417364 | Swedish national genomic surveillance program of SARS-CoV-2 | The Public Health Agency of Sweden | Swedish national genomic surveillance program of SARS-CoV-2 |
| EPI_ISL_1418326 | National Institute of Infectious Diseases-Prof. Dr. Matei Bals Molecular Diagnostics Laboratory | National Institute of Infectious Diseases-Prof. Dr. Matei Bals Molecular Diagnostics Laboratory | Corina Casangiu, Leontina Banica, Marius Surleac, Ovidiu Vlaicu, Ionelia Nicolae, Simona Paraschiv, Dan Otelea |
| EPI_ISL_1419251, EPI_ISL_1419496 | INSACOG-WB | National Institute of Biomedical Genomics - INSACOG | Arindam Maitra, Bhaswati Bandyopadhyay, Nidhan Kumar Biswas, Tamal Ghosh, Sreedhar Chinnaswamy, Ajay Chakraborti, Saumitra Das |
| EPI_ISL_1420668, EPI_ISL_1420672, EPI_ISL_1420695, EPI_ISL_1420706, EPI_ISL_1420741, EPI_ISL_1420753 | Biomedical Research Foundation of the Academy of Athens (BRFAA) | Greek Genome Center, Biomedical Research Foundation of the Academy of Athens (BRFAA) | Emmanouil Athanasiadis, Ioannis Vatsellas, Theodoros Loupis, Katerina Zoi, Dimitrios Thanos |
| EPI_ISL_1423505 | Unidad de Patologia Clinica | Instituto de Diagnostico y Referencia Epidemiologicos (INDRE) | Claudia Wong-Arambula, Abril Rodriguez-Maldonado, Vanessa Rivero-Arredondo, Ariadna Medina-Benitez, Joaquin Quiroz-Mercado, Sergio Rangel-Guerrero, Natividad Cruz-Ortiz, Tatiana Nunez-Garcia, Gisela Barrera-Badillo, Lucia Hernandez-Rivas, Irma Lopez-Martinez, Ernesto Ramirez-Gonzalez. |
| EPI_ISL_1423506, EPI_ISL_1423507, EPI_ISL_1423508, EPI_ISL_1423509 | LESP Chihuahua | Instituto de Diagnostico y Referencia Epidemiologicos (INDRE) | Claudia Wong-Arambula, Abril Rodriguez-Maldonado, Vanessa Rivero-Arredondo, Ariadna Medina-Benitez, Joaquin Quiroz-Mercado, Sergio Rangel-Guerrero, Natividad Cruz-Ortiz, Tatiana Nunez-Garcia, Gisela Barrera-Badillo, Lucia Hernandez-Rivas, Irma Lopez-Martinez, Ernesto Ramirez-Gonzalez. |
| EPI_ISL_1423510 | LESP Ciudad de Mexico | Instituto de Diagnostico y Referencia Epidemiologicos (INDRE) | Claudia Wong-Arambula, Abril Rodriguez-Maldonado, Vanessa Rivero-Arredondo, Ariadna Medina-Benitez, Joaquin Quiroz-Mercado, Sergio Rangel-Guerrero, Natividad Cruz-Ortiz, Tatiana Nunez-Garcia, Gisela Barrera-Badillo, Lucia Hernandez-Rivas, Irma Lopez-Martinez, Ernesto Ramirez-Gonzalez. |
| EPI_ISL_1423511 | LESP Tamaulipas | Instituto de Diagnostico y Referencia Epidemiologicos (INDRE) | Claudia Wong-Arambula, Abril Rodriguez-Maldonado, Vanessa Rivero-Arredondo, Ariadna Medina-Benitez, Joaquin Quiroz-Mercado, Sergio Rangel-Guerrero, Natividad Cruz-Ortiz, Tatiana Nunez-Garcia, Gisela Barrera-Badillo, Lucia Hernandez-Rivas, Irma Lopez-Martinez, Ernesto Ramirez-Gonzalez. |
| EPI_ISL_1423512 | LESP Chihuahua | Instituto de Diagnostico y Referencia Epidemiologicos (INDRE) | Claudia Wong-Arambula, Abril Rodriguez-Maldonado, Vanessa Rivero-Arredondo, Ariadna Medina-Benitez, Joaquin Quiroz-Mercado, Sergio Rangel-Guerrero, Natividad Cruz-Ortiz, Tatiana Nunez-Garcia, Gisela Barrera-Badillo, Lucia Hernandez-Rivas, Irma Lopez-Martinez, Ernesto Ramirez-Gonzalez. |
| EPI_ISL_1423513 | LESP Queretaro | Instituto de Diagnostico y Referencia Epidemiologicos (INDRE) | Claudia Wong-Arambula, Abril Rodriguez-Maldonado, Vanessa Rivero-Arredondo, Ariadna Medina-Benitez, Joaquin Quiroz-Mercado, Sergio Rangel-Guerrero, Natividad Cruz-Ortiz, Tatiana Nunez-Garcia, Gisela Barrera-Badillo, Lucia Hernandez-Rivas, Irma Lopez-Martinez, Ernesto Ramirez-Gonzalez. |
| EPI_ISL_1423514 | LESP Yucatan | Instituto de Diagnostico y Referencia Epidemiologicos (INDRE) | Claudia Wong-Arambula, Abril Rodriguez-Maldonado, Vanessa Rivero-Arredondo, Ariadna Medina-Benitez, Joaquin Quiroz-Mercado, Sergio Rangel-Guerrero, Natividad Cruz-Ortiz, Tatiana Nunez-Garcia, Gisela Barrera-Badillo, Lucia Hernandez-Rivas, Irma Lopez-Martinez, Ernesto Ramirez-Gonzalez. |
| EPI_ISL_1424043 | LESP Guerrero | Instituto de Diagnostico y Referencia Epidemiologicos (INDRE) | Claudia Wong-Arambula, Abril Rodriguez-Maldonado, Vanessa Rivero-Arredondo, Ariadna Medina-Benitez, Joaquin Quiroz-Mercado, Sergio Rangel-Guerrero, Natividad Cruz-Ortiz, Tatiana Nunez-Garcia, Gisela Barrera-Badillo, Lucia Hernandez-Rivas, Irma Lopez-Martinez, Ernesto Ramirez-Gonzalez. |
| EPI_ISL_1424063 | Instituto Nacional de Salud- Dirección de Investigación en Salud Pública | Instituto Nacional de Salud- Dirección de Investigación en Salud Pública | Katherine Laiton-Donato, Carlos Franco-Muñoz, Diego A. Álvarez-Díaz, Mauricio Pacheco, Jhonnatan Reales-González, Diego Andrés Prada, Sheryll Corchuelo, Maria T. Herrera-Sepúlveda, Julian Naizaque, Gerardo Santamaría, Magdalena Wiesner, Martha Lucia Ospina Martínez, Marcela Mercado-Reyes. |
| EPI_ISL_1424433 | Università Federico II - Dipartimento di scienze mediche traslazionali - Napoli | TIGEM | Antonio Grimaldi Patrizia Annunziata Francesco Panariello Michele Cennamo Valentina Bouche Chiara Colantuono Lucio Di Filippo Mariano Fiorenza Anna Manfredi Marcello Salvi Giuseppe Portella Andrea Ballabio Davide Cacchiarelli |
| EPI_ISL_1424519, EPI_ISL_1424520, EPI_ISL_1424521, EPI_ISL_1424522, EPI_ISL_1424523, EPI_ISL_1424535, EPI_ISL_1424564, EPI_ISL_1424566, EPI_ISL_1424576, EPI_ISL_1424589, EPI_ISL_1424599, EPI_ISL_1424607, EPI_ISL_1424624, EPI_ISL_1424625, EPI_ISL_1424627, EPI_ISL_1424632, EPI_ISL_1424633, EPI_ISL_1424634 | | | |
| see above | Queensland Medical Laboratories | Victorian Infectious Diseases Reference Laboratory (VIDRL) and the Melbourne Diagnostic Unit Public Health Laboratory (MDU-PHL) | Palou, T., Vaccher, S., Seemann, T., Sherry, N.L. |
| EPI_ISL_1424680, EPI_ISL_1424835, EPI_ISL_1424841 | Queensland Medical Laboratories | Melbourne Diagnostic Unit Public Health Laboratory (MDU-PHL) | Palou, T., Vaccher, S., Seemann, T., Sherry, N.L. |
| EPI_ISL_1424924 | Clalit Health Services Laboratories, Israel | Stern Lab | Stern Lab |
| EPI_ISL_1425560 | WSSE w Krakowie | 1. National Institute of Public Health - National Institute of Hygiene; 2. Eurofins Genomics Europe Sequencing GmbH | Wokowicz Tomasz, Zaczarczuk Katarzyna, Sadkowska-Todys Magorzata, Gierczyki Rafa, Eurofins Genomics Europe Sequencing Team, ECDC COVID-19 WGS support team |
| EPI_ISL_1426831, EPI_ISL_1430678, EPI_ISL_1430730, EPI_ISL_1430834, EPI_ISL_1431510, EPI_ISL_1432172, EPI_ISL_1432850, EPI_ISL_1432862 | Pathogen Genomics Center, National Institute of Infectious Diseases | Pathogen Genomics Center, National Institute of Infectious Diseases | Tsuyoshi Sekizuka, Kentaro Itokawa, Rina Tanaka, Masanori Hashino, Makoto Kuroda |
| EPI_ISL_1433132, EPI_ISL_1434443 | Institut National d'Hygiène | Unité Mixte Internationale TransVIHMI (UMI 233 IRD - U1175 INSERM - Université de Montpellier) IRD (Institut de recherche pour le développement) | Mounerou SALOU, Christelle BUTEL, Wembo A. HALATOKO,Issaka Maman, Abla A. KONOU, Amivi EHLAN, Adodo SADJI, Kokou TEGUENI,Sidonie A.M.KAGNISSODE, Akoélé SILIADIN, Alassane OURO-MEDELI, Messanh DOUFFAN,Déléma MABA,Sika DOSSIM, Améyo DORKENOO, Mireille PRINCE-DAVID,Anoumou DAGNRA,Laetitia SERRANO,Ahidjo AYOUBA,Eric DELAPORTE, Martine PEETERS |
| EPI_ISL_1437273 | SYNLAB MVZ Leinfelden-Echterdingen | Robert Koch Institute | unknown |

| | | | |
|---|---|---|---|
| EPI_ISL_1437749 | Institut National d'Hygiène | Unité Mixte Internationale TransVIHMI (UMI 233 IRD - U1175 INSERM - Université de Montpellier) IRD (Institut de recherche pour le développement) | Mounerou SALOU, Christelle BUTEL, Wembo A. HALATOKO,Issaka Maman, Abla A. KONOU, Amivi EHLAN, Adodo SADJI, Kokou TEGUENI,Sidonie A.M.KAGNISSODE, Akoélé SILIADIN, Alassane OURO-MEDELI, Messanh DOUFFAN,Déléma MABA,Sika DOSSIM, Améyo DORKENOO, Mireille PRINCE-DAVID,Anoumou DAGNRA,Laetitia SERRANO,Ahidjo AYOUBA,Eric DELAPORTE, Martine PEETERS |
| EPI_ISL_1440087, EPI_ISL_1440112, EPI_ISL_1440118, EPI_ISL_1440298 | KEMRI-Wellcome Trust Research Programme,Kilifi | KEMRI-Wellcome Trust Research Programme,Kilifi | Githinji G.,Mohamed K.S.,deLaurent Z.,Mburu M.W. |
| EPI_ISL_1443001, EPI_ISL_1443002 | Institut National d'hygiène | "Unité Mixte Internationale TransVIHMI (UMI 233 IRD - U1175 INSERM - Université de Montpellier) IRD (Institut de recherche pour le développement)" | Mounerou SALOU, Christelle BUTEL, Wembo A. HALATOKO,Issaka Maman, Abla A. KONOU, Amivi EHLAN, Adodo SADJI, Kokou TEGUENI,Sidonie A.M.KAGNISSODE, Akoélé SILIADIN, Alassane OURO-MEDELI, Messanh DOUFFAN,Déléma MABA,Sika DOSSIM, Améyo DORKENOO, Mireille PRINCE-DAVID,Anoumou DAGNRA,Laetitia SERRANO,Ahidjo AYOUBA,Eric DELAPORTE, Martine PEETERS |
| EPI_ISL_1443153 | TYKS, Kliininen mikrobiologia | Expert Microbiology, National Institute for Health and Welfare | Soile Blomqvist, Jani Halkilahti, Kirsi Liitsola, Haider al-Hello, Päivi Laurila, Teemu Smura, Sari Hannula, Pekka Ellonen, Niina Ikonen, Carita Savolainen-Kopra |
| EPI_ISL_1443196, EPI_ISL_1443197 | Hospital Aliança | Hospital São Rafael - IDOR | Isadora Cristina de Siqueira, Aquiles Assunção Camelier, Elves A.P. Maciel, Margarida Celia L. C. Neves, Carolina Kymie Vasques Nonaka, Karoline Almeida Félix de Sousa, Victor Costa Araujo, Yasmin Santos Freitas Macêdo, Aurea Angelica Paste, Bruno Solano de Freitas Souza, Tiago Gräf |
| EPI_ISL_1443641, EPI_ISL_1443642, EPI_ISL_1443643, EPI_ISL_1443650, EPI_ISL_1443651, EPI_ISL_1443652, EPI_ISL_1443656, EPI_ISL_1443658 | Institute of Microbiology, Universidad San Francisco de Quito | Omics Sciences Laboratory | Derly Andrade Molina, Rubén Armas González, Gabriel Morey León, Darlyn Amaya, Katheryn Sacheri Viteri, Emily Sulay Saltos Montalvo, Paula Juliana Gavilanes Jarrín, Sully Márquez,, Fernanda Zurita, Juan José Guadalupe, Monica Becerra-Wong, Belén Prado-Vivar, Bernardo Gutiérrez, Andrea Cunguan, Nabih Dahik, Dayron Brossad, Patricio Rojas-Silva, Gabriel Trueba, Michelle Grunauer, Verónica Barragán, Paúl Cárdenas, Juan Carlos Fernández Cadena |
| EPI_ISL_1443668, EPI_ISL_1443669, EPI_ISL_1443670, EPI_ISL_1443671, EPI_ISL_1443672, EPI_ISL_1443677, EPI_ISL_1443682 | Omics Sciences Laboratory | Omics Sciences Laboratory | Derly Andrade Molina, Rubén Armas González, Gabriel Morey León, Darlyn Amaya, Katheryn Sacheri Viteri, Emily Sulay Saltos Montalvo, Paula Juliana Gavilanes Jarrín, Juan Carlos Fernández Cadena |
| EPI_ISL_1444238 | Helix/Illumina | Centers for Disease Control and Prevention Division of Viral Diseases, Pathogen Discovery | Peter W. Cook, Dakota Howard, Dhwani Batra, Ben L. Rambo-Martin, Eileen de Feo, Jan Antico, Christine Tran, Matthew Tolentino, Shannon Wickline, Kim Gietzen, Brad Sickler, Jingtao Liu, Eric Allen, Phil Febbo, Summer Galloway, Nicole L. Washington, Simon White, Geraint Levan, Kelly Schiabor Barrett, Elizabeth Cirulli, Alexandre Bolze, Ary Ascencio, Charlotte Rivera-Garcia, Ryan Cho, Jason Nguyen, Sherry Wang, Jimmy Ramirez, Tyler Cassens, Efren Sandoval, Magnus Isaksson, William Lee, David Becker, Marc Laurent, James Lu, Clinton R. Paden, Suxiang Tong, Duncan MacCannell |
| EPI_ISL_1445115 | COMPLEXO HOSPITALAR OURO VERDE DE CAMPINAS | Instituto Butantan / Mendelics | Dimas Tadeu Covas, Sandra Coccuzzo Sampaio, Maria Carolina Elias, José Salvatore Leister Patané, Vincent Louis Viala, Antonio Jorge Martins, Ricardo Haddad, Claudia Renata dos Santos Barros, Elaine Cristina Marqueze, Raul Machado Neto, Debora Botequio Moretti, Bibiana Santos, João Paulo Kitajima, Erika Freitas, David Schlesinger, Simone Kashima, Evandra Strazza Rodrigues, Svetoslav Nanev Slavov, Elaine Vieira dos Santos, Rafael dos Santos Bezerra, Luiz Carlos Junior de Alcantara, Marta Giovanetti, Vagner Fonseca, Flavia Aburjaile, Rodrigo Tocantins Calado. |
| EPI_ISL_1445335, EPI_ISL_1445587 | Aegis Sciences Corporation | Centers for Disease Control and Prevention Division of Viral Diseases, Pathogen Discovery | Dakota Howard, Dhwani Batra, Peter W. Cook, Kara Moser, Adrian Paskey, Jason Caravas, Benjamin Rambo-Martin, Shatavia Morrison, Christopher Gulvick, Scott Sammons, Yvette Unoarumhi, Darlene Wagner, Matthew Schmerer, Cyndi Clark, Patrick Campbell, Rob Case, Vikramsinha Ghorpade, Holly Houdeshell, Ola Kvalvaag, Dillon Nall, Ethan Sanders, Alec Vest, Shaun Westlund, Matthew Hardison, Clinton R. Paden, Duncan MacCannell |
| EPI_ISL_1447306, EPI_ISL_1447307 | National Public Health Center, COVID Laboratory | National Public Health Center, National Biosafety Laboratory | Bernadett Pályi, Zoltán Kis, Nóra Magyar, Judit Henczkó, Dániel Déri, Norbert Solymosi |
| EPI_ISL_1448017, EPI_ISL_1448018, EPI_ISL_1448020 | Laboratory of virology, National center of expertise | RSE "National Center for Biotechnology" and RSE "National Center of Expertise" | Shevtsov Alexandr, Amirgazin Asylulan, Kamalova Dinara, Abdaliyev Askar, Tungushbayev Talgat, Sharipova Saule, Balykbaev Kanat, Ramankulov Yerlan |
| EPI_ISL_1456453, EPI_ISL_1456463, EPI_ISL_1456466, EPI_ISL_1456499, EPI_ISL_1457450, EPI_ISL_1457711 | Dutch COVID-19 response team | National Institute for Public Health and the Environment (RIVM) | Adam Meijer, Harry Vennema, Dirk Eggink, Jeroen Cremer, Sharon van den Brink, Bas van der Veer, AnneMarie van den Brandt, Lisa Wijsman, Kim Freriks, Ryanne Jaarsma, Euníce Then, Jolienke Hardeman, Lynn Aarts, Sanne Bos, Melissa van Tuil, Robert Kohl, Linda van de Nes, Sjoerd Kuiling, James Groot, Florian Zwagemaker, Dennis Schmitz, Annelies Kroneman, Karim Hajji, Chantal Reusken, on behalf of the national COVID-19 response team |
| EPI_ISL_1462855 | Laboratory Corporation of America | Centers for Disease Control and Prevention Division of Viral Diseases, Pathogen Discovery | Dakota Howard, Dhwani Batra, Peter W. Cook, Kara Moser, Adrian Paskey, Jason Caravas, Benjamin Rambo-Martin, Shatavia Morrison, Christopher Gulvick, Scott Sammons, Yvette Unoarumhi, Darlene Wagner, Matthew Schmerer, Minoo Agarwal, Eyad Almasri, Debbie Boles, Ayla Burns, Nuthawin Charoensri, Oren Cohen, Susan Countryman, Mary Ann Cristobal, Bobbi Croy, Suzanne Dale, Hrushikesh Deshmukh, Amanda Douglas, Vincent Drouillon, Marcia Eisenberg, Howard Engler, Rama Ghatti, Prashant Gupta, Susan Hicks, Jake Humphrey, Lax Iyer, Manoj Jain, Mohan Kolli, Brian Krueger, Tim Kuphal, Stanley Letovsky, Michael Levandoski, Craig Lukasik, Jonathan Meltzer, Brian Norvell, Mindy Nye, Scott Parker, Christos Petropoulos, John Pruitt, Steven Ragan, Scott Ryan, Mike Sapeta, Jana Schroth, Suresh Babu Selvaraju, Goran Stevovic, Amanda Suchanek, Andrea Throop, Lyndon Tilson, Thomas Urban, Joe Voshell, Kimberly Wagner, Jonathan Williams, Mary Williamson, Qian Zeng, Tricia Zwiefelhofer, Clinton R. Paden, Duncan MacCannell |
| EPI_ISL_1463468 | Labo Analyses Med | National Reference Center for Viruses of Respiratory Infections, Institut Pasteur, Paris | Marion Barbet, Sylvie Behillil,Frédéric Lemoine,Corinne Maufrais,Christophe Malabat, Méline Bizard, Angela Brisebarre, Camille Capel, Louise Lefrançois, Etienne Simon-Lorière, Vincent Enouf, Maud Vanpeene, Sylvie van der Werf,Nabil Gastli |
| EPI_ISL_1464648, EPI_ISL_1464661 | Laboratório de Virologia - UNIFESP | Laboratory of Respiratory Viruses and Measles, Oswaldo Cruz Institute, FIOCRUZ | Paola Resende, Nancy Belei, Luciana Appolinario, Fernando Motta, Anna Carolina Paixao, Ana Carolina Mendonca, Alice Sampaio Rocha, Renata Serrano Lopes, Marilda Siqueira on behalf of the Fiocruz COVID-19 Genomic Surveillance Network |
| EPI_ISL_1465248 | Laboratorio Central de Saude Publica do Estado do Maranhao (LACEN-MA) | Laboratory of Respiratory Viruses and Measles, Oswaldo Cruz Institute, FIOCRUZ | Paola Resende, Luciana Appolinario, Fernando Motta, Anna Carolina Paixao, Ana Carolina Mendonca, Alice Sampaio Rocha, Renata Serrano Lopes, Lidio Gonçalves Lima Neto, Marilda Siqueira on behalf of the Fiocruz COVID-19 Genomic Surveillance Network |
| EPI_ISL_1469093, EPI_ISL_1469095 | LabPLUS | Institute of Environmental Science and Research (ESR) | Rachel Boyle, SallyAnn Harbison, Olivia Stroeven, Xiaoyun Ren, Matt Storey, Nikki Freed, Muhammad Faisal, Jing Wang, Hermes Perez, Anja Werno, Antje van der Linden, Arlo Upton, Chris Mansell, David Hammer, Dragana Drinkovic, Gary McAuliffe, Hana Sofia Andersson, James Ussher, Jill Sherwood, Josh Freeman, Julia Howard, Juliet Elvy, Mary DeAlmeida, Matt Blakiston, Matthew Rogers, Max Bloomfield, Michael Addidle, Michelle Balm, Sally Roberts, Sarah Jefferies, Sharmini Muttaiyah, Susan Morpeth, Susan Taylor, Timothy Blackmore, Vani Sathyendran, Veronica Playle, Virginia Hope, Erasmus Smit, Lauren Jelly, Olin Silander, Joep de Ligt |
| EPI_ISL_1469116 | Centro de Investigacion en Ciencias de la Salud y Biomedicina | CINVESTAV | Sofia Bernal, Andreu Comas |
| EPI_ISL_1469314, EPI_ISL_1469319, EPI_ISL_1469325, EPI_ISL_1469339, EPI_ISL_1469359 | MRC/UVRI & LSHTM Uganda Research Unit | Where sequence data have been generated and submitted to GISAID | Matthew Cotten, Dan Lule Bugembe, My V.T. Phan, Isaac Sseeewanyana, Patrick Semanda, Susan Nabadda, Pontiano Kaleebu |
| EPI_ISL_1469610 | Diretoria de Vigilância em Saúde | Epiclin | Fernando Hayashi Sant'Anna, Ana Paula Muterle, Janira Prichula, Juliana Comerlato, Carolina Comerlato, Eliana Márcia Da Ros Wendland |
| EPI_ISL_1469685 | HOSPITAL SAO FRANCISCO DE ASSIS | Epiclin | Fernando Hayashi Sant'Anna, Ana Paula Muterle, Janira Prichula, Juliana Comerlato, Carolina Comerlato, Eliana Márcia Da Ros Wendland |
| EPI_ISL_1469778 | Hospital Municipal Getúlio Vargas | Epiclin | Fernando Hayashi Sant'Anna, Ana Paula Muterle, Janira Prichula, Juliana Comerlato, Carolina Comerlato, Eliana Márcia Da Ros Wendland |
| EPI_ISL_1469819 | Diretoria de Vigilância em Saúde | Epiclin | Fernando Hayashi Sant'Anna, Ana Paula Muterle, Janira Prichula, Juliana Comerlato, Carolina Comerlato, Eliana Márcia Da Ros Wendland |
| EPI_ISL_1470040, EPI_ISL_1470056 | Synlab Eesti OÜ | 1. Laboratory of Communicable Diseases (Estonia); 2. Eurofins Genomics Europe Sequencing GmbH | Liidia Dotsenko et al. |
| EPI_ISL_1470494, EPI_ISL_1470516 | Genetica Molecular and Subdepartamento de Virologia ISP Chile | Instituto de Salud Publica de Chile | Javier Tognarelli, Karen Orostica, Barbara Parra, Loredana Arata, Jaime Lagos, Gisselle Barra, Patricia Bustos, Rodrigo Fasce, Andres Castillo, Jorge Fernandez |
| EPI_ISL_1472376 | Department of Health Technology and Informatics, The Hong Kong Polytechnic University | Department of Health Technology and Informatics, The Hong Kong Polytechnic University | Siu,G.K.-H., Lee,L.-K., Leung,K.S.-S., Leung,J.S.-L., Ng,T.T.-L., Chan,C.T.-M., Tam,K.K.-G., Lao,H.-Y., Wong,D.S.-H., Lo,W.-H., Wu,A.K.-L., Yau,M.C.-Y. and Lai,Y.W.-M. |
| EPI_ISL_1472377 | Department of Health Technology and Informatics, The Hong Kong Polytechnic University | Department of Health Technology and Informatics, The Hong Kong Polytechnic University | Siu,G.K.-H., Lee,L.-K., Leung,K.S.-S., Leung,J.S.-L., Ng,T.T.-L.,Chan,C.T.-M., Tam,K.K.-G., Lao,H.-Y., Wong,D.S.-H., Lo,W.-H.,Wu,A.K.-L., Yau,M.C.-Y. and Lai,Y.W.-M. |
| EPI_ISL_1475583 | Oxford Viromics, NDM, University of Oxford; Oxford University | COVID-19 Genomics UK (COG-UK) Consortium | Tanya Golubchik, David Bonsall, George Macintyre, Amy Trebes, Mariateresa de Cesare, Catrin Moore, Alex Mobbs, Anita Justice, Robert Shaw, Monique |

| | | | |
|---|---|---|---|
| | Hospitals; Basingstoke and North Hampshire Hospital | | Andersson, Timothy Peto, Emma Wise, Nathan Moore, Jessica Lynch, Nick Cortes, Matilde Mori, Stephen Kidd, David Buck, John Todd, Christophe Fraser |
| EPI_ISL_1476996 | National Public Health Laboratory, National Centre for Infectious Diseases | National Public Health Laboratory, National Centre for Infectious Diseases | Tze Minn Mak, Zhenyang Zhou, Grace Jie Yin Ngan, Royce Ang, Lin Cui, Raymond Tzer Pin Lin |
| EPI_ISL_1477047 | Sanitary-Epidemiological And Public Health Department Of Tashkent Region, Uzbekistan | Center of Genomics and bioinformatics, Bioinformatics laboratory | Mirzakamol S Ayubov, Zabardast T Buriev, Mukhammadjon H Mirzakhmedov, Abdurakhmon N Yusupov, Shukhrat E Shermatov, Ibrokhim Y Abdurakhmonov. |
| EPI_ISL_1480306 | Helix/Illumina | Centers for Disease Control and Prevention Division of Viral Diseases, Pathogen Discovery | Dakota Howard, Dhwani Batra, Peter W. Cook, Kara Moser, Adrian Paskey, Jason Caravas, Benjamin Rambo-Martin, Shatavia Morrison, Christopher Gulvick, Scott Sammons, Yvette Unoarumhi, Darlene Wagner, Matthew Schmerer, Eileen de Feo, Jan Antico, Christine Tran, Matthew Tolentino, Shannon Wickline, Kim Gietzen, Brad Sickler, Jingtao Liu, Eric Allen, Phil Febbo, Nicole L. Washington, Simon White, Geraint Levan, Kelly Schiabor Barrett, Elizabeth Cirulli, Alexandre Bolze, Ary Ascencio, Charlotte Rivera-Garcia, Ryan Cho, Jason Nguyen, Sherry Wang, Jimmy Ramirez, Tyler Cassens, Efren Sandoval, Magnus Isaksson, William Lee, David Becker, Marc Laurent, James Lu, Clinton R. Paden, Duncan MacCannell |
| EPI_ISL_1482832 | MUSC Molecular Pathology Laboratory | MUSC Molecular Pathology Laboratory | Julie W. Hirschhorn, W. Bailey Glen Jr, Dariusz Pytel, Jaclyn Dunne, Kristen Maurer, Frederick S. Nolte |
| EPI_ISL_1483029 | Public Health Virology-Forensic and Scientific Services (PHV-FSS) | Public Health Virology-Forensic and Scientific Services (PHV-FSS) | Son Nguyen |
| EPI_ISL_1483588 | Alaska State Virology Laboratory | Alaska State Virology Laboratory | Stephanie DeRonde, Elva House, Lisa Smith, Ph.D., Jack Chen, Ph.D. |
| EPI_ISL_1487570 | Lighthouse Lab in Alderley Park | Wellcome Sanger Institute for the COVID-19 Genomics UK (COG-UK) Consortium | Jacquelyn Wynn, Mairead Hyland, The Lighthouse Lab in Alderley Park and Alex Alderton, Roberto Amato, Jeffrey Barrett, Sonia Goncalves, Ewan Harrison, David K. Jackson, Ian Johnston, Dominic Kwiatkowski, Cordelia Langford, John Sillitoe on behalf of the Wellcome Sanger Institute COVID-19 Surveillance Team |
| EPI_ISL_1489534, EPI_ISL_1489570, EPI_ISL_1489574, EPI_ISL_1489594 | Division of Emerging Infectious Diseases, Bureau of Infectious Diseases Diagnosis Control, Korea Disease Control and Prevention Agency | Division of Emerging Infectious Diseases, Bureau of Infectious Diseases Diagnosis Control, Korea Disease Control and Prevention Agency | Ae Kyung Park, Il-Hwan Kim, Heui Man Kim, Jeong-Min Kim, Jeong-Ah Kim, Chae Young Lee, Jin Sun No, Eun-Jin Kim |
| EPI_ISL_1489932 | Laboratory of Virology, National center of expertise | RSE "National Center of Expertise" and RSE "National center for Biotechnology" | Abdaliyev Askar, Tungushbayev Talgat, Sharipova Saule, Shevtsov Alexandr, Amirgazin Asylulan, Kamalova Dinara, Ramankulov Erlan, Balykbaev Kanat |
| EPI_ISL_1490030, EPI_ISL_1490161 | Division of Emerging Infectious Diseases, Bureau of Infectious Diseases Diagnosis Control, Korea Disease Control and Prevention Agency | Division of Emerging Infectious Diseases, Bureau of Infectious Diseases Diagnosis Control, Korea Disease Control and Prevention Agency | Ae Kyung Park, Il-Hwan Kim, Heui Man Kim, Jeong-Min Kim, Jeong-Ah Kim, Chae Young Lee, Jin Sun No, Eun-Jin Kim |
| EPI_ISL_1490666 | The Caribbean Public Health Agency | Carrington Lab, Department of PreClinical Sciences, Faculty of Medical Sciences, The University of the West Indies | Nikita S. D. Sahadeo, Arianne Brown-Jordan, Sarah Hill, Vernie Ramkissoon, Roshan Parasram, Naresh Nandram, Avery Hinds, Jerome Foster, Stanley Giddings, Karla Georges, Marsha Ivey, Rahul Naidu, Risha Singh, SueMin Nathaniel, Rajini Haraksingh, Jaya Jayaraman, Chinna Chinnadurai, Adesh Ramsubhag, Nuno Faria, Oliver Pybus, Christopher Oura, Gabriel Escobar, Christine V. F. Carrington |
| EPI_ISL_1491182 | National Laboratory for Health, Environment and Food, OMM, Maribor | CISLD (Clinical Institute of Special Laboratory Diagnostics), University Children's Hospital, University Medical Center Ljubljana | Jernej Kova, Barbara Jenko Bizjan, Tine Tesovnik, Robert Šket, Katarina Kozmos, Ana Grom, Maruša Debeljak, Marko Pokorn, Tadej Battelino |
| EPI_ISL_1491364 | Hospital Sharp | Microbial Genomics Laboratory | Bruno Gomez-Gil, Julissa Enciso-Ibarra, Alejandra Garcia-Gasca, Daniel Fregoso-Rueda |
| EPI_ISL_1491538, EPI_ISL_1491541 | Institute for Urban Disease Control and Prevention | COVID-19 Network Investigations (CONI) Alliance | Elizabeth Batty, Wasun Chantratita, Thanat Chookajorn, Stefan Fernandez, Angkana Huang, Anthony R. Jones, Khajohn Joonlasak, Chonticha Klungtong, Theerarat Kochakarn, Namfon Kotanan, Krittikorn Kumpornsin, Duangkamon Loesbanluechai, Wudtichai Manasatienkij, Bhakbhoom Panthan, Ekawat Pasomsub, Kingkan Rakmanee, Insee Sensorn, Janjira Thaipadungpanit, Arporn Wangwiwatsin, Treewat Watthanachockchai, Kamolthip Atsawawaranunt, Prayuth Kaewmalang, Amornmas Kongklieng, Pukkaporn Parnwijitkul, Vichan Pawun |
| EPI_ISL_1491561, EPI_ISL_1491565 | Division of Epidemiology | COVID-19 Network Investigations (CONI) Alliance | Elizabeth Batty, Wasun Chantratita, Thanat Chookajorn, Stefan Fernandez, Angkana Huang, Anthony R. Jones, Khajohn Joonlasak, Chonticha Klungtong, Theerarat Kochakarn, Namfon Kotanan, Krittikorn Kumpornsin, Duangkamon Loesbanluechai, Wudtichai Manasatienkij, Bhakbhoom Panthan, Ekawat Pasomsub, Kingkan Rakmanee, Insee Sensorn, Janjira Thaipadungpanit, Arporn Wangwiwatsin, Treewat Watthanachockchai, Atika Awae, Wachirapun Chainontee, Sawanya Chantutanon, Saraiya Chete, Kamonchanok Chuayjan, Kawinna Kerdsalung, Araya Keskesorn, Rusmanira Khwankerd, Rattanapong Kompao, Prapai Kreerat, Sawvanit Lamun, Sitipatimoh Mina, Prangsiri Nalaem, Chodmanee Sukunpun, Chattiwutti Tanabat, Pantila Taweewigyakarn, Juraiwan Ungsathaphornphon, Arthicha Wongkumma |
| EPI_ISL_1491577, EPI_ISL_1491581 | Faculty of Medicine Vajira Hospital | COVID-19 Network Investigations (CONI) Alliance | Elizabeth Batty, Wasun Chantratita, Thanat Chookajorn, Stefan Fernandez, Angkana Huang, Anthony R. Jones, Khajohn Joonlasak, Chonticha Klungtong, Theerarat Kochakarn, Namfon Kotanan, Krittikorn Kumpornsin, Duangkamon Loesbanluechai, Wudtichai Manasatienkij, Bhakbhoom Panthan, Ekawat Pasomsub, Kingkan Rakmanee, Insee Sensorn, Janjira Thaipadungpanit, Arporn Wangwiwatsin, Treewat Watthanachockchai, Sunisa Dongphooyao, Tonsan Hansirisathit, Jakravoot Maneerit, Anan Manomaipiboon, Uraporn Phumisantiphong, Chayanit Phutthanu, Wipawee Thongsopa, Ampan Vimonvattana |
| EPI_ISL_1492070, EPI_ISL_1492238, EPI_ISL_1492290, EPI_ISL_1492334, EPI_ISL_1492341 | National Institute of Public Health | National Institute of Public Health | Helena Jirincova, Jaromira Vecerova, Timotej Suri, Dusan Trnka, Alexander Nagy |
| EPI_ISL_1492644, EPI_ISL_1492645, EPI_ISL_1492647, EPI_ISL_1492648, EPI_ISL_1492649, EPI_ISL_1492650, EPI_ISL_1492651, EPI_ISL_1492652, EPI_ISL_1492657, EPI_ISL_1492659, EPI_ISL_1492660, EPI_ISL_1492664 | | | |
| see above | Laboratorio Central de Salud Publica de Paraguay | Laboratorio Central de Salud Publica de Paraguay | Marta Giovanetti, María José Ortega, Andrea Gómez de la Fuente, Shirley Villalba, Juan Torales, María Liz Gamarra, Vagner Fonseca, Flavia Aburjaile, Talita Adelino, Luiz Carlos Junior Alcantara, Cynthia Vázquez |
| EPI_ISL_1493343 | Wojewodzka Stacja Sanitarno-Epidemiologiczna w Olsztynie, Laboratorium Badan Epidemiologiczno-Klinicznych | Wojewodzka Stacja Sanitarno-Epidemiologiczna w Olsztynie, Laboratorium Badan Epidemiologiczno-Klinicznych | Sylwia Krzetowska, Monika Czerminska, Ewa Liszewska, Tomasz Jakubczak, Marta Lukian, Patryk Bielecki, Aleksandra Kobiatko, Emilia Tarabasz, Paulina Rozycka, Barbara Dolinska |
| EPI_ISL_1494567 | Virginia Division of Consolidated Laboratory Services | Virginia Division of Consolidated Laboratory Services | Virginia DCLS |
| EPI_ISL_1494660 | LESP Guanajuato | Instituto de Diagnostico y Referencia Epidemiologicos (INDRE) | Claudia Wong-Arambula, Abril Rodriguez-Maldonado, Vanessa Rivero-Arredondo, Ariadna Medina-Benitez, Joaquin Quiroz-Mercado, Sergio Rangel-Guerrero, Natividad Cruz-Ortiz, Tatiana Nunez-Garcia, Gisela Barrera-Badillo, Lucia Hernandez-Rivas, Irma Lopez-Martinez, Ernesto Ramirez-Gonzalez. |
| EPI_ISL_1494709 | Laboratorio Aziendale di Microbiologia e Virologia, Azienda Sanitaria dell'Alto Adige | Laboratorio Aziendale di Microbiologia e Virologia, Azienda Sanitaria dell'Alto Adige | Irene Bianconi, Bartolomeo Mobilio Rodriguez, Elisabetta Giacobazzi, Elisa Masi, Stefanie Wieser, Anne Picard, Claudia Volpato, Chiara Cantaloni, Elisabetta Pagani |
| EPI_ISL_1494952 | Comfandi | Instituto Nacional de Salud- Dirección de Investigación en Salud Pública | Katherine Laiton-Donato, Diego A. Álvarez-Díaz, Carlos Franco-Muñoz, Hector Alejandro Ruiz-Moreno, Paola Rojas, Maria T. Herrera-Sepúlveda, Diego Andrés Prada, Jhonnatan Reales-González, Sheryll Corchuelo, Julian Naizaque, Gerardo Santamaría, Sergio Gomez, Lisseth Pardo, Juan Camilo Martinez, Marta Lopez Blanco, Ángela Alarcon Cruz, Diana Malo, Carmen Osorio, Magdalena Wiesner, Martha Lucia Ospina Martinez, Marcela Mercado-Reyes |
| EPI_ISL_1495128, EPI_ISL_1495138 | State Institution «Public Health Center of Ministry of Health of Ukraine» | Robert Koch Institute, ZBS1 Highly Pathogenic Viruses, Berlin, Germany | Annika Brinkmann, Steven Uddin, Roman Rodyna, Liudmyla Chernenko, Janine Michel, Andreas Nitsche, Iryna Demchyshyna |
| EPI_ISL_1495210, EPI_ISL_1495233, EPI_ISL_1495239, EPI_ISL_1495241 | Public Health Authority of the Slovak Republic | Bergthaler laboratory, CeMM Research Center for Molecular Medicine of the Austrian Academy of Sciences | Lukas Endler, Anna Schedl, Fabian Amman, Petr Triska, Thomas Penz, Benedikt Agerer, Maelle Le Moing, Michael Schuster, Bekir Erguner, Jan Laine, Martin Senekowitsch, Christoph Bock, Andreas Bergthaler |
| EPI_ISL_1495730 | Elling group, Institute of Molecular Biotechnology (IMBA) | Bergthaler laboratory, CeMM Research Center for Molecular Medicine of the Austrian Academy of Sciences | Lukas Endler, Anna Schedl, Fabian Amman, Petr Triska, Thomas Penz, Benedikt Agerer, Maelle Le Moing, Michael Schuster, Bekir Erguner, Jan Laine, Martin Senekowitsch, Christoph Bock, Andreas Bergthaler |
| EPI_ISL_1496732, EPI_ISL_1496735, EPI_ISL_1496826 | Department of Virology and Immunology, University of Helsinki and Helsinki University Hospital, Huslab Finland | Department of Virology, Faculty of Medicine, University of Helsinki, Helsinki, Finland | Teemu Smura, Ravi Kant, Phuoc Truong, Hussein Alburkat, Hannimari Kallio-Kokko, Jenni Virtanen, Maija Suvanto, Essi Korhonen, Sari Hannula, Harri Kangas, Hanna Liimatainen, Satu Kurkela, Hanna Jarva, Maija Lappalainen, Pekka Ellonen, Olli Vapalahti |

| | | | |
|---|---|---|---|
| EPI_ISL_1497322, EPI_ISL_1497365, EPI_ISL_1497376, EPI_ISL_1497438, EPI_ISL_1497491, EPI_ISL_1497500 | National Public Health Organization | National Public Health Organization | Kyriaki Tryfinopoulou et al |
| EPI_ISL_1497703 | 1. Glowny Inspektorat Sanitarny. 2. Diagnostyka. Laboratoria Medyczne. | 1. ViroGenetics - BSL3 Laboratory of Virology, Maopolska Centre of Biotechnology, Jagiellonian University; 2. genXone SA, Research & Development Laboratory | Mazur-Panasiuk,N., Grzegorz Nowicki, Gromowski,T., Natalia Drweska-Matelska, Jakub Grabowski, Anna Brylak, Aleksandra Gidlewicz, Karol Szeszko, Maciej Sykulski, ukasz Krych, Kowalski,M., Szulc,P., Sylwia Januszczak, Labaj,P.P., Micha Kaszuba, Pyrc,K. |
| EPI_ISL_1498123, EPI_ISL_1498125 | National Reference Laboratory | National Reference Laboratory | Enatha Mukantwari, Jeanne d'Arc Umuringa, Yvan Butera |
| EPI_ISL_1498130 | Institute of Epidemiology, Disease Control and Research (IEDCR) | Institute for Developing Science and Health Initiatives (ideSHi) | Hassan Afrad, Sadia Rahman, Fidausi Qadri, Tahmina Shirin |
| EPI_ISL_1498379 | The Caribbean Public Health Agency | Carrington Lab, Department of PreClinical Sciences, Faculty of Medical Sciences, The University of the West Indies | Nikita S. D. Sahadeo, Arianne Brown-Jordan, Sarah Hill, Vernie Ramkissoon, Roshan Parasram, Naresh Nandram, Avery Hinds, Jerome Foster, Stanley Giddings, Karla Georges, Marsha Ivey, Rahul Naidu, Risha Singh, SueMin Nathaniel, Rajini Haraksingh, Jaya Jayaraman, Chinna Chinnadurai, Adesh Ramsubhag, Nuno Faria, Oliver Pybus, Christopher Oura, Gabriel Escobar, Christine V. F. Carrington |
| EPI_ISL_1498420, EPI_ISL_1498512, EPI_ISL_1498649 | BioneXt Lab | Laboratoire national de sante, Microbiology, Microbial Genomics Platform | Anke Wienecke-Baldacchino, Catherine Ragimbeau,Jessica Tapp, Fatu Djabi, Lise Pignon, Raoul Salmon, Thibault Ferrandon, Tamir Abdelrahman |
| EPI_ISL_1498840 | National Institute of Public Health | National Institute of Public Health | Helena Jirincova, Jaromira Vecerova, Timotej Suri, Dusan Trnka, Alexander Nagy |
| EPI_ISL_1498952, EPI_ISL_1499006, EPI_ISL_1499016, EPI_ISL_1499058 | Institute of Microbiology and Immunology, Faculty of Medicine, University of Ljubljana | Institute of Microbiology and Immunology, Faculty of Medicine, University of Ljubljana | Alen Sulji, Samo Zakotnik, Tomaž Mark Zorec, Matic Brvar, Doroteja Vlaj, Andraž Celar, Dominika Šturm, Patricija Pozvek, Špela Pleh, Miša Korva, Mario Poljak, Tatjana Avši - Županc |
| EPI_ISL_1499270 | 1. Glowny Inspektorat Sanitarny. 2. Diagnostyka. Laboratoria Medyczne. | 1. ViroGenetics - BSL3 Laboratory of Virology, Maopolska Centre of Biotechnology, Jagiellonian University; 2. genXone SA, Research & Development Laboratory | Mazur-Panasiuk,N., Grzegorz Nowicki, Gromowski,T., Natalia Drweska-Matelska, Jakub Grabowski, Anna Brylak, Aleksandra Gidlewicz, Karol Szeszko, Maciej Sykulski, ukasz Krych, Kowalski,M., Szulc,P., Sylwia Januszczak, Labaj,P.P., Micha Kaszuba, Pyrc,K. |
| EPI_ISL_1499537 | Institute of Public Health of Dubrovnik-Neretva County | Croatian Institute of Public Health | Irena Tabain, Ivana Ferenak |
| EPI_ISL_1500948 | Institut National d'Hygiène (INH) | Unité Mixte Internationale TransVIHMI (UMI 233 IRD - U1175 INSERM - Université de Montpellier)IRD (Institut de recherche pour le développement) | Mounerou SALOU, Christelle BUTEL, Wembo A. HALATOKO,Issaka Maman, Abla A. KONOU, Amivi EHLAN, Adodo SADJI, Kokou TEGUENI,Sidonie A.M.KAGNISSODE, Akoélé SILIADIN, Alassane OURO-MEDELI, Messanh DOUFFAN,Déléma MABA,Sika DOSSIM, Améyo DORKENOO, Mireille PRINCE-DAVID,Anoumou DAGNRA,Laetitia SERRANO,Ahidjo AYOUBA,Eric DELAPORTE, Martine PEETERS |
| EPI_ISL_1501715 | Lurie Children's Hospital of Chicago | Northwestern University - Ozer Lab | Ramon Lorenzo-Redondo, Lacy M. Simons, Taylor J. Dean, Michael G. Ison, Xiaotian, Zheng, William J. Muller, Larry K. Kociolek, Judd F. Hultquist, Egon A. Ozer |
| EPI_ISL_1502156 | Division of Medical Virology, National Health Laboratory Service (NHLS), Tygerberg Hospital / Stellenbosch University | Division of Medical Virology, Stellenbosch University and NHLS Tygerberg Hospital | Susan Engelbrecht, Bronwyn Kleinhans, Gert van Zyl, Wolfgang Preiser |
| EPI_ISL_1502580 | Biochemistry and Molecular Biology Department-Faculty of Medicine, Al-Quds University | Biochemistry and Molecular Biology Department-Faculty of Medicine, Al-Quds University | Nasereddin,A., Ereqat,S., Al-Jawabreh,A., Dumaidi,K. |
| EPI_ISL_1502834 | Hospital Regional Dr. Luis "Chicho" Fábrega | Gorgas Memorial Laboratory of Health Studies | Gonzalez Claudia, Herrera Melissa, Herrera Genarino, Leyda Abrego, Moreno Ambar, Oris Chavarria, Jessica Gondola, Marlenne Castillo, Ortiz Alma, Castillo Jorge, Moreno Brechla, Franco Danilo, Lopez-Verges Sandra, Martinez Alexander |
| EPI_ISL_1502933, EPI_ISL_1502937, EPI_ISL_1502939, EPI_ISL_1502943, EPI_ISL_1502957, EPI_ISL_1503012, EPI_ISL_1503030, EPI_ISL_1503042, EPI_ISL_1503102, EPI_ISL_1503109, EPI_ISL_1503119, EPI_ISL_1503132, EPI_ISL_1503139 | | | |
| see above | Gorgas Memorial Laboratory of Health Studies | Gorgas Memorial Laboratory of Health Studies | Gonzalez Claudia, Leyda Abrego, Moreno Ambar, Oris Chavarria, Jessica Gondola, Marlenne Castillo, Ortiz Alma, Castillo Jorge, Moreno Brechla, Franco Danilo, Lopez-Verges Sandra, Martinez Alexander |
| EPI_ISL_1503171, EPI_ISL_1503205 | National Public Health Organization | National Public Health Organization | Kyriaki Tryfinopoulou et al |
| EPI_ISL_1503485 | Massachusetts State Public Health Laboratory | Massachusetts State Public Health Laboratory | Andrew Lang, Timelia Fink, Glen Gallagher, Sandra Smole |
| EPI_ISL_1508826 | Medical Genetics Laboratory, Regional Centre of Medical Genetics, Emergency County Hospital Craiova | Medical Genetics Laboratory, Regional Centre of Medical Genetics, Emergency County Hospital Craiova | Anca-Lelia (Riza) Costache, Ioana Streata, Stefania Dorobantu, Adina Dragos, Mihai Cucu, Ana-Maria Buga, Razvan Plesea, Andrei Pirvu, Elena Plesea, Monica Cara, Mihai Ioana |
| EPI_ISL_1508895 | Laboratory for HIV and opportunistic infections diagnosis The Republican Research and Practical Center for Epidemiology and Microbiology (RRPCEM) | Laboratory for HIV and opportunistic infections diagnosis The Republican Research and Practical Center for Epidemiology and Microbiology (RRPCEM) | Elena Gasich, Kirill Bulda, Yauhen Sysaliatsin, Leonid Valentovich, Alina Drozd, Nastassia Kabankova, Anatoly Krasko, Vladimir Gorbunov, Alena Mikhalenka, Alexander Kilchevcky |
| EPI_ISL_1508896 | Laboratoire Biolim/FSS/UL | Unité Mixte Internationale TransVIHMI (UMI 233 IRD - U1175 INSERM - Université de Montpellier) IRD (Institut de recherche pour le développement) | Mounerou SALOU, Christelle BUTEL, Wembo A. HALATOKO, Amivi EHLAN, Abla A. KONOU, Issaka Maman,Syntyche DEVATCHAGNI, Adodo SADJI, Kokou TEGUENI,Koku AGBODEKA, Sidonie A.M.KAGNISSODE, Akoélé SILIADIN, Alassane OURO-MEDELI, Messanh DOUFFAN,Déléma MABA,Sika DOSSIM, Améyo DORKENOO, Mireille PRINCE-DAVID,Anoumou DAGNRA, Laetitia SERRANO,Ahidjo AYOUBA, Eric DELAPORTE, Martine PEETERS |
| EPI_ISL_1508945 | Laboratoire Biolim/FSS/UL | Unité Mixte Internationale TransVIHMI (UMI 233 IRD - U1175 INSERM - Université de Montpellier) IRD (Institut de recherche pour le développement) | Mounerou SALOU, Christelle BUTEL, Wembo A. HALATOKO, Amivi EHLAN, Abla A. KONOU, Issaka Maman, Syntyche DEVATCHAGNI, Adodo SADJI, Kokou TEGUENI,Koku AGBODEKA, Sidonie A.M.KAGNISSODE, Akoélé SILIADIN, Alassane OURO-MEDELI, Messanh DOUFFAN,Déléma MABA,Sika DOSSIM, Améyo DORKENOO, Mireille PRINCE-DAVID,Anoumou DAGNRA, Laetitia SERRANO,Ahidjo AYOUBA, Eric DELAPORTE, Martine PEETERS |
| EPI_ISL_1508953 | Institut National d'hygiène | Unité Mixte Internationale TransVIHMI (UMI 233 IRD - U1175 INSERM - Université de Montpellier) IRD (Institut de recherche pour le développement) | Mounerou SALOU, Christelle BUTEL, Wembo A. HALATOKO, Amivi EHLAN, Abla A. KONOU, Issaka Maman,Syntyche DEVATCHAGNI, Adodo SADJI, Kokou TEGUENI,Koku AGBODEKA, Sidonie A.M.KAGNISSODE, Akoélé SILIADIN, Alassane OURO-MEDELI, Messanh DOUFFAN,Déléma MABA,Sika DOSSIM, Améyo DORKENOO, Mireille PRINCE-DAVID,Anoumou DAGNRA, Laetitia SERRANO, Ahidjo AYOUBA, Eric DELAPORTE, Martine PEETERS |
| EPI_ISL_1508960 | Institut National d'hygiène | Unité Mixte Internationale TransVIHMI (UMI 233 IRD - U1175 INSERM - Université de Montpellier) IRD (Institut de recherche pour le développement) | Mounerou SALOU, Christelle BUTEL, Wembo A. HALATOKO, Amivi EHLAN, Abla A. KONOU, Issaka Maman, Syntyche DEVATCHAGNI, Adodo SADJI, Kokou TEGUENI,Koku AGBODEKA, Sidonie A.M.KAGNISSODE, Akoélé SILIADIN, Alassane OURO-MEDELI, Messanh DOUFFAN,Déléma MABA,Sika DOSSIM, Améyo DORKENOO, Mireille PRINCE-DAVID,Anoumou DAGNRA, Laetitia SERRANO,Ahidjo AYOUBA, Eric DELAPORTE, Martine PEETERS |
| EPI_ISL_1508962 | Laboratoire Biolim/FSS/UL | Unité Mixte Internationale TransVIHMI (UMI 233 IRD - U1175 INSERM - Université de Montpellier) IRD (Institut de recherche pour le développement) | Mounerou SALOU, Christelle BUTEL, Wembo A. HALATOKO, Amivi EHLAN, Abla A. KONOU, Issaka Maman, Syntyche DEVATCHAGNI, Adodo SADJI, Kokou TEGUENI,Koku AGBODEKA, Sidonie A.M.KAGNISSODE, Akoélé SILIADIN, Alassane OURO-MEDELI, Messanh DOUFFAN,Déléma MABA,Sika DOSSIM, Améyo DORKENOO, Mireille PRINCE-DAVID,Anoumou DAGNRA, Laetitia SERRANO,Ahidjo AYOUBA, Eric DELAPORTE, Martine PEETERS |
| EPI_ISL_1508998 | Institut National d'hygiène | Unité Mixte Internationale TransVIHMI (UMI 233 IRD - U1175 INSERM - Université de Montpellier) IRD (Institut de recherche pour le développement) | Mounerou SALOU, Christelle BUTEL, Wembo A. HALATOKO,Amivi EHLAN, Abla A. KONOU, Issaka Maman,Syntyche DEVATCHAGNI, Adodo SADJI, Kokou TEGUENI,Koku AGBODEKA, Sidonie A.M.KAGNISSODE, Akoélé SILIADIN, Alassane OURO-MEDELI, Messanh DOUFFAN,Déléma MABA,Sika DOSSIM, Améyo DORKENOO, Mireille PRINCE-DAVID,Anoumou DAGNRA, Laetitia SERRANO,Ahidjo AYOUBA, Eric DELAPORTE, Martine PEETERS |
| EPI_ISL_1509001 | National Institute of Laboratory Medicine and Referral Center | Genomic Research Lab, BCSIR | Tasnim Nafisa, Md. Murshed Hasan Sarkar, Shahina Akter, Abu Sayeed Mohammad Mahmud, Mohammad Samir Uzzaman, Eshrar Osman, Md. Ahasan Habib, Tanjina Akhter Banu, Barna Goswami, Iffat Jahan, Md. Saddam Hossain, Md. Maruf Ahmed Molla, Mahmuda Yeasmin, Asish Kumar Ghosh, Arifa Akram, A. K. M. Shamsuzzaman, Md. Salim Khan |
| EPI_ISL_1509638 | Laboratoire Biolim/FSS/UL | Unité Mixte Internationale TransVIHMI (UMI 233 IRD - U1175 INSERM - Université de Montpellier) IRD (Institut de recherche pour le développement) | Mounerou SALOU, Christelle BUTEL, Wembo A. HALATOKO, Amivi EHLAN, Abla A. KONOU, Issaka Maman, Syntyche DEVATCHAGNI, Adodo SADJI, Kokou TEGUENI,Koku AGBODEKA, Sidonie A.M.KAGNISSODE, Akoélé SILIADIN, Alassane OURO-MEDELI, Messanh DOUFFAN,Déléma MABA,Sika DOSSIM, Améyo DORKENOO, Mireille PRINCE-DAVID,Anoumou DAGNRA, Laetitia SERRANO,Ahidjo AYOUBA, Eric DELAPORTE, Martine PEETERS |
| EPI_ISL_1509848 | National Center of Infectious and Parasitic Diseases | National Center of Infectious and Parasitic Diseases | Alexiev, Ivanov, Korsun, Stoitsova, Philipova, Dimitrova, Grigorova L., Hristova, Donchev, Stoykov, Trifonova, Dobrinov, Grigorova I., Kantardjiev |
| EPI_ISL_1510263 | Idaho Bureau of Laboratories | Idaho Bureau of Laboratories | "R. Beukelman, Matthew Charles Burns, Aimee Ceniseros, Robert L. Voermans, Christopher Ball" |
| EPI_ISL_1510286, EPI_ISL_1510382 | National Institute of Public Health | State Veterinary Institute Prague | Nagy,A;Jirincova,H;Suri,T;Trnka,D;Vecerova,J |
| EPI_ISL_1511126, EPI_ISL_1511131, EPI_ISL_1511135 | Institute for Urban Disease Control and Prevention | COVID-19 Network Investigations (CONI) Alliance | Elizabeth Batty, Wasun Chantratita, Thanat Chookajorn, Stefan Fernandez, Angkana Huang, Anthony R. Jones, Khajohn Joonlasak, Chonticha Klungtong, Theerarat Kochakarn, Namfon Kotanan, Krittikorn Kumpornsin, Duangkamon Loesbanluechai, Wudtichai Manasatienkij, Bhakbhoom Panthan, Ekawat Pasomsub, Kingkan Rakmanee, Insee Sensorn, Janjira Thaipadungpanit, Arporn Wangwiwatsin, Treewat Watthanachockchai, Kamolthip Atsawawaranunt, |

| | | | |
|---|---|---|---|
| | | | Prayuth Kaewmalang, Amornmas Kongklieng, Pukkaporn Parnwijitkul, Vichan Pawun |
| EPI_ISL_1511196 | DPHL | Delaware Public Health Lab | Rebecca Savage |
| EPI_ISL_1511469, EPI_ISL_1511475 | Hospital of the University of Pennsylvania Molecular Pathology Lab | Bushman Lab - University of Pennsylvania | John Everett, Kyle Rodino, Shantan Reddy, Pascha Hokama, Aoife M. Roche, Young Hwang, Abigail Glascock, Scott Sherrill-Mix, Samantha A. Whiteside, Jevon Graham-Wooten, Layla A. Khatib, Ayannah S. Fitzgerald, Arupa Ganguly, Mike Feldman, Brendan Kelly, Ronald G. Collman and Frederic Bushman |
| EPI_ISL_1513154, EPI_ISL_1513219, EPI_ISL_1513383 | Aegis Sciences Corporation | Centers for Disease Control and Prevention Division of Viral Diseases, Pathogen Discovery | Dakota Howard, Dhwani Batra, Peter W. Cook, Kara Moser, Adrian Paskey, Jason Caravas, Benjamin Rambo-Martin, Shatavia Morrison, Christopher Gulvick, Scott Sammons, Yvette Unoarumhi, Darlene Wagner, Matthew Schmerer, Cyndi Clark, Patrick Campbell, Rob Case, Vikramsinha Ghorpade, Holly Houdeshell, Ola Kvalvaag, Dillon Nall, Ethan Sanders, Alec Vest, Shaun Westlund, Matthew Hardison, Clinton R. Paden, Duncan MacCannell |
| EPI_ISL_1516782 | LESP Aguascalientes | Instituto de Diagnostico y Referencia Epidemiologicos (INDRE) | Claudia Wong-Arambula, Abril Rodriguez-Maldonado, Vanessa Rivero-Arredondo, Ariadna Medina-Benitez, Joaquin Quiroz-Mercado, Sergio Rangel-Guerrero, Natividad Cruz-Ortiz, Tatiana Nunez-Garcia, Gisela Barrera-Badillo, Lucia Hernandez-Rivas, Irma Lopez-Martinez, Ernesto Ramirez-Gonzalez. |
| EPI_ISL_1516836, EPI_ISL_1516840, EPI_ISL_1516844, EPI_ISL_1516853 | Botswana Harvard HIV Reference Laboratory | Botswana Harvard HIV Reference Laboratory | Sikhulile Wonderful T. Choga, Dorcas Maruapula, Thongbotho Mphoyakgosi, Boitumelo Zuze, Botshelo Radibe, Legodile Kooepile, David Lawrence, Roger Shapiro, Shahin Lockman, Mosepele Mosepele, Joseph Makhema, Simani Gaseitsiwe |
| EPI_ISL_1516859, EPI_ISL_1516860, EPI_ISL_1516862 | Botswana Harvard HIV Reference Laboratory | Botswana Harvard HIV Reference Laboratory | Sikhulile Dorcas Maruapula, Wonderful T. Choga, Thongbotho Mphoyakgosi, Boitumelo Zuze, Botshelo Radibe, Legodile Kooepile, David Lawrence, Roger Shapiro, Shahin Lockman, Mosepele Mosepele, Joseph Makhema, Simani Gaseitsiwe |
| EPI_ISL_1516883, EPI_ISL_1516884 | Public Health Virology-Forensic and Scientific Services (PHV-FSS) | Public Health Virology-Forensic and Scientific Services (PHV-FSS) | Son Nguyen |
| EPI_ISL_1516885 | PHV-FSS | PHV-FSS | Son Nguyen |
| EPI_ISL_1517393, EPI_ISL_1517398, EPI_ISL_1517403, EPI_ISL_1517409 | Inciensa, Instituto Costarricense de Investigación y Enseñanza en Nutrición y Salud | Inciensa, Instituto Costarricense de Investigación y Enseñanza en Nutrición y Salud | Cristian Peréz-Corrales & Valeria Peralta-Barquero |
| EPI_ISL_1517410 | Inciensa, Instituto Costarricense de Investigación y Enseñanza en Nutrición y Salud | Inciensa, Instituto Costarricense de Investigación y Enseñanza en Nutrición y Salud | Cristian Peréz-Corrales, Valeria Peralta-Barquero & César Cerdas Quesada |
| EPI_ISL_1517425 | Inciensa, Instituto Costarricense de Investigación y Enseñanza en Nutrición y Salud | Inciensa, Instituto Costarricense de Investigación y Enseñanza en Nutrición y Salud | Cristian Peréz-Corrales, Valeria Peralta-Barquero & Salas-Abarca P |
| EPI_ISL_1519361 | National Public Health Laboratory, National Centre for Infectious Diseases | National Public Health Laboratory, National Centre for Infectious Diseases | Tze Minn Mak, Zhenyang Zhou, Grace Jie Yin Ngan, Royce Ang, Lin Cui, Raymond Tzer Pin Lin |
| EPI_ISL_1520866, EPI_ISL_1520868, EPI_ISL_1521070, EPI_ISL_1521073, EPI_ISL_1521080, EPI_ISL_1521145, EPI_ISL_1521161, EPI_ISL_1521170, EPI_ISL_1521321, EPI_ISL_1521322, EPI_ISL_1521324, EPI_ISL_1521839, EPI_ISL_1521845, EPI_ISL_1521867, EPI_ISL_1521875, EPI_ISL_1521876, EPI_ISL_1522076, EPI_ISL_1522125, EPI_ISL_1522126, EPI_ISL_1522127, EPI_ISL_1522128, EPI_ISL_1522177, EPI_ISL_1522181, EPI_ISL_1522188, EPI_ISL_1522193, EPI_ISL_1522194, EPI_ISL_1522198, EPI_ISL_1522199, EPI_ISL_1522201, EPI_ISL_1522202, EPI_ISL_1522205, EPI_ISL_1522211, EPI_ISL_1522215, EPI_ISL_1522217 | | | |
| see above | Dutch COVID-19 response team | National Institute for Public Health and the Environment (RIVM) | Adam Meijer, Harry Vennema, Dirk Eggink, Jeroen Cremer, Sharon van den Brink, Bas van der Veer, AnneMarie van den Brandt, Lisa Wijsman, Kim Freriks, Ryanne Jaarsma, Euníce Then, Jolienke Hardeman, Lynn Aarts, Sanne Bos, Melissa van Tuil, Robert Kohl, Linda van de Nes, Sjoerd Kuiling, James Groot, Florian Zwagemaker, Dennis Schmitz, Annelies Kroneman, Karim Hajji, Chantal Reusken, on behalf of the national COVID-19 response team |
| EPI_ISL_1522598 | Hospital Center Emile Mayrisch | Laboratoire national de sante, Microbiology, Microbial Genomics Platform | Anke Wienecke-Baldacchino, Catherine Ragimbeau,Jessica Tapp, Fatu Djabi, Lise Pignon, Raoul Salmon, Cynthia Oxacelay, Tamir Abdelrahman |
| EPI_ISL_1523084 | Laboratoires d'analyses medicales - Ketterthill | Laboratoire national de sante, Microbiology, Microbial Genomics Platform | Anke Wienecke-Baldacchino, Catherine Ragimbeau,Jessica Tapp, Fatu Djabi, Lise Pignon, Raoul Salmon, Serge Vedy, Caroline Scheiber, Tamir Abdelrahman |
| EPI_ISL_1524117 | Public Health Authority of the Slovak Republic | Laboratory of Genomics and Bioinformatics, Comenius University Science Park | Tatiana Sedláková, Diana Rusáková, Miroslav Böhmer, Anna Giová, Jaroslav Budiš, Tomáš Szemes |
| EPI_ISL_1524330, EPI_ISL_1524344, EPI_ISL_1524347, EPI_ISL_1524353, EPI_ISL_1524381 | Biology Department, College of Science, Al Muthanna University and Public Health Laboratory, Al-Muthanna Health Directorate | Department of Virology, Faculty of Medicine, University of Helsinki, Helsinki, Finland generated and submitted to GISAID | Nihad Al-Rashedi, Hussein Alburkat, Murad Munahi, Alaa Hameed, Ali Jasim, Olli Vapalahti, Tarja Sironen,Teemu Smura |
| EPI_ISL_1524723 | Laboratoire Biolim/FSS/UL | Unité Mixte Internationale TransVIHMI (UMI 233 IRD - U1175 INSERM - Université de Montpellier) IRD (Institut de recherche pour le développement) | Mounerou SALOU, Christelle BUTEL, Wembo A. HALATOKO, Amivi EHLAN, Abla A. KONOU, Issaka Maman, Syntyche DEVATCHAGNI, Adodo SADJI, Kokou TEGUENI,Koku AGBODEKA, Sidonie A.M.KAGNISSODE, Akoélé SILIADIN, Alassane OURO-MEDELI, Messanh DOUFFAN,Déléma MABA,Sika DOSSIM, Améyo DORKENOO, Mireille PRINCE-DAVID,Anoumou DAGNRA, Laetitia SERRANO,Ahidjo AYOUBA, Eric DELAPORTE, Martine PEETERS |
| EPI_ISL_1524877, EPI_ISL_1524885 | Botswana Harvard HIV Reference Laboratory | Botswana Harvard HIV Reference Laboratory | Sikhulile Wonderful T. Choga, Dorcas Maruapula, Thongbotho Mphoyakgosi, Boitumelo Zuze, Botshelo Radibe, Legodile Kooepile, David Lawrence, Roger Shapiro, Shahin Lockman, Mosepele Mosepele, Joseph Makhema, Simani Gaseitsiwe |
| EPI_ISL_1526045 | Quest Diagnostics Incorporated | Centers for Disease Control and Prevention Division of Viral Diseases, Pathogen Discovery | Dakota Howard, Dhwani Batra, Peter W. Cook, Kara Moser, Adrian Paskey, Jason Caravas, Benjamin Rambo-Martin, Shatavia Morrison, Christopher Gulvick, Scott Sammons, Yvette Unoarumhi, Darlene Wagner, Matthew Schmerer, S. H. Rosenthal, A. Gerasimova, R. M. Kagan, B. Anderson, M. Hua, Y. Liu, L.E. Bernstein, K.E. Livingston, A. Perez, I. A. Shlyakhter, R. V. Rolando, R. Owen, P. Tanpaiboon, F. Lacbawan, Clinton R. Paden, Duncan MacCannell |
| EPI_ISL_1526648, EPI_ISL_1526651 | Public Health Authority of the Slovak Republic | Laboratory of Genomics and Bioinformatics, Comenius University Science Park | Tatiana Sedláková, Diana Rusáková, Miroslav Böhmer, Anna Giová, Jaroslav Budiš, Tomáš Szemes |
| EPI_ISL_1527024 | AS Belén Flores | Inciensa, Instituto Costarricense de Investigación y Enseñanza en Nutrición y Salud | Pérez-Corrales C, Barboza-Arguedas E & Centeno-Miranda M |
| EPI_ISL_1527025 | unknown | Inciensa, Instituto Costarricense de Investigación y Enseñanza en Nutrición y Salud | Pérez-Corrales C & Barboza-Arguedas E |
| EPI_ISL_1528197 | Hospital | National Reference Center for Viruses of Respiratory Infections, Institut Pasteur, Paris | Marion Barbet, Sylvie Behillil, Frédéric Lemoine, Corinne Maufrais, Christophe Malabat,Adrien Pain, Méline Bizard, Angela Brisebarre, Camille Capel, Louise Lefrançois, Etienne Simon-Lorière, Vincent Enouf, Maud Vanpeene, Sylvie van der Werf,Sylvaine Bastian |
| EPI_ISL_1530876 | Genome Analysis Center, Kamma Memorial Hospital | Genome Analysis Center, Kamma Memorial Hospital | Hanako Yazawa, Jun Ishii, Satoko Soma, Kaori Watanabe, Takuya Yazawa, Michiaki Masuda, Hiroshi Kamma |
| EPI_ISL_1531458 | US Air Force School of Aerospace Medicine | US Air Force School of Aerospace Medicine | Anthony Fries, Jennifer Meyer, William Gruner, William Buggele, Amanda Javorina, Sarah Purves, Clarise Starr, Elizabeth Macias |
| EPI_ISL_1531816 | Centro de Diagnostico COVID-19 UABC Tijuana | Andersen lab at Scripps Research | SEARCH Alliance San Diego with Idanya Rubi Serafin Higuera, Manuel Sanchez Alavez, Jorge Luis Jimenez Niebla, German Ibarra, Jonathan Vincent Baena, Oscar Efren Zazueta Fierro |
| EPI_ISL_1532183 | Laboratorio de Referencia Nacional de Virus Respiratorio. Instituto Nacional de Salud Perú | Laboratorio de Referencia Nacional de Biotecnología y Biología Molecular. Instituto Nacional de Salud Perú | Carlos Padilla Rojas, Karolyn Vega Chozo, Luis Barcena, Priscila Lope Pari, Omar Caceres Rey, Marco Galarza Perez, Maribel Huaringa Nuñez, Johanna Balbuena Torrez, Henri Bailon Calderon, Nancy Rojas Serrano |
| EPI_ISL_1532238 | LESP Guanajuato | Instituto de Diagnostico y Referencia Epidemiologicos (INDRE) | Claudia Wong-Arambula, Abril Rodriguez-Maldonado, Vanessa Rivero-Arredondo, Ariadna Medina-Benitez, Joaquin Quiroz-Mercado, Sergio Rangel-Guerrero, Natividad Cruz-Ortiz, Tatiana Nunez-Garcia, Gisela Barrera-Badillo, Lucia Hernandez-Rivas, Irma Lopez-Martinez, Ernesto Ramirez-Gonzalez. |
| EPI_ISL_1532246 | LESP Chiapas | Instituto de Diagnostico y Referencia Epidemiologicos (INDRE) | Claudia Wong-Arambula, Abril Rodriguez-Maldonado, Vanessa Rivero-Arredondo, Ariadna Medina-Benitez, Joaquin Quiroz-Mercado, Sergio Rangel-Guerrero, Natividad Cruz-Ortiz, Tatiana Nunez-Garcia, Gisela Barrera-Badillo, Lucia Hernandez-Rivas, Irma Lopez-Martinez, Ernesto Ramirez-Gonzalez. |
| EPI_ISL_1532285 | Oman-National Influenza Center | Biotechnology & OMICs Laboratory | Ahmed Al Harrasi, Aisha Al-Amri, Intisar Al-Shukri, Amal Al-Maani, Sajjad Asaf, Bilal Hussain, Samiya Al-Zadjali, Ahmed N Al-Rawahi, Saqib Bilal, Abdul Latif Khan, Samira Al-Mahruqi, Ahmed Al-Rawahi, Hanan Al-Kindi, Amina Al-Jardani. |
| EPI_ISL_1532287 | Oman-National Influenza Center | Biotechnology & OMICs Laboratory | Intisar Al-Shukri, Ahmed Al Harrasi, Aisha Al-Amri, Amal Al-Maani, Sajjad Asaf, Fahad Al-Balushi, Laila Al-Balushi, Ahmed N Al-Rawahi, Saqib Bilal, Abdul |

| | | | |
|---|---|---|---|
| | | | Latif Khan, Samira Al-Mahruqi, Ahmed Al-Rawahi, Hanan Al-Kindi, Amina Al-Jardani. |
| EPI_ISL_1532293 | Oman-National Influenza Center | Biotechnology & OMICs Laboratory | Aisha Al-Amri, Ahmed Al Harrasi, Intisar Al-Shukri, Amal Al-Maani, Sajjad Asaf, Bilal Hussain, Samiya Al-Zadjali, Ahmed N Al-Rawahi, Saqib Bilal, Abdul Latif Khan, Samira Al-Mahruqi, Ahmed Al-Rawahi, Hanan Al-Kindi, Amina Al-Jardani. |
| EPI_ISL_1532808, EPI_ISL_1532809 | Virology Unit, Institut Pasteur du Cambodge | Virology Unit, Institut Pasteur du Cambodge | Sokhoun Yann, Teyputita Ou, Leakhena Pum, Ly Sovann, Kraing Sidonn, Yi Sengdoeurn, Chin Savuth, Chau Darapheak, Veasna Duong, Erik A Karlsson |
| EPI_ISL_1532854 | Botswana Harvard HIV Reference Laboratory | Botswana Harvard HIV Reference Laboratory | Sikhulile Wonderful T. Choga, Dorcas Maruapula, Thongbotho Mphoyakgosi, Boitumelo Zuze, Botshelo Radibe, Legodile Kooepile, David Lawrence, Roger Shapiro, Shahin Lockman, Mosepele Mosepele, Joseph Makhema, Simani Gaseitsiwe |
| EPI_ISL_1532958 | Lab voor klinische biologie | Lab voor klinische biologie | Marija Janevska, Hannelore Hamerlinck, Bruno Verhasselt |
| EPI_ISL_1533046, EPI_ISL_1533060 | Diagnostic and Research Center of Infectious Diseases, Medical Faculty, Andalas University | Diagnostic and Research Center of Infectious Diseases, Medical Faculty, Andalas University | Andani Eka Putra, Syafrizayanti, Ikhwan R. Sudji, Linosefa, Dede Rahman Agustian, Ayu Novita Trisnawati, Siskalil Fahma, Nia Ayuni Putri, Sekar Asri Tresnaningtyas, SM Rezvi, Fauzul Azhim, Dessy Arisanty, Gestina Aliska, Desmawati, Nita Afriani ,Syandrez Prima Putra, Juane Plantika Menra, Yolani Syaputri, Mutia Lailani |
| EPI_ISL_1533513 | Atlanta VA Medical Center | Genomics and Discovery, Respiratory Viruses Branch, Division of Viral Diseases, Centers for Disease Control and Prevention | Yan Li, Ying Tao, Anna Kelleher, Jing Zhang, Anna Montmayeur, Brian Lynch, Krista Queen, Anna Uehara, Peter Cook, Han Jia Justin Ng, Rachel Marine, Clinton R. Paden, Haibin Wang, Mark Burroughs, Justin Lee, Adam Retchless, Suxiang Tong |
| EPI_ISL_1533822, EPI_ISL_1533829 | Centre for Dengue Research and AICBU, Department of Immunology and Molecular Medicine | Centre for Dengue Research and AICBU, Department of Immunology and Molecular Medicine | Chandima Jeewandara, Deshni Jayathilaka, Dinuka Ariyaratne, Deshan Madhusanka, Diyanath Ranasinghe, Laksiri Gomes, Gathsaurie Neelika Malavige |
| EPI_ISL_1534254 | Groote Schuur Hospital wc GSH | NHLS/UCT | Arash Iranzadeh, Deelan Doolabh, Lynn Tyers, Bruna Galvao, Innocent Mudau, Marvin Hsiao, Kruger Marais, Diana Hardie, Stephen Korsman, Carolyn Williamson |
| EPI_ISL_1534279 | Ocean View CDC wc OCE | NHLS/UCT | Arash Iranzadeh, Deelan Doolabh, Lynn Tyers, Bruna Galvao, Innocent Mudau, Marvin Hsiao, Kruger Marais, Diana Hardie, Stephen Korsman, Carolyn Williamson |
| EPI_ISL_1534292 | Victoria Hospital wc VHW | NHLS/UCT | Arash Iranzadeh, Deelan Doolabh, Lynn Tyers, Bruna Galvao, Innocent Mudau, Marvin Hsiao, Kruger Marais, Diana Hardie, Stephen Korsman, Carolyn Williamson |
| EPI_ISL_1534309 | Groote Schuur Hospital wc GSH | NHLS/UCT | Arash Iranzadeh, Deelan Doolabh, Lynn Tyers, Bruna Galvao, Innocent Mudau, Marvin Hsiao, Kruger Marais, Diana Hardie, Stephen Korsman, Carolyn Williamson |
| EPI_ISL_1534345 | Groote Schuur Hospital wc GSH | NHLS/UCT | Arash Iranzadeh, Deelan Doolabh, Lynn Tyers, Bruna Galvao, Innocent Mudau, Marvin Hsiao, Kruger Marais, Diana Hardie, Stephen Korsman, Emmanuel SJ, Tegally H, de Oliveira T, Carolyn Williamson |
| EPI_ISL_1534355 | CoVid WC Garden Route | NHLS/UCT | Arash Iranzadeh, Deelan Doolabh, Lynn Tyers, Bruna Galvao, Innocent Mudau, Marvin Hsiao, Kruger Marais, Diana Hardie, Stephen Korsman, Emmanuel SJ, Tegally H, de Oliveira T, Carolyn Williamson |
| EPI_ISL_1534438 | New Horizon Clinic wc NZC | NHLS/UCT | Arash Iranzadeh, Deelan Doolabh, Lynn Tyers, Bruna Galvao, Innocent Mudau, Marvin Hsiao, Kruger Marais, Diana Hardie, Stephen Korsman, Emmanuel SJ, Tegally H, de Oliveira T, Carolyn Williamson |
| EPI_ISL_1534497 | Ministry of Health Turkey | Ministry of Health Turkey | Fatma Bayrakdar, Yasemin Cosgun, Suleyman Yalcin, Gulay Korukluoglu |
| EPI_ISL_1534538, EPI_ISL_1534541, EPI_ISL_1534543 | Virology Unit, Institut Pasteur du Cambodge | Virology Unit, Institut Pasteur du Cambodge | Sokhoun Yann, Teyputita Ou, Leakhena Pum, Ly Sovann, Kraing Sidonn, Yi Sengdoeurn, Chin Savuth, Chau Darapheak, Veasna Duong, Erik A Karlsson |
| EPI_ISL_1534666, EPI_ISL_1534673, EPI_ISL_1534685, EPI_ISL_1534688, EPI_ISL_1534689 | Laboratorio de Referencia Nacional de Virus Respiratorio. Instituto Nacional de Salud Perú | Laboratorio de Referencia Nacional de Biotecnología y Biología Molecular. Instituto Nacional de Salud Perú | Carlos Padilla Rojas, Karolyn Vega Chozo, Luis Barcena, Priscila Lope Pari, Omar Caceres Rey, Marco Galarza Perez, Maribel Huaringa Nuñez, Johanna Balbuena Torrez, Henri Bailon Calderon, Nancy Rojas Serrano |
| EPI_ISL_1534690 | BTKLPP kelas 1 Batam | National Institute of Health Research and Development | Hana Apsari Pawestri, Subangkit, Kartika Dewi Puspa, Arie Ardiansyah Nugraha, Hartanti Dian Ikawati, Nelly Puspandari, Krisna Pangesti, Budi Santosa, Ismail, Ida Neni Haryanti, Vivi Setiawaty |
| EPI_ISL_1534699 | Ciputra Hospital Citra Garden City | National Institute of Health Research and Development | Vivi Setiawaty, Hana Apsari Pawestri, Subangkit, Kartika Dewi Puspa, Arie Ardiansyah Nugraha, Hartanti Dian Ikawati, Nelly Puspandari, Krisna Pangesti |
| EPI_ISL_1538423, EPI_ISL_1538426 | Institute for Urban Disease Control and Prevention | COVID-19 Network Investigations (CONI) Alliance | Elizabeth Batty, Wasun Chantratita, Thanat Chookajorn, Stefan Fernandez, Angkana Huang, Anthony R. Jones, Khajohn Joonlasak, Chonticha Klungtong, Theerarat Kochakarn, Namfon Kotanan, Krittikorn Kumpornsin, Duangkamon Loesbanluechai, Wudtichai Manasatienkij, Bhakbhoom Panthan, Ekawat Pasomsub, Kingkan Rakmanee, Insee Sensorn, Janjira Thaipadungpanit, Arporn Wangwiwatsin, Treewat Watthanachockchai, Kamolthip Atsawawaranunt, Prayuth Kaewmalang, Amornmas Kongklieng, Pukkaporn Parnwijitkul, Vichan Pawun |
| EPI_ISL_1538479 | LABORATORIUM BADA KLINICZNYCH WSSE w OPOLU | 1. National Institute of Public Health - National Institute of Hygiene; 2. Eurofins Genomics Europe Sequencing GmbH | Wokowicz Tomasz, Zacharczuk Katarzyna, Sadkowska-Todys Magorzata, Gierczyki Rafa, Eurofins Genomics Europe Sequencing Team, ECDC COVID-19 WGS support team |
| EPI_ISL_1539426 | Colorado Department of Public Health and Environment | Colorado Department of Public Health and Environment | Laura Bankers, Molly C. Hetherington-Rauth, Diana Ir, Shannon Ely, Shannon R. Matzinger, Sarah Elizabeth Totten, Emily A. Travanty |
| EPI_ISL_1540707 | Universidad Tecnica Particular de Loja | Institute of Microbiology, Universidad San Francisco de Quito | Belén Prado-Vivar, Sully Márquez, Juan José Guadalupe, Monica Becerra-Wong, Bernardo Gutiérrez, Paola Dalgo, Fernando Serrano, Katherine Ojeda, David Zuñiga, Melissa Ortega, Raiza Briceño, Luis Flores, Verónica Barragán, Patricio Rojas-Silva, Gabriel Trueba, Michelle Grunauer, Paúl Cárdenas |
| EPI_ISL_1541011, EPI_ISL_1541041 | Genetica Molecular and Subdepartamento de Virologia ISP Chile | Instituto de Salud Publica de Chile | Javier Tognarelli, Karen Orostica, Barbara Parra, Loredana Arata, Jaime Lagos, Gisselle Barra, Patricia Bustos, Rodrigo Fasce, Andres Castillo, Jorge Fernandez |
| EPI_ISL_1543959 | National Public Health Laboratory, National Centre for Infectious Diseases | National Public Health Laboratory, National Centre for Infectious Diseases | Tze Minn Mak, Zhenyang Zhou, Grace Jie Yin Ngan, Royce Ang, Lin Cui, Raymond Tzer Pin Lin |
| EPI_ISL_1544019 | National Centre For Cell Science | National Centre For Cell Science - INSACOG | Dhiraj Paul, Mohak P Gujare, Shivang P. Bhanushali, Mitali Inamdar, Sonal Manik Chavan, Manoj Kumar Bhat, Ajay Pillai, INSACOG Consortium team, Yogesh Shouche |
| EPI_ISL_1545287, EPI_ISL_1545302, EPI_ISL_1545310, EPI_ISL_1545314, EPI_ISL_1545315, EPI_ISL_1545318, EPI_ISL_1545320, EPI_ISL_1545331, EPI_ISL_1545332 | Instituto Nacional de Investigação em Saúde | KRISP, KZN Research Innovation and Sequencing Platform | Morais J, Neto Z, Afonso P, Miranda J, David K, Inglês L, Pereira A, Paulo A Carralero RR Paixão JP, Freitas RH, Mufinda M, Lutucuta S, Giandhari J, Pillay S, Naidoo Y, Emmanuel SJ, Tegally H, Wilkinson E, de Oliveira T |
| EPI_ISL_1547424 | Centre Hospitalier Universitaire Clermont-Ferrand | CHU Clermont-Ferrand, service de virologie | Bisseux Maxime, Mirand Audrey, Combes Patricia, Henquell Cécile |
| EPI_ISL_1547465 | HOPITAL DE VICHY | CHU Clermont-Ferrand, service de virologie | Bisseux Maxime, Mirand Audrey, Combes Patricia, Henquell Cécile |
| EPI_ISL_1547711 | Microbiology Department, Laboratori Clínic Metropolitana Nord. Hospital Universitari Germans Trias i Pujol. | Can Ruti SARS-CoV-2 Sequencing Hub (HUGTiP/IrsiCaixa/IGTP) | Marc Noguera-Julian, Pilar Armengol, Ignacio Blanco, Antoni E Bordoy, Francesc Catala-Moll, Pere-Joan Cardona, Maria Casadellà, Cristina Casañ, Gemma Clara, Bonaventura Clotet, Cristina Esteban, Montserrat Giménez, Mercedes Guerrero, Anna Not, Roger Paredes, Mariona Parera, Verónica Saludes, Alba Sánchez, and Elisa Martró on behalf of the Can Ruti SARS-CoV-2 Sequencing Hub. |
| EPI_ISL_1548116 | Montana Public Health Laboratory | Montana Public Health Laboratory | Joy Ritter, Michelle Mozer, Carrie Biskupiak, Deborah Gibson |
| EPI_ISL_1550920, EPI_ISL_1550921 | National Health Laboratory Service, South Africa | KRISP, KZN Research Innovation and Sequencing Platform | Giandhari J, Pillay S, Naidoo Y, Ramphal U, Lessells R, Mdlalose K, York D, Khan S, Emmanuel SJ, Tegally H, Wilkinson E, de Oliveira T |
| EPI_ISL_1551146 | Aegis Sciences Corporation | Centers for Disease Control and Prevention Division of Viral Diseases, Pathogen Discovery | Dakota Howard, Dhwani Batra, Peter W. Cook, Kara Moser, Adrian Paskey, Jason Caravas, Benjamin Rambo-Martin, Shatavia Morrison, Christopher Gulvick, Scott Sammons, Yvette Unoarumhi, Darlene Wagner, Matthew Schmerer, Cyndi Clark, Patrick Campbell, Rob Case, Vikramsinha Ghorpade, Holly Houdeshell, Ola Kvalvaag, Dillon Nall, Ethan Sanders, Alec Vest, Shaun Westlund, Matthew Hardison, Clinton R. Paden, Duncan MacCannell |
| EPI_ISL_1551655 | Quest Diagnostics Incorporated | Centers for Disease Control and Prevention Division of Viral Diseases, Pathogen Discovery | Peter W. Cook, Dakota Howard, Dhwani Batra, Ben L. Rambo-Martin, S. H. Rosenthal, A. Gerasimova, R. M. Kagan, B. Anderson, M. Hua, Y. Liu, L.E. Bernstein, K.E. Livingston, A. Perez, I. A. Shlyakhter, R. V. Rolando, R. Owen, P. Tanpaiboon, F. Lacbawan, Clinton R. Paden, Suxiang Tong, Duncan MacCannell |

| | | | |
|---|---|---|---|
| EPI_ISL_1553125 | Quest Diagnostics Incorporated | Centers for Disease Control and Prevention Division of Viral Diseases, Pathogen Discovery | Dakota Howard, Dhwani Batra, Peter W. Cook, Kara Moser, Adrian Paskey, Jason Caravas, Benjamin Rambo-Martin, Shatavia Morrison, Christopher Gulvick, Scott Sammons, Yvette Unoarumhi, Darlene Wagner, Matthew Schmerer, S. H. Rosenthal, A. Gerasimova, R. M. Kagan, B. Anderson, M. Hua, Y. Liu, L.E. Bernstein, K.E. Livingston, A. Perez, I. A. Shlyakhter, R. V. Rolando, R. Owen, P. Tanpaiboon, F. Lacbawan, Clinton R. Paden, Duncan MacCannell |
| EPI_ISL_1554344 | Helix/Illumina | Centers for Disease Control and Prevention Division of Viral Diseases, Pathogen Discovery | Dakota Howard, Dhwani Batra, Peter W. Cook, Kara Moser, Adrian Paskey, Jason Caravas, Benjamin Rambo-Martin, Shatavia Morrison, Christopher Gulvick, Scott Sammons, Yvette Unoarumhi, Darlene Wagner, Matthew Schmerer, Eileen de Feo, Jan Antico, Christine Tran, Matthew Tolentino, Shannon Wickline, Kim Gietzen, Brad Sickler, Jingtao Liu, Eric Allen, Phil Febbo, Nicole L. Washington, Simon White, Geraint Levan, Kelly Schiabor Barrett, Elizabeth Cirulli, Alexandre Bolze, Ary Ascencio, Charlotte Rivera-Garcia, Ryan Cho, Jason Nguyen, Sherry Wang, Jimmy Ramirez, Tyler Cassens, Efren Sandoval, Magnus Isaksson, William Lee, David Becker, Marc Laurent, James Lu, Clinton R. Paden, Duncan MacCannell |
| EPI_ISL_1555852 | Fulgent Genetics | Centers for Disease Control and Prevention Division of Viral Diseases, Pathogen Discovery | Dakota Howard, Dhwani Batra, Peter W. Cook, Kara Moser, Adrian Paskey, Jason Caravas, Benjamin Rambo-Martin, Shatavia Morrison, Christopher Gulvick, Scott Sammons, Yvette Unoarumhi, Darlene Wagner, Matthew Schmerer, Harry Gao, Mickey Li, John Gao, Joseph Fierro, Benafsh Sapra, Becky Tsai, Yan Meng, Doreen Ng, James Xie, Clinton R. Paden, Duncan MacCannell |
| EPI_ISL_1557692 | ASL Napoli 1 Centro | AMES Centro Polidiagnostico Strumentale S.r.l. | "Giovanni Savarese, Raffaella Ruggiero, Eloisa Evangelista, Antonella Di Carlo, Luisa Circelli, Luigi D'Amore, Nadia Petrillo, Monica Ianniello, Roberto Sirica,Maurizio D'Amora, Antonio Fico" |
| EPI_ISL_1558828 | LESP Jalisco/UdeG | Instituto de Diagnostico y Referencia Epidemiologicos (INDRE) | Claudia Wong-Arambula, Abril Rodriguez-Maldonado, Vanessa Rivero-Arredondo, Ariadna Medina-Benitez, Joaquin Quiroz-Mercado, Sergio Rangel-Guerrero, Natividad Cruz-Ortiz, Tatiana Nunez-Garcia, Gisela Barrera-Badillo, Lucia Hernandez-Rivas, Irma Lopez-Martinez, Ernesto Ramirez-Gonzalez. |
| EPI_ISL_1562715 | Aegis Sciences Corporation | Centers for Disease Control and Prevention Division of Viral Diseases, Pathogen Discovery | Dakota Howard, Dhwani Batra, Peter W. Cook, Kara Moser, Adrian Paskey, Jason Caravas, Benjamin Rambo-Martin, Shatavia Morrison, Christopher Gulvick, Scott Sammons, Yvette Unoarumhi, Darlene Wagner, Matthew Schmerer, Cyndi Clark, Patrick Campbell, Rob Case, Vikramsinha Ghorpade, Holly Houdeshell, Ola Kvalvaag, Dillon Nall, Ethan Sanders, Alec Vest, Shaun Westlund, Matthew Hardison, Clinton R. Paden, Duncan MacCannell |
| EPI_ISL_1563675 | PHV-FSS | PHV-FSS | Son Nguyen |
| EPI_ISL_1568432 | National Public Health Laboratory, National Centre for Infectious Diseases | National Public Health Laboratory, National Centre for Infectious Diseases | Tze Minn Mak, Zhenyang Zhou, Grace Jie Yin Ngan, Royce Ang, Lin Cui, Raymond Tzer Pin Lin |
| EPI_ISL_1570309 | Eurofins LifeCodexx GmbH | Robert Koch Institute | unknown |
| EPI_ISL_1570317 | UniversitÃ¤tsklinikum Heidelberg | Robert Koch Institute | unknown |
| EPI_ISL_1571815 | Wyoming Public Health Laboratory | Wyoming Public Health Laboratory | Jim Mildenberger, Wanda Manley, Noah Hull, Taylor Fearing, Lynette Gumbleton, Channing Weber, Ashley Norberg, Chayse Rowley, Marley Goetz, Brian Dominguez, Elliot Thomasson, Cari Sloma, and Rob Christensen |
| EPI_ISL_1575423, EPI_ISL_1575431 | Kansas Health and Environmental Lab | Kansas Health and Environmental Lab | Mike Grose, Jonathan Barnell, Ben Olsen, and Phil Adam |
| EPI_ISL_1575943 | Laboratorio di Microbiologia | Laboratorio di Microbiologia | Martinetti Lucchini Gladys, Valeria Spina |
| EPI_ISL_1576638 | Helix/Illumina | Centers for Disease Control and Prevention Division of Viral Diseases, Pathogen Discovery | Dakota Howard, Dhwani Batra, Peter W. Cook, Kara Moser, Adrian Paskey, Jason Caravas, Benjamin Rambo-Martin, Shatavia Morrison, Christopher Gulvick, Scott Sammons, Yvette Unoarumhi, Darlene Wagner, Matthew Schmerer, Eileen de Feo, Jan Antico, Christine Tran, Matthew Tolentino, Shannon Wickline, Kim Gietzen, Brad Sickler, Jingtao Liu, Eric Allen, Phil Febbo, Nicole L. Washington, Simon White, Geraint Levan, Kelly Schiabor Barrett, Elizabeth Cirulli, Alexandre Bolze, Ary Ascencio, Charlotte Rivera-Garcia, Ryan Cho, Jason Nguyen, Sherry Wang, Jimmy Ramirez, Tyler Cassens, Efren Sandoval, Magnus Isaksson, William Lee, David Becker, Marc Laurent, James Lu, Clinton R. Paden, Duncan MacCannell |
| EPI_ISL_1577752, EPI_ISL_1577816, EPI_ISL_1577817 | INHRR | Laboratorio de Virología Molecular | Loureiro CL, Jaspe RC, D Angelo P, Zambrano JL, Rodriguez L, Alarcon V, Delgado M, Aguilar M, Garzaro D, Rangel HR, Pujol FH |
| EPI_ISL_1578115, EPI_ISL_1578116 | Genome Analysis Center, Yamanashi Central Hospital | Genome Analysis Center, Yamanashi Central Hospital | Yosuke Hirotsu |
| EPI_ISL_1578214 | Rhode Island Department of Health | Infectious Disease Program, Broad Institute of Harvard and MIT | Siddle,K.J., Azevedo,K., Miller,A., Adams,G., Pearlman,L., Gladden-Young,A., Lagerborg,K., Rudy,M., DeRuff,K., Carter,A., Normandin,E., Bauer,M., Reilly,S., Tomkins-Tinch,C., Loreth,C., Chaluvadi,S., Lemieux,J.E., Birren,B.W., Sabeti,P.C., Huard,R., King,E., Park,D.J., and MacInnis,B.L. |
| EPI_ISL_1578512 | Institute for Public Health of Zagreb County | Croatian Institute of Public Health | Irena Tabain, Ivana Ferenak |
| EPI_ISL_1579241 | Nebraska Public Health Laboratory | NPHL COVID-19 Response Team | NPHL COVID-19 Response Team |
| EPI_ISL_1581250 | Helix/Illumina | Centers for Disease Control and Prevention Division of Viral Diseases, Pathogen Discovery | Dakota Howard, Dhwani Batra, Peter W. Cook, Kara Moser, Adrian Paskey, Jason Caravas, Benjamin Rambo-Martin, Shatavia Morrison, Christopher Gulvick, Scott Sammons, Yvette Unoarumhi, Darlene Wagner, Matthew Schmerer, Eileen de Feo, Jan Antico, Christine Tran, Matthew Tolentino, Shannon Wickline, Kim Gietzen, Brad Sickler, Jingtao Liu, Eric Allen, Phil Febbo, Nicole L. Washington, Simon White, Geraint Levan, Kelly Schiabor Barrett, Elizabeth Cirulli, Alexandre Bolze, Ary Ascencio, Charlotte Rivera-Garcia, Ryan Cho, Jason Nguyen, Sherry Wang, Jimmy Ramirez, Tyler Cassens, Efren Sandoval, Magnus Isaksson, William Lee, David Becker, Marc Laurent, James Lu, Clinton R. Paden, Duncan MacCannell |
| EPI_ISL_1581645 | National Food and Veterinary Risk Assessment Institute | Vilnius University Hospital Santaros Klinikos, Center of Laboratory Medicine | Rimvydas Norvilas, Daniel Naumovas, Dovile Ezerskyte, Gytis Dudas, Ingrida Olendraite, Ligita Raugaite, Monika Katenaite, Mindaugas Stoskus, Laimonas Griskevicius |
| EPI_ISL_1582396 | Institute of Epidemiology, Disease Control and Research (IEDCR) | Institute for Developing Science and Health Initiatives (ideSHi) | Hassan Afrad, Sadia Rahman, Fidausi Qadri, Tahmina Shirin |
| EPI_ISL_1582412 | Centre for Dengue Research and AICBU, Department of Immunology and Molecular Medicine | Centre for Dengue Research and AICBU, Department of Immunology and Molecular Medicine | Chandima Jeewandara, Deshni Jayathilaka, Dinuka Ariyaratne, Deshan Madhusanka, Diyanath Ranasinghe, Laksiri Gomes, Gathsaurie Neelika Malavige |
| EPI_ISL_1582848, EPI_ISL_1582926 | Hospital Universitari Vall d'Hebron - Vall Hebron Institut de Recerca | Hospital Universitari Vall d'Hebron - Vall Hebron Institut de Recerca | Cristina Andrés, Maria Piñana, Josep F Abril, Damir Garcia-Cehic, Ariadna Rando, Juliana Esperalba, Maria Gema Codina, Carla Castillo, Maria Carmen Martin, Tomàs Pumarola, Josep Quer, Andrés Antón |
| EPI_ISL_1582983 | HOSPITAL DEPARTAMENTAL DE VILLAVICENCIO | Instituto Nacional de Salud- Dirección de Investigación en Salud Pública | Katherine Laiton-Donato, Diego A. Álvarez-Díaz, Carlos Franco-Muñoz, Hector Alejandro Ruiz-Moreno, Paola Rojas, Maria T. Herrera-Sepúlveda, Diego Andrés Prada, Jhonnatan Reales-González, Sheryll Corchuelo, Julian Naizaque, Gerardo Santamaría, Sergio Gomez, Lisseth Pardo, Juan Camilo Martinez, Marta Lopez Blanco, Ángela Alarcon Cruz, Diana Malo, Carmen Osorio, Magdalena Wiesner, Martha Lucia Ospina Martinez, Marcela Mercado-Reyes |
| EPI_ISL_1583252 | Armed Forces Institute of Pathology (AFIP), Dhaka Cantonment | Genomic Research Lab, BCSIR | Md. Murshed Hasan Sarkar, Abu Sayeed Mohammad Mahmud, Mohammad Samir Uzzaman, Eshrar Osman, Md. Ahasan Habib, Shahina Akter, Tanjina Akhter Banu, Barna Goswami, Iffat Jahan, Md. Saddam Hossain, Mohammad Mohi Uddin, Md. Kamrul Islam, Mohammad Mizanur Rahman, Susane Giti, Md. Salim Khan |
| EPI_ISL_1583287 | Klinikum Wels-Grieskirchen | Bergthaler laboratory, CeMM Research Center for Molecular Medicine of the Austrian Academy of Sciences | Lukas Endler, Anna Schedl, Fabian Amman, Petr Triska, Thomas Penz, Benedikt Agerer, Maelle Le Moing, Michael Schuster, Bekir Erguner, Jan Laine, Martin Senekowitsch, Christoph Bock, Andreas Bergthaler |
| EPI_ISL_1583464 | Austrian Agency for Health and Food Safety (AGES) | Bergthaler laboratory, CeMM Research Center for Molecular Medicine of the Austrian Academy of Sciences | Lukas Endler, Anna Schedl, Fabian Amman, Petr Triska, Thomas Penz, Benedikt Agerer, Maelle Le Moing, Michael Schuster, Bekir Erguner, Jan Laine, Martin Senekowitsch, Christoph Bock, Andreas Bergthaler |
| EPI_ISL_1583691, EPI_ISL_1583727 | Central Public Health Laboratory - LACEN -Bahia, Salvador, Brazil | Central Public Health Laboratory - LACEN -Bahia, Salvador, Brazil | Stephane Tosta, Luciana Oliveira, Vanessa Nardy,Patrícia Cajado,Marcela Gómez, Breno Dominguez, Jaqueline Gomes, Vagner Fonseca,Marta Giovanetti,Luiz Alcantara, Felicidade Pereira, Arabela Leal |
| EPI_ISL_1585392, EPI_ISL_1585393, EPI_ISL_1585394, EPI_ISL_1585400, EPI_ISL_1585426 | Centro de Investigación Biomédica del Noreste (CIBIN) | Instituto Nacional de Enfermedades Respiratorias (INER): Centro de Investigación en Enfermedades Infecciosas (CIENI) | Consorcio Mexicano de Vigilancia Genómica (CoViGen-Mex). Authors (in alphabetical order): Julio Elias Alvarado-Yaah, Carlos F. Arias, Santiago Ávila-Ríos, Víctor Hugo Borja-Aburto, Celia Boukadida, Juan Bautista Chale-Dzul, Célida Duque Molina, José Antonio Enciso-Moreno, Gloria Elena Espinosa-Ayala, Fernando Fontove-Herrera, Victor Eduardo Garcia-Arias, Concepción Grajales-Muñiz, Ricardo Grande, Alfredo Herrera-Estrella, Carla Ivón Herrera-Najera, Pavel Isa, Brenda Irasema Maldonado-Meza, Bernardo Martínez-Miguel, Margarita Matías-Florentino, María Guadalupe de Jesús Mireles-Rivera, Gloria María Molina-Salinas, Hector Montoya-Fuentes, José Esteban Muñoz-Medina, José de Jesús Nuñez-Contreras, Alicia Ocaña-Mondragón, Luis Alberto Ochoa-Carrera, Hector Esteban Paz-Juárez, Francisco Pulido, Helen Haydee Fernanda Ramírez-Plascencia, Angel |

| | | | |
|---|---|---|---|
| | | | Gustavo Salas-Lais, Alejandro Sanchez-Flores, Clara Esperanza Santacruz-Tinoco, María Guadalupe Santiago-Mauricio, Nelly Sélem-Mojica, Blanca Taboada, Gloria Vazquez |
| EPI_ISL_1585428, EPI_ISL_1585430, EPI_ISL_1585434, EPI_ISL_1585444 | Unidad de Investigación Médica de Yucatán (UIMY) | Instituto Nacional de Enfermedades Respiratorias (INER): Centro de Investigación en Enfermedades Infecciosas (CIENI) | Consorcio Mexicano de Vigilancia Genómica (CoViGen-Mex). Authors (in alphabetical order): Julio Elias Alvarado-Yaah, Carlos F. Arias, Santiago Ávila-Ríos, Víctor Hugo Borja-Aburto, Celia Boukadida, Juan Bautista Chale-Dzul, Célida Duque Molina, José Antonio Enciso-Moreno, Gloria Elena Espinosa-Ayala, Fernando Fontove-Herrera, Victor Eduardo Garcia-Arias, Concepción Grajales-Muñiz, Ricardo Grande, Alfredo Herrera-Estrella, Carla Ivón Herrera-Najera, Pavel Isa, Brenda Irasema Maldonado-Meza, Bernardo Martínez-Miguel, Margarita Matías-Florentino, María Guadalupe de Jesús Mireles-Rivera, Gloria María Molina-Salinas, Hector Montoya-Fuentes, José Esteban Muñoz-Medina, José de Jesús Nuñez-Contreras, Alicia Ocaña-Mondragón, Luis Alberto Ochoa-Carrera, Hector Esteban Paz-Juárez, Francisco Pulido, Helen Haydee Fernanda Ramírez-Plascencia, Angel Gustavo Salas-Lais, Alejandro Sanchez-Flores, Clara Esperanza Santacruz-Tinoco, María Guadalupe Santiago-Mauricio, Nelly Sélem-Mojica, Blanca Taboada, Gloria Vazquez |
| EPI_ISL_1585480, EPI_ISL_1585499 | Centro de Investigación Biomédica de Occidente (CIBO) | Instituto Nacional de Enfermedades Respiratorias (INER): Centro de Investigación en Enfermedades Infecciosas (CIENI) | Consorcio Mexicano de Vigilancia Genómica (CoViGen-Mex). Authors (in alphabetical order): Julio Elias Alvarado-Yaah, Carlos F. Arias, Santiago Ávila-Ríos, Víctor Hugo Borja-Aburto, Celia Boukadida, Juan Bautista Chale-Dzul, Célida Duque Molina, José Antonio Enciso-Moreno, Gloria Elena Espinosa-Ayala, Fernando Fontove-Herrera, Victor Eduardo Garcia-Arias, Concepción Grajales-Muñiz, Ricardo Grande, Alfredo Herrera-Estrella, Carla Ivón Herrera-Najera, Pavel Isa, Brenda Irasema Maldonado-Meza, Bernardo Martínez-Miguel, Margarita Matías-Florentino, María Guadalupe de Jesús Mireles-Rivera, Gloria María Molina-Salinas, Hector Montoya-Fuentes, José Esteban Muñoz-Medina, José de Jesús Nuñez-Contreras, Alicia Ocaña-Mondragón, Luis Alberto Ochoa-Carrera, Hector Esteban Paz-Juárez, Francisco Pulido, Helen Haydee Fernanda Ramírez-Plascencia, Angel Gustavo Salas-Lais, Alejandro Sanchez-Flores, Clara Esperanza Santacruz-Tinoco, María Guadalupe Santiago-Mauricio, Nelly Sélem-Mojica, Blanca Taboada, Gloria Vazquez |
| EPI_ISL_1585583, EPI_ISL_1585620, EPI_ISL_1585647, EPI_ISL_1585658 | Laboratorio Central de Epidemiología (LCE) | Instituto Nacional de Enfermedades Respiratorias (INER): Centro de Investigación en Enfermedades Infecciosas (CIENI) | Consorcio Mexicano de Vigilancia Genómica (CoViGen-Mex). Authors (in alphabetical order): Julio Elias Alvarado-Yaah, Carlos F. Arias, Santiago Ávila-Ríos, Víctor Hugo Borja-Aburto, Celia Boukadida, Juan Bautista Chale-Dzul, Célida Duque Molina, José Antonio Enciso-Moreno, Gloria Elena Espinosa-Ayala, Fernando Fontove-Herrera, Victor Eduardo Garcia-Arias, Concepción Grajales-Muñiz, Ricardo Grande, Alfredo Herrera-Estrella, Carla Ivón Herrera-Najera, Pavel Isa, Brenda Irasema Maldonado-Meza, Bernardo Martínez-Miguel, Margarita Matías-Florentino, María Guadalupe de Jesús Mireles-Rivera, Gloria María Molina-Salinas, Hector Montoya-Fuentes, José Esteban Muñoz-Medina, José de Jesús Nuñez-Contreras, Alicia Ocaña-Mondragón, Luis Alberto Ochoa-Carrera, Hector Esteban Paz-Juárez, Francisco Pulido, Helen Haydee Fernanda Ramírez-Plascencia, Angel Gustavo Salas-Lais, Alejandro Sanchez-Flores, Clara Esperanza Santacruz-Tinoco, María Guadalupe Santiago-Mauricio, Nelly Sélem-Mojica, Blanca Taboada, Gloria Vazquez |
| EPI_ISL_1585917 | Instituto Nacional de Medicina Genomica | Instituto Nacional de Medicina Genomica | Hidalgo-Miranda A, Mendoza-Vargas A, Reyes-Grajeda JP, Cedro-Tanda A, Alcaraz N, Gonzalez-Barrera D, Rangel-DeLeon D, Miranda-Ortiz H, Rosas-Escobar P, Canseco Mendez JC, Ramirez-Vega O, Munguia-Garza P, Garcia-Cardenas FJ, Gonzalez-Woge MA, Herrera-Montalvo LA |
| EPI_ISL_1587503, EPI_ISL_1587511, EPI_ISL_1587529, EPI_ISL_1587531, EPI_ISL_1587539, EPI_ISL_1587542 | Division of Emerging Infectious Diseases, Bureau of Infectious Diseases Diagnosis Control, Korea Disease Control and Prevention Agency | Division of Emerging Infectious Diseases, Bureau of Infectious Diseases Diagnosis Control, Korea Disease Control and Prevention Agency | Ae Kyung Park, Il-Hwan Kim, Heui Man Kim, Jeong-Min Kim, Jeong-Ah Kim, Chae Young Lee, Jin Sun No, Eun-Jin Kim |
| EPI_ISL_1587580, EPI_ISL_1587582 | Genome Analysis Center, Yamanashi Central Hospital | Genome Analysis Center, Yamanashi Central Hospital | Yosuke Hirotsu |
| EPI_ISL_1587755, EPI_ISL_1588030 | Infinity Biologix | Centers for Disease Control and Prevention Division of Viral Diseases, Pathogen Discovery | Dakota Howard, Dhwani Batra, Peter W. Cook, Kara Moser, Adrian Paskey, Jason Caravas, Benjamin Rambo-Martin, Shatavia Morrison, Christopher Gulvick, Scott Sammons, Yvette Unoarumhi, Darlene Wagner, Matthew Schmerer, Christian Bixby, Yihe Wang, Jonathan Schultz, Chirayu Goswami, Russ Hager, Robin Grimwood, Clinton R. Paden, Duncan MacCannell |
| EPI_ISL_1588073, EPI_ISL_1588074 | NB-Hôpital Georges L. Dumont | National Microbiology Laboratory (NML) | Anna Majer, Shari Tyson, Grace Seo, Philip Mabon, Elsie Grudeski, Rhiannon Huzarewich, Russell Mandes, Anneliese Landgraff, Jennifer Tanner, Natalie Knox, Morag Graham, Gary Van Domselaar, Richard Garceau, Guillaume Desnoyers, Nathalie Bastien, Yan Li, Timothy Booth, Darian Hole, Madison Chapel, Kirsten Biggar, CanCOGeN's metadata curation team, Public Health Agency of Canada CanCOGeN team |
| EPI_ISL_1588899, EPI_ISL_1589003, EPI_ISL_1589004, EPI_ISL_1589115 | Caribbean Public Health Agency | Carrington Lab, Department of PreClinical Sciences, Faculty of Medical Sciences, The University of the West Indies | Nikita S. D. Sahadeo, Arianne Brown-Jordan, Sarah Hill, Vernie Ramkissoon, Roshan Parasram, Naresh Nandram, Avery Hinds, Jerome Foster, Stanley Giddings, Karla Georges, Marsha Ivey, Rahul Naidu, Risha Singh, SueMin Nathaniel, Rajini Haraksingh, Jaya Jayaraman, Chinna Chinnadurai, Adesh Ramsubhag, Nuno Faria, Oliver Pybus, Christopher Oura, Gabriel Escobar, Christine V. F. Carrington |
| EPI_ISL_1591097, EPI_ISL_1591098 | NAMRU-6 | Pathogen Discovery, Respiratory Viruses Branch, Division of Viral Diseases, Centers for Disease Control and Prevention | Yan Li, Ying Tao, Anna Kelleher, Jing Zhang, Anna Montmayeur, Brian Lynch, Krista Queen, Anna Uehara, Peter Cook, Han Jia Justin Ng, Rachel Marine, Clinton R. Paden, Haibin Wang, Mark Burroughs, Justin Lee, Adam Retchless, Suxiang Tong |
| EPI_ISL_1591356, EPI_ISL_1591422, EPI_ISL_1591438 | Division of Medical Virology, National Health Laboratory Service (NHLS), Tygerberg Hospital / Stellenbosch University | Division of Medical Virology, Stellenbosch University and NHLS Tygerberg Hospital | Susan Engelbrecht, Bronwyn Kleinhans, Gert van Zyl, Wolfgang Preiser |
| EPI_ISL_1591718 | Caribbean Public Health Agency | Carrington Lab, Department of PreClinical Sciences, Faculty of Medical Sciences, The University of the West Indies | Nikita S. D. Sahadeo, Arianne Brown-Jordan, Sarah Hill, Vernie Ramkissoon, Roshan Parasram, Naresh Nandram, Avery Hinds, Jerome Foster, Stanley Giddings, Karla Georges, Marsha Ivey, Rahul Naidu, Risha Singh, SueMin Nathaniel, Rajini Haraksingh, Jaya Jayaraman, Chinna Chinnadurai, Adesh Ramsubhag, Nuno Faria, Oliver Pybus, Christopher Oura, Gabriel Escobar, Christine V. F. Carrington |
| EPI_ISL_1593728 | NAMRU-6 | Pathogen Discovery, Respiratory Viruses Branch, Division of Viral Diseases, Centers for Disease Control and Prevention | Yan Li, Ying Tao, Anna Kelleher, Jing Zhang, Anna Montmayeur, Brian Lynch, Krista Queen, Anna Uehara, Peter Cook, Han Jia Justin Ng, Rachel Marine, Clinton R. Paden, Haibin Wang, Mark Burroughs, Justin Lee, Adam Retchless, Suxiang Tong |
| EPI_ISL_1593785 | Wyoming Public Health Laboratory | Wyoming Public Health Laboratory | Jim Mildenberger, Wanda Manley, Noah Hull, Taylor Fearing, Lynette Gumbleton, Channing Weber, Ashley Norberg, Chayse Rowley, Marley Goetz, Brian Dominguez, Elliot Thomasson, Sam Britz, Cari Sloma, and Rob Christensen |
| EPI_ISL_1593853 | Institute for Developing Science and Health Initiatives (ideSHi) | Institute for Developing Science and Health Initiatives (ideSHi) | Hassan Afrad, Sadia Rahman, Fidausi Qadri, Tahmina Shirin |
| EPI_ISL_1593856 | Armed Forces Institute of Pathology (AFIP), Dhaka Cantonment | Genomic Research Lab, BCSIR | Mohammad Mizanur Rahman, Md. Murshed Hasan Sarkar, Mohammad Samir Uzzaman, Eshrar Osman, Md. Ahasan Habib, Shahina Akter, Tanjina Akhter Banu, Abu Sayeed Mohammad Mahmud, Barna Goswami, Iffat Jahan, Md. Saddam Hossain, Mohammad Mohi Uddin, Md. Kamrul Islam, Susane Giti, Md. Salim Khan |
| EPI_ISL_1593946 | Missouri State Public Health Laboratory | Missouri State Public Health Laboratory | Matthew Sinn, Joshua Barry, Ashley New |
| EPI_ISL_1594149, EPI_ISL_1594166, EPI_ISL_1594241, EPI_ISL_1594253 | MB-Cadham Provincial laboratory | National Microbiology Laboratory (NML) | Anna Majer, Shari Tyson, Grace Seo, Philip Mabon, Elsie Grudeski, Rhiannon Huzarewich, Russell Mandes, Anneliese Landgraff, Jennifer Tanner, Natalie Knox, Morag Graham, Gary Van Domselaar, Paul Van Caeseele, Jared Bullard, David Alexander, Kerry Dust, Nathalie Bastien, Yan Li, Timothy Booth, Darian Hole, Madison Chapel, Kirsten Biggar, CanCOGeN's metadata curation team, Public Health Agency of Canada CanCOGeN team |
| EPI_ISL_1594355 | Center for Laboratory Medicine St. Gallen | Center for Laboratory Medicine St. Gallen | Yannick Gerth |
| EPI_ISL_1595604 | Centro de Investigación Biomédica de Occidente (CIBO) | Instituto Nacional de Enfermedades Respiratorias (INER): Centro de Investigación en Enfermedades Infecciosas (CIENI) | Consorcio Mexicano de Vigilancia Genómica (CoViGen-Mex). Authors (in alphabetical order): Julio Elias Alvarado-Yaah, Carlos F. Arias, Santiago Ávila-Ríos, Víctor Hugo Borja-Aburto, Celia Boukadida, Juan Bautista Chale-Dzul, Célida Duque Molina, José Antonio Enciso-Moreno, Gloria Elena Espinosa-Ayala, Fernando Fontove-Herrera, Victor Eduardo Garcia-Arias, Concepción Grajales-Muñiz, Ricardo Grande, Alfredo Herrera-Estrella, Carla Ivón Herrera-Najera, Pavel Isa, Brenda Irasema Maldonado-Meza, Bernardo Martínez-Miguel, Margarita Matías-Florentino, María Guadalupe de Jesús Mireles-Rivera, Gloria María Molina-Salinas, Hector Montoya-Fuentes, José Esteban Muñoz-Medina, José de Jesús Nuñez-Contreras, Alicia Ocaña-Mondragón, Luis Alberto Ochoa-Carrera, Hector Esteban Paz-Juárez, Francisco Pulido, Helen Haydee Fernanda Ramírez-Plascencia, Angel Gustavo Salas-Lais, Alejandro Sanchez-Flores, Clara Esperanza Santacruz-Tinoco, María Guadalupe Santiago-Mauricio, Nelly Sélem-Mojica, Blanca Taboada, Gloria Vazquez |
| EPI_ISL_1595804 | Department of Virology and Immunology, University of Helsinki and Helsinki University Hospital, Huslab Finland | Department of Virology, Faculty of Medicine, University of Helsinki, Helsinki, Finland | Teemu Smura, Ravi Kant, Phuoc Truong, Hussein Alburkat, Hannimari Kallio-Kokko, Jenni Virtanen, fathiah Zakham, Maija Suvanto, Essi Korhonen, Sari Hannula, Harri Kangas, Hanna Liimatainen, Satu Kurkela, Hanna Jarva, Maija Lappalainen, Pekka Ellonen, Olli Vapalahti |
| EPI_ISL_1595846, EPI_ISL_1595850, EPI_ISL_1595851 | Genome Analysis Center, Yamanashi Central Hospital | Genome Analysis Center, Yamanashi Central Hospital | Yosuke Hirotsu |
| EPI_ISL_1595853 | Yunan Center for Disease Control and Prevention | National Institute for Viral Disease Control and Prevention, China CDC | Xiaoqing Fu, Xiang Zhao, Meiling Zhang, Jienan Zhou, Yenan Feng, Zhixiao Chen, Yuchao Wu, Senquan Jia |

| | | | |
|---|---|---|---|
| EPI_ISL_1596057, EPI_ISL_1596090, EPI_ISL_1596107, EPI_ISL_1596113, EPI_ISL_1596236, EPI_ISL_1596268, EPI_ISL_1596400 | Dutch COVID-19 response team | National Institute for Public Health and the Environment (RIVM) | Adam Meijer, Harry Vennema, Dirk Eggink, Jeroen Cremer, Sharon van den Brink, Bas van der Veer, AnneMarie van den Brandt, Lisa Wijsman, Kim Freriks, Ryanne Jaarsma, Euníce Then, Jolienke Hardeman, Lynn Aarts, Sanne Bos, Melissa van Tuil, Robert Kohl, Linda van de Nes, Sjoerd Kuiling, James Groot, Florian Zwagemaker, Dennis Schmitz, Annelies Kroneman, Karim Hajji, Chantal Reusken, on behalf of the national COVID-19 response team |
| EPI_ISL_1599388 | Nordland Hospital - Bodo, Laboratory Department, Molecular Biology Unit | Norwegian Institute of Public Health, Department of Virology | Kathrine Stene-Johansen, Kamilla Heddeland Instefjord, Hilde Elshaug, Garcia Llorente Ignacio, Jon Bråte, Engebretsen Serina Beate,Pedersen Benedikte Nevjen, Debech Nadia, Atiya R Ali,Marie Paulsen Madsen, Rasmus Riis Kopperud, Hilde Vollan, Karoline Bragstad, Olav Hungnes |
| EPI_ISL_1600170 | LHUB-ULB | Labo Klinische Biologie, UZA | Marie Le Mercier, Jasmine Coppens, Basil Britto Xavier, Christine Lammens, Veerle Matheeussen, Herman Goossens |
| EPI_ISL_1608168 | Hospital São Rafael - IDOR | Hospital São Rafael - IDOR | Carolina Kymie Vasques Nonaka, Tiago Gräf, Camila Araújo de Lorenzo Barcia, Vanessa Ferreira Costa, Janderson Lopes de Oliveira, Rogério da Hora Passos, Clarissa Araújo Gurgel Rocha, Iasmin Nogueira Bastos, Maria Clara Brito de Santana, Ian Marinho Santos, Karoline Almeida Felix de Sousa, Isadora Cristina de Siqueira, Thamires Gomes Lopes Weber, Ana Verena Almeida Mendes, Bruno Solano de Freitas Souza |
| EPI_ISL_1610352, EPI_ISL_1610527 | Laboratory Corporation of America | Centers for Disease Control and Prevention Division of Viral Diseases, Pathogen Discovery | Dakota Howard, Dhwani Batra, Peter W. Cook, Kara Moser, Adrian Paskey, Jason Caravas, Benjamin Rambo-Martin, Shatavia Morrison, Christopher Gulvick, Scott Sammons, Yvette Unoarumhi, Darlene Wagner, Matthew Schmerer, Minoo Agarwal, Eyad Almasri, Debbie Boles, Ayla Burns, Nuthawin Charoensri, Oren Cohen, Susan Countryman, Mary Ann Cristobal, Bobbi Croy, Suzanne Dale, Hrushikesh Deshmukh, Amanda Douglas, Vincent Drouillon, Marcia Eisenberg, Howard Engler, Rama Ghatti, Prashant Gupta, Susan Hicks, Jake Humphrey, Lax Iyer, Manoj Jain, Mohan Kolli, Brian Krueger, Tim Kuphal, Stanley Letovsky, Michael Levandoski, Craig Lukasik, Jonathan Meltzer, Brian Norvell, Mindy Nye, Scott Parker, Christos Petropoulos, John Pruitt, Steven Ragan, Scott Ryan, Mike Sapeta, Jana Schroth, Suresh Babu Selvaraju, Goran Stevovic, Amanda Suchanek, Andrea Throop, Lyndon Tilson, Thomas Urban, Joe Voshell, Kimberly Wagner, Jonathan Williams, Mary Williamson, Qian Zeng, Tricia Zwiefelhofer, Clinton R. Paden, Duncan MacCannell |
| EPI_ISL_1610543, EPI_ISL_1610548 | KEMRI-Wellcome Trust Research Programme,Kilifi | KEMRI-Wellcome Trust Research Programme,Kilifi | Githinji G.,Matoke D.,Mohamed K.S.,de Laurent Z.,Mburu M.W.,Thiongo K. |
| EPI_ISL_1610600, EPI_ISL_1610605, EPI_ISL_1610637 | KEMRI Center for Biotechnology Research and Development | KEMRI-Wellcome Trust Research Programme,Kilifi | Githinji G.,Matoke D.,Mohamed K.S.,de Laurent Z.,Mburu M.W.,Thiongo K. |
| EPI_ISL_1610786, EPI_ISL_1611092, EPI_ISL_1612264, EPI_ISL_1612774 | Laboratory Corporation of America | Centers for Disease Control and Prevention Division of Viral Diseases, Pathogen Discovery | Dakota Howard, Dhwani Batra, Peter W. Cook, Kara Moser, Adrian Paskey, Jason Caravas, Benjamin Rambo-Martin, Shatavia Morrison, Christopher Gulvick, Scott Sammons, Yvette Unoarumhi, Darlene Wagner, Matthew Schmerer, Minoo Agarwal, Eyad Almasri, Debbie Boles, Ayla Burns, Nuthawin Charoensri, Oren Cohen, Susan Countryman, Mary Ann Cristobal, Bobbi Croy, Suzanne Dale, Hrushikesh Deshmukh, Amanda Douglas, Vincent Drouillon, Marcia Eisenberg, Howard Engler, Rama Ghatti, Prashant Gupta, Susan Hicks, Jake Humphrey, Lax Iyer, Manoj Jain, Mohan Kolli, Brian Krueger, Tim Kuphal, Stanley Letovsky, Michael Levandoski, Craig Lukasik, Jonathan Meltzer, Brian Norvell, Mindy Nye, Scott Parker, Christos Petropoulos, John Pruitt, Steven Ragan, Scott Ryan, Mike Sapeta, Jana Schroth, Suresh Babu Selvaraju, Goran Stevovic, Amanda Suchanek, Andrea Throop, Lyndon Tilson, Thomas Urban, Joe Voshell, Kimberly Wagner, Jonathan Williams, Mary Williamson, Qian Zeng, Tricia Zwiefelhofer, Clinton R. Paden, Duncan MacCannell |
| EPI_ISL_1613127 | LIC | Latvian Biomedical Research and Study Centre | Janis Pjalkovskis, Nikita Zrelovs, Monta Ustinova, Ivars Silamikelis, Liga Birzniece, Kaspars Megnis, Una Krumina, Guntars Zarins, Vita Rovite, Lauma Freimane, Laila Silamikele, Laura Ansone, Davids Fridmanis, Elina Dimina, Reinis Zeltmatis, Diana Dusacka, Jurijs Perevoscikovs, Uga Dumpis, Janis Klovins |
| EPI_ISL_1613130, EPI_ISL_1613133 | E. Gulbja laboratorija | Latvian Biomedical Research and Study Centre | Janis Pjalkovskis, Nikita Zrelovs, Monta Ustinova, Ivars Silamikelis, Liga Birzniece, Kaspars Megnis, Una Krumina, Guntars Zarins, Vita Rovite, Lauma Freimane, Laila Silamikele, Laura Ansone, Davids Fridmanis, Elina Dimina, Mikus Gavars, Dmitrijs Perminovs, Jurijs Perevoscikovs, Uga Dumpis, Janis Klovins |
| EPI_ISL_1613237, EPI_ISL_1613314 | BIOR | Latvian Biomedical Research and Study Centre | Janis Pjalkovskis, Nikita Zrelovs, Monta Ustinova, Ivars Silamikelis, Liga Birzniece, Kaspars Megnis, Una Krumina, Guntars Zarins, Vita Rovite, Lauma Freimane, Laila Silamikele, Laura Ansone, Davids Fridmanis, Elina Dimina, Irena Meistere, Daina Pule, Jurijs Perevoscikovs, Uga Dumpis, Janis Klovins |
| EPI_ISL_1613343 | E. Gulbja laboratorija | Latvian Biomedical Research and Study Centre | Janis Pjalkovskis, Nikita Zrelovs, Monta Ustinova, Ivars Silamikelis, Liga Birzniece, Kaspars Megnis, Una Krumina, Guntars Zarins, Vita Rovite, Lauma Freimane, Laila Silamikele, Laura Ansone, Davids Fridmanis, Elina Dimina, Mikus Gavars, Dmitrijs Perminovs, Jurijs Perevoscikovs, Uga Dumpis, Janis Klovins |
| EPI_ISL_1613535 | BIOR | Latvian Biomedical Research and Study Centre | Janis Pjalkovskis, Nikita Zrelovs, Monta Ustinova, Ivars Silamikelis, Liga Birzniece, Kaspars Megnis, Una Krumina, Guntars Zarins, Vita Rovite, Lauma Freimane, Laila Silamikele, Laura Ansone, Davids Fridmanis, Elina Dimina, Irena Meistere, Daina Pule, Jurijs Perevoscikovs, Uga Dumpis, Janis Klovins |
| EPI_ISL_1613636 | E. Gulbja laboratorija | Latvian Biomedical Research and Study Centre | Janis Pjalkovskis, Nikita Zrelovs, Monta Ustinova, Ivars Silamikelis, Liga Birzniece, Kaspars Megnis, Una Krumina, Guntars Zarins, Vita Rovite, Lauma Freimane, Laila Silamikele, Laura Ansone, Davids Fridmanis, Elina Dimina, Mikus Gavars, Dmitrijs Perminovs, Jurijs Perevoscikovs, Uga Dumpis, Janis Klovins |
| EPI_ISL_1613848 | BIOR | Latvian Biomedical Research and Study Centre | Janis Pjalkovskis, Nikita Zrelovs, Monta Ustinova, Ivars Silamikelis, Liga Birzniece, Kaspars Megnis, Una Krumina, Guntars Zarins, Vita Rovite, Lauma Freimane, Laila Silamikele, Laura Ansone, Davids Fridmanis, Elina Dimina, Irena Meistere, Daina Pule, Jurijs Perevoscikovs, Uga Dumpis, Janis Klovins |
| EPI_ISL_1614079 | Fulgent Genetics | Centers for Disease Control and Prevention Division of Viral Diseases, Pathogen Discovery | Dakota Howard, Dhwani Batra, Peter W. Cook, Kara Moser, Adrian Paskey, Jason Caravas, Benjamin Rambo-Martin, Shatavia Morrison, Christopher Gulvick, Scott Sammons, Yvette Unoarumhi, Darlene Wagner, Matthew Schmerer, Harry Gao, Mickey Li, John Gao, Joseph Fierro, Benafsh Sapra, Becky Tsai, Yan Meng, Doreen Ng, James Xie, Clinton R. Paden, Duncan MacCannell |
| EPI_ISL_1615301 | Helix/Illumina | Centers for Disease Control and Prevention Division of Viral Diseases, Pathogen Discovery | Dakota Howard, Dhwani Batra, Peter W. Cook, Kara Moser, Adrian Paskey, Jason Caravas, Benjamin Rambo-Martin, Shatavia Morrison, Christopher Gulvick, Scott Sammons, Yvette Unoarumhi, Darlene Wagner, Matthew Schmerer, Eileen de Feo, Jan Antico, Christine Tran, Matthew Tolentino, Shannon Wickline, Kim Gietzen, Brad Sickler, Jingtao Liu, Eric Allen, Phil Febbo, Nicole L. Washington, Simon White, Geraint Levan, Kelly Schiabor Barrett, Elizabeth Cirulli, Alexandre Bolze, Ary Ascencio, Charlotte Rivera-Garcia, Ryan Cho, Jason Nguyen, Sherry Wang, Jimmy Ramirez, Tyler Cassens, Efren Sandoval, Magnus Isaksson, William Lee, David Becker, Marc Laurent, James Lu, Clinton R. Paden, Duncan MacCannell |
| EPI_ISL_1615587 | Bandung Public Health | West Java Health Laboratory; School of Life Sciences and Technology, Institut Teknologi Bandung | Azzania Fibriani, Ema Rahmawati, Ryan Bayusantika Ristandi, Rifky Waluyajati Rachman, Cut Nur Cinthia Alamanda, Rini Robiani, Miftahul Faridl, Karimatu Khoirunnisa, Aulia Saraswati Wicaksono |
| EPI_ISL_1615590 | Subang Public Health | West Java Health Laboratory; School of Life Sciences and Technology, Institut Teknologi Bandung | Azzania Fibriani, Ema Rahmawati, Ryan Bayusantika Ristandi, Rifky Waluyajati Rachman, Cut Nur Cinthia Alamanda, Rini Robiani, Miftahul Faridl, Karimatu Khoirunnisa, Aulia Saraswati Wicaksono |
| EPI_ISL_1615664 | Armed Forces Institute of Pathology (AFIP), Dhaka Cantonment | Genomic Research Lab, BCSIR | Mohammad Mizanur Rahman, Md. Murshed Hasan Sarkar, Mohammad Samir Uzzaman, Eshrar Osman, Md. Ahasan Habib, Shahina Akter, Tanjina Akhter Banu, Abu Sayeed Mohammad Mahmud, Barna Goswami, Iffat Jahan, Md. Saddam Hossain, Mohammad Mohi Uddin, Md. Kamrul Islam, Susane Giti, Md. Salim Khan |
| EPI_ISL_1616873 | Colorado Department of Public Health and Environment | Colorado Department of Public Health and Environment | Laura Bankers, Molly C. Hetherington-Rauth, Diana Ir, Shannon Ely, Shannon R. Matzinger, Sarah Elizabeth Totten, Emily A. Travanty |
| EPI_ISL_1617610, EPI_ISL_1617659, EPI_ISL_1617810, EPI_ISL_1618225, EPI_ISL_1618228, EPI_ISL_1619245, EPI_ISL_1619292 | Swedish national genomic surveillance program of SARS-CoV-2 | The Public Health Agency of Sweden | Swedish national genomic surveillance program of SARS-CoV-2 |
| EPI_ISL_1620132 | National Public Health Laboratory, National Centre for Infectious Diseases | National Public Health Laboratory, National Centre for Infectious Diseases | Tze Minn Mak, Zhenyang Zhou, Grace Jie Yin Ngan, Royce Ang, Lin Cui, Raymond Tzer Pin Lin |
| EPI_ISL_1620175, EPI_ISL_1620176, EPI_ISL_1620178 | MRCG at LSHTM Genomics lab | MRCG at LSHTM Genomics lab | Abdul Karim sesay, Abdoulie Kanteh, Jarra Manneh, Mariama Kujabi, Bakary Sanyang |
| EPI_ISL_1621167 | Alaska State Virology Laboratory | Alaska State Virology Laboratory | Stephanie DeRonde, Elva House, Lisa Smith, Ph.D., Jack Chen, Ph.D. |
| EPI_ISL_1621297 | Middlemore Hospital | Institute of Environmental Science and Research (ESR) | Rachel Boyle, SallyAnn Harbison, Olivia Stroeven, Xiaoyun Ren, Matt Storey, Nikki Freed, Muhammad Faisal, Jing Wang, Hermes Perez, Anja Werno, Antje van der Linden, Arlo Upton, Chris Mansell, David Hammer, Dragana Drinkovic, Gary McAuliffe, Hana Sofia Andersson, James Ussher, Jill Sherwood, Josh Freeman, Julia Howard, Juliet Elvy, Mary DeAlmeida, Matt Blakiston, Matthew Rogers, Max Bloomfield, Michael Addidle, Michelle Balm, Sally Roberts, Sarah Jefferies, Sharmini Muttaiyah, Susan Morpeth, Susan Taylor, Timothy Blackmore, Vani Sathyendran, Veronica Playle, Virginia Hope, Erasmus Smit, Lauren Jelly, Olin Silander, Joep de Ligt |

| | | | |
|---|---|---|---|
| EPI_ISL_1621298, EPI_ISL_1621307, EPI_ISL_1621313, EPI_ISL_1621316, EPI_ISL_1621317 | LabPLUS | Institute of Environmental Science and Research (ESR) | Rachel Boyle, SallyAnn Harbison, Olivia Stroeven, Xiaoyun Ren, Matt Storey, Nikki Freed, Muhammad Faisal, Jing Wang, Hermes Perez, Anja Werno, Antje van der Linden, Arlo Upton, Chris Mansell, David Hammer, Dragana Drinkovic, Gary McAuliffe, Hana Sofia Andersson, James Ussher, Jill Sherwood, Josh Freeman, Julia Howard, Juliet Elvy, Mary DeAlmeida, Matt Blakiston, Matthew Rogers, Max Bloomfield, Michael Addidle, Michelle Balm, Sally Roberts, Sarah Jefferies, Sharmini Muttaiyah, Susan Morpeth, Susan Taylor, Timothy Blackmore, Vani Sathyendran, Veronica Playle, Virginia Hope, Erasmus Smit, Lauren Jelly, Olin Silander, Joep de Ligt |
| EPI_ISL_1622351 | Division of Emerging Infectious Diseases, Bureau of Infectious Diseases Diagnosis Control, Korea Disease Control and Prevention Agency | Division of Emerging Infectious Diseases, Bureau of Infectious Diseases Diagnosis Control, Korea Disease Control and Prevention Agency | Ae Kyung Park, Il-Hwan Kim, Heui Man Kim, Jeong-Min Kim, Jeong-Ah Kim, Chae Young Lee, Jin Sun No, Eun-Jin Kim |
| EPI_ISL_1622366 | RSUD Cileungsi | Eijkman Institute for Molecular Biology, Ministry of Research and Technology/National Agency for Research and Innovation | Willy Agustine, Hidayat Trimarsanto, Lydia V. Panggalo, Iskandar Adnan, Sukma Oktavianthi, Edison Johar, Frilasita A Yudhaputri, Muhammad Rezki Rasyak, Safarina G Malik, Khin Saw Myint, Amin Soebandrio |
| EPI_ISL_1622386 | RSJ Dr. Soeharto Heerdjan | Eijkman Institute for Molecular Biology, Ministry of Research and Technology/National Agency for Research and Innovation | Lydia V. Panggalo, Iskandar Adnan, Sukma Oktavianthi, Edison Johar, Frilasita A Yudhaputri, Muhammad Rezki Rasyak, Willy Agustine, Hidayat Trimarsanto, Safarina G Malik, Khin Saw Myint, Amin Soebandrio |
| EPI_ISL_1623057 | Centre Hospitalier Universitaire Clermont-Ferrand | CHU Clermont-Ferrand, service de virologie | Bisseux Maxime, Mirand Audrey, Combes Patricia, Henquell Cécile |
| EPI_ISL_1623733 | MD PHL | MD PHL | Maryland Department of Health Laboratories Administration |
| EPI_ISL_1623756, EPI_ISL_1623758 | Viral Respiratory Lab, National Institute for Biomedical Research (INRB) | Pathogen Sequencing Lab, National Institute for Biomedical Research (INRB) | Placide Mbala-Kingebeni, Edith Nkwembe, Eddy Kinganda-Lusamaki, Amuri Aziza, Francisca Muyembe Mawete, Emmanuel Lokilo Lofiko, Jean Claude Makangara, Catherine Pratt, Matthias Pauthner, Josh Quick, Allison Black, James Hadfield, Trevor Bedford, Ian Goodfellow, Andrew Rambaut, Nick Loman, Kristian Andersen, Michael Wiley, Steve Ahuka-Mundeke, Jean-Jacques Muyembe Tamfum |
| EPI_ISL_1624305 | Naval Health Research Center | Naval Medical Research Center Biological Defense Research Directorate | Logan Voegtly, Lindsay Glang, Francisco Malagon Bautista, Kyle Long, Regina Cer, Kimberly Bishop-Lilly |
| EPI_ISL_1626141, EPI_ISL_1626451 | Wisconsin State Laboratory of Hygiene Communicable Disease Division | Wisconsin State Laboratory of Hygiene Communicable Disease Division | Kelsey R. Florek, Abigail C. Shockey |
| EPI_ISL_1626501, EPI_ISL_1626519 | NSTU COVID-19 Diagnostic Center | NSU Genome Research Institute (NGRI) | Maqsud Hossain, Tahrima Saiha Huq, Aura Rahman, Md. Aminul Islam, Syeda Naushin Tabassum, Kazi Nadim Hasan, Abdul Khaleque, Abdus Sadique, Mohammad Salim Hossain, Newaz Mohammed Bahadur, Firoz Ahmed, Hasan Mahmud Reza |
| EPI_ISL_1626614 | Instituto Nacional de Medicina Genomica | Instituto Nacional de Medicina Genomica | Hidalgo-Miranda A, Mendoza-Vargas A, Reyes-Grajeda JP, Cedro-Tanda A, Alcaraz N, Gonzalez-Barrera D, Rangel-DeLeon D, Miranda-Ortiz H, Rosas-Escobar P, Canseco Mendez JC, Munguia-Garza P, Ramirez-Vega O, Garcia-Cardenas FJ, Gonzalez-Woge MA, Herrera-Montalvo LA |
| EPI_ISL_1626618, EPI_ISL_1626619 | Laboratorio de Salud Pública Bogota | Gencore - Universidad de los Andes | Luisa Sacristan, David Gonzalez, Silvia Restrepo, Marcela Guevara, Ana Maria Palacio, Johana Hernandez, Gabriela Delgado, Alejandro Gomez |
| EPI_ISL_1626722 | Institute of Virology, Biomedical Research Center of the Slovak Academy of Sciences, Bratislava | Faculty of Natural Sciences, Comenius University, Bratislava | Kristina Borsova, Viktoria Cabanova, Brona Brejova, Viktoria Hodorova, Sabina Fumacova Havlikova, Juraj Kopacek, Martina Lickova, Lubomira Lukacikova, Martina Nebohacova, Monika Slavikova, Tomas Vinar, Boris Klempa, Jozef Nosek |
| EPI_ISL_1626794, EPI_ISL_1626795, EPI_ISL_1626796, EPI_ISL_1626797, EPI_ISL_1626798, EPI_ISL_1626799, EPI_ISL_1626800, EPI_ISL_1626801 | LESP Chihuahua | Instituto de Diagnostico y Referencia Epidemiologicos (INDRE) | Claudia Wong-Arambula, Abril Rodriguez-Maldonado, Vanessa Rivero-Arredondo, Ariadna Medina-Benitez, Joaquin Quiroz-Mercado, Sergio Rangel-Guerrero, Natividad Cruz-Ortiz, Tatiana Nunez-Garcia, Gisela Barrera-Badillo, Lucia Hernandez-Rivas, Irma Lopez-Martinez, Ernesto Ramirez-Gonzalez. |
| EPI_ISL_1626802 | LESP Yucatan | Instituto de Diagnostico y Referencia Epidemiologicos (INDRE) | Claudia Wong-Arambula, Abril Rodriguez-Maldonado, Vanessa Rivero-Arredondo, Ariadna Medina-Benitez, Joaquin Quiroz-Mercado, Sergio Rangel-Guerrero, Natividad Cruz-Ortiz, Tatiana Nunez-Garcia, Gisela Barrera-Badillo, Lucia Hernandez-Rivas, Irma Lopez-Martinez, Ernesto Ramirez-Gonzalez. |
| EPI_ISL_1626803, EPI_ISL_1626804, EPI_ISL_1626805 | LESP Chihuahua | Instituto de Diagnostico y Referencia Epidemiologicos (INDRE) | Claudia Wong-Arambula, Abril Rodriguez-Maldonado, Vanessa Rivero-Arredondo, Ariadna Medina-Benitez, Joaquin Quiroz-Mercado, Sergio Rangel-Guerrero, Natividad Cruz-Ortiz, Tatiana Nunez-Garcia, Gisela Barrera-Badillo, Lucia Hernandez-Rivas, Irma Lopez-Martinez, Ernesto Ramirez-Gonzalez. |
| EPI_ISL_1626806 | LESP Nuevo Leon | Instituto de Diagnostico y Referencia Epidemiologicos (INDRE) | Claudia Wong-Arambula, Abril Rodriguez-Maldonado, Vanessa Rivero-Arredondo, Ariadna Medina-Benitez, Joaquin Quiroz-Mercado, Sergio Rangel-Guerrero, Natividad Cruz-Ortiz, Tatiana Nunez-Garcia, Gisela Barrera-Badillo, Lucia Hernandez-Rivas, Irma Lopez-Martinez, Ernesto Ramirez-Gonzalez. |
| EPI_ISL_1626807 | LESP Ciudad de Mexico | Instituto de Diagnostico y Referencia Epidemiologicos (INDRE) | Claudia Wong-Arambula, Abril Rodriguez-Maldonado, Vanessa Rivero-Arredondo, Ariadna Medina-Benitez, Joaquin Quiroz-Mercado, Sergio Rangel-Guerrero, Natividad Cruz-Ortiz, Tatiana Nunez-Garcia, Gisela Barrera-Badillo, Lucia Hernandez-Rivas, Irma Lopez-Martinez, Ernesto Ramirez-Gonzalez. |
| EPI_ISL_1626808 | LESP Hidalgo | Instituto de Diagnostico y Referencia Epidemiologicos (INDRE) | Claudia Wong-Arambula, Abril Rodriguez-Maldonado, Vanessa Rivero-Arredondo, Ariadna Medina-Benitez, Joaquin Quiroz-Mercado, Sergio Rangel-Guerrero, Natividad Cruz-Ortiz, Tatiana Nunez-Garcia, Gisela Barrera-Badillo, Lucia Hernandez-Rivas, Irma Lopez-Martinez, Ernesto Ramirez-Gonzalez. |
| EPI_ISL_1626809 | LESP Guanajuato | Instituto de Diagnostico y Referencia Epidemiologicos (INDRE) | Claudia Wong-Arambula, Abril Rodriguez-Maldonado, Vanessa Rivero-Arredondo, Ariadna Medina-Benitez, Joaquin Quiroz-Mercado, Sergio Rangel-Guerrero, Natividad Cruz-Ortiz, Tatiana Nunez-Garcia, Gisela Barrera-Badillo, Lucia Hernandez-Rivas, Irma Lopez-Martinez, Ernesto Ramirez-Gonzalez. |
| EPI_ISL_1626810 | LESP Chiapas | Instituto de Diagnostico y Referencia Epidemiologicos (INDRE) | Claudia Wong-Arambula, Abril Rodriguez-Maldonado, Vanessa Rivero-Arredondo, Ariadna Medina-Benitez, Joaquin Quiroz-Mercado, Sergio Rangel-Guerrero, Natividad Cruz-Ortiz, Tatiana Nunez-Garcia, Gisela Barrera-Badillo, Lucia Hernandez-Rivas, Irma Lopez-Martinez, Ernesto Ramirez-Gonzalez. |
| EPI_ISL_1627208 | Wojewodzka Stacja Sanitarno-Epidemiologiczna w Olsztynie, Laboratorium Badan Epidemiologiczno-Klinicznych | Wojewodzka Stacja Sanitarno-Epidemiologiczna w Olsztynie, Laboratorium Badan Epidemiologiczno-Klinicznych | Sylwia Krzetowska, Monika Czerminska, Ewa Liszewska, Tomasz Jakubczak, Marta Lukian, Patryk Bielecki, Aleksandra Kobiatko, Emilia Tarabasz, Paulina Rozycka, Barbara Dolinska |
| EPI_ISL_1627299 | Idaho Bureau of Laboratories | Idaho Bureau of Laboratories | R. Beukelman, Matthew Charles Burns, Aimee Ceniseros, Robert L. Voermans, Christopher Ball |
| EPI_ISL_1627393 | North Dakota Department of Health, Public Health Laboratory | North Dakota Department of Health, Public Health Laboratory | Lisa Wingerter |
| EPI_ISL_1627706 | LABORATORIUM BADA KLINICZNYCH WSSE w OPOLU | 1. National Institute of Public Health - National Institute of Hygiene; 2. Eurofins Genomics Europe Sequencing GmbH | Wokowicz Tomasz, Zacharczuk Katarzyna, Sadkowska-Todys Magorzata, Gierczyki Rafa, Eurofins Genomics Europe Sequencing Team, ECDC COVID-19 WGS support team |
| EPI_ISL_1627927 | Laboratorium Diagnostyki Molekularnej,Cytogenetyki i Cytometrii | 1. National Institute of Public Health - National Institute of Hygiene; 2. Eurofins Genomics Europe Sequencing GmbH | Wokowicz Tomasz, Zacharczuk Katarzyna, Sadkowska-Todys Magorzata, Gierczyki Rafa, Eurofins Genomics Europe Sequencing Team, ECDC COVID-19 WGS support team |
| EPI_ISL_1628482 | University of Mississippi Medical Center, Department of Pathology | University of Mississippi Medical Center, Molecular and Genomics Core Facility | Ashley C. Johnson, Ithiel J. Frame, Krishna K. Ayyalasomayajula, Michael R. Garrett, D. Ashley Robinson |
| EPI_ISL_1628499 | Universidad Industrial de Santander (Laboratorio Central de Investigaciones - Clínica Chicamocha). | Universidad Industrial de Santander | Francisco Martinez-Perez, Cristian E. Cadena-Caballero, Diego Rueda-Plata, Carolina S. Torres-Jiménez, Lizeth J. Forero-Buitrago, Erika Lizarazo-Gutiérrez, Carlos Barrios-Hernández, Lina M. Vera-Cala. |
| EPI_ISL_1628597, EPI_ISL_1628598, EPI_ISL_1628605, EPI_ISL_1628609, EPI_ISL_1628624, EPI_ISL_1628645, EPI_ISL_1628683 | InDRE | Instituto Nacional de Medicina Genomica | Hidalgo-Miranda A, Mendoza-Vargas A, Reyes-Grajeda JP, Cedro-Tanda A, Alcaraz N, Gisela Barrera-Badillo, Irma Lopez-Martinez, Jose Ernesto Ramirez González, Gonzalez-Barrera D, Rangel-DeLeon D, Miranda-Ortiz H, Rosas-Escobar P, Canseco Mendez JC, Munguia-Garza P, Ramirez-Vega O, Garcia-Cardenas FJ, Gonzalez-Woge MA, Herrera-Montalvo LA. |
| EPI_ISL_1629710 | Universidad Industrial de Santander (Laboratorio Central de Investigaciones - Clínica Chicamocha). | Universidad Industrial de Santander. | Francisco Martinez-Perez, Cristian E. Cadena-Caballero, Diego Rueda-Plata, Carolina S. Torres-Jiménez, Lizeth J. Forero-Buitrago, Erika Lizarazo-Gutiérrez, Carlos Barrios-Hernández, Lina M. Vera-Cala. |
| EPI_ISL_1629731 | UDEA | Universidad Nacional de Colombia - Laboratorio Genómico | Laura Silvana Perez, Andres F. Cardona-Rios, Daniel O. Maldonado-Perez, Maria Stella López, Simón Villegas Velásquez, Karl A Ciuoderis, Idabely |

| | | | |
|---|---|---|---|
| | | One Health | Betancur Ortiz, Sandra Ines Cano, Eliana Patricia Calvo Tapiero, Diego A. Álvarez-Díaz, Carlos Franco-Muñoz, Hector Alejandro Ruiz-Moreno, Marcela Mercado-Reyes, Jorge E. Osorio, Juan P. Hernandez-Ortiz |
| EPI_ISL_1629734, EPI_ISL_1629735 | SURA | Universidad Nacional de Colombia - Laboratorio Genómico One Health | Laura Silvana Perez, Andres F. Cardona-Rios, Daniel O. Maldonado-Perez, Maria Stella López, Simón Villegas Velásquez, Karl A Ciuoderis, Idabely Betancur Ortiz, Sandra Ines Cano, Eliana Patricia Calvo Tapiero, Diego A. Álvarez-Díaz, Carlos Franco-Muñoz, Hector Alejandro Ruiz-Moreno, Marcela Mercado-Reyes, Jorge E. Osorio, Juan P. Hernandez-Ortiz |
| EPI_ISL_1629746 | LABORATORIO ECHAVARRIA | Universidad Nacional de Colombia - Laboratorio Genómico One Health | Andres F. Cardona-Rios, Daniel O. Maldonado-Perez, Maria Stella López, Simón Villegas Velásquez, Laura Silvana Perez, Karl A Ciuoderis, Idabely Betancur Ortiz, Sandra Ines Cano, Eliana Patricia Calvo Tapiero, Diego A. Álvarez-Díaz, Carlos Franco-Muñoz, Hector Alejandro Ruiz-Moreno, Marcela Mercado-Reyes, Jorge E. Osorio, Juan P. Hernandez-Ortiz |
| EPI_ISL_1629779 | Hospital Carlos Alberto Seguín Escobedo - EsSalud | Laboratorio de Genómica Microbiana, Universidad Peruana Cayetano Heredia | Lenin Maturrano, Pablo Tsukayama, Alejandra Dávila-Barclay, Guillermo Salvatierra, Luis González, Pedro E. Romero, Janet Huancachoque, Pool Marcos, Fernando Sánchez Fragoso, Lin Zevallos Cuarite |
| EPI_ISL_1629784 | Laboratorio de Referencia Nacional de Virus Respiratorios, Instituto Nacional de Salud Peru | Laboratorio de Genómica Microbiana, Universidad Peruana Cayetano Heredia | Lenin Maturrano, Pablo Tsukayama, Alejandra Dávila-Barclay, Guillermo Salvatierra, Luis González, Pedro E. Romero, Diego Cuicapuza, Janet Huancachoque, Pool Marcos, Maribel Huaringa, Priscilla Lope, Nancy Rojas |
| EPI_ISL_1629791, EPI_ISL_1629793, EPI_ISL_1629794, EPI_ISL_1629795, EPI_ISL_1629797, EPI_ISL_1629798 | Instituto de Medicina Tropical Alexander Von Humboldt, Universidad Peruana Cayetano Heredia | Laboratorio de Genómica Microbiana, Universidad Peruana Cayetano Heredia | Lenin Maturrano, Pablo Tsukayama, Alejandra Dávila-Barclay, Guillermo Salvatierra, Luis González, Pedro E. Romero, Diego Cuicapuza, Janet Huancachoque, Pool Marcos |
| EPI_ISL_1629808 | Laboratorio de Referencia Nacional de Virus Respiratorios, Instituto Nacional de Salud Peru | Laboratorio de Genómica Microbiana, Universidad Peruana Cayetano Heredia | Lenin Maturrano, Pablo Tsukayama, Alejandra Dávila-Barclay, Guillermo Salvatierra, Luis González, Pedro E. Romero, Diego Cuicapuza, Janet Huancachoque, Pool Marcos |
| EPI_ISL_1630256, EPI_ISL_1630257 | IRESSEF | Abbott | Souleymane Mboup, Ambroise Ahouidi, Adbou Padane, Nafissatou Leye, Moustapha Mbow, Aminata Mboup, Papa Alassane Diaw, Cyrille Diedhiou, Aminata Dia , Anna julienne selbe Ndiaye, Ndeye Diabou Diagne, Ana Olivo, Todd Meyer, Barbara Harris, Mary Rodgers, Gavin Cloherty |
| EPI_ISL_1631790 | Randox Laboratories | Wellcome Sanger Institute for the COVID-19 Genomics UK (COG-UK) Consortium | Randox Laboratories and Alex Alderton, Roberto Amato, Jeffrey Barrett, Sonia Goncalves, Ewan Harrison, David K. Jackson, Ian Johnston, Dominic Kwiatkowski, Cordelia Langford, John Sillitoe on behalf of the Wellcome Sanger Institute COVID-19 Surveillance Team |
| EPI_ISL_1632531 | UNIVERSIDAD DEL SINU | Instituto Nacional de Salud- Dirección de Investigación en Salud Pública | Katherine Laiton-Donato, Diego A. Álvarez-Díaz, Carlos Franco-Muñoz, Hector Alejandro Ruiz-Moreno, Paola Rojas, Maria T. Herrera-Sepúlveda, Diego Andrés Prada, Jhonnatan Reales-González, Sheryll Corchuelo, Julian Naizaque, Jorge Rivera, Gerardo Santamaría, Sergio Gomez, Lisseth Pardo, Juan Camilo Martinez, Marta Lopez Blanco, Ángela Alarcon Cruz, Diana Malo, Carmen Osorio, Magdalena Wiesner, Martha Lucia Ospina Martinez, Marcela Mercado-Reyes |
| EPI_ISL_1633305 | Central Public Health Lab, National Public Health Organization | Central Public Health Lab, National Public Health Organization | Kyriaki Tryfinopoulou et al |
| EPI_ISL_1633504 | Genetica Molecular and Subdepartamento de Virologia ISP Chile | Instituto de Salud Publica de Chile | Javier Tognarelli, Karen Orostica, Barbara Parra, Loredana Arata, Jaime Lagos, Gisselle Barra, Patricia Bustos, Rodrigo Fasce, Andres Castillo, Jorge Fernandez |
| EPI_ISL_1633656, EPI_ISL_1633658 | Public Health Virology-Forensic and Scientific Services (PHV-FSS) | Public Health Virology-Forensic and Scientific Services (PHV-FSS) | Son Nguyen |
| EPI_ISL_1633674, EPI_ISL_1633675, EPI_ISL_1633677, EPI_ISL_1633681 | PathWest Laboratory Medicine WA | PathWest Laboratory Medicine WA Microbial Surveillance Unit | PathWest Laboratory Medicine WA Microbial Surveillance Unit |
| EPI_ISL_1634442 | Mitra Kasih Hospital | West Java Health Laboratory; School of Life Sciences and Technology, Institut Teknologi Bandung | Azzania Fibriani, Ema Rahmawati, Ryan Bayusantika Ristandi, Rifky Waluyajati Rachman, Cut Nur Cinthia Alamanda, Rini Robiani, Miftahul Faridl, Karimatu Khoirunnisa, Aulia Saraswati Wicaksono |
| EPI_ISL_1634470 | Tasikmalaya Public Health | West Java Health Laboratory; School of Life Sciences and Technology, Institut Teknologi Bandung | Azzania Fibriani, Ema Rahmawati, Ryan Bayusantika Ristandi, Rifky Waluyajati Rachman, Cut Nur Cinthia Alamanda, Rini Robiani, Miftahul Faridl, Karimatu Khoirunnisa, Aulia Saraswati Wicaksono |
| EPI_ISL_1634474 | Cimahi Public Health | West Java Health Laboratory; School of Life Sciences and Technology, Institut Teknologi Bandung | Azzania Fibriani, Ema Rahmawati, Ryan Bayusantika Ristandi, Rifky Waluyajati Rachman, Cut Nur Cinthia Alamanda, Rini Robiani, Miftahul Faridl, Karimatu Khoirunnisa, Aulia Saraswati Wicaksono |
| EPI_ISL_1636127 | Pandemic Response Lab - NYC | Pandemic Response Lab, R&D | Henry Lee, Michael Hammerling, Melissa Hopkins, Cybill del Castillo, Shinyoung Clair Kang, William Ward, Pradeep Bugga, Sol Rey, Dylan Law, Katharine Nelson, Haiping Hao, Jon Laurent |
| EPI_ISL_1639329 | Chemische und VeterinÃ¤runtersuchungsamt MÃ¼nsterland-Emscher-Lippe (CVUA-MEL) | Robert Koch Institute | unknown |
| EPI_ISL_1647656, EPI_ISL_1647661, EPI_ISL_1647681, EPI_ISL_1647710 | Public Health Authority of the Slovak Republic | Laboratory of Genomics and Bioinformatics, Comenius University Science Park | Tatiana Sedláková, Diana Rusáková, Miroslav Böhmer, Anna Giová, Jaroslav Budiš, Tomáš Szemes |
| EPI_ISL_1649083, EPI_ISL_1649398, EPI_ISL_1649560 | Aegis Sciences Corporation | Centers for Disease Control and Prevention Division of Viral Diseases, Pathogen Discovery | Dakota Howard, Dhwani Batra, Peter W. Cook, Kara Moser, Adrian Paskey, Jason Caravas, Benjamin Rambo-Martin, Shatavia Morrison, Christopher Gulvick, Scott Sammons, Yvette Unoarumhi, Darlene Wagner, Matthew Schmerer, Cyndi Clark, Patrick Campbell, Rob Case, Vikramsinha Ghorpade, Holly Houdeshell, Ola Kvalvaag, Dillon Nall, Ethan Sanders, Alec Vest, Shaun Westlund, Matthew Hardison, Clinton R. Paden, Duncan MacCannell |
| EPI_ISL_1651264 | LESP Morelos | Instituto de Diagnostico y Referencia Epidemiologicos (INDRE) | Claudia Wong-Arambula, Abril Rodriguez-Maldonado, Vanessa Rivero-Arredondo, Ariadna Medina-Benitez, Joaquin Quiroz-Mercado, Sergio Rangel-Guerrero, Natividad Cruz-Ortiz, Tatiana Nunez-Garcia, Gisela Barrera-Badillo, Lucia Hernandez-Rivas, Irma Lopez-Martinez, Ernesto Ramirez-Gonzalez. |
| EPI_ISL_1652056, EPI_ISL_1652058 | Integrated Biorepository of H3Africa Uganda - IBRH3AU | Molecular Biology Laboratory | Savannah Mwesigwa, Eric Katagirya, Gerald Mboowa, David Patrick Kateete, Misaki Wayengera, Emmanuel Nasinghe, Ashaba Fred Katabazi, Edgar Kigozi, Samuel Kirimunda, Rogers Kamulegeya, Moses Luutu, Lwanga Newton, Nsubuga Gideon, Bernard Ssentalo Bagaya, Sarah Stanley, Moses Joloba |
| EPI_ISL_1652637, EPI_ISL_1652640, EPI_ISL_1652682 | HELIX LLC | WHO National Influenza Centre Russian Federation | Andrey Komissarov, Artem Fadeev, Anna Ivanova, Kseniya Komissarova, Tamila Musaeva, Maria Timofeeva, Veronika Eder, Maria Pisareva, Daria Danilenko, Ksenia Safina, Elena Nabieva, Georgii Bazykin, Dmitry Lioznov |
| EPI_ISL_1653484 | Lighthouse Lab in Alderley Park | Wellcome Sanger Institute for the COVID-19 Genomics UK (COG-UK) Consortium | Jacquelyn Wynn, Mairead Hyland, The Lighthouse Lab in Alderley Park and Alex Alderton, Roberto Amato, Jeffrey Barrett, Sonia Goncalves, Ewan Harrison, David K. Jackson, Ian Johnston, Dominic Kwiatkowski, Cordelia Langford, John Sillitoe on behalf of the Wellcome Sanger Institute COVID-19 Surveillance Team |
| EPI_ISL_1653933 | Laboratory for HIV and opportunistic infections diagnosis The Republican Research and Practical Center for Epidemiology and Microbiology (RRPCEM) | Laboratory for HIV and opportunistic infections diagnosis The Republican Research and Practical Center for Epidemiology and Microbiology (RRPCEM) | Elena Gasich, Kirill Bulda, Yauhen Sysaliatsin, Leonid Valentovich, Alina Drozd, Nastassia Kabankova, Katsiaryna Belyakova, Anatoly Krasko, Vladimir Gorbunov, Alena Mikhalenka, Alexander Kilchevcky |
| EPI_ISL_1654212, EPI_ISL_1654215, EPI_ISL_1654217, EPI_ISL_1654218 | Fondation Congolaise pour la recherche medicale (FCRM), Francine Ntoumi | Institute of Tropical Medicine | Prof. Francine Ntoumi and Prof. Dr. Thirumalaisamy P. Velavan |
| EPI_ISL_1654357 | Department of Public Health Microbiology Ljubljana, National Laboratory for Health, Environment and Food | Department for Public Health Microbiology Ljubljana, National Laboratory for Health, Environment and Food | Tom Koritnik, José Gonçalves, Martin Bosilj, Katarina Prosenc, Natasa Berginc, Metka Paragi |
| EPI_ISL_1654710 | NVMRVI | National Public Health Surveillance Laboratory | Lukas Zemaitis, Migle Gabrielaite, Jelena Razmuk, Svajune Muralyte, Ana Steponkiene, Lukas Vasionis, Danas Baksa |
| EPI_ISL_1654812, EPI_ISL_1654813, EPI_ISL_1654814, EPI_ISL_1654815, EPI_ISL_1654827, EPI_ISL_1654839 | Institute of Virology, Vaccines and Sera "Torlak" | Institute of microbiology and Immunology, Faculty of Medicine, University of Belgrade | Knezevic,A., Jankovic,M., Vidanovic,D., Milicevic,O., Tesovic,B., Sekler,M., Jovanovic,T. |
| EPI_ISL_1654842 | Laboratory for COVID19 diagnostics, Clinical Centre of Serbia | Institute of microbiology and Immunology, Faculty of Medicine, University of Belgrade | Knezevic,A., Jankovic,M., Vidanovic,D., Milicevic,O., Tesovic,B., Sekler,M., Jovanovic,T. |
| EPI_ISL_1656108 | NMVRVI | National Public Health Surveillance Laboratory | Lukas Zemaitis, Migle Gabrielaite, Jelena Razmuk, Svajune Muralyte, Ana Steponkiene, Lukas Vasionis, Danas Baksa |

| | | | |
|---|---|---|---|
| EPI_ISL_1656146 | Universidad Tecnica Particular de Loja | Institute of Microbiology, Universidad San Francisco de Quito | Belén Prado-Vivar, Sully Márquez, Juan José Guadalupe, Monica Becerra-Wong, Bernardo Gutiérrez, Paola Dalgo, Fernando Serrano, Katherine Ojeda, David Zuñiga, Melissa Ortega, Raiza Briceño, Luis Flores, Verónica Barragán, Patricio Rojas-Silva, Gabriel Trueba, Michelle Grunauer, Paúl Cárdenas |
| EPI_ISL_1656147 | Institute of Microbiology, Universidad San Francisco de Quito | Institute of Microbiology, Universidad San Francisco de Quito | Belén Prado-Vivar, Sully Márquez, Juan José Guadalupe, Monica Becerra-Wong, Bernardo Gutiérrez, Mayra Perero Intriago, Freddy Saldarriaga-Mera , Miguel Sacoto Mazini, Verónica Barragán, Patricio Rojas-Silva, Gabriel Trueba, Michelle Grunauer, Paúl Cárdenas |
| EPI_ISL_1658994 | Viollier AG | Department of Biosystems Science and Engineering, ETH Zurich | Chaoran Chen, Sarah Nadeau, Catharine Aquino, Ivan Topolsky, Philipp Jablonski, Lara Fuhrmann, David Dreifuss, Katharina Jahn, Andreia Cabral de Gouvea, Maria Domenica Moccia, Simon Gruter, Timothy Sykes, Lennart Opitz, Griffin White, Laura Neff, Doris Popovic, Andrea Patrignani, Jay Tracy, Ralph Schlapbach, Christiane Beckmann, Maurice Redondo, Olivier Kobel, Christoph Noppen, Sophie Seidel, Noemie Santamaria de Souza, Niko Beerenwinkel, Tanja Stadler |
| EPI_ISL_1660233, EPI_ISL_1660234, EPI_ISL_1660242, EPI_ISL_1660248, EPI_ISL_1660251, EPI_ISL_1660252, EPI_ISL_1660256, EPI_ISL_1660272, EPI_ISL_1660286, EPI_ISL_1660288, EPI_ISL_1660289, EPI_ISL_1660290 | | | |
| see above | Virology Unit, Institut Pasteur de Madagascar | Virology Unit, Institut Pasteur de Madagascar | Christian Ranaivoson, Cara E. Brook, Vida Ahyong, Soa Fy Andriamandimby, Vololoniaina Raharinosy, Tsiry Randriambolamanantsoa, Helisoa Razafimanjato, Norosoa Razanajatovo, Michelle Tan, Cristina M. Tato, Joseph L. DeRisi, Jean-Michel Heraud, Philippe Dussart |
| EPI_ISL_1660296, EPI_ISL_1660300, EPI_ISL_1660302, EPI_ISL_1660303, EPI_ISL_1660307, EPI_ISL_1660310, EPI_ISL_1660331, EPI_ISL_1660332 | Virology Unit, Institut Pasteur de Madagascar | Virology Unit, Institut Pasteur de Madagascar | Marion Barbet, Sylvie Behillil, Frédéric Lemoine, Corinne Maufrais, Christophe Malabat, Emmanuelle Permal, Méline Bizard, Angela Brisebarre, Camille Capel, Louise Lefrançois, Etienne Simon-Lorière, Vincent Enouf, Maud Vanpeene, Christian Ranaivoson, Cara E. Brook, Soa Fy Andriamandimby, Vololoniaina Raharinosy, Tsiry Randriambolamanantsoa, Helisoa Razafimanjato, Norosoa Razanajatovo, Sylvie van der Werf, Philippe Dussart |
| EPI_ISL_1660398, EPI_ISL_1660411 | New South Wales Health Pathology Royal Prince Alfred Hospital | Microbiology RPAH | Foster, C.; Au, J.; Ruiz Silva, M.; Deveson, I.; Bull, R.; Van Hal, S.; Rawlinson, W. |
| EPI_ISL_1660419 | Santosa Kopo Hospital | West Java Health Laboratory; School of Life Sciences and Technology, Institut Teknologi Bandung | Azzania Fibriani, Ema Rahmawati, Ryan Bayusantika Ristandi, Rifky Waluyajati Rachman, Cut Nur Cinthia Alamanda, Rini Robiani, Miftahul Faridl, Karimatu Khoirunnisa, Aulia Saraswati Wicaksono |
| EPI_ISL_1660439, EPI_ISL_1660446, EPI_ISL_1660464, EPI_ISL_1660465 | Communicable Disease Laboratory, Public Health Directorate | Communicable Disease Laboratory, Public Health Directorate | Alwasti,H., AlHujairi,Z., AlAbbas,Z., Marhoon,A., Almoamen,G. |
| EPI_ISL_1660476 | New South Wales Health Pathology Royal Prince Alfred Hospital | Microbiology RPAH | Foster, C.; Au, J.; Ruiz Silva, M.; Deveson, I.; Bull, R.; Van Hal, S.; Rawlinson, W. |
| EPI_ISL_1661091 | Platform BIS UZA/UAntwerpen | Labo Klinische Biologie, UZA | Jasmine Coppens, Marie Le Mercier, Basil Britto Xavier, Christine Lammens, Veerle Matheeussen, Herman Goossens |
| EPI_ISL_1661496 | LESP Guanajuato | Instituto de Diagnostico y Referencia Epidemiologicos (INDRE) | Claudia Wong-Arambula, Abril Rodriguez-Maldonado, Vanessa Rivero-Arredondo, Ariadna Medina-Benitez, Joaquin Quiroz-Mercado, Sergio Rangel-Guerrero, Natividad Cruz-Ortiz, Tatiana Nunez-Garcia, Gisela Barrera-Badillo, Lucia Hernandez-Rivas, Irma Lopez-Martinez, Ernesto Ramirez-Gonzalez. |
| EPI_ISL_1661551 | LESP Hidalgo | Instituto de Diagnostico y Referencia Epidemiologicos (INDRE) | Claudia Wong-Arambula, Abril Rodriguez-Maldonado, Vanessa Rivero-Arredondo, Ariadna Medina-Benitez, Joaquin Quiroz-Mercado, Sergio Rangel-Guerrero, Natividad Cruz-Ortiz, Tatiana Nunez-Garcia, Gisela Barrera-Badillo, Lucia Hernandez-Rivas, Irma Lopez-Martinez, Ernesto Ramirez-Gonzalez. |
| EPI_ISL_1661649, EPI_ISL_1661650 | UAB Medicina practica laboratorija | Vilnius University Hospital Santaros Klinikos, Center of Laboratory Medicine | Gytis Dudas, Daniel Naumovas, Dovile Ezerskyte, Ingrida Olendraite, Rimvydas Norvilas, Ligita Raugaite, Monika Katenaite, Mindaugas Stoskus, Laimonas Griskevicius |
| EPI_ISL_1661724, EPI_ISL_1661758 | Anteja laboratorija (UAB Diagnostikos laboratorija) | Vilnius University Hospital Santaros Klinikos, Center of Laboratory Medicine | Gytis Dudas, Daniel Naumovas, Dovile Ezerskyte, Ingrida Olendraite, Rimvydas Norvilas, Ligita Raugaite, Monika Katenaite, Mindaugas Stoskus, Laimonas Griskevicius |
| EPI_ISL_1661789 | UAB "Rezus.lt" | Vilnius University Hospital Santaros Klinikos, Center of Laboratory Medicine | Gytis Dudas, Daniel Naumovas, Dovile Ezerskyte, Ingrida Olendraite, Rimvydas Norvilas, Ligita Raugaite, Monika Katenaite, Mindaugas Stoskus, Laimonas Griskevicius |
| EPI_ISL_1661919 | IU-Cerrahpasa, Cerrahpasa School of Medicine, COVID-19 Lab | IU-Cerrahpasa, Cerrahpasa School of Medicine, COVID-19 Lab | Mert Kuskucu, Yesim Tuyji Tok, Kenan Midilli |
| EPI_ISL_1661936 | Laboratori Clínic Territorial de Girona | Can Ruti SARS-CoV-2 Sequencing Hub (HUGTiP/IrsiCaixa/IGTP) | Marc Noguera-Julian, Pilar Armengol, Ignacio Blanco, Antoni E Bordoy, Francesc Catala-Moll, Pere-Joan Cardona, Maria Casadellà, Cristina Casañ, Gemma Clara, Bonaventura Clotet, Cristina Esteban, Montserrat Giménez, Mercedes Guerrero, Anna Not, Roger Paredes, Mariona Parera, Verónica Saludes, Alba Sánchez, and Elisa Martró on behalf of the Can Ruti SARS-CoV-2 Sequencing Hub. |
| EPI_ISL_1661984, EPI_ISL_1661987, EPI_ISL_1661988, EPI_ISL_1662047, EPI_ISL_1662048 | Centro de Investigacion Biomedica del Noreste (CIBIN) | Unidad de Genomica Avanzada | Consorcio Mexicano de Vigilancia Genomica (CoViGen-Mex). Authors (in alphabetical order): Julio Elias Alvarado-Yaah, Carlos F. Arias, Santiago Ávila-Rios, Victor Hugo Borja-Aburto, Celia Boukadida, Juan Bautista Chale-Dzul, Celida Duque Molina, Jose Antonio Enciso-Moreno, Gloria Elena Espinosa-Ayala, Fernando Fontove-Herrera, Victor Eduardo Garcia-Arias,Concepcion Grajales-Muniz, Ricardo Grande, Alfredo Herrera-Estrella, Carla Ivon Herrera-Najera, Pavel Isa, Brenda Irasema Maldonado-Meza, Bernardo Martinez-Miguel, Margarita Matias-Florentino, Maria Guadalupe de Jesus Mireles-Rivera, Gloria Maria Molina-Salinas, Hector Montoya-Fuentes, Jose Esteban Munoz-Medina, Jose de Jesus Nunez-Contreras, Alicia Ocana-Mondragon, Luis Alberto Ochoa-Carrera, Hector Esteban Paz-Juarez, Francisco Pulido, Helen Haydee Fernanda Ramirez-Plascencia, Angel Gustavo Salas-Lais, Alejandro Sanchez-Flores, Clara Esperanza Santacruz-Tinoco, Maria Guadalupe Santiago-Mauricio, Nelly Selem-Mojica, Blanca Taboada, Gloria Vazquez |
| EPI_ISL_1662060, EPI_ISL_1662061, EPI_ISL_1662073, EPI_ISL_1662074, EPI_ISL_1662119, EPI_ISL_1662121, EPI_ISL_1662170, EPI_ISL_1662171, EPI_ISL_1662210 | Laboratorio Central de Epidemiologia (LCE) | Unidad de Genomica Avanzada | Consorcio Mexicano de Vigilancia Genomica (CoViGen-Mex). Authors (in alphabetical order): Julio Elias Alvarado-Yaah, Carlos F. Arias, Santiago Ávila-Rios, Victor Hugo Borja-Aburto, Celia Boukadida, Juan Bautista Chale-Dzul, Celida Duque Molina, Jose Antonio Enciso-Moreno, Gloria Elena Espinosa-Ayala, Fernando Fontove-Herrera, Victor Eduardo Garcia-Arias,Concepcion Grajales-Muniz, Ricardo Grande, Alfredo Herrera-Estrella, Carla Ivon Herrera-Najera, Pavel Isa, Brenda Irasema Maldonado-Meza, Bernardo Martinez-Miguel, Margarita Matias-Florentino, Maria Guadalupe de Jesus Mireles-Rivera, Gloria Maria Molina-Salinas, Hector Montoya-Fuentes, Jose Esteban Munoz-Medina, Jose de Jesus Nunez-Contreras, Alicia Ocana-Mondragon, Luis Alberto Ochoa-Carrera, Hector Esteban Paz-Juarez, Francisco Pulido, Helen Haydee Fernanda Ramirez-Plascencia, Angel Gustavo Salas-Lais, Alejandro Sanchez-Flores, Clara Esperanza Santacruz-Tinoco, Maria Guadalupe Santiago-Mauricio, Nelly Selem-Mojica, Blanca Taboada, Gloria Vazquez |
| EPI_ISL_1662215 | Unidad de Investigacion Medica de Yucatan (UIMY) | Unidad de Genomica Avanzada | Consorcio Mexicano de Vigilancia Genomica (CoViGen-Mex). Authors (in alphabetical order): Julio Elias Alvarado-Yaah, Carlos F. Arias, Santiago Ávila-Rios, Victor Hugo Borja-Aburto, Celia Boukadida, Juan Bautista Chale-Dzul, Celida Duque Molina, Jose Antonio Enciso-Moreno, Gloria Elena Espinosa-Ayala, Fernando Fontove-Herrera, Victor Eduardo Garcia-Arias,Concepcion Grajales-Muniz, Ricardo Grande, Alfredo Herrera-Estrella, Carla Ivon Herrera-Najera, Pavel Isa, Brenda Irasema Maldonado-Meza, Bernardo Martinez-Miguel, Margarita Matias-Florentino, Maria Guadalupe de Jesus Mireles-Rivera, Gloria Maria Molina-Salinas, Hector Montoya-Fuentes, Jose Esteban Munoz-Medina, Jose de Jesus Nunez-Contreras, Alicia Ocana-Mondragon, Luis Alberto Ochoa-Carrera, Hector Esteban Paz-Juarez, Francisco Pulido, Helen Haydee Fernanda Ramirez-Plascencia, Angel Gustavo Salas-Lais, Alejandro Sanchez-Flores, Clara Esperanza Santacruz-Tinoco, Maria Guadalupe Santiago-Mauricio, Nelly Selem-Mojica, Blanca Taboada, Gloria Vazquez |
| EPI_ISL_1662225, EPI_ISL_1662226, EPI_ISL_1662230, EPI_ISL_1662231, EPI_ISL_1662232 | Unidad de Investigacion Biomedica de Zacatecas (UIBZ) | Unidad de Genomica Avanzada | Consorcio Mexicano de Vigilancia Genomica (CoViGen-Mex). Authors (in alphabetical order): Julio Elias Alvarado-Yaah, Carlos F. Arias, Santiago Ávila-Rios, Victor Hugo Borja-Aburto, Celia Boukadida, Juan Bautista Chale-Dzul, Celida Duque Molina, Jose Antonio Enciso-Moreno, Gloria Elena Espinosa-Ayala, Fernando Fontove-Herrera, Victor Eduardo Garcia-Arias,Concepcion Grajales-Muniz, Ricardo Grande, Alfredo Herrera-Estrella, Carla Ivon Herrera-Najera, Pavel Isa, Brenda Irasema Maldonado-Meza, Bernardo Martinez-Miguel, Margarita Matias-Florentino, Maria Guadalupe de Jesus Mireles-Rivera, Gloria Maria Molina-Salinas, Hector Montoya-Fuentes, Jose Esteban Munoz-Medina, Jose de Jesus Nunez-Contreras, Alicia Ocana-Mondragon, Luis Alberto Ochoa-Carrera, Hector Esteban Paz-Juarez, Francisco Pulido, Helen Haydee Fernanda Ramirez-Plascencia, Angel Gustavo Salas-Lais, Alejandro Sanchez-Flores, Clara Esperanza Santacruz-Tinoco, Maria Guadalupe Santiago-Mauricio, Nelly Selem-Mojica, Blanca Taboada, Gloria Vazquez |
| EPI_ISL_1662620 | IU-Cerrahpasa, Cerrahpasa School of Medicine, COVID-19 Lab | IU-Cerrahpasa, Cerrahpasa School of Medicine, COVID-19 Lab | Mert Kuskucu, Yesim Tuyji Tok, Kenan Midilli |
| EPI_ISL_1662645 | Delaware Public Health Lab | Delaware Public Health Lab | Rebecca Savage |

| | | | |
|---|---|---|---|
| EPI_ISL_1663061 | IU-Cerrahpasa, Cerrahpasa School of Medicine, COVID-19 Lab | IU-Cerrahpasa, Cerrahpasa School of Medicine, COVID-19 Lab | Mert Kuskucu, Yesim Tuyji Tok, Kenan Midilli |
| EPI_ISL_1663062 | IU-Cerrahpasa, Cerrahpasa School of Medicine, COVID-19 Lab | Illumina Miseq | Mert Kuskucu, Yesim Tuyji Tok, Kenan Midilli |
| EPI_ISL_1663231 | Fimlab Laboratories | Fimlab Laboratories | Leena Huhti, Minna Paloniemi, Sara Lehtinen, Bruno Luukinen, Mauri Keinänen, Tapio Seiskari |
| EPI_ISL_1663379 | NCCS, Pune | Institute of Life Sciences - INSACOG | Sunil K. Raghav, Safal Walia, Arup Ghosh, Atimukta Jha, Amol M. Kanampalliwar, Shifu Aggarwal, Rupesh Dash, Rajeeb Swain, Punit Prasad, INSACOG Consortium, Ajay Parida |
| EPI_ISL_1663493 | Capital hospital, Bhubaneswar | Institute of Life Sciences - INSACOG | Sunil K. Raghav, Safal Walia, Arup Ghosh, Atimukta Jha, Amol M. Kanampalliwar, Shifu Aggarwal, Rupesh Dash, Rajeeb Swain, Punit Prasad, INSACOG Consortium, Ajay Parida |
| EPI_ISL_1663570 | Fimlab Laboratories | Fimlab Laboratories | Minna Paloniemi, Sara Lehtinen, Leena Huhti, Bruno Luukinen, Tapio Seiskari, Mauri Keinäne |
| EPI_ISL_1663577, EPI_ISL_1663579, EPI_ISL_1663581 | IU-Cerrahpasa, Cerrahpasa School of Medicine, COVID-19 Lab | IU-Cerrahpasa, Cerrahpasa School of Medicine, COVID-19 Lab | Mert Kuskucu, Yesim Tuyji Tok, Kenan Midilli |
| EPI_ISL_1663659, EPI_ISL_1663660, EPI_ISL_1663666, EPI_ISL_1663674, EPI_ISL_1663676, EPI_ISL_1663677, EPI_ISL_1663678, EPI_ISL_1663679, EPI_ISL_1663680 | Molecular diagnostic unit for viral haemorrhagic fevers and emerging viruses, Bouaké CHU Laboratory | Molecular diagnostic unit for viral haemorrhagic fevers and emerging viruses, Bouaké CHU Laboratory | Chantal Akoua-Koffi, Diané Bamourou, Etilé Anoh, Oby Wayoro, Safiatou Karidioula, Adjaratou Traoré, Soundélé Maïté, Monemo Pacome, Coulibaly Mbegnan, Bamba Fatoumata Touré, Kra Ouffoué, Grit Schubert, Essia Belarbi, Fabian Leendertz |
| EPI_ISL_1664175 | Laboratorio Central Noel Nutels | Bioinformatics Laboratory / LNCC | Luiz G P de Almeida, Alessandra P Lamarca, Ronaldo da Silva F Jr, Liliane Cavalcante, Alexandra L Gerber, Ana Paula de C Guimarães, Douglas Terra Machado, Cassia Alves, Diana Mariani, Thais Felix Cruz, Mario Sergio Ribeiro, Silvia Carvalho, Flávio Dias da Silva, Marcio Henrique de Oliveira Garcia, Leandro Magalhães de Souza, Cristiane Gomes da Silva, Caio Luiz Pereira Ribeiro, Andréa Cony Cavalcanti, Claudia Maria Braga de Mello, Amilcar Tanuri, Ana Tereza R Vasconcelos |
| EPI_ISL_1664478 | New Mexico Department of Health Scientific Laboratory | Center for Global Health, University of New Mexico Health Sciences Center | Daryl Domman, Kurt Schwalm, Twila Kunde, Joseph Hicks, Anastacia Griego, Michael Edwards, Valerie Morley, Darrell Dinwiddie |
| EPI_ISL_1664670, EPI_ISL_1664723 | HELIX LLC | WHO National Influenza Centre Russian Federation | Andrey Komissarov, Artem Fadeev, Anna Ivanova, Kseniya Komissarova, Mikhail Bakaev, Tamila Musaeva, Maria Timofeeva, Veronika Eder, Maria Pisareva, Daria Danilenko, Ksenia Safina, Elena Nabieva, Georgii Bazykin, Dmitry Lioznov |
| EPI_ISL_1664740 | UMMC-Health | WHO National Influenza Centre Russian Federation | Andrey Komissarov, Artem Fadeev, Tatiana Platonova, Anna Ivanova, Kseniya Komissarova, Mikhail Bakaev, Tamila Musaeva, Maria Timofeeva, Veronika Eder, Maria Pisareva, Daria Danilenko, Ksenia Safina, Elena Nabieva, Georgii Bazykin, Dmitry Lioznov |
| EPI_ISL_1664809, EPI_ISL_1664971 | National Laboratory for Health, Environment and Food, OMM, Koper | NLZOH (National Laboratory for Health, Environment and Food) / CISLD (Clinical Institute of Special Laboratory Diagnostics), University Children's Hospital, University Medical Center Ljubljana | Sandra Janezic, Aleksander Mahnic, Maja Rupnik, Tjasa Žohar retnik, Alenka Štorman, Nika Gobec, Aleksander Kocuvan, Kaja Tominc, Maša Jari, Gašper Strugar, Tina Cvetkovi, Mitja Rak / Jernej Kova, Barbara Jenko Bizjan, Tine Tesovnik, Robert Šket, Katarina Kozmos, Ana Grom, Maruša Debeljak, Marko Pokorn, Tadej Battelino |
| EPI_ISL_1666881, EPI_ISL_1666908, EPI_ISL_1666917, EPI_ISL_1666918, EPI_ISL_1666956, EPI_ISL_1666982 | NS-QEII Health Sciences Centre | National Microbiology Laboratory (NML) | Anna Majer, Shari Tyson, Grace Seo, Philip Mabon, Elsie Grudeski, Rhiannon Huzarewich, Russell Mandes, Anneliese Landgraff, Jennifer Tanner, Natalie Knox, Morag Graham, Gary Van Domselaar, Todd Hatchette, Jason LeBlanc, Janice Pettipas, Dan Gaston, Nathalie Bastien, Yan Li, Timothy Booth, Darian Hole, Madison Chapel, Kirsten Biggar, CanCOGeN's metadata curation team, Public Health Agency of Canada CanCOGeN team |
| EPI_ISL_1667469, EPI_ISL_1667471, EPI_ISL_1667474, EPI_ISL_1667475 | Department of Laboratory Medicine, National Taiwan University Hospital | Microbial Genomics Core Lab, National Taiwan University Centers of Genomic and Precision Medicine | Shiou-Hwei Yeh, You-Yu Lin, Ya-Yun Lai, Chiao-Ling Li, Shan-Chwen Chang, Pei-Jer Chen, Sui-Yuan Chang |
| EPI_ISL_1667807 | Aegis Sciences Corporation | Centers for Disease Control and Prevention Division of Viral Diseases, Pathogen Discovery | Dakota Howard, Dhwani Batra, Peter W. Cook, Kara Moser, Adrian Paskey, Jason Caravas, Benjamin Rambo-Martin, Shatavia Morrison, Christopher Gulvick, Scott Sammons, Yvette Unoarumhi, Darlene Wagner, Matthew Schmerer, Cyndi Clark, Patrick Campbell, Rob Case, Vikramsinha Ghorpade, Holly Houdeshell, Ola Kvalvaag, Dillon Nall, Ethan Sanders, Alec Vest, Shaun Westlund, Matthew Hardison, Clinton R. Paden, Duncan MacCannell |
| EPI_ISL_1669105 | Quest Diagnostics Incorporated | Centers for Disease Control and Prevention Division of Viral Diseases, Pathogen Discovery | Dakota Howard, Dhwani Batra, Peter W. Cook, Kara Moser, Adrian Paskey, Jason Caravas, Benjamin Rambo-Martin, Shatavia Morrison, Christopher Gulvick, Scott Sammons, Yvette Unoarumhi, Darlene Wagner, Matthew Schmerer, S. H. Rosenthal, A. Gerasimova, R. M. Kagan, B. Anderson, M. Hua, Y. Liu, L.E. Bernstein, K.E. Livingston, A. Perez, I. A. Shlyakhter, R. V. Rolando, R. Owen, P. Tanpaiboon, F. Lacbawan, Clinton R. Paden, Duncan MacCannell |
| EPI_ISL_1669115, EPI_ISL_1669123 | PathWest Laboratory Medicine WA | PathWest Laboratory Medicine WA Microbial Surveillance Unit | PathWest Laboratory Medicine WA Microbial Surveillance Unit |
| EPI_ISL_1669760, EPI_ISL_1669764 | Genome Analysis Center, Yamanashi Central Hospital | Genome Analysis Center, Yamanashi Central Hospital | Yosuke Hirotsu |
| EPI_ISL_1669954, EPI_ISL_1669995 | Laboratory of virology and molecular diagnostics, Institute of Public Health | Laboratory of virology and molecular diagnostics, Institute of Public Health | Kuzmanovska M, Boshevska G, Janchevska E. |
| EPI_ISL_1669999 | General Hospital - Gostivar | Laboratory of virology and molecular diagnostics, Institute of Public Health | Kuzmanovska M, Boshevska G, Janchevska E. |
| EPI_ISL_1671016, EPI_ISL_1671053, EPI_ISL_1671065 | Austrian Agency for Health and Food Safety (AGES) | Bergthaler laboratory, CeMM Research Center for Molecular Medicine of the Austrian Academy of Sciences | Lukas Endler, Anna Schedl, Fabian Amman, Petr Triska, Thomas Penz, Benedikt Agerer, Maelle Le Moing, Michael Schuster, Bekir Erguner, Jan Laine, Martin Senekowitsch, Christoph Bock, Andreas Bergthaler |
| EPI_ISL_1671117 | Institute for Laboratory Diagnostics and Microbiology, Klinikum Klagenfurt am Wörthersee | Bergthaler laboratory, CeMM Research Center for Molecular Medicine of the Austrian Academy of Sciences | Lukas Endler, Anna Schedl, Fabian Amman, Petr Triska, Thomas Penz, Benedikt Agerer, Maelle Le Moing, Michael Schuster, Bekir Erguner, Jan Laine, Martin Senekowitsch, Christoph Bock, Andreas Bergthaler |
| EPI_ISL_1671922, EPI_ISL_1671923, EPI_ISL_1671925, EPI_ISL_1671927, EPI_ISL_1671929, EPI_ISL_1671930 | Fondation Congolaise pour la recherche medicale (FCRM), Francine Ntoumi | Institute of Tropical Medicine | Prof. Dr. Thirumalaisamy P. Velavan and Prof. Francine Ntoumi |
| EPI_ISL_1672036 | SYNLAB | GIGA Medical Genomics | Keith Durkin, Maria Artesi, Sébastien Bontems, Raphaël Boreux, Bouchra Boujemla, Nathalie Renotte, Cécile Meex, Pierrette Melin, Marie-Pierre Hayette, Vincent Bours |
| EPI_ISL_1672414 | Life Sciences Center, Vilnius University | Institute of Biotechnology, Life Sciences Center, Vilnius University | Emilija Vasiliunaite, Milda Norkiene, Danguole Ziogiene, Albertas Timinskas, Alma Gedvilaite |
| EPI_ISL_1673257 | Hospital Lahad Datu | Institute for Medical Research, Infectious Disease Research Centre, National Institutes of Health, Ministry of Health Malaysia Jalan Setia Murni U13/52, Bandar Setia Alam, 40170 Selangor, Malaysia | Suppiah J, Kamel K, Mohd Zawawi Z, Thayan R |
| EPI_ISL_1673299 | Institute for Medical Research, Infectious Disease Research Centre, National Institutes of Health, Ministry of Health Malaysia | Institute for Medical Research, Infectious Disease Research Centre, National Institutes of Health, Ministry of Health Malaysia | Suppiah J, Kamel K, Thayan R |
| EPI_ISL_1673315, EPI_ISL_1673318, EPI_ISL_1673319, EPI_ISL_1673325, EPI_ISL_1673328, EPI_ISL_1673330 | Laboratorio de Investigaciones de Baney | Swiss Tropical and Public Health Institute | Salome Hosch, Carlos Cortes, Claudia Daubenberger, Guillermo Garcia, Bonifacio Manguire Nlavo, Maximilian Mpina, Elizabeth Nyakarungu, Diosdado Odjama Nseng Ada, Mitoha Ondo O Ayekaba, Tobias Schindler, Philipp Wagner, Philip Wonder Phiri |
| EPI_ISL_1673336 | Institute for Medical Research, Infectious Disease Research Centre, National Institutes of Health, Ministry of Health Malaysia | Institute for Medical Research, Infectious Disease Research Centre, National Institutes of Health, Ministry of Health Malaysia | Suppiah J, Kamel K, Thayan R |

| | | | |
|---|---|---|---|
| EPI_ISL_1673345, EPI_ISL_1673420 | Institute for Medical Research, Infectious Disease Research Centre, National Institutes of Health, Ministry of Health Malaysia | Institute for Medical Research, Infectious Disease Research Centre, National Institutes of Health, Ministry of Health Malaysia | Suppiah J, Kamel K, Mohd Zawawi Z, Thayan R |
| EPI_ISL_1673830 | Israel Central Virology laboratory | Israel National Consortium for SARS-CoV-2 sequencing | Neta Zuckerman, Efrat Dahan Bucris, Michal Mandelboim, Dana Bar-Ilan, Oran Erster, Tzvia Mann, Omer Murik, David A. Zeevi, Assaf Rokney, Joseph Jaffe, Eva Nachum, Maya Davidovich Cohen, Ephraim Fass, Gal Zizelski Valenci, Mor Rubinstein, Efrat Rorman, Israel Nissan, Efrat Glick-Saar, Omri Nayshool, Gideon Rechavi, Ella Mendelson, Orna Mor |
| EPI_ISL_1675130 | Alaska State Virology Laboratory | Alaska State Virology Laboratory | Stephanie DeRonde, Elva House, Lisa Smith, Ph.D., Jack Chen, Ph.D. |
| EPI_ISL_1675319 | SURA | Universidad Nacional de Colombia - Laboratorio Genómico One Health | Andres F. Cardona-Rios, Maria Stella López, Daniel O. Maldonado-Perez, Simón Villegas Velásquez, Lina M. Hurtado, Laura Silvana Perez, Karl A Ciuoderis, Maria Angélica Maya, Rita Almanza Payares, Idabely Betancur Ortiz, Sandra Ines Cano, Diego A. Álvarez-Díaz, Carlos Franco-Muñoz, Katherine Laiton-Donato, Hector Alejandro Ruiz-Moreno, Marcela Mercado-Reyes, Jorge E. Osorio, Juan P. Hernandez-Ortiz |
| EPI_ISL_1676017 | Swedish national genomic surveillance program of SARS-CoV-2 | The Public Health Agency of Sweden | Maximilian Riess, Maria Lind Karlberg, Alma Brolund, Swedish national genomic surveillance program of SARS-CoV-2 |
| EPI_ISL_1677700, EPI_ISL_1677702, EPI_ISL_1677703, EPI_ISL_1677704, EPI_ISL_1677705, EPI_ISL_1677707, EPI_ISL_1677714, EPI_ISL_1677716, EPI_ISL_1677723, EPI_ISL_1677731, EPI_ISL_1677732, EPI_ISL_1677733, EPI_ISL_1677735, EPI_ISL_1677737 | | | |
| see above | National Health Laboratory | Botswana Institute for Technology Research and Innovation | Kefentse Arnold Tumedi, Madisa Mine, Dineo Emang Tshiamo. Tefelo Thela, Gape Nyepetsi, Thongbotho Mphoyakgosi, Malebogo Kebabonye, Maitshwarelo Ignatius Matsheka |
| EPI_ISL_1677770 | GMERS Medical Collage, GMERS Medical College,Civil Hospital, Sola, Ahmedabad | Gujarat Biotechnology Research Centre | Twinkle Soni,Sonal Sharma ,Zuber Saiyed,Ramesh Pandit,Janvi Raval,Zarna Patel,Nitin Savaliya,Dinesh Kumar,Nidhi Sood,Umang Mishra,Nitesh Shah,Chaitanya Joshi,Madhvi Joshi |
| EPI_ISL_1677893 | Department of Public Health Microbiology Ljubljana, National Laboratory for Health, Environment and Food | Department for Public Health Microbiology Ljubljana, National Laboratory for Health, Environment and Food | Tom Koritnik, José Gonçalves, Martin Bosilj, Natasa Berginc, Katarina Prosenc |
| EPI_ISL_1678504 | Ministry of Health Turkey | Ministry of Health Turkey | Fatma Bayrakdar, Yasemin Cogun, Süleyman Yalcin, Gülay Korukluolu |
| EPI_ISL_1678578 | HG Pharma GmbH | Bergthaler laboratory, CeMM Research Center for Molecular Medicine of the Austrian Academy of Sciences | Lukas Endler, Anna Schedl, Fabian Amman, Petr Triska, Thomas Penz, Benedikt Agerer, Maelle Le Moing, Michael Schuster, Bekir Erguner, Jan Laine, Martin Senekowitsch, Christoph Bock, Andreas Bergthaler |
| EPI_ISL_1680029 | UW Virology Lab | UW Virology Lab | Pavitra Roychoudhury, Hong Xie, Lasata Shrestha, Shah Mohamed Bakhash, Michelle Lin, Noah R. Baker, Sean Ellis, Meei-Li Huang, Keith R Jerome, Alexander Greninger |
| EPI_ISL_1681373, EPI_ISL_1681539, EPI_ISL_1681598, EPI_ISL_1681908, EPI_ISL_1682218 | Laboratory Corporation of America | Centers for Disease Control and Prevention Division of Viral Diseases, Pathogen Discovery | Dakota Howard, Dhwani Batra, Peter W. Cook, Kara Moser, Adrian Paskey, Jason Caravas, Benjamin Rambo-Martin, Shatavia Morrison, Christopher Gulvick, Scott Sammons, Yvette Unoarumhi, Darlene Wagner, Matthew Schmerer, Minoo Agarwal, Eyad Almasri, Debbie Boles, Ayla Burns, Nuthawin Charoensri, Oren Cohen, Susan Countryman, Mary Ann Cristobal, Bobbi Croy, Suzanne Dale, Hrushikesh Deshmukh, Amanda Douglas, Vincent Drouillon, Marcia Eisenberg, Howard Engler, Rama Ghatti, Prashant Gupta, Susan Hicks, Jake Humphrey, Lax Iyer, Manoj Jain, Mohan Kolli, Brian Krueger, Tim Kuphal, Stanley Letovsky, Michael Levandoski, Craig Lukasik, Jonathan Meltzer, Brian Norvell, Mindy Nye, Scott Parker, Christos Petropoulos, John Pruitt, Steven Ragan, Scott Ryan, Mike Sapeta, Jana Schroth, Suresh Babu Selvaraju, Goran Stevovic, Amanda Suchanek, Andrea Throop, Lyndon Tilson, Thomas Urban, Joe Voshell, Kimberly Wagner, Jonathan Williams, Mary Williamson, Qian Zeng, Tricia Zwiefelhofer, Clinton R. Paden, Duncan MacCannell |
| EPI_ISL_1682548 | Viollier AG | Department of Biosystems Science and Engineering, ETH Zürich | Chaoran Chen, Sarah Nadeau, Catharine Aquino, Ivan Topolsky, Philipp Jablonski, Lara Fuhrmann, David Dreifuss, Katharina Jahn, Andreia Cabral de Gouvea, Maria Domenica Moccia, Simon Grüter, Timothy Sykes, Lennart Opitz, Griffin White, Laura Neff, Doris Popovic, Andrea Patrignani, Jay Tracy, Ralph Schlapbach, Christiane Beckmann, Maurice Redondo, Olivier Kobel, Christoph Noppen, Sophie Seidel, Noemie Santamaria de Souza, Niko Beerenwinkel, Tanja Stadler |
| EPI_ISL_1683818 | Laboratory Corporation of America | Centers for Disease Control and Prevention Division of Viral Diseases, Pathogen Discovery | Dakota Howard, Dhwani Batra, Peter W. Cook, Kara Moser, Adrian Paskey, Jason Caravas, Benjamin Rambo-Martin, Shatavia Morrison, Christopher Gulvick, Scott Sammons, Yvette Unoarumhi, Darlene Wagner, Matthew Schmerer, Minoo Agarwal, Eyad Almasri, Debbie Boles, Ayla Burns, Nuthawin Charoensri, Oren Cohen, Susan Countryman, Mary Ann Cristobal, Bobbi Croy, Suzanne Dale, Hrushikesh Deshmukh, Amanda Douglas, Vincent Drouillon, Marcia Eisenberg, Howard Engler, Rama Ghatti, Prashant Gupta, Susan Hicks, Jake Humphrey, Lax Iyer, Manoj Jain, Mohan Kolli, Brian Krueger, Tim Kuphal, Stanley Letovsky, Michael Levandoski, Craig Lukasik, Jonathan Meltzer, Brian Norvell, Mindy Nye, Scott Parker, Christos Petropoulos, John Pruitt, Steven Ragan, Scott Ryan, Mike Sapeta, Jana Schroth, Suresh Babu Selvaraju, Goran Stevovic, Amanda Suchanek, Andrea Throop, Lyndon Tilson, Thomas Urban, Joe Voshell, Kimberly Wagner, Jonathan Williams, Mary Williamson, Qian Zeng, Tricia Zwiefelhofer, Clinton R. Paden, Duncan MacCannell |
| EPI_ISL_1684154 | CONSEJERIA DE SANIDAD Y ASUNTOS SOCIALES | Instituto de Salud Carlos III | Iglesias-Caballero, M. Sandonís,V. Vázquez-Morón, S. Camarero, S. Pozo, F. Casas, I. Jiménez, P. Zaballos, A. Monzón, S. Varona, S. Cuesta, I.GUTIERREZ AVILA, GONZALO |
| EPI_ISL_1684589, EPI_ISL_1684627, EPI_ISL_1684634, EPI_ISL_1684719 | Laboratory Corporation of America | Centers for Disease Control and Prevention Division of Viral Diseases, Pathogen Discovery | Dakota Howard, Dhwani Batra, Peter W. Cook, Kara Moser, Adrian Paskey, Jason Caravas, Benjamin Rambo-Martin, Shatavia Morrison, Christopher Gulvick, Scott Sammons, Yvette Unoarumhi, Darlene Wagner, Matthew Schmerer, Minoo Agarwal, Eyad Almasri, Debbie Boles, Ayla Burns, Nuthawin Charoensri, Oren Cohen, Susan Countryman, Mary Ann Cristobal, Bobbi Croy, Suzanne Dale, Hrushikesh Deshmukh, Amanda Douglas, Vincent Drouillon, Marcia Eisenberg, Howard Engler, Rama Ghatti, Prashant Gupta, Susan Hicks, Jake Humphrey, Lax Iyer, Manoj Jain, Mohan Kolli, Brian Krueger, Tim Kuphal, Stanley Letovsky, Michael Levandoski, Craig Lukasik, Jonathan Meltzer, Brian Norvell, Mindy Nye, Scott Parker, Christos Petropoulos, John Pruitt, Steven Ragan, Scott Ryan, Mike Sapeta, Jana Schroth, Suresh Babu Selvaraju, Goran Stevovic, Amanda Suchanek, Andrea Throop, Lyndon Tilson, Thomas Urban, Joe Voshell, Kimberly Wagner, Jonathan Williams, Mary Williamson, Qian Zeng, Tricia Zwiefelhofer, Clinton R. Paden, Duncan MacCannell |
| EPI_ISL_1685404 | Synlab Eesti OÜ | 1. Laboratory of Communicable Diseases (Estonia); 2. Eurofins Genomics Europe Sequencing GmbH | Liidia Dotsenko et al. |
| EPI_ISL_1687168, EPI_ISL_1687465, EPI_ISL_1687558 | Aegis Sciences Corporation | Centers for Disease Control and Prevention Division of Viral Diseases, Pathogen Discovery | Dakota Howard, Dhwani Batra, Peter W. Cook, Kara Moser, Adrian Paskey, Jason Caravas, Benjamin Rambo-Martin, Shatavia Morrison, Christopher Gulvick, Scott Sammons, Yvette Unoarumhi, Darlene Wagner, Matthew Schmerer, Cyndi Clark, Patrick Campbell, Rob Case, Vikramsinha Ghorpade, Holly Houdeshell, Ola Kvalvaag, Dillon Nall, Ethan Sanders, Alec Vest, Shaun Westlund, Matthew Hardison, Clinton R. Paden, Duncan MacCannell |
| EPI_ISL_1689242 | Lab voor klinische biologie | Lab voor klinische biologie | Marija Janevska, Hannelore Hamerlinck, Bruno Verhasselt |
| EPI_ISL_1689605, EPI_ISL_1690037, EPI_ISL_1690069, EPI_ISL_1690222, EPI_ISL_1690252 | Aegis Sciences Corporation | Centers for Disease Control and Prevention Division of Viral Diseases, Pathogen Discovery | Dakota Howard, Dhwani Batra, Peter W. Cook, Kara Moser, Adrian Paskey, Jason Caravas, Benjamin Rambo-Martin, Shatavia Morrison, Christopher Gulvick, Scott Sammons, Yvette Unoarumhi, Darlene Wagner, Matthew Schmerer, Cyndi Clark, Patrick Campbell, Rob Case, Vikramsinha Ghorpade, Holly Houdeshell, Ola Kvalvaag, Dillon Nall, Ethan Sanders, Alec Vest, Shaun Westlund, Matthew Hardison, Clinton R. Paden, Duncan MacCannell |
| EPI_ISL_1690395 | Helix/Illumina | Centers for Disease Control and Prevention Division of Viral Diseases, Pathogen Discovery | Dakota Howard, Dhwani Batra, Peter W. Cook, Kara Moser, Adrian Paskey, Jason Caravas, Benjamin Rambo-Martin, Shatavia Morrison, Christopher Gulvick, Scott Sammons, Yvette Unoarumhi, Darlene Wagner, Matthew Schmerer, Eileen de Feo, Jan Antico, Christine Tran, Matthew Tolentino, Shannon Wickline, Kim Gietzen, Brad Sickler, Jingtao Liu, Eric Allen, Phil Febbo, Nicole L. Washington, Simon White, Geraint Levan, Kelly Schiabor Barrett, Elizabeth Cirulli, Alexandre Bolze, Ary Ascencio, Charlotte Rivera-Garcia, Ryan Cho, Jason Nguyen, Sherry Wang, Jimmy Ramirez, Tyler Cassens, Efren Sandoval, Magnus Isaksson, William Lee, David Becker, Marc Laurent, James Lu, Clinton R. Paden, Duncan MacCannell |
| EPI_ISL_1692751 | Università Federico II - Dipartimento di scienze mediche traslazionali - Napoli | Telethon Institute of Genetics and Medicine (TIGEM) | Antonio Grimaldi Patrizia Annunziata Francesco Panariello Teresa Giuliano Michele Cennamo Valentina Bouche Chiara Colantuono Lucio Di Filippo Mariano Fiorenza Anna Manfredi Marcello Salvi Giuseppe Portella Andrea Ballabio Davide Cacchiarelli |
| EPI_ISL_1694212 | Infinity Biologix | Centers for Disease Control and Prevention Division of Viral Diseases, Pathogen Discovery | Dakota Howard, Dhwani Batra, Peter W. Cook, Kara Moser, Adrian Paskey, Jason Caravas, Benjamin Rambo-Martin, Shatavia Morrison, Christopher Gulvick, Scott Sammons, Yvette Unoarumhi, Darlene Wagner, Matthew Schmerer, Christian Bixby, Yihe Wang, Jonathan Schultz, Chirayu Goswami, Russ Hager, Robin Grimwood, Clinton R. Paden, Duncan MacCannell |
| EPI_ISL_1694617 | Croatian Institute for Public Health | Croatian Institute of Public Health | Irena Tabain, Ivana Ferenak |
| EPI_ISL_1695619 | Quest Diagnostics Incorporated | Centers for Disease Control and Prevention Division of Viral Diseases, Pathogen Discovery | Dakota Howard, Dhwani Batra, Peter W. Cook, Kara Moser, Adrian Paskey, Jason Caravas, Benjamin Rambo-Martin, Shatavia Morrison, Christopher Gulvick, Scott Sammons, Yvette Unoarumhi, Darlene Wagner, Matthew Schmerer, S. H. Rosenthal, A. Gerasimova, R. M. Kagan, B. Anderson, M. Hua, Y. Liu, L.E. Bernstein, K.E. Livingston, A. Perez, I. A. Shlyakhter, R. V. Rolando, R. Owen, P. Tanpaiboon, F. Lacbawan, Clinton R. Paden, Duncan MacCannell |

| | | | |
|---|---|---|---|
| EPI_ISL_1696496 | Idaho Bureau of Laboratories | Idaho Bureau of Laboratories | R. Beukelman, Matthew Charles Burns, Aimee Ceniseros, Robert L. Voermans, Christopher Ball |
| EPI_ISL_1696661 | National Virus Reference Laboratory | National Virus Reference Laboratory | Zoe Yandle, Charlene Bennett, Gabriel Gonzalez, Michael Carr, Jonathan Dean, Cillian F De Gascun |
| EPI_ISL_1696750, EPI_ISL_1696751 | Institute for Medical Research, Infectious Disease Research Centre, National Institutes of Health, Ministry of Health Malaysia | Institute for Medical Research, Infectious Disease Research Centre, National Institutes of Health, Ministry of Health Malaysia | Suppiah J, Kamel K, Mohd Zawawi Z, Thayan R |
| EPI_ISL_1697055 | Laboratoire de santé publique du Québec | Laboratoire de santé publique du Québec | Sandrine Moreira, Ioannis Ragoussis, Guillaume Bourque, Jesse Shapiro, Mark Lathrop and Michel Roger on behalf of the CoVSeQ research group (http://covseq.ca/researchgroup) |
| EPI_ISL_1697281 | New South Wales Health Pathology Royal Prince Alfred Hospital | Microbiology RPAH | Foster, C.; Au, J.; Ruiz Silva, M.; Deveson, I.; Bull, R.; Van Hal, S.; Rawlinson, W. |
| EPI_ISL_1697434 | Bioptická laborato, s.r.o. | Bioptická laborato, s.r.o. | Petr Šteiner, Tomáš Vanek, Nikola Bílá, Martina Putzová, Michaela íhová, Silva Vondráková |
| EPI_ISL_1700594 | Hospital | National Reference Center for Viruses of Respiratory Infections, Institut Pasteur, Paris | Marion Barbet, Sylvie Behillil, Méline Bizard, Angela Brisebarre, Camille Capel, Vincent Enouf, Louise Lefrançois, Frédéric Lemoine, Christophe Malabat, Corinne Maufrais, Etienne Simon-Lorière, Maud Vanpeene, Sylvie Van der Werf, Sylvaine Bastian |
| EPI_ISL_1700675, EPI_ISL_1700681, EPI_ISL_1700683, EPI_ISL_1700686, EPI_ISL_1700687 | Laboratorio de Investigaciones de Baney | Swiss Tropical and Public Health Institute | Salome Hosch, Carlos Cortes, Claudia Daubenberger, Guillermo Garcia, Bonifacio Manguire Nlavo, Maximilian Mpina, Elizabeth Nyakarungu, Diosdado Odjama Nseng Ada, Mitoha Ondo O Ayekaba, Tobias Schindler, Philipp Wagner, Philip Wonder Phiri |
| EPI_ISL_1700786, EPI_ISL_1700787 | Department of Virology | Department of Virology | Massab Umair, Aamer Ikram, Muhammad Salman, Nazish Badar, Sana Tamim, Zaira Rehman, Abdul Ahad, Shannon Whitmer, Melissa Mobley, Austin Leach, Ketan Patel, Joel Montgomery, John Klena |
| EPI_ISL_1700816 | LESP Campeche | Instituto de Diagnostico y Referencia Epidemiologicos (INDRE) | Claudia Wong-Arambula, Abril Rodriguez-Maldonado, Vanessa Rivero-Arredondo, Ariadna Medina-Benitez, Joaquin Quiroz-Mercado, Sergio Rangel-Guerrero, Natividad Cruz-Ortiz, Tatiana Nunez-Garcia, Gisela Barrera-Badillo, Lucia Hernandez-Rivas, Irma Lopez-Martinez, Ernesto Ramirez-Gonzalez. |
| EPI_ISL_1702110, EPI_ISL_1702123 | Aegis Sciences Corporation | Centers for Disease Control and Prevention Division of Viral Diseases, Pathogen Discovery | Dakota Howard, Dhwani Batra, Peter W. Cook, Kara Moser, Adrian Paskey, Jason Caravas, Benjamin Rambo-Martin, Shatavia Morrison, Christopher Gulvick, Scott Sammons, Yvette Unoarumhi, Darlene Wagner, Matthew Schmerer, Cyndi Clark, Patrick Campbell, Rob Case, Vikramsinha Ghorpade, Holly Houdeshell, Ola Kvalvaag, Dillon Nall, Ethan Sanders, Alec Vest, Shaun Westlund, Matthew Hardison, Clinton R. Paden, Duncan MacCannell |
| EPI_ISL_1702148 | Helix/Illumina | Centers for Disease Control and Prevention Division of Viral Diseases, Pathogen Discovery | Dakota Howard, Dhwani Batra, Peter W. Cook, Kara Moser, Adrian Paskey, Jason Caravas, Benjamin Rambo-Martin, Shatavia Morrison, Christopher Gulvick, Scott Sammons, Yvette Unoarumhi, Darlene Wagner, Matthew Schmerer, Eileen de Feo, Jan Antico, Christine Tran, Matthew Tolentino, Shannon Wickline, Kim Gietzen, Brad Sickler, Jingtao Liu, Eric Allen, Phil Febbo, Nicole L. Washington, Simon White, Geraint Levan, Kelly Schiabor Barrett, Elizabeth Cirulli, Alexandre Bolze, Ary Ascencio, Charlotte Rivera-Garcia, Ryan Cho, Jason Nguyen, Sherry Wang, Jimmy Ramirez, Tyler Cassens, Efren Sandoval, Magnus Isaksson, William Lee, David Becker, Marc Laurent, James Lu, Clinton R. Paden, Duncan MacCannell |
| EPI_ISL_1703064, EPI_ISL_1703434, EPI_ISL_1703524 | Dutch COVID-19 response team | National Institute for Public Health and the Environment (RIVM) | Adam Meijer, Harry Vennema, Dirk Eggink, Jeroen Cremer, Sharon van den Brink, Bas van der Veer, AnneMarie van den Brandt, Lisa Wijsman, Kim Freriks, Ryanne Jaarsma, Euníce Then, Jolienke Hardeman, Lynn Aarts, Sanne Bos, Melissa van Tuil, Robert Kohl, Linda van de Nes, Sjoerd Kuiling, James Groot, Florian Zwagemaker, Dennis Schmitz, Annelies Kroneman, Karim Hajji, Chantal Reusken, on behalf of the national COVID-19 response team |
| EPI_ISL_1704158 | ICMR-National Institute of Virology - INSACOG | NIV Influenza | Dr. Varsha Potdar |
| EPI_ISL_1704676 | SA Pathology | SA Pathology | Lex Leong, Julien Soubrier, Chuan Kok Lim, Song Gao, Mark Turra, Karin Kassahn, Ivan Bastian, Geoff Higgins |
| EPI_ISL_1704792, EPI_ISL_1704793, EPI_ISL_1704796, EPI_ISL_1704797, EPI_ISL_1704800, EPI_ISL_1704801, EPI_ISL_1704804, EPI_ISL_1704805, EPI_ISL_1704808 | SA Pathology | SA Pathology | Lex Leong, Chuan Kok Lim, Mark Turra, Ivan Bastian, Geoff Higgins |
| EPI_ISL_1704921, EPI_ISL_1704937, EPI_ISL_1704945, EPI_ISL_1704954, EPI_ISL_1704955, EPI_ISL_1704956, EPI_ISL_1704964, EPI_ISL_1704967, EPI_ISL_1704971, EPI_ISL_1704976, EPI_ISL_1704977, EPI_ISL_1704979, EPI_ISL_1704988, EPI_ISL_1705160, EPI_ISL_1705180, EPI_ISL_1705186, EPI_ISL_1705242, EPI_ISL_1705251, EPI_ISL_1705268, EPI_ISL_1705285, EPI_ISL_1705293, EPI_ISL_1705301, EPI_ISL_1705606, EPI_ISL_1705852, EPI_ISL_1705853, EPI_ISL_1705885, EPI_ISL_1705886, EPI_ISL_1705887, EPI_ISL_1705893, EPI_ISL_1705928 | | | |
| see above | Dutch COVID-19 response team | National Institute for Public Health and the Environment (RIVM) | Adam Meijer, Harry Vennema, Dirk Eggink, Jeroen Cremer, Sharon van den Brink, Bas van der Veer, AnneMarie van den Brandt, Lisa Wijsman, Kim Freriks, Ryanne Jaarsma, Euníce Then, Jolienke Hardeman, Lynn Aarts, Sanne Bos, Melissa van Tuil, Robert Kohl, Linda van de Nes, Sjoerd Kuiling, James Groot, Florian Zwagemaker, Dennis Schmitz, Annelies Kroneman, Karim Hajji, Chantal Reusken, on behalf of the national COVID-19 response team |
| EPI_ISL_1706278 | Department of Virology | Department of Virology | Massab Umair, Aamer Ikram, Muhammad Salman, Nazish Badar, Sana Tamim, Zaira Rehman, Abdul Ahad, Shannon Whitmer, Melissa Mobley, Austin Leach, Ketan Patel, Joel Montgomery, John Klena |
| EPI_ISL_1706366, EPI_ISL_1706367 | SA Pathology | SA Pathology | Lex Leong, Chuan Kok Lim, Mark Turra, Ivan Bastian, Geoff Higgins |
| EPI_ISL_1706602 | Virology Unit, Institut Pasteur du Cambodge | Virology Unit, Institut Pasteur du Cambodge | Jurre Y Siegers, Teyputita Ou, Leakhena Pum, Cecile Troupin, Ly Sovann, Kraing Sidonn, Yi Sengdoeurn, Chin Savuth, Chau Darapheak, Veasna Duong, Erik A Karlsson |
| EPI_ISL_1708008 | University Hospitals of Geneva, Laboratory of Virology | HUG, Laboratory of Virology and the Health2030 Genome Center | Samuel Cordey, Ana Rita Goncalves, Laurent Kaiser, Lorenzo Cerutti, Henri Pegeot, Melyssa Elies, Deborah Penet, Keith Harshman, Ioannis Xenarios, Emmanouil Dermitzakis |
| EPI_ISL_1708623 | Hospital | National Reference Center for Viruses of Respiratory Infections, Institut Pasteur, Paris | Marion Barbet, Sylvie Behillil, Méline Bizard, Angela Brisebarre, Camille Capel, Vincent Enouf, Louise Lefrançois, Frédéric Lemoine, Christophe Malabat, Corinne Maufrais, Etienne Simon-Lorière, Maud Vanpeene, Sylvie Van der Werf ,Pierre Alain Billy |
| EPI_ISL_1710858 | Pathogenic Microorganisms Variability Laboratory | Pathogenic Microorganisms Variability Laboratory | Alexey Shchetinin, Maria Nikiforova, Andrei Siniavin, Andrey Pochtovyy, Evgeny Usachev, Elena Shidlovskaya, Elizaveta Divisenko, Nadezhda Kuznetsova, Valeria Bacalin, Olga Burgasova, Ludmila Kolobukhina, Svetlana Smetanina, Denis Logunov, Vladimir Gushchin, Alexander Gintsburg |
| EPI_ISL_1711063 | University of Mississippi Medical Center, Department of Pathology | University of Mississippi Medical Center, Molecular and Genomics Core Facility | Ashley C. Johnson, Ithiel J. Frame, Krishna K. Ayyalasomayajula, Michael R. Garrett, D. Ashley Robinson |
| EPI_ISL_1711297, EPI_ISL_1711298, EPI_ISL_1711299, EPI_ISL_1711300, EPI_ISL_1711301, EPI_ISL_1711302 | Instituto Nacional de Medicina Genomica | Instituto Nacional de Medicina Genomica | Hidalgo-Miranda A, Mendoza-Vargas A, Reyes-Grajeda JP, Cedro-Tanda A, Alcaraz N, Gonzalez-Barrera D, Rangel-DeLeon D, Miranda-Ortiz H, Rosas-Escobar P, Canseco Mendez JC, Munguia-Garza P, Ramirez-Vega O, Garcia-Cardenas FJ, Gonzalez-Woge MA, Herrera-Montalvo LA |
| EPI_ISL_1711358 | Wichita State University - Molecular Diagnostics Lab | Kansas Health and Environmental Lab | Mike Grose, Jonathan Barnell, Ben Olsen, and Phil Adam |
| EPI_ISL_1711669, EPI_ISL_1711671, EPI_ISL_1711674 | Department of Virology | Department of Virology | Massab Umair, Aamer Ikram, Muhammad Salman, Nazish Badar, Sana Tamim, Zaira Rehman, Abdul Ahad, Shannon Whitmer, Melissa Mobley, Austin Leach, Ketan Patel, Joel Montgomery, John Klena |
| EPI_ISL_1711693, EPI_ISL_1711698 | "AR Dept. of Health-PHL, Molecular Diagnostics" | Centers for Disease Control and Prevention Division of Viral Diseases, Pathogen Discovery | Mili Sheth, Sarah Nobles, Jasmine Padilla, Mark Burroughs, Shoshona Le, Katie Dillon, Peter Cook, Clinton R. Paden, Dhwani Batra, Krista Queen, Kristen Knipe, Dakota Howard, Yvette Unoarumhi, Darlene Wagner, Matthew Schmerer, Ben L. Rambo-Martin, Kristine Lacek, Sam Shepard, Alison Laufer Halpin, Dave Wentworth, Vivien Dugan, Suxiang Tong, Justin Lee |
| EPI_ISL_1711812 | "OK Public Health Laboratory, Oklahoma State DOH" | Centers for Disease Control and Prevention Division of Viral Diseases, Pathogen Discovery | Mili Sheth, Sarah Nobles, Jasmine Padilla, Mark Burroughs, Shoshona Le, Katie Dillon, Peter Cook, Clinton R. Paden, Dhwani Batra, Krista Queen, Kristen Knipe, Dakota Howard, Yvette Unoarumhi, Darlene Wagner, Matthew Schmerer, Ben L. Rambo-Martin, Kristine Lacek, Sam Shepard, Alison Laufer Halpin, Dave Wentworth, Vivien Dugan, Suxiang Tong, Justin Lee |
| EPI_ISL_1711929 | SC Dept of Health and Env. Control-Bureau of Laboratories | Centers for Disease Control and Prevention Division of Viral Diseases, Pathogen Discovery | Mili Sheth, Sarah Nobles, Jasmine Padilla, Mark Burroughs, Shoshona Le, Katie Dillon, Peter Cook, Clinton R. Paden, Dhwani Batra, Krista Queen, Kristen Knipe, Dakota Howard, Yvette Unoarumhi, Darlene Wagner, Matthew Schmerer, Ben L. Rambo-Martin, Kristine Lacek, Sam Shepard, Alison Laufer Halpin, Dave Wentworth, Vivien Dugan, Suxiang Tong, Justin Lee |
| EPI_ISL_1711976, EPI_ISL_1711987, | Virology Unit, Institut Pasteur du Cambodge | Virology Unit, Institut Pasteur du Cambodge | Jurre Y Siegers, Teyputita Ou, Leakhena Pum, Cecile Troupin, Ly Sovann, Kraing Sidonn, Yi Sengdoeurn, Chin Savuth, Chau Darapheak, Veasna Duong, |

| | | | |
|---|---|---|---|
| EPI_ISL_1711988, EPI_ISL_1711992, EPI_ISL_1711996 | | | Erik A Karlsson |
| EPI_ISL_1712082 | Clinical Microbiology, Infection Prevention and Control | Section for Molecular Diagnostics | Björn Hallström, Jonas Björkman |
| EPI_ISL_1712296 | FSBI «NATIONAL MEDICAL RESEARCH CENTER FOR OBSTETRICS, GYNECOLOGY AND PERINATOLOGY NAMED AFTER ACADEMICIAN V.I.KULAKOV» MINISTRY OF HEALTHCARE OF THE RUSSIAN FEDERATION | Center for Precision Genome Editing and Genetic Technologies for Biomedicine, Pirogov Medical University, Moscow, Russian Federation | Yegor Botsmanov, Andrey Krivoy, Vera Belova, Anastasia Shut, Margarita Korzhanova, Dmitriy Korostin |
| EPI_ISL_1712321, EPI_ISL_1712323 | Genetica Molecular and Subdepartamento de Virologia ISP Chile | Instituto de Salud Publica de Chile | Javier Tognarelli, Karen Orostica, Barbara Parra, Loredana Arata, Gisselle Barra, Patricia Bustos, Rodrigo Fasce, Andres Castillo, Soledad Ulloa, Jorge Fernandez |
| EPI_ISL_1712385 | HOSPITAL MEXICO | Inciensa, Instituto Costarricense de Investigación y Enseñanza en Nutrición y Salud | Francisco Duarte, Hebleen Porras, Claudio Soto-Garita, Estela Cordero, Adriana Godínez, Melany Calderón, José Luis Vargas, Mariela Gutiérrez, Joselyn Prado & Teresita Somogyi |
| EPI_ISL_1712386 | AREA DE SALUD OREAMUNO-PACAYAS-TIERRA BLANCA | Inciensa, Instituto Costarricense de Investigación y Enseñanza en Nutrición y Salud | Francisco Duarte, Hebleen Porras, Claudio Soto-Garita, Estela Cordero, Adriana Godínez, Melany Calderón, José Luis Vargas, Mariela Gutiérrez, Joselyn Prado & Carolina Loría Acosta |
| EPI_ISL_1712389 | AREA DE SALUD TURRIALBA-JIMENEZ | Inciensa, Instituto Costarricense de Investigación y Enseñanza en Nutrición y Salud | Francisco Duarte, Hebleen Porras, Claudio Soto-Garita, Estela Cordero, Adriana Godínez, Melany Calderón, José Luis Vargas, Mariela Gutiérrez, Joselyn Prado & Mónica Charpentier-Artavia |
| EPI_ISL_1712393 | HOSPITAL DR. TONY FACIO CASTRO | Inciensa, Instituto Costarricense de Investigación y Enseñanza en Nutrición y Salud | Francisco Duarte, Hebleen Porras, Claudio Soto-Garita, Estela Cordero, Adriana Godínez, Melany Calderón, José Luis Vargas, Mariela Gutiérrez, Joselyn Prado & Marcela Rodríguez-Gómez |
| EPI_ISL_1712410 | AREA DE SALUD ALAJUELITA | Inciensa, Instituto Costarricense de Investigación y Enseñanza en Nutrición y Salud | Francisco Duarte, Hebleen Porras, Claudio Soto-Garita, Estela Cordero, Adriana Godínez, Melany Calderón, José Luis Vargas, Mariela Gutiérrez, Joselyn Prado & Jonathan Rojas |
| EPI_ISL_1712418 | AREA DE SALUD NARANJO | Inciensa, Instituto Costarricense de Investigación y Enseñanza en Nutrición y Salud | Francisco Duarte, Hebleen Porras, Claudio Soto-Garita, Estela Cordero, Adriana Godínez, Melany Calderón, José Luis Vargas, Mariela Gutiérrez, Joselyn Prado & Teresita Somogyi |
| EPI_ISL_1713239, EPI_ISL_1713251, EPI_ISL_1713266, EPI_ISL_1713267, EPI_ISL_1713268, EPI_ISL_1713421, EPI_ISL_1713422, EPI_ISL_1713424, EPI_ISL_1713430, EPI_ISL_1713457, EPI_ISL_1713464, EPI_ISL_1713465, EPI_ISL_1713516, EPI_ISL_1713553, EPI_ISL_1713555, EPI_ISL_1713557, EPI_ISL_1713558, EPI_ISL_1713559, EPI_ISL_1713592, EPI_ISL_1713708, EPI_ISL_1713746, EPI_ISL_1713854, EPI_ISL_1713924 | | | |
| see above | Ministry of Public Health / Hamad Medical Corporation | Biomedical Research Center (BRC), Qatar University / Qatar Genome Project (QGP) | BRC: Fatiha M. Benslimane, Heba A. Al-Khatib, Oal Al-Jamal, Dana Al-Batesh, Hadi M. Yassine, Asmaa A. Al-Thani. MOPH and HMC: Abdullatif Al-Khal, Muna A. S. Al-Maslamani, Mashael A. Al-Bader, Hamda Alromaihi, Roberto Bertollini, Peter V. Coyle, Einas A. E. Al-Kuwari, Hamad E. Al-Romaihi, Salih Al-Marri, Mohammed Al-Thani, Reham A. El-Kahlout. QBB: Tasneem Al-Hamad, Dina Elgakhlab QGP: Fatima H. Al-Kuwari, Chadi Saad |
| EPI_ISL_1714174, EPI_ISL_1714289, EPI_ISL_1714300, EPI_ISL_1714328, EPI_ISL_1714343, EPI_ISL_1714495, EPI_ISL_1714520, EPI_ISL_1714539 | Ministry of Public Health / Hamad Medical Corporation | Weill Cornell Medical College - Qatar (WCM-Q), Genomics Core Laboratory / Qatar Genome Project (QGP) | WCMQ: Ayeda A. Ahmed, Meryem Bensaad, Shameem Younuskunju, Yasmin Mohamoud, Laith Abu-Raddad, Joel A Malek. QGP: Fatima H. Al-Kuwari, Chadi Saad MOPH and HMC: Abdullatif Al-Khal, Muna A. S. Al-Maslamani, Mashael A. Al-Bader, Hamda Alromaihi, Roberto Bertollini, Peter V. Coyle, Einas A. E. Al-Kuwari, Hamad E. Al-Romaihi, Salih Al-Marri, Mohammed Al-Thani, Reham A. El-Kahlout. QBB: Tasneem Al-Hamad, Dina Elgakhlab |
| EPI_ISL_1715135 | CS II Dr Jose Ferreira Telles | Instituto Adolfo Lutz, Interdiciplinary Procedures Center, Strategic Laboratory | Claudio Tavares Sacchi, Claudia Regina Gonçalves, Erica Valessa Ramos Gomes, Karoline Rodrigues Campos, Caio Vinicius Dias Lopes, Leonardo Jose Tadeu de Araujo, Katia Correa de Oliveira Santos |
| EPI_ISL_1715169 | Virology Laboratory, International Centre for Diarrhoeal Disease Research, Bangladesh (ICDDR,B) | Virology Laboratory, International Centre for Diarrhoeal Disease Research, Bangladesh (ICDDR,B) | Mohammad Enayet Hossain, Mojnu Miah, Rashedul Hasan, Md. Mahfuzur Rahman, Mohammed Ziaur Rahman, Mustafizur Rahman |
| EPI_ISL_1715196 | National Public Health Laboratory, Cameroon | African Centre of Excellence for Genomics of Infectious Diseases, Redeemer's University | Olawoye, I.B. et al |
| EPI_ISL_1715236 | Zakad Genetyki Synevo | 39. Tricity SARS-CoV-2 sequencing consortium: University of Gdansk, Medical University of Gdansk, Vaxican Ltd., Invicta Ltd. 2. National Institute of Public Health - National Institute of Hygiene, Warsaw, Poland | Maciej Kosinski, Celina Cybulska, Krystyna Bienkowska Szewczyk, Maciej Grzybek, Karolina Gackowska, Katarzyna Groth, Lukasz Rabalski, Katarzyna Zacharczuk, Magdalena Nowakowska, Magorzata Sadkowska-Todys, Tomasz Wokowicz |
| EPI_ISL_1715359 | WSSE w Gdasku | 183. Tricity SARS-CoV-2 sequencing consortium: University of Gdansk, Medical University of Gdansk, Vaxican Ltd., Invicta Ltd. 2. National Institute of Public Health - National Institute of Hygiene, Warsaw, Poland | Maciej Kosinski, Celina Cybulska, Krystyna Bienkowska Szewczyk, Maciej Grzybek, Karolina Gackowska, Katarzyna Groth, Lukasz Rabalski, Katarzyna Zacharczuk, Magdalena Nowakowska, Magorzata Sadkowska-Todys, Tomasz Wokowicz |
| EPI_ISL_1715390 | Nigerian Centre for Disease Control (NCDC) | African Centre of Excellence for Genomics of Infectious Diseases, Redeemer's University | Olawoye, I.B. et al |
| EPI_ISL_1716438 | Laboratorio Saude | LGBio (Laboratorio de Genetica & Biodiversidade) | Mariana Pires de Campos Telles, Daniela de Melo e Silva, Elisangela de Paula Silveira Lacerda, Renata de Oliveira Dias, Rhewter Nunes, Cintia Pelegrineti Targueta de Azevedo Brito, Ramilla dos Santos Braga, Thais Guimarães Castro, Thays Millena Alves Pedroso, Amanda Alves de Melo, Francyelli Mello Andrade |
| EPI_ISL_1716727 | Biomedical Research Foundation of the Academy of Athens (BRFAA) | Greek Genome Center, Biomedical Research Foundation of the Academy of Athens (BRFAA) | Emmanouil Athanasiadis, Ioannis Vatsellas, Theodoros Loupis, Katerina Zoi, Dimitrios Thanos |
| EPI_ISL_1717033, EPI_ISL_1717036, EPI_ISL_1717039, EPI_ISL_1717041, EPI_ISL_1717042, EPI_ISL_1717065, EPI_ISL_1718053, EPI_ISL_1718056 | Centre for Dengue Research and AICBU, Department of Immunology and Molecular Medicine | Centre for Dengue Research and AICBU, Department of Immunology and Molecular Medicine | Chandima Jeewandara, Deshni Jayathilaka, Dinuka Ariyaratne, Tibutius Thanesh Pramanayagam, Diyanath Ranasinghe, Laksiri Gomes, Gathsaurie Neelika Malavige |
| EPI_ISL_1718180 | Pandemic Response Lab - NYC | Pandemic Response Lab, R&D | Henry Lee, Michael Hammerling, Melissa Hopkins, Cybill del Castillo, Shinyoung Clair Kang, William Ward, Pradeep Bugga, Sol Rey, Dylan Law, Katharine Nelson, Haiping Hao, Jon Laurent |
| EPI_ISL_1718284, EPI_ISL_1718285, EPI_ISL_1718286, EPI_ISL_1718290, EPI_ISL_1718293 | National Center of Disease Control and Prevention of the Republic of Armenia | Institute of Molecular Biology NAS RA, Republic of Armenia, Department of Bioengineering, BioinformaticsInstitute and Molecular Biology IBMPh RAU, Republic of Armenia | Arsen Arakelyan, Diana Avetyan, Siras Hakobyan, Gisane Khachatyan, Maria Nikoghosyan, Tamara Sirunyan, Nelli Muradyan, Andranik Chavushyan, Hovsep Ghazaryan, Roksana Zakharyan, Shushan Sargrsyan, Gayane Melik-Pashayan |
| EPI_ISL_1719883 | National Public Health Laboratory, National Centre for Infectious Diseases | National Public Health Laboratory, National Centre for Infectious Diseases | Tze Minn Mak, Zhenyang Zhou, Grace Jie Yin Ngan, Royce Ang, Lin Cui, Raymond Tzer Pin Lin |
| EPI_ISL_1722306 | GRMDCR & JU | Genomic Research Lab, BCSIR | Barna Goswami, Md. Murshed Hasan Sarkar, Mohammad Samir Uzzaman, Eshrar Osman, Md. Ahasan Habib, Shahina Akter, Tanjina Akhter Banu, Abu Sayeed Mohammad Mahmud, Iffat Jahan, Md. Saddam Hossain, Mohammad Mohi Uddin, Md. Kamrul Islam, Nihad Adnan, Mohd Raeed Jamiruddin, M Ahsanul Haq, Mohib Ullah Khondoker, Maha Jamiruddin, Md. Rubel Hossain, Nowshin Jahan, Tamanna Ali, Shahad Saif Khandker, M Firoz Ahmed, Md. Salim Khan |
| EPI_ISL_1729652, EPI_ISL_1729672 | SYNLAB MVZ Trier | Robert Koch Institute | unknown |
| EPI_ISL_1731212 | National Virus Reference Laboratory | National Virus Reference Laboratory | Zoe Yandle, Charlene Bennett, Gabriel Gonzalez, Michael Carr, Jonathan Dean, Cillian F De Gascun |
| EPI_ISL_1731549, EPI_ISL_1731563 | MRCG at LSHTM Genomics lab | MRCG at LSHTM Genomics lab | Abdul Karim sesay, Abdoulie Kanteh, Jarra Manneh, Mariama Kujabi, Bakary Sanyang |
| EPI_ISL_1731579 | Instituto Adolfo Lutz - Regional de Aracatuba | Instituto Adolfo Lutz, Interdiciplinary Procedures Center, Strategic Laboratory | Claudio Tavares Sacchi, Claudia Regina Gonçalves, Erica Valessa Ramos Gomes, Karoline Rodrigues Campos, Caio Vinicius Dias Lopes, Leonardo Jose Tadeu de Araujo, Katia Correa de Oliveira Santos |
| EPI_ISL_1731586, EPI_ISL_1731593 | Instituto Adolfo Lutz Central | Instituto Adolfo Lutz, Interdiciplinary Procedures Center, Strategic Laboratory | Claudio Tavares Sacchi, Claudia Regina Gonçalves, Erica Valessa Ramos Gomes, Karoline Rodrigues Campos, Caio Vinicius Dias Lopes, Leonardo Jose Tadeu de Araujo, Katia Correa de Oliveira Santos |

| | | | |
|---|---|---|---|
| EPI_ISL_1731851 | Synlab Eesti OÜ | 1. Laboratory of Communicable Diseases (Estonia); 2. Eurofins Genomics Europe Sequencing GmbH | Liidia Dotsenko et al. |
| EPI_ISL_1732070 | North Estonia Medical Centre | 1. Laboratory of Communicable Diseases (Estonia); 2. Eurofins Genomics Europe Sequencing GmbH | Liidia Dotsenko et al. |
| EPI_ISL_1732274, EPI_ISL_1732277 | Department of Virology | Department of Virology | Massab Umair, Aamer Ikram, Muhammad Salman, Nazish Badar, Sana Tamim, Zaira Rehman, Abdul Ahad, Ammar Amjad, Adnan Haider, Qasim Ali |
| EPI_ISL_1732291 | Kansas Health and Environmental Lab | Kansas Health and Environmental Lab | Mike Grose, Jonathan Barnell, Ben Olsen, and Phil Adam |
| EPI_ISL_1732294 | Wichita State University - Molecular Diagnostics Lab | Kansas Health and Environmental Lab | Mike Grose, Jonathan Barnell, Ben Olsen, and Phil Adam |
| EPI_ISL_1734084 | Fulgent Genetics | Centers for Disease Control and Prevention Division of Viral Diseases, Pathogen Discovery | Dakota Howard, Dhwani Batra, Peter W. Cook, Kara Moser, Adrian Paskey, Jason Caravas, Benjamin Rambo-Martin, Shatavia Morrison, Christopher Gulvick, Scott Sammons, Yvette Unoarumhi, Darlene Wagner, Matthew Schmerer, Harry Gao, Mickey Li, John Gao, Joseph Fierro, Benafsh Sapra, Becky Tsai, Yan Meng, Doreen Ng, James Xie, Clinton R. Paden, Duncan MacCannell |
| EPI_ISL_1734604 | Helix/Illumina | Centers for Disease Control and Prevention Division of Viral Diseases, Pathogen Discovery | Dakota Howard, Dhwani Batra, Peter W. Cook, Kara Moser, Adrian Paskey, Jason Caravas, Benjamin Rambo-Martin, Shatavia Morrison, Christopher Gulvick, Scott Sammons, Yvette Unoarumhi, Darlene Wagner, Matthew Schmerer, Eileen de Feo, Jan Antico, Christine Tran, Matthew Tolentino, Shannon Wickline, Kim Gietzen, Brad Sickler, Jingtao Liu, Eric Allen, Phil Febbo, Nicole L. Washington, Simon White, Geraint Levan, Kelly Schiabor Barrett, Elizabeth Cirulli, Alexandre Bolze, Ary Ascencio, Charlotte Rivera-Garcia, Ryan Cho, Jason Nguyen, Sherry Wang, Jimmy Ramirez, Tyler Cassens, Efren Sandoval, Magnus Isaksson, William Lee, David Becker, Marc Laurent, James Lu, Clinton R. Paden, Duncan MacCannell |
| EPI_ISL_1734850, EPI_ISL_1734853, EPI_ISL_1734857, EPI_ISL_1734874, EPI_ISL_1734877, EPI_ISL_1734883 | Instituto de Biotecnologia - UNESP-Botucatu-SP | Instituto de Biotecnologia - UNESP-Botucatu-SP | Fábio Sossai Possebon; Leila Sabrina Ullmann; Cecília Artico Banho; Cíntia Bittar; Guilherme Campos; Helena Lage Ferreira; Jorge A. Petroli Marchesi; Lívia Sacchetto; Maisa C. Pereira Parra; Marília Moraes; Maurício L. Nogueira; Paula Rahal; Paulo Inacio da Costa; João Pessoa Araújo Jr. |
| EPI_ISL_1735269 | Helix/Illumina | Centers for Disease Control and Prevention Division of Viral Diseases, Pathogen Discovery | Dakota Howard, Dhwani Batra, Peter W. Cook, Kara Moser, Adrian Paskey, Jason Caravas, Benjamin Rambo-Martin, Shatavia Morrison, Christopher Gulvick, Scott Sammons, Yvette Unoarumhi, Darlene Wagner, Matthew Schmerer, Eileen de Feo, Jan Antico, Christine Tran, Matthew Tolentino, Shannon Wickline, Kim Gietzen, Brad Sickler, Jingtao Liu, Eric Allen, Phil Febbo, Nicole L. Washington, Simon White, Geraint Levan, Kelly Schiabor Barrett, Elizabeth Cirulli, Alexandre Bolze, Ary Ascencio, Charlotte Rivera-Garcia, Ryan Cho, Jason Nguyen, Sherry Wang, Jimmy Ramirez, Tyler Cassens, Efren Sandoval, Magnus Isaksson, William Lee, David Becker, Marc Laurent, James Lu, Clinton R. Paden, Duncan MacCannell |
| EPI_ISL_1736692, EPI_ISL_1737433, EPI_ISL_1737545, EPI_ISL_1737567, EPI_ISL_1737850 | Aegis Sciences Corporation | Centers for Disease Control and Prevention Division of Viral Diseases, Pathogen Discovery | Dakota Howard, Dhwani Batra, Peter W. Cook, Kara Moser, Adrian Paskey, Jason Caravas, Benjamin Rambo-Martin, Shatavia Morrison, Christopher Gulvick, Scott Sammons, Yvette Unoarumhi, Darlene Wagner, Matthew Schmerer, Cyndi Clark, Patrick Campbell, Rob Case, Vikramsinha Ghorpade, Holly Houdeshell, Ola Kvalvaag, Dillon Nall, Ethan Sanders, Alec Vest, Shaun Westlund, Matthew Hardison, Clinton R. Paden, Duncan MacCannell |
| EPI_ISL_1737927, EPI_ISL_1738322, EPI_ISL_1738363, EPI_ISL_1738604 | Quest Diagnostics Incorporated | Centers for Disease Control and Prevention Division of Viral Diseases, Pathogen Discovery | Dakota Howard, Dhwani Batra, Peter W. Cook, Kara Moser, Adrian Paskey, Jason Caravas, Benjamin Rambo-Martin, Shatavia Morrison, Christopher Gulvick, Scott Sammons, Yvette Unoarumhi, Darlene Wagner, Matthew Schmerer, S. H. Rosenthal, A. Gerasimova, R. M. Kagan, B. Anderson, M. Hua, Y. Liu, L.E. Bernstein, K.E. Livingston, A. Perez, I. A. Shlyakhter, R. V. Rolando, R. Owen, P. Tanpaiboon, F. Lacbawan, Clinton R. Paden, Duncan MacCannell |
| EPI_ISL_1738785, EPI_ISL_1738787, EPI_ISL_1738789, EPI_ISL_1738790 | Laboratorio de Salud Pública Bogota | Gencore - Universidad de los Andes | Marcela Guevara, Luisa Sacristan, David Gonzalez, Silvia Restrepo, Ana Maria Palacio, Johana Hernandez, Gabriela Delgado, Alejandro Gomez |
| EPI_ISL_1738804 | Nebraska Public Health Laboratory | NPHL COVID-19 Response Team | NPHL COVID-19 Response Team |
| EPI_ISL_1738807 | Institute of Microbiology, Universidad San Francisco de Quito | Institute of Microbiology, Universidad San Francisco de Quito | Sully Márquez, Belén Prado-Vivar, Juan José Guadalupe, Monica Becerra-Wong, Bernardo Gutiérrez, Manuel Jaramillo, Verónica Barragán, Patricio Rojas-Silva, Gabriel Trueba, Michelle Grunauer, Paúl Cárdenas |
| EPI_ISL_1739220 | unknown | Instituto Nacional de Saude (INSA) | Borges et al |
| EPI_ISL_1739244 | Institute of Microbiology, Universidad San Francisco de Quito | Institute of Microbiology, Universidad San Francisco de Quito | Sully Márquez, Belén Prado-Vivar, Juan José Guadalupe, Monica Becerra-Wong, Bernardo Gutiérrez, Gabriela Zambrano, Verónica Barragán, Patricio Rojas-Silva, Gabriel Trueba, Michelle Grunauer, Paúl Cárdenas |
| EPI_ISL_1739309 | Hospital | National Reference Center for Viruses of Respiratory Infections, Institut Pasteur, Paris | Marion Barbet, Sylvie Behillil, Méline Bizard, Angela Brisebarre, Camille Capel, Vincent Enouf, Louise Lefrançois, Frédéric Lemoine, Christophe Malabat, Corinne Maufrais, Gael Millot, Etienne Simon-Lorière, Maud Vanpeene, Sylvie Van der Werf ,Nabil Gastli |
| EPI_ISL_1740138, EPI_ISL_1740369 | Ontario's COVID-19 Genomics Rapid Response Coalition | McMaster University | Allison McGeer, Patryk Aftanas, Hooman Derakhshani, Angel Li, Kuganya Nirmalarajah, Emily Panousis, Ahmed Draia, Jalees Nasir, Michael Surette, Samira Mubareka, Andrew G. McArthur |
| EPI_ISL_1742835 | GRMDCR & JU | Genomic Research Lab, BCSIR | Md. Murshed Hasan Sarkar, Mohammad Samir Uzzaman, Eshrar Osman, Md. Ahasan Habib, Shahina Akter, Tanjina Akhter Banu, Abu Sayeed Mohammad Mahmud, Barna Goswami, Iffat Jahan, Mohammad Mohi Uddin, Md. Kamrul Islam, Nihad Adnan, Mohd Raeed Jamiruddin, M Ahsanul Haq, Mohib Ullah Khondoker, Maha Jamiruddin, Md. Rubel Hossain, Nowshin Jahan, Tamanna Ali, Shahad Saif Khandker, M Firoz Ahmed, Md. Salim Khan |
| EPI_ISL_1742838 | GRMDCR & JU | Genomic Research Lab, BCSIR | Md. Murshed Hasan Sarkar, Mohammad Samir Uzzaman, Eshrar Osman, Md. Ahasan Habib, Shahina Akter, Tanjina Akhter Banu, Abu Sayeed Mohammad Mahmud, Barna Goswami, Iffat Jahan, Md. Saddam Hossain, Mohammad Mohi Uddin, Md. Kamrul Islam, Nihad Adnan, Mohd Raeed Jamiruddin, M Ahsanul Haq, Mohib Ullah Khondoker, Maha Jamiruddin, Md. Rubel Hossain, Nowshin Jahan, Tamanna Ali, Shahad Saif Khandker, M Firoz Ahmed, Md. Salim Khan |
| EPI_ISL_1744304 | Broad Institute Clinical Research Sequencing Platform | Infectious Disease Program, Broad Institute of Harvard and MIT | Siddle,K.J., Adams,G., Pearlman,L., Gladden-Young,A., Vicente,G., Blumenstiel,B., DeFelice,M., Lee,M., McGovern,S., Lagerborg,K., Rudy,M., DeRuff,K., Carter,A., Normandin,E., Bauer,M., Reilly,S., Tomkins-Tinch,C., Loreth,C., Chaluvadi,S., Meldrim,J., Granger,B., Lemieux,J.E., Birren,B.W., Sabeti,P.C., Larkin,K., Dodge,S., Lennon,N., Madoff,L., Brown,C., Gallagher,G., Smole,S., Park,D.J., Gabriel,S., and MacInnis,B.L. |
| EPI_ISL_1745724 | WSSE Laboratorium Mikrobiologii i Parazytologii | 1. National Institute of Public Health - National Institute of Hygiene; 2. Eurofins Genomics Europe Sequencing GmbH | Wokowicz Tomasz, Zacharczuk Katarzyna, Sadkowska-Todys Magorzata, Gierczyki Rafa, Eurofins Genomics Europe Sequencing Team, ECDC COVID-19 WGS support team |
| EPI_ISL_1746434 | Servicio de Microbiología Clínica (Complejo Hospitalario de Navarra, Pamplona) | Centro de Secuenciación NASERTIC | Carmen Ezpeleta Baquedano, Ana Navascués, Ana Miqueleiz |
| EPI_ISL_1747987 | Viollier AG | Clinical Bacteriology | Tim Roloff, Madlen Stange, Helena MB Seth-Smith, Alfredo Mari, Karoline Leuzinger, Julia Bielicki, Christiane Beckmann, Manuel Battegay, Hans Hirsch, Adrian Egli |
| EPI_ISL_1749448, EPI_ISL_1749520, EPI_ISL_1749579, EPI_ISL_1749583 | Public Health Authority of the Slovak Republic | Laboratory of Genomics and Bioinformatics, Comenius University Science Park | Tatiana Sedláková, Diana Rusáková, Miroslav Böhmer, Anna Giová, Jaroslav Budiš, Tomáš Szemes |
| EPI_ISL_1750958 | Institute of Epidemiology, Disease Control and Research (IEDCR) | Institute for Developing Science and Health Initiatives (ideSHi) | Hassan Afrad, Sadia Rahman, Fidausi Qadri, Tahmina Shirin |
| EPI_ISL_1750965 | PathWest Laboratory Medicine WA | PathWest Laboratory Medicine WA Microbial Surveillance Unit | PathWest Laboratory Medicine WA Microbial Surveillance Unit |
| EPI_ISL_1752643 | Instituto Adolfo Lutz - Regional de Aracatuba | Instituto Adolfo Lutz, Interdiciplinary Procedures Center, Strategic Laboratory | Claudio Tavares Sacchi, Claudia Regina Gonçalves, Erica Valessa Ramos Gomes, Karoline Rodrigues Campos, Caio Vinicius Dias Lopes, Leonardo Jose Tadeu de Araujo, Katia Correa de Oliveira Santos |
| EPI_ISL_1752690 | Child Health Research Foundation | Child Health Research Foundation | CHRF Bangladesh Genomics Team |
| EPI_ISL_1753025 | Laboratorio de Investigaciones de Baney | Swiss Tropical and Public Health Institute | Salome Hosch, Carlos Cortes, Claudia Daubenberger, Guillermo Garcia, Bonifacio Manguire Nlavo, Maximilian Mpina, Elizabeth Nyakarungu, Diosdado Odjama Nseng Ada, Mitoha Ondo O Ayekaba, Tobias Schindler, Philipp Wagner, Philip Wonder Phiri |
| EPI_ISL_1753834 | Jessa | Jessa | Cruys et al. on behalf of the Jessa_cmdLab |
| EPI_ISL_1754139, EPI_ISL_1754140 | LESP Campeche | Instituto de Diagnostico y Referencia Epidemiologicos (INDRE) | Claudia Wong-Arambula, Abril Rodriguez-Maldonado, Vanessa Rivero-Arredondo, Ariadna Medina-Benitez, Joaquin Quiroz-Mercado, Sergio Rangel-Guerrero, Natividad Cruz-Ortiz, Tatiana Nunez-Garcia, Gisela Barrera-Badillo, Lucia Hernandez-Rivas, Irma Lopez-Martinez, Ernesto Ramirez-Gonzalez. |

| | | | |
|---|---|---|---|
| EPI_ISL_1754141 | LESP Baja California | Instituto de Diagnostico y Referencia Epidemiologicos (INDRE) | Claudia Wong-Arambula, Abril Rodriguez-Maldonado, Vanessa Rivero-Arredondo, Ariadna Medina-Benitez, Joaquin Quiroz-Mercado, Sergio Rangel-Guerrero, Natividad Cruz-Ortiz, Tatiana Nunez-Garcia, Gisela Barrera-Badillo, Lucia Hernandez-Rivas, Irma Lopez-Martinez, Ernesto Ramirez-Gonzalez. |
| EPI_ISL_1789696, EPI_ISL_1789697, EPI_ISL_1789698, EPI_ISL_1789699 | Laboratorio PGM | Laboratorio de Infectología Molecular, Departamento de Bioquímica y Medicina Molecular,Facultad de Medicina - Universidad Autónoma de Nuevo León | Kame A. Galán-Huerta, María F. Herrera-Saldivar, Natalia Martínez-Acuña, Sonia A. Lozano-Sepúlveda, Daniel Arellanos-Soto, Ana M. Rivas-Estilla, Javier Ramos-Jimenez, Gabriela Elizondo, Eduardo Garza-de-la-Peña |
| EPI_ISL_402125 | National Institute for Communicable Disease Control and Prevention (ICDC) Chinese Center for Disease Control and Prevention (China CDC) | National Institute for Communicable Disease Control and Prevention (ICDC) Chinese Center for Disease Control and Prevention (China CDC) | Zhang,Y.-Z., Wu,F., Chen,Y.-M., Pei,Y.-Y., Xu,L., Wang,W., Zhao,S., Yu,B., Hu,Y., Tao,Z.-W., Song,Z.-G., Tian,J.-H., Zhang,Y.-L., Liu,Y., Zheng,J.-J., Dai,F.-H., Wang,Q.-M., She,J.-L. and Zhu,T.-Y. |
| EPI_ISL_406798 | General Hospital of Central Theater Command of People's Liberation Army of China | BGI & Institute of Microbiology, Chinese Academy of Sciences & Shandong First Medical University & Shandong Academy of Medical Sciences & General Hospital of Central Theater Command of People's Liberation Army of China | Weijun Chen, Yuhai Bi, Weifeng Shi and Zhenhong Hu |
| EPI_ISL_407071 | Respiratory Virus Unit, Microbiology Services Colindale, Public Health England | Respiratory Virus Unit, Microbiology Services Colindale, Public Health England | Monica Galiano, Shahjahan Miah, Richard Myers, Angie Lackenby, Omolola Akinbami, Tiina Talts, Leena Bhaw, Kirstin Edwards, Jonathan Hubb, Joanna Ellis, Maria Zambon |
| EPI_ISL_407073 | Respiratory Virus Unit, Microbiology Services Colindale, Public Health England | Respiratory Virus Unit, Microbiology Services Colindale, Public Health England | Monica Galiano, Shahjahan Miah, Richard Myers, Angie Lackenby, Omolola Akinbami, Tiina Talts, Leena Bhaw, Kirstin Edwards, Jonathan Hubb, Joanna Ellis, Maria Zambon. |
| EPI_ISL_407079 | Lapland Central Hospital | Department of Virology, University of Helsinki and Helsinki University Hospital, Helsinki, Finland | Teemu Smura, Suvi Kuivanen, Hannimari Kallio-Kokko, Olli Vapalahti |
| EPI_ISL_407193 | Korea Centers for Disease Control & Prevention (KCDC) Center for Laboratory Control of Infectious Diseases Division of Viral Diseases | Korea Centers for Disease Control & Prevention (KCDC) Center for Laboratory Control of Infectious Diseases Division of Viral Diseases | Jeong-Min Kim, Yoon-Seok Chung, Namjoo Lee, Mi-Seon Kim, SangHee Woo, Hye-Joon Jo, Sehee Park, Heui Man Kim, Myung Guk Han |
| EPI_ISL_407894 | Pathology Queensland | Public Health Virology Laboratory | Ben Huang, Alyssa Pyke, Amanda De Jong, Andrew Van Den Hurk, Carmel Taylor, David Warrilow, Doris Genge, Elisabeth Gamez, Glen Hewitson, Ian Maxwell Mackay, Inga Sultana, Jamie McMahon, Jean Barcelon, Judy Northill, Mitchell Finger, Natalie Simpson, Neelima Nair, Peter Burtonclay, Peter Moore, Sarah Wheatley, Sean Moody, Sonja Hall-Mendelin, Timothy Gardam, and Frederick Moore. |
| EPI_ISL_407976 | KU Leuven, Clinical and Epidemiological Virology | KU Leuven, Clinical and Epidemiological Virology | Bert Vanmechelen, Elke Wollants, Annabel Rector, Els Keyaerts, Lies Laenen, Marc Van Ranst, and Piet Maes |
| EPI_ISL_408431 | Sorbonne Université, Inserm et Assistance Publique-Hôpitaux de Paris (Pitié Salpétrière) | National Reference Center for Viruses of Respiratory Infections, Institut Pasteur, Paris | Mélanie Albert, Marion Barbet, Sylvie Behillil, Méline Bizard, Angela Brisebarre, Flora Donati, Vincent Enouf, Maud Vanpeene, Sylvie van der Werf, Sonia Burrel, Anne-Geneviève Marcelin, Vincent Calvez, David Boutolleau, Elise Klément, Valérie Pourcher, Eric Caumes. |
| EPI_ISL_408665, EPI_ISL_408666 | Dept. of Virology III, National Institute of Infectious Diseases | Pathogen Genomics Center, National Institute of Infectious Diseases | Tsuyoshi Sekizuka, Shutoku Matsuyama, Naganori Nao, Kazuya Shirato, Makoto Takeda, Makoto Kuroda |
| EPI_ISL_410301 | National Influenza Centre, National Public Health Laboratory, Kathmandu, Nepal | The University of Hong Kong | Ranjit Sah , Runa Jha, Daniel Chu, Haogao Gu, Malik Peiris, Anup Bastola, Alfonso J. Rodriguez-Morales, Bibek Kumar Lal, Basu Dev Pandey, Leo Poon |
| EPI_ISL_410532 | Dept. of Pathology, National Institute of Infectious Diseases | Pathogen Genomics Center, National Institute of Infectious Diseases | Tsuyoshi Sekizuka, Harutaka Katano, Shutoku Matsuyama, Naganori Nao, Kazuya Shirato, Motoi Suzuki, Hideki Hasegawa, Takaji Wakita, Makoto Takeda, Tadaki Suzuki, Makoto Kuroda |
| EPI_ISL_410536 | Singapore General Hospital, Molecular Laboratory, Division of Pathology | Programme in Emerging Infectious Diseases, Duke-NUS Medical School | Danielle E Anderson, Martin Linster, Yan Zhuang, Jayanthi Jayakumar, Kian Sing Chan, Lynette LE Oon, Shirin Kalimuddin, Jenny GH Low, Yvonne CF Su, Gavin JD Smith |
| EPI_ISL_412869, EPI_ISL_412870 | Division of Viral Diseases, Center for Laboratory Control of Infectious Diseases, Korea Centers for Diseases Control and Prevention | Division of Viral Diseases, Center for Laboratory Control of Infectious Diseases, Korea Centers for Diseases Control and Prevention | Jeong-Min Kim, Yoon-Seok Chung, Namjoo Lee, Mi-Seon Kim, Sang Hee Woo, Hye-Jun Jo, Sehee Park, Heui Man Kim, Myung Guk Han |
| EPI_ISL_412972 | Instituto Nacional de Enfermedades Respiratorias | Instituto de Diagnostico y Referencia Epidemiologicos (INDRE) | Ramirez-Gonzalez Ernesto, Garces-Ayala Fabiola, Araiza-Rodriguez Adnan, Mendieta-Condado Edgar, Rodriguez-Maldonado Abril, Wong-Arambula Claudia, Vazquez-Perez Joel, Martinez Arturo, Boukadida Celia, Munoz-Medina Esteban, Sanchez Alejandro, Isa Pavel, Taboada Blanca, Lopez Susana, Arias Carlos, Barrera-Badillo Gisela, Hernandez-Rivas Lucia, Lopez-Martinez Irma |
| EPI_ISL_413016 | Hospital Israelita Albert Einstein | Instituto Adolfo Lutz, Interdisciplinary Procedures Center, Strategic Laboratory | Jaqueline Goes de Jesus, Claudio Tavares Sacchi, Fabiana Cristina Pereira dos Santos, Ingra Morales Claro, Flávia Cristina da Silva Sales, Claudia Regina Gonçalves, Joshua Quick, Maria do Carmo Sampaio Tavares Timenetsky, Nicholas James Loman, Andrew Rambaut, Ester Cerdeira Sabino, Nuno Rodrigues Faria |
| EPI_ISL_413490 | Auckland Hospital | Institute of Environmental Science and Research (ESR) | Matt Storey, Xiaoyun Ren, Gary McAuliffe, Sally Roberts, Matthew Blakiston, Erasmus Smit, Lauren Jelly, Joep de Ligt |
| EPI_ISL_413523 | Indian Council of Medical Research-National Institute of Virology | National Influenza Center, Indian Council of Medical Research-National Institute of Virology | Potdar V, Yadav PD, Choudhary ML, Shete-Aich A |
| EPI_ISL_413570 | RIVM | Erasmus Medical Center | David Nieuwenhuijse, Bas Oude Munnink, Reina Sikkema, Claudia Schapendonk, Irina Chestakova, Anne van der Linden, Mark Pronk, Pascal Lexmond, Corien Swaan, Manon Haverkate, Madelief Mollers, Mart Stein, Sandra Kengne Kamga Mobou, Jeroen van Kampen, Jolanda Voermans, Aura Timen, Corine GeurtsvanKessel, Annemiek van der Eijk, Richard Molenkamp, Marion Koopmans, on behalf of the Dutch national COVID-19 response team. |
| EPI_ISL_413589 | MHC Utrecht | Erasmus Medical Center | David Nieuwenhuijse, Bas Oude Munnink, Reina Sikkema, Claudia Schapendonk, Irina Chestakova, Anne van der Linden, Mark Pronk, Pascal Lexmond, Corien Swaan, Manon Haverkate, Madelief Mollers, Mart Stein, Sandra Kengne Kamga Mobou, Jeroen van Kampen, Jolanda Voermans, Aura Timen, Corine GeurtsvanKessel, Annemiek van der Eijk, Richard Molenkamp, Marion Koopmans, on behalf of the Dutch national COVID-19 response team. |
| EPI_ISL_413596 | Centre for Infectious Diseases and Microbiology - Public Health | NSW Health Pathology - Institute of Clinical Pathology and Medical Research; Westmead Hospital; University of Sydney | Rockett R, Eden J-S, Lam C, Gray K, Timms, V, Gall, M, Carter I, Rahman H, Holmes EC, O'Sullivan MV, Sintchenko V, Chen SC, Maddocks S, Kok J and Dwyer DE for the 2019-nCoV Study Group* |
| EPI_ISL_414015 | Hospital São Joaquim Beneficencia Portuguesa | Instituto Adolfo Lutz, Interdiciplinary Procedures Center, Strategic Laboratory | Claudio Tavares Sacchi, Claudia Regina Gonçalves, SimoneGuadagnucci Morillo, Carlos Henrique Camargo, Maria do Carmo Sampaito Tavares Timenetsky, Fabiana Cristina Pereira dos Santos Terezinha Maria de Paiva, Ester Cerdeira Sabino |
| EPI_ISL_414414 | Pathology Queensland | Public Health Virology Laboratory | Bixing Huang, Alyssa Pyke, Amanda De Jong, Andrew Van Den Hurk, Carmel Taylor, David Warrilow, Doris Genge, Elisabeth Gamez, Glen Hewitson, Ian Maxwell Mackay, Inga Sultana, Jamie McMahon, Jean Barcelon, Judy Northill, Mitchell Finger, Natalie Simpson, Neelima Nair, Peter Burtonclay, Peter Moore, Sarah Wheatley, Sean Moody, Sonja Hall-Mendelin, Timothy Gardam, and Frederick Moore |
| EPI_ISL_414498, EPI_ISL_414499 | Center of Medical Microbiology, Virology, and Hospital Hygiene, University of Duesseldorf | Center of Medical Microbiology, Virology, and Hospital Hygiene, University of Duesseldorf | Ortwin Adams, Marcel Andree, Alexander Dilthey, Torsten Feldt, Sandra Hauka, Torsten Houwaart, Björn-Erik Jensen, Detlef Kindgen-Milles, Malte Kohns Vasconcelos, Klaus Pfeffer, Tina Senff, Daniel Strelow, Jörg Timm, Andreas Walker, Tobias Wienemann |
| EPI_ISL_415580 | BCCDC Public Health Laboratory | BCCDC Public Health Laboratory | Harrigan, Prystajecky, Krajden, Lee, Kamelian, Lapointe, Choi, Hoang, Sekirov, Levett, Tyson, Snutch, Loman, Quick, Li, Gilmour |
| EPI_ISL_415641 | R. G. Lugar Center for Public Health Research, National Center for Disease Control and Public Health (NCDC) of Georgia. | R. G. Lugar Center for Public Health Research, National Center for Disease Control and Public Health (NCDC) of Georgia. | Nato Kotaria, Marine Murtskhvaladze, Ann Machablishvili, Lela Sabadze, Mari Gavashelidze, Ana Papkiauri, Meri Pantsulaia, Gvantsa Brachveli, Tata Imnadze, Tamar Jashiashvili, Tea Tevdoradze, Ketevan Sidamonidze, Ekaterine Khmaladze, Ekaterine Zhghenti, Roena Sukhiashvili, Mariam Zakalashvili, Lela Urushadze, Magda Dgebuadze, Giorgi Tomashvili, Davit Tsaguria, Ekaterine Zangaladze, Nino Berishvili, Gvantsa Chanturia, Adam Kotorashvili, Maia Alkhazashvili, Irma Burjanadze, Anna Kasradze, Khatuna Zakhashvili, Paata Imnadze, Amiran Gamkrelidze. |
| EPI_ISL_416324, EPI_ISL_416370 | Shanghai Public Health Clinical Center, Shanghai Medical College, Fudan University | National Research Center for Translational Medicine (Shanghai), Ruijin Hospital affiliated to Shanghai Jiao Tong University School of Medicine & Shanghai Public Health Clinical Center | Shengyue Wang, Xiaonan Zhang, Gang Lu, Yun Tan, Yun Ling, Hongzhou Lu, Saijuan Chen |
| EPI_ISL_416429 | National Influenza Center, National Institute of Hygiene and | National Influenza Center, National Institute of Hygiene and | Le Quynh Mai, Taichiro Takemura, Meng Ling Moi, Takeshi Nabeshima, Nguyen Le Khanh Hang, Hoang Vu Mai Phuong, Ung Thi Hong Trang, Le Thi |

| | | | |
|---|---|---|---|
| | Epidemiology (NIHE) | Epidemiology (NIHE) | Thanh, Nguyen Vu Son, Vuong Duc Cuong, Pham Thi Hien, Tran Thu Huong, Nguyen Phuong Anh, Pham Hong Quynh Anh, Kouichi Morita, Futoshi Hasebe, Dang Duc Anh |
| EPI_ISL_416482 | R. G. Lugar Center for Public Health Research, National Center for Disease Control and Public Health (NCDC) of Georgia. | R. G. Lugar Center for Public Health Research, National Center for Disease Control and Public Health (NCDC) of Georgia. | Adam Kotorashvili, Marine Murtskhvaladze, Nato Kotaria, Ann Machablishvili, Lela Sabadze, Mari Gavashelidze, Ana Papkiauri, Meri Pantsulaia, Gvantsa Brachveli, Tata Imnadze, Tamar Jashiashvili, Tea Tevdoradze, Ketevan Sidamonidze, Ekaterine Khmaladze, Ekaterine Zhghenti, Roena Sukhiashvili, Mariam Zakalashvili, Lela Urushadze, Magda Dgebuadze, Giorgi Tomashvili, Davit Tsaguria, Ekaterine Zangaladze, Nino Berishvili, Gvantsa Chanturia, Maia Alkhazashvili, Irma Burjanadze, Anna Kasradze, Khatuna Zakhashvili, Paata Imnadze, Amiran Gamkrelidze. |
| EPI_ISL_416594 | Japanese Quarantine Stations | Pathogen Genomics Center, National Institute of Infectious Diseases | Tsuyoshi Sekizuka, Kentaro Itokawa, Rina Tanaka, Masanori Hashino, Tsutomu Kageyama, Shinji Saito, Ikuyo Takayama, Hideki Hasegawa, Takuri Takahashi, Hajime Kamiya, Takuya Yamagishi, Motoi Suzuki, Takaji Wakita, Makoto Kuroda |
| EPI_ISL_417033 | Sullivan Nicolaides Pathology | Public Health Virology Laboratory | Bixing Huang, Alyssa Pyke, Amanda De Jong, Andrew Van Den Hurk, Carmel Taylor, David Warrilow, Doris Genge, Elisabeth Gamez, Glen Hewitson, Ian Maxwell Mackay, Inga Sultana, Jamie McMahon, Jean Barcelon, Judy Northill, Mitchell Finger, Natalie Simpson, Neelima Nair, Peter Burtonclay, Peter Moore, Sarah Wheatley, Sean Moody, Sonja Hall-Mendelin, Timothy Gardam, and Frederick Moore |
| EPI_ISL_417211, EPI_ISL_417212 | Dunedin Hospital | University of Otago | M.E. Quiñones-Mateu, B. Lawley, J. Grant, R. Harfoot, J. Ussher |
| EPI_ISL_417444 | Department of Healthcare Biotechnology, National University of Sciences and Technology (NUST) | Department of Healthcare Biotechnology, National University of Sciences and Technology (NUST) | Javed,A., Niazi,S.K., Ghani,E., Saqib,M., Janjua,H.A., Corman,V.M. and Zohaib,A. |
| EPI_ISL_417577, EPI_ISL_417765 | The National University Hospital of Iceland | deCODE genetics | Daniel F Gudbjartsson; Agnar Helgason; Hakon Jonsson; Olafur T Magnusson; Pall Melsted; Gudmundur L Norddahl; Jona Saemundsdottir; Asgeir Sigurdsson; Patrick Sulem; Arna B Agustsdottir; Berglind Eiriksdottir; Run Fridriksdottir; Elisabet E Gardarsdottir; Gudmundur Georgsson; Olafia S Gretarsdottir; Kjartan R Gudmundsson; Thora R Gunnarsdottir; Arnaldur Gylfason; Hilma Holm; Brynjar O Jensson; Aslaug Jonasdottir; Kamilla S Josefsdottir; Thordur Kristjansson; Droplaug N Magnusdottir; Louise le Roux; Gudrun Sigmundsdottir; Gardar Sveinbjornsson; Kristin E Sveinsdottir; Maney Sveinsdottir; Emil A Thorarensen; Bjarni Thorbjornsson; Gisli Masson; Ingileif Jonsdottir; Alma Moller; Thorolfur Gudnason; Karl G Kristinsson; Unnur Thorsteinsdottir; Kari Stefansson |
| EPI_ISL_417878, EPI_ISL_417879, EPI_ISL_417880 | Institute of Virology, Biomedical Research Center of the Slovak Academy of Sciences, Bratislava; Public Health Authority of the Slovak Republic, Bratislava | Institute of Virology, Biomedical Research Center of the Slovak Academy of Sciences, Bratislava; Comenius University Science Park, Bratislava | Monika Slávikova, Martina Liková, Sabina Fumaová Havlíková, Juraj Koi, Juraj Kopáek, Elena Tichá, Edita Staroová, Jaroslav Budiš, Werner Krampl, Miroslav Böhmer, Diana Rusáková, Tomáš Szemeš, Boris Klempa |
| EPI_ISL_418212 | Institut Pasteur Dakar | Institut Pasteur de Dakar | Ndongo Dia, Ousmane Faye, Amadou Alpha sall |
| EPI_ISL_418269 | Microbiology and Immunology department, Pasteur institute in Ho Chi Minh city | Microbiology and Immunology department, Pasteur institute in Ho Chi Minh city | Cao,T.M., Nguyen,H.T., Pham,H.T.T., Vu,N.P.H., Dao,M.H., Huynh,L.T.K., Nguyen,L.T., Nguyen,N.T., Nguyen,T.T.N., Nguyen,A.H., Luong,Q.C., Nguyen,T.V., Tran,K.C., Pham,Q.D., Tran,T., Hoang,C.Q., Nguyen,T.T., Le,H.Q., Phung,T.M., Vo,T.N.A., Nguyen,S.N., Pham,D.T., Phan,L.T. and Nguyen,T.V. |
| EPI_ISL_418799 | Mater Pathology | Public Health Virology Laboratory | Bixing Huang, Alyssa Pyke, Amanda De Jong, Andrew Van Den Hurk, Carmel Taylor, David Warrilow, Doris Genge, Elisabeth Gamez, Glen Hewitson, Ian Maxwell Mackay, Inga Sultana, Jamie McMahon, Jean Barcelon, Judy Northill, Mitchell Finger, Natalie Simpson, Neelima Nair, Peter Burtonclay, Peter Moore, Sarah Wheatley, Sean Moody, Sonja Hall-Mendelin, Timothy Gardam, and Frederick Moore |
| EPI_ISL_418817 | BCCDC Public Health Laboratory | BCCDC Public Health Laboratory | Harrigan, Prystajecky, Krajden, Lee, Kamelian, Lapointe, Choi, Hoang, Sekirov, Levett, Tyson, Snutch, Loman, Quick, Li, Gilmour |
| EPI_ISL_419558 | OR State PHL-Virology/Immunology Section | Pathogen Discovery, Respiratory Viruses Branch, Division of Viral Diseases, Centers for Disease Control and Prevention | Ying Tao, Jing Zhang, Krista Queen, Anna Uehara, Clinton R. Paden, Yan Li, Haibin Wang, Jasmine Padilla, Justin Lee, Suxiang Tong |
| EPI_ISL_419596 | Laboratoire National de Santé, Microbiology, Virology | Laboratoire National de Santé, Microbiology, Epidemiology and Microbial Genomics | Anke Wienecke-Baldacchino, Ardashel Latsuzbaia, Jessica Tapp, Catherine Ragimbeau, Guillaume Fournier, Tamir Abdelrahman, Trung Nguyen Nguyen, Joel Mossong |
| EPI_ISL_419734, EPI_ISL_419868 | Victorian Infectious Diseases Reference Laboratory (VIDRL) | Victorian Infectious Diseases Reference Laboratory and Microbiological Diagnostic Unit Public Health Laboratory, Doherty Institute | Caly L., Seemann T., Sait, M., Schultz M., Druce J., Sherry, N. |
| EPI_ISL_420037 | NIC Viral Respiratory Unit - Institut Pasteur of Algeria | National Reference Center for Viruses of Respiratory Infections, Institut Pasteur, Paris | Mélanie Albert, Marion Barbet, Sylvie Behillil, Méline Bizard, Angela Brisebarre, Flora Donati, Etienne Simon-Lorière, Vincent Enouf, Maud Vanpeene, Sylvie van der Werf, Fawzi Derrar |
| EPI_ISL_420138 | University Hospital of Northern Norway, Department for Microbiology and Infectious Disease Control | Norwegian Institute of Public Health, Department of Virology | Kathrine Stene-Johansen, Kamilla Heddeland Instefjord, Hilde Elshaug, Karoline Bragstad, Olav Hungnes |
| EPI_ISL_420144 | Department for Virology, Molecular Biology and Genome Research, R. G. Lugar Center for Public Health Research, National Center for Disease Control and Public Health (NCDC) of Georgia. | Department for Virology, Molecular Biology and Genome Research, R. G. Lugar Center for Public Health Research, National Center for Disease Control and Public Health (NCDC) of Georgia. | Gvantsa Chanturia, Ann Machablishvili, Nato Kotaria, Marine Murtskhvaladze, Lela Sabadze, Mari Gavashelidze, Ana Papkiauri, Meri Pantsulaia, Gvantsa Brachveli, Tata Imnadze, Tamar Jashiashvili, Tea Tevdoradze, Ketevan Sidamonidze, Ekaterine Khmaladze, Ekaterine Zhghenti, Roena Sukhiashvili, Mariam Zakalashvili, Lela Urushadze, Magda Dgebuadze, Giorgi Tomashvili, Davit Tsaguria, Ekaterine Zangaladze, Nino Berishvili, Adam Kotorashvili, Maia Alkhazashvili, Irma Burjanadze, Anna Kasradze, Khatuna Zakhashvili, Paata Imnadze, Amiran Gamkrelidze. |
| EPI_ISL_420317 | KU Leuven, Clinical and Epidemiological Virology | KU Leuven, Clinical and Epidemiological Virology | Joan Marti-Carreras, Bert Vanmechelen, Tony Wawina, Piet Maes |
| EPI_ISL_420456 | PathWest Laboratory Medicine WA | PathWest Laboratory Medicine WA | Chisha Sikazwe, Jurissa Lang, Avram Levy, David Smith and David Speers |
| EPI_ISL_420791 | NH Department of Health and Human Services Public Health Labs | Pathogen Discovery, Respiratory Viruses Branch, Division of Viral Diseases, Centers for Disease Control and Prevention | Krista Queen, Yan Li, Ying Tao, Jing Zhang, Anne Uehara, Clinton R. Paden, Haibin Wang, Rachel Marine, Mary S. Keckler, Alison S. Laufer Halpin, Jasmine Padilla, Justin Lee, Christopher A. Elkins, Suxiang Tong |
| EPI_ISL_421223 | Hangzhou Center for Diseases Control and Prevention | Hangzhou Center for Diseases Control and Prevention | Jun Li, Haoqiu Wang, Lingfeng Mao, Hua Yu, Xinfen Yu, Zhou Sun, Xin Qian, Shuchang Chen, Junfang Chen, Xuchu Wang |
| EPI_ISL_422347 | Wales Specialist Virology Centre | Public Health Wales Microbiology Cardiff | Catherine Moore, Johnathan Evans, Malorie Perry, Simon Cottrell, Alec Birchley, Alexander Adams, Amy Gaskin, Bree Gatica-Wilcox, Jason Coombes, Lauren Gilbert, Lee Graham, Nicole Pacchiarini, Sara Kumziene-Summerhayes, Sarah Taylor, Sophie Jones, Sara Rey, Matthew Bull, Joanne Watkins, Sally Corden, Tom Connor |
| EPI_ISL_422405 | NMIMR, Department of Virology | WACCBIP, University of Ghana | Joyce M. Ngoi, Bright Adu, Collins M. Morang'a, Selassie Kumordjie, Miriam Eshun, Linda Boatemaa, Vanessa Magnussen, Erasmus Kotey, Fred Tei-Maya, Dominic S. Y. Amuzu, Peter Quashie, Augustina Arjarquah, Ivy Asante, Evelyn Bonney, George B. Kyei, Kofi Bonney, Abraham Kwabena Anang, Gordon A. Awandare, William Ampofo |
| EPI_ISL_422427 | JABER AL AHMAD AL SABAH HOSPITAL - KUWAIT CITY | Dasman Diabetes Institute | Fahd Al-Mulla, Rasheeba Iqbal, Sumi John, Ebaa Al-Ozairi, Qais Al-Duwairi |
| EPI_ISL_424916 | MA State Public Health Laboratory | Pathogen Discovery, Respiratory Viruses Branch, Division of Viral Diseases, Centers for Disease Control and Prevention | Ying Tao, Clinton R. Paden, Jing Zhang, Krista Queen, Anna Uehara, Yan Li, Haibin Wang, Mary S. Keckler, Alison S. Laufer Halpin, Christopher A. Elkins, Suxiang Tong |
| EPI_ISL_426629 | TSGH-CP molecular lab | TSGH-CP molecular lab | Cherng-Lih Perng, Ming-Jr Jian, Chih-Kai Chang, Jung-Chung Lin, Kuo-Ming Yeh, Chien-Wen Chen, Sheng-Kang Chiu, Hsing-Yi Chung, Shih-Hung Tsai, Kuo-Sheng Hung, Feng-Yee Chang, Hung-Sheng Shang |
| EPI_ISL_427047, EPI_ISL_427128 | Victorian Infectious Diseases Reference Laboratory (VIDRL) | Microbiological Diagnostic Unit Public Health Laboratory and Victorian Infectious Diseases Reference Laboratory, Doherty Institute | Caly L., Seemann T., Sait, M., Schultz M., Druce J., Sherry, N. |
| EPI_ISL_427203 | UW Virology Lab | UW Virology Lab | Pavitra Roychoudhury, Hong Xie, Keith Jerome, Alexander Greninger |
| EPI_ISL_428671 | Centre for Dengue Research | Centre for Dengue Research | Chandima Jeewandara, Dinuka Ariyatane, Laksiri Gomes, Deshni Jayathilaka, Diyanath Ranasinghe, Ananda Wijewickrama, Eranga Narangoda, Damayanthi Tdampitiya, Neelika Malavige |
| EPI_ISL_428673 | Centre for Dengue Research | Centre for Dengue Research | Chandima Jeewandara, Dinuka Ariyaratne, Laksiri Gomes, Deshni Jayathilaka, Diyanath Ranasinghe, Ananda Wijewickrama, Eranga Narangoda, Damayanthi Idampitiya, Neelika Malavige |
| EPI_ISL_428855 | MRCG at LSHTM Geomics lab | MRCG at LSHTM Genomics lab | Sesay et al |
| EPI_ISL_428901 | State Research Center of Virology and Biotechnology | State Research Center of Virology and Biotechnology | Sergey A. Bodnev, Oleg V. Pyankov, Tatyana V. Tregubchak, Alexander N. Shvalov, Elena V. Gavrilova, Rinat A. Maksyutov |

| | | | |
|---|---|---|---|
| | VECTOR, Department of Collection of Microorganisms | VECTOR, Department of Collection of Microorganisms | |
| EPI_ISL_429629 | UW Virology Lab | UW Virology Lab | Pavitra Roychoudhury, Hong Xie, Keith Jerome, Alexander Greninger |
| EPI_ISL_429998, EPI_ISL_430000 | Biolab Diagnostic Laboratories | Andersen lab at Scripps Research | Issa Abu-Dayyeh, Ahmad Tibi, Lama Hussein, Lina Mohammad, Zein Naber, Amid Abdelnour with SEARCH Alliance San Diego |
| EPI_ISL_430292 | Washington State Department of Health | Seattle Flu Study | Chu et al |
| EPI_ISL_434564 | unknown | Microbiology, The University of Hong Kong | To,K.K.W. and Yuen,K.-Y. |
| EPI_ISL_435114 | Viral Respiratory Lab, National Institute for Biomedical Research (INRB) | Pathogen Sequencing Lab, National Institute for Biomedical Research (INRB) | Placide Mbala-Kingebeni, Edith Nkwembe, Eddy Kinganda-Lusamaki, Adrienne Amuri Aziza, Francisca Muyembe Mawete, Catherine Pratt, Matthias Pauthner, Josh Quick, Allison Black, James Hadfield, Trevor Bedford, Ian Goodfellow, Andrew Rambaut, Nick Loman, Kristian Andersen, Michael Wiley, Steve Ahuka-Mundeke, Jean-Jacques Muyembe Tamfum |
| EPI_ISL_436120 | Victorian Infectious Diseases Reference Laboratory (VIDRL) | Microbiological Diagnostic Unit Public Health Laboratory and Victorian Infectious Diseases Reference Laboratory, Doherty Institute | Caly L., Seemann T., Sait, M., Schultz M., Druce J., Sherry, N. |
| EPI_ISL_437440 | Department of MicroBiology, Government Medical College, Surat | Gujarat Biotechnology Research Centre | Nitin Savaliya, Raghawendra Kumar, Dinesh Kumar, Zuber Saiyed, Dipa Kinariwala, Disha Patel, Binita Aring, Neeta Khandelwal, Geeta Vaghela, Sonia Barve, Bhavesh Modi, Kairavi Joshi, Gaurishankar Shrimali, Nidhi Sood, Pranay Shah, R D Dixit, Snehal Bagatharia, Kamlesh J Upadhyay, Ramesh Pandit, Tejas Shah, Ankit Hinsu, Pritesh Sabara, Apurvasinh Puvar, Janvi Raval, Monika Gandhi, Pinal Trivedi, Maharshi Pandya, Amit Kanani, Akanksha Verma, Bhavya Jindal, Chaitanya Joshi, Madhvi Joshi |
| EPI_ISL_444610 | U.S. Naval Medical Research Center Biological Defense Research Directorate | U.S. Naval Medical Research Center Biological Defense Research Directorate | Voegtly,L.J., Cer,R.Z., Pena-Gomez,D., Paskey,A.C., Long,K.A., Hollis,E.M., Pan,R.W., Balansy-Ames,M.S., Myers,C.A., Christy,N.C. and Bishop-Lilly,K.A. |
| EPI_ISL_444998, EPI_ISL_444999 | Naval Health Research Center | Naval Medical Research Center Biological Defense Research Directorate | Logan Voegtly, Regina Cer, Dessiree Pena-Gomez, Adrian Paskey,Kyle Long, Roger Pan, Melinda Balansay-Ames, Chris Myers, Ewell Hollis, Nathaniel Christy, Kimberly Bishop-Lilly |
| EPI_ISL_445072 | Laboratoire National de Sante, Microbiology, Virology | Laboratoire National de Sante, Microbiology, Epidemiology and Microbial Genomics | Anke Wienecke-Baldacchino, Ardashel Latsuzbaia, Jessica Tapp, Catherine Ragimbeau, Guillaume Fournier, Tamir Abdelrahman, Trung Nguyen Nguyen, Joel Mossong |
| EPI_ISL_446556 | Wales Specialist Virology Centre | Public Health Wales Microbiology Cardiff | Catherine Moore, Johnathan Evans, Laura Gifford, Malorie Perry, Simon Cottrell, Alec Birchley, Alexander Adams, Amy Gaskin, Bree Gatica-Wilcox, Jason Coombes, Lauren Gilbert, Lee Graham, Nicole Pacchiarini, Sara Kumziene-Summerhayes, Sarah Taylor, Sophie Jones, Sara Rey, Matthew Bull, Joanne Watkins, Sally Corden, Tom Connor |
| EPI_ISL_447232 | Viral Respiratory Lab, National Institute for Biomedical Research (INRB) | Pathogen Sequencing Lab, National Institute for Biomedical Research (INRB) | Placide Mbala-Kingebeni, Edith Nkwembe, Eddy Kinganda-Lusamaki, Amuri Aziza, Francisca Muyembe Mawete, Catherine Pratt, Matthias Pauthner, Josh Quick, Allison Black, James Hadfield, Trevor Bedford, Ian Goodfellow, Andrew Rambaut, Nick Loman, Kristian Andersen, Michael Wiley, Steve Ahuka-Mundeke, Jean-Jacques Muyembe Tamfum |
| EPI_ISL_447268 | Microbiology laboratory, Assuta Ashdod University-Affiliated Hospital | Stern Lab | Stern Lab |
| EPI_ISL_447331, EPI_ISL_447413 | Clinical Virology Unit, Hadassah Hebrew University Medical Center | Stern Lab | Stern Lab |
| EPI_ISL_447593 | TSGH-CP molecular lab | TSGH-CP molecular lab | Cherng-Lih Perng, Ming-Jr JIAN, Chih-Kai Chang, Jung-Chung Lin, Kuo-Ming Yeh, Chien-Wen Chen, Sheng-Kang Chiu, Hsing-Yi Chung, Shih-Hung Tsai, Kuo-Sheng Hung, Tien-Yao Chang, Feng-Yee Chang, Hung-Sheng Shang |
| EPI_ISL_447750 | Grupo de Investigaciones Microbiológicas-UR (GIMUR), Departamento de Biología, Facultad de Ciencias Naturales, Universidad del Rosario, Bogotá, Colombia | Grupo de Investigaciones Microbiológicas-UR (GIMUR), Departamento de Biología, Facultad de Ciencias Naturales, Universidad del Rosario, Bogotá, Colombia Instituto Nacional de Salud, Bogotá, Colombia Icahn School of Medicine at Mount Sinai, New York, USA | Juan David Ramírez, Carolina Florez, Marina Muñoz, Carolina Hernandez, Adriana Castillo, Sergio Castañeda, Nathalia Ballesteros, David Martínez, Laura Vega, Jesús E. Jaimes, Sergio Gomez, Angelica Rico, Lisseth Pardo, Esther C. Barros, Martha L. Ospina, Aníbal A. Teherán, Ana S. Gonzalez-Reiche, Matthew M. Hernandez, Emilia Mia Sordillo, Viviana Simon, Harm van Bakel, Alberto Paniz-Mondolfi |
| EPI_ISL_447761, EPI_ISL_447797, EPI_ISL_447800, EPI_ISL_447807 | Instituto Nacional de Salud, Bogotá, Colombia | Grupo de Investigaciones Microbiológicas-UR (GIMUR), Departamento de Biología, Facultad de Ciencias Naturales, Universidad del Rosario, Bogotá, Colombia Instituto Nacional de Salud, Bogotá, Colombia Icahn School of Medicine at Mount Sinai, New York, USA | Juan David Ramírez, Carolina Florez, Marina Muñoz, Carolina Hernandez, Adriana Castillo, Sergio Castañeda, Nathalia Ballesteros, David Martínez, Laura Vega, Jesús E. Jaimes, Sergio Gomez, Angelica Rico, Lisseth Pardo, Esther C. Barros, Martha L. Ospina, Aníbal A. Teherán, Ana S. Gonzalez-Reiche, Matthew M. Hernandez, Emilia Mia Sordillo, Viviana Simon, Harm van Bakel, Alberto Paniz-Mondolfi |
| EPI_ISL_450209 | Department of Virology | Department of Virology | Boehmer,M.M., Buchholz,U., Corman,V.M., Hoch,M., Katz,K., Marosevic,D.V., Boehm,S., Woudenberg,T., Ackermann,N., Konrad,R., Eberle,U., Treis,B., Dangel,A., Bengs,K., Fingerle,V., Berger,A., Hoermansdorfer,S., Ippisch,S., Wicklein,B., Grahl,A., Poertner,K., Muller,N., Zeitlmann,N., Boender,T.S., Cai,W., Reich,A., an der Heiden,M., Rexroth,U., Hamouda,O., Schneider,J., Veith,T., Muehlemann,B., Woelfel,R., Antwerpen,M., Walter,M., Protzer,U., Liebl,B., Haas,W., Sing,A., Drosten,C., Zapf,A., Jones,T.C. |
| EPI_ISL_450350 | St.Olavs hospital/NTNU | Institute of Genomics Core Facility, University of Tartu | Aleksandr Ianevski, Tuuli Reisberg, Janne-Fossum Malmring, Svein Arne Nordbø, Denis Kainov |
| EPI_ISL_450774 | Minnesota Department of Health, Public Health Laboratory | Minnesota Department of Health, Public Health Laboratory | Matt Plumb, Jacob Garfin, and Xiong Wang |
| EPI_ISL_450800 | AR Dept. of Health-Public Health Lab | Pathogen Discovery, Respiratory Viruses Branch, Division of Viral Diseases, Centers for Disease Control and Prevention | Yan Li, Anna Montmayeur, Ying Tao, Krista Queen, Jing Zhang, Anna Uehara, Clinton R. Paden, Rachel Marine, Haibin Wang, Zachary Weiner, Bettina Bankamp, Suxiang Tong |
| EPI_ISL_451532 | Pathology West - NSW Health Pathology | NSW Health Pathology - Institute of Clinical Pathology and Medical Research; Westmead Hospital; University of Sydney | CIDM-PH et al. |
| EPI_ISL_452122 | VI-US Virgin Islands Department of Health | Pathogen Discovery, Respiratory Viruses Branch, Division of Viral Diseases, Centers for Disease Control and Prevention | Jing Zhang, Anna Montmayeur, Yan Li, Ying Tao, Krista Queen, Anna Uehara, Clinton R. Paden, Rachel Marine, Mary S. Keckler, Alison S. Laufer Halpin, Haibin Wang, Christopher A. Elkins, Zachary Weiner, Suxiang Tong |
| EPI_ISL_454497 | RSE "National Center for Biotechnology" | RSE "National Center for Biotechnology" | Alexandr Shevtsov, Ilyas Akhmetollayev, Viktoriya Lutsay, Asylulan Amirgazin, Askar Abdaliyev, Akbota Rakhmetova, Zabira Aushakhmetova, Ruslan Kalendar, Yerlan Ramankulov |
| EPI_ISL_454923, EPI_ISL_454979 | Wuhan Chain Medical Labs (CMLabs) | State Key Laboratory of Biotherapy of Sichuan University | Baowen Du, Minjin Wang, Chao Tang, Chuan Chen, Yongzhao Zhou, Mingxia Yu, Hancheng Wei, Weimin Li, Jing-wen Lin, Jia Geng, Binwu Ying, Lu Chen |
| EPI_ISL_455099 | South Eastern Area Laboratory Services | NSW Health Pathology - Institute of Clinical Pathology and Medical Research; Westmead Hospital; University of Sydney | CIDM-PH et al. |
| EPI_ISL_455415 | Nigeria Centre for Disease Control (NCDC) | African Centre of Excellence for Genomics of Infectious Diseases (ACEGID), Redeemer's University, Ede, Osun State, Nigeria | Oluniyi P.E., Ajogbasile F.V., Kayode A., Oguzie J., Olawoye I., Uwanibe J., Olumade T., Folarin O.A., Ihekweazu C., Happi C.T. |
| EPI_ISL_455633 | KRISP, KZN Research Innovation and Sequencing Platform | KRISP, KZN Research Innovation and Sequencing Platform | Giandhari J, Pillay S, Lessells R, Chimukangara B, Deforche K, Tegally H, Wilkinson E, de Oliveira T |
| EPI_ISL_455791 | Institute for Medical Research, Infectious Disease Research Centre, National Institutes of Health, Ministry of Health Malaysia | Malaysia Genome Institute | Mohd Noor Mat Isa, Irni Suhayu Sapian, Yusuf Muhammad Noor, Jeyanthi Suppiah, Nurhezreen Md Iqbal, Enizza Kasim, Zarina Mohd Zawawi, Siti Noraini Othman, Mohd Faizal Abu Bakar, Shamsidar Sopie, Azrin Ahmad, Ravindran Thayan, Norazah Ahmad, Tahir Aris, Shahrul Hisham Zainal Ariffin |
| EPI_ISL_456163 | PathLab Bay of Plenty | Institute of Environmental Science and Research (ESR) | Matt Storey, Xiaoyun Ren, Anja Werno, Antje van der Linden, Arlo Upton, Chris Mansell, David Hammer, Dragana Drinkovic, Erasmus Smit, Gary McAuliffe, Hana Sofia Andersson, James Ussher, Jill Sherwood, Josh Freeman, Julia Howard, Juliet Elvy, Matt Blakiston, Matthew Rogers, Max Bloomfield, Michael Addidle, Michelle Balm, Sally Roberts, Sarah Jefferies, Sharmini Muttaiyah, Susan Morpeth, Susan Taylor, Timothy Blackmore, Vani Sathyendran, Veronica Playle, Virginia Hope, Erasmus Smit, Lauren Jelly, Joep de Ligt |
| EPI_ISL_456169, EPI_ISL_456184, | Wellington SCL | Institute of Environmental Science and Research (ESR) | Matt Storey, Xiaoyun Ren, Anja Werno, Antje van der Linden, Arlo Upton, Chris Mansell, David Hammer, Dragana Drinkovic, Erasmus Smit, Gary |

| | | | |
|---|---|---|---|
| EPI_ISL_456186 | | | McAuliffe, Hana Sofia Andersson, James Ussher, Jill Sherwood, Josh Freeman, Julia Howard, Juliet Elvy, Mary DeAlmeida, Matt Blakiston, Matthew Rogers, Max Bloomfield, Michael Addidle, Michelle Balm, Sally Roberts, Sarah Jefferies, Sharmini Muttaiyah, Susan Morpeth, Susan Taylor, Timothy Blackmore, Vani Sathyendran, Veronica Playle, Virginia Hope, Erasmus Smit, Lauren Jelly, Joep de Ligt |
| EPI_ISL_456193 | Waikato Hospital | Institute of Environmental Science and Research (ESR) | Matt Storey, Xiaoyun Ren, Anja Werno, Antje van der Linden, Arlo Upton, Chris Mansell, David Hammer, Dragana Drinkovic, Erasmus Smit, Gary McAuliffe, Hana Sofia Andersson, James Ussher, Jill Sherwood, Josh Freeman, Julia Howard, Juliet Elvy, Mary DeAlmeida, Matt Blakiston, Matthew Rogers, Max Bloomfield, Michael Addidle, Michelle Balm, Sally Roberts, Sarah Jefferies, Sharmini Muttaiyah, Susan Morpeth, Susan Taylor, Timothy Blackmore, Vani Sathyendran, Veronica Playle, Virginia Hope, Erasmus Smit, Lauren Jelly, Joep de Ligt |
| EPI_ISL_456195, EPI_ISL_456223 | PathLab Bay of Plenty | Institute of Environmental Science and Research (ESR) | Matt Storey, Xiaoyun Ren, Anja Werno, Antje van der Linden, Arlo Upton, Chris Mansell, David Hammer, Dragana Drinkovic, Erasmus Smit, Gary McAuliffe, Hana Sofia Andersson, James Ussher, Jill Sherwood, Josh Freeman, Julia Howard, Juliet Elvy, Mary DeAlmeida, Matt Blakiston, Matthew Rogers, Max Bloomfield, Michael Addidle, Michelle Balm, Sally Roberts, Sarah Jefferies, Sharmini Muttaiyah, Susan Morpeth, Susan Taylor, Timothy Blackmore, Vani Sathyendran, Veronica Playle, Virginia Hope, Erasmus Smit, Lauren Jelly, Joep de Ligt |
| EPI_ISL_456238, EPI_ISL_456251 | Southern Community Labs Dunedin | Institute of Environmental Science and Research (ESR) | Matt Storey, Xiaoyun Ren, Anja Werno, Antje van der Linden, Arlo Upton, Chris Mansell, David Hammer, Dragana Drinkovic, Erasmus Smit, Gary McAuliffe, Hana Sofia Andersson, James Ussher, Jill Sherwood, Josh Freeman, Julia Howard, Juliet Elvy, Mary DeAlmeida, Matt Blakiston, Matthew Rogers, Max Bloomfield, Michael Addidle, Michelle Balm, Sally Roberts, Sarah Jefferies, Sharmini Muttaiyah, Susan Morpeth, Susan Taylor, Timothy Blackmore, Vani Sathyendran, Veronica Playle, Virginia Hope, Erasmus Smit, Lauren Jelly, Joep de Ligt |
| EPI_ISL_456319, EPI_ISL_456325, EPI_ISL_456326, EPI_ISL_456328, EPI_ISL_456340 | Wellington SCL | Institute of Environmental Science and Research (ESR) | Matt Storey, Xiaoyun Ren, Anja Werno, Antje van der Linden, Arlo Upton, Chris Mansell, David Hammer, Dragana Drinkovic, Erasmus Smit, Gary McAuliffe, Hana Sofia Andersson, James Ussher, Jill Sherwood, Josh Freeman, Julia Howard, Juliet Elvy, Mary DeAlmeida, Matt Blakiston, Matthew Rogers, Max Bloomfield, Michael Addidle, Michelle Balm, Sally Roberts, Sarah Jefferies, Sharmini Muttaiyah, Susan Morpeth, Susan Taylor, Timothy Blackmore, Vani Sathyendran, Veronica Playle, Virginia Hope, Erasmus Smit, Lauren Jelly, Joep de Ligt |
| EPI_ISL_456365 | Canterbury Health Laboratories | Institute of Environmental Science and Research (ESR) | Matt Storey, Xiaoyun Ren, Anja Werno, Antje van der Linden, Arlo Upton, Chris Mansell, David Hammer, Dragana Drinkovic, Erasmus Smit, Gary McAuliffe, Hana Sofia Andersson, James Ussher, Jill Sherwood, Josh Freeman, Julia Howard, Juliet Elvy, Mary DeAlmeida, Matt Blakiston, Matthew Rogers, Max Bloomfield, Michael Addidle, Michelle Balm, Sally Roberts, Sarah Jefferies, Sharmini Muttaiyah, Susan Morpeth, Susan Taylor, Timothy Blackmore, Vani Sathyendran, Veronica Playle, Virginia Hope, Erasmus Smit, Lauren Jelly, Joep de Ligt |
| EPI_ISL_456377 | MedLab Central Ltd | Institute of Environmental Science and Research (ESR) | Matt Storey, Xiaoyun Ren, Anja Werno, Antje van der Linden, Arlo Upton, Chris Mansell, David Hammer, Dragana Drinkovic, Erasmus Smit, Gary McAuliffe, Hana Sofia Andersson, James Ussher, Jill Sherwood, Josh Freeman, Julia Howard, Juliet Elvy, Mary DeAlmeida, Matt Blakiston, Matthew Rogers, Max Bloomfield, Michael Addidle, Michelle Balm, Sally Roberts, Sarah Jefferies, Sharmini Muttaiyah, Susan Morpeth, Susan Taylor, Timothy Blackmore, Vani Sathyendran, Veronica Playle, Virginia Hope, Erasmus Smit, Lauren Jelly, Joep de Ligt |
| EPI_ISL_456402 | Wellington SCL | Institute of Environmental Science and Research (ESR) | Matt Storey, Xiaoyun Ren, Anja Werno, Antje van der Linden, Arlo Upton, Chris Mansell, David Hammer, Dragana Drinkovic, Erasmus Smit, Gary McAuliffe, Hana Sofia Andersson, James Ussher, Jill Sherwood, Josh Freeman, Julia Howard, Juliet Elvy, Mary DeAlmeida, Matt Blakiston, Matthew Rogers, Max Bloomfield, Michael Addidle, Michelle Balm, Sally Roberts, Sarah Jefferies, Sharmini Muttaiyah, Susan Morpeth, Susan Taylor, Timothy Blackmore, Vani Sathyendran, Veronica Playle, Virginia Hope, Erasmus Smit, Lauren Jelly, Joep de Ligt |
| EPI_ISL_456562 | Victorian Infectious Diseases Reference Laboratory (VIDRL) | Microbiological Diagnostic Unit Public Health Laboratory and Victorian Infectious Diseases Reference Laboratory, Doherty Institute | Caly L., Seemann T., Sait, M., Schultz M., Druce J., Sherry, N. |
| EPI_ISL_456597, EPI_ISL_456600 | National Health Laboratory, Timor-Leste | Microbiological Diagnostic Unit Public Health Laboratory, The Peter Doherty Institute for Infection and Immunity | Soares da Silva, E., Dolores de Jesus da Costa, M., Salles de Sousa, A., Jayanti Pereira Tilman, A., Antonia da Costa, E., Barreto, I., Marr, I., Wapling, J., Francis, J., Ximenes, J., Canisia, D., Freeman, K., Dakh, F., Douglas, N., Baird, R., Caly, L., Seemann, T., Sait, M., Schultz, M., Sherry, N. |
| EPI_ISL_456642 | Victorian Infectious Diseases Reference Laboratory (VIDRL) | Microbiological Diagnostic Unit Public Health Laboratory and Victorian Infectious Diseases Reference Laboratory, Doherty Institute | Caly L., Seemann T., Sait, M., Schultz M., Druce J., Sherry, N. |
| EPI_ISL_456821 | West of Scotland Specialist Virology Centre, NHSGGC / MRC-University of Glasgow Centre for Virus Research | COVID-19 Genomics UK (COG-UK) Consortium | Ana da Silva Filipe, Natasha Johnson, Kathy Smollett, Daniel Mair, Stephen Carmichael, Lily Tong, Jenna Nichols, Elihu Aranday-Cortes, Kirstyn Brunker, Yasmin Parr, Kyriaki Nomikou; Sarah McDonald, Marc Niebel, Patawee Asamaphan; Richard Orton, Joseph Hughes, Sreenu Vattipally, David L Robertson; Alasdair MacLean, Rory Gunson; Kathy Li, Natasha Jesudason, Rajiv Shah, James Shepherd, Antonia Ho, Emma Thomson |
| EPI_ISL_457845, EPI_ISL_457852, EPI_ISL_457910 | KEMRI-CGMR-C | KEMRI-Wellcome Trust Research Programme/KEMRI-CGMR-C Kilifi | Githinji G. et al 2020 |
| EPI_ISL_457942, EPI_ISL_457973 | Laboratorio de Biología Molecular Asociación Española Primera en Salud | Departments of Pathology and Medicine, New York University School of Medicine | Maria Victoria Elizondo, Maria Noel Zubillaga, Gonzalo Manrique, Paul Zappile, Gael Westby, Matthew T Maurano, Christian Marier, Adriana Heguy |
| EPI_ISL_459980, EPI_ISL_459983 | Institut Pasteur du Maroc | Institut Pasteur du Maroc | Marion Barbet, Sylvie Behillil, Méline Bizard, Angela Brisebarre, Camille Capel, Etienne Simon-Lorière, Vincent Enouf, Maud Vanpeene, Sylvie van der Werf, Latifa Anga, Abdellah Faouzi, Anass Abbad, Mjid Eloualid, Jalal Nourlil, Anderrahmane Maaroufi |
| EPI_ISL_460111 | Massachusetts General Hospital | Infectious Disease Program, Broad Institute of Harvard and MIT | Lemieux,J.E., Siddle,K.J., Shaw,B., Adams,G., Pierce,V., Turbett,S., Anahtar,M., Branda,J., Slater,D., Harris,J., Lin,A.E., Gladden-Young,A., Lagerborg,K., Rudy,M., DeRuff,K., Carter,A., Normandin,E., Bauer,M., Reilly,S., Tomkins-Tinch,C., Loreth,C., Chaluvadi,S., Neumann,A., Cusick,C., Chapman,S.B., Gnirke,A., Flowers,K., Cerrato,F., Birren,B.W., Gallagher,G., Smole,S., Park,D.J., MacInnis,B.L., Ryan,E., LaRocque,R., Rosenberg,E., Sabeti,P.C. |
| EPI_ISL_461950 | Queens Medical Centre, Clinical Microbiology Department / DeepSeq Nottingham | COVID-19 Genomics UK (COG-UK) Consortium | Gemma Clark, Wendy Smith, Manjinder Khakh, Hannah Howson-Wells, Jonathan Ball, Patrick McClure, Joseph Chappell, Theocharis Tsoleridis, Nadine Holmes, Matthew Carlisle, Christopher Moore, Fei Sang, Johnny Debebe, Victoria Wright, Matthew Loose |
| EPI_ISL_462462 | Clinical Center, University of Sarajevo | Charite Universitatsmedizin Berlin, Institute of Virology | Victor M Corman, Jorn Beheim-Schwarzbach, Barbara Muehlemann, Talitha Veith, Julia Schneider, Terry Jones, Amela Dedeic-Ljubovic, Irma Salimovic-Besic, Suzana Arapcic, Almedina Hadzihasanovic-Moro, Selma Mutevelic, Christian Drosten |
| EPI_ISL_462753 | University Clinical Hospital of Mostar | University of Sarajevo Veterinary Faculty | Goletic, T., Softic, A., Goletic, S., Ostojic, M., Hukic, M., Eterovic, T., Seho-Alic, A. |
| EPI_ISL_463002, EPI_ISL_463003 | unknown | Clinical virology | Fares,W., Triki,H. |
| EPI_ISL_463747 | Department of Molecular Virology, Cyprus Institute of Neurology and Genetics | Department of Molecular Virology, Cyprus Institute of Neurology and Genetics | Jan Richter, George Krashias, Christina Tryfonos, Stavros Bashiardes, Dana Koptides, Christina Christodoulou |
| EPI_ISL_464070 | KU Leuven, Rega Institute, Clinical and Epidemiological Virology | KU Leuven, Rega Institute, Clinical and Epidemiological Virology | Tony Wawina-Bokalanga, Bert Vanmechelen, Joan Marti-Carerras, Piet Maes |
| EPI_ISL_466636 | National Institute of Laboratory Medicine and Referral Center | Genomic Research Lab, BCSIR | Abu Sayeed Mohammad Mahmud, Mohammad Samir Uzzaman, Eshrar Osman, Md. Ahasan Habib, Shahina Akter, Tanjina Akhter Banu, Md. Murshed Hasan Sarker, Iffat Jahan, Barna Goswami, Md. Saddam Hossain, Tasnim Nafisa, Md. Maruf Ahmed Molla, Mahmuda Yeasmin, Asish Kumar Ghosh, Arifa Akram, A. K. M. Shamsuzzaman, Sheikh Md. Selim Al Din, Utpal Chandra Ray, Salek Ahmed Sajib, Md. Salim Khan |
| EPI_ISL_467374 | Dinkes Samarinda | Eijkman Institute for Molecular Biology, Ministry of Research and Technology/National Agency for Research and Innovation | Edison Johar, Frilasita A Yudhaputri, Hidayat Trimarsanto, David H Muljono, Safarina G Malik, Khin Saw Myint, Amin Soebandrio |
| EPI_ISL_467666 | Virology lab, NIC, NCCD, Ulaanbaatar, Mongolia | National Centre for Communicable Deseases (NCCD) | Naranzul Ts,Darmaa B,Bayasgalan N,Ankhbayar S,Tsogtbaatar B, Erdene-Ochir Ts,Nymadawa P |
| EPI_ISL_467987, EPI_ISL_467989, EPI_ISL_468024, EPI_ISL_468039 | SA Pathology | SA Pathology | Lex Leong, Chuan Kok Lim, Mark Turra, Ivan Bastian, Geoff Higgins |
| EPI_ISL_468119 | OHSU Lab Services Molecular Microbiology Lab | Oregon SARS-CoV-2 Genome Sequencing Center | Brendan L. O'Connell, Ruth V. Nichols, Alec J. Hirsch, Guang Fan, Daniel N. Streblow, William B. Messer, Andrew C. Adey, Benjamin N. Bimber, Brian J. O'Roak |
| EPI_ISL_469048 | Banas Medical College and Research Institute | Gujarat Biotechnology Research Centre | Radhika Khara, Sunil R Joshi, Viren s Doshi, Zarna Patel, Monika Gandhi, Pinal Trivedi, Maharshi Pandya, Nidhi Patel, Nitin Savaliya, Raghawendra Kumar, Dinesh Kumar, Zuber Saiyed, Komal Patel, Labdhi Pandya, Snehal Bagatharia, Tejas Shah, Ankit Hinsu, Pritesh Sabara, Apurvasinh Puvar, Janvi Raval, Priti Pandita, R D Dixit, A M Kadri, Harsh Bakshi, Chaitanya Joshi, Madhvi Joshi |

| | | | |
|---|---|---|---|
| EPI_ISL_469095 | National Public Health Laboratory, National Centre for Infectious Diseases | National Public Health Laboratory, National Centre for Infectious Diseases | Mak TM, Octavia S, Chavatte JM, Cui L, Lin RTP |
| EPI_ISL_470848, EPI_ISL_470851, EPI_ISL_470855 | PathWest Laboratory Medicine WA | PathWest Laboratory Medicine WA | Chisha Sikazwe, Jurissa Lang, Avram Levy, David Smith and David Speers |
| EPI_ISL_470876 | Department for Virology, Molecular Biology and Genome Research, R. G. Lugar Center for Public Health Research, National Center for Disease Control and Public Health (NCDC) of Georgia. | Department for Virology, Molecular Biology and Genome Research, R. G. Lugar Center for Public Health Research, National Center for Disease Control and Public Health (NCDC) of Georgia. | Giorgi Tomashvili, Meri Pantsulaia, Gvantsa Brachveli, Gvantsa Chanturia, Ann Machablishvili, Nato Kotaria, Marine Murtskhvaladze, Lela Sabadze, Mari Gavashelidze, Ana Papkiauri, Tata Imnadze, Tamar Jashiashvili, Tea Tevdoradze, Ketevan Sidamonidze, Ekaterine Khmaladze, Ekaterine Zhghenti, Roena Sukhiashvili, Mariam Zakalashvili, Lela Urushadze, Magda Dgebuadze, Davit Tsaguria, Ekaterine Zangaladze, Nino Berishvili, Adam Kotorashvili, Maia Alkhazashvili, Irma Burjanadze, Anna Kasradze, Khatuna Zakhashvili, Paata Imnadze, Amiran Gamkrelidze. |
| EPI_ISL_471163, EPI_ISL_471165 | MRCG at LSHTM Genomics lab | MRCG at LSHTM Genomics lab | Sesay et al |
| EPI_ISL_471446 | Division of Viral Diseases, Center for Laboratory Control of Infectious Diseases, Korea Centers for Diseases Control and Prevention | Division of Viral Diseases, Center for Laboratory Control of Infectious Diseases, Korea Centers for Diseases Control and Prevention | Jeong-Min Kim, Yoon-Seok Chung, Namjoo Lee, Sang Hee Woo, Hye-Jun Jo, Heui Man Kim, Jun-Sub Kim, Dong Hyun Song, Daesang Lee, Seong Tae Jeong, Myung Guk Han |
| EPI_ISL_475009 | Israel Central Virology laboratory | Israel Central Virology laboratory | Neta Zuckerman, Efrat Dahan Bucris, Oran Erster, Ella Mendelson, Michal Mandelboim |
| EPI_ISL_475545 | Karolinska Universitetslaboratoriet | The Public Health Agency of Sweden | Oskar Karlsson Lindsjo, Maria Lind Karlberg, Mattias Haukland, Reza Advani, Olov Svartstrom, Anna-Malin Linde, Sandra Broddesson, Shaman Muradrasoli, Anna Risberg, Karin Tegmark-Wisell |
| EPI_ISL_476155 | Laboratório de Patologia Clínica - UNICAMP | Laboratório de Estudos de Vírus Emergentes - UNICAMP | José Luiz Proença-Modena, Magnun Nueldo Nunes dos Santos, Angelica Schreiber, Julia Forato,Camila Simeoni, Marcilio Jorge Fumagalli, Mariene Ribeiro Amorim, Darlan da Silva Candido, Nuno Rodrigues Faria, Julien Theze, Luiz Gonzaga,Jaqueline Goes Jesus e William Marciel de Souza |
| EPI_ISL_476443 | Hospital da Clínicas da Faculdade de Medicina da Universidade de São Paulo | Instituto de Medicina Tropical da Univesidade de São Paulo | Samples: Ingra Morales Claro, Erika Regina Manuli, Cecilia Salete Alencar, Carolina S. Lazar, Silvia F. Costa; Sequencing: Ingra Morales Claro, Jaqueline Goes de Jesus, Erika Regina Manuli, Flavia Cristina da Silva Sales, Thais de Moura Coletti, Camila Alves Maia da Silva, Mariana Severo Ramundo, Giula Magalhaes Ferreira, Darlan da Silva Candido, Julien Theze, Nuno Faria, Ester Sabino |
| EPI_ISL_476559 | unknown | Laboratoire Sciences et Technologies de la Santé (STS) Institut Supérieur des Sciences de la Santé Université Hassan 1er, Settat, Morocco | Hajar Lemriss, Sanaâ Lemriss, Amal Souiri, Narjis Amar, Mustapha Mouallif, Touria Essayagh, Jawad Bouzid, Saâd EL Kabbaj, Abderraouf Hilali |
| EPI_ISL_476702, EPI_ISL_476704 | Incubadora Venezolana de Ciencia, Venezuela | Incubadora Venezolana de Ciencia, Venezuela / Instituto Nacional de Salud, Bogotá, Colombia / Grupo de Investigaciones Microbiológicas-UR (GIMUR), Departamento de Biología, Facultad de Ciencias Naturales, Universidad del Rosario, Bogotá, Colombia / Icahn School of Medicine at Mount Sinai, New York, USA | Alberto Paniz-Mondolfi, Marina Muñoz, Luis Perez-Garcia, Lourdes Delgado, Carolina Florez, Sergio Gomez, Angelica Rico, Lisseth Pardo, Esther C. Barros, Carolina Hernández, Jesús E. Jaimes, Aníbal A. Teherán, Ana S. Gonzalez-Reiche, Matthew M. Hernandez, Emilia Mia Sordillo, Viviana Simon, Harm van Bakel, Juan David Ramírez |
| EPI_ISL_476961 | KU Leuven, Rega Institute, Clinical and Epidemiological Virology | KU Leuven, Rega Institute, Clinical and Epidemiological Virology | Tony Wawina-Bokalanga, Joan Marti-Carerras, Bert Vanmechelen, Piet Maes |
| EPI_ISL_477169 | Department for Virology, Molecular Biology and Genome Research, R. G. Lugar Center for Public Health Research, National Center for Disease Control and Public Health (NCDC) of Georgia. | Department for Virology, Molecular Biology and Genome Research, R. G. Lugar Center for Public Health Research, National Center for Disease Control and Public Health (NCDC) of Georgia. | Tata Imnadze, Giorgi Tomashvili, Meri Pantsulaia, Gvantsa Brachveli, Gvantsa Chanturia, Ann Machablishvili, Nato Kotaria, Marine Murtskhvaladze, Lela Sabadze, Mari Gavashelidze, Ana Papkiauri, Tamar Jashiashvili, Tea Tevdoradze, Ketevan Sidamonidze, Ekaterine Khmaladze, Ekaterine Zhghenti, Roena Sukhiashvili, Mariam Zakalashvili, Lela Urushadze, Magda Dgebuadze, Davit Tsaguria, Ekaterine Zangaladze, Nino Berishvili, Adam Kotorashvili, Maia Alkhazashvili, Irma Burjanadze, Anna Kasradze, Khatuna Zakhashvili, Paata Imnadze, Amiran Gamkrelidze. |
| EPI_ISL_478718 | Sydney South West Pathology Service (SSWPS) - Concord Repatriation General Hospital - NSW Health Pathology | NSW Health Pathology - Institute of Clinical Pathology and Medical Research; Westmead Hospital; University of Sydney | CIDM-PH et al. |
| EPI_ISL_479800 | Sapporo City Institute of Public Health | Pathogen Genomics Center, National Institute of Infectious Diseases | Tsuyoshi Sekizuka, Asami Ohnishi, Kentaro Itokawa, Rina Tanaka, Masanori Hashino, Hajime Kamiya, Motoi Suzuki, Makoto Kuroda |
| EPI_ISL_480031 | Tochigi Prefectural Institute of Public Health and Environmental Science | Pathogen Genomics Center, National Institute of Infectious Diseases | Tsuyoshi Sekizuka, Ako Nakajima, Kentaro Itokawa, Rina Tanaka, Masanori Hashino, Hajime Kamiya, Motoi Suzuki, Makoto Kuroda |
| EPI_ISL_480298 | National Reference Laboratory "Influenza and acute respiratory diseases" | NRL-HIV | Ivan Ivanov, Ivailo Alexiev, Ivva Philipova |
| EPI_ISL_480326 | Hospital Nacional de Niños | Charité Virology-University of Costa Rica | Andres Moreira-Soto, Eugenia Corrales-Aguilar, Ignacio Postigo-Hidalgo, Cristian Pérez Corrales, Andrei Montero Bonilla, Jan Felix Drexler |
| EPI_ISL_480328 | Laboratorio LABIN | Charité Virology-University of Costa Rica | Andres Moreira-Soto, Eugenia Corrales-Aguilar, Ignacio Postigo-Hidalgo, Ignacio Soto Pacheco, Jan Felix Drexler |
| EPI_ISL_480332, EPI_ISL_480333, EPI_ISL_480335 | Microbial Genomics Laboratory, Institut Pasteur de Montevideo | Microbial Genomics Laboratory, Institut Pasteur de Montevideo | Cecilia Salazar, Marianoel Pereira, Ignacio Ferrés, Gonzalo Moratorio, Pilar Moreno, Gregorio Iraola |
| EPI_ISL_480435 | Laboratorio de Biología Molecular Asociación Española Primera en Salud | Departments of Pathology and Medicine, New York University School of Medicine | Maria Victoria Elizondo, Maria Noel Zubillaga, Gonzalo Manrique, Paul Zappile, Gael Westby, Matthew T Maurano, Christian Marier, Adriana Heguy |
| EPI_ISL_481207, EPI_ISL_481210 | Ostfold Hospital Trust - Kalnes, Centre for Laboratory Medicine, Section for gene technology and infection serology | Norwegian Institute of Public Health, Department of Virology | Kathrine Stene-Johansen, Kamilla Heddeland Instefjord, Hilde Elshaug, Rasmus Riis Kopperud, Karoline Bragstad, Olav Hungnes |
| EPI_ISL_481240 | Institut Pasteur Dakar | Institut Pasteur de Dakar | Ndongo Dia, Moussa Moise Diagne, Mamadou Diop, Marie Henriette Dior Ndione, Mamadou Malado Jallow, Safietou Sanke, Ousmane Faye, Amadou Alpha Sall. |
| EPI_ISL_481245 | Hospital IESS Babahoyo | Institute of Microbiology, Universidad San Francisco de Quito | Belén Prado-Vivar, Sully Márquez, Juan José Guadalupe, Monica Becerra-Wong, Carla Torres, Bernardo Gutiérrez, Francisco Cordova, Ninfa Henriquez, Killen Briones-Zamora, Killen Briones-Claudette, Verónica Barragán, Patricio Rojas-Silva, Gabriel Trueba, Michelle Grunauer, Paúl Cárdenas |
| EPI_ISL_481584, EPI_ISL_481653 | Department of Virology and Immunology, University of Helsinki and Helsinki University Hospital, Huslab Finland | Department of Virology, Faculty of Medicine, University of Helsinki, Helsinki, Finland | Teemu Smura, Hannimari Kallio-Kokko, Jenni Virtanen, Maija Suvanto, Sari Hannula, Harri Kangas, Pekka Ellonen, Olli Vapalahti |
| EPI_ISL_482394 | Providence St. Joseph Health Molecular Genomics Laboratory | Providence St. Joseph Health Molecular Genomics Laboratory | Alexa K Dowdell, Brian D Piening, Fred L Robinson, Carlo B Bifulco, Mary Campbell |
| EPI_ISL_483645 | National Institute of Laboratory Medicine and Referral Center | Genomic Research Lab, BCSIR | Md. Saddam Hossain, Abu Sayeed Mohammad Mahmud, Mohammad Samir Uzzaman, Eshrar Osman, Md. Ahasan Habib, Shahina Akter, Tanjina Akhter Banu, Md. Murshed Hasan Sarkar, Barna Goswami, Iffat Jahan, Tasnim Nafisa, Md. Maruf Ahmed Molla, Mahmuda Yeasmin, Asish Kumar Ghosh, A. K. M. Shamsuzzaman, Sheikh Md. Selim Al Din, Utpal Chandra Ray, Salek Ahmed Sajib, Md. Salim Khan |
| EPI_ISL_485399 | Institute of Human Genetics, Polish Academy of Sciences | Institute of Human Genetics, Polish Academy of Sciences | Szymon Hryhorowicz, Adam Ustaszewski, Marta Kaczmarek-Ry, Emilia Lis, Ewa Zitkiewicz, Micha Witt, Andrzej Pawski |
| EPI_ISL_486412, EPI_ISL_486418 | Centrl laboratorija | Latvian Biomedical Research and Study Centre | Ivars Silamielis, Kaspars Megnis, Monta Ustinova, ikita Zrelovs, Vita Rovte, Stella Lapia, Jana Oste, Marta Priedte, Uga Dumpis, Jnis Kloviš |
| EPI_ISL_487453 | CICM-Mali | Bundeswehr Institut of Microbiology | Kouriba, Dürr, Sangaré, Rehn, Traoré, Bestehorn-Willmann, Walter, Quedraogo, Zimmermann, Maiga, Heitzer, Sogodogo, Antwerpen, Wölfel |
| EPI_ISL_489973 | Viollier AG | Department of Biosystems Science and Engineering, ETH Zürich | Christian Beisel, Sarah Nadeau, Ivan Topolsky, Pedro Ferreira, Philipp Jablonski, Susana Posada-Céspedes, Tobias Schär, Ina Nissen, Natascha Santacroce, Elodie Burcklen, Christiane Beckmann, Maurice Redondo, Olivier Kobel, Christoph Noppen, Sophie Seidel, Noemie Santamaria de Souza, Niko Beerenwinkel, Tanja Stadler |
| EPI_ISL_489996, EPI_ISL_490001, EPI_ISL_490003 | King Fahad Medical City | King Fahad Medical City | Alosaimi,B., Naeem,A., Alghoraibi,M., Enani,M. |
| EPI_ISL_490036 | Pathology West - NSW Health Pathology | NSW Health Pathology - Institute of Clinical Pathology and Medical Research; Westmead Hospital; University of Sydney | CIDM-PH et al. |

| | | | |
|---|---|---|---|
| EPI_ISL_490078 | National Public Health Laboratory, National Centre for Infectious Diseases | National Public Health Laboratory, National Centre for Infectious Diseases | Mak TM, Octavia S, Zhou Z, Chavatte JM, Cui L, Lin RTP |
| EPI_ISL_490090, EPI_ISL_490103 | Institute for Medical Research, Infectious Disease Research Centre, National Institutes of Health, Ministry of Health Malaysia | Institute for Medical Research, Infectious Disease Research Centre, National Institutes of Health, Ministry of Health Malaysia | Suppiah J, Mohd-Zawawi Z, Kamel K, Kalyanasundram J, Thayan R |
| EPI_ISL_490105 | South Eastern Area Laboratory Services (SEALS) | NSW Health Pathology - Institute of Clinical Pathology and Medical Research; Westmead Hospital; University of Sydney | CIDM-PH et al. |
| EPI_ISL_491051, EPI_ISL_491056, EPI_ISL_491057 | Suceava County Emergency Hospital | "Stefan cel Mare" University Metagenomics Lab | Lobiuc Andrei et al. |
| EPI_ISL_491064, EPI_ISL_491079 | Suceava County Emergency Hospital | "Stefan cel Mare" University Metagenomics Lab | Lobiuc Andrei, Antoniadis Panagiotis et al. |
| EPI_ISL_491298 | Rural Health Unit - Calauan, Laguna | Research Institute for Tropical Medicine | Tujan, M.A.A., Onza,O.J.T., Polotan,F.G.M., Medado,I.A.P., Bautista,C.T., Brunker, K., Mercado,E.S., Manalo, D.L., Demetria, C.S. |
| EPI_ISL_492075 | Institute for Public Health of the Republic of North Macedonia | Charite Universitatsmedizin Berlin, Institute of Virology | Victor M Corman, Joern Beheim-Schwarzbach, Barbara Muhlemann, Talitha Veith, Julia Schneider, Elizabeta Jancheska, Maja Kuzmanovska, Golubinka Bosevska, Terry Jones, Christian Drosten |
| EPI_ISL_493338 | Instituto de Diagnostico y Referencia Epidemiologicos (INDRE) | Instituto de Diagnostico y Referencia Epidemiologicos (INDRE) | Gisela Barrera-Badillo , Abril Rodriguez-Maldonado, Claudia Wong-Arambula , Natividad Cruz-Ortiz, Tatiana Nunez-Garcia, Dayanira Arellano-Suarez, Fabiola Garces-Ayala, Edgar Mendieta-Condado, Lucia Hernandez-Rivas, Irma Lopez-Martinez, Ernesto Ramirez-Gonzalez. |
| EPI_ISL_495411 | Kafkas University, Faculty of Medicine, Department of Medical Microbiology | Kafkas University, Faculty of Medicine, Department of Medical Microbiology | Murat Karamese, Didem Ozgur, E. Ediz Tutuncu |
| EPI_ISL_496772 | Gorgas Memorial Laboratory of Health Studies | Gorgas Memorial Laboratory of Health Studies | Danilo Franco, Claudia Gonzalez Sandra Lopez-Verges, Alexander A Martinez |
| EPI_ISL_497823, EPI_ISL_497824, EPI_ISL_497827 | Department of Microbiology, The University of Hong Kong | Department of Microbiology, The University of Hong Kong | Kelvin K.W. To, Kwok-Yung Yuen |
| EPI_ISL_497874 | National Centre For Cell Science | National Centre For Cell Science | Dhiraj Paul, Kunal Jani, Radha Chauhan, Janesh Kumar, Vasudevan Seshadri, Girdhari Lal, Rajesh Karyakarte, Suvarna Joshi, Murlidhar Tambe, Sourav Sen, Santosh Karade, Kavita Bala Anand, Shelinder Pal Singh Shergill, Rajiv Mohan Gupta, Manoj Kumar Bhat, Arvind Sahu, Maharashtra COVID-19 Study Group, DBT's PAN-INDIA 1000 SARS-CoV2 RNA genome sequencing consortium, Yogesh S Shouche |
| EPI_ISL_498000 | Division of Viral Diseases, Center for Laboratory Control of Infectious Diseases, Korea Centers for Diseases Control and Prevention | Division of Viral Diseases, Center for Laboratory Control of Infectious Diseases, Korea Centers for Diseases Control and Prevention | Jeong-Min Kim, Yoon-Seok Chung, Namjoo Lee, Sang Hee Woo, Hye-Jun Jo, Heui Man Kim, Jun-Sub Kim, Myung Guk Han |
| EPI_ISL_498239 | Institut Pasteur de Dakar | Institut Pasteur de Dakar | Ndongo Dia, Moussa Moise Diagne, Mamadou Diop, Marie Henriette Dior Ndione, Mamadou Malado Jallow, Safietou Sankhe Mbengue, Ousmane Faye, Amadou Alpha Sall. |
| EPI_ISL_498526, EPI_ISL_498534, EPI_ISL_498543, EPI_ISL_498548 | ACT Pathology | Schwessinger Lab | Ashley Jones, Benjamin Schwessinger, Robert Lanfear, Robyn N Hall, Megan McDonald, Ming-Dao Chia, Kevin Murray, Craig Kennedy, Karina Kennedy |
| EPI_ISL_498817 | National Institute of Laboratory Medicine and Referral Center | Genomic Research Lab, BCSIR | Abu Sayeed Mohammad Mahmud, Mohammad Samir Uzzaman, Eshrar Osman, Md. Ahasan Habib, Shahina Akter, Tanjina Akhter Banu, Md. Murshed Hasan Sarkar, Barna Goswami, Iffat Jahan, Md. Saddam Hossain, Tasnim Nafisa, Md. Maruf Ahmed Molla, Mahmuda Yeasmin, Asish Kumar Ghosh, A. K. M. Shamsuzzaman, Sheikh Md. Selim Al Din, Utpal Chandra Ray, Salek Ahmed Sajib, Md. Salim Khan |
| EPI_ISL_500530 | Mayo Clinic Laboratories | University of Washington Virology Lab | Pavitra Roychoudhury, Hong Xie, Lasata Shrestha, Amin Addetia, Truong Nguyen, Victoria M Rachleff, Meei-Li Huang, Keith R Jerome, Alexander Greninger |
| EPI_ISL_500548 | Singapore General Hospital | Department of Microbiology | Nurdyana Abdul Rahman, Kun Lee Lim, Chenhao Li, Kian Sing Chan, Lynette Oon, Kern Rei Chng, Niranjan Nagarajan, Karrie Ko |
| EPI_ISL_500940 | Viollier AG | Department of Biosystems Science and Engineering, ETH Zürich | Christian Beisel, Sarah Nadeau, Ivan Topolsky, Pedro Ferreira, Philipp Jablonski, Susana Posada-Céspedes, Tobias Schär, Ina Nissen, Natascha Santacroce, Elodie Burcklen, Christiane Beckmann, Maurice Redondo, Olivier Kobel, Christoph Noppen, Sophie Seidel, Noemie Santamaria de Souza, Niko Beerenwinkel, Tanja Stadler |
| EPI_ISL_501190, EPI_ISL_501204, EPI_ISL_501207, EPI_ISL_501222 | Department of Medical Microbiology, University Malaya Medical Centre | Department of Medical Microbiology, Faculty of Medicine, University of Malaya | Yoong Min CHONG, Jennifer Chong, I-Ching SAM, Yoke Fun CHAN, University Malaya Medical Centre COVID Team |
| EPI_ISL_501231, EPI_ISL_501233, EPI_ISL_501236 | Hellenic Pasteur Institute, National Influenza Reference laboratory of Southern Greece & Unit of Bioinformatics and Applied Genomics | Hellenic Pasteur Institute, National Influenza Reference laboratory of Southern Greece & Unit of Bioinformatics and Applied Genomics | Vasiliki Pogka, Timokratis Karamitros, Athanasios Kossyvakis, Antonios Kalliaropoulos, Horefti Elina, Evangelidou Maria, Androniki Voulgari-Kokota, Aspasia Kontou, Andreas Mentis |
| EPI_ISL_504203 | National Institute for Communicable Diseases of the National Health Laboratory Service | National Institute for Communicable Diseases of the National Health Laboratory Service | Allam M, Ismail A, Khumalo Z, Kwenda S, Mtshali P, Mnyameni F, Mohale T, Bhiman JN |
| EPI_ISL_508134, EPI_ISL_508605, EPI_ISL_508606, EPI_ISL_508607 | SA Pathology | SA Pathology | Lex Leong, Chuan Kok Lim, Mark Turra, Ivan Bastian, Geoff Higgins |
| EPI_ISL_508862, EPI_ISL_508863 | Virology Unit, Institut Pasteur de Madagascar | Virology Unit, Institut Pasteur de Madagascar | Christian Ranaivoson, Cara Brook, Norosoa Razanajatovo, Vida Ahyong, Tsiry Randriambolamanantsoa, Michelle Tan, Vololoniaina Raharinosy, Helisoa Razafimanjato, Cristina M. Tato, Joseph L. DeRisi, Soa Fy Andriamandimby, Jean-Michel Heraud |
| EPI_ISL_509498, EPI_ISL_509505, EPI_ISL_509516, EPI_ISL_509523 | Area of Virology, Serology and Virology Division (SAViD), New South Wales Health Pathology Randwick | Area of Virology, Serology and Virology Division (SAViD), New South Wales Health Pathology Randwick | Rawlinson, W. |
| EPI_ISL_510430 | Hospital Universitario Virgen de las Nieves de Granada-SAS | SeqCOVID-SPAIN consortium/IBV(CSIC) | Mercedes Pérez Ruiz, Sara Sanbonmatsu Gámez, Irene Pedrosa Corral, José M. Navarro-Marí and SeqCOVID-SPAIN consortium |
| EPI_ISL_510544 | SA Pathology | SA Pathology | Lex Leong, Chuan Kok Lim, Mark Turra, Ivan Bastian, Geoff Higgins |
| EPI_ISL_510569, EPI_ISL_510646 | Division of Viral Diseases, Center for Laboratory Control of Infectious Diseases, Korea Centers for Diseases Control and Prevention | Division of Viral Diseases, Center for Laboratory Control of Infectious Diseases, Korea Centers for Diseases Control and Prevention | Jeong-Min Kim, Yoon-Seok Chung, Namjoo Lee, Sang Hee Woo, Hye-Jun Jo, Heui Man Kim, Jun-Sub Kim, Myung Guk Han |
| EPI_ISL_511105, EPI_ISL_511171 | Instituto Nacional de Saude (INSA) | Instituto Nacional de Saude (INSA) | Borges et al |
| EPI_ISL_511229 | Instituto Nacional de Saude (INSA) and Instituto Gulbenkian de Ciencia (IGC) | Instituto Nacional de Saude (INSA) and Instituto Gulbenkian de Ciencia (IGC) | Borges et al |
| EPI_ISL_512618 | National Laboratory for Influenza/Virology reference laboratory, Public Health Center of the Ministry of Health of Ukraine | Respiratory Virus Unit, Microbiology Services Colindale, Public Health England | PHE Covid Sequencing Team, Dr. Iryna Demchyshyna |
| EPI_ISL_512647, EPI_ISL_512649 | Latvijas Infektoloijas centrs | Latvian Biomedical Research and Study Centre | Ivars Silamielis, Kaspars Megnis, Monta Ustinova, ikita Zrelovs, Vita Rovte, Jeena Storoženko, Tatjana Kolupajeva, Oksana Savicka, Uga Dumpis, Jnis Kloviš |
| EPI_ISL_512668 | Area De Salud Corredores | Inciensa, Instituto Costarricense de Investigación y Enseñanza en Nutrición y Salud | Francisco Duarte, Hebleen Porras, Claudio Soto-Garita, Estela Cordero, Adriana Godinez & Melany Calderon |
| EPI_ISL_512742 | PathWest Laboratory Medicine WA | PathWest Laboratory Medicine WA Microbial Surveillance Unit | PathWest Laboratory Medicine WA Microbial Surveillance Unit |
| EPI_ISL_512811, EPI_ISL_512812 | Kenema Government Hospital, Ministry of Health and Sanitation | Kenema Government Hospital, Ministry of Health and Sanitation | Goba,A., Momoh,M., Sandi,J., Tomkins-Tinch,C., Siddle,K., Mehta,S., Oluniyi,P., Jalloh,S., Park,D., Andersen,K., Garry,R., Happi,C., Grant,D., Olawoye,I. |

| | | | |
|---|---|---|---|
| EPI_ISL_512832 | National Public Health Laboratory, National Centre for Infectious Diseases | National Public Health Laboratory, National Centre for Infectious Diseases | Mak TM, Octavia S, Zhou Z, Chavatte JM, Cui L, Lin RTP |
| EPI_ISL_512844 | Department of Medical Research | DMR_Myanmar | Myat Htut Nyunt, Hnin Ohnmar Soe, Kay Thi Aye, Wah Wah Aung,Yi Yi Kyaw, Aung Kyaw Kyaw, Theingi Win Myat, Phyu Win Ei, Aung Zaw Latt, Nan Aye Thida Oo, Lai Lai San, Su Mon Win, Ni Ni Zaw, Htin Lin, Hlaing Myat Thu, Zaw Than Htun |
| EPI_ISL_513338 | Pathology West - NSW Health Pathology | NSW Health Pathology - Institute of Clinical Pathology and Medical Research; Westmead Hospital; University of Sydney | CIDM-PH et al. |
| EPI_ISL_513580 | Programa de Oncovirologia, Instituto Nacional de Câncer | Programa de Oncovirologia, Instituto Nacional de Câncer | Juliana D. Siqueira, Livia R. Goes, Brunna M. Alves, Claudia Cicala,James Arthos, João P.B. Viola, Andreia C. de Melo, Marcelo A. Soares |
| EPI_ISL_514118, EPI_ISL_514124 | Viral Respiratory Lab, National Institute for Biomedical Research (INRB) | Pathogen Sequencing Lab, National Institute for Biomedical Research (INRB) | Placide Mbala-Kingebeni, Edith Nkwembe, Eddy Kinganda-Lusamaki, Amuri Aziza, Francisca Muyembe Mawete, Emmanuel Lokilo Lofiko, Catherine Pratt, Matthias Pauthner, Josh Quick, Allison Black, James Hadfield, Trevor Bedford, Ian Goodfellow, Andrew Rambaut, Nick Loman, Kristian Andersen, Michael Wiley, Steve Ahuka-Mundeke, Jean-Jacques Muyembe Tamfum |
| EPI_ISL_514226 | Laboratorio de Referencia Nacional de Virus Respiratorio. Instituto Nacional de Salud Perú | Laboratorio de Referencia Nacional de Biotecnología y Biología Molecular. Instituto Nacional de Salud Perú | Carlos Padilla Rojas, Karolyn Vega Chozo, Priscila Lope Pari, Omar Caceres Rey, Marco Galarza Perez, Maribel Huaringa Nuñez, Johanna Balbuena Torrez, Henri Bailon Calderon, Nancy Rojas Serrano |
| EPI_ISL_514300 | Israel Central Virology laboratory | Israel Central Virology laboratory | Neta Zuckerman, Efrat Dahan Bucris, Oran Erster, Ella Mendelson, Michal Mandelboim |
| EPI_ISL_514615 | NSTU COVID-19 Diagnostic Center | NSU Genome Research Institute (NGRI), North South University | Dr. Muhammad Maqsud Hossain, Aura Rahman, Prof. Firoz Ahmed, Tahrima Huq, Abdus Sadique, Jahidul Alam, Tamanna Afroze, Md Aminul Islam, Prof. Md. Didar-Ul-Alam, Prof. Kazi Nadim Hasan, Prof. Abdul Khaleque, Prof, Hasan Mahmud Reza |
| EPI_ISL_515040 | Division of Viral Diseases, Center for Laboratory Control of Infectious Diseases, Korea Centers for Diseases Control and Prevention | Division of Viral Diseases, Center for Laboratory Control of Infectious Diseases, Korea Centers for Diseases Control and Prevention | Jeong-Min Kim, Yoon-Seok Chung, Namjoo Lee, Sang Hee Woo, Hye-Jun Jo, Heui Man Kim, Jun-Sub Kim, Myung Guk Han |
| EPI_ISL_515094, EPI_ISL_515103 | Department of Biochemistry, Cell and Molecular Biology | WACCBIP, University of Ghana | Ngoi,J.M., Quashie,P., Morang'a,C.M., Amuzu,D.S., Adu,B., Kumordjie,S., Eshun,M., Boatemaa,L., Magnussen,V., Kotey,E., Tei-Maya,F., Arjarquah,A., Mutungi,J.K., Bediako,Y., Asante,I., Bonney,E., Kyei,G.B., Bonney,K., Amenga-Etego,L.N., Anang,A.K., Awandare,G.A., Ampofo,W. |
| EPI_ISL_515466 | Discovery DNA | Discovery DNA | Dustin Hittel, Marina Kerr, Leo Dimnik, Desmond Koo, Alice Li, Aneal Khan |
| EPI_ISL_515851 | Medical Disagnostics Services (MDS) | KRISP, KZN Research Innovation and Sequencing Platform | Giandhari J, Pillay S, Lessells R, ChimukangaraB, Mdlalose K, York D, Khan S, Tegally H, Wilkinson E, de Oliveira T |
| EPI_ISL_516622 | Instituto de Diagnostico y Referencia Epidemiologicos (INDRE) | Instituto de Diagnostico y Referencia Epidemiologicos (INDRE) | Gisela Barrera-Badillo , Abril Rodriguez-Maldonado, Claudia Wong-Arambula , Natividad Cruz-Ortiz, Tatiana Nunez-Garcia, Dayanira Arellano-Suarez, Fabiola Garces-Ayala, Edgar Mendieta-Condado, Lucia Hernandez-Rivas, Irma Lopez-Martinez, Ernesto Ramirez-Gonzalez. |
| EPI_ISL_516650 | Institute of Microbiology, Universidad San Francisco de Quito | Institute of Microbiology, Universidad San Francisco de Quito | Prado-Vivar, Sully Márquez, Juan José Guadalupe, Monica Becerra-Wong, Bernardo Gutiérrez, Verónica Barragán, Patricio Rojas-Silva, Gabriel Trueba, Michelle Grunauer, Paúl Cárdenas |
| EPI_ISL_516930 | Department for Molecular Diagnostics, Centre for Medical Microbiology, Institute of Public Health of Montenegro | Charite Universitatsmedizin Berlin, Institut fur Virologie | Victor M Corman, Terry Jones, Jörn Beheim-Schwarzbach, Barbara Muehlemann, Talitha Veith, Julia Schneider, Marija Govedarica and Danijela Vujoševi, Christian Drosten |
| EPI_ISL_516938 | Nicolae Testemitanu State University of Medicine and Pharmacy | International Centre for Genetic Engineering and Biotechnology (ICGEB) and ARGO Open Lab Platform for Genome Sequencing | Ulinici M, Licastro D, Dal Monego S, Rajasekharan S, Marcello A |
| EPI_ISL_517616, EPI_ISL_517627, EPI_ISL_517644, EPI_ISL_517661, EPI_ISL_517662 | Academic Hospital Paramaribo | Erasmus Medical Center | Bas Oude Munnink, Dion Gajadin, Ed Ijzerman, Emmanuelle Munger, Gary Gummels, Ingrid Krishnadath, Lycke Woittiez, Marion Koopmans, Mireille Van de Veer, Princes Wongsowidjojo, Radjesh Ori, Rohma Banwari, Stephen Vreden |
| EPI_ISL_518835 | Oman-National Influenza Center | Biotechnology & OMICs Laboratory, Natural & Medical Sciences Research Center, University of Nizwa | Samira Al-Mahruqi, Abdul Latif Khan, Samiha Al-Kharusi, Adil Khan , Ahmed Al-Rawahi, Sajjad Asaf, Amina Al-Jardani, Hanan Al-Kindi, Intisar Al-Shukri, Adil Al-Wahaibi, Seif Al-Abri, Ahmed Al-Harrasi |
| EPI_ISL_521860 | Royal Darwin Hospital Pathology | MDU-PHL | Meumann, E., Caly L., Seemann T., Sait, M., Schultz M., Druce J., Sherry, N. |
| EPI_ISL_522568, EPI_ISL_522590, EPI_ISL_522705 | Royal Hobart Hospital Microbiology Department | MDU-PHL | Cooley L., van Haeften R., Seemann T., Sait, M., Schultz, M.B., Sherry N. |
| EPI_ISL_523255 | Dutch COVID-19 response team | Erasmus Medical Center | Bas Oude Munnink, David Nieuwenhuijse, Reina Sikkema, Claudia Schapendonk, Irina Chestakova, Anne van der Linden, Theo Bestebroer, Stefan van Nieuwkoop, Mark Pronk, Pascal Lexmond, Corien Swaan, Manon Haverkate, Madelief Mollers, Mart Stein, Sandra Kengne Kamga Mobou, Jeroen van Kampen, Jolanda Voermans, Aura Timen, Corine GeurtsvanKessel, Annemiek van der Eijk, Richard Molenkamp, Marion Koopmans, on behalf of the Dutch national COVID-19 response team. |
| EPI_ISL_525353 | National Virus Reference Laboratory | National Virus Reference Laboratory | Michael Carr, Gabriel Gonzalez, Jonathan Dean, Aditi Chaturvedi, Suzie Coughlan, Cillian F De Gascun |
| EPI_ISL_525432 | Institute of Microbiology, Universidad San Francisco de Quito | Institute of Microbiology, Universidad San Francisco de Quito | Juan José Guadalupe, Monica Becerra-Wong, Belén Prado-Vivar, Sully Márquez, Ligia Briceño, Carlos Mena, Nabih Dahik, Bernardo Gutiérrez, Verónica Barragán, Patricio Rojas-Silva, Gabriel Trueba, Michelle Grunauer, Paúl Cárdenas |
| EPI_ISL_525469 | Universidad Iberoamericana | International Centre for Genetic Engineering and Biotechnology (ICGEB) and ARGO Open Lab Platform | Robert Paulino-Ramirez, Eileen Riego, Alejandro Vallejo Degaudenzi, Victor Virgilio Calderon, Leandro Tapia, Patricia Leon, Danilo Licastro, Simeone Dal Monego, Sreejith Rajasekharan and Alessandro Marcello. |
| EPI_ISL_525474 | Centre for Dengue Research | Centre for Dengue Research, USJ, SL | Chandima Jeewandara, Deshni Jayathilaka, Dinuka Ariyaratne, Laksiri Gomes, Diyanath Ranasinghe, Dinuka Guruge, Ruwan Wijayamuni, Gathsaurie Neelika Malavige |
| EPI_ISL_527012, EPI_ISL_527024 | Area of Virology, Serology and Virology Division (SAViD), New South Wales Health Pathology Randwick | Area of Virology, Serology and Virology Division (SAViD), New South Wales Health Pathology Randwick | Rawlinson, W. |
| EPI_ISL_527815 | Institute of Microbiology, Universidad San Francisco de Quito | Institute of Microbiology, Universidad San Francisco de Quito | Belén Prado-Vivar, Sully Márquez, Juan José Guadalupe, Monica Becerra-Wong, Bernardo Gutiérrez, Khurram Mahbbob, Verónica Barragán, Patricio Rojas-Silva, Gabriel Trueba, Michelle Grunauer, Paúl Cárdenas |
| EPI_ISL_527875 | Nigeria Centre for Disease Control (NCDC) | African Centre of Excellence for Genomics of Infectious Diseases (ACEGID), Redeemer's University, Ede, Osun State, Nigeria | Oluniyi P.E. et al |
| EPI_ISL_528382 | Translational Health Science and Technology Institute -ESIC medical college and hospital, Faridabad | THSTI Bioassay laboratory | Saurabh Kumar, Jigme Wangchuk, Anil Kumar Pandey, Asim Das, Guruprasad R. Medigeshi |
| EPI_ISL_528743 | Malaysia Genome Institute | Malaysia Genome Institute | Mohd Noor Mat Isa, Irni Suhayu Sapian, Yusuf Muhammad Noor, Nurhezreen Md Iqbal, Mohd Faizal Abu Bakar, Enizza Kasim, Shamsidar Sopie, Siti Noraini Othman, Azrin Ahmad, Nor Azfa Johari, Shahrul Hisham Zainal Ariffin |
| EPI_ISL_528943 | Agenzia di Tutela della Salute di Bergamo | Istituto Zooprofilattico Sperimentale dell'Abruzzo e Molise "G.Caporale" | Lorusso A, Marcacci M, Di Domenico M, Curini V, Ancora M, Cammà C, Rinaldi A, Mangone I, Di Pasquale A, Puglia I, Savini G. |
| EPI_ISL_529010 | Servizio Igiene Epidemiologia e Sanità Pubblica (SIESP)-L'Aquila | Istituto Zooprofilattico Sperimentale dell'Abruzzo e Molise "G.Caporale" | Lorusso A, Marcacci M, Di Domenico M, Curini V, Ancora M, Cammà C, Rinaldi A, Mangone I, Di Pasquale A, Puglia I, Savini G. |
| EPI_ISL_530232, EPI_ISL_530234, EPI_ISL_530236, EPI_ISL_530239, EPI_ISL_530240, EPI_ISL_530244, EPI_ISL_530246, EPI_ISL_530250, EPI_ISL_530251, EPI_ISL_530252, EPI_ISL_530256, EPI_ISL_530276, EPI_ISL_530277 | | | |
| see above | Queensland Health Forensic and Scientific Services, Public Health Virology | Public Health Virology Laboratory, Forensic and Scientific Services, Queensland Health | Son Nguyen et al |
| EPI_ISL_533115 | Lighthouse Lab in Glasgow | Wellcome Sanger Institute for the COVID-19 Genomics UK (COG-UK) consortium | Harper VanSteenhouse, Yumi Kasai, David Gray, Carol Clugston, Anna Dominiczak and Alex Alderton, Roberto Amato, Sonia Goncalves, Ewan Harrison, David K. Jackson, Ian Johnston, Dominic Kwiatkowski, Cordelia Langford, John Sillitoe |

| | | | |
|---|---|---|---|
| EPI_ISL_534248 | Laboratoriemedicin Vasternorrland | The Public Health Agency of Sweden | Anna-Malin Linde, Maria Lind Karlberg, Mattias Haukland, Reza Advani, Olov Svartstrom, Oskar Karlsson Lindsjo, Sandra Broddesson, Petra Edquist, Mia Brytting, Anna Risberg, Karin Tegmark-Wisell |
| EPI_ISL_534336 | Department of Laboratory Medicine, National Taiwan University Hospital | Microbial Genomics Core Lab, National Taiwan University Centers of Genomic and Precision Medicine | Shiou-Hwei Yeh, You-Yu Lin, Ya-Yun Lai, Chiao-Ling Li, Shan-Chwen Chang, Pei-Jer Chen, Sui-Yuan Chang |
| EPI_ISL_535263, EPI_ISL_535274 | New Mexico Department of Health Scientific Laboratory | New Mexico Department of Health Scientific Laboratory | Ellie Johnson, Anastacia Griego-Fisher, D'Eldra Malone |
| EPI_ISL_535362 | Oklahoma Animal Disease Diagnostic Laboratory | Oklahoma Animal Disease Diagnostic Laboratory | Sai Narayanan, John C Ritchey, Girish Patil, Teluguakula Narasaraju, Sunil More, Jerry Malayer, Jeremiah Saliki, Anil Kaul, Akhilesh Ramachandran |
| EPI_ISL_535527 | NHLS-IALCH | KRISP, KZN Research Innovation and Sequencing Platform | Giandhari J, Pillay S, Lessells R, Mdlalose K, York D, Khan S, Tegally H, Wilkinson E, de Oliveira T |
| EPI_ISL_536736 | University of Wisconsin-Madison AIDS Vaccine Research Laboratories | University of Wisconsin-Madison AIDS Vaccine Research Laboratories | Gage Moreno, Katarina Braun, et al. AIDS Vaccine Research Laboratories |
| EPI_ISL_538365 | Kingston Health Sciences Centre / Queen's University | Ontario Institute for Cancer Research | Prameet M. Sheth, Calvin Sjaarda, Robert Colautti, Katya Douchant, Ilinca Lungu, Bernard Lam, Paul Krzyzanowski, Michael Laszloffy, Lawrence E Heisler, Richard de Borja, Jared T. Simpson |
| EPI_ISL_539496 | Hospital Nostra Senyora de Meritxell | Instituto de Salud Carlos III | Iglesias-Caballero, M. Molinero Calamita, M. González-Esguevillas, M. Camarero, S. Pozo, F. Casas, I. Jiménez, P. Jiménez, M. Zaballos, A. Monzón, S. Varona, S. Juliá, M. Cuesta, I, F. Fernández |
| EPI_ISL_539587 | ZOTZ KLIMAS MVZ Düsseldorf-Centrum GbR ÜBAG für Labormedizin, Genetik, Zytologie, Pathologie | Center of Medical Microbiology, Virology, and Hospital Hygiene, University of Duesseldorf | Maximilian Damagnez, Alexander Dilthey, Ashley-Jane Duplessis, Patrick Finzer, Katrin Hoffmann, Torsten Houwaart, Malte Kohns Vasconcelos, Marek Korencak, Nadine Lübke, Jessica Nicolai, Klaus Pfeffer, Daniel Strelow, Jörg Timm, Andreas Walker, Tobias Wienemann, Rainer Zotz |
| EPI_ISL_539793 | Institute of Microbiology, Universidad San Francisco de Quito | Institute of Microbiology, Universidad San Francisco de Quito | Belén Prado-Vivar, Sully Márquez, Juan José Guadalupe, Monica Becerra-Wong, Bernardo Gutiérrez, Ligia Briceño, Nabih Dahik, Verónica Barragán, Patricio Rojas-Silva, Gabriel Trueba, Michelle Grunauer, Paúl Cárdenas |
| EPI_ISL_539836 | Minnesota Department of Health, Public Health Laboratory | Minnesota Department of Health, Public Health Laboratory | Matt Plumb, Jacob Garfin, and Xiong Wang |
| EPI_ISL_539879 | Microbiology Division, South Carolina Department of Health and Environmental Control | Microbiology Division, South Carolina Department of Health and Environmental Control | Flores,H. |
| EPI_ISL_540935 | Laboratorio de Referencia Nacional de Virus Respiratorios, Instituto Nacional de Salud Peru | Laboratorio de Genómica Microbiana, Universidad Peruana Cayetano Heredia | Pablo Tsukayama, Alejandra Dávila-Barclay, Luis González, Pedro E. Romero, Brenda Ayzanoa, Janet Huancachoque, Pool Marcos, Maribel Huaringa, Camila Castillo-Vilcahuaman, Guillermo Salvatierra |
| EPI_ISL_541332 | The National Institute of Public Health | State Veterinary Institute Prague | Nagy,A; Jirincova,H; Novakova,L; Trnka,D; Vecerova,J |
| EPI_ISL_541551 | University of Wisconsin-Madison Campus AIDS Vaccine Research Laboratories | University of Wisconsin-Madison AIDS Vaccine Research Laboratories | Gage Moreno, Katarina Braun, et al. AIDS Vaccine Research Laboratories |
| EPI_ISL_541655 | Laboratory Diagnostic, Veterinary Specialized Institute Kraljevo | Laboratory Diagnostic, Veterinary Specialized Institute Kraljevo | Vidanovic,D., Tesovic,B., Knezevic,A., Jovanovic,T., Jankovic,M., Sekler,M., Banovic Djeri,B., Volkening,J., Afonso,C., Petrovic,T. |
| EPI_ISL_542235 | ASST GOM Niguarda | Dep. Of Oncology and Hemato-Oncology University of Milan | Claudia Alteri, Valeria Cento, Antonio Piralla, Valentino Costabile, Monica Tallarita, Luna Colagrossi, Silvia Renica, Federica Giardina, Federica Novazzi, Stefano Gaiarsa, Elisa Matarazzo, Maria Antonello, Chiara Vismara, Roberto Fumagalli, Oscar Massimiliano Epis, Massimo Puoti, Carlo Federico Perno, Fausto Baldanti |
| EPI_ISL_545007 | Histopath | NSW Health Pathology - Institute of Clinical Pathology and Medical Research; Westmead Hospital; University of Sydney | CIDM-PH et al. |
| EPI_ISL_545033 | Pathology West - NSW Health Pathology | NSW Health Pathology - Institute of Clinical Pathology and Medical Research; Westmead Hospital; University of Sydney | CIDM-PH et al. |
| EPI_ISL_547445, EPI_ISL_547449, EPI_ISL_547452 | Dutch COVID-19 response team | National Institute for Public Health and the Environment (RIVM) | Adam Meijer, Harry Vennema, Jeroen Cremer, Sharon van den Brink, Bas van der Veer, AnneMarie van den Brandt, Florian Zwagemaker, Dennis Schmitz, Chantal Reusken, on behalf of the national COVID-19 response team |
| EPI_ISL_547985 | Canterbury Health Laboratories | Institute of Environmental Science and Research (ESR) | Xiaoyun Ren, Matt Storey, Nikki Freed, Muhammad Faisal, Jing Wang, Hermes Perez, Anja Werno, Antje van der Linden, Arlo Upton, Chris Mansell, David Hammer, Dragana Drinkovic, Gary McAuliffe, Hana Sofia Andersson, James Ussher, Jill Sherwood, Josh Freeman, Julia Howard, Juliet Elvy, Mary DeAlmeida, Matt Blakiston, Matthew Rogers, Max Bloomfield, Michael Addidle, Michelle Balm, Sally Roberts, Sarah Jefferies, Sharmini Muttaiyah, Susan Morpeth, Susan Taylor, Timothy Blackmore, Vani Sathyendran, Veronica Playle, Virginia Hope, Erasmus Smit, Lauren Jelly, Olin Silander, Joep de Ligt |
| EPI_ISL_548012 | Middlemore Hospital | Institute of Environmental Science and Research (ESR) | Xiaoyun Ren, Matt Storey, Nikki Freed, Muhammad Faisal, Jing Wang, Hermes Perez, Anja Werno, Antje van der Linden, Arlo Upton, Chris Mansell, David Hammer, Dragana Drinkovic, Gary McAuliffe, Hana Sofia Andersson, James Ussher, Jill Sherwood, Josh Freeman, Julia Howard, Juliet Elvy, Mary DeAlmeida, Matt Blakiston, Matthew Rogers, Max Bloomfield, Michael Addidle, Michelle Balm, Sally Roberts, Sarah Jefferies, Sharmini Muttaiyah, Susan Morpeth, Susan Taylor, Timothy Blackmore, Vani Sathyendran, Veronica Playle, Virginia Hope, Erasmus Smit, Lauren Jelly, Olin Silander, Joep de Ligt |
| EPI_ISL_548064 | Waikato Hospital | Institute of Environmental Science and Research (ESR) | Xiaoyun Ren, Matt Storey, Nikki Freed, Muhammad Faisal, Jing Wang, Hermes Perez, Anja Werno, Antje van der Linden, Arlo Upton, Chris Mansell, David Hammer, Dragana Drinkovic, Gary McAuliffe, Hana Sofia Andersson, James Ussher, Jill Sherwood, Josh Freeman, Julia Howard, Juliet Elvy, Mary DeAlmeida, Matt Blakiston, Matthew Rogers, Max Bloomfield, Michael Addidle, Michelle Balm, Sally Roberts, Sarah Jefferies, Sharmini Muttaiyah, Susan Morpeth, Susan Taylor, Timothy Blackmore, Vani Sathyendran, Veronica Playle, Virginia Hope, Erasmus Smit, Lauren Jelly, Olin Silander, Joep de Ligt |
| EPI_ISL_548065 | North Shore Hospital | Institute of Environmental Science and Research (ESR) | Xiaoyun Ren, Matt Storey, Nikki Freed, Muhammad Faisal, Jing Wang, Hermes Perez, Anja Werno, Antje van der Linden, Arlo Upton, Chris Mansell, David Hammer, Dragana Drinkovic, Gary McAuliffe, Hana Sofia Andersson, James Ussher, Jill Sherwood, Josh Freeman, Julia Howard, Juliet Elvy, Mary DeAlmeida, Matt Blakiston, Matthew Rogers, Max Bloomfield, Michael Addidle, Michelle Balm, Sally Roberts, Sarah Jefferies, Sharmini Muttaiyah, Susan Morpeth, Susan Taylor, Timothy Blackmore, Vani Sathyendran, Veronica Playle, Virginia Hope, Erasmus Smit, Lauren Jelly, Olin Silander, Joep de Ligt |
| EPI_ISL_548105 | LabTests | Institute of Environmental Science and Research (ESR) | Xiaoyun Ren, Matt Storey, Nikki Freed, Muhammad Faisal, Jing Wang, Hermes Perez, Anja Werno, Antje van der Linden, Arlo Upton, Chris Mansell, David Hammer, Dragana Drinkovic, Gary McAuliffe, Hana Sofia Andersson, James Ussher, Jill Sherwood, Josh Freeman, Julia Howard, Juliet Elvy, Mary DeAlmeida, Matt Blakiston, Matthew Rogers, Max Bloomfield, Michael Addidle, Michelle Balm, Sally Roberts, Sarah Jefferies, Sharmini Muttaiyah, Susan Morpeth, Susan Taylor, Timothy Blackmore, Vani Sathyendran, Veronica Playle, Virginia Hope, Erasmus Smit, Lauren Jelly, Olin Silander, Joep de Ligt |
| EPI_ISL_548118 | Canterbury Health Laboratories | Institute of Environmental Science and Research (ESR) | Xiaoyun Ren, Matt Storey, Nikki Freed, Muhammad Faisal, Jing Wang, Hermes Perez, Anja Werno, Antje van der Linden, Arlo Upton, Chris Mansell, David Hammer, Dragana Drinkovic, Gary McAuliffe, Hana Sofia Andersson, James Ussher, Jill Sherwood, Josh Freeman, Julia Howard, Juliet Elvy, Mary DeAlmeida, Matt Blakiston, Matthew Rogers, Max Bloomfield, Michael Addidle, Michelle Balm, Sally Roberts, Sarah Jefferies, Sharmini Muttaiyah, Susan Morpeth, Susan Taylor, Timothy Blackmore, Vani Sathyendran, Veronica Playle, Virginia Hope, Erasmus Smit, Lauren Jelly, Olin Silander, Joep de Ligt |
| EPI_ISL_548130, EPI_ISL_548133 | Middlemore Hospital | Institute of Environmental Science and Research (ESR) | Xiaoyun Ren, Matt Storey, Nikki Freed, Muhammad Faisal, Jing Wang, Hermes Perez, Anja Werno, Antje van der Linden, Arlo Upton, Chris Mansell, David Hammer, Dragana Drinkovic, Gary McAuliffe, Hana Sofia Andersson, James Ussher, Jill Sherwood, Josh Freeman, Julia Howard, Juliet Elvy, Mary DeAlmeida, Matt Blakiston, Matthew Rogers, Max Bloomfield, Michael Addidle, Michelle Balm, Sally Roberts, Sarah Jefferies, Sharmini Muttaiyah, Susan Morpeth, Susan Taylor, Timothy Blackmore, Vani Sathyendran, Veronica Playle, Virginia Hope, Erasmus Smit, Lauren Jelly, Olin Silander, Joep de Ligt |
| EPI_ISL_548140 | Canterbury Health Laboratories | Institute of Environmental Science and Research (ESR) | Xiaoyun Ren, Matt Storey, Nikki Freed, Muhammad Faisal, Jing Wang, Hermes Perez, Anja Werno, Antje van der Linden, Arlo Upton, Chris Mansell, David Hammer, Dragana Drinkovic, Gary McAuliffe, Hana Sofia Andersson, James Ussher, Jill Sherwood, Josh Freeman, Julia Howard, Juliet Elvy, Mary DeAlmeida, Matt Blakiston, Matthew Rogers, Max Bloomfield, Michael Addidle, Michelle Balm, Sally Roberts, Sarah Jefferies, Sharmini Muttaiyah, Susan Morpeth, Susan Taylor, Timothy Blackmore, Vani Sathyendran, Veronica Playle, Virginia Hope, Erasmus Smit, Lauren Jelly, Olin Silander, Joep de Ligt |
| EPI_ISL_548220 | Laboratoire de Virologie, HUG | Swiss National Reference Centre for Influenza | LAUBSCHER F. |
| EPI_ISL_550381 | Lighthouse Lab in Milton Keynes | Wellcome Sanger Institute for the COVID-19 Genomics UK (COG-UK) consortium | The Lighthouse Lab in Milton Keynes and Alex Alderton, Roberto Amato, Sonia Goncalves, Ewan Harrison, David K. Jackson, Ian Johnston, Dominic Kwiatkowski, Cordelia Langford, John Sillitoe on behalf of the Wellcome Sanger Institute COVID-19 Surveillance Team (http://www.sanger.ac.uk/covid-team) |
| EPI_ISL_560385 | National Health Laboratory | Botswana Institute for Technology Research and Innovation | Kefentse Arnold Tumedi, Madisa Mine, Dineo Emang Tshiamo, Gape Nyepetsi, Thongbotho Mphoyakgosi, Maitshwarelo Ignatius Matsheka |

| | | | |
|---|---|---|---|
| EPI_ISL_560386 | National Health Laboratory | Botswana Institute for Technology Research and innovation | Kefentse Arnold Tumedi, Madisa Mine, Dineo Emang Tshiamo. Gape Nyepetsi, Thongbotho Mphoyakgosi, Maitshwarelo Ignatius Matsheka |
| EPI_ISL_560390 | National Health Laboratory | Botswana Institute for Technology Research and Innovation | Kefentse Arnold Tumedi, Madisa Mine, Dineo Emang Tshiamo. Gape Nyepetsi, Thongbotho Mphoyakgosi, Maitshwarelo Ignatius Matsheka |
| EPI_ISL_560405 | Vilnius University Hospital Santaros Klinikos, Vilnius University | Institute of Biotechnology, Life Sciences Center, Vilnius University and Thermo Fisher Scientific | Justinas Slikas, Albertas Timinskas, Alma Gedvilaite, Aurelija Zvirbliene, Daniel Naumovas, Laimonas Griskevicius, Ligita Jancioriene, Mindaugas Paulauskas |
| EPI_ISL_561000, EPI_ISL_561008, EPI_ISL_561014, EPI_ISL_561034, EPI_ISL_561038, EPI_ISL_561086 | MRCG at LSHTM Genomics lab | MRCG at LSHTM Genomics lab | Abdul Karim sesay, Abdoulie Kanteh, Jarra Manneh, Mariama Kujabi, Bakary Sanyang |
| EPI_ISL_562120 | Microbiological Diagnostic Unit - Public Health Laboratory (MDU-PHL) | MDU-PHL | Seemann, T., Schultz M. B., Sait, M., Sherry, N. |
| EPI_ISL_564370 | Victorian Infectious Diseases Reference Laboratory (VIDRL) | VIDRL and MDU-PHL | Caly, L., Seemann, T., Sait, M., Schultz, M. B., Druce J., Sherry, N. |
| EPI_ISL_564759, EPI_ISL_565124, EPI_ISL_565610 | Microbiological Diagnostic Unit - Public Health Laboratory (MDU-PHL) | MDU-PHL | Seemann, T., Schultz M. B., Sait, M., Sherry, N. |
| EPI_ISL_568900, EPI_ISL_568969, EPI_ISL_569427 | MEPHI, Aix Marseille University | MEPHI, Aix Marseille University | Anthony LEVASSEUR |
| EPI_ISL_569769, EPI_ISL_569794, EPI_ISL_569803 | Omsk Research Institute of Natural Focal Infections | WHO National Influenza Centre Russian Federation | Artem Fadeev, Ekaterina Gradoboeva, Ekaterina Savkina, Daria Nashatyreva, Elena Poleshchuk, Aleksei Vasilenko, Valery Yakimenko, Andrey Komissarov |
| EPI_ISL_571746 | Quest Diagnostics | Quest Diagnostics | Rosenthal,S.H., Gerasimova,A., Kagan,R.M., Anderson, B., Grover, D., Livingston, K.E., Hua, M., Liu Y., Shalhout, D.F., Owen, R., Lacbawan, F. |
| EPI_ISL_574519 | National Public Health Laboratory, National Centre for Infectious Diseases | National Public Health Laboratory, National Centre for Infectious Diseases | Tze Minn Mak, Sophie Octavia, Zhenyang Zhou, Lin Cui, Raymond Tzer Pin Lin |
| EPI_ISL_575213 | Utah Public Health Laboratory | Utah Public Health Laboratory | Erin Young, Kelly Oakeson |
| EPI_ISL_575332 | Israel Central Virology laboratory | Israel Central Virology laboratory | Neta Zuckerman, Efrat Dahan Bucris, Oran Erster, Ella Mendelson, Michal Mandelboim |
| EPI_ISL_576117 | Laboratory, The Bio Arte Limited | Laboratory, The Bio Arte Limited | Biazzo,M., Madeddu,S., Santoro,F., Pinzauti,D. |
| EPI_ISL_576131, EPI_ISL_576136, EPI_ISL_576137 | Public Health Virology Laboratory, Forensic and Scientific Services (PHV-FSS) | Public Health Virology Laboratory, Forensic and Scientific Services (PHV-FSS) | Son Nguyen et al |
| EPI_ISL_576178 | DC Public Health Lab/ Dept. of Forensic Sciences | Pathogen Discovery, Respiratory Viruses Branch, Division of Viral Diseases, Centers for Disease Control and Prevention | Ying Tao, Jing Zhang, Brian Lynch, Yan Li, Krista Queen, Anna Uehara, Clinton R. Paden, Peter Cook, Haibin Wang, Suxiang Tong |
| EPI_ISL_576233 | Minnesota Department of Health, Public Health Laboratory | Minnesota Department of Health, Public Health Laboratory | Matt Plumb, Jacob Garfin, Alexandra Lorentz, and Xiong Wang |
| EPI_ISL_576265 | Instituto de Diagnostico y Referencia Epidemiologicos (INDRE) | Instituto de Diagnostico y Referencia Epidemiologicos (INDRE) | Ernesto Ramirez-Gonzalez, Abril Rodriguez-Maldonado, Claudia Wong-Arambula , Natividad Cruz-Ortiz, Tatiana Nunez-Garcia, Dayanira Arellano-Suarez, Adnan Araiza-Rodriguez, Edgar Mendieta-Condado, Lucia Hernandez-Rivas, Irma Lopez-Martinez, Gisela Barrera-Badillo. |
| EPI_ISL_577607 | Royal Hobart Hospital | Royal Hobart Hospital | Cooley L., van Haeften R. |
| EPI_ISL_577739 | Institute of Virology, Biomedical Research Center of the Slovak Academy of Sciences, Bratislava | Faculty of Natural Sciences, Comenius University, Bratislava | Kristína Boršová, Viktória Hodorová, Broa Brejová, Viktória abanová, Dominika Friová, Sabina Fumaová Havlíková, Juraj Kopáek, Martina Liková, ubomíra Lukáiková, Martina Neboháová, Monika Sláviková, Edita Staroová, Elena Tichá, Tomáš Vina, Boris Klempa, Jozef Nosek |
| EPI_ISL_579069, EPI_ISL_579072, EPI_ISL_579088, EPI_ISL_579091 | Canterbury Health Laboratories | Institute of Environmental Science and Research (ESR) | Xiaoyun Ren, Matt Storey, Nikki Freed, Muhammad Faisal, Jing Wang, Hermes Perez, Anja Werno, Antje van der Linden, Arlo Upton, Chris Mansell, David Hammer, Dragana Drinkovic, Gary McAuliffe, Hana Sofia Andersson, James Ussher, Jill Sherwood, Josh Freeman, Julia Howard, Juliet Elvy, Mary DeAlmeida, Matt Blakiston, Matthew Rogers, Max Bloomfield, Michael Addidle, Michelle Balm, Sally Roberts, Sarah Jefferies, Sharmini Muttaiyah, Susan Morpeth, Susan Taylor, Timothy Blackmore, Vani Sathyendran, Veronica Playle, Virginia Hope, Erasmus Smit, Lauren Jelly, Olin Silander, Joep de Ligt |
| EPI_ISL_579093 | North Shore Hospital | Institute of Environmental Science and Research (ESR) | Xiaoyun Ren, Matt Storey, Nikki Freed, Muhammad Faisal, Jing Wang, Hermes Perez, Anja Werno, Antje van der Linden, Arlo Upton, Chris Mansell, David Hammer, Dragana Drinkovic, Gary McAuliffe, Hana Sofia Andersson, James Ussher, Jill Sherwood, Josh Freeman, Julia Howard, Juliet Elvy, Mary DeAlmeida, Matt Blakiston, Matthew Rogers, Max Bloomfield, Michael Addidle, Michelle Balm, Sally Roberts, Sarah Jefferies, Sharmini Muttaiyah, Susan Morpeth, Susan Taylor, Timothy Blackmore, Vani Sathyendran, Veronica Playle, Virginia Hope, Erasmus Smit, Lauren Jelly, Olin Silander, Joep de Ligt |
| EPI_ISL_579118 | Waikato Hospital | Institute of Environmental Science and Research (ESR) | Xiaoyun Ren, Matt Storey, Nikki Freed, Muhammad Faisal, Jing Wang, Hermes Perez, Anja Werno, Antje van der Linden, Arlo Upton, Chris Mansell, David Hammer, Dragana Drinkovic, Gary McAuliffe, Hana Sofia Andersson, James Ussher, Jill Sherwood, Josh Freeman, Julia Howard, Juliet Elvy, Mary DeAlmeida, Matt Blakiston, Matthew Rogers, Max Bloomfield, Michael Addidle, Michelle Balm, Sally Roberts, Sarah Jefferies, Sharmini Muttaiyah, Susan Morpeth, Susan Taylor, Timothy Blackmore, Vani Sathyendran, Veronica Playle, Virginia Hope, Erasmus Smit, Lauren Jelly, Olin Silander, Joep de Ligt |
| EPI_ISL_579170, EPI_ISL_579172, EPI_ISL_579189 | Southern Community Labs Dunedin | Institute of Environmental Science and Research (ESR) | Xiaoyun Ren, Matt Storey, Nikki Freed, Muhammad Faisal, Jing Wang, Hermes Perez, Anja Werno, Antje van der Linden, Arlo Upton, Chris Mansell, David Hammer, Dragana Drinkovic, Gary McAuliffe, Hana Sofia Andersson, James Ussher, Jill Sherwood, Josh Freeman, Julia Howard, Juliet Elvy, Mary DeAlmeida, Matt Blakiston, Matthew Rogers, Max Bloomfield, Michael Addidle, Michelle Balm, Sally Roberts, Sarah Jefferies, Sharmini Muttaiyah, Susan Morpeth, Susan Taylor, Timothy Blackmore, Vani Sathyendran, Veronica Playle, Virginia Hope, Erasmus Smit, Lauren Jelly, Olin Silander, Joep de Ligt |
| EPI_ISL_579314 | Middlemore Hospital | Institute of Environmental Science and Research (ESR) | Xiaoyun Ren, Matt Storey, Nikki Freed, Muhammad Faisal, Jing Wang, Hermes Perez, Anja Werno, Antje van der Linden, Arlo Upton, Chris Mansell, David Hammer, Dragana Drinkovic, Gary McAuliffe, Hana Sofia Andersson, James Ussher, Jill Sherwood, Josh Freeman, Julia Howard, Juliet Elvy, Mary DeAlmeida, Matt Blakiston, Matthew Rogers, Max Bloomfield, Michael Addidle, Michelle Balm, Sally Roberts, Sarah Jefferies, Sharmini Muttaiyah, Susan Morpeth, Susan Taylor, Timothy Blackmore, Vani Sathyendran, Veronica Playle, Virginia Hope, Erasmus Smit, Lauren Jelly, Olin Silander, Joep de Ligt |
| EPI_ISL_579406, EPI_ISL_579407 | North Shore Hospital | Institute of Environmental Science and Research (ESR) | Xiaoyun Ren, Matt Storey, Nikki Freed, Muhammad Faisal, Jing Wang, Hermes Perez, Anja Werno, Antje van der Linden, Arlo Upton, Chris Mansell, David Hammer, Dragana Drinkovic, Gary McAuliffe, Hana Sofia Andersson, James Ussher, Jill Sherwood, Josh Freeman, Julia Howard, Juliet Elvy, Mary DeAlmeida, Matt Blakiston, Matthew Rogers, Max Bloomfield, Michael Addidle, Michelle Balm, Sally Roberts, Sarah Jefferies, Sharmini Muttaiyah, Susan Morpeth, Susan Taylor, Timothy Blackmore, Vani Sathyendran, Veronica Playle, Virginia Hope, Erasmus Smit, Lauren Jelly, Olin Silander, Joep de Ligt |
| EPI_ISL_579422 | LabTests | Institute of Environmental Science and Research (ESR) | Xiaoyun Ren, Matt Storey, Nikki Freed, Muhammad Faisal, Jing Wang, Hermes Perez, Anja Werno, Antje van der Linden, Arlo Upton, Chris Mansell, David Hammer, Dragana Drinkovic, Gary McAuliffe, Hana Sofia Andersson, James Ussher, Jill Sherwood, Josh Freeman, Julia Howard, Juliet Elvy, Mary DeAlmeida, Matt Blakiston, Matthew Rogers, Max Bloomfield, Michael Addidle, Michelle Balm, Sally Roberts, Sarah Jefferies, Sharmini Muttaiyah, Susan Morpeth, Susan Taylor, Timothy Blackmore, Vani Sathyendran, Veronica Playle, Virginia Hope, Erasmus Smit, Lauren Jelly, Olin Silander, Joep de Ligt |
| EPI_ISL_579454, EPI_ISL_579474 | Canterbury Health Laboratories | Institute of Environmental Science and Research (ESR) | Xiaoyun Ren, Matt Storey, Nikki Freed, Muhammad Faisal, Jing Wang, Hermes Perez, Anja Werno, Antje van der Linden, Arlo Upton, Chris Mansell, David Hammer, Dragana Drinkovic, Gary McAuliffe, Hana Sofia Andersson, James Ussher, Jill Sherwood, Josh Freeman, Julia Howard, Juliet Elvy, Mary DeAlmeida, Matt Blakiston, Matthew Rogers, Max Bloomfield, Michael Addidle, Michelle Balm, Sally Roberts, Sarah Jefferies, Sharmini Muttaiyah, Susan Morpeth, Susan Taylor, Timothy Blackmore, Vani Sathyendran, Veronica Playle, Virginia Hope, Erasmus Smit, Lauren Jelly, Olin Silander, Joep de Ligt |
| EPI_ISL_582018, EPI_ISL_582019, EPI_ISL_582021 | PathLab Bay of Plenty | Institute of Environmental Science and Research (ESR) | Xiaoyun Ren, Matt Storey, Nikki Freed, Muhammad Faisal, Jing Wang, Hermes Perez, Anja Werno, Antje van der Linden, Arlo Upton, Chris Mansell, David Hammer, Dragana Drinkovic, Gary McAuliffe, Hana Sofia Andersson, James Ussher, Jill Sherwood, Josh Freeman, Julia Howard, Juliet Elvy, Mary DeAlmeida, Matt Blakiston, Matthew Rogers, Max Bloomfield, Michael Addidle, Michelle Balm, Sally Roberts, Sarah Jefferies, Sharmini Muttaiyah, Susan Morpeth, Susan Taylor, Timothy Blackmore, Vani Sathyendran, Veronica Playle, Virginia Hope, Erasmus Smit, Lauren Jelly, Olin Silander, Joep de Ligt |
| EPI_ISL_582032 | Institute of Human Genetics, Polish Academy of Sciences | Institute of Human Genetics, Polish Academy of Sciences | Szymon Hryhorowicz, Adam Ustaszewski, Marta Kaczmarek-Ry, Emilia Lis, Ewa Zitkiewicz, Micha Witt, Andrzej Pawski |
| EPI_ISL_582033 | Department of Pathology, School of Medicine, Imam Khomeini Hospital, Tehran University of Medical Sciences | Genetics Research Center. University Of Social Welfare And Rehabilitation Sciences | Zohreh Fattahi, Marzieh Mohseni, Khadijeh Jalalvand, Azam Ghaziasadi, Seyedeh elham Mortazavi, Ali Jafarpour, Azar Hadadi, Alireza Abdollahi, Ali Jafarpour, Azam Ghaziasad, Seyedeh elham Mortazavi, Saber Soltani, Reza Najafipour , Kimia Kahrizi, Seyed Mohammad Jazayeri, Hossein Najmabadi |
| EPI_ISL_582126 | Sheikh Khalifa Medical City | Molecular Surveillance lab Sheikh Khalifa Medical City | Amirtharaj Francis, Sajeed Abdul, Hala Imambaccus, Sahar Almarzooqi, Hiba Saud, Stefan Weber |

| | | | |
|---|---|---|---|
| EPI_ISL_582302, EPI_ISL_582450 | Cadham Provincial Laboratory | National Microbiology Laboratory (NML) | Anna Majer, Shari Tyson, Grace Seo, Philip Mabon, Elsie Grudeski, Rhiannon Huzarewich, Russell Mandes, Anneliese Landgraff, Jennifer Tanner, Natalie Knox, Morag Graham, Gary Van Domselaar, Paul Van Caeseele, Jared Bullard, David Alexander, Kerry Dust, Nathalie Bastien, Yan Li, Timothy Booth, Darian Hole, Madison Chapel, CanCOGeN's metadata curation team, Public Health Agency of Canada CanCOGeN team |
| EPI_ISL_582610, EPI_ISL_582621, EPI_ISL_582625, EPI_ISL_582627, EPI_ISL_582656 | Sheikh Khalifa Medical City | Molecular/Surveillance lab Sheikh Khalifa Medical City | Amirtharaj Francis, Sajeed Abdul, Hala Imambaccus, Sahar Almarzooqi, Hiba Saud, Stefan Weber |
| EPI_ISL_583623 | Pathologie-Labor Dr. Obrist- Dr. Brunhuber | Bergthaler laboratory, CeMM Research Center for Molecular Medicine of the Austrian Academy of Sciences | Alexandra Popa, Benedikt Agerer, Henrique Colaco, Lukas Endler, Jakob-Wendelin Genger, Alexander Lercher, Mark Smyth, Thomas Penz, Michael Schuster, Jan Laine, Martin Senekowitsch, Judith Aberle, Stephan Aberle, Peter Hufnagl, Daniela Schmid, Franz Allerberger, Elisabeth Puchhammer-Stoeckl, Manfred Nairz, Guenter Weiss, Gregor Hörmann, Kinga Rigler-Hohenwarter, Rainer Gattringer, Wegene Borena, Dorothee von Laer, Gernot Walder, Peter Obrist, Christian Paar, Sabine Sussitz-Rack, Gunther Vogl, Adi Steinrigl, Christoph Bock, Andreas Bergthaler |
| EPI_ISL_583884 | Austrian Agency for Health and Food Safety (AGES) | Bergthaler laboratory, CeMM Research Center for Molecular Medicine of the Austrian Academy of Sciences | Alexandra Popa, Benedikt Agerer, Henrique Colaco, Lukas Endler, Jakob-Wendelin Genger, Alexander Lercher, Mark Smyth, Thomas Penz, Michael Schuster, Jan Laine, Martin Senekowitsch, Judith Aberle, Stephan Aberle, Peter Hufnagl, Daniela Schmid, Franz Allerberger, Elisabeth Puchhammer-Stoeckl, Manfred Nairz, Guenter Weiss, Gregor Hörmann, Kinga Rigler-Hohenwarter, Rainer Gattringer, Wegene Borena, Dorothee von Laer, Gernot Walder, Peter Obrist, Christian Paar, Sabine Sussitz-Rack, Gunther Vogl, Adi Steinrigl, Christoph Bock, Andreas Bergthaler |
| EPI_ISL_584075 | The National Institute of Public Health | State Veterinary Institute Prague | Nagy,A;Jirincova,H;Novakova,L;Trnka,D;Vecerova,J |
| EPI_ISL_584621 | Liverpool Clinical Laboratories | COVID-19 Genomics UK (COG-UK) Consortium | Sam Haldenby, Anita Lucaci, Steve Paterson, Julian Hiscox, Alistair Darby, M Almsaud, A Alrezaihi, Muhannad Alruwaili, Stuart D Armstrong, Jones Benjamin, Eleanor G Bentley, Anu Chawla, Jordan J Clark, Angela Cowell, Richard Eccles, Isabel García-Dorival, Matthew Gemmell, Alessandro Gerada, PKF Gilmore, Richard Gregory, Ximeng Han, Catherine Hartley, Margaret Hughes, Miren Iturriza-Gomara, James Johnson, L Luu, Jenifer Manson, Charlotte Nelson, Elaine O'Toole, Cassie Olateju, Rebekah Penrice-Randal , Lucille Rainbow, N.P Randle, Trevor Ian Robinson, Parul Sharma, Ghada T Shawli, James P Stewart, Neil Swainston, Ecaterina Vamos, Joanne Watts, Mark Whitehead |
| EPI_ISL_590820 | Minnesota Department of Health, Public Health Laboratory | Minnesota Department of Health, Public Health Laboratory | Matt Plumb, Jacob Garfin, Alexandra Lorentz, and Xiong Wang |
| EPI_ISL_590886 | Vestfold Hospital, Toensberg Department of Microbiology | Norwegian Institute of Public Health, Department of Virology | Kathrine Stene-Johansen, Kamilla Heddeland Instefjord, Hilde Elshaug, Rasmus Riis Kopperud, Hilde Vollan, Karoline Bragstad, Olav Hungnes |
| EPI_ISL_591083 | Viral Respiratory Lab, National Institute for Biomedical Research (INRB) | Pathogen Sequencing Lab, National Institute for Biomedical Research (INRB) | Placide Mbala-Kingebeni, Edith Nkwembe, Eddy Kinganda-Lusamaki, Amuri Aziza, Francisca Muyembe Mawete, Emmanuel Lokilo Lofiko, Jean Claude Makangara, Catherine Pratt, Matthias Pauthner, Josh Quick, Allison Black, James Hadfield, Trevor Bedford, Ian Goodfellow, Andrew Rambaut, Nick Loman, Kristian Andersen, Michael Wiley, Steve Ahuka-Mundeke, Jean-Jacques Muyembe Tamfum |
| EPI_ISL_591280 | National Institute for Viral Disease Control and Prevention, China CDC | National Institute for Viral Disease Control and Prevention, China CDC | Huilai Ma, Zhaoguo Wang, Xiang Zhao, Jun Han, Yong Zhang, Hong Wang, Cao Chen, Ji Wang, Jingdong Song, Yao Meng, Yuchao Wu, Zhixiao Chen, Dayan Wang, Ruqin Gao, George F.Gao, Wenbo Xu |
| EPI_ISL_591489 | Laverty Pathology | NSW Health Pathology - Institute of Clinical Pathology and Medical Research; Westmead Hospital; University of Sydney | CIDM-PH et al. |
| EPI_ISL_593001, EPI_ISL_593410 | Microbiological Diagnostic Unit - Public Health Laboratory (MDU-PHL) | MDU-PHL | Seemann T., Schultz, M. B., Sait, M., Sherry, N. |
| EPI_ISL_593622, EPI_ISL_593642 | Public Health Virology Laboratory, Forensic and Scientific Services (PHV-FSS) | Public Health Virology Laboratory, Forensic and Scientific Services (PHV-FSS) | Son Nguyen et al |
| EPI_ISL_593656 | Pathology North - Hunter - NSW Health Pathology | NSW Health Pathology - Institute of Clinical Pathology and Medical Research; Westmead Hospital; University of Sydney | CIDM-PH et al. |
| EPI_ISL_593660 | Pathology West - NSW Health Pathology | NSW Health Pathology - Institute of Clinical Pathology and Medical Research; Westmead Hospital; University of Sydney | CIDM-PH et al. |
| EPI_ISL_593685, EPI_ISL_593692 | South Eastern Area Laboratory Services (SEALS) | NSW Health Pathology - Institute of Clinical Pathology and Medical Research; Westmead Hospital; University of Sydney | CIDM-PH et al. |
| EPI_ISL_594133 | Yale COVID-19 Biorepository | Grubaugh Lab - Yale School of Public Health | Joseph Fauver, Tara Alpert, Anderson Brito, Anne Wyllie, Chantal Vogels, Mary Petrone, Chaney Kalinich, Isabel Ott, Arnau Casanovas, Catherine Muenker, Adam Moore, Alice Lu, Maria Tokuyama, Patrick Wong, Peiwen Lu, Saad Omer, Richard Martinello, Allison Nelson, Shelli Farhadian, Akiko Iwasaki, Charlese Dela Cruz, Albert Ko, Nathan Grubaugh |
| EPI_ISL_594185, EPI_ISL_594188, EPI_ISL_596451 | Department of Pathology, School of Medicine, Imam Khomeini Hospital, Tehran University of Medical Sciences | Genetics Research Center, University of Social Welfare and Rehabilitation Sciences | Zohreh Fattahi, Marzieh Mohseni, Khadijeh Jalalvand, Azam Ghaziasadi, Seyedeh elham Mortazavi, Ali Jafarpour, Azar Hadadi, Alireza Abdollahi, Ali Jafarpour, Azam Ghaziasad, Seyedeh elham Mortazavi, Saber Soltani, Reza Najafipour, Kimia Kahrizi, Seyed Mohammad Jazayeri, Hossein Najmabadi |
| EPI_ISL_596457 | National Public Health Laboratory, National Centre for Infectious Diseases | National Public Health Laboratory, National Centre for Infectious Diseases | Tze Minn Mak, Sophie Octavia, Zhenyang Zhou, Lin Cui, Raymond Tzer Pin Lin |
| EPI_ISL_596504, EPI_ISL_596517, EPI_ISL_596534, EPI_ISL_596558 | Palestinian Ministry of Health | Molecular Genetics Lab | Nouar Qutob, Zaidoun Salah, Damien Richard, Hisham Darwish, Husam Sallam, Issa Shtayeh, Osama Najjar, Mahmoud Ruzayqat, Dana Najjar, Francois Balloux, Lucy van Dorp |
| EPI_ISL_596703, EPI_ISL_596743, EPI_ISL_596752, EPI_ISL_596866 | PathWest Laboratory Medicine WA | PathWest Laboratory Medicine WA Microbial Surveillance Unit | PathWest Laboratory Medicine WA Microbial Surveillance Unit |
| EPI_ISL_602358 | HELIX LLC | WHO National Influenza Centre Russian Federation | Andrey Komissarov, Artem Fadeev, Kseniya Komissarova, Anna Ivanova, Dmitry Bazhenov, Daria Danilenko |
| EPI_ISL_603249 | National Institute of Laboratory Medicine and Referral Center | Genomic Research Lab, BCSIR | Md. Ahashan Habib, Abu Sayeed Mohammad Mahmud, Mohammad Samir Uzzaman, Eshrar Osman, Shahina Akter, Tanjina Akhter Banu, Md. Murshed Hasan Sarkar, Barna Goswami, Iffat Jahan, Md. Saddam Hossain, Tasnim Nafisa, Md. Maruf Ahmed Molla, Mahmuda Yeasmin, Asish Kumar Ghosh, A. K. M. Shamsuzzaman, Monira Parveen, Md. Masum Hossain Arif, Md. Salim Khan |
| EPI_ISL_605843 | PathWest Laboratory Medicine WA | PathWest Laboratory Medicine WA Microbial Surveillance Unit | PathWest Laboratory Medicine WA Microbial Surveillance Unit |
| EPI_ISL_605907 | NGS Lab, DNA SOLUTION LTD. | NGS Lab, DNA SOLUTION LTD. | Khan,M.I., Hasan,K.N., Sufian,A., Polol,M.N.I., Khaleque,A., Rahman,M., Chowdhury,M., Haider,H.U., Razu,M.H., Khan,M., Rabbi,M.F.A. |
| EPI_ISL_610194, EPI_ISL_610200 | Department of Health Technology and Informatics, The Hong Kong Polytechnic University | Department of Health Technology and Informatics, The Hong Kong Polytechnic University | Siu,G.K.-H., Lee,L.-K., Leung,K.S.-S., Leung,J.S.-L., Ng,T.T.-L., Chan,C.T.-M., Tam,K.K.-G., Lao,H.-Y., Wu,A.K.-L., Yau,M.C.-Y., Lai,Y.W.-M., Fung,K.S.-C., Chau,S.K.-Y., Wong,B.K.-C., To,W.-K., Luk,K., Ho,A.Y.-M., Que,T.-L., Yip,K.-T., Yam,W.C., Shum,D.H.-K., Yip,S.P. |
| EPI_ISL_613425, EPI_ISL_613431 | Institut Pasteur de la Guadeloupe | Institut Pasteur de la Guadeloupe | Marion Barbet, Sylvie Behillil, Méline Bizard, Angela Brisebarre, Camille Capel, Etienne Simon-Lorière, Vincent Enouf, Maud Vanpeene, Sylvie van der Werf, Stéphanie Guyomard, Sébastien Breurec, Antoine Talarmin |
| EPI_ISL_613448 | Institut Pasteur de la Guadeloupe | Institut Pasteur de la Guadeloupe | Marion Barbet, Sylvie Behillil, Méline Bizard, Angela Brisebarre, Camille Capel, Etienne Simon-Lorière, Vincent Enouf, Maud Vanpeene, Sylvie van der Werf, Stéphanie Guyomard, Sébastien Breurec, Radjin Steingrover, Cherina Fleming, Antoine Talarmin |
| EPI_ISL_614357, EPI_ISL_614389, EPI_ISL_614392 | Molecular diagnostic unit for viral haemorrhagic fevers and emerging viruses, Bouaké CHU Laboratory | Project group Epidemology of Highly Pathogenic Microorganisms, Robert Koch-Institute | Chantal Akoua-Koffi, Diané Bamourou, Etilé Anoh, Essia Belarbi, Safiatou Karidioula, Grit Schubert, Adjaratou Traoré, Soundélé Maïté, Monemo Pacome, Coulibaly Mbegnan, Bamba Fatoumata Touré, Kra Ouffoué, Fabian Leendertz |
| EPI_ISL_618115, EPI_ISL_618660, EPI_ISL_622747 | Department of Virus and Microbiological Special Diagnostics, Statens Serum Institut, Denmark | Albertsen lab, Department of Chemistry and Bioscience, Aalborg University, Denmark | Danish Covid-19 Genome Consortia |
| EPI_ISL_622795, EPI_ISL_622796, EPI_ISL_622797, EPI_ISL_622799 | Canterbury Health Laboratories | Institute of Environmental Science and Research (ESR) | Xiaoyun Ren, Matt Storey, Nikki Freed, Muhammad Faisal, Jing Wang, Hermes Perez, Anja Werno, Antje van der Linden, Arlo Upton, Chris Mansell, David Hammer, Dragana Drinkovic, Gary McAuliffe, Hana Sofia Andersson, James Ussher, Jill Sherwood, Josh Freeman, Julia Howard, Juliet Elvy, Mary DeAlmeida, Matt Blakiston, Matthew Rogers, Max Bloomfield, Michael Addidle, Michelle Balm, Sally Roberts, Sarah Jefferies, Sharmini Muttaiyah, Susan Morpeth, Susan Taylor, Timothy Blackmore, Vani Sathyendran, Veronica Playle, Virginia Hope, Erasmus Smit, Lauren Jelly, Olin Silander, Joep de Ligt |
| EPI_ISL_622806 | PathLab Bay of Plenty | Institute of Environmental Science and Research (ESR) | Xiaoyun Ren, Matt Storey, Nikki Freed, Muhammad Faisal, Jing Wang, Hermes Perez, Anja Werno, Antje van der Linden, Arlo Upton, Chris Mansell, David Hammer, Dragana Drinkovic, Gary McAuliffe, Hana Sofia Andersson, James Ussher, Jill Sherwood, Josh Freeman, Julia Howard, Juliet Elvy, Mary |

| | | | |
|---|---|---|---|
| | | | DeAlmeida, Matt Blakiston, Matthew Rogers, Max Bloomfield, Michael Addidle, Michelle Balm, Sally Roberts, Sarah Jefferies, Sharmini Muttaiyah, Susan Morpeth, Susan Taylor, Timothy Blackmore, Vani Sathyendran, Veronica Playle, Virginia Hope, Erasmus Smit, Lauren Jelly, Olin Silander, Joep de Ligt |
| EPI_ISL_622810, EPI_ISL_622812 | Canterbury Health Laboratories | Institute of Environmental Science and Research (ESR) | Xiaoyun Ren, Matt Storey, Nikki Freed, Muhammad Faisal, Jing Wang, Hermes Perez, Anja Werno, Antje van der Linden, Arlo Upton, Chris Mansell, David Hammer, Dragana Drinkovic, Gary McAuliffe, Hana Sofia Andersson, James Ussher, Jill Sherwood, Josh Freeman, Julia Howard, Juliet Elvy, Mary DeAlmeida, Matt Blakiston, Matthew Rogers, Max Bloomfield, Michael Addidle, Michelle Balm, Sally Roberts, Sarah Jefferies, Sharmini Muttaiyah, Susan Morpeth, Susan Taylor, Timothy Blackmore, Vani Sathyendran, Veronica Playle, Virginia Hope, Erasmus Smit, Lauren Jelly, Olin Silander, Joep de Ligt |
| EPI_ISL_625456 | Virology Unit, Institut Pasteur de Madagascar | Virology Unit, Institut Pasteur de Madagascar | Christian Ranaivoson, Cara Brook, Norosoa Razanajatovo, Vida Ahyong, Tsiry Randriambolamanantsoa, Michelle Tan, Vololoniaina Raharinosy, Helisoa Razafimanjato, Cristina M. Tato, Joseph L. DeRisi, Soa Fy Andriamandimby, Jean-Michel Heraud, Philippe Dussart |
| EPI_ISL_625475 | Child Health Research Foundation | Child Health Research Foundation | Senjuti Saha, Md Saiful Islam Sajib, Nikkon Sarkar, Syed Muktadir Al Sium, Afroza Akter Tanni, Roly Malaker, Arif Mohammad Tanmoy, Md Hafizur Rahman, Samir K Saha |
| EPI_ISL_625683 | National Reference Laboratory for COVID-19, Pasteur Institute of Iran | National Reference Laboratory for COVID-19, Pasteur Institute of Iran | Zahra Ahmadi, Zahra Fereydouni, Tahmineh Jalali, Mohammad Hassan Pouriayevali, Mahsa Tavakkoli, Marzieh Sajjadi, Setareh Kashanian, Sanam Azad-Manjiri, Heasam Nemati, Tahereh Mohammadi, Kayhan Azadmanesh, Zabihollah Shoja, Sana Eybpoosh, Ahmad Ghasemi, Parastoo Yekta, Sepideh Gerdooei, Farideh Niknam, Mostafa Salehi-Vaziri |
| EPI_ISL_626352 | LabPLUS | Institute of Environmental Science and Research (ESR) | Xiaoyun Ren, Matt Storey, Nikki Freed, Muhammad Faisal, Jing Wang, Hermes Perez, Anja Werno, Antje van der Linden, Arlo Upton, Chris Mansell, David Hammer, Dragana Drinkovic, Gary McAuliffe, Hana Sofia Andersson, James Ussher, Jill Sherwood, Josh Freeman, Julia Howard, Juliet Elvy, Mary DeAlmeida, Matt Blakiston, Matthew Rogers, Max Bloomfield, Michael Addidle, Michelle Balm, Sally Roberts, Sarah Jefferies, Sharmini Muttaiyah, Susan Morpeth, Susan Taylor, Timothy Blackmore, Vani Sathyendran, Veronica Playle, Virginia Hope, Erasmus Smit, Lauren Jelly, Olin Silander, Joep de Ligt |
| EPI_ISL_626563 | Laboratorio de Biología Molecular, Facultad de Medicina, Universidad de Atacama, Copiapo, Chile/ FONDAP CRG, Universidad Andrés Bello, Santiago, Chile | Center for Mathematical Modeling and Center for Genome Regulation. Santiago, Chile | Echeverría C, Manríquez R, Bastias M, Sanhueza D, Travisany D, Allende ML, Maass A, González M, Montecino, M, Orellana A, Castro E, Meneses C. |
| EPI_ISL_626633 | National Public Health Laboratory, National Centre for Infectious Diseases | National Public Health Laboratory, National Centre for Infectious Diseases | Tze Minn Mak, Sophie Octavia, Zhenyang Zhou, Lin Cui, Raymond Tzer Pin Lin |
| EPI_ISL_632261, EPI_ISL_632263, EPI_ISL_632285 | Communicable Disease Laboratory, Public Health Directorate | Communicable Disease Laboratory, Public Health Directorate | AlWasti,H., AlTaif,Z., AlHujairi,Z., AlAbbas,Z. |
| EPI_ISL_632908 | Genomic Sciences, Rehman Medical Institute | Genomic Sciences, Rehman Medical Institute | Ali,J., Afridi,U.K., Haider,S.A., Sabiha,B., Jan,H. and Jehanzeb,V. |
| EPI_ISL_632934 | Department of Acute Infectious Diseases Control and Prevention,Yunnan Provincial Center for Disease Control and Prevention | Department of Acute Infectious Diseases Control and Prevention,Yunnan Provincial Center for Disease Control and Prevention | Meiling Zhang,Jienan Zhou,Senquan Jia,Xiaonan Zhao,Xiaoqing Fu |
| EPI_ISL_635038 | National Health Laboratory Service - Inkosi Albert Luthuli Central Hospital (NHLS-IALCH) | KRISP, KZN Research Innovation and Sequencing Platform | Giandhari J, Pillay S, Lessells R, Mdlalose K, York D, Khan S, Tegally H, Wilkinson E, de Oliveira T |
| EPI_ISL_635061 | 1-Laboratory of Microbiology, National Reference Lab, Charles Nicolle Hospital; 2-University of Tunis ElManar, Faculty of Medicine of Tunis, LR99ES09, Tunis, Tunisia | 1-Clinical and Experimental Pharmacology Lab, LR16SP02, National Center of Pharmacovigilance, University of Tunis El Manar, Tunis, Tunisia. 2-Neurodegenerative diseases and psychiatric troubles, LR18SP03, Razi Hospital, University of Tunis El Manar, Tunis, Tunisia. 3- Ministry of Health, National Observatory of New and Emerging Diseases, 1006, Tunis, Tunisia | Ilhem Boutiba-Ben Boubaker, Sameh Trabelsi, Nissaf Ben Alaya, Maher Kharrat, Alia Ben Kahla, Jalila Ben Khelil, Salma Abid, Sana Ferjani, Mouna Ben Sassi, Mouna Safer, Imen Mkada, Imen Kacem, Gaies Emna, Soumaya Rammeh, Riadh Daghfous, Riadh Gouider. |
| EPI_ISL_635284, EPI_ISL_635299 | Institute of Microbiology and Immunology, Faculty of Medicine, University of Ljubljana | Institute of Microbiology and Immunology, Faculty of Medicine, University of Ljubljana | Tomaž Mark Zorec, Samo Zakotnik, Miša Korva, Tatjana Avši - Županc, Mario Poljak |
| EPI_ISL_636514, EPI_ISL_636516, EPI_ISL_636520 | Dutch COVID-19 response team | National Institute for Public Health and the Environment (RIVM) | Adam Meijer, Harry Vennema, Jeroen Cremer, Sharon van den Brink, Bas van der Veer, AnneMarie van den Brandt, Florian Zwagemaker, Dennis Schmitz, Chantal Reusken, on behalf of the national COVID-19 response team |
| EPI_ISL_636889 | Lithuanian University of Health Sciences Hospital, Department of Laboratory Medicine | Lithuanian University of Health Sciences, Molecular cardiology lab. | Lukas Zemaitis, Ingrida Olendrait, Arnoldas Pautienius, Kamile Tamusauskaite, Dovydas Gecys, Laura Pareckaite, Vaiva Lesauskaite, Astra Vitkauskiene |
| EPI_ISL_636966 | Pathogen Genomics Lab King Abdullah University of Science and Technology(KAUST) | Pathogen Genomics Lab King Abdullah University of Science and Technology(KAUST) | Fathia Ben Rached, Raeece Naeem, Sharif Hala, Fadwa Alofi, Rahul P Salunke, Sara Mfarrej, Amit Kumar Subudhi, Afrah Alsomali, Asim Khogeer, Ahmad Bakur Mahmoud, Anwar Hashem, Naif Almontashiri, Arnab Pain |
| EPI_ISL_637088 | PathLab Bay of Plenty | Institute of Environmental Science and Research (ESR) | Xiaoyun Ren, Matt Storey, Nikki Freed, Muhammad Faisal, Jing Wang, Hermes Perez, Anja Werno, Antje van der Linden, Arlo Upton, Chris Mansell, David Hammer, Dragana Drinkovic, Gary McAuliffe, Hana Sofia Andersson, James Ussher, Jill Sherwood, Josh Freeman, Julia Howard, Juliet Elvy, Mary DeAlmeida, Matt Blakiston, Matthew Rogers, Max Bloomfield, Michael Addidle, Michelle Balm, Sally Roberts, Sarah Jefferies, Sharmini Muttaiyah, Susan Morpeth, Susan Taylor, Timothy Blackmore, Vani Sathyendran, Veronica Playle, Virginia Hope, Erasmus Smit, Lauren Jelly, Olin Silander, Joep de Ligt |
| EPI_ISL_639758 | Public Health Virology Laboratory, Forensic and Scientific Services (PHV-FSS) | Public Health Virology Laboratory, Forensic and Scientific Services (PHV-FSS) | Son Nguyen et al |
| EPI_ISL_639927 | Omsk Research Institute of Natural Focal Infections | WHO National Influenza Centre Russian Federation | Artem Fadeev, Ekaterina Gradoboeva, Ekaterina Savkina, Daria Nashatyreva, Elena Poleshchuk, Aleksei Vasilenko, Valery Yakimenko, Andrey Komissarov |
| EPI_ISL_641050 | Microbiological Diagnostic Unit - Public Health Laboratory (MDU-PHL) | MDU-PHL | Seemann T., Schultz M.B., Sait, M.L., Sherry, N.L. |
| EPI_ISL_645091 | Human Genome Variation Research Group, Malopolska Centre of Biotechnology | Human Genome Variation Research Group, Malopolska Centre of Biotechnology | Kowalski,M., Pospiech,E., Klajmon,A., Gromowski,T., Pisarek,A., Marszalek,K., Kopera,K., Foremny,J., Swadzba,J., Sanak,M., Owczarek,K., Dabrowska,A., Szczepanski,A., Botwina,P., Labaj,P.P., Pyrc,K., Branicki,W. |
| EPI_ISL_647977, EPI_ISL_647978 | National Microbiology Reference Laboratory | Quadram Institute Bioscience | Thanh Le Viet, Andrew J. Page, Justin O'Grady, Gemma Kay, David Baker, Gaetan Thilliez, Ana-Victoria Gutierrez, Robert Kingsley, Leonardo de Oliveira Martins, Sekesai Zinyowera, Tatenda Takawira, Muchaneta Mugabe, Gibson Mhlanga, Portia Manangazira, Andrew Tarupiwa, Hlanai Gumbo, Agnes Juru, Charles Nyagupe, Alexander Goredema, Isaac Phiri, Barbra Murwira, Beuty Makamure, Tapfumanei Mashe |
| EPI_ISL_648125 | UHAS COVID-19 Lab | UHAS COVID-19 Lab | Kwabena O. Duedu, Jones Gyamfi, Reuben Ayivor-Djanie, John O. Gyapong and the UHAS COVID-19 Lab Team |
| EPI_ISL_648318, EPI_ISL_648321 | Laboratorio de Investigaciones de Baney | University Hospital Basel, Clinical Bacteriology | Carlos Cortes, Claudia Daubenberger, Adrian Egli, Guillermo Garcia, Salome Hosch, Bonifacio Manguire Nlavo, Alfredo Mari, Maximilian Mpina, Elizabeth Nyakarungu, Diosdado Odjama Nseng Ada, Mitoha Ondo O Ayekaba, Tim Roloff, Tobias Schindler, Helena Seth-Smith, Madlen Stange, Philip Wonder Phiri |
| EPI_ISL_649094 | Israel Central Virology laboratory | Israel Central Virology laboratory | Neta Zuckerman, Efrat Dahan Bucris, Oran Erster, Ella Mendelson, Michal Mandelboim |
| EPI_ISL_649123 | Wellington SCL (WN) | Institute of Environmental Science and Research (ESR) | Xiaoyun Ren, Matt Storey, Nikki Freed, Muhammad Faisal, Jing Wang, Hermes Perez, Anja Werno, Antje van der Linden, Arlo Upton, Chris Mansell, David Hammer, Dragana Drinkovic, Gary McAuliffe, Hana Sofia Andersson, James Ussher, Jill Sherwood, Josh Freeman, Julia Howard, Juliet Elvy, Mary DeAlmeida, Matt Blakiston, Matthew Rogers, Max Bloomfield, Michael Addidle, Michelle Balm, Sally Roberts, Sarah Jefferies, Sharmini Muttaiyah, Susan Morpeth, Susan Taylor, Timothy Blackmore, Vani Sathyendran, Veronica Playle, Virginia Hope, Erasmus Smit, Lauren Jelly, Olin Silander, Joep de Ligt |
| EPI_ISL_649124 | Waikato Hospital | Institute of Environmental Science and Research (ESR) | Xiaoyun Ren, Matt Storey, Nikki Freed, Muhammad Faisal, Jing Wang, Hermes Perez, Anja Werno, Antje van der Linden, Arlo Upton, Chris Mansell, David Hammer, Dragana Drinkovic, Gary McAuliffe, Hana Sofia Andersson, James Ussher, Jill Sherwood, Josh Freeman, Julia Howard, Juliet Elvy, Mary DeAlmeida, Matt Blakiston, Matthew Rogers, Max Bloomfield, Michael Addidle, Michelle Balm, Sally Roberts, Sarah Jefferies, Sharmini Muttaiyah, Susan Morpeth, Susan Taylor, Timothy Blackmore, Vani Sathyendran, Veronica Playle, Virginia Hope, Erasmus Smit, Lauren Jelly, Olin Silander, Joep de Ligt |

| | | | |
|---|---|---|---|
| EPI_ISL_653762 | Instituto Nacional de Salud, Bogotá, Colombia | Instituto Nacional de Salud, Bogotá, Colombia | Katherine Laiton-Donato, Diego A. Álvarez-Díaz, Carlos Franco-Muñoz, Mauricio Pacheco-Montealegre, Jonathan Reales, Diego Andrés Prada, Jose A. Usme-Ciro, Zulma M. Cucunubá, Christian Julian VillabonaArenas, Liz Villabona-Arenas, Sussy Echeverria, Astrid C. Flórez, Carolina Ferro, Diana Marcela Walteros-Acero, Franklin Prieto, Carlos Andrés Durán, Martha Lucia Ospina Martinez, Marcela Mercado-Reyes |
| EPI_ISL_654803 | SA Pathology | SA Pathology | Lex Leong, Julien Soubrier, Chuan Kok Lim, Song Gao, Mark Turra, Karin Kassahn, Ivan Bastian, Geoff Higgins |
| EPI_ISL_654873 | Pasteur Institute in Ho Chi Minh city | Department of Microbiology and Immunology - Pasteur Institute in Ho Chi Minh city | Phm Th Thu Hng, ào Huy Mnh, V Phm Hng Nhung, Cao Minh Thng, Hunh Th Kim Loan, Nguyn Hoàng Quân, Hunh Phng Tho, Hoàng Nh ào, Nguyn Trung Hiu, Nguyn Th Thanh Tho, Nguyn Thu Ngc, Phm Th Nhung, Nguyn Th Ngc Tho, ng Thanh Giang, Nguyn Th Thanh Thng, Hoàng Minh, Trn Th Hng Kim, Nguyn Thanh Long, Phm Duy Quang, Lng Chn Quang, Hoàng Quc Cng, Nguyn V Thng, Phan Trng Lân |
| EPI_ISL_660181 | NHLS-IALCH | KRISP, KZN Research Innovation and Sequencing Platform | Gazy I, Sigal A, Karim F, Cele S, Giandhari J, Pillay S, Tegally H, Wilkinson E, de Oliveira T |
| EPI_ISL_660446, EPI_ISL_660447, EPI_ISL_660450, EPI_ISL_660451, EPI_ISL_660454, EPI_ISL_660473, EPI_ISL_660474, EPI_ISL_660476, EPI_ISL_660478, EPI_ISL_660479, EPI_ISL_660519 | | | |
| see above | Laboratoire de Microbiologie CHU Souro Sanou | Centre Muraz | Abdoul-Salam Ouedraogo, Yacouba Sawadogo, Essia Belarbi, Grit Schubert, Fabian Leendertz, Arsène Zongo, Soumeya Ouangraoua, Zekiba Tarnagda, Lassana Sangaré, Halidou Tinto |
| EPI_ISL_660531 | Institute of Microbiology, Universidad San Francisco de Quito | Institute of Microbiology, Universidad San Francisco de Quito | Sully Márquez, Belén Prado-Vivar, Juan José Guadalupe, Monica Becerra-Wong, Bernardo Gutiérrez, Manuel Jaramillo, Verónica Barragán, Patricio Rojas-Silva, Gabriel Trueba, Michelle Grunauer, Paúl Cárdenas |
| EPI_ISL_660543, EPI_ISL_660544 | Laboratory Medicine | Department of Laboratory Medicine, Lin-Kou Chang Gung Memorial Hospital, Taoyuan, Taiwan | Kuo-Chien Tsao, Yu-Nong Gong, Shu-Li Yang, Yi-Chun Liu, Chung-Guei Huang, Mei-Jen Hsiao, Po-Wei Huang, Cheng-Ta Yang, Cheng-Hsun Chiu, Peng-Nien Huang, Kuo-Ming Lee, Guang-Wu Chen, Shin-Ru Shih |
| EPI_ISL_660728 | CHU Nîmes | CNR Virus des Infections Respiratoires - France SUD | Antonin Bal, Gregory Destras, Gwendolyne Burfin, Hadrien Règue, Quentin Semanas, Martine Valette, Bruno Lina, Jean-Philippe Lavigne, Stephan Robin, Maxence Lotellier, Marie-Josée Carles, Laurence Josset |
| EPI_ISL_661196 | Scientific Veterinary Institute Novi Sad | Veterinary Specialized Institute "Kraljevo", Serbia | Vidanovic,D., Tesovic,B., Knezevic,A., Jovanovic,T., Jankovic,M., Sekler,M., Banovic Djeri,B., Petrovic,T., Volkening,J., Afonso,C. |
| EPI_ISL_661210 | Department of Clinical Microbiology | GIGA Medical Genomics | Keith Durkin, Maria Artesi, Sébastien Bontems, Raphaël Boreux, Bouchra Boujemla, Cécile Meex, Pierrette Melin, Marie-Pierre Hayette, Vincent Bours |
| EPI_ISL_663940 | Microbiological Diagnostic Unit - Public Health Laboratory (MDU-PHL) | MDU-PHL | Seemann T., Schultz M.B., Sait, M.L., Sherry, N.L. |
| EPI_ISL_667333 | OHSU Lab Services Molecular Microbiology Lab | Oregon SARS-CoV-2 Genome Sequencing Center | Brendan L. O'Connell, Ruth V. Nichols, Sally Grindstaff, Alec J. Hirsch, Donna Hansel, Guang Fan, Daniel N. Streblow, William B. Messer, Andrew C. Adey, Benjamin N. Bimber, Brian J. O'Roak |
| EPI_ISL_667774 | Microbiology, Infectious Diseases and Immunology, Centre de Recherche du Centre Hospitalier de l'Universite de Montreal | Microbiology, Infectious Diseases and Immunology, Centre de Recherche du Centre Hospitalier de l'Universite de Montreal | Benoit,P., Point,F., Gagnon,S., Hardy,I., Kaufmann,D., Tremblay,C., Coutlee,F., Grandjean-Lapierre,S. |
| EPI_ISL_667809 | Laboratory Medicine | Department of Laboratory Medicine, Lin-Kou Chang Gung Memorial Hospital, Taoyuan, Taiwan | Kuo-Chien Tsao, Yu-Nong Gong, Shu-Li Yang, Yi-Chun Liu, Chung-Guei Huang, Mei-Jen Hsiao, Po-Wei Huang, Cheng-Ta Yang, Cheng-Hsun Chiu, Peng-Nien Huang, Kuo-Ming Lee, Guang-Wu Chen, Shin-Ru Shih |
| EPI_ISL_668411 | Norwegian Institute of Public Health, Department of Virology | Norwegian Institute of Public Health, Department of Virology | Kathrine Stene-Johansen, Kamilla Heddeland Instefjord, Hilde Elshaug, Marie Paulsen Madsen, Rasmus Riis Kopperud, Hilde Vollan, Karoline Bragstad, Olav Hungnes |
| EPI_ISL_668449, EPI_ISL_668451 | Centre for Dengue Research, Department of Immunology and Molecular Medicine | Centre for Dengue Research, Department of Immunology and Molecular Medicine | Chandima Jeewandara, Deshni Jayathilaka, Dinuka Ariyaratne, Diyanath Ranasinghe, Laksiri Gomes, Gathsaurie Neelika Malavige |
| EPI_ISL_671360, EPI_ISL_671391 | National Virus Reference Laboratory | Irish Coronavirus Sequencing Consortium - Teagasc Moorepark | Calum Walsh, Fiona Crispie, John Kenny, Paul Cotter |
| EPI_ISL_672625 | PathWest Laboratory Medicine WA | PathWest Laboratory Medicine WA Microbial Surveillance Unit | PathWest Laboratory Medicine WA Microbial Surveillance Unit |
| EPI_ISL_676592 | Scientific Veterinary Institute Novi Sad | Veterinary Specialized Institute "Kraljevo", Serbia | Vidanovic,D., Tesovic,B., Knezevic,A., Jovanovic,T., Jankovic,M., Sekler,M., Banovic Djeri,B., Petrovic,T., Volkening,J., Afonso,C. |
| EPI_ISL_676989 | Wadsworth Center, New York State Department.of Health | Wadsworth Center, New York State Department.of Health | Kirsten St. George, Daryl M. Lamson, Alexis Russel, Jonathan Plitnick, Navjot Singh, John Kelly, Sara Griesemer, Erasmus Schneider, Erica Lasek-Nesselquist |
| EPI_ISL_677287 | Colorado Department of Public Health and Environment | Colorado Department of Puplic Health and Environment | Laura Bankers, Molly Hetherington-Rauth, Shannon Ely, Shannon R. Matzinger, Sarah Elizabeth Totten, Emily A. Travanty |
| EPI_ISL_677439 | University of Wisconsin-Madison AIDS Vaccine Research Laboratories | University of Wisconsin-Madison AIDS Vaccine Research Laboratories | Gage Moreno, Katarina Braun, et al. AIDS Vaccine Research Laboratories |
| EPI_ISL_677634, EPI_ISL_677635, EPI_ISL_677636 | Virology Unit, Institut Pasteur de Madagascar | Virology Unit, Institut Pasteur de Madagascar | Christian Ranaivoson, Cara E. Brook, Vida Ahyong, Soa Fy Andriamandimby, Vololoniaina Raharinosy, Tsiry Randriambolamanantsoa, Helisoa Razafimanjato, Norosoa Razanajatovo, Michelle Tan, Cristina M. Tato, Joseph L. DeRisi, Jean-Michel Heraud, Philippe Dussart |
| EPI_ISL_677674 | General Hospital - Prilep | Research Center for Genetic Engineering and Biotechnology "Georgi D. Efremov" , Macedonian Academy of Sciences and Arts | RCGEB - MASA |
| EPI_ISL_677715 | Clinical Hospital - Bitola | Research Center for Genetic Engineering and Biotechnology "Georgi D. Efremov" , Macedonian Academy of Sciences and Arts | RCGEB - MASA |
| EPI_ISL_678101 | Pathogen Genomics Lab King Abdullah University of Science and Technology(KAUST) | Pathogen Genomics Lab King Abdullah University of Science and Technology(KAUST) | Amanda Ooi, Luke Esau, Sara Mfarrej, Raeece Naeem, Sharif Hala, Asim Khogeer, Fadwa Alofi, Afrah Alsomali, Jumana Taha, Abdulaziz Alahmadi, Kahled Algithami, Anwar Hashem, Naif Almontashiri, Arnab Pain |
| EPI_ISL_678115 | Pathogen Genomics Lab King Abdullah University of Science and Technology(KAUST) | Pathogen Genomics Lab King Abdullah University of Science and Technology(KAUST) | Luke Esau, Amanda Ooi, Sharif Hala, Raeece Naeem, Sara Mfarrej, Asim Khogeer, Fadwa Alofi, Afrah Alsomali, Jumana Taha, Abdulaziz Alahmadi, Kahled Algithami, Anwar Hashem, Naif Almontashiri, Arnab Pain |
| EPI_ISL_678178, EPI_ISL_678179 | Pathogen Genomics Lab King Abdullah University of Science and Technology(KAUST) | Pathogen Genomics Lab King Abdullah University of Science and Technology(KAUST) | Sara Mfarrej, Sharif Hala, Luke Esau, Amanda Ooi, Raeece Naeem, Awad Al-Omari, Samer Salih, Abbas Al Mutair, Arnab Pain |
| EPI_ISL_678256 | General Hospital - Ohrid | Research Center for Genetic Engineering and Biotechnology "Georgi D. Efremov" , Macedonian Academy of Sciences and Arts | RCGEB - MASA |
| EPI_ISL_678377 | Area of Virology, Serology and Virology Division (SAViD), New South Wales Health Pathology Randwick | Virology Research Laboratory; Area of Virology, Serology and Virology Division (SAViD), New South Wales Health Pathology Randwick | Foster, C.; Au, J.; Ruiz Silva, M.; Deveson, I.; Bull, R.; Van Hal, S.; Rawlinson, W. |
| EPI_ISL_681309 | Communicable Disease Laboratory, Public Health Directorate | Communicable Disease Laboratory, Public Health Directorate | Alwasti,H., Altaif,Z., AlHujairi,Z., AlAbbas,Z. |
| EPI_ISL_681680 | Molecular Medicine Laboratory, University of Magallanes | Centro Asistencial Docente y de Investigacion, Universidad de Magallanes | Jorge González, Jacqueline Aldridge, Diego Alvarez, Marco Montes de Oca, Hermy Alvarez, Roberto Uribe-Paredes, Marcelo Navarrete |
| EPI_ISL_682248 | AREA DE SALUD CIUDAD QUESADA | Inciensa, Instituto Costarricense de Investigación y Enseñanza en Nutrición y Salud | Francisco Duarte, Hebleen Porras, Claudio Soto-Garita, Estela Cordero, Adriana Godinez, Melany Calderon & Mariel López |
| EPI_ISL_682272 | HOSPITAL SAN JUAN DE DIOS | Inciensa, Instituto Costarricense de Investigación y Enseñanza en Nutrición y Salud | Francisco Duarte, Hebleen Porras, Claudio Soto-Garita, Estela Cordero, Adriana Godinez & Melany Calderon |
| EPI_ISL_682276 | LABORATORIOS LABIN | Inciensa, Instituto Costarricense de Investigación y Enseñanza en Nutrición y Salud | Francisco Duarte, Hebleen Porras, Claudio Soto-Garita, Estela Cordero, Adriana Godinez, Melany Calderon & Pei Ling Chan |
| EPI_ISL_682285 | Canterbury Health Laboratories | Institute of Environmental Science and Research (ESR) | Xiaoyun Ren, Matt Storey, Nikki Freed, Muhammad Faisal, Jing Wang, Hermes Perez, Anja Werno, Antje van der Linden, Arlo Upton, Chris Mansell, David |

| | | | |
|---|---|---|---|
| | | | Hammer, Dragana Drinkovic, Gary McAuliffe, Hana Sofia Andersson, James Ussher, Jill Sherwood, Josh Freeman, Julia Howard, Juliet Elvy, Mary DeAlmeida, Matt Blakiston, Matthew Rogers, Max Bloomfield, Michael Addidle, Michelle Balm, Sally Roberts, Sarah Jefferies, Sharmini Muttaiyah, Susan Morpeth, Susan Taylor, Timothy Blackmore, Vani Sathyendran, Veronica Playle, Virginia Hope, Erasmus Smit, Lauren Jelly, Olin Silander, Joep de Ligt |
| EPI_ISL_682300, EPI_ISL_682322 | Communicable Disease Laboratory, Public Health Directorate | Communicable Disease Laboratory, Public Health Directorate | Alwasti,H., Altaif,Z., AlHujairi,Z., AlAbbas,Z. |
| EPI_ISL_682350 | NHLS Universitas Academic | UFS Virology | PA Bester, MM Nyaga, P Nthiga, MT Mogotsi, D Goedhals, T de Oliveira |
| EPI_ISL_683355 | CNR Virus des Infections Respiratoires - France SUD | CNR Virus des Infections Respiratoires - France SUD | Antonin Bal, Gregory Destras, Gwendolyne Burfin, Quentin Semanas, Martine Valette, Bruno Lina, Laurence Josset |
| EPI_ISL_683835 | CICM | Malaria Research and Training Center (MRTC-Parasito) | Antoine Dara, Abdoulaye Djimde |
| EPI_ISL_686846, EPI_ISL_688071, EPI_ISL_688493, EPI_ISL_690706 | Pathogen Genomics Center, National Institute of Infectious Diseases | Pathogen Genomics Center, National Institute of Infectious Diseases | Tsuyoshi Sekizuka, Kentaro Itokawa, Rina Tanaka, Masanori Hashino, Makoto Kuroda |
| EPI_ISL_693283, EPI_ISL_693284, EPI_ISL_693285, EPI_ISL_693290 | Public Health Virology Laboratory, Forensic and Scientific Services (PHV-FSS) | Public Health Virology Laboratory, Forensic and Scientific Services (PHV-FSS) | Son Nguyen et al |
| EPI_ISL_693305 | Department of Laboratory Medicine, National Taiwan University Hospital | Microbial Genomics Core Lab, National Taiwan University Centers of Genomic and Precision Medicine | Shiou-Hwei Yeh, You-Yu Lin, Ya-Yun Lai, Chiao-Ling Li, Shan-Chwen Chang, Pei-Jer Chen, Sui-Yuan Chang |
| EPI_ISL_693472, EPI_ISL_693473, EPI_ISL_693477, EPI_ISL_693482, EPI_ISL_693483 | Central Public Health Laboratory | National Public Health Laboratory, National Centre for Infectious Diseases | Tze Minn Mak, Sophie Octavia, Zhenyang Zhou, Esorom Daoni, Theresa Palou, Lin Cui, Raymond Tzer Pin Lin |
| EPI_ISL_694532 | AZ SPHL, Arizona Department of Health Services | TGen North | Jolene Bowers, Megan Folkerts, Chris French, Hayley Yaglom, Ashlyn Pfeiffer, Darrin Lemmer, Dave Engelthaler, The Arizona COVID Genomics Union (ACGU) |
| EPI_ISL_697785 | Institute of Microbiology, Universidad San Francisco de Quito | Institute of Microbiology, Universidad San Francisco de Quito | Belén Prado-Vivar, Sully Márquez, Juan José Guadalupe, Monica Becerra-Wong, Bernardo Gutiérrez, Jonathan Araujo, Verónica Barragán, Patricio Rojas-Silva, Gabriel Trueba, Michelle Grunauer, Paúl Cárdenas |
| EPI_ISL_697795 | Institute of Microbiology, Universidad San Francisco de Quito | Institute of Microbiology, Universidad San Francisco de Quito | Andrea Macias, Belén Prado-Vivar, Sully Márquez, Juan José Guadalupe, Monica Becerra-Wong, Bernardo Gutiérrez, Verónica Barragán, Patricio Rojas-Silva, Gabriel Trueba, Michelle Grunauer, Paúl Cárdenas |
| EPI_ISL_700102 | Hematopathology Laboratory, ACTREC, TMC | Hematopathology Laboratory, ACTREC, TMC | Hematopathology Laboratory, ACTREC |
| EPI_ISL_700336 | Child Health Research Foundation | Child Health Research Foundation | Senjuti Saha, Afroza Akter Tanni, Syed Muktadir Al Sium, Roly Malaker, Sharmistha Goswami, Arif Mohammad Tanmoy, Md Hafizur Rahman, Samir K Saha |
| EPI_ISL_707794 | Waikato Hospital | Institute of Environmental Science and Research (ESR) | Xiaoyun Ren, Matt Storey, Nikki Freed, Muhammad Faisal, Jing Wang, Hermes Perez, Anja Werno, Antje van der Linden, Arlo Upton, Chris Mansell, David Hammer, Dragana Drinkovic, Gary McAuliffe, Hana Sofia Andersson, James Ussher, Jill Sherwood, Josh Freeman, Julia Howard, Juliet Elvy, Mary DeAlmeida, Matt Blakiston, Matthew Rogers, Max Bloomfield, Michael Addidle, Michelle Balm, Sally Roberts, Sarah Jefferies, Sharmini Muttaiyah, Susan Morpeth, Susan Taylor, Timothy Blackmore, Vani Sathyendran, Veronica Playle, Virginia Hope, Erasmus Smit, Lauren Jelly, Olin Silander, Joep de Ligt |
| EPI_ISL_708751 | PathWest Laboratory Medicine WA | PathWest Laboratory Medicine WA Microbial Surveillance Unit | PathWest Laboratory Medicine WA Microbial Surveillance Unit |
| EPI_ISL_708800 | Regional medical sciences center 6 chonburi | National Institute of Health, Department of Medical Sciences, Ministry of Public Health, Thailand | Pilailuk Okada; Siripaporn Phuygun; Thanutsapa Thanadachakul; Sittiporn Parnmen; Warawan Wongboot; Sunthareeya Waicharoen; Malinee Chittaganpitch |
| EPI_ISL_708801, EPI_ISL_708804 | Regional medical sciences center 6 chonburi | National Institute of Health, Department of Medical Sciences, Ministry of Public Health, Thailand | Pilailuk Okada; Siripaporn Phuygun; Thanutsapa Thanadachakul; Sittiporn Parnmen; Pakorn Piromtong; Warawan Wongboot; Sunthareeya Waicharoen; Malinee Chittaganpitch |
| EPI_ISL_708812 | World Medical Hospital | National Institute of Health, Department of Medical Sciences, Ministry of Public Health, Thailand | Pilailuk Okada; Siripaporn Phuygun; Thanutsapa Thanadachakul; Sittiporn Parnmen; Pakorn Piromtong; Warawan Wongboot; Sunthareeya Waicharoen; Malinee Chittaganpitch |
| EPI_ISL_708815 | Urban Institute for Disease Prevention and Control | National Institute of Health, Department of Medical Sciences, Ministry of Public Health, Thailand | Pilailuk Okada; Siripaporn Phuygun; Thanutsapa Thanadachakul; Sittiporn Parnmen; Pakorn Piromtong; Warawan Wongboot; Sunthareeya Waicharoen; Malinee Chittaganpitch |
| EPI_ISL_708821 | Synphaet Hospital | National Institute of Health, Department of Medical Sciences, Ministry of Public Health, Thailand | Pilailuk Okada; Siripaporn Phuygun; Thanutsapa Thanadachakul; Sittiporn Parnmen; Pakorn Piromtong; Warawan Wongboot; Sunthareeya Waicharoen; Malinee Chittaganpitch |
| EPI_ISL_709542 | National Institute of Blood Diseases (NIBD), Molecular Biology Lab | Genomics Lab NIBD | Samina Naz Mukry, Sayed Ali Raza, Shariq Ahmed, Aneeta Shahni, Gul Sufaida, Arshi Naz , Tahir Sultan Shamsi |
| EPI_ISL_710483 | Department of Medical Laboratory Sciences, Arab American University | Department of Medical Laboratory Sciences, Arab American University | Al-Jawabreh,A., Nasereddin,A., Dumaidi,K., Al-Jawabreh,H., Ereqat,S. |
| EPI_ISL_710572, EPI_ISL_710573 | University Hospital Dubrava | Ruer Boškovic Institute; Forensic Science Centre Ivan Vueti; University of Zagreb Faculty of Science | Robert Beluži, Marina Korolija, Ana Livun, Vjekoslav Tomai, Dunja Glavaš, Maja Kuzman, Paula Štancl, Lucija Markulin, Lucija Basi, Antonela Blažekovi, Fran Boroveki, Lidija Cvetko-Krajinovi, Ivana elap, Fuad osovi, Mirjana Domazet-Lošo, Tomislav Domazet-Lošo, Valentina umlijan-Combaj, Kristina Gotovac Jerei, Jasna Kašman, Vladimir Krajinovi, Danilo Licastro, Boris Maek, Željka Maak Šafranko, Gordana Maravi Vlahoviek, Senica Pejša, Josipa Skelin, Ivan Šamija, Mario Štefanovi, Sanja Tadinac, Katarina Marija Tupek, Petra Vrabec, Rosa Karli, Kristian Vlahoviek |
| EPI_ISL_710581 | The Public Health Agency of Sweden | The Public Health Agency of Sweden | Department of Microbiology, The Public Health Agency of Sweden |
| EPI_ISL_712034 | Department of Virus and Microbiological Special Diagnostics, Statens Serum Institut, Copenhagen, Denmark | Albertsen Lab, Department of Chemistry and Bioscience, Aalborg University, Denmark | Danish Covid-19 Genome Consortium |
| EPI_ISL_717692, EPI_ISL_717697 | Trinidad Public Health Laboratory | Carrington Lab, Department of PreClinical Sciences, Faculty of Medical Sciences, The University of the West Indies | Nikita S. D. Sahadeo, Arianne Brown-Jordan, Sarah Hill, Vernie Ramkissoon, Naresh Nandram, Avery Hinds, Jerome Foster, Stanley Giddings, Karla Georges, Marsha Ivey, Rahul Naidu, Risha Singh, SueMin Nathaniel, Rajini Haraksingh, Jaya Jayaraman, Chinna Chinnadurai, Adesh Ramsubhag, Nuno Faria, Oliver Pybus, Christopher Oura, Gabriel Escobar, Christine V. F. Carrington |
| EPI_ISL_717893, EPI_ISL_717957 | Laboratorio de Virologia Molecular / UFRJ | Bioinformatics Laboratory / LNCC | Carolina M Voloch, Ronaldo da Silva F Jr, Luiz G P de Almeida, Cynthia C Cardoso, Otavio Bustrolini, Alexandra L Gerber, Ana Paula de C Guimarães, Diana Mariani, Andréa Cony Cavalcanti, Claudia dos Santos Rodrigues, Terezinha M P P Castiñeira, Amilcar Tanuri, Ana Tereza R de Vasconcelos |
| EPI_ISL_717975 | Institute of Virology, Biomedical Research Center of the Slovak Academy of Sciences, Bratislava | Faculty of Natural Sciences, Comenius University, Bratislava | Broa Brejová, Viktória Hodorová, Kristína Boršová, Viktória abanová, Dominika Friová, Sabina Fumaová Havlíková, Juraj Kopáek, Martina Liková, ubomíra Lukáiková, Martina Neboháová, Monika Sláviková, Edita Staroová, Elena Tichá, Tomáš Vina, Jozef Nosek, Boris Klempa |
| EPI_ISL_718177, EPI_ISL_718189, EPI_ISL_718193, EPI_ISL_718213 | Ministry of Health Hospitals | Institute of Health and Community Medicine | David Perera, Ooi Mong How, Chua Hock Hin, Tonnii Sia Loong Loong, Wong Jyn Shan, Wong Kiing Aik, Chan Chia Jui |
| EPI_ISL_718254 | Institute of Virology, Biomedical Research Center of the Slovak Academy of Sciences, Bratislava | Faculty of Natural Sciences, Comenius University, Bratislava | Kristína Boršová, Viktória Hodorová, Broa Brejová, Viktória abanová, Sabina Fumaová Havlíková, Juraj Kopáek, Martina Liková, ubomíra Lukáiková, Martina Neboháová, Monika Sláviková, Tomáš Vina, Boris Klempa, Jozef Nosek |
| EPI_ISL_718280, EPI_ISL_718301 | Institute for Medical Research, Infectious Disease Research Centre, National Institutes of Health, Ministry of Health Malaysia | Institute for Medical Research, Infectious Disease Research Centre, National Institutes of Health, Ministry of Health Malaysia | Suppiah J, Kamel K, Mohd-Zawawi Z, Thayan R |
| EPI_ISL_719202 | Lighthouse Lab in Cambridge | Wellcome Sanger Institute for the COVID-19 Genomics UK (COG-UK) Consortium | Rob Howes, The Lighthouse Lab in Cambridge and Alex Alderton, Roberto Amato, Sonia Goncalves, Ewan Harrison, David K. Jackson, Ian Johnston, Dominic Kwiatkowski, Cordelia Langford, John Sillitoe on behalf of the Wellcome Sanger Institute COVID-19 Surveillance Team |
| EPI_ISL_722014 | Hospital das Clínicas Universidade de São Paulo Medical School | Laboratório de Parasitologia Médica - Instituto de Medicina Tropical - Universidade de São Paulo | Brazil-UK Centre for Arbovirus Discovery Diagnosis Genomics and Epidemiology (CADDE) Genomic Network - Instituto de Medicina Tropical |
| EPI_ISL_722201 | Vitalis Mostar | Alea Genetic Center | Salihefendic L., Pecar D., Konjhodzic R. |

| | | | |
|---|---|---|---|
| EPI_ISL_728161 | Institute for Medical Research, Infectious Disease Research Centre, National Institutes of Health, Ministry of Health Malaysia | Institute for Medical Research, Infectious Disease Research Centre, National Institutes of Health, Ministry of Health Malaysia | Suppiah J, Kamel K, Mohd-Zawawi Z, Thayan R |
| EPI_ISL_729577 | A. Krumbholz, Labor Dr. Krause und Kollegen MVZ GmbH, Kiel | Charité Universitätsmedizin Berlin, Institut für Virologie | Victor M Corman, Barbara Mühlemann, Jörn Beheim-Schwarzbach, Talitha Veith, Julia Schneider, Terry Jones, Christian Drosten |
| EPI_ISL_729815 | Laboratório Central de Saúde Pública do Estado do Rio Grande do Sul (LACEN-RS) | Laboratory of Respiratory Viruses and Measles, Oswaldo Cruz Institute, FIOCRUZ | Paola Resende, Luciana Appolinario, Fernando Motta, Anna Carolina Paixão, Ana Carolina Mendonça, Tatiana Schaffer Gregianini, Marilda Tereza Mar da Rosa, Marilda Siqueira on behalf of the Fiocruz COVID-19 Genomic Surveillance Network |
| EPI_ISL_729880 | Laboratorio de Referencia Nacional de Virus Respiratorios, Instituto Nacional de Salud Peru | Laboratorio de Genómica Microbiana, Universidad Peruana Cayetano Heredia | Pablo Tsukayama, Alejandra Dávila-Barclay, Luis González, Guillermo Salvatierra, Pedro E. Romero, Brenda Ayzanoa, Janet Huancachoque, Pool Marcos, Marco Galarza, Priscila Lope, Nancy Rojas |
| EPI_ISL_729911 | Instituto de Medicina Tropical, Universidad Nacional Toribio Rodríguez de Mendoza de Amazonas | Laboratorio de Genómica Microbiana, Universidad Peruana Cayetano Heredia | Pablo Tsukayama, Alejandra Dávila-Barclay, Luis González, Pedro E. Romero, Brenda Ayzanoa, Janet Huancachoque, Pool Marcos, Stella Chenet, Rafael Tapia, Cecilia Pajuelo, Carla Montenegro |
| EPI_ISL_729982, EPI_ISL_729984, EPI_ISL_729985 | Nigeria Centre for Disease Control (NCDC) | African Centre of Excellence for Genomics of Infectious Diseases (ACEGID), Redeemer's University, Ede, Osun State, Nigeria | Oluniyi P.E. et al |
| EPI_ISL_730259, EPI_ISL_730553 | Biolab Diagnostic Laboratories | Andersen lab at Scripps Research | Issa Abu-Dayyeh, Ahmad Tibi, Lama Hussein, Lina Mohammad, Zein Naber, Amid Abdelnour with SEARCH Alliance San Diego |
| EPI_ISL_732530 | Bundeswehr Institute of Microbiology | Bundeswehr Institute of Microbiology | Markus Antwerpen, Alexandra Rehn, Mathias Walter, Malena Bestehorn-Willmann, Sabine Zange, Enrico Georgi, Roman Wölfel |
| EPI_ISL_732957, EPI_ISL_732959 | Department of Tropical Parasitology | Laboratory of Recombinant Vaccines | Lukasz Rabalski, Maciej Kosinski, Teemu Smura, Kirsi Aaltonen, Ravi Kant, Tarja Sironen, Boguslaw Szewczyk, Maciej Grzybek |
| EPI_ISL_732960, EPI_ISL_732961 | SA Pathology | SA Pathology | Lex Leong, Julien Soubrier, Chuan Kok Lim, Song Gao, Mark Turra, Karin Kassahn, Ivan Bastian, Geoff Higgins |
| EPI_ISL_732963 | Canterbury Health Laboratories | Institute of Environmental Science and Research (ESR) | Xiaoyun Ren, Matt Storey, Nikki Freed, Muhammad Faisal, Jing Wang, Hermes Perez, Anja Werno, Antje van der Linden, Arlo Upton, Chris Mansell, David Hammer, Dragana Drinkovic, Gary McAuliffe, Hana Sofia Andersson, James Ussher, Jill Sherwood, Josh Freeman, Julia Howard, Juliet Elvy, Mary DeAlmeida, Matt Blakiston, Matthew Rogers, Max Bloomfield, Michael Addidle, Michelle Balm, Sally Roberts, Sarah Jefferies, Sharmini Muttaiyah, Susan Morpeth, Susan Taylor, Timothy Blackmore, Vani Sathyendran, Veronica Playle, Virginia Hope, Erasmus Smit, Lauren Jelly, Olin Silander, Joep de Ligt |
| EPI_ISL_733220, EPI_ISL_733222 | Pathogenic Microorganisms Variability Laboratory | WHO National Influenza Centre Russian Federation | Andrey Komissarov, Artem Fadeev, Anna Ivanova, Kseniya Komissarova, Dmitry Bazhenov, Daria Danilenko, Ksenia Safina, Elena Nabieva, Georgii Bazykin, Nadezhda Kuznetsova, Elena Shidlovskaya, Sergey Alkhovsky, Tatyana Vishnevskaya, Elizaveta Divisenko, Alexey Shchetinin, Maria Nikiforova, Andrey Pochtovyy, Evgeny Usachev, Elena Vokalova, Maxim Rubalsky, Oleg Rubalsky, Artem Tkachuk, Vladimir Gushchin, Alexander Gintsburg, Dmitry Lioznov |
| EPI_ISL_733433 | HELIX LLC | WHO National Influenza Centre Russian Federation | Andrey Komissarov, Artem Fadeev, Anna Ivanova, Kseniya Komissarova, Dmitry Bazhenov, Daria Danilenko, Ksenia Safina, Elena Nabieva, Georgii Bazykin, Dmitry Lioznov |
| EPI_ISL_733500 | 1-Laboratory of Microbiology, National Reference Lab, Charles Nicolle Hospital; 2-University of Tunis ElManar, Faculty of Medicine of Tunis, LR99ES09, Tunis, Tunisia | 1-Clinical and Experimental Pharmacology Lab, LR16SP02, National Center of Pharmacovigilance, University of Tunis El Manar, Tunis, Tunisia. 2-Neurodegenerative diseases and psychiatric troubles, LR18SP03, Razi Hospital, University of Tunis El Manar, Tunis, Tunisia. 3- Ministry of Health, National Observatory of New and Emerging Diseases, 1006, Tunis, Tunisia | Ilhem Boutiba-Ben Boubaker, Sameh Trabelsi, Nissaf Ben Alaya, Maher Kharrat, Alia Ben Kahla, Jalila Ben Khelil, Salma Abid, Sana Ferjani, Mouna Ben Sassi, Mouna Safer, Guedi Ali Barreh, Habiba Ben Romdhane, Souissi Amira, Sarra Chamman, Hanen El Jebari, Asma Ferjani, Gaies Emna, Riadh Daghfous, Riadh Gouider. |
| EPI_ISL_733528, EPI_ISL_733561 | Institute for Urban Disease Control and Prevention | COVID-19 Network Investigations (CONI) Alliance | Kamolthip Atsawawaranunt, Elizabeth Batty, Wasun Chantratita, Thanat Chookajorn, Stefan Fernandez, Angkana Huang, Anthony R. Jones, Khajohn Joonsalak, Chonticha Klungtong, Theerarat Kochakarn, Prayuth Kaewmalang, Amornmas Kongklieng, Namfon Kotanan, Krittikorn Kumpornsin, Wudtichai Manasatienkij, Anek Mungaomklang, Bhakbhoom Panthan, Pukkaporn Parnwijitkul, Ekawat Pasomsub, Kingkan Rakmanee, Insee Sensorn, Janjira Thaipadungpanit, Arporn Wangwiwatsin, Treewat Watthanachockchai |
| EPI_ISL_735319 | Genomic Laboratory (GLAB) (Conjoint lab of Health Directorate of Istanbul and Istanbul Technical University) | Genomic Laboratory (GLAB), Istanbul Technical University | Ilker Karacan, Tugba Kizilboga Akgun, Nihat Bugra Agaoglu, Payam Zolfagharian, Mehtap Aydin, Gizem Alkurt, Jale Yildiz, Betsi Kose, Nisan Denizce Can, Ayse Serra Ozel, Nilsun Altunal, Arzu Irvem, Yasemin Kendir Demirkol, Ozlem Akgun Dogan, Levent Doganay, Gizem Dinler Doganay |
| EPI_ISL_735402 | Instituto Adolfo Lutz - Regional de Rio Claro | Instituto Adolfo Lutz, Interdiciplinary Procedures Center, Strategic Laboratory | Claudio Tavares Sacchi, Claudia Regina Gonçalves, Erica Valessa Ramos Gomes, Karoline Rodrigues Campos |
| EPI_ISL_735502 | Abdul Malek Ukil Medical College, Noakhali | Central Biological Research Laboratory and Department of Biochemistry and Molecular Biology | H. M. Abdullah Al Masud, Mohammad Omar Faruque, Sajib Rudra, Md. Khondakar Raziur Rahman, Imam Hossen, Md. Arif Hossain, Shanta Paul, Md. Omer Faruq, Md. Imranul Hoq, Robiul Hasan Bhuiyan |
| EPI_ISL_735503 | ACT Pathology | Schwessinger Lab | Robyn N Hall, Benjamin Schwessinger, Ashley Jones, Robert Lanfear, Megan McDonald, Ming-Dao Chia, Kevin Murray, Craig Kennedy, Karina Kennedy |
| EPI_ISL_737078 | Department of Virology and Immunology, University of Helsinki and Helsinki University Hospital, Huslab Finland | Department of Virology, Faculty of Medicine, University of Helsinki, Helsinki, Finland | Teemu Smura, Ravi Kant, Phuoc Truong, Hussein Alburkat, Hannimari Kallio-Kokko, Jenni Virtanen, Maija Suvanto, Sari Hannula, Harri Kangas, Pekka Ellonen, Olli Vapalahti |
| EPI_ISL_737330 | Department of Clinical Microbiology | GIGA Medical Genomics | Keith Durkin, Maria Artesi, Sébastien Bontems, Raphaël Boreux, Bouchra Boujemla, Cécile Meex, Pierrette Melin, Marie-Pierre Hayette, Vincent Bours |
| EPI_ISL_737955, EPI_ISL_737963, EPI_ISL_737970, EPI_ISL_737973, EPI_ISL_737974, EPI_ISL_737979, EPI_ISL_738040, EPI_ISL_738041 | Uganda Central Public Health Lab and Uganda Virus Research Institute | MRC/UVRI & LSHTM Uganda Research Unit | Matthew Cotten, Dan Lule Bugembe, My V.T. Phan, Pontiano Kaleebu et al. |
| EPI_ISL_738137 | Area of Virology, Serology and Virology Division (SAViD), New South Wales Health Pathology Randwick | Area of Virology, Serology and Virology Division (SAViD), New South Wales Health Pathology Randwick | Rawlinson, W., Deveson, I., Van Haal, S., Foster, C. |
| EPI_ISL_739659 | Al-Quds Nutrition and Health Research Institute, Al-Quds University | Al-Quds Nutrition and Health Research Institute, Al-Quds University | Nasereddin,A., Ereqat,S., Al-Jawabreh,A., Rishmawi,C. |
| EPI_ISL_739675 | Instituto Nacional de Salud, Bogotá, Colombia | Instituto Nacional de Salud, Bogotá, Colombia | Katherine Laiton-Donato, Diego A. Álvarez-Díaz, Carlos Franco-Muñoz, Mauricio Pacheco-Montealegre, Jonathan Reales, Diego Andrés Prada, Sheryl Corchuelo, Magdalena Weisner, Martha Lucia Ospina Martinez, Marcela Mercado-Reyes |
| EPI_ISL_740259 | Laboratoire national de santé, Microbiology, Virology | Laboratoire national de santé, Microbiology, Microbial Genomics Platform | Anke Wienecke-Baldacchino, Catherine Ragimbeau, Tamir Abdelrahman, Jessica Tapp, Fatu Djabi |
| EPI_ISL_740547 | Department of Laboratory Medicine, National Taiwan University Hospital | Microbial Genomics Core Lab, National Taiwan University Centers of Genomic and Precision Medicine | Shiou-Hwei Yeh, You-Yu Lin, Ya-Yun Lai, Chiao-Ling Li, Shan-Chwen Chang, Pei-Jer Chen, Sui-Yuan Chang |
| EPI_ISL_744367 | Laboratoire national de santé, Microbiology, Virology | Laboratoire national de santé, Microbiology, Microbial Genomics Platform | Anke Wienecke-Baldacchino, Catherine Ragimbeau, Tamir Abdelrahman, Jessica Tapp, Fatu Djabi |
| EPI_ISL_744489 | Laboratoire national de santé, Microbiology, Virology | Laboratoire national de santé, Microbiology, Microbial Genomics Platform | Anke Wienecke-Baldacchino, Catherine Ragimbeau,Jessica Tapp, Fatu Djabi, Lise Pignon, Raoul Salmon, Tamir Abdelrahman |
| EPI_ISL_745030 | Area of Virology, Serology and Virology Division (SAViD), New South Wales Health Pathology Randwick | Area of Virology, Serology and Virology Division (SAViD), New South Wales Health Pathology Randwick | Rawlinson, W. |
| EPI_ISL_745072 | Israel Central Virology laboratory | Israel Central Virology laboratory | Neta Zuckerman, Efrat Dahan Bucris, Oran Erster, Michal Mandelboim, Orna Mor, Ella Mendelson |
| EPI_ISL_746507, EPI_ISL_746516, EPI_ISL_746525, EPI_ISL_746580, | Genetica Molecular and Subdepartamento de Virologia ISP Chile | Instituto de Salud Publica de Chile | Javier Tognarelli, Barbara Parra, Loredana Arata, Jaime Lagos, Gisselle Barra, Patricia Bustos, Rodrigo Fasce, Andres Castillo, Jorge Fernandez |

| | | | |
|---|---|---|---|
| EPI_ISL_746586, EPI_ISL_746591, EPI_ISL_746675, EPI_ISL_746733, EPI_ISL_746747, EPI_ISL_746764 | | | |
| EPI_ISL_747067, EPI_ISL_747127 | Respiratory Viruses Branch, Centers for Disease Control and Prevention | Respiratory Viruses Branch, Centers for Disease Control and Prevention | Queen,K., Li,Y., Tao,Y., Uehara,A., Montmayeur,A., Paden,C.R., Cook,P.W., Marine,R., Sheth,M., Wang,H., Lee,J., Tong,S. |
| EPI_ISL_748139 | Instituto de Investigaciones Biologicas Clemente Estable | Institut Pasteur de Montevideo | Daiana Mir, Natalia Rego, Paola Cristina Resende, Fernando Lopez-Tort, Tamara Fernandez-Calero, Veronica Noya, Mariana Brandes, Tania Possi, Mailen Arleo, Natalia Reyes, Matias Victoria, Andres Lizasoain, Matias Castells, Leticia Maya, Matías Salvo, Tatiana Schäffer Gregianini, Marilda Tereza Mar da Rosa, Letícia Garay Martins, Cecilia Alonso, Yasser Vega, Cecilia Salazar, Ignacio Ferrés, Pablo Smircich, Jose Sotelo, Ighor Arantes, Luciana Appolinario, Ana Carolina Mendonça, Maria Jose Benitez-Galeano, Martín Graña, Camila Simoes, Fernando Motta, Marilda Mendonça Siqueira, Gonzalo Bello, Rodney Colina, Lucia Spangenberg |
| EPI_ISL_750108, EPI_ISL_750161, EPI_ISL_750173, EPI_ISL_750174, EPI_ISL_751011 | Sanatorio Americano | Institut Pasteur de Montevideo | Daiana Mir, Natalia Rego, Paola Cristina Resende, Fernando Lopez-Tort, Tamara Fernandez-Calero, Veronica Noya, Mariana Brandes, Tania Possi, Mailen Arleo, Natalia Reyes, Matias Victoria, Andres Lizasoain, Matias Castells, Leticia Maya, Matías Salvo, Tatiana Schäffer Gregianini, Marilda Tereza Mar da Rosa, Letícia Garay Martins, Cecilia Alonso, Yasser Vega, Cecilia Salazar, Ignacio Ferrés, Pablo Smircich, Jose Sotelo, Ighor Arantes, Luciana Appolinario, Ana Carolina Mendonça, Maria Jose Benitez-Galeano, Martín Graña, Camila Simoes, Fernando Motta, Marilda Mendonça Siqueira, Gonzalo Bello, Rodney Colina, Lucia Spangenberg |
| EPI_ISL_751229 | Pathogen Genomics Lab King Abdullah University of Science and Technology(KAUST) | Pathogen Genomics Lab King Abdullah University of Science and Technology(KAUST) | Sara Mfarrej, Sharif Hala, Raushan Nugmanova, Olga Douvropoulou, Raeece Naeem, Fadwa Alofi, Asim Khogeer, Afrah Alsomali, Jumana Taha, Abdulaziz Alahmadi, Kahled Algithami, Anwar Hashem, Naif Almontashiri, Arnab Pain |
| EPI_ISL_751563 | NE Public Health Laboratory | Genomics and Discovery, Respiratory Viruses Branch, Division of Viral Diseases, Centers for Disease Control and Prevention | Krista Queen, Yan Li, Ying Tao, Jing Zhang, Anna Uehara, Anna Montmayeur, Clinton R. Paden, Peter W. Cook,Rachel Marine, Mili Sheth, Haibin Wang, Justin Lee, Suxiang Tong |
| EPI_ISL_752544 | National Virus Reference Laboratory | National Virus Reference Laboratory | Michael Carr, Gabriel Gonzalez, Jonathan Dean, Daniel Hare, Cillian F De Gascun |
| EPI_ISL_752598, EPI_ISL_752599 | SA Pathology | SA Pathology | Lex Leong, Julien Soubrier, Chuan Kok Lim, Song Gao, Mark Turra, Karin Kassahn, Ivan Bastian, Geoff Higgins |
| EPI_ISL_752663, EPI_ISL_752963 | State Laboratories Division, Hawaii State Department of Health | State Laboratories Division, Hawaii State Department of Health | Pamela O'Brien, Sabrina Diemert, Drew Kuwazaki, Razvan Sultana, Edward Desmond |
| EPI_ISL_754080, EPI_ISL_754082, EPI_ISL_754087 | National Public Health Laboratory, National Centre for Infectious Diseases | National Public Health Laboratory, National Centre for Infectious Diseases | Tze Minn Mak, Sophie Octavia, Zhenyang Zhou, Lin Cui, Raymond Tzer Pin Lin |
| EPI_ISL_754193 | Charité Universitätsmedizin Berlin, Institut für Virologie/Labor Berlin | Charité Universitätsmedizin Berlin, Institut für Virologie | Victor M Corman, Jörn Beheim-Schwarzbach, Barbara Mühlemann, Julia Schneider, Talitha Veith, Terry Jones, Christian Drosten |
| EPI_ISL_754757 | Emory Molecular Diagnostics Laboratory, Emory Healthcare | Piantadosi Lab, Emory Department of Pathology | Ahmed Babiker, Anne Piantadosi |
| EPI_ISL_755126, EPI_ISL_755129, EPI_ISL_755236, EPI_ISL_755239, EPI_ISL_755242, EPI_ISL_755244 | Biolab Diagnostic Laboratories | Andersen lab at Scripps Research | Issa Abu-Dayyeh, Ahmad Tibi, Lama Hussein, Lina Mohammad, Zein Naber, Amid Abdelnour with SEARCH Alliance San Diego |
| EPI_ISL_755425 | Maine Health and Environmental Testing Laboratory | Tewhey Lab, The Jackson Laboratory | Matluk,N., Dewey,H., Iosue,F., Barter,M., Lynch,R., Munger,H. and Tewhey,R. |
| EPI_ISL_755568, EPI_ISL_755569, EPI_ISL_755570, EPI_ISL_755571 | SA Pathology | SA Pathology | Lex Leong, Julien Soubrier, Chuan Kok Lim, Song Gao, Mark Turra, Karin Kassahn, Ivan Bastian, Geoff Higgins |
| EPI_ISL_755629, EPI_ISL_755633 | LabPLUS | Institute of Environmental Science and Research (ESR) | Xiaoyun Ren, Matt Storey, Nikki Freed, Muhammad Faisal, Jing Wang, Hermes Perez, Anja Werno, Antje van der Linden, Arlo Upton, Chris Mansell, David Hammer, Dragana Drinkovic, Gary McAuliffe, Hana Sofia Andersson, James Ussher, Jill Sherwood, Josh Freeman, Julia Howard, Juliet Elvy, Mary DeAlmeida, Matt Blakiston, Matthew Rogers, Max Bloomfield, Michael Addidle, Michelle Balm, Sally Roberts, Sarah Jefferies, Sharmini Muttaiyah, Susan Morpeth, Susan Taylor, Timothy Blackmore, Vani Sathyendran, Veronica Playle, Virginia Hope, Erasmus Smit, Lauren Jelly, Olin Silander, Joep de Ligt |
| EPI_ISL_755634 | Middlemore Hospital | Institute of Environmental Science and Research (ESR) | Xiaoyun Ren, Matt Storey, Nikki Freed, Muhammad Faisal, Jing Wang, Hermes Perez, Anja Werno, Antje van der Linden, Arlo Upton, Chris Mansell, David Hammer, Dragana Drinkovic, Gary McAuliffe, Hana Sofia Andersson, James Ussher, Jill Sherwood, Josh Freeman, Julia Howard, Juliet Elvy, Mary DeAlmeida, Matt Blakiston, Matthew Rogers, Max Bloomfield, Michael Addidle, Michelle Balm, Sally Roberts, Sarah Jefferies, Sharmini Muttaiyah, Susan Morpeth, Susan Taylor, Timothy Blackmore, Vani Sathyendran, Veronica Playle, Virginia Hope, Erasmus Smit, Lauren Jelly, Olin Silander, Joep de Ligt |
| EPI_ISL_755637 | LabPLUS | Institute of Environmental Science and Research (ESR) | Xiaoyun Ren, Matt Storey, Nikki Freed, Muhammad Faisal, Jing Wang, Hermes Perez, Anja Werno, Antje van der Linden, Arlo Upton, Chris Mansell, David Hammer, Dragana Drinkovic, Gary McAuliffe, Hana Sofia Andersson, James Ussher, Jill Sherwood, Josh Freeman, Julia Howard, Juliet Elvy, Mary DeAlmeida, Matt Blakiston, Matthew Rogers, Max Bloomfield, Michael Addidle, Michelle Balm, Sally Roberts, Sarah Jefferies, Sharmini Muttaiyah, Susan Morpeth, Susan Taylor, Timothy Blackmore, Vani Sathyendran, Veronica Playle, Virginia Hope, Erasmus Smit, Lauren Jelly, Olin Silander, Joep de Ligt |
| EPI_ISL_755638 | Canterbury Health Laboratories | Institute of Environmental Science and Research (ESR) | Xiaoyun Ren, Matt Storey, Nikki Freed, Muhammad Faisal, Jing Wang, Hermes Perez, Anja Werno, Antje van der Linden, Arlo Upton, Chris Mansell, David Hammer, Dragana Drinkovic, Gary McAuliffe, Hana Sofia Andersson, James Ussher, Jill Sherwood, Josh Freeman, Julia Howard, Juliet Elvy, Mary DeAlmeida, Matt Blakiston, Matthew Rogers, Max Bloomfield, Michael Addidle, Michelle Balm, Sally Roberts, Sarah Jefferies, Sharmini Muttaiyah, Susan Morpeth, Susan Taylor, Timothy Blackmore, Vani Sathyendran, Veronica Playle, Virginia Hope, Erasmus Smit, Lauren Jelly, Olin Silander, Joep de Ligt |
| EPI_ISL_756165 | Department of Virology and Immunology, University of Helsinki and Helsinki University Hospital, Huslab Finland | Department of Virology, Faculty of Medicine, University of Helsinki, Helsinki, Finland | Teemu Smura, Ravi Kant, Phuoc Truong, Hussein Alburkat, Hannimari Kallio-Kokko, Jenni Virtanen, Maija Suvanto, Sari Hannula, Harri Kangas, Pekka Ellonen, Olli Vapalahti |
| EPI_ISL_756375 | Siriraj Hospital, Mahidol University | National Institute of Health, Department of Medical Sciences, Ministry of Public Health, Thailand | Pilailuk Okada; Siripaporn Phuygun; Sittiporn Parnmen; Navin Horthongkham; Ratana Tacharoenmuang; Pakorn Piromtong; Natchaya Khiadsang; Thanutsapa Thanadachakul; Warawan Wongboot; Sirikanda wimol; Sunthareeya Waicharoen; Niracha Athipanyasilp; Archiraya Pattama; Chutikarn Chaimayo; Ruengpung Sutthent; Wannee Kantakamalakul. |
| EPI_ISL_757286 | Department of Virology, Public Health Laboratories Division | Department of Virology, Public Health Laboratories Division | Massab Umair, Aamer Ikram, Muhammad Salman |
| EPI_ISL_759705 | RSUD Bung Karno, Surakarta | Universitas Sebelas Maret (UNS); Rumah Sakit Universitas Sebelas Maret (RS-UNS), Surakarta; National Institute of Health Research and Development, Indonesian Ministry of Health , Jakarta). | Betty Suryawati, Yulia Sari, Hartono, Maryani, Afif Avicena Gufron, Hana Apsari Pawestri, Kartika, Dewi Puspa, Hartanti Dian Ika, Arie Ardiansyah Nugraha, Vivi Setiawaty. |
| EPI_ISL_759724 | University of Wisconsin-Madison AIDS Vaccine Research Laboratories | University of Wisconsin-Madison AIDS Vaccine Research Laboratories | Gage Moreno, Katarina Braun, et al. AIDS Vaccine Research Laboratories |
| EPI_ISL_759811 | Department of Virology and Immunology, University of Helsinki and Helsinki University Hospital, Huslab Finland | Department of Virology, Faculty of Medicine, University of Helsinki, Helsinki, Finland | Teemu Smura, Ravi Kant, Phuoc Truong, Hussein Alburkat, Hannimari Kallio-Kokko, Jenni Virtanen, Maija Suvanto, Sari Hannula, Harri Kangas, Pekka Ellonen, Olli Vapalahti |
| EPI_ISL_760053 | Hong Kong Department of Health | School of Public Health, The University of Hong Kong | Daniel Chu, Haogao Gu, Pavithra Krishnan, Daisy Ng, Gigi Liu, Carrie Wan, Malik Peiris, Leo Poon |
| EPI_ISL_763065 | 1-Laboratory of Microbiology, National Reference Lab, Charles Nicolle Hospital; 2-University of Tunis ElManar, Faculty of Medicine of Tunis, LR99ES09, Tunis, Tunisia | 1-Clinical and Experimental Pharmacology Lab, LR16SP02, National Center of Pharmacovigilance, University of Tunis El Manar, Tunis, Tunisia. 2-Neurodegenerative diseases and psychiatric troubles, LR18SP03, Razi Hospital, University of Tunis El Manar, Tunis, Tunisia. 3- Ministry of Health, National Observatory of New and Emerging Diseases, 1006, Tunis, Tunisia | Ilhem Boutiba-Ben Boubaker, Sameh Trabelsi, Nissaf Ben Alaya, Maher Kharrat, Alia Ben Kahla, Jalila Ben Khelil, Salma Abid, Sana Ferjani, Mouna Ben Sassi, Mouna Safer, Awatef El Moussi, Habiba Ben Romdhane, Souissi Amira, Ines Mdini, Hanen El Jebari, Asma Ferjani, Gaies Emna, Riadh Daghfous, Riadh Gouider. |
| EPI_ISL_765704 | Massachusetts General Hospital | Infectious Disease Program, Broad Institute of Harvard and | Lemieux,J.E., Siddle,K.J., Shaw,B., Adams,G., Pierce,V., Turbett,S., Anahtar,M., Branda,J., Slater,D., Harris,J., Lin,A.E., Gladden-Young,A., Lagerborg,K., |

| | | | |
|---|---|---|---|
| | | MIT | Rudy,M., DeRuff,K., Carter,A., Normandin,E., Bauer,M., Reilly,S., Tomkins-Tinch,C., Loreth,C., Chaluvadi,S., Neumann,A., Cusick,C., Chapman,S.B., Gnirke,A., Flowers,K., Cerrato,F., Birren,B.W., Gallagher,G., Smole,S., Park,D.J., MacInnis,B.L., Ryan,E., LaRocque,R., Rosenberg,E. and Sabeti,P.C. |
| EPI_ISL_766046 | RSUP Fatmawati | Eijkman Institute for Molecular Biology, Ministry of Research and Technology/National Agency for Research and Innovation | Frilasita A Yudhaputri, Edison Johar, Hidayat Trimarsanto, David H Muljono, Safarina G Malik, Khin Saw Myint, Amin Soebandrio |
| EPI_ISL_766569 | Oman-National Influenza Center | Oman-National Influenza Center | Samiha Al-Kharusi, Laila Al-Balushi, Hamida Al-Barwani, Aisha Al-Busaidi, Intisar Al-Shukri, Samira Al-Mahruqi, Hanan Al-Kindi, Amina Al-Jardani |
| EPI_ISL_766718 | A05 Biomedicum | The Public Health Agency of Sweden | Department of Microbiology, The Public Health Agency of Sweden |
| EPI_ISL_766864, EPI_ISL_766866 | NIC Viral Respiratory Unit - Institut Pasteur of Algeria | National Reference Center for Viruses of Respiratory Infections, Institut Pasteur, Paris | Mélanie Albert, Marion Barbet, Sylvie Behillil, Méline Bizard, Angela Brisebarre, Flora Donati, Etienne Simon-Lorière, Vincent Enouf, Maud Vanpeene, Sylvie van der Werf, Fawzi Derrar |
| EPI_ISL_767880 | Laverty Pathology | NSW Health Pathology - Institute of Clinical Pathology and Medical Research; Westmead Hospital; University of Sydney | CIDM-PH et al. |
| EPI_ISL_767896 | Australian Clinical Labs | NSW Health Pathology - Institute of Clinical Pathology and Medical Research; Westmead Hospital; University of Sydney | CIDM-PH et al. |
| EPI_ISL_767899 | Laverty Pathology | NSW Health Pathology - Institute of Clinical Pathology and Medical Research; Westmead Hospital; University of Sydney | CIDM-PH et al. |
| EPI_ISL_767901, EPI_ISL_767904 | Sydney South West Pathology Service (SSWPS) - Liverpool Hospital - NSW Health Pathology | NSW Health Pathology - Institute of Clinical Pathology and Medical Research; Westmead Hospital; University of Sydney | CIDM-PH et al. |
| EPI_ISL_767906, EPI_ISL_767909 | Sydney South West Pathology Service (SSWPS) - Royal Prince Alfred Hospital - NSW Health Pathology | NSW Health Pathology - Institute of Clinical Pathology and Medical Research; Westmead Hospital; University of Sydney | CIDM-PH et al. |
| EPI_ISL_767912 | Sydney South West Pathology Service (SSWPS) - Liverpool Hospital - NSW Health Pathology | NSW Health Pathology - Institute of Clinical Pathology and Medical Research; Westmead Hospital; University of Sydney | CIDM-PH et al. |
| EPI_ISL_767922 | South Eastern Area Laboratory Services (SEALS) | NSW Health Pathology - Institute of Clinical Pathology and Medical Research; Westmead Hospital; University of Sydney | CIDM-PH et al. |
| EPI_ISL_767932 | Southern. IML Pathology | NSW Health Pathology - Institute of Clinical Pathology and Medical Research; Westmead Hospital; University of Sydney | CIDM-PH et al. |
| EPI_ISL_768324 | Singapore General Hospital | Department of Microbiology | Nurdyana Abdul Rahman, Kun Lee Lim, Chenhao Li, Sui Sin Goh, Kenneth Xin Long Chan, Kian Sing Chan, Lynette Oon, Kern Rei Chng, Niranjan Nagarajan, Karrie Ko |
| EPI_ISL_768620, EPI_ISL_768622, EPI_ISL_768631 | National Public Health Laboratory, National Centre for Infectious Diseases | National Public Health Laboratory, National Centre for Infectious Diseases | Tze Minn Mak, Sophie Octavia, Zhenyang Zhou, Lin Cui, Raymond Tzer Pin Lin |
| EPI_ISL_769994 | Area De Salud Catedral Noreste | Inciensa, Instituto Costarricense de Investigación y Enseñanza en Nutrición y Salud | Francisco Duarte, Hebleen Porras, Claudio Soto-Garita, Estela Cordero, Adriana Godínez, Melany Calderón & Mariel López |
| EPI_ISL_770002 | Area De Salud La Cruz | Inciensa, Instituto Costarricense de Investigación y Enseñanza en Nutrición y Salud | Francisco Duarte, Hebleen Porras, Claudio Soto-Garita, Estela Cordero, Adriana Godínez, Melany Calderón & Mariel López |
| EPI_ISL_770018 | Laboratorio Cenahce | Inciensa, Instituto Costarricense de Investigación y Enseñanza en Nutrición y Salud | Francisco Duarte, Hebleen Porras, Claudio Soto-Garita, Estela Cordero, Adriana Godínez, Melany Calderón & Andrés Feoli-Grant |
| EPI_ISL_770023 | Area De Salud San Francisco-San Antonio (Coopesana) | Inciensa, Instituto Costarricense de Investigación y Enseñanza en Nutrición y Salud | Francisco Duarte, Hebleen Porras, Claudio Soto-Garita, Estela Cordero, Adriana Godínez, Melany Calderón & Mariel López |
| EPI_ISL_770026 | Area De Salud Catedral Noreste | Inciensa, Instituto Costarricense de Investigación y Enseñanza en Nutrición y Salud | Francisco Duarte, Hebleen Porras, Claudio Soto-Garita, Estela Cordero, Adriana Godínez, Melany Calderón & Mariel López |
| EPI_ISL_770470 | National Health laboratory | Botswana Institute for Technology Research and Innovation | Kefentse Arnold Tumedi, Madisa Mine, Dineo Emang Tshiamo. Gape Nyepetsi, Thongbotho Mphoyakgosi, Malebogo Kebabonye, Maitshwarelo Ignatius Matsheka |
| EPI_ISL_770481 | Wyoming Public Health Laboratory | Wyoming Public Health Laboratory | Noah Hull, Taylor Fearing, Lynette Gumbleton, Channing Weber, Ashley Norberg, Bailey Bowcutt, and Wanda Manley |
| EPI_ISL_770902, EPI_ISL_771092, EPI_ISL_771139 | Laboratoire national de santé, Microbiology, Virology | Laboratoire national de santé, Microbiology, Microbial Genomics Platform | Anke Wienecke-Baldacchino, Catherine Ragimbeau,Jessica Tapp, Fatu Djabi, Lise Pignon, Raoul Salmon, Tamir Abdelrahman |
| EPI_ISL_771368 | SA Pathology | SA Pathology | Lex Leong, Julien Soubrier, Chuan Kok Lim, Song Gao, Mark Turra, Karin Kassahn, Ivan Bastian, Geoff Higgins |
| EPI_ISL_774901 | Designated Reference Institute for Chemical Measurements (DRICM) | DNA SOLUTION LTD. | Md. Imran Khan, Kazi Nadim Hasan, Abu Sufian, Jannatun Naima, Abdul Khaleque, Mizanur Rahman, MSM Chowdhury, Hasan Ul Haider, Mamudul Hasan Razu, Mala Khan, Mohammad Fazle Alam Rabbi |
| EPI_ISL_775019 | Gonoshasthya-RNA Molecular Research Center | Gonoshasthya-RNA Molecular Research Center | Nihad Adnan, Mohd. Raeed Jamiruddin, Md. Ahsanul Haq, Mohib Ullah Khondoker, Nafisa Azmuda, Firoz Ahmed, Shahana Sharmin, Salma Akter, Taslin Jahan Mou, Mahfuza Marzan, Sayeda Moriam Liza, Nowshin Jahan, Tamanna Ali, Maha Jamiruddin, Mousumi Chaity, Shahad Saif Khandker, Mumtarin Jannat Oishee |
| EPI_ISL_775219, EPI_ISL_775220 | Laboratoire Biolife | Laboratoire de Biotechnologie | Mouna Ouadghiri, Tarik Aanniz, Mohammed Walid Chemao Elfihri, Mohamed Chenaoui, Hanae Dakka, Afaf Alaoui, Otmane Touzani, Amina Benouda, Bouchra Belfquih, Lahcen belyamani, Saaid Amzazi and Azeddine Ibrahimi |
| EPI_ISL_779256, EPI_ISL_779267 | Jamil-ur-Rahman Center for Genome Research, Dr. Panjwani Center for Molecular Medicine and Drug Research | Jamil-ur-Rahman Center for Genome Research, Dr. Panjwani Center for Molecular Medicine and Drug Research | Shakeel,M., Irfan,M., Nisa,Z., Rashid,M., Ansari,S., Khan,I. |
| EPI_ISL_779402 | South Eastern Area Laboratory Services (SEALS) | NSW Health Pathology - Institute of Clinical Pathology and Medical Research; Westmead Hospital; University of Sydney | CIDM-PH et al. |
| EPI_ISL_779408, EPI_ISL_779409, EPI_ISL_779410, EPI_ISL_779413 | Royal Darwin Hospital Pathology | MDU-PHL | Meumann, E., Caly L., Seemann T., Sait, M.L., Druce J., Sherry, N.L. |
| EPI_ISL_779628 | Microbiological Diagnostic Unit - Public Health Laboratory (MDU-PHL) | MDU-PHL | Seemann T., Sait, M.L., Sherry, N.L. |
| EPI_ISL_779633 | Victorian Infectious Diseases Reference Laboratory (VIDRL) | VIDRL and MDU-PHL | Caly L., Seemann T., Sait, M.L., Druce J., Sherry, N.L. |
| EPI_ISL_779642, EPI_ISL_779643, EPI_ISL_779647 | Microbiological Diagnostic Unit - Public Health Laboratory (MDU-PHL) | MDU-PHL | Seemann T., Sait, M.L., Sherry, N.L. |
| EPI_ISL_779649 | Victorian Infectious Diseases Reference Laboratory (VIDRL) | VIDRL and MDU-PHL | Caly L., Seemann T., Sait, M.L., Druce J., Sherry, N.L. |
| EPI_ISL_779651 | Institute of Virology, Biomedical Research Center of the Slovak Academy of Sciences, Bratislava | Faculty of Natural Sciences, Comenius University, Bratislava | Viktória abanová, Kristína Boršová, Broa Brejová, Viktória Hodorová, Sabina Fumaová Havlíková, Juraj Kopáek, Martina Liková, ubomíra Lukáiková, Martina Neboháová, Monika Sláviková, Andrej Belák, Tomáš Vina, Jozef Nosek, Boris Klempa |
| EPI_ISL_779652 | Institute of Virology, Biomedical Research Center of the Slovak Academy of Sciences, Bratislava | Faculty of Natural Sciences, Comenius University, Bratislava | Kristína Boršová, Viktória abanová, Broa Brejová, Viktória Hodorová, Sabina Fumaová Havlíková, Juraj Kopáek, Martina Liková, ubomíra Lukáiková, Martina Neboháová, Monika Sláviková, Andrej Belák, Tomáš Vina, Boris Klempa, Jozef Nosek |
| EPI_ISL_779653 | Institute of Virology, Biomedical Research Center of the Slovak Academy of Sciences, Bratislava | Faculty of Natural Sciences, Comenius University, Bratislava | Viktória abanová, Kristína Boršová, Broa Brejová, Viktória Hodorová, Sabina Fumaová Havlíková, Juraj Kopáek, Martina Liková, ubomíra Lukáiková, Martina Neboháová, Monika Sláviková, Andrej Belák, Tomáš Vina, Jozef Nosek, Boris Klempa |

| | | | |
|---|---|---|---|
| EPI_ISL_779654 | Institute of Virology, Biomedical Research Center of the Slovak Academy of Sciences, Bratislava | Faculty of Natural Sciences, Comenius University, Bratislava | Kristína Boršová, Viktória abanová, Broa Brejová, Viktória Hodorová, Sabina Fumaová Havlíková, Juraj Kopáek, Martina Liková, ubomíra Lukáiková, Martina Neboháová, Monika Sláviková, Andrej Belák, Tomáš Vina, Boris Klempa, Jozef Nosek |
| EPI_ISL_779659 | Institute of Virology, Biomedical Research Center of the Slovak Academy of Sciences, Bratislava | Faculty of Natural Sciences, Comenius University, Bratislava | Broa Brejová, Viktória abanová, Kristína Boršová, Viktória Hodorová, Sabina Fumaová Havlíková, Juraj Kopáek, Martina Liková, ubomíra Lukáiková, Martina Neboháová, Monika Sláviková, Andrej Belák, Tomáš Vina, Jozef Nosek, Boris Klempa |
| EPI_ISL_779660 | Institute of Virology, Biomedical Research Center of the Slovak Academy of Sciences, Bratislava | Faculty of Natural Sciences, Comenius University, Bratislava | Viktória abanová, Kristína Boršová, Broa Brejová, Viktória Hodorová, Sabina Fumaová Havlíková, Juraj Kopáek, Martina Liková, ubomíra Lukáiková, Martina Neboháová, Monika Sláviková, Andrej Belák, Tomáš Vina, Jozef Nosek, Boris Klempa |
| EPI_ISL_788937 | Centre de Recherche et de Formation en Infectiologie Guinée | TransVIHMI, IRD/INSERM/Monpellier University | Alpha Kabinet KEITA, Abdoul Karim SOUMAH, Abdoulaye TOURE, Moriba POVOGUI, Joel KOIVOGUI, Jean-louis MONEMOU, Mamadou Saliou SOW, Penda Malhado DIALLO, Mamadou Bhoye KEITA, Alimou CAMARA, Kaba KOUROUMA, Mandiou DIAKITE, Mamadou Saliou BAH, Sakoba KEITA, Bouna |
| EPI_ISL_788938 | Centre de Recherche et de Formation en Infectiologie Guinée | TransVIHMI, IRD/INSERM/Monpellier University | Alpha Kabinet KEITA, Abdoul Karim SOUMAH, Abdoulaye TOURE, Moriba POVOGUI, Joel KOIVOGUI, Jean-louis MONEMOU, Mamadou Saliou SOW, Penda Malhado DIALLO, Mamadou Bhoye KEITA, Alimou CAMARA, Kaba KOUROUMA, Mandiou DIAKITE, Mamadou Saliou BAH, Sakoba KEITA, Bouna Yatassaye, Christelle BUTEL, Laetitia SERRANO, Ahidjo AYOUBA, Eric DELAPORTE, Martine PEETERS |
| EPI_ISL_789054 | Unilabs AB | The Public Health Agency of Sweden | Department of Microbiology, The Public Health Agency of Sweden |
| EPI_ISL_791088 | Instituto Nacional de Salud - Unidad de Secuenciación y Análisis Genómico | Instituto Nacional de Salud - Dirección de Investigación en Salud Pública | Katherine Laiton-Donato, Diego A. Álvarez-Díaz, Carlos Franco-Muñoz, Mauricio Pacheco-Montealegre, Jonathan Reales, Sheryl Corchuelo, Maria T. Herrera, Julian Naizaque, Gerardo Santamaría, Paola Muñoz-Laiton, Diego Andrés Prada, Magdalena Wiesner, Martha Lucia Ospina Martinez, Marcela Mercado-Reyes |
| EPI_ISL_791947 | Massachusetts General Hospital | Infectious Disease Program, Broad Institute of Harvard and MIT | Lemieux,J.E., Siddle,K.J., Shaw,B., Adams,G., Pierce,V., Turbett,S., Anahtar,M., Branda,J., Slater,D., Harris,J., Lin,A.E., Gladden-Young,A., Lagerborg,K., Rudy,M., DeRuff,K., Carter,A., Normandin,E., Bauer,M., Reilly,S., Tomkins-Tinch,C., Loreth,C., Chaluvadi,S., Neumann,A., Cusick,C., Chapman,S.B., Gnirke,A., Flowers,K., Cerrato,F., Birren,B.W., Gallagher,G., Smole,S., Park,D.J., MacInnis,B.L., Ryan,E., LaRocque,R., Rosenberg,E. and Sabeti,P.C. |
| EPI_ISL_792125 | Laboratorio de Virología del Hospital de Niños Dr. Ricardo Gutierrez | Área de Secuenciación del Laboratorio de Virología del Hospital de Niños Dr. Ricardo Gutierrez on behalf of 'Proyecto Argentino Interinstitucional de genomica de SARS-CoV-2' (PAIS Consortium) | Nabaes Jodar, MS; Goya, S; Natale, MI; Lusso, S; Gravis, E; Acevedo, ME; Alvarez Lopez, C; Alexay, S; Jacques, O; Mistchenko, AS; Valinotto, LE; Viegas, M. |
| EPI_ISL_792388 | Plataforma de Servicios Biotecnólogicos: UTTIPP/PSB , Universidad Nacional de Quilmes. | Área de Secuenciación del Laboratorio de Virología del Hospital de Niños Dr. Ricardo Gutierrez on behalf of 'Proyecto Argentino Interinstitucional de genomica de SARS-CoV-2' (PAIS Consortium) | Nabaes Jodar, MS; Goya, S; Natale, MI; Lusso, S; Farina, H; Goñi, S; Cardama, G; Castello, A; Valinotto, LE; Viegas, M. |
| EPI_ISL_792495 | Laboratorio Central Mg. Luis Alfredo Pianciola | Hospital Regional Ushuaia - Centro Austral De Investigaciones Científicas - Universidad Nacional De Tierra Del Fuego on behalf of 'Proyecto Argentino Interinstitucional de genomica de SARS-CoV-2' (PAIS Consortium) | Ceballos, SG; Nardi, CF; Gramundi, ID; Gallego, F; Pinto, C; Ziehm, MC; Mazzeo, M; Pianciola, L. |
| EPI_ISL_792512 | Laboratorio Central de la Ciudad de Santa Fe | Grupo de Genómica y Bioinformática del Instituto de Investigación de la Cadena Láctea CONICET-INTA on behalf of 'Proyecto Argentino Interinstitucional de genomica de SARS-CoV-2' (PAIS Consortium) | Eberhardt, MF; Irazoqui, JM; Ojeda, G; Rompato, G; Mugna, V; Pastor, C; Amadio, AF |
| EPI_ISL_792572 | Laboratório Central de Saúde Pública do Estado da Paraíba (LACEN-PB) | Laboratory of Respiratory Viruses and Measles, Oswaldo Cruz Institute, FIOCRUZ | Paola Resende, Luciana Appolinario, Fernando Motta, Anna Carolina Paixao, Ana Carolina Mendonca, João Felipe Bezerra, Romero Henrique Teixeira de Vasconcelos, Dalane Loudal Florentino Teixeira, Thiago Franco de Oliveira Carneiro, Marilda Siqueira on behalf of the Fiocruz COVID-19 Genomic Surveillance Network |
| EPI_ISL_793594 | Department of Virus and Microbiological Special Diagnostics, Statens Serum Institut, Copenhagen, Denmark | Albertsen Lab, Department of Chemistry and Bioscience, Aalborg University, Denmark | Danish Covid-19 Genome Consortium |
| EPI_ISL_794592 | Laboratorio Estatal de Salud Pública de Tamaulipas | Instituto de diagnóstico y Referencia Epidemiologicos (INDRE) | Abril Rodriguez-Maldonado, Claudia Wong-Arambula, Fabiola Garces-Ayala, Gisela Barrera-Badillo, Ana Maria Cortez-Calderon, Bernardita Reyes-Berrones, Hilda del Carmen-Selvera, Gloria Molina-Gamboa, Lucia Hernandez-Rivas, Irma Lopez-Martinez, Celia Alpuche-Aranda, Jose Luis Alomia-Zegarra, Hugo Lopez Gatell-Ramirez, Ernesto Ramirez-Gonzalez. |
| EPI_ISL_794600, EPI_ISL_794603 | Central Laboratories, Egyptian Ministry of Health and Population | Central Laboratories, Egyptian Ministry of Health and Population | Kayed,A.E., Roshdy,W.H., El-Shesheny,R., Mostafa,A., Khalifa,M.K., Shehata,M., Shawky,S., Saleh,M., Gomaa,M., El Taweel,A., Mahmoud,S.H., Moatasim,Y., Kutkat,O., Kamel,M.N., Mahrous,N., El Sayes,M.A., El Guindy,N.M., Naguib,A., Kandeil,A., Kayali,G., Ali,M.A. |
| EPI_ISL_794606 | Central Public Health Laboratories, Egyptian Ministry of Health and Population | Central Public Health Laboratories, Egyptian Ministry of Health and Population | El-Shesheny,R., Roshdy,W.H., Kayed,A.E., Mostafa,A., Khalifa,M.K., Shehata,M., Shawky,S., Saleh,M., Gomaa,M., El Taweel,A., Mahmoud,S.H., Moatasim,Y., Kutkat,O., Kamel,M.N., Mahrous,N., El Sayes,M.A., El Guindy,N.M., Naguib,A., Kandeil,A., Kayali,G., Ali,M.A. |
| EPI_ISL_794610, EPI_ISL_794614, EPI_ISL_794620, EPI_ISL_794621, EPI_ISL_794622 | LabPLUS | Institute of Environmental Science and Research (ESR) | Xiaoyun Ren, Matt Storey, Nikki Freed, Muhammad Faisal, Jing Wang, Hermes Perez, Anja Werno, Antje van der Linden, Arlo Upton, Chris Mansell, David Hammer, Dragana Drinkovic, Gary McAuliffe, Hana Sofia Andersson, James Ussher, Jill Sherwood, Josh Freeman, Julia Howard, Juliet Elvy, Mary DeAlmeida, Matt Blakiston, Matthew Rogers, Max Bloomfield, Michael Addidle, Michelle Balm, Sally Roberts, Sarah Jefferies, Sharmini Muttaiyah, Susan Morpeth, Susan Taylor, Timothy Blackmore, Vani Sathyendran, Veronica Playle, Virginia Hope, Erasmus Smit, Lauren Jelly, Olin Silander, Joep de Ligt |
| EPI_ISL_794624 | Middlemore Hospital | Institute of Environmental Science and Research (ESR) | Xiaoyun Ren, Matt Storey, Nikki Freed, Muhammad Faisal, Jing Wang, Hermes Perez, Anja Werno, Antje van der Linden, Arlo Upton, Chris Mansell, David Hammer, Dragana Drinkovic, Gary McAuliffe, Hana Sofia Andersson, James Ussher, Jill Sherwood, Josh Freeman, Julia Howard, Juliet Elvy, Mary DeAlmeida, Matt Blakiston, Matthew Rogers, Max Bloomfield, Michael Addidle, Michelle Balm, Sally Roberts, Sarah Jefferies, Sharmini Muttaiyah, Susan Morpeth, Susan Taylor, Timothy Blackmore, Vani Sathyendran, Veronica Playle, Virginia Hope, Erasmus Smit, Lauren Jelly, Olin Silander, Joep de Ligt |
| EPI_ISL_794627 | Canterbury Health Laboratories | Institute of Environmental Science and Research (ESR) | Xiaoyun Ren, Matt Storey, Nikki Freed, Muhammad Faisal, Jing Wang, Hermes Perez, Anja Werno, Antje van der Linden, Arlo Upton, Chris Mansell, David Hammer, Dragana Drinkovic, Gary McAuliffe, Hana Sofia Andersson, James Ussher, Jill Sherwood, Josh Freeman, Julia Howard, Juliet Elvy, Mary DeAlmeida, Matt Blakiston, Matthew Rogers, Max Bloomfield, Michael Addidle, Michelle Balm, Sally Roberts, Sarah Jefferies, Sharmini Muttaiyah, Susan Morpeth, Susan Taylor, Timothy Blackmore, Vani Sathyendran, Veronica Playle, Virginia Hope, Erasmus Smit, Lauren Jelly, Olin Silander, Joep de Ligt |
| EPI_ISL_794634 | Biology, MCL | Biology, MCL | Seadawy,M.G., Shamel,M.D., EL-hosieny,M.F., Gad,A.F., EL-harty,B.S. and EL-Safty,A.S. |
| EPI_ISL_794663 | Carvajal Laboratorios | Instituto Nacional de Salud - Dirección de Investigación en Salud Pública | Katherine Laiton-Donato, Diego A. Álvarez-Díaz, Carlos Franco-Muñoz, Mauricio Pacheco-Montealegre, Jonathan Reales, Sheryl Corchuelo, Maria T. Herrera, Julian Naizaque, Gerardo Santamaría, Paola Muñoz-Laiton, Diego Andrés Prada, Magdalena Wiesner, Martha Lucia Ospina Martinez, Marcela Mercado-Reyes |
| EPI_ISL_794672, EPI_ISL_794673 | PathWest Laboratory Medicine WA | PathWest Laboratory Medicine WA Microbial Surveillance Unit | PathWest Laboratory Medicine WA Microbial Surveillance Unit |
| EPI_ISL_794735, EPI_ISL_794737, EPI_ISL_794738 | 1-Laboratory of Microbiology, National Reference Lab, Charles Nicolle Hospital; 2-University of Tunis ElManar, Faculty of Medicine of Tunis, LR99ES09, Tunis, Tunisia | 1-Clinical and Experimental Pharmacology Lab, LR16SP02, National Center of Pharmacovigilance, University of Tunis El Manar, Tunis, Tunisia. 2-Neurodegenerative diseases and psychiatric troubles, LR18SP03, Razi Hospital, University of Tunis El Manar, Tunis, Tunisia. 3- Ministry of Health, National Observatory of New and Emerging Diseases, 1006, Tunis, Tunisia | Ilhem Boutiba-Ben Boubaker, Sameh Trabelsi, Nissaf Ben Alaya, Maher Kharrat, Alia BenKahla, Jalila Ben Khelil, Salma Abid, Sana Ferjani, Mouna Ben Sassi, Mouna Safer, Zaineb Hamzaoui, Guedi Ali Barreh, Habiba Ben Romdhane, Souissi Amira, Sarra Chamman, Hanen El Jebari, Asma Ferjani, Gaies Emna, Riadh Daghfous, Riadh Gouider. |
| EPI_ISL_795797 | Department of Virus and Microbiological Special Diagnostics, Statens Serum Institut, Copenhagen, Denmark | Albertsen Lab, Department of Chemistry and Bioscience, Aalborg University, Denmark | Danish Covid-19 Genome Consortium |
| EPI_ISL_796012, EPI_ISL_796018, EPI_ISL_796019, EPI_ISL_796025 | Hebei Provincial Center for Disease Control and Prevention, Shijiazhuang, Hebei Province; National Institute for Viral Disease Control and Prevention, China CDC | Hebei Provincial Center for Disease Control and Prevention, Shijiazhuang, Hebei Province; National Institute for Viral Disease Control and Prevention, China CDC | Shunxiang Qi, Xiang Zhao, Nankun Liu, George F. Gao, Yang Song, Wenbo Xu, Qi Li |

| | | | |
|---|---|---|---|
| EPI_ISL_796782 | Laboratory of Microbiology, National Reference Lab, Charles Nicolle Hospital; 2-University of Tunis ElManar, Faculty of Medicine of Tunis, LR99ES09, Tunis, Tunisia | Clinical and Experimental Pharmacology Lab, LR16SP02, National Center of Pharmacovigilance, University of Tunis El Manar, Tunis, Tunisia. 2-Neurodegenerative diseases and psychiatric troubles, LR18SP03, Razi Hospital, University of Tunis El Manar, Tunis, Tunisia. 3- Ministry of Health, National Observatory of New and Emerging Diseases, 1006, Tunis, Tunisia | Ilhem Boutiba-Ben Boubaker, Sameh Trabelsi, Nissaf Ben Alaya, Maher Kharrat, Alia BenKahla, Jalila Ben Khelil, Salma Abid, Sana Ferjani, Mouna Ben Sassi, Mouna Safer, Zaineb Hamzaoui, Guedi Ali Barreh, Habiba Ben Romdhane, Souissi Amira, Sarra Chamman, Hanen El Jebari, Ahmed Fakhfakh, Gaies Emna, Riadh Daghfous, Riadh Gouider. |
| EPI_ISL_801566 | Laboratory of Molecular Virology, Pontificia Universidad Católica de Chile | MSHS Pathogen Surveillance Program | Leonardo I. Almonacid, Ana S. Gonzalez-Reiche, Matthew M. Hernandez, Jorge Levican, Tamara García-Salum, Zenab Khan, Adriana van De Guchte, Ajay Obla, Jayeeta Dutta, Bremy Alburquerque, Eileen Serrano, Erick Salinas, Manuel Ampuero, Hala Alshammary, Juan Soto, Shwetha Hara Sridhar, Ying-Chih Wang, Melissa Smith, Robert Sebra, Fernando Valiente, Aldo Gaggero, Adolfo Garcia-Sastre, Edward C. Holmes, Viviana Simon, Harm van Bakel, Rafael A. Medina. |
| EPI_ISL_801702, EPI_ISL_801751, EPI_ISL_801767 | Laboratory of Molecular Virology, Pontificia Universidad Católica de Chile | MSHS Pathogen Surveillance Program | Leonardo I. Almonacid, Ana S. Gonzalez-Reiche, Matthew M. Hernandez, Jorge Levican, Ana Maria Contreras, Carlos Palma, Tamara García-Salum, Zenab Khan, Adriana van De Guchte, Ajay Obla, Jayeeta Dutta, Bremy Alburquerque, Eileen Serrano, Constanza Maldonado, M. Belen Leyton, Erick Salinas, Hala Alshammary, Juan Soto, Shwetha Hara Sridhar, Ying-Chih Wang, Melissa Smith, Robert Sebra, Marcela Ferres, Adolfo Garcia-Sastre, Edward C. Holmes, Viviana Simon, Harm van Bakel, Rafael A. Medina. |
| EPI_ISL_803430, EPI_ISL_803851 | Laboratory of Microbiology, National Reference Lab, Charles Nicolle Hospital; 2-University of Tunis ElManar, Faculty of Medicine of Tunis, LR99ES09, Tunis, Tunisia | Clinical and Experimental Pharmacology Lab, LR16SP02, National Center of Pharmacovigilance, University of Tunis El Manar, Tunis, Tunisia. 2-Neurodegenerative diseases and psychiatric troubles, LR18SP03, Razi Hospital, University of Tunis El Manar, Tunis, Tunisia. 3- Ministry of Health, National Observatory of New and Emerging Diseases, 1006, Tunis, Tunisia | Ilhem Boutiba-Ben Boubaker, Sameh Trabelsi, Nissaf Ben Alaya, Maher Kharrat, Alia BenKahla, Jalila Ben Khelil, Salma Abid, Sana Ferjani, Mouna Ben Sassi, Mouna Safer, Zaineb Hamzaoui, Habiba Ben Romdhane, Souissi Amira, Sarra Chamman, Hanen El Jebari, Ahmed Fakhfakh, Gaies Emna, Riadh Daghfous, Riadh Gouider. |
| EPI_ISL_803961, EPI_ISL_803987 | National Public Health Laboratory, National Centre for Infectious Diseases | National Public Health Laboratory, National Centre for Infectious Diseases | Tze Minn Mak, Sophie Octavia, Zhenyang Zhou, Lin Cui, Raymond Tzer Pin Lin |
| EPI_ISL_804030 | Maryland Public Health Laboratory | Maryland Public Health Laboratory | Maryland Department of Health Laboratories Administration |
| EPI_ISL_804054 | SC (UCO) Igiene e Sanità Pubblica (funzione integrata con SC Microbiologia e Virologia) e Laboratory of Molecular Virology of the International Centre for Genetic Engineering and Biotechnology (ICGEB) | ARGO Laboratorio Genomica ed Epigenomica | Licastro D, Dal Monego S, Degasperi M, Marcello A, D'Agaro P |
| EPI_ISL_804067 | Israel Central Virology laboratory | Israel National Consortium for SARS-CoV-2 sequencing | Neta Zuckerman, Efrat Dahan Bucris, Michal Mandelboim, Dana Bar-Ilan, Oran Erster, Tzvia Mann, Omer Murik, David A. Zeevi, Assaf Rokney, Joseph Jaffe, Eva Nachum, Maya Davidovich Cohen, Ephraim Fass, Gal Zizelski Valenci, Mor Rubinstein, Efrat Rorman, Israel Nissan, Efrat Glick-Saar, Omri Nayshool, Gideon Rechavi, Ella Mendelson, Orna Mor |
| EPI_ISL_804507 | MEPHI, Aix Marseille University | MEPHI, Aix Marseille University | Anthony LEVASSEUR |
| EPI_ISL_804863 | DC Public Health Lab/ Dept. of Forensic Sciences | DC Public Health Lab/ Dept. of Forensic Sciences | Scott Nguyen, Elizabeth Zelaya, Connie Maza, Monica Mann, Brittany Hamilton, David Payne, Jocelyn Hauser |
| EPI_ISL_806625, EPI_ISL_806698 | KEMRI-Wellcome Trust Research Programme/KEMRI-CGMR-C Kilifi | KEMRI-Wellcome Trust Research Programme/KEMRI-CGMR-C Kilifi | Githinji et al |
| EPI_ISL_806719 | Pathology North - Royal North Shore Hospital - NSW Health Pathology | NSW Health Pathology - Institute of Clinical Pathology and Medical Research; Westmead Hospital; University of Sydney | CIDM-PH et al. |
| EPI_ISL_806721 | Histopath | NSW Health Pathology - Institute of Clinical Pathology and Medical Research; Westmead Hospital; University of Sydney | CIDM-PH et al. |
| EPI_ISL_806729 | Sydney South West Pathology Service (SSWPS) - Royal Prince Alfred Hospital - NSW Health Pathology | NSW Health Pathology - Institute of Clinical Pathology and Medical Research; Westmead Hospital; University of Sydney | CIDM-PH et al. |
| EPI_ISL_807154, EPI_ISL_807156 | Deva County Emergency Hospital | National Institute of Infectious Diseases-Prof. Dr. Matei Bals Molecular Diagnostics Laboratory | Leontina Banica, Marius Surleac, Corina Casangiu, Petre Milu, Andreea Tudor, Simona Paraschiv, Dan Otelea |
| EPI_ISL_810801, EPI_ISL_810804, EPI_ISL_810965, EPI_ISL_810969 | PathWest Laboratory Medicine WA | PathWest Laboratory Medicine WA Microbial Surveillance Unit | PathWest Laboratory Medicine WA Microbial Surveillance Unit |
| EPI_ISL_812045 | Lighthouse Lab in Milton Keynes | Wellcome Sanger Institute for the COVID-19 Genomics UK (COG-UK) Consortium | The Lighthouse Lab in Milton Keynes and Alex Alderton, Roberto Amato, Sonia Goncalves, Ewan Harrison, David K. Jackson, Ian Johnston, Dominic Kwiatkowski, Cordelia Langford, John Sillitoe on behalf of the Wellcome Sanger Institute COVID-19 Surveillance Team |
| EPI_ISL_812248 | North Dakota Department of Health, Public Health Laboratory | North Dakota Department of Health, Public Health Laboratory | Lisa Wingerter |
| EPI_ISL_812266 | Landstuhl Regional Medical Center | United States Air Force School of Aerospace Medicine | Anthony Fries, Jennifer Meyer, Amanda Javorina, Sarah Purves, William Gruner, Clarise Starr, Elizabeth Macias, Fritz Castillo, Cole Anderson |
| EPI_ISL_812300 | Delaware Public Health Lab | Delaware Public Health Lab | Gregory Hovan |
| EPI_ISL_812423 | Royal Darwin Hospital Pathology | MDU-PHL | Meumann, E., Caly L., Seemann T., Sait, M.L., Druce J., Sherry, N.L. |
| EPI_ISL_812424, EPI_ISL_812426, EPI_ISL_812428, EPI_ISL_812430, EPI_ISL_812431, EPI_ISL_812432 | Microbiological Diagnostic Unit - Public Health Laboratory (MDU-PHL) | MDU-PHL | Seemann T., Sait, M.L., Sherry, N.L. |
| EPI_ISL_812437 | Victorian Infectious Diseases Reference Laboratory (VIDRL) | VIDRL and MDU-PHL | Caly L., Seemann T., Sait, M.L., Druce J., Sherry, N.L. |
| EPI_ISL_812513 | Laboratorio de Referencia Nacional de Virus Respiratorios, Instituto Nacional de Salud Peru | Laboratorio de Genómica Microbiana, Universidad Peruana Cayetano Heredia | Pablo Tsukayama, Alejandra Dávila-Barclay, Guillermo Salvatierra, Luis González, Pedro E. Romero, Brenda Ayzanoa, Janet Huancachoque, Pool Marcos, Camila Castillo-Vilcahuamán, Oscar Escalante, Priscila Lope, Nancy Rojas |
| EPI_ISL_812521 | Pathology North - Royal North Shore Hospital - NSW Health Pathology | NSW Health Pathology - Institute of Clinical Pathology and Medical Research; Westmead Hospital; University of Sydney | CIDM-PH et al. |
| EPI_ISL_812561, EPI_ISL_812648, EPI_ISL_812698 | United States Air Force School of Aerospace Medicine | United States Air Force School of Aerospace Medicine | Anthony Fries, Jennifer Meyer, Amanda Javorina, Sarah Purves, William Gruner, Clarise Starr, Elizabeth Macias |
| EPI_ISL_812766, EPI_ISL_812767, EPI_ISL_812775, EPI_ISL_812780 | Ministry of Health Turkey | Ministry of Health Turkey | Fatma Bayrakdar, Yasemin Cogun, Süleyman Yalcin, Aye Baak Alta, Gülay Korukluolu |
| EPI_ISL_812787, EPI_ISL_812790, EPI_ISL_812814, EPI_ISL_812828 | Genomics Program, Children Cancer Hospital | Genomics Program, Children Cancer Hospital | Hatem,A., Hadad,A., Abouelnaga,S., Amer,K., Salah,H., Farawyla,H., Halafawy,A., Mansour,T., shalaby,L., Hassan,W., Soliman,M., Gomaa,C., Hassan,R., Soliman,S., Monuir,G., Hammad,M., Hussein,S., Abdo,I., Jalal,D., El-Zayat,M., El-Shaqnqery,H., Diab,A., Bakry,U., Samir,O., Magdeldin,S., Sayed,A. |
| EPI_ISL_812876 | Ministry of Health Turkey | Ministry of Health Turkey | Fatma Bayrakdar, Yasemin Cogun, Süleyman Yalcin, Aye Baak Alta, Gülay Korukluolu |
| EPI_ISL_812922 | Pasteur Institute in Ho Chi Minh city | Department of Microbiology and Immunology - Pasteur Institute in Ho Chi Minh city | Manh Huy Dao, Hang Thi Thu Pham, Nhung Pham Hong Vu, Hieu Minh Le, Thang Minh Cao, Loan Thi Kim Huynh, Long Thanh Nguyen, Anh Hoang Nguyen, Hieu Trung Nguyen, Thao Thi Ngoc Nguyen, Thinh Viet Nguyen, Quang Duy Pham, Quang Chan Luong, Lan Trong Phan, Thuong Vu Nguyen. |
| EPI_ISL_812933, EPI_ISL_812935, EPI_ISL_812936, EPI_ISL_812938, EPI_ISL_812942 | Institute for Urban Disease Control and Prevention | COVID-19 Network Investigations (CONI) Alliance | Kamolthip Atsawawaranunt, Elizabeth Batty, Wasun Chantratita, Thanat Chookajorn, Stefan Fernandez, Angkana Huang, Anthony R. Jones, Khajohn Joonsalak, Chonticha Klungtong, Theerarat Kochakarn, Prayuth Kaewmalang, Amornmas Kongklieng, Namfon Kotanan, Krittikorn Kumpornsin, Duangkamon Loesbanluechai, Wudtichai Manasatienkij, Anek Mungaomklang, Bhakbhoom Panthan, Pukkaporn Parnwijitkul, Ekawat Pasomsub, Kingkan |

| | | | |
|---|---|---|---|
| | | | Rakmanee, Insee Sensorn, Janjira Thaipadungpanit, Arporn Wangwiwatsin, Treewat Watthanachockchai |
| EPI_ISL_812967 | University Clinical Research Center, University of Sciences | University Clinical Research Center, University of Sciences | Diarra,B., Kone,A., Guindo,I., Bane,S., Diakite,M., Dao,S., Iknane,A.A., Doumbia,S. |
| EPI_ISL_813173 | Department of Pathology, University of Cambridge | COVID-19 Genomics UK (COG-UK) Consortium | Aminu S. Jahun, Yasmin Chaudhry, Grant Hall, Iliana Georgana, Myra Hosmillo, Martin D. Curran, Malte Pinckert, Surendra Parmar, Ian Goodfellow |
| EPI_ISL_813636 | University of Exeter | COVID-19 Genomics UK (COG-UK) Consortium | Ben Temperton,Aaron Jeffries,Michelle Michelsen,Joanna Warwick-Dugdale,Audrey Farbos,Robyn Manley,Stephen Michell,Jane Masoli |
| EPI_ISL_813997 | Hospital General Universitario Gregorio Marañón | SeqCOVID-SPAIN consortium/IBV(CSIC) | Darío García de Viedma, Laura Pérez-Lago, Marta Herranz, Jon Sicilia, Julia Suárez, Pilar Catalán, Patricia Muñoz and SeqCOVID-SPAIN consortium |
| EPI_ISL_815297 | Centogene | Centogene | Peter Bauer, Krishna Kumar Kandaswamy, Vivi Hue-Trang Lieu |
| EPI_ISL_824286, EPI_ISL_824287 | Institute of Microbiology, Universidad San Francisco de Quito | Institute of Microbiology, Universidad San Francisco de Quito | Belén Prado-Vivar, Sully Márquez, Juan José Guadalupe, Monica Becerra-Wong, Bernardo Gutiérrez, Andrea Cunguan, Nabih Dahik, Verónica Barragán, Patricio Rojas-Silva, Gabriel Trueba, Michelle Grunauer, Paúl Cárdenas |
| EPI_ISL_824288, EPI_ISL_824289 | Institute of Microbiology, Universidad San Francisco de Quito | Institute of Microbiology, Universidad San Francisco de Quito | Belén Prado-Vivar, Sully Márquez, Juan José Guadalupe, Monica Becerra-Wong, Bernardo Gutiérrez, Tania Guayasamin, Patricio Reyes, Verónica Barragán, Patricio Rojas-Silva, Gabriel Trueba, Michelle Grunauer, Paúl Cárdenas |
| EPI_ISL_824290, EPI_ISL_824291 | Institute of Microbiology, Universidad San Francisco de Quito | Institute of Microbiology, Universidad San Francisco de Quito | Belén Prado-Vivar, Sully Márquez, Juan José Guadalupe, Monica Becerra-Wong, Bernardo Gutiérrez, Andrea Cunguan, Nabih Dahik, Verónica Barragán, Patricio Rojas-Silva, Gabriel Trueba, Michelle Grunauer, Paúl Cárdenas |
| EPI_ISL_824292 | Institute of Microbiology, Universidad San Francisco de Quito | Institute of Microbiology, Universidad San Francisco de Quito | Belén Prado-Vivar, Sully Márquez, Juan José Guadalupe, Monica Becerra-Wong, Bernardo Gutiérrez, Tania Guayasamin, Patricio Reyes, Verónica Barragán, Patricio Rojas-Silva, Gabriel Trueba, Michelle Grunauer, Paúl Cárdenas |
| EPI_ISL_824293, EPI_ISL_824294 | Institute of Microbiology, Universidad San Francisco de Quito | Institute of Microbiology, Universidad San Francisco de Quito | Belén Prado-Vivar, Sully Márquez, Juan José Guadalupe, Monica Becerra-Wong, Bernardo Gutiérrez, Eulalia Pazmiño, Katalina Pacheco, Verónica Barragán, Patricio Rojas-Silva, Gabriel Trueba, Michelle Grunauer, Paúl Cárdenas |
| EPI_ISL_824814 | Department of Clinical Microbiology | GIGA Medical Genomics | Keith Durkin, Maria Artesi, Sébastien Bontems, Raphaël Boreux, Bouchra Boujemla, Cécile Meex, Pierrette Melin, Marie-Pierre Hayette, Vincent Bours |
| EPI_ISL_825075 | National Public Health Laboratory, National Centre for Infectious Diseases | National Public Health Laboratory, National Centre for Infectious Diseases | Tze Minn Mak, Sophie Octavia, Zhenyang Zhou, Lin Cui, Raymond Tzer Pin Lin |
| EPI_ISL_827031, EPI_ISL_827168, EPI_ISL_827358 | deCODE genetics | deCODE genetics | Daniel F Gudbjartsson; Agnar Helgason; Hakon Jonsson; Olafur T Magnusson; Pall Melsted; Gudmundur L Norddahl; Jona Saemundsdottir; Asgeir Sigurdsson; Patrick Sulem; Arna B Agustsdottir; Hannes Eggertsson; Berglind Eiriksdottir; Run Fridriksdottir; Elisabet E Gardarsdottir; Gudmundur Georgsson; Olafia S Gretarsdottir; Kjartan R Gudmundsson; Thora R Gunnarsdottir; Arnaldur Gylfason; Hilma Holm; Brynjar O Jensson; Aslaug Jonasdottir; Kamilla S Josefsdottir; Thordur Kristjansson; Droplaug N Magnusdottir; Solvi Rognvaldsson; Louise le Roux; Gudrun Sigmundsdottir; Gardar Sveinbjornsson; Kristin E Sveinsdottir; Maney Sveinsdottir; Emil A Thorarensen; Bjarni Thorbjornsson; Gisli Masson; Ingileif Jonsdottir; Alma Moller; Thorolfur Gudnason; Karl G Kristinsson; Unnur Thorsteinsdottir; Kari Stefansson |
| EPI_ISL_827383 | The National University Hospital of Iceland | deCODE genetics | Daniel F Gudbjartsson; Agnar Helgason; Hakon Jonsson; Olafur T Magnusson; Pall Melsted; Gudmundur L Norddahl; Jona Saemundsdottir; Asgeir Sigurdsson; Patrick Sulem; Arna B Agustsdottir; Hannes Eggertsson; Berglind Eiriksdottir; Run Fridriksdottir; Elisabet E Gardarsdottir; Gudmundur Georgsson; Olafia S Gretarsdottir; Kjartan R Gudmundsson; Thora R Gunnarsdottir; Arnaldur Gylfason; Hilma Holm; Brynjar O Jensson; Aslaug Jonasdottir; Kamilla S Josefsdottir; Thordur Kristjansson; Droplaug N Magnusdottir; Solvi Rognvaldsson; Louise le Roux; Gudrun Sigmundsdottir; Gardar Sveinbjornsson; Kristin E Sveinsdottir; Maney Sveinsdottir; Emil A Thorarensen; Bjarni Thorbjornsson; Gisli Masson; Ingileif Jonsdottir; Alma Moller; Thorolfur Gudnason; Karl G Kristinsson; Unnur Thorsteinsdottir; Kari Stefansson |
| EPI_ISL_828255, EPI_ISL_828256 | deCODE genetics | deCODE genetics | Daniel F Gudbjartsson; Agnar Helgason; Hakon Jonsson; Olafur T Magnusson; Pall Melsted; Gudmundur L Norddahl; Jona Saemundsdottir; Asgeir Sigurdsson; Patrick Sulem; Arna B Agustsdottir; Hannes Eggertsson; Berglind Eiriksdottir; Run Fridriksdottir; Elisabet E Gardarsdottir; Gudmundur Georgsson; Olafia S Gretarsdottir; Kjartan R Gudmundsson; Thora R Gunnarsdottir; Arnaldur Gylfason; Hilma Holm; Brynjar O Jensson; Aslaug Jonasdottir; Kamilla S Josefsdottir; Thordur Kristjansson; Droplaug N Magnusdottir; Solvi Rognvaldsson; Louise le Roux; Gudrun Sigmundsdottir; Gardar Sveinbjornsson; Kristin E Sveinsdottir; Maney Sveinsdottir; Emil A Thorarensen; Bjarni Thorbjornsson; Gisli Masson; Ingileif Jonsdottir; Alma Moller; Thorolfur Gudnason; Karl G Kristinsson; Unnur Thorsteinsdottir; Kari Stefansson |
| EPI_ISL_831775, EPI_ISL_831794 | United States Air Force School of Aerospace Medicine | United States Air Force School of Aerospace Medicine | Anthony Fries, Jennifer Meyer, William Gruner, Amanda Javorina, Sarah Purves, Clarise Starr, Elizabeth Macias |
| EPI_ISL_831938 | Laboratório de Microbiologia Molecular - Universidade FEEVALE | Universidade Federal de Ciências da Saúde de Porto Alegre | Vinícius Bonetti Franceschi, Amanda de Menezes Mayer, Gabriel Dickin Caldana, Carla Andretta Moreira Neves, Patrícia Aline Gröhs Ferrareze, Gabriela Bettella Cybis, Ricardo Ariel Zimerman, Lívia Kmetzsch, Fernando Rosado Spilki, Claudia Elizabeth Thompson |
| EPI_ISL_832919 | Maine HETL | Tewhey Lab, The Jackson Laboratory | Matluk,N., Dewey,H., Iosue,F., Barter,M., Lynch,R., Munger,H. and Tewhey,R. |
| EPI_ISL_833041 | Defence Services Medical Research Center, Biological Research Laboratory | Defence Services Medical Research Center, Biological Research Laboratory | Khine Zaw Oo, Nay Myo Aung, Ko Ko Win, Phyo Kyaw Aung, Zaw Win Htun, Sat Paing Htoo, Kyaw Wanna, Thein Zaw, Kyee Myint, Ko Ko Lwin |
| EPI_ISL_833210, EPI_ISL_833211, EPI_ISL_833219 | Department of Virology and Immunology, University of Helsinki and Helsinki University Hospital, Huslab Finland | Department of Virology, Faculty of Medicine, University of Helsinki, Helsinki, Finland | Teemu Smura, Ravi Kant, Phuoc Truong, Hussein Alburkat, Hannimari Kallio-Kokko, Jenni Virtanen, Maija Suvanto, Fathiah Zakham, Essi Korhonen, Sari Hannula, Harri Kangas, Pekka Ellonen, Olli Vapalahti |
| EPI_ISL_833381 | National Public Health Laboratory, National Centre for Infectious Diseases | National Public Health Laboratory, National Centre for Infectious Diseases | Tze Minn Mak, Sophie Octavia, Zhenyang Zhou, Lin Cui, Raymond Tzer Pin Lin |
| EPI_ISL_833499 | RS Qadr, Tangerang, Indonesia | Biosafety Level-3 Laboratory, Indonesian Institute of Sciences (LIPI) | Isa Nuryana, Ade Andriani, Ahmad Fathoni, Anik Budhi Dharmayanthi, Syam Budi Iryanto, Andri Wardiana, Anggia Prasetyoputri, Ratna Dwi Ramadani, Sugiyono Saputra, Ratih Asmana Ningrum |
| EPI_ISL_837299 | Istituto Zooprofilattico Sperimentale del Mezzogiorno | TIGEM | Antonio Grimaldi, Patrizia Annunziata, Francesco Panariello, Biancamaria Pierri, Valentina Bouche, Chiara Colantuono, Maria Concetta Cuomo, Denise Di Concilio, Lucio Di Filippo, Anna Manfredi, Marcello Salvi, Antonio Limone, Pellegrino Cerino, Andrea Ballabio, Davide Cacchiarelli. |
| EPI_ISL_837394 | National Virus Reference Laboratory | National Virus Reference Laboratory | Michael Carr, Gabriel Gonzalez, Jonathan Dean, Cillian F De Gascun |
| EPI_ISL_837556, EPI_ISL_837557, EPI_ISL_837558, EPI_ISL_837560, EPI_ISL_837563, EPI_ISL_837572, EPI_ISL_837573, EPI_ISL_837575, EPI_ISL_837576, EPI_ISL_837577, EPI_ISL_837578 | | | |
| see above | Centro Nacional de Enfermedades Tropicales (CENETROP) | Laboratory of Respiratory Viruses and Measles, Oswaldo Cruz Institute, FIOCRUZ | Paola Resende, Roxana Loayza, Cinthia Avila, Luciana Appolinario, Fernando Motta, Anna Carolina Paixao, Ana Carolina Mendonca, Marilda Siqueira on behalf of the Fiocruz COVID-19 Genomic Surveillance Network |
| EPI_ISL_843199 | Canterbury Health Laboratories | Institute of Environmental Science and Research (ESR) | Xiaoyun Ren, Matt Storey, Nikki Freed, Muhammad Faisal, Jing Wang, Hermes Perez, Anja Werno, Antje van der Linden, Arlo Upton, Chris Mansell, David Hammer, Dragana Drinkovic, Gary McAuliffe, Hana Sofia Andersson, James Ussher, Jill Sherwood, Josh Freeman, Julia Howard, Juliet Elvy, Mary DeAlmeida, Matt Blakiston, Matthew Rogers, Max Bloomfield, Michael Addidle, Michelle Balm, Sally Roberts, Sarah Jefferies, Sharmini Muttaiyah, Susan Morpeth, Susan Taylor, Timothy Blackmore, Vani Sathyendran, Veronica Playle, Virginia Hope, Erasmus Smit, Lauren Jelly, Olin Silander, Joep de Ligt |
| EPI_ISL_843607, EPI_ISL_844001 | Department of Virus and Microbiological Special Diagnostics, Statens Serum Institut, Copenhagen, Denmark | Albertsen Lab, Department of Chemistry and Bioscience, Aalborg University, Denmark | Danish Covid-19 Genome Consortium |
| EPI_ISL_845546 | National Public Health Laboratory, Cameroon | African Centre of Excellence for Genomics of Infectious Diseases (ACEGID), Redeemer's University | Oluniyi P.E. et al |
| EPI_ISL_845663 | Quest Diagnostics | Quest Diagnostics | Rosenthal,S.H., Gerasimova,A., Kagan,R.M., Anderson, B., Bernstein, L.E., Livingston, K.E., Hua, M., Liu Y., Shalhout, D.F., Shlyakhter, I.A., Owen, R., Lacbawan, F. |
| EPI_ISL_847877 | University Hospitals of Geneva, Laboratory of Virology | HUG, Laboratory of Virology and the Health2030 Genome Center | Samuel Cordey, Ana Rita Goncalves, Laurent Kaiser, Lorenzo Cerutti, Henri Pegeot, Melyssa Elies, Keith Harshman, Ioannis Xenarios, Emmanouil Dermitzakis |
| EPI_ISL_848112 | National Virus Reference Laboratory | Irish Coronavirus Sequencing Consortium - National University of Ireland Galway | Kate Reddington, Grainne Mc Andrew, Simone Coughlan |
| EPI_ISL_848783 | Florida Bureau of Public Health Laboratories | Florida Bureau of Public Health Laboratories | Sarah Schmedes, Jason Blanton |
| EPI_ISL_849190, EPI_ISL_849284, | Servicio Virosis Respiratorias-Departamento Virología-INEI | Instituto Nacional Enfermedades Infecciosas C.G.Malbran | Baumeister E., Avaro M., Benedetti E., Russo M., Dattero ME, Pontoriero A., Cisterna D., Molina V., Perandones C., Tuduri E., Lorenzo F., Poklepovich T., |

| | | | |
|---|---|---|---|
| EPI_ISL_849287 | | | Campos J. |
| EPI_ISL_849644 | SIESP DIPARTIMENTO DI PREVENZIONE CHIETI | Istituto Zooprofilattico Sperimentale dell'Abruzzo e Molise "G.Caporale" | Lorusso A, Marcacci M, Di Domenico M, Curini V, Ancora M, Cammà C, Rinaldi A, Mangone I, Di Pasquale A, Puglia I, Savini G. |
| EPI_ISL_849683, EPI_ISL_849686, EPI_ISL_849693, EPI_ISL_849694, EPI_ISL_849731, EPI_ISL_849734 | Public Health Virology Laboratory, Forensic and Scientific Services (PHV-FSS) | Public Health Virology Laboratory, Forensic and Scientific Services (PHV-FSS) | Son Nguyen et al |
| EPI_ISL_849737, EPI_ISL_849739, EPI_ISL_849742 | Special Operations Medical Research Division, Defence Services Medical Research Centre | Special Operations Medical Research Division, Defence Services Medical Research Centre | Oo,K.Z., Aung,N.M., Win,K.K., Aung,P.K., Htun,Z.W., Zaw,T., Myint,K., Lwin,K.K. |
| EPI_ISL_849745, EPI_ISL_849746, EPI_ISL_849747, EPI_ISL_849748, EPI_ISL_849749, EPI_ISL_849752, EPI_ISL_849756, EPI_ISL_849761 | Public Health Virology Laboratory, Forensic and Scientific Services (PHV-FSS) | Public Health Virology Laboratory, Forensic and Scientific Services (PHV-FSS) | Son Nguyen et al |
| EPI_ISL_849852 | A. Krumbholz, Labor Dr. Krause und Kollegen MVZ GmbH, Kiel | Charité Universitätsmedizin Berlin, Institut für Virologie | Victor M Corman, Jörn Beheim-Schwarzbach, Talitha Veith, Julia Schneider, Tobias Bleicker, Julia Tesch, Barbara Mühlemann, Terry Jones, Christian Drosten |
| EPI_ISL_849895 | Utah Public Health Laboratory | Utah Public Health Laboratory | Erin L. Young, Kelly F. Oakeson, Tara Gallagher |
| EPI_ISL_850449 | Division of Emerging Infectious Diseases, Bureau of Infectious Diseases Diagnosis Control, Korea Disease Control and Prevention Agency | Division of Emerging Infectious Diseases, Bureau of Infectious Diseases Diagnosis Control, Korea Disease Control and Prevention Agency | Ae Kyung Park, Il-Hwan Kim, Heui Man Kim, Jeong-Min Kim, Namjoo Lee, Chaeyoung Lee, Sang Hee Woo, Eun-Jin Kim |
| EPI_ISL_850947, EPI_ISL_850949, EPI_ISL_850950 | National Institute for Viral Disease Control and Prevention, China CDC | National Institute for Viral Disease Control and Prevention, China CDC | Xiang Zhao, Yenan Feng, Zhixiao Chen, Yao Meng, Yuchao Wu, Yang Song, Ji Wang, Kai Nie, Yong Zhang, Yanhai Wang, Weimin Zhou, Wenjie Tan, Jun Han, Shiwen Wang, Wenbo Xu, Cao Chen, Dayan Wang |
| EPI_ISL_852826 | Microbiology Lab | NBCC Sequencing Facility | Dr. Jeff Wrana |
| EPI_ISL_854745 | Royal Darwin Hospital Pathology | MDU-PHL | Meumann, E., Caly L., Seemann T., Sait, M.L., Druce J., Sherry, N.L. |
| EPI_ISL_854748 | Victorian Infectious Diseases Reference Laboratory (VIDRL) | VIDRL and MDU-PHL | Caly L., Seemann T., Sait, M.L., Druce J., Sherry, N.L. |
| EPI_ISL_854755 | Microbiological Diagnostic Unit - Public Health Laboratory (MDU-PHL) | MDU-PHL | Seemann T., Sait, M.L., Sherry, N.L. |
| EPI_ISL_854762 | Victorian Infectious Diseases Reference Laboratory (VIDRL) | VIDRL and MDU-PHL | Caly L., Seemann T., Sait, M.L., Druce J., Sherry, N.L. |
| EPI_ISL_855763 | Department of Virus and Microbiological Special Diagnostics, Statens Serum Institut, Copenhagen, Denmark | Aalborg University | Danish Covid-19 Genome Consortium |
| EPI_ISL_856757 | Servicio Virosis Respiratorias-Departamento Virología-INEI | Instituto Nacional Enfermedades Infecciosas C.G.Malbran | Baumeister E., Avaro M., Benedetti E., Russo M., Dattero ME, Pontoriero A., Cisterna D., Molina V., Perandones C., Tuduri E., Lorenzo F., Poklepovich T., Campos J. |
| EPI_ISL_857092 | DOHMH Riverside | New York City Public Health Laboratory | Jade Wang, et al. |
| EPI_ISL_857309 | Medlab Pathology | NSW Health Pathology - Institute of Clinical Pathology and Medical Research; Westmead Hospital; University of Sydney | CIDM-PH et al. |
| EPI_ISL_857312 | South Eastern Area Laboratory Services (SEALS) | NSW Health Pathology - Institute of Clinical Pathology and Medical Research; Westmead Hospital; University of Sydney | CIDM-PH et al. |
| EPI_ISL_857317, EPI_ISL_857319, EPI_ISL_857320, EPI_ISL_857328, EPI_ISL_857346 | Cancer Biology Department, National Cancer Institute | Cancer Biology Department, National Cancer Institute | Zekri,A.N., Sedawy,M.G., Ahmed,O.S., Hafez,M.M., Soliman,H.K., Bahnassy,A.A., Elhosieny,F.W., Gad,A.E., Hamdy,M.S., Soliman,M.S., Soliman,L., Abouelhoda,M. |
| EPI_ISL_857424 | Maine HETL | Tewhey Lab, The Jackson Laboratory | Matluk,N., Dewey,H., Iosue,F., Barter,M., Lynch,R., Munger,H. and Tewhey,R. |
| EPI_ISL_857468 | PathWest Laboratory Medicine WA | PathWest Laboratory Medicine WA Microbial Surveillance Unit | PathWest Laboratory Medicine WA Microbial Surveillance Unit |
| EPI_ISL_859623, EPI_ISL_859691, EPI_ISL_859772, EPI_ISL_859838, EPI_ISL_859997, EPI_ISL_860033, EPI_ISL_860039, EPI_ISL_860044, EPI_ISL_860059, EPI_ISL_860091 | BTC, Khalifa University | BTC, Khalifa University | Al Safar et al |
| EPI_ISL_860235 | Oslo University Hospital, Department of Medical Microbiology | Norwegian Institute of Public Health, Department of Virology | Kathrine Stene-Johansen, Kamilla Heddeland Instefjord, Hilde Elshaug, Atiya R Ali,Marie Paulsen Madsen, Rasmus Riis Kopperud, Hilde Vollan, Karoline Bragstad, Olav Hungnes |
| EPI_ISL_860600 | NHLS-IALCH | KRISP, KZn Research Innovation and Sequencing Platform | Giandhari J, Pillay S, Lessells R, Mdlalose K, York D, Khan S, Tegally H, Wilkinson E, de Oliveira T |
| EPI_ISL_860802 | WHO/Minsk | Charité Universitätsmedizin Berlin, Institut für Virologie | Victor M Corman, Barbara Mühlemann, Jörn Beheim-Schwarzbach, Talitha Veith, Julia Tesch, Tobias Bleicker, Julia Schneider, Shmialiova Natallia, Sivets Natallia, Terry Jones, Christian Drosten |
| EPI_ISL_860910 | CH du MANS - Lab. Bio. Moléculaire | National Reference Center for Viruses of Respiratory Infections, Institut Pasteur, Paris | Marion Barbet, Sylvie Behillil, Méline Bizard, Angela Brisebarre, Camille Capel, Etienne Simon-Lorière, Vincent Enouf, Maud Vanpeene, Sylvie van der Werf,Ramanantsoa CéLine |
| EPI_ISL_861457 | WHO/Minsk | Charité Universitätsmedizin Berlin, Institut für Virologie | Victor M Corman, Barbara Mühlemann, Jörn Beheim-Schwarzbach, Talitha Veith, Julia Tesch, Tobias Bleicker, Julia Schneider, Shmialiova Natallia, Sivets Natallia, Terry Jones, Christian Drosten |
| EPI_ISL_861906 | LATE - Laboratório de Técnicas Especiais - Hospital Israelita Albert Einstein | LATE - Laboratório de Técnicas Especiais - Hospital Israelita Albert Einstein | Deyvid Amgarten, Fernanda de Mello Malta, Raquel Riyuzo, Ana Paula Moreira Salles, Pedro Henrique Sebe Rodrigues, João Renato Rebello Pinho |
| EPI_ISL_862039 | RSUP Dr. Sardjito | Genetics Working Group (Pokja Genetik) Faculty of Medicine, Public Health and Nursing Universitas Gadjah Mada (FK-KMK UGM); Disease Investigation Center Wates Ministry of Agriculture Indonesia; Department of Microbiology FK-KMK UGM; Laboratorium Diagnostik Yayasan Tahija World Mosquito Program (WMP) Yogyakarta Center for Tropical Medicine FK-KMK UGM; Integrated Research Center FK-KMK UGM; Department of Computer Science and Electronics FMIPA UGM; RSUP Dr. Sardjito | Gunadi, Hendra Wibawa, . Marcellus, Mohamad S. Hakim, Edwin W. Daniwijaya, Ludhang P. Rizki, Endah Supriyati, Eggi Arguni, Titik Nuryastuti, Tri Wibawa, Dwi AA Nugrahaningsih, Afiahayati, Siswanto, Kristy Iskandar, Nungki Anggorowati, Ika Trisnawati, Sumardi, Eko Budiono, Bambang Sigit Riyanto, Heni Retnowulan, Munawar Gani, Satria Maulana, Nur Rahmi Ananda, Riat El Khair, Yunika Puspadewi, Osman Sianipar, Umi Solekhah Intansari, Elizabeth Henny Herningtiyas, Ira Puspitawati, Nur Imma Fatimah Harahap,William Widitjiarso, Maria Patricia Inggriani |
| EPI_ISL_862040 | RSUP Dr. Sardjito | Genetics Working Group (Pokja Genetik) Faculty of Medicine, Public Health and Nursing Universitas Gadjah Mada (FK-KMK UGM); Disease Investigation Center Wates Ministry of Agriculture Indonesia; Department of Microbiology FK-KMK UGM; Laboratorium Diagnostik Yayasan Tahija World Mosquito Program (WMP) Yogyakarta Center for Tropical | Gunadi, Hendra Wibawa, . Marcellus, Mohamad S. Hakim, Edwin W. Daniwijaya, Ludhang P. Rizki, Endah Supriyati, Eggi Arguni, Titik Nuryastuti, Tri Wibawa, Dwi AA Nugrahaningsih, Afiahayati, Siswanto, Kristy Iskandar, Nungki Anggorowati, Ika Trisnawati, Sumardi, Eko Budiono, Bambang Sigit Riyanto, Heni Retnowulan, Munawar Gani, Satria Maulana, Nur Rahmi Ananda, Riat El Khair, Yunika Puspadewi, Osman Sianipar, Umi Solekhah Intansari, Elizabeth Henny Herningtiyas, Ira Puspitawati, Nur Imma Fatimah Harahap,Alvin Santoso Kalim, Untung Riawan |

| | | | |
|---|---|---|---|
| | | Medicine FK-KMK UGM; Integrated Research Center FK-KMK UGM; Department of Computer Science and Electronics FMIPA UGM; RSUP Dr. Sardjito | |
| EPI_ISL_862041 | RSUP Dr. Sardjito | Genetics Working Group (Pokja Genetik) Faculty of Medicine, Public Health and Nursing Universitas Gadjah Mada (FK-KMK UGM); Disease Investigation Center Wates Ministry of Agriculture Indonesia; Department of Microbiology FK-KMK UGM; Laboratorium Diagnostik Yayasan Tahija World Mosquito Program (WMP) Yogyakarta Center for Tropical Medicine FK-KMK UGM; Integrated Research Center FK-KMK UGM; Department of Computer Science and Electronics FMIPA UGM; RSUP Dr. Sardjito | Gunadi, Hendra Wibawa, . Marcellus, Mohamad S. Hakim, Edwin W. Daniwijaya, Ludhang P. Rizki, Endah Supriyati, Eggi Arguni, Titik Nuryastuti, Tri Wibawa, Dwi AA Nugrahaningsih, Afiahayati, Siswanto, Kristy Iskandar, Nungki Anggorowati, Ika Trisnawati, Sumardi, Eko Budiono, Bambang Sigit Riyanto, Heni Retnowulan, Munawar Gani, Satria Maulana, Nur Rahmi Ananda, Riat El Khair, Yunika Puspadewi, Osman Sianipar, Umi Solekhah Intansari, Elizabeth Henny Herningtiyas, Ira Puspitawati, Nur Imma Fatimah Harahap, Dyah Ayu Puspitarani, Audric Kenny Tedja |
| EPI_ISL_862072 | Department of Virology and Immunology, University of Helsinki and Helsinki University Hospital, Huslab Finland | Department of Virology, Faculty of Medicine, University of Helsinki, Helsinki, Finland | Teemu Smura, Ravi Kant, Phuoc Truong, Hussein Alburkat, Hanna Liimatainen, Hannimari Kallio-Kokko, Jenni Virtanen, Maija Suvanto, Sari Hannula, Harri Kangas, Pekka Ellonen, Olli Vapalahti |
| EPI_ISL_862075 | National Influenza Center, Virology Department | National Influenza Center | V Salimi,NZ Shafiei Jandaghi, J Yavarian, A Nejati, K Sadeghi, N Ghavvami,F Ajaminejad and T Mokhtari Azad |
| EPI_ISL_862078 | National Influenza Center, Virology Department | National Influenza Center | A Nejati, J Yavarian,K Sadeghi, NZ Shafiei Jandaghi, V Salimi, N Ghavvami,F Ajaminejad and T Mokhtari Azad |
| EPI_ISL_862079 | National Influenza Center, Virology Department | National Influenza Center | K Sadeghi, A Nejati, J Yavarian, NZ Shafiei Jandaghi, V Salimi, F Ajaminejad,N Ghavvami and T Mokhtari Azad |
| EPI_ISL_862094 | Department of Virology and Immunology, University of Helsinki and Helsinki University Hospital, Huslab Finland | Department of Virology, Faculty of Medicine, University of Helsinki, Helsinki, Finland | Teemu Smura, Ravi Kant, Phuoc Truong, Hussein Alburkat, Hanna Liimatainen, Hannimari Kallio-Kokko, Jenni Virtanen, Maija Suvanto, Sari Hannula, Harri Kangas, Pekka Ellonen, Olli Vapalahti |
| EPI_ISL_862311, EPI_ISL_862415 | Kurnool Medical College (KMC) | CSIR Institute of Genomics and Integrative Biology | Pallavali Roja Rani, Mohamed Imran, J. Vijaya Lakshmi, Bani Jolly, S. Afsar, Abhinav Jain, Mohit Kumar Divakar, Panyam Suresh, Disha Sharma, Nambi Rajesh, Rahul C Bhoyar, Dasari Ankaiah, Sanaga Shanthi Kumari, Gyan Ranjan, Valluri Anitha Lavanya, Mercy Rophina, S. Umadevi, Paras Sehgal, Avula Renuka Devi, A. Surekha, Pulala Chandra, Rajamadugu Hymavathy, P R Vanaja, Vinod Scaria, Sridhar Sivasubbu |
| EPI_ISL_862812 | Cancer Biology Department, National Cancer Institute | Cancer Biology Department, National Cancer Institute | Zekri,A.N., Sedawy,M.G., Ahmed,O.S., Hafez,M.M., Soliman,H.K., Bahnassy,A.A., Elhosieny,F.W., Gad,A.E., Hamdy,M.S., Soliman,M.S., Soliman,L., Abouelhoda,M. |
| EPI_ISL_862832, EPI_ISL_862834, EPI_ISL_862835 | National Institute of Infectious Diseases-Prof. Dr. Matei Bals Molecular Diagnostics Laboratory | National Institute of Infectious Diseases-Prof. Dr. Matei Bals Molecular Diagnostics Laboratory | Leontina Banica, Marius Surleac, Corina Casangiu, Petre Milu, Andreea Tudor, Simona Paraschiv, Dan Otelea |
| EPI_ISL_869241 | Suceava County Emergency Hospital | "Stefan cel Mare" University Metagenomics Laboratory | Lobiuc Andrei, Puscaselu Roxana |
| EPI_ISL_871505 | Department of Virus and Microbiological Special Diagnostics, Statens Serum Institut, Copenhagen, Denmark | Aalborg University | Danish Covid-19 Genome Consortium |
| EPI_ISL_871870 | Wyoming Public Health Laboratory | Wyoming Public Health Laboratory | Noah Hull, Taylor Fearing, Lynette Gumbleton, Channing Weber, Ashley Norberg, Bailey Bowcutt, and Wanda Manley |
| EPI_ISL_871908 | Servicio de Microbiología Clínica (Complejo Hospitalario de Navarra, Pamplona), Instituto de Investigación Sanitaria de Navarra (IdiSNA) | SeqCOVID-SPAIN consortium/IBV(CSIC) | Carmen Ezpeleta Baquedano, Ana Navascués, Ana Miqueleiz and SeqCOVID-SPAIN consortium |
| EPI_ISL_872025 | Ospedale "Di Venere" | Beaconlab (Bioinformatics, Evolution and Comparative Genomics lab), Dept of Biosciences, University on Milan | Iacobellis M, d'Avenia M, Piluscio R, Parisi A, Chiara M, Manzari C, Pesole G |
| EPI_ISL_872040 | Department of Clinical Microbiology | GIGA Medical Genomics | Keith Durkin, Maria Artesi, Sébastien Bontems, Raphaël Boreux, Bouchra Boujemla, Cécile Meex, Pierrette Melin, Marie-Pierre Hayette, Vincent Bours |
| EPI_ISL_872103 | Chu Tivoli | GIGA Medical Genomics | Keith Durkin, Maria Artesi, Sébastien Bontems, Raphaël Boreux, Bouchra Boujemla, Cécile Meex, Pierrette Melin, Marie-Pierre Hayette, Vincent Bours |
| EPI_ISL_872194 | Caribbean Public Health Agency | Carrington Lab, Department of PreClinical Sciences, Faculty of Medical Sciences, The University of the West Indies | Nikita S. D. Sahadeo, Arianne Brown-Jordan, Vernie Ramkissoon, Sarah Hill, Naresh Nandram, Avery Hinds, Jerome Foster, Stanley Giddings, Karla Georges, Marsha Ivey, Rahul Naidu, Risha Singh, SueMin Nathaniel, Rajini Haraksingh, Jaya Jayaraman, Chinna Chinnadurai, Adesh Ramsubhag, Nuno Faria, Oliver Pybus, Christopher Oura, Gabriel Escobar, Christine V. F. Carrington |
| EPI_ISL_872323 | Labo Analyses Med | National Reference Center for Viruses of Respiratory Infections, Institut Pasteur, Paris | Marion Barbet, Sylvie Behillil, Méline Bizard, Angela Brisebarre, Camille Capel, Etienne Simon-Lorière, Vincent Enouf, Maud Vanpeene, Sylvie van der Werf,Rouah Raquel |
| EPI_ISL_872534 | New Mexico Department of Health Scientific Laboratory | Center for Global Health, University of New Mexico Health Sciences Center | Daryl Domman, Kurt Schwalm, Twila Kunde, Joseph Hicks, Anastacia Griego, Michael Edwards, Darrell Dinwiddie |
| EPI_ISL_872584 | South Eastern Area Laboratory Services (SEALS) | NSW Health Pathology - Institute of Clinical Pathology and Medical Research; Westmead Hospital; University of Sydney | CIDM-PH et al. |
| EPI_ISL_872586 | Sydney South West Pathology Service (SSWPS) - Royal Prince Alfred Hospital - NSW Health Pathology | NSW Health Pathology - Institute of Clinical Pathology and Medical Research; Westmead Hospital; University of Sydney | CIDM-PH et al. |
| EPI_ISL_872604, EPI_ISL_872626 | Nigeria Centre for Disease Control (NCDC) | African Centre of Excellence for Genomics of Infectious Diseases (ACEGID), Redeemer's University | Oluniyi P.E. et al |
| EPI_ISL_872681 | Rhode Island Department of Health | Infectious Disease Program, Broad Institute of Harvard and MIT | Lemieux,J.E., Siddle,K.J., Huard,R., King,E., Azevedo,K., Miller,A., Adams,G., Gladden-Young,A., Lagerborg,K., Rudy,M., DeRuff,K., Carter,A., Normandin,E., Bauer,M., Reilly,S., Tomkins-Tinch,C., Loreth,C., Chaluvadi,S., Birren,B.W., Gallagher,G., Smole,S., Park,D.J., MacInnis,B.L., and Sabeti,P.C. |
| EPI_ISL_872984 | HELIX LLC | WHO National Influenza Centre Russian Federation | Andrey Komissarov, Artem Fadeev, Anna Ivanova, Kseniya Komissarova, Dmitry Bazhenov, Mikhail Bakaev, Daria Danilenko, Ksenia Safina, Elena Nabieva, Georgii Bazykin, Dmitry Lioznov |
| EPI_ISL_873164 | Medical Laboratory Sciences, Arab American University | Medical Laboratory Sciences, Arab American University | Dumaidi,k., Al-Jawabreh,A., Ereqat,S., Al-Jawabreh,H., Nasereddin,A. |
| EPI_ISL_875975, EPI_ISL_876011 | Laboratory of Molecular Biology, Diagnostyka sp. z o.o. | genXone SA, Research & Development Laboratory | Maciej Sykulski, Grzegorz Nowicki, Monika Makowska-Woniak, Jakub Grabowski, Natalia Drwska-Matelska, ukasz Krych, Micha Kaszuba |
| EPI_ISL_877127 | Academic Hospital of Gajdah Mada University (RSA UGM) | Genetics Working Group (Pokja Genetik) Faculty of Medicine, Public Health and Nursing Universitas Gadjah Mada (FK-KMK UGM); Disease Investigation Center Wates Ministry of Agriculture Indonesia; Department of Microbiology FK-KMK UGM; Laboratorium Diagnostik Yayasan Tahija World Mosquito Program (WMP) Yogyakarta Center for Tropical Medicine FK-KMK UGM; Integrated Research center FK-KMK UGM; Department of Computer Science and Electronics FMIPA UGM | Gunadi, Hendra Wibawa, Marcellus, Mohamad S. Hakim, Edwin W. Daniwijaya, Ludhang P. Rizki, Endah Supriyati, Eggi Arguni, Titik Nuryastuti, Tri Wibawa, Dwi AA Nugrahaningsih, Afiahayati, Siswanto,Audric Kenny Tedja, Maria Patricia Inggriani |
| EPI_ISL_877206 | LabPLUS | Institute of Environmental Science and Research (ESR) | Xiaoyun Ren, Matt Storey, Nikki Freed, Muhammad Faisal, Jing Wang, Hermes Perez, Anja Werno, Antje van der Linden, Arlo Upton, Chris Mansell, David Hammer, Dragana Drinkovic, Gary McAuliffe, Hana Sofia Andersson, James Ussher, Jill Sherwood, Josh Freeman, Julia Howard, Juliet Elvy, Mary DeAlmeida, Matt Blakiston, Matthew Rogers, Max Bloomfield, Michael Addidle, Michelle Balm, Sally Roberts, Sarah Jefferies, Sharmini Muttaiyah, Susan Morpeth, Susan Taylor, Timothy Blackmore, Vani Sathyendran, Veronica Playle, Virginia Hope, Erasmus Smit, Lauren Jelly, Olin Silander, Joep de Ligt |
| EPI_ISL_877557, EPI_ISL_877558, EPI_ISL_877560 | Institute of Microbiology, Universidad San Francisco de Quito | Institute of Microbiology, Universidad San Francisco de Quito | Belén Prado-Vivar, Sully Márquez, Juan José Guadalupe, Monica Becerra-Wong, Bernardo Gutiérrez, Eulalia Pazmiño, Katalina Pacheco, Verónica Barragán, Patricio Rojas-Silva, Gabriel Trueba, Michelle Grunauer, Paúl Cárdenas |
| EPI_ISL_877562 | Institute of Microbiology, Universidad San Francisco de Quito | Institute of Microbiology, Universidad San Francisco de Quito | Belén Prado-Vivar, Sully Márquez, Juan José Guadalupe, Monica Becerra-Wong, Bernardo Gutiérrez, Piedad Villacis, Rosa Avila, Adriana Arnao, David |

| | | | |
|---|---|---|---|
| | | | Acosta-España, Alvaro Davalos de Castro, Verónica Barragán, Patricio Rojas-Silva, Gabriel Trueba, Michelle Grunauer, Paúl Cárdenas |
| EPI_ISL_878443, EPI_ISL_878463, EPI_ISL_878571 | Biolab Diagnostic Laboratories | Andersen lab at Scripps Research | Issa Abu-Dayyeh, Ahmad Tibi, Lama Hussein, Lina Mohammad, Zein Naber, Amid Abdelnour with SEARCH Alliance San Diego |
| EPI_ISL_882637 | Azerbaijan National Hematology Center Division of Medical Genetics | Azerbaijan National Hematology Center Division of Medical Genetics | Aghayev Agha Rza, Bayraml Ramin |
| EPI_ISL_882642, EPI_ISL_882643 | Azerbaijan National Hematology Center Division of Medical Genetics | Azerbaijan National Hematology Center Division of Medical Genetics | Aghayev Agha Rza |
| EPI_ISL_882663 | LACEN do Distrito Federal | Instituto Adolfo Lutz, Interdiciplinary Procedures Center, Strategic Laboratory | Claudio Tavares Sacchi, Claudia Regina Gonçalves, Erica Valessa Ramos Gomes, Karoline Rodrigues Campos |
| EPI_ISL_882768 | 1.AO Universitaria 'S. Giovanni di Dio e Ruggi D'Aragona, Scuola Medica Salernitana' Hospital / 2.UOC di Virologia e Microbiologia, Università della Campania 'L. Vanvitelli' / 3.AO Universitaria 'Federico II' Napoli Hospital / 4.AORN 'San Giuseppe Moscati' Avellino Hospital / 5.AO 'San Pio - presidio G. Rummo' Benevento Hospital / 6.AO 'Sant'Anna e San Sebastiano' Caserta Hospital / 7.PO 'Maria Santissima Addolorata' Eboli Hospital / 8.Biogem Istituto di Ricerche Genetiche | 1. Genome Research Center for Health (CRGS) / 2. Laboratory of Molecular Medicine and Genomics(LMMGe) / 3. Center for Research in Pure and Applied Mathematics (CRMPA) | Giorgio Giurato, Francesca Rizzo, Alessandro Weisz, Gianluigi Franci, Giovanni Nassa, Pasquale Pagliano, Roberta Tarallo, Elena Alexandrova, Ylenia D'Agostino, Carlo Ferravante, Jessica Lamberti, Viola Melone, Domenico Memoli, Valeria Mirici Cappa, Domenico Palumbo, Giovanni Pecoraro, Assunta Sellitto, Oriana Strianese, Ilaria Terenzi, Giuseppe Fenza, Aniello Gentile, Antonello Saccomanno, Sonia Amabile, Teresa Rocco, Annamaria Salvati, Emilia Vaccaro, Massimiliano Galdiero, Michele Cennamo, Giuseppe Portella, Maria Grazia Foti, Mariarosaria Ingino, Maria Landi, Maurizio Fumi, Vincenzo Rocco, Rita Greco, Vittoria Letizia, Arnolfo Petruzziello, Maddalena Schioppa, Gregorio Goffredi, Francesca Marciano, Michele Caraglia, Alessia Cossu, Marianna Scrima |
| EPI_ISL_882771 | Siti Khodijah Hospital | Institute of Tropical Disease, Universitas Airlangga | Kazufumi Shimizu, Krisnoadi Rahardjo, Aldise M Nastri, Jezzy R Dewantari, Rima R Prasetya, Muhammad Hamdan, Gatot Soegiarto, Laksmi Wulandari, Resti Yudhawati, Yasuko Mori, Soetjipto, Maria I Lusida |
| EPI_ISL_882776, EPI_ISL_882793 | Institute for Urban Disease Control and Prevention | COVID-19 Network Investigations (CONI) Alliance | Kamolthip Atsawawaranunt, Elizabeth Batty, Wasun Chantratita, Thanat Chookajorn, Stefan Fernandez, Angkana Huang, Anthony R. Jones, Khajohn Joonsalak, Chonticha Klungtong, Theerarat Kochakarn, Prayuth Kaewmalang, Amornmas Kongklieng, Namfon Kotanan, Krittikorn Kumpornsin, Duangkamon Loesbanluechai, Wudtichai Manasatienkij, Anek Mungaomklang, Bhakbhoom Panthan, Pukkaporn Parnwijitkul, Ekawat Pasomsub, Vichan Pawun, Kingkan Rakmanee, Insee Sensorn, Janjira Thaipadungpanit, Arporn Wangwiwatsin, Treewat Watthanachockchai |
| EPI_ISL_882943 | The National Institute of Public Health | State Veterinary Institute Prague | Nagy,A;Jirincova,H;Vecerova,JTrnka,D |
| EPI_ISL_882955 | State Veterinary Institute Prague | State Veterinary Institute Prague | Nagy,A;Cernikova,L;Stara,M |
| EPI_ISL_882960 | Laboratorium Diagnostyi Mikrobiologicznej z Pracowni Prtka Grulicy SPSzW im. Jana Boego w Lublinie | National Institute of Public Health - National Institute of Hygiene | Wokowicz Tomasz, Zacharczuk Katarzyna, Gawor Jan |
| EPI_ISL_883065 | Kansas Health and Environmental Lab | Kansas Health and Environmental Lab | Mike Grose, Carissa Robertson, Ben Olsen, and Phil Adam |
| EPI_ISL_884096 | Eurofins Diatherix | Hudsonalpha Genome Sequencing Center | Jane Grimwood, Melissa Williams, Lori H. Handley, Joshua Stough, Leslie Malone, Stefan Brzezinski, Ada Stewart, Teresa Jones, Jenell Webber, John Lovell, Jennifer Cart, and Jeremy Schmutz |
| EPI_ISL_884240 | Kansas Health and Environmental Lab | Kansas Health and Environmental Lab | Mike Grose, Paige Drury, Carissa Robertson, Ben Olsen, and Phil Adam |
| EPI_ISL_884262, EPI_ISL_884267 | Institute of Medical Microbiology and Hospital Hygiene | Institute of Medical Microbiology and Hospital Hygiene | Prof. Dr. Achim Kaasch, Aljoscha Tersteegen |
| EPI_ISL_884350, EPI_ISL_884401 | Infectious Diseases, Quest Diagnostics | Infectious Diseases, Quest Diagnostics | Rosenthal,S.H., Gerasimova,A., Kagan,R.M., Anderson,B., Bernstein,L.E., Livingston,K.E., Hua,M., Liu,Y., Shalhout,D.F., Owen,R., Lacbawan,F. |
| EPI_ISL_884846 | Department of Biochemistry, Cell and Molecular Biology, West African Centre for Cell Biology of Infectious Pathogens (WACCBIP), University of Ghana | Department of Biochemistry, Cell and Molecular Biology, West African Centre for Cell Biology of Infectious Pathogens (WACCBIP), University of Ghana | Ngoi,J.M., Tei-Maya,F., Morang'a,C.M., Magnussen,V., Amuzu,D.S., Mohammed,A., Tapela,K., Kibinge,N., Diallo,A.B., Kumi-Ansah,F., Odoom,T., Boakye,O.D., Amoako,E., Abass,A.-K., Quashie,P., Amenga-Etego,L.N., Akoriyea,S.K., Awandare,G.A., Bediako,Y. |
| EPI_ISL_885139 | PathWest Laboratory Medicine WA | PathWest Laboratory Medicine WA Microbial Surveillance Unit | PathWest Laboratory Medicine WA Microbial Surveillance Unit |
| EPI_ISL_885143 | Central public health laboratory | Molecular Diagnostics Department, Central public health laboratory | Dler,H., Dlshad,H., Furat,S., Sharmeen,F.-A., Dalia,F., Mohsen,A.,Hemdad,A., Fahmi,A., Hemn,M. and Idrees,H. |
| EPI_ISL_886249, EPI_ISL_886371, EPI_ISL_886433 | Labcorp | Genomics and Discovery, Respiratory Viruses Branch, Division of Viral Diseases, Centers for Disease Control and Prevention | Peter W. Cook,Dhwani Batra,Ben L. Rambo-Martin,Summer Galloway,Brian Krueger,Minoo Agarwal,Eyad Almasri,Debbie Boles,Ayla Burns,Nuthawin Charoensri,Oren Cohen,Susan Countryman,Mary Ann Cristobal,Bobbi Croy,Suzanne Dale,Hrushikesh Deshmukh,Amanda Douglas,Vincent Drouillon,Marcia Eisenberg,Howard Engler,Rama Ghatti,Prashant Gupta,Susan Hicks,Jake Humphrey,Lax Iyer,Manoj Jain,Mohan Kolli,Tim Kuphal,Stanley Letovsky,Michael Levandoski,Craig Lukasik,Jonathan Meltzer,Brian Norvell,Mindy Nye,Scott Parker,Christos Petropoulos,John Pruitt,Steven Ragan,Scott Ryan,Mike Sapeta,Jana Schroth,Suresh Babu Selvaraju,Goran Stevovic,Amanda Suchanek,Andrea Throop,Lyndon Tilson,Thomas Urban,Joe Voshell,Kimberly Wagner,Jonathan Williams,Mary Williamson,Qian Zeng,Tricia Zwiefelhofer,Clinton R. Paden,Suxiang Tong,Duncan MacCannell, |
| EPI_ISL_887419, EPI_ISL_887428, EPI_ISL_887437, EPI_ISL_887439, EPI_ISL_887440, EPI_ISL_887443, EPI_ISL_887450, EPI_ISL_887476, EPI_ISL_887484, EPI_ISL_887488, EPI_ISL_887489, EPI_ISL_887490, EPI_ISL_887491, EPI_ISL_887492, EPI_ISL_887495, EPI_ISL_887503 | | | |
| see above | Instituto Nacional de Saude (INS), Mozambique | KRISP, KZN Research Innovation and Sequencing Platform | Nalia Ismael, Nadia Sitoe, Paulo Arnaldo, Nedio Mabunda, Giandhari J, Pillay S, Tegally H, Wilkinson E, de Oliveira T |
| EPI_ISL_888265, EPI_ISL_888378, EPI_ISL_888428, EPI_ISL_888433, EPI_ISL_888575 | Labcorp | Genomics and Discovery, Respiratory Viruses Branch, Division of Viral Diseases, Centers for Disease Control and Prevention | Peter W. Cook,Dhwani Batra,Ben L. Rambo-Martin,Summer Galloway,Brian Krueger,Minoo Agarwal,Eyad Almasri,Debbie Boles,Ayla Burns,Nuthawin Charoensri,Oren Cohen,Susan Countryman,Mary Ann Cristobal,Bobbi Croy,Suzanne Dale,Hrushikesh Deshmukh,Amanda Douglas,Vincent Drouillon,Marcia Eisenberg,Howard Engler,Rama Ghatti,Prashant Gupta,Susan Hicks,Jake Humphrey,Lax Iyer,Manoj Jain,Mohan Kolli,Tim Kuphal,Stanley Letovsky,Michael Levandoski,Craig Lukasik,Jonathan Meltzer,Brian Norvell,Mindy Nye,Scott Parker,Christos Petropoulos,John Pruitt,Steven Ragan,Scott Ryan,Mike Sapeta,Jana Schroth,Suresh Babu Selvaraju,Goran Stevovic,Amanda Suchanek,Andrea Throop,Lyndon Tilson,Thomas Urban,Joe Voshell,Kimberly Wagner,Jonathan Williams,Mary Williamson,Qian Zeng,Tricia Zwiefelhofer,Clinton R. Paden,Suxiang Tong,Duncan MacCannell, |
| EPI_ISL_889051, EPI_ISL_889141, EPI_ISL_889144, EPI_ISL_889234, EPI_ISL_889283 | Israel Central Virology laboratory | Israel National Consortium for SARS-CoV-2 sequencing | Neta Zuckerman, Efrat Dahan Bucris, Michal Mandelboim, Dana Bar-Ilan, Oran Erster, Tzvia Mann, Omer Murik, David A. Zeevi, Assaf Rokney, Joseph Jaffe, Eva Nachum, Maya Davidovich Cohen, Ephraim Fass, Gal Zizelski Valenci, Mor Rubinstein, Efrat Rorman, Israel Nissan, Efrat Glick-Saar, Omri Nayshool, Gideon Rechavi, Ella Mendelson, Orna Mor |
| EPI_ISL_889390 | Olomouc University Hospital | Institute of Applied Biotechnologies a.s. | Petr Klempt, Ondej Brzo, Martin Kašný, Kateina Kvapilová, Marián Hajdúch, Petr Kvapil |
| EPI_ISL_889616, EPI_ISL_889672 | LSUHS Emerging Viral Threat Laboratory | Microbial Genome Sequencing Center | Jeremy P. Kamil, Jennifer L. Carroll, Camille F. Abshire, Maarten Van Diest, Mohammed N.A. Siddiquey, Andrew D. Yurochko, Martin J. Sapp, Rona S. Scott, Christopher G. Kevil, Daniel J. Snyder, Vaughn S. Cooper, John A. Vanchiere |
| EPI_ISL_890185 | Academic Hospital of Gadjah Mada University (RSA UGM) | Genetics Working Group (Pokja Genetik) Faculty of Medicine, Public Health and Nursing Universitas Gadjah Mada (FK-KMK UGM); Disease Investigation Center Wates Ministry of Agriculture Indonesia; Department of Microbiology FK-KMK UGM; Laboratorium Diagnostik Yayasan Tahija World Mosquito Program (WMP) Yogyakarta Center for Tropical Medicine FK-KMK UGM; Integrated Research center FK-KMK UGM; Department of Computer Science and Electronics FMIPA UGM | Gunadi, Hendra Wibawa, Marcellus, Mohamad S. Hakim, Edwin W. Daniwijaya, Ludhang P. Rizki, Endah Supriyati, Eggi Arguni, Titik Nuryastuti, Tri Wibawa, Dwi AA Nugrahaningsih, Afiahayati, Siswanto, Kristy Iskandar, Nungki Anggorowati, Aditya Rifqi Fauzi, Fadil Fahri |
| EPI_ISL_890208 | Cancer Biology Department, National Cancer Institute | Cancer Biology Department, National Cancer Institute | Zekri,A.N., Sedawy,M.G., Ahmed,O.S., Hafez,M.M., Soliman,H.K., Bahnassy,A.A., Elhosieny,F.W., Gad,A.E., Hamdy,M.S., Soliman,M.S., Soliman,L., Abouelhoda,M. |
| EPI_ISL_891031, EPI_ISL_891041 | Washington State Department of Health | Seattle Flu Study | Deborah A. Nickerson, Chris D. Frazar, Jover Lee, Benjamin Pelle, Erica Ryke, Matthew Richardson, Amanda Adler, Elisabeth Brandstetter, Peter D. Han, Kairsten Fay, Misja Ilcisin, Kirsten Lacombe, Thomas R. Sibley, Melissa Truong, Caitlin R. Wolf, Romesh Gautom, Geoff Melly, Brian Hiatt, Philip Dykema, |

| | | | |
|---|---|---|---|
| | | | Scott Lindquist, Michael Boeckh, Janet A. Englund, Michael Famulare, Barry R. Lutz, Mark J. Rieder, Lea M. Starita, Matthew Thompson, Helen Y. Chu, Jay Shendure, Trevor Bedford |
| EPI_ISL_891136 | The Jackson Laboratory | The Jackson Laboratory | Lloyd M, Sanderson B, Srivastava A, Maurya R, Renzette N, Omerza G, Kelly K, Li L, Wei C L, Adams M |
| EPI_ISL_891231, EPI_ISL_891247 | National Public Health Laboratory, National Centre for Infectious Diseases | National Public Health Laboratory, National Centre for Infectious Diseases | Tze Minn Mak, Zhenyang Zhou, Lin Cui, Raymond Tzer Pin Lin |
| EPI_ISL_891384 | Lighthouse Lab in Alderley Park | Wellcome Sanger Institute for the COVID-19 Genomics UK (COG-UK) Consortium | Jacquelyn Wynn, Mairead Hyland, The Lighthouse Lab in Alderley Park and Alex Alderton, Roberto Amato, Sonia Goncalves, Ewan Harrison, David K. Jackson, Ian Johnston, Dominic Kwiatkowski, Cordelia Langford, John Sillitoe on behalf of the Wellcome Sanger Institute COVID-19 Surveillance Team |
| EPI_ISL_892244 | National Laboratory for Health, Environment and Food | National Laboratory for Health, Environment and Food | Aleksander Mahnic, Sandra Janezic, Maja Rupnik |
| EPI_ISL_893362, EPI_ISL_894295 | Pathogen Genomics Center, National Institute of Infectious Diseases | Pathogen Genomics Center, National Institute of Infectious Diseases | Tsuyoshi Sekizuka, Kentaro Itokawa, Rina Tanaka, Masanori Hashino, Makoto Kuroda |
| EPI_ISL_896374 | University of Medicine and Pharmacy of Craiova | "Stefan cel Mare" University Metagenomics Lab | Lobiuc Andrei, Gheorghita Roxana |
| EPI_ISL_896380 | Suceava County Emergency Hospital | "Stefan cel Mare" University Metagenomics Lab | Lobiuc Andrei, Gheorghita Roxana |
| EPI_ISL_898026 | KU Leuven, Rega Institute, Clinical and Epidemiological Virology | KU Leuven, Rega Institute, Clinical and Epidemiological Virology | Tony Wawina-Bokalanga, Bert Vanmechelen, Joan Marti-Carerras, Piet Maes |
| EPI_ISL_899154, EPI_ISL_899300, EPI_ISL_899580 | Viollier AG | Department of Biosystems Science and Engineering, ETH Zürich | Christian Beisel, Sarah Nadeau, Chaoran Chen, Ivan Topolsky, Philipp Jablonski, Lara Fuhrmann, David Dreifuss, Katharina Jahn, Tobias Schär, Ina Nissen, Natascha Santacroce, Elodie Burcklen, Christiane Beckmann, Maurice Redondo, Olivier Kobel, Christoph Noppen, Sophie Seidel, Noemie Santamaria de Souza, Niko Beerenwinkel, Tanja Stadler |
| EPI_ISL_900578 | IZSM | TIGEM | Antonio Grimaldi, Patrizia Annunziata, Francesco Panariello, Biancamaria Pierri, Valentina Bouche, Chiara Colantuono, Maria Concetta Cuomo, Denise Di Concilio, Lucio Di Filippo, Anna Manfredi, Marcello Salvi, Antonio Limone, Pellegrino Cerino, Andrea Ballabio, Davide Cacchiarelli. |
| EPI_ISL_902820, EPI_ISL_902859, EPI_ISL_902861, EPI_ISL_902869 | Department of Virology and Immunology, University of Helsinki and Helsinki University Hospital, Huslab Finland | Department of Virology, Faculty of Medicine, University of Helsinki, Helsinki, Finland | Teemu Smura, Ravi Kant, Phuoc Truong, Hussein Alburkat, Hannimari Kallio-Kokko, Jenni Virtanen, Maija Suvanto, Essi Korhonen, Sari Hannula, Harri Kangas, Hanna Liimatainen, Satu Kurkela, Hanna Jarva, Maija Lappalainen, Pekka Ellonen, Olli Vapalahti |
| EPI_ISL_903083 | Seattle Flu Study | Seattle Flu Study | Deborah A. Nickerson, Chris D. Frazar, Jover Lee, Benjamin Pelle, Erica Ryke, Matthew Richardson, Amanda Adler, Elisabeth Brandstetter, Peter D. Han, Kairsten Fay, Misja Ilcisin, Kirsten Lacombe, Thomas R. Sibley, Melissa Truong, Caitlin R. Wolf, Karen Cowgill, Stephanie Schrag, Jeff Duchin, Michael Boeckh, Janet A. Englund, Michael Famulare, Barry R. Lutz, Mark J. Rieder, Lea M. Starita, Matthew Thompson, Helen Y. Chu, Trevor Bedford, Jay Shendure |
| EPI_ISL_903313 | University of Iowa Hospitals & Clinics, Microbiology Laboratory | University or Iowa, Lung Biology and Cystic Fibrosis Research Center, Pezzulo Lab | Miguel E Ortiz, Alejandro A Pezzulo |
| EPI_ISL_903379, EPI_ISL_903382, EPI_ISL_903383 | MOH - Jaber Al-Ahmad Hospital (Innovation Research Laboratory) | MOH - Jaber Al-Ahmad Hospital (Innovation Research Laboratory) | Salman Al-Sabah , Mohammad Alghounaim |
| EPI_ISL_903657, EPI_ISL_903696, EPI_ISL_903730, EPI_ISL_903816 | VT Dept. of Health Laboratory | Genomics and Discovery, Respiratory Viruses Branch, Division of Viral Diseases, Centers for Disease Control and Prevention | Krista Queen, Yan Li, Ying Tao, Jing Zhang, Anna Uehara, Anna Montmayeur, Clinton R. Paden, Peter W. Cook, Rachel Marine, Mili Sheth, Jasmine Padilla, Sarah Nobles, Mark Burroughs, Lori Rowe, Haibin Wang, Ben L. Rambo-Martin, Dhwani Batra, Justin Lee, Suxiang Tong |
| EPI_ISL_903860 | NH Dept. of Health and Human Services Public Health Labs | Genomics and Discovery, Respiratory Viruses Branch, Division of Viral Diseases, Centers for Disease Control and Prevention | Krista Queen, Yan Li, Ying Tao, Jing Zhang, Anna Uehara, Anna Montmayeur, Clinton R. Paden, Peter W. Cook, Rachel Marine, Mili Sheth, Jasmine Padilla, Sarah Nobles, Mark Burroughs, Lori Rowe, Haibin Wang, Ben L. Rambo-Martin, Dhwani Batra, Justin Lee, Suxiang Tong |
| EPI_ISL_903900, EPI_ISL_903920 | IA State Hygienic Laboratory | Genomics and Discovery, Respiratory Viruses Branch, Division of Viral Diseases, Centers for Disease Control and Prevention | Krista Queen, Yan Li, Ying Tao, Jing Zhang, Anna Uehara, Anna Montmayeur, Clinton R. Paden, Peter W. Cook, Rachel Marine, Mili Sheth, Jasmine Padilla, Sarah Nobles, Mark Burroughs, Lori Rowe, Haibin Wang, Ben L. Rambo-Martin, Dhwani Batra, Justin Lee, Suxiang Tong |
| EPI_ISL_903929 | VT Dept. of Health Laboratory | Genomics and Discovery, Respiratory Viruses Branch, Division of Viral Diseases, Centers for Disease Control and Prevention | Krista Queen, Yan Li, Ying Tao, Jing Zhang, Anna Uehara, Anna Montmayeur, Clinton R. Paden, Peter W. Cook, Rachel Marine, Mili Sheth, Jasmine Padilla, Sarah Nobles, Mark Burroughs, Lori Rowe, Haibin Wang, Ben L. Rambo-Martin, Dhwani Batra, Justin Lee, Suxiang Tong |
| EPI_ISL_904004, EPI_ISL_904006 | Scientific Veterinary Institute Novi Sad | Veterinary Specialized Institute "Kraljevo", Serbia | Vidanovic,D., Tesovic,B., Knezevic,A., Jovanovic,T., Jankovic,M., Sekler,M., Banovic Djeri,B., Petrovic,T., Volkening,J., Afonso,C. |
| EPI_ISL_904010 | Veterinary Specialized Institute Kraljevo | Veterinary Specialized Institute "Kraljevo", Serbia | Vidanovic,D., Tesovic,B., Knezevic,A., Jovanovic,T., Jankovic,M., Sekler,M., Banovic Djeri,B., Petrovic,T., Volkening,J., Afonso,C. |
| EPI_ISL_904016 | Molecular Biology and Virology lab, Faculty of Veterinary Medicine, Jordan University of Science and Technology | Molecular Biology and Virology lab, Faculty of Veterinary Medicine, Jordan University of Science and Technology | Mohammad Hussien Alboom, Dr.Mahmoud Hamad Gazo, Suhaila Ibrahim Khalil, Dr.Saied Jaradat, Hazaem Haddad, Dr. Moh"D Borhan Al-Zghoul , Dr.Mustafa Ababneh |
| EPI_ISL_904370 | Dutch COVID-19 response team | Erasmus Medical Center | Bas Oude Munnink, Reina Sikkema, David Nieuwenhuijse, Irina Chestakova, Anne van der Linden, Marjan Boter, Emmanuelle Munger, Corine GeurtsvanKessel, Annemiek van der Eijk, Richard Molenkamp, Marion Koopmans, on behalf of the Dutch national COVID-19 response team. |
| EPI_ISL_904761, EPI_ISL_905562 | Dutch COVID-19 response team | National Institute for Public Health and the Environment (RIVM) | Adam Meijer, Harry Vennema, Dirk Eggink, Jeroen Cremer, Sharon van den Brink, Bas van der Veer, AnneMarie van den Brandt, Florian Zwagemaker, Dennis Schmitz, Chantal Reusken, on behalf of the national COVID-19 response team |
| EPI_ISL_905735 | National Institute of Public Health - National Institute of Hygiene | National Institute of Public Health - National Institute of Hygiene | Wokowicz Tomasz, Zacharczuk Katarzyna |
| EPI_ISL_905820 | OHSU Lab Services Molecular Microbiology Lab | Oregon SARS-CoV-2 Genome Sequencing Center | Brendan L. O'Connell, Sally Grindstaff, Kayla Carter, Ruth V. Nichols, Alec J. Hirsch, Donna Hansel, Guang Fan, Xuan, Qin, Daniel N. Streblow, William B. Messer, Andrew C. Adey, Benjamin N. Bimber, Brian J. O'Roak |
| EPI_ISL_906058 | Tilia Laboratories s.r.o. | Tilia Laboratories s.r.o. | Sona Pekova, MD, PhD. |
| EPI_ISL_906284 | Nigeria Centre for Disease Control (NCDC) | African Centre of Excellence for Genomics of Infectious Diseases (ACEGID), Redeemer's University | Oluniyi P.E. et al |
| EPI_ISL_907075 | Department of Biology, University of Basrah | Department of Biology, University of Basrah | Abu-Ali,H.M. and Al-Badran,I.F. |
| EPI_ISL_907079 | Biology, MCL | Biology, MCL | Seadawy,M.G., Shamel,M.D., Gad,A.F., Elhoseiny,M.F., Zekri,A.N. |
| EPI_ISL_907095, EPI_ISL_907117 | Cancer Biology Department, National Cancer Institute | Cancer Biology Department, National Cancer Institute | Zekri,A.N., Sedawy,M.G., Ahmed,O.S., Hafez,M.M., Soliman,H.K., Bahnassy,A.A., Elhosieny,F.W., Gad,A.E., Hamdy,M.S., Soliman,M.S., Soliman,L., Abouelhoda,M. |
| EPI_ISL_910552, EPI_ISL_910565, EPI_ISL_911130 | Laboratoire national de sante, Microbiology, Virology | Laboratoire national de sante, Microbiology, Microbial Genomics Platform | Anke Wienecke-Baldacchino, Catherine Ragimbeau,Jessica Tapp, Fatu Djabi, Lise Pignon, Raoul Salmon, Tamir Abdelrahman |
| EPI_ISL_911439 | AZDelta | AZDelta | Geert Martens; Dieter De Smet |
| EPI_ISL_911708 | Tanjungpura University Hospital | Tanjungpura University Hospital | Puji Astuti; Andriani; Mahyarudin; Delima Fajar Liana; Virhan Novianry ; Sofi Siti Shofiyah |
| EPI_ISL_912354, EPI_ISL_912372, EPI_ISL_912373, EPI_ISL_912376, EPI_ISL_912377, EPI_ISL_912378, EPI_ISL_912380 | Fondation Congolaise pour la recherche medicale (FCRM), Francine Ntoumi | NGS Competence Center Tuebingen, Institut für Medizinische Mikrobiologie und Hygiene, Universitaetsklinikum Tübingen | Angel Angelov |
| EPI_ISL_912411 | KU Leuven, Rega Institute, Clinical and Epidemiological Virology | KU Leuven, Rega Institute, Clinical and Epidemiological Virology | Tony Wawina-Bokalanga, Bert Vanmechelen, Joan Marti-Carerras, Piet Maes |

| | | | |
|---|---|---|---|
| EPI_ISL_912481 | NHLS Universitas Academic | UFS Virology | PA Bester, MM Nyaga, P Nthiga, MT Mogotsi, D Goedhals, T de Oliveira |
| EPI_ISL_912750 | Hôpital Henri Mondor | Department of Virology, Henri Mondor University Hospital, Assistance Publique Hôpitaux de Paris, Université Paris-Est Créteil, INSERM U955 | Christophe Rodriguez, Slim Fourati, Vanessa Demontant, Guillaume Gricourt, Melissa N'Debi, Alexandre Soulier, Elisabeth Trawinski, Jean-Michel Pawlotsky |
| EPI_ISL_913022 | Hospital San Pedro de Alcántara | Instituto de Salud Carlos III | Iglesias-Caballero, M. Camarero, S. Sandonís,V. Vázquez, S. Pozo, F. Casas, I. Jiménez, P. Zaballos, A. Monzón, S. Varona, S. Cuesta, I. Rodríguez, G. |
| EPI_ISL_913040 | Hospital Universitario de Guadalajara | Instituto de Salud Carlos III | Iglesias-Caballero, M. Camarero, S. Sandonís,V. Vázquez, S. Pozo, F. Casas, I. Jiménez, P. Zaballos, A. Monzón, S. Varona, S. Cuesta, I. González, A. |
| EPI_ISL_913084 | Center for Virology | Center for Virology | Jeremy V. Camp, Irene Goerzer, Monika Redlberger-Fritz, Stephan W. Aberle |
| EPI_ISL_913462 | Klinisk mikrobiologi | The Public Health Agency of Sweden | Anna-Malin Linde, Maria Lind Karlberg, Carlo Berg, Oskar Karlsson Lindsjo, Sofia Stamouli, Reza Advani, Mattias Haukland, Petra Holmstrom, Noura Walai, Petra Edquist, Mia Brytting, Anna Risberg, Karin Tegmark-Wisell |
| EPI_ISL_913846 | TGen North | TGen North | "Jolene Bowers, Megan Folkerts, Chris French, Hayley Yaglom, Ashlyn Pfeiffer, Darrin Lemmer, Dave Engelthaler, The Arizona COVID Genomics Union (ACGU)" |
| EPI_ISL_913939, EPI_ISL_913957, EPI_ISL_913977 | Instituto de Diagnostico y Referencia Epidemiologicos INDRE_RNLSP | Instituto de Diagnostico y Referencia Epidemiologicos (INDRE) | Claudia Wong-Arambula, Abril Rodriguez-Maldonado, Fabiola Garces-Ayala, Adnan Araiza-Rodriguez, David Fragoso-Fonseca, Sergio Rangel-Guerrero, Mayra Jimenez-Morales, Nancy Munoz-Hernandez, Natividad Cruz-Ortiz, Tatiana Nunez-Garcia, Gisela Barrera-Badillo, Lucia Hernandez-Rivas, Irma Lopez-Martinez, Ernesto Ramirez-Gonzalez. |
| EPI_ISL_914802 | AREA DE SALUD CATEDRAL NORESTE | Inciensa, Instituto Costarricense de Investigación y Enseñanza en Nutrición y Salud | Francisco Duarte, Hebleen Porras, Claudio Soto-Garita, Estela Cordero, Adriana Godínez, Melany Calderón & Mariel López |
| EPI_ISL_914812 | AREA DE SALUD GOICOECHEA 2 - CLINICA DR. JIMENEZ NUÑEZ | Inciensa, Instituto Costarricense de Investigación y Enseñanza en Nutrición y Salud | Francisco Duarte, Hebleen Porras, Claudio Soto-Garita, Estela Cordero, Adriana Godínez, Melany Calderón & Mariel López |
| EPI_ISL_914813 | TAMIZAJE COMUNITARIO - PASO CANOAS | Inciensa, Instituto Costarricense de Investigación y Enseñanza en Nutrición y Salud | Francisco Duarte, Hebleen Porras, Claudio Soto-Garita, Estela Cordero, Adriana Godínez, Melany Calderón & Mariel López |
| EPI_ISL_914814 | HOSPITAL METROPOLITANO | Inciensa, Instituto Costarricense de Investigación y Enseñanza en Nutrición y Salud | Francisco Duarte, Hebleen Porras, Claudio Soto-Garita, Estela Cordero, Adriana Godínez, Melany Calderón & Margarita Lee-Lui |
| EPI_ISL_914815 | HOSPITAL DE NIÑOS DR. CARLOS SAENZ HERRERA | Inciensa, Instituto Costarricense de Investigación y Enseñanza en Nutrición y Salud | Francisco Duarte, Hebleen Porras, Claudio Soto-Garita, Estela Cordero, Adriana Godínez, Melany Calderón & Cristian Pérez-Corrales |
| EPI_ISL_914820 | AREA DE SALUD SAN JUAN-SAN DIEGO-CONCEPCION 2 | Inciensa, Instituto Costarricense de Investigación y Enseñanza en Nutrición y Salud | Francisco Duarte, Hebleen Porras, Claudio Soto-Garita, Estela Cordero, Adriana Godínez, Melany Calderón & Mariel López |
| EPI_ISL_914822 | HOSPITAL DR. ENRIQUE BALTODANO BRICEÑO | Inciensa, Instituto Costarricense de Investigación y Enseñanza en Nutrición y Salud | Francisco Duarte, Hebleen Porras, Claudio Soto-Garita, Estela Cordero, Adriana Godínez, Melany Calderón & Mariel López |
| EPI_ISL_914877 | Sydney South West Pathology Service (SSWPS) - Royal Prince Alfred Hospital - NSW Health Pathology | NSW Health Pathology - Institute of Clinical Pathology and Medical Research; Westmead Hospital; University of Sydney | CIDM-PH et al. |
| EPI_ISL_915421 | MRCG at LSHTM Genomics lab | MRCG at LSHTM Genomics lab | Abdul Karim sesay, Abdoulie Kanteh, Jarra Manneh, Mariama Kujabi, Bakary Sanyang |
| EPI_ISL_918367, EPI_ISL_918372 | Virology Unit, Institut Pasteur du Cambodge | Virology Unit, Institut Pasteur du Cambodge | Sokhoun Yann, Ly Sovann, Kraing Sidonn, Yi Sengdoeurn, Chin Savuth, Chau Darapheak, Etienne Simon-Loriere, Veasna Duong, Erik A Karlsson |
| EPI_ISL_918560 | LACEN - Laboratório Central de Saúde Pública do Amapa | Evandro Chagas Institute | Santos, M.C.; Silva, A.M.; Junior, W.D.C.; Barbagelata, L.S.; Ferreira, J.A.; Sousa, E.M.A.; da Silva, P.S.; Pinheiro, K.C.; L.C.; Sousa Junior, E.C. |
| EPI_ISL_922614 | Wales Specialist Virology Centre Sequencing lab: Pathogen Genomics Unit | Public Health Wales Microbiology Cardiff Wales Specialist Virology Centre | Catherine Moore, Johnathan Evans, Laura Gifford, Malorie Perry, Simon Cottrell, Angela Marchbank, Alec Birchley, Alexander Adams, Amy Gaskin, Bree Gatica-Wilcox, Jason Coombes, Joel Southgate, Lauren Gilbert, Lee Graham, Nicole Pacchiarini, Sara Kumziene-Summerhayes, Sarah Taylor, Sophie Jones, Sara Rey, Matthew Bull, Joanne Watkins, Sally Corden, Tom Connor |
| EPI_ISL_925851, EPI_ISL_925870, EPI_ISL_925903, EPI_ISL_925908, EPI_ISL_925913, EPI_ISL_925915 | Nucleic Acid Testing, National Reference Laboratory | GIGA Medical Genomics | Yvan Butera, Keith Durkin, Maria Artesi, Bouchra Boujemla, Robert Rutayisire, Patrick Tuyisenge, Esperence Umumararungu, Sébastien Bontems, Marie-Pierre Hayette, Nathalie Renotte, Swaibu Gatare, Jacob Souopgui, Sabin Nsanzimana, Vincent Bours, Léon Mutesa |
| EPI_ISL_929262 | Department of Virus and Microbiological Special Diagnostics, Statens Serum Institut, Copenhagen, Denmark | Aalborg University | Danish Covid-19 Genome Consortium |
| EPI_ISL_933648 | Toronto Invasive Bacterial Diseases Network | McMaster University | Allison McGeer, Patryk Aftanas, Hooman Derakhshani, Angel Li, Kuganya Nirmalarajah, Emily Panousis, Ahmed Draia, Jalees Nasir, Michael Surette, Samira Mubareka, Andrew G. McArthur |
| EPI_ISL_933780, EPI_ISL_933781, EPI_ISL_933782, EPI_ISL_933783, EPI_ISL_933784 | Virology Unit, Institut Pasteur du Cambodge | Virology Unit, Institut Pasteur du Cambodge | Sokhoun Yann, Ly Sovann, Kraing Sidonn, Yi Sengdoeurn, Chin Savuth, Chau Darapheak, Etienne Simon-Loriere, Veasna Duong, Erik A Karlsson |
| EPI_ISL_933790, EPI_ISL_933798 | PathWest Laboratory Medicine WA | PathWest Laboratory Medicine WA Microbial Surveillance Unit | PathWest Laboratory Medicine WA Microbial Surveillance Unit |
| EPI_ISL_934102, EPI_ISL_934180 | Vilnius university hospital Santaros Klinikos, Center of Laboratory Medicine | Vilnius university hospital Santaros Klinikos, Center of Laboratory Medicine | Ingrida Olendraite, Daniel Naumovas, Rimvydas Norvilas, Dovile Ezerskyte, Justinas Slikas, Gytis Dudas |
| EPI_ISL_934601 | Austrian Agency for Health and Food Safety (AGES) | Bergthaler laboratory, CeMM Research Center for Molecular Medicine of the Austrian Academy of Sciences | Lukas Endler, Anna Schedl, Thomas Penz, Benedikt Agerer, Maelle Le Moing, Michael Schuster, Bekir Erguner, Jan Laine, Martin Senekowitsch, Christoph Bock, Andreas Bergthaler |
| EPI_ISL_935087 | Ministry of Health Turkey | Ministry of Health Turkey | Fatma Bayrakdar, Yasemin Cogun, Süleyman Yalcin, Aye Baak Alta, Gülay Korukluolu |
| EPI_ISL_936607 | Northwestern Memorial Hospital | Ozer Lab | Ramon Lorenzo-Redondo, Lacy M. Simons, Chad J. Achenbach, Lawrence J. Jennings, Michael G. Ison, Judd F. Hultquist, Egon A. Ozer |
| EPI_ISL_940157 | Laboratory of Virology and Molecular Diagnostics | Institute of Public Health of Republic of North Macedonia Laboratory of Virology and Molecular Diagnostics | Maja Kuzmanovska, Golubinka Bosehvska, Elizabeta Janchevska |
| EPI_ISL_940279, EPI_ISL_940347 | Hôpital Bichat Claude Bernard, Laboratoire de Virologie | IAME UMR1137 Inserm, Université de Paris, Hôpital Bichat | Antoine Bridier-Nahmias, Amélie Recoing, Quentin Le Hingrat, Lena Daniel, Siham Hamri, Gilles Collin, Alexandre Storto, Mélanie Bertine, Charlotte Charpentier, Nadhira Houhou-Fidouh, Diane Descamps, Benoit Visseaux |
| EPI_ISL_940769, EPI_ISL_940773, EPI_ISL_940775 | INSPI-CRN de Influenza y otros virus respiratorios | INSPI-Centro de Investigación Multidisciplinaria de la DTIDI | Leandro Patiño, Doménica de Mora, Maritza Olmedo, Andrés Carrazco-Montalvo, Orson Mestanza, Mary Regato-Arrata, Melissa Zambrano, Manuel González, Alfredo Bruno, Alberto Orlando. |
| EPI_ISL_940889 | Vaccines and Infectious Diseases Analytics Research Unit (VIDA) | KRISP, KZN Research Innovation and Sequencing Platform | Baillie Vicky, du Plessis Jeanine, Giandhari Jennifer, Pillay Sureshnee, Naidoo Yeshnee, Tegally Houriiyah, de Oliveira Tulio, Madhi Shabir |
| EPI_ISL_940912, EPI_ISL_940986 | Centers for Disease Control and Prevention, Dengue Branch | Centers for Disease Control and Prevention, Dengue Branch | Gilberto A. Santiago, Glenda Gonzalez, Betzabel Flores, Keyla Charriez, Gabriela Paz-Bailey, Jorge L. Munoz-Jordan |
| EPI_ISL_941276 | Nigeria Centre for Disease Control (NCDC) | African Centre of Excellence for Genomics of Infectious Diseases (ACEGID), Redeemer's University | Oluniyi P.E. et al |
| EPI_ISL_941442, EPI_ISL_941638 | Instituto Nacional de Saude (INSA) | Instituto Nacional de Saude (INSA) | Borges et al |
| EPI_ISL_941761, EPI_ISL_941860 | Instituto Nacional de Saude (INSA) and Instituto Gulbenkian de Ciencia (IGC) | Instituto Nacional de Saude (INSA) and Instituto Gulbenkian de Ciencia (IGC) | Borges et al |
| EPI_ISL_941943, EPI_ISL_941994 | Instituto Nacional de Salud, Bogotá, Colombia | Centro de Investigaciones en Microbiología y Biotecnología-UR (CIMBIUR), Facultad de Ciencias | Luz Helena Patiño, Marina Muñoz, Nathalia Ballesteros, Carolina Hernández, Carolina Flórez, Sergio Gomez, Adriana van de Guchte, Zenab Khan, Jayeeta Dutta, Hala Alejel Alshammary, Ana S. Gonzalez-Reiche, Matthew M. Hernandez, Emilia Mia Sordillo, Viviana Simon, Harm van Bakel, Alberto |

| | | | |
|---|---|---|---|
| | | Naturales, Universidad del Rosario, Bogotá, Colombia Instituto Nacional de Salud, Bogotá, Colombia Icahn School of Medicine at Mount Sinai, New York, USA | Paniz-Mondolfi, Juan David Ramírez |
| EPI_ISL_941996, EPI_ISL_942000, EPI_ISL_942002 | Centro de Investigaciones en Microbiología y Biotecnología-UR (CIMBIUR), Facultad de Ciencias Naturales, Universidad del Rosario, Bogotá, Colombia | Centro de Investigaciones en Microbiología y Biotecnología-UR (CIMBIUR), Facultad de Ciencias Naturales, Universidad del Rosario, Bogotá, Colombia Instituto Nacional de Salud, Bogotá, Colombia Icahn School of Medicine at Mount Sinai, New York, USA | Luz Helena Patiño, Marina Muñoz, Nathalia Ballesteros, Carolina Hernández, Carolina Flórez, Sergio Gomez, Adriana van de Guchte, Zenab Khan, Jayeeta Dutta, Hala Alejel Alshammary, Ana S. Gonzalez-Reiche, Matthew M. Hernandez, Emilia Mia Sordillo, Viviana Simon, Harm van Bakel, Alberto Paniz-Mondolfi, Juan David Ramírez |
| EPI_ISL_942749, EPI_ISL_942860 | Gundersen Molecular Diagnostics Laboratory | Kabara Cancer Research Institute | Craig S. Richmond, Paraic A. Kenny |
| EPI_ISL_943096 | Dutch COVID-19 response team | National Institute for Public Health and the Environment (RIVM) | Adam Meijer, Harry Vennema, Dirk Eggink, Jeroen Cremer, Sharon van den Brink, Bas van der Veer, AnneMarie van den Brandt, Florian Zwagemaker, Dennis Schmitz, Chantal Reusken, on behalf of the national COVID-19 response team |
| EPI_ISL_943559 | National Institute of Laboratory Medicine and Referral Center | Genomic Research Lab, BCSIR | Shahina Akter, Mohammad Samir Uzzaman, Eshrar Osman, Md. Ahashan Habib,Tanjina Akhtar Banu, Abu Sayeed Mohammad Mahmud, Md. Murshed Hasan Sarkar, Barna Goswami, Iffat Jahan, Md. Saddam Hossain, Tasnim Nafisa, Md. Maruf Ahmed Molla, Mahmuda Yeasmin, Asish Kumar Ghosh, Arifa Akram, A. K. M. Shamsuzzaman, Md. Salim Khan |
| EPI_ISL_943585 | Central Laboratory of Public Health of Rio Grande do Sul (Lacen-RS) | State Center for Health Surveillance of the Health Department of the State of Rio Grande do Sul (CEVS/SES-RS) | Aline Campos, Amanda da Silva, Anelise Schaurich, Claudia Dornelles, Cynthia Molina, Fernanda Godinho, Lara Crescente, Leticia Garay, Regina Barcellos, Richard Salvato, Tatiana Gregianini, Vagner Fonseca |
| EPI_ISL_944154 | National Health Laboratory Service, South Africa | KRISP, KZN Research Innovation and Sequencing Platform | Laguda-Akingba O, Giandhari J, Pillay S, Lessells R, Mdlalose K, York D, Khan S, Emmanuel SJ, Tegally H, Wilkinson E, de Oliveira T |
| EPI_ISL_944555 | Israel Central Virology laboratory | Israel National Consortium for SARS-CoV-2 sequencing | Neta Zuckerman, Efrat Dahan Bucris, Michal Mandelboim, Dana Bar-Ilan, Oran Erster, Tzvia Mann, Omer Murik, David A. Zeevi, Assaf Rokney, Joseph Jaffe, Eva Nachum, Maya Davidovich Cohen, Ephraim Fass, Gal Zizelski Valenci, Mor Rubinstein, Efrat Rorman, Israel Nissan, Efrat Glick-Saar, Omri Nayshool, Gideon Rechavi, Ella Mendelson, Orna Mor |
| EPI_ISL_944663, EPI_ISL_944667, EPI_ISL_944714 | Department of Biochemistry, Cell and Molecular Biology, West African Centre for Cell Biology of Infectious Pathogens (WACCBIP), University of Ghana | Department of Biochemistry, Cell and Molecular Biology, West African Centre for Cell Biology of Infectious Pathogens (WACCBIP), University of Ghana | Morang'a,C.M., Ngoi,J.M., Quansah,E.B., Saiid,S., Amuzu,D.S., Asante,I., Bonney,J.H., Bonney,E., Odoom,J.K., Ndam,N.T., Tei-Maya,F., Adusei-Poku,M., Ofori-Boadu,L., Ampofo,W.K., Amenga-Etego,L.N., Quashie,P., Bediako,Y., Awandare,G.A. |
| EPI_ISL_944736, EPI_ISL_944737, EPI_ISL_944744, EPI_ISL_944748 | Public Health Virology-Forensic and Scientific Services (PHV-FSS) | Public Health Virology-Forensic and Scientific Services (PHV-FSS) | Son Nguyen et al |
| EPI_ISL_954227, EPI_ISL_954243, EPI_ISL_954261, EPI_ISL_954264, EPI_ISL_954267, EPI_ISL_954270, EPI_ISL_954279, EPI_ISL_954287, EPI_ISL_954291 | MRC/UVRI & LSHTM Uganda Research Unit | Where sequence data have been generated and submitted to GISAID | Matthew Cotten, Dan Lule Bugembe, My V.T. Phan, Isaac Sseeewanyana, Patrick Semanda, Susan Nabadda, Pontiano Kaleebu |
| EPI_ISL_954745 | General Hospital - Ohrid | Research Center for Genetic Engineering and Biotechnology "Georgi D. Efremov" , Macedonian Academy of Sciences and Arts | Aleksandar J. Dimovski, Dijana Plasheska-Karanfilska, Predrag Noveski, Gjorgji Bozinovski, Milena Jakimovska |
| EPI_ISL_955148 | University of Sarajevo, Veterinary Faculty, Laboratory for Molecular Diagnostic and Research Laboratory | University of Sarajevo, Veterinary Faculty, Laboratory for Molecular Diagnostic and Research Laboratory | Goleti T., Goleti Š., Softi A., Ali-Šeho A., Jaži A., Šabi E., Terzi I., Nicevi M., Hodži A. |
| EPI_ISL_955161, EPI_ISL_955162 | Institute for Biocides and Medical Ecology | Institute of microbiology and Immunology, Faculty of Medicine, University of Belgrade | Knezevic,A., Jankovic,M., Vidanovic,D., Milicevic,O., Tesovic,B., Sekler,M., Jovanovic,T. |
| EPI_ISL_955184 | University of Sarajevo, Veterinary Faculty, Laboratory for Molecular Diagnostic and Research Laboratory | University of Sarajevo, Veterinary Faculty, Laboratory for Molecular Diagnostic and Research Laboratory | Goleti Š., Goleti T., Softi A., Ali-Šeho A., Nicevi M., Hodži A., Terzi I., Šabi E., Jaži A. |
| EPI_ISL_955272 | AR Dept. of Health-PHL, Molecular Diagnostics | Pathogen Discovery, Respiratory Viruses Branch, Division of Viral Diseases, Centers for Disease Control and Prevention | Ying Tao, Jing Zhang, Yan Li, Krista Queen, Anna Uehara, Peter Cook, Clinton R. Paden, Haibin Wang, Suxiang Tong |
| EPI_ISL_955325 | American Esoteric Laboratory | Pathogen Discovery, Respiratory Viruses Branch, Division of Viral Diseases, Centers for Disease Control and Prevention | Ying Tao, Jing Zhang, Yan Li, Krista Queen, Anna Uehara, Peter Cook, Clinton R. Paden, Haibin Wang, Suxiang Tong |
| EPI_ISL_956286 | Instituto Nacional de Salud- Dirección de Redes de Laboratorios de Salud Pública | Instituto Nacional de Salud- Dirección de Investigación en Salud Pública | Katherine Laiton-Donato, Diego A. Álvarez-Díaz, Carlos Franco-Muñoz, Mauricio Pacheco-Montealegre, Hector Alejandro Ruiz-Moreno, Maria T. Herrera-Sepúlveda, Diego Andrés Prada, Jhonnatan Reales-González, Sheryll Corchuelo, Julian Naizaque, Gerardo Santamaría, Magdalena Wiesner, Martha Lucia Ospina Martinez, Marcela Mercado-Reyes |
| EPI_ISL_956317, EPI_ISL_956325, EPI_ISL_956326, EPI_ISL_956330 | Laboratory Medicine | Department of Laboratory Medicine, Lin-Kou Chang Gung Memorial Hospital, Taoyuan, Taiwan | Kuo-Chien Tsao, Yu-Nong Gong, Shu-Li Yang, Yi-Chun Liu, Chung-Guei Huang, Mei-Jen Hsiao, Po-Wei Huang, Cheng-Ta Yang, Cheng-Hsun Chiu, Peng-Nien Huang, Kuo-Ming Lee, Guang-Wu Chen, Shin-Ru Shih |
| EPI_ISL_956367, EPI_ISL_956382 | General Hospital - Prilep | Research Center for Genetic Engineering and Biotechnology "Georgi D. Efremov" , Macedonian Academy of Sciences and Arts | Aleksandar J. Dimovski, Dijana Plasheska-Karanfilska, Predrag Noveski, Gjorgji Bozinovski, Milena Jakimovska |
| EPI_ISL_959277 | National Influenza Center, Virology Department | National Influenza Center | J Yavarian,K Sadeghi, NZ Shafiei Jandaghi, V Salimi, A Nejati, N Ghavvami,F Ajaminejad and T Mokhtari Azad |
| EPI_ISL_959280, EPI_ISL_959282 | National Influenza Center, Virology Department | National Influenza Center | J Yavarian, NZ Shafiei Jandaghi, V Salimi, A Nejati, K Sadeghi, F Ajaminejad,N Ghavvami and T Mokhtari Azad |
| EPI_ISL_959642 | Institute of Virology, Biomedical Research Center of the Slovak Academy of Sciences, Bratislava | Faculty of Natural Sciences, Comenius University, Bratislava | Kristína Boršová, Viktória abanová, Broa Brejová, Viktória Hodorová, Sabina Fumaová Havlíková, Juraj Kopáek, Martina Liková, ubomíra Lukáiková, Martina Neboháová, Monika Sláviková, Tomáš Vina, Boris Klempa, Jozef Nosek |
| EPI_ISL_959650 | General Hospital "Abdulah Nakas" Sarajevo | Alea Genetic Centre | Dino Pecar, Lana Salihefendic, Sead Jazic, Rijad Konjhodzic |
| EPI_ISL_960117 | Groote Schuur Hospital wc GSH | National Health Laboratory Service/UCT | Arash Iranzadeh, Deelan Doolabh, Lynn Tyers, Bruna Galvao, Innocent Mudau, Marvin Hsiao, Kruger Marais, Diana Hardie, Stephen Korsman, Carolyn Williamson |
| EPI_ISL_960228, EPI_ISL_960235, EPI_ISL_960300 | Nucleic Acid Testing, National Reference Laboratory | GIGA Medical Genomics | Yvan Butera, Keith Durkin, Maria Artesi, Bouchra Boujemla, Robert Rutayisire, Patrick Tuyisenge, Esperence Umumararungu, Sébastien Bontems, Marie-Pierre Hayette, Nathalie Renotte, Corinne Fasquelle, Swaibu Gatare, Jacob Souopgui, Sabin Nsanzimana, Vincent Bours, Léon Mutesa |
| EPI_ISL_960404, EPI_ISL_960418, EPI_ISL_960433 | The National Institute of Public Health | State Veterinary Institute Prague | Nagy,A;Vecerova,J;Cernikova,L;Stara,M;Jirincova,H;Trnka,D |
| EPI_ISL_960782 | Germano de sousa | Instituto Gulbenkian de Ciencia | João Costa, João Sobral, Maria Costa, Susana Ladeiro, Cathy Paulino, Ricardo Leite |
| EPI_ISL_960783 | Germano de sousa | Instituto Gulbenkian de Ciencia | Susana Ladeiro, Cathy Paulino, João Costa, João Sobral, Maria Costa, Ricardo Leite |
| EPI_ISL_960795 | Germano de sousa | Instituto Gulbenkian de Ciencia | João Costa, João Sobral, Maria Costa, Susana Ladeiro, Cathy Paulino, Ricardo Leite |
| EPI_ISL_961466 | Laboratorios Lister | Instituto de Diagnostico y Referencia Epidemiologicos (INDRE) | Claudia Wong-Arambula, Abril Rodriguez-Maldonado, Fabiola Garces-Ayala, Natividad Cruz-Ortiz, Tatiana Nunez-Garcia, Gisela Barrera-Badillo, Lucia Hernandez-Rivas, Irma Lopez-Martinez, Ernesto Ramirez-Gonzalez. |
| EPI_ISL_961467, EPI_ISL_961468, EPI_ISL_961469 | LESP Nuevo Leon/Grupo de Diagnostico ARIES | Instituto de Diagnostico y Referencia Epidemiologicos (INDRE) | Claudia Wong-Arambula, Abril Rodriguez-Maldonado, Fabiola Garces-Ayala, Natividad Cruz-Ortiz, Tatiana Nunez-Garcia, Gisela Barrera-Badillo, Lucia Hernandez-Rivas, Irma Lopez-Martinez, Ernesto Ramirez-Gonzalez. |
| EPI_ISL_961480 | Michigan Department of Health and Human Services, Bureau of Laboratories | Michigan Department of Health and Human Services, Bureau of Laboratories | Blankenship HM, Riner D, Soehnlen MK |

| | | | |
|---|---|---|---|
| EPI_ISL_961575 | Hôpital Georges L. Dumont | National Microbiology Laboratory (NML) | Anna Majer, Shari Tyson, Grace Seo, Philip Mabon, Elsie Grudeski, Rhiannon Huzarewich, Russell Mandes, Anneliese Landgraff, Jennifer Tanner, Natalie Knox, Morag Graham, Gary Van Domselaar, Richard Garceau, Guillaume Desnoyers, Nathalie Bastien, Yan Li, Timothy Booth, Darian Hole, Madison Chapel, Kirsten Biggar, CanCOGeN's metadata curation team, Public Health Agency of Canada CanCOGeN team |
| EPI_ISL_962205 | Institute for Medical Research, Infectious Disease Research Centre, National Institutes of Health, Ministry of Health Malaysia | Institute for Medical Research, Infectious Disease Research Centre, National Institutes of Health, Ministry of Health Malaysia | Suppiah J, Kamel K, Azizan MA, Thayan R |
| EPI_ISL_962536 | Infectious Disease Control and Prevention Institute | Infectious Disease Control and Prevention Institute | Xiaolin Jiang, Mingxiao Yao, Yuwei Zhang, Yujie He, Zengqiang Kou, Ti Liu, Yan Li, Jianxing Wang, Bo Pang, Julong Wu |
| EPI_ISL_962822 | Microbiological Diagnostic Unit - Public Health Laboratory (MDU-PHL) | MDU-PHL | Seemann T., Sait, M.L., Sherry, N.L. |
| EPI_ISL_962877, EPI_ISL_962878 | National Virology Reference Laboratory | National Public Health Laboratory, National Centre for Infectious Diseases | Tze Minn Mak, Zhenyang Zhou, Zaini Zainun, Taib Surita, Lin Cui, Raymond Tzer Pin Lin |
| EPI_ISL_962960 | Hospital Universitario de Gran Canaria Dr. Negrín | SeqCOVID-SPAIN consortium/IBV(CSIC) | M. Carmen Pérez González, Francisco J. Chamizo López, Ana Bordes Benítez and SeqCOVID-SPAIN consortium |
| EPI_ISL_964882 | General Hospital "Abdulah Nakas" Sarajevo | Alea Genetic Centre | Lana Salihefendic, Dino Pecar, Adis Kandic, Sead Jazic, Rijad Konjhodzic |
| EPI_ISL_964918, EPI_ISL_964924 | Instituto Nacional de Saude (INS), Mozambique | KRISP, KZN Research Innovation and Sequencing Platform | Nalia Ismael, Nadia Sitoe, Paulo Arnaldo, Nedio Mabunda, Giandhari J, Pillay S, Emmanuel S, Tegally H, Wilkinson E, de Oliveira T |
| EPI_ISL_965122 | Viral Respiratory Lab, National Institute for Biomedical Research (INRB) | Pathogen Sequencing Lab, National Institute for Biomedical Research (INRB) | Placide Mbala-Kingebeni, Edith Nkwembe, Eddy Kinganda-Lusamaki, Amuri Aziza, Francisca Muyembe Mawete, Emmanuel Lokilo Lofiko, Jean-Claude Makangara Cigolo, Catherine Pratt, Matthias Pauthner, Josh Quick, Allison Black, James Hadfield, Trevor Bedford, Ian Goodfellow, Andrew Rambaut, Nick Loman, Kristian Andersen, Michael Wiley, Steve Ahuka-Mundeke, Jean-Jacques Muyembe Tamfum |
| EPI_ISL_965854 | GA Department of Public Health | GA Department of Public Health | Stacy Reeves, Jonathan Edwards, Cynthia Dixey, Tonia Parrott |
| EPI_ISL_966832 | Maine HETL | Tewhey Lab, The Jackson Laboratory | Matluk,N., Dewey,H., Iosue,F., Barter,M., Lynch,R., Munger,H. and Tewhey,R. |
| EPI_ISL_966962, EPI_ISL_967019, EPI_ISL_967290 | Helix/Illumina | Respiratory Viruses Branch, Division of Viral Diseases, Centers for Disease Control and Prevention | Peter W. Cook,Dakota Howard,Dhwani Batra,Ben L. Rambo-Martin,Eileen de Feo,Jan Antico,Christine Tran,Matthew Tolentino,Shannon Wickline,Kim Gietzen,Brad Sickler,Jingtao Liu,Eric Allen,Phil Febbo,Summer Galloway,Nicole L. Washington,Simon White,Geraint Levan,Kelly Schiabor Barrett,Elizabeth Cirulli,Alexandre Bolze,Ary Ascencio,Charlotte Rivera-Garcia,Ryan Cho,Jason Nguyen,Sherry Wang,Jimmy Ramirez,Tyler Cassens,Efren Sandoval,Magnus Isaksson,William Lee,David Becker,Marc Laurent,James Lu,Clinton R. Paden,Suxiang Tong,Duncan MacCannell, |
| EPI_ISL_967723, EPI_ISL_967751 | State Laboratories Division, Hawaii State Department of Health | State Laboratories Division, Hawaii State Department of Health | Pamela O'Brien, Drew Kuwazaki, Ayana Garnet, Razvan Sultana, Edward Desmond |
| EPI_ISL_968081 | Public Health Virology-Forensic and Scientific Services (PHV-FSS) | Public Health Virology-Forensic and Scientific Services (PHV-FSS) | Alyssa T. Pyke et al. |
| EPI_ISL_968181 | Clinical Molecular Microbiology Laboratory, UNC Hospital | Dirk Dittmer | Justin T. Landis , Razia Moorad , Brent A. Eason, Melissa B. Miller, Linda Pluta, Dirk Dittmer, Angelica Juarez, Cecilia Thompson, Shawn Hawken, Cameroon Grant, Evelyn Hoffman, Patricio Cano, Jason Wong, Carolina Caro-Vegas, Ryan McNamara, Blossom Damania. |
| EPI_ISL_968213 | PHV-FSS | PHV-FSS | Son Nguyen et al. |
| EPI_ISL_969034 | Botswana Harvard HIV Reference Laboratory | Botswana Harvard HIV Reference Laboratory | Sikhulile Moyo, Dorcas Maruapula, Wonderful Choga, Botshelo Radibe, Boitumelo Zuze, David Lawrence, Roger Shapiro, Shahin Lockman, Mosepele Mosepele, Joseph Makhema, Simani Gaseitsiwe |
| EPI_ISL_971451 | Cell culture Unit at CV-MIT belonging to HIMMV | Functional Genomic Platform UATRS-biology, CNRST | Nadia Touil, Abderrazzak Rfak, Mouhssine Hemlali, Sanaa ALAOUI-Amine, Marouane MELLOUL, Mly Abdelaziz ELALAOUI, Hicham Elannaz, Amine Idriss Lahlou, Mostafa ELOUENNASS, Khalid ENNIBI, Elmostafa EL FAHIME |
| EPI_ISL_973147, EPI_ISL_973471 | Department of Virus and Microbiological Special Diagnostics, Statens Serum Institut, Copenhagen, Denmark | Aalborg University | Danish Covid-19 Genome Consortium |
| EPI_ISL_976824 | BCCDC Public Health Laboratory | BCCDC Public Health Laboratory | Prystajecky Natalie, Linda Hoang, Dan Fornika, John Tyson, Shannon Russell, Kim Macdonald, Kimia Kamelian, Ana Pacagnella, Corrinne Ng, Loretta Janz, Robert Azana Terry Snutch, Mel Krajden |
| EPI_ISL_977270, EPI_ISL_977278, EPI_ISL_977292, EPI_ISL_977334, EPI_ISL_977346, EPI_ISL_977347, EPI_ISL_977348, EPI_ISL_977349, EPI_ISL_977350, EPI_ISL_977351, EPI_ISL_977357, EPI_ISL_977362, EPI_ISL_977421, EPI_ISL_977463, EPI_ISL_977468 | | | |
| see above | University of Zambia, School of Veterinary Medicine | UNZAVET and PATH | Mulenga Mwenda-Chimfwembe, Ngonda Saasa, Daniel Bridges |
| EPI_ISL_977581 | Caribbean Public Health Agency | Carrington Lab, Department of PreClinical Sciences | Nikita S. D. Sahadeo, Arianne Brown-Jordan, Vernie Ramkissoon, Sarah Hill, Naresh Nandram, Avery Hinds, Dr. Sharon Belmar-George, Jerome Foster, Stanley Giddings, Karla Georges, Marsha Ivey, Rahul Naidu, Risha Singh, SueMin Nathaniel, Rajini Haraksingh, Jaya Jayaraman, Chinna Chinnadurai, Adesh Ramsubhag, Nuno Faria, Oliver Pybus, Christopher Oura, Gabriel Escobar, Christine V. F. Carrington |
| EPI_ISL_977587 | Department of Medical Microbiology, Hospital Pengajar Universiti Putra Malaysia | Malaysia Genome Institute | Mohd Noor Mat Isa, Syafinaz Amin-Nordin, Irni Suhayu Sapian, Hui-Yee Chee, Yusuf Muhammad Noor, Nurhezreen Md Iqbal, Enizza Kasim, Siti Noraini Othman, Mohd Faizal Abu Bakar, Shamsidar Sopie, Azrin Ahmad, Narcisse Joseph, Muhammad MI, Avisha Richards, Nor Zahrin Hasran, Nor Azfa Johari |
| EPI_ISL_977655, EPI_ISL_977659 | Caribbean Public Health Agency | Carrington Lab, Department of PreClinical Sciences, Building 36, First Floor Biochemistry Unit, Faculty of Medical Sciences, The University of the West Indies | Nikita S. D. Sahadeo, Arianne Brown-Jordan, Vernie Ramkissoon, Sarah Hill, Naresh Nandram, Avery Hinds, Kenneth George, Jerome Foster, Stanley Giddings, Karla Georges, Marsha Ivey, Rahul Naidu, Risha Singh, SueMin Nathaniel, Rajini Haraksingh, Jaya Jayaraman, Chinna Chinnadurai, Adesh Ramsubhag, Nuno Faria, Oliver Pybus, Christopher Oura, Gabriel Escobar, Christine V. F. Carrington |
| EPI_ISL_978544 | Centre de Virologie des Maladies infectueuses Tropicales | Functional Genomic Platform UATRS-biology, CNRST | Elmostafa EL FAHIME, Elmostafa BENAISSA, Marouane MELLOUL, Abdelilah LARAQUI, Nadia Touil, Mly Abdelaziz ELALAOUI, Sanaa ALAOUI-Amine, Yassine SEKHSOKH, Farida HILALI, Hemlali Mouhssine, Abderrazzak Rfaki, , Mostafa ELOUENNASS, Khalid ENNIBI, Tahar BAJJOU |
| EPI_ISL_979360 | Microbiological Diagnostic Unit - Public Health Laboratory (MDU-PHL) | MDU-PHL | Seemann T., Sait, M.L., Sherry, N.L. |
| EPI_ISL_979362 | Victorian Infectious Diseases Reference Laboratory (VIDRL) | VIDRL and MDU-PHL | Caly L., Seemann T., Sait, M.L., Druce J., Sherry, N.L. |
| EPI_ISL_979800, EPI_ISL_979893 | National Institute of Infectious Diseases-Prof. Dr. Matei Bals Molecular Diagnostics Laboratory | National Institute of Infectious Diseases-Prof. Dr. Matei Bals Molecular Diagnostics Laboratory | Leontina Banica, Marius Surleac, Corina Casangiu, Petre Milu, Andreea Tudor, Simona Paraschiv, Dan Otelea |
| EPI_ISL_981020 | National Public Health Laboratory, National Centre for Infectious Diseases | National Public Health Laboratory, National Centre for Infectious Diseases | Tze Minn Mak, Zhenyang Zhou, Lin Cui, Raymond Tzer Pin Lin |
| EPI_ISL_982930 | MEPHI Aix Marseille University (AMU) | MEPHI Aix Marseille University (AMU) | Anthony LEVASSEUR |
| EPI_ISL_983994 | Wisconsin State Laboratory of Hygiene Communicable Disease Division | Wisconsin State Laboratory of Hygiene Communicable Disease Division | Kelsey R. Florek, Abigail C. Shockey |
| EPI_ISL_985079 | Biorepository and Clinical Virology Laboratory | Ozer Lab | Ramon Lorenzo-Redondo, Adeola A. Fowotade, Ewan C. Omoruyi, Johnson A. Adeniji, Lacy M. Simons, Judd F. Hultquist, Babafemi O. Taiwo, Olubusuyi M. Adewumi, Egon A. Ozer |
| EPI_ISL_985161 | Instituto Nacional de Medicina Genomica | Instituto Nacional de Medicina Genomica | Hidalgo-Miranda A, Mendoza-Vargas A, Reyes-Grajeda JP, Cisneros-Villanueva M, Cedro-Tanda A,Peñaloza-Figueroa F, Herrera-Montalvo LA |
| EPI_ISL_985234, EPI_ISL_985235 | Public Health Virology-Forensic and Scientific Services | Public Health Virology-Forensic and Scientific Services | Son Nguyen et al |
| EPI_ISL_985246 | Delaware Public Health Laboratory (DPHL) | Delaware Public Health Lab | Gregory Hovan |
| EPI_ISL_985395 | National Institute of Infectious Diseases-Prof. Dr. Matei Bals Molecular Diagnostics Laboratory | National Institute of Infectious Diseases-Prof. Dr. Matei Bals Molecular Diagnostics Laboratory | Leontina Banica, Marius Surleac, Corina Casangiu, Petre Milu, Andreea Tudor, Simona Paraschiv, Dan Otelea |
| EPI_ISL_986782 | Lighthouse Lab in Cambridge | Wellcome Sanger Institute for the COVID-19 Genomics UK (COG-UK) Consortium | Rob Howes, The Lighthouse Lab in Cambridge and Alex Alderton, Roberto Amato, Sonia Goncalves, Ewan Harrison, David K. Jackson, Ian Johnston, Dominic Kwiatkowski, Cordelia Langford, John Sillitoe on behalf of the Wellcome Sanger Institute COVID-19 Surveillance Team |

| | | | |
|---|---|---|---|
| | | | (http://www.sanger.ac.uk/covid-team) |
| EPI_ISL_990439 | Lighthouse Lab in Alderley Park | Wellcome Sanger Institute for the COVID-19 Genomics UK (COG-UK) Consortium | Jacquelyn Wynn, Mairead Hyland, The Lighthouse Lab in Alderley Park and Alex Alderton, Roberto Amato, Sonia Goncalves, Ewan Harrison, David K. Jackson, Ian Johnston, Dominic Kwiatkowski, Cordelia Langford, John Sillitoe on behalf of the Wellcome Sanger Institute COVID-19 Surveillance Team |
| EPI_ISL_994665 | "Dr. Andrija Stampar" Teaching Institute of Public Health, Department of Clinical Microbiology | Institute of Applied Genomics | Jasmina Vranes, Michele Morgante, Federica Cattonaro |
| EPI_ISL_995221, EPI_ISL_995248, EPI_ISL_995255, EPI_ISL_995288 | Division of Emerging Infectious Diseases, Bureau of Infectious Diseases Diagnosis Control, Korea Disease Control and Prevention Agency | Division of Emerging Infectious Diseases, Bureau of Infectious Diseases Diagnosis Control, Korea Disease Control and Prevention Agency | Ae Kyung Park, Il-Hwan Kim, Heui Man Kim, Jeong-Min Kim, Namjoo Lee, Chae Young Lee, Sang Hee Woo, Eun-Jin Kim |
| EPI_ISL_995302 | National Public Health Laboratory, National Centre for Infectious Diseases | National Public Health Laboratory, National Centre for Infectious Diseases | Tze Minn Mak, Zhenyang Zhou, Lin Cui, Raymond Tzer Pin Lin |
| EPI_ISL_995599, EPI_ISL_995606 | KU Leuven, Rega Institute, Clinical and Epidemiological Virology | KU Leuven, Rega Institute, Clinical and Epidemiological Virology | Tony Wawina-Bokalanga, Bert Vanmechelen, Joan Marti-Carerras, Piet Maes |
| EPI_ISL_995787 | Division of Emerging Infectious Diseases, Bureau of Infectious Diseases Diagnosis Control, Korea Disease Control and Prevention Agency | Division of Emerging Infectious Diseases, Bureau of Infectious Diseases Diagnosis Control, Korea Disease Control and Prevention Agency | Ae Kyung Park, Il-Hwan Kim, Heui Man Kim, Jeong-Min Kim, Namjoo Lee, Chae Young Lee, Sang Hee Woo, Eun-Jin Kim |
